# Variants in the SOX9 transactivation middle domain induce axial skeleton dysplasia and scoliosis

**DOI:** 10.1101/2023.05.29.23290174

**Authors:** Lianlei Wang, Zhaoyang Liu, Sen Zhao, Kexin Xu, Valeria Aceves, Cheng Qiu, Benjamin Troutwine, Lian Liu, Samuel Ma, Yuchen Niu, Shengru Wang, Suomao Yuan, Xiaoxin Li, Lina Zhao, Xinyu Liu, Zhihong Wu, Terry Jianguo Zhang, Ryan S. Gray, Nan Wu

**Affiliations:** Department of Orthopedic Surgery, State Key Laboratory of Complex Severe and Rare Diseases, Peking Union Medical College Hospital, Peking Union Medical College, and Chinese Academy of Medical Sciences, Beijing, China; Department of Orthopedic Surgery, Qilu Hospital of Shandong University, Jinan, Shandong, China; Beijing Key Laboratory for Genetic Research of Skeletal Deformity, Beijing, China; Center for Craniofacial Molecular Biology, Herman Ostrow School of Dentistry, University of Southern California, Los Angeles, CA, USA; Department of Nutritional Sciences, Dell Pediatric Research Institute, University of Texas at Austin, Austin, TX, USA; Department of Nutritional Sciences, Dell Pediatric Research Institute, The University of Texas at Austin, Dell Medical School, Austin, TX, USA; Key laboratory of big data for spinal deformities, Chinese Academy of Medical Sciences, Beijing, China; Medical Research Center, Peking Union Medical College Hospital, Peking Union Medical College, and Chinese Academy of Medical Sciences, Beijing, China

**Keywords:** *SOX9* (*SRY-Box 9*), Campomelic Dysplasia, Skeletal Dysplasia, Congenital Vertebral Malformations, Scoliosis, Protein Stability

## Abstract

SOX9 is an essential transcriptional regulator of cartilage development and homeostasis. In humans, dysregulation of *SOX9* is associated with a wide spectrum of skeletal disorders, including campomelic and acampomelic dysplasia, and scoliosis. The mechanism of how *SOX9* variants contribute to the spectrum of axial skeletal disorders is not well understood. Here, we report four novel pathogenic variants of *SOX9* identified in a large cohort of patients with congenital vertebral malformations. Three of these heterozygous variants are in the HMG and DIM domains, and for the first time, we report a pathogenic variant within the transactivation middle (TAM) domain of *SOX9*. Probands with these variants exhibit variable skeletal dysplasia, ranging from isolated vertebral malformation to acampomelic dysplasia. We also generated a *Sox9* hypomorphic mutant mouse model bearing a microdeletion within the TAM domain (*Sox9^Asp272del^*). We demonstrated that disturbance of the TAM domain with missense mutation or microdeletion results in reduced protein stability but does not affect the transcriptional activity of SOX9. Homozygous *Sox9^Asp272del^* mice exhibited axial skeletal dysplasia including kinked tails, ribcage anomalies, and scoliosis, recapitulating phenotypes observed in human, while heterozygous mutants display a milder phenotype. Analysis of primary chondrocytes and the intervertebral discs in *Sox9^Asp272del^* mutant mice revealed dysregulation of a panel of genes with major contributions of the extracellular matrix, angiogenesis, and ossification-related processes. In summary, our work identified the first pathologic variant of *SOX9* within the TAM domain and demonstrated that this variant is associated with reduced SOX9 protein stability. Our finding suggests that reduced SOX9 stability caused by variants in the TAM domain may be responsible for the milder forms of axial skeleton dysplasia in humans.

## INTRODUCTION

*SRY-Box 9* (*SOX9*) encodes a transcriptional regulator essential for the development and homeostasis of cartilaginous tissues (1). SOX9 is required to secure the commitment of skeletogenic progenitor cells to the chondrocyte lineage and direct the formation of multiple elements of the axial and appendicular skeleton (2). Ablation of *Sox9* in mesenchymal progenitors of the limb bud or during the early stages of chondrogenesis results in a complete failure of cartilage and bone generation or severe chondrodysplasia (3). SOX9 is also required for axial skeletogenesis. Deletion of *Sox9* in the notochord prevents the formation of its cartilage-rich perinotochordal sheath and results in notochordal cell death (4). Conditional inactivation of *Sox9* in the *Scx*+/*Sox9*+ cell populations cause severe hypoplasia of the ribcage and sternum and defective formation of the vertebral bodies and intervertebral discs of the spine (5). Recent studies also demonstrate that *SOX9* plays a crucial role in maintaining the homeostasis of postnatal intervertebral discs (6, 7). These studies affirm that *SOX9* is critical in vertebral column development and maintenance.

*SOX9* is featured with a highly conserved SRY-related high-mobility-group-box (HMGbox) DNA-binding domain, which is found in all SOX family proteins that act as transcriptional regulators in cell fate determination and differentiation in a variety of lineages (2, 8). Two molecules of SOX9 can form homodimer via the self-dimerization domain (DIM) upon binding in the minor groove of a DNA sequence composed of two SOX recognition motifs, oriented head-to-head and separated by 3-4 nucleotides (9). In addition to the N-terminal HMG and DIM domains, SOX9 is also composed of three additional domains: the transactivation domain in the middle of the protein (TAM), the transactivation domain at the C-terminal (TAC), and the PQA (Proline, Glutamine, and Alanine-rich) domain (8). In vitro analyses show that TAM and TAC domains can work independently and synergistically to control the transactivation of SOX9 (8). PQA domain has no major role in SOX9 transactivation but may help mediate SOX9 activity in specific contexts (8, 10). The molecular functions of these C-terminal domains are proposed to provide synergistic binding of transcriptional co-activators and basal transcriptional machinery (8). For instance, the CREB (cAMP response element-binding protein) transcription factor can be co-immunoprecipitated using the C-terminal transactivation domains but not with the N-terminal domains of SOX9 (11).

Variants in *SOX9* are associated with a range of skeletal deformities including campomelic dysplasia (CMPD, OMIM 114290), acampomelic dysplasia (acampomelic CMPD), and scoliosis (1, 12–15). Heterozygous point mutations (missense, nonsense, frameshift, and splice site) in *SOX9* cause CMPD, characterized by congenital shortening and bending of long bones, vertebral malformation, rib anomalies, dysplasia of scapula, and dysmorphic facial features (14, 16–22). Heterozygous *Sox9* knockout mice phenocopy most of the skeletal abnormalities of this syndrome (23). Though it is generally believed that haploinsufficiency for *SOX9* can underlie CMPD, a recent study implicates that combined haploinsufficiency/hypomorphic and dominant-negative mechanisms can apply for some *SOX9* CMPD mutations in a target gene and cell context-dependent manner (24). Roughly 10% of CMPD patients who do not display bending of long bones are subclassified as acampomelic CMPD (25, 26). Many of the *SOX9* variants causing CMPD are located in DIM and HMG domains, which are essential for the dimerization and DNA binding properties of the SOX9 protein (19, 27) and SOX9-dependent regulation of genes such as *Col2a1* (28, 29) and *Col11a2* (30). These patients usually exhibit severe skeletal abnormalities, including deformities of the long tubular bones, and some of them may present with sex reversal. CMPD can also arise from variants in the TAC and PQA domains of SOX9 (31, 32). Frameshift and nonsense mutations reported in these domains usually result in a truncated SOX9 protein and severe CMPD and sex reversal (32, 33). On the other hand, patients with acampomelic CMPD are more likely to have missense variants, genomic rearrangement with breakpoints upstream of *SOX9*, or a *SOX9* upstream deletion (34). We have previously identified a recurrent missense variant at the prolines/glutamines/serines (PQS) domain (now called TAC domain) of *SOX9* in patients with congenital vertebral malformations but without other systematic deformities (35), which is one of the first non-truncating variants reported in the TAC domain that is associated with a milder form of axial skeleton dysplasia. No missense or nonsense variant in the TAM (previously called K2) domain has been reported so far (36, 37). These observations suggest that locations of variants in different domains of SOX9 might underline a spectrum of skeletal dysplasia from CMPD to milder forms that primarily or exclusively affect the axial skeleton.

To this end, we investigated ultra-rare SOX9 variants within a large cohort of congenital vertebral malformations and identified four novel variants of *SOX9*. We report the first pathologic missense variant of *SOX9* within the TAM domain associated with a milder form of axial skeletal dysplasia and demonstrated that variants in the TAM domains result in reduced SOX9 protein stability. We also generated a *Sox9* hypomorphic mouse model with in-frame microdeletion in the TAM domain which recapitulates some of the skeletal phenotypes observed in the human patient. This work enhances our understanding of SOX9 function in the axial skeleton and provides new insights into the roles of different *SOX9* domains in the spectrum of skeletal dysplasia.

## RESULTS

### Rare and *de novo SOX9* variants identified in individuals with various severity of skeletal dysplasia

We investigated ultra-rare *SOX9* variants with a cohort of 424 patients of congenital vertebral malformations, which consists of a broad spectrum of syndromic and non-syndromic vertebral deformities. Four novel heterozygous missense variants were identified from four probands, including three *de novo* variants (c.218T>C, p.Ile73Thr; c.337A>C, p.Met113Leu and c.826G>T, p.Gly276Cys) and one variant (c.398C>G, p.Ala133Gly) with unknown inheritance (Figure 1, A and B, and Table 1). The clinical and genetic features of these probands are summarized in Table 1. No pathogenic variants in other genes were identified. Family history assessments of the probands show that their parents and siblings are all uneventful.

**Figure 1.**
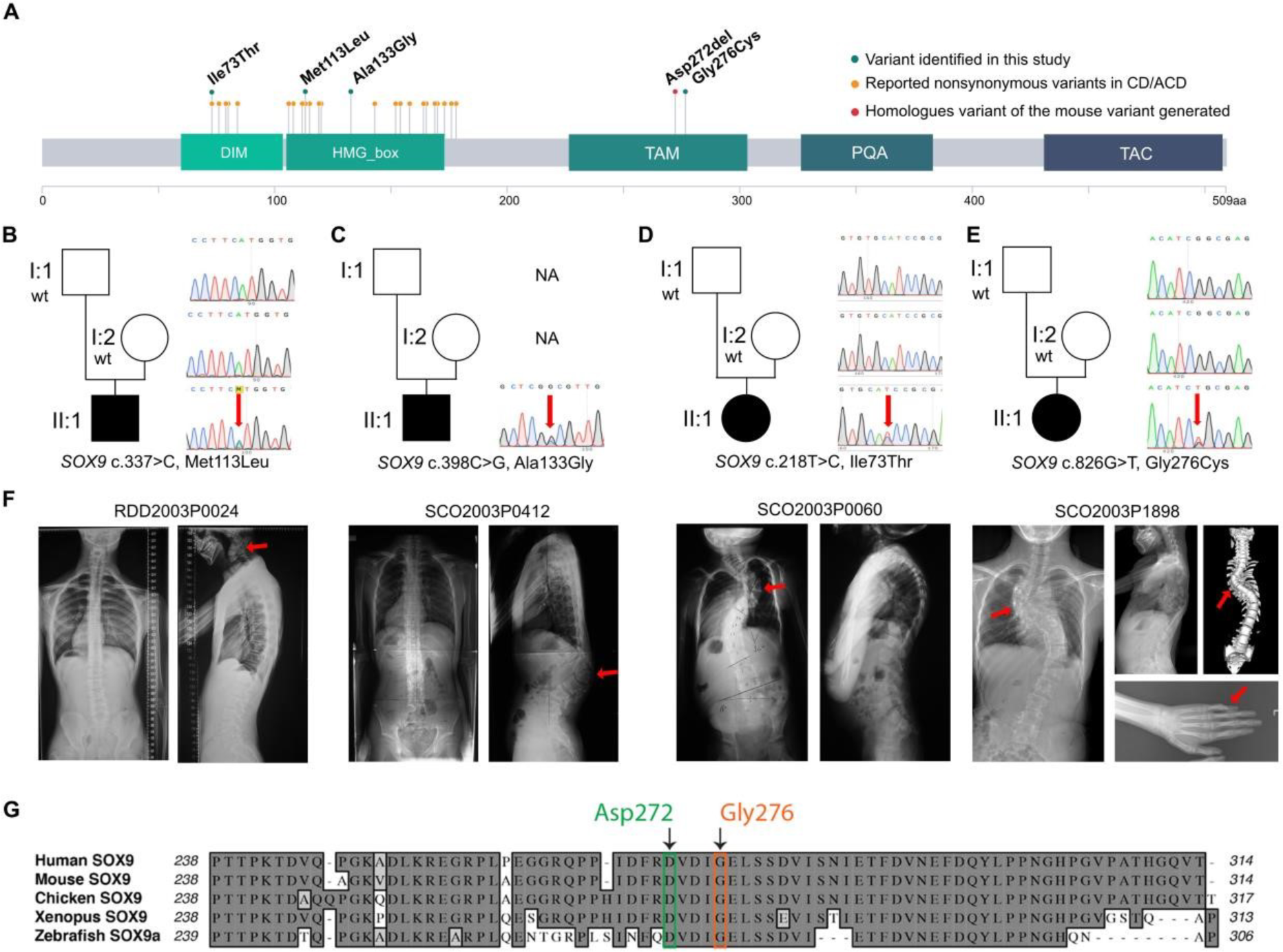
Identification of *de novo* or sporadic *SOX9* variants in four individuals with skeletal dysplasia. (**A**) A schematic presentation of the SOX9 protein. *SOX9* variants identified in this study (green dots), previous studies (orange dots), and the homologs of the mouse variant generated in this study (the red dot) are labeled. (**B-E**) Sanger sequencing results of *de novo SOX9* variants in three trio families and one sporadic case. (**F**) Clinical presentation of the four individuals carrying the *SOX9* variants. Red arrows indicate skeletal malformations. G. Both the Asp272 and Gly276 variants are highly conserved residues. *CMPD: Campomelic Dysplasia; acampomelic CMPD: Acampomelic Dysplasia; DIM: dimerization domain; HMG: high mobility group domain; TAM: transactivation middle, PQA: proline, glutamine, and alanine-rich domain; TAC: transactivation C-terminal.*

**Table 1.** Clinical and genetic features of the four probands

| ID |  | RDD2003P0397 | SCO2003P0412 | SCO2003P0060 | SCO2003P1898 |
| --- | --- | --- | --- | --- | --- |
| Variant information |  |  |  |  |  |
| HGVS |  | c.337A>C | c.398C>G | c.218T>C | c.826G>T |
|  |  | (p.Met113Leu) | (p.Ala133Gly) | (p.Ile73Thr) | (p.Gly276Cys) |
| Origin |  | *de novo* | Unknown | *de novo* | *de novo* |
| Domain |  | HMG | HMG | DIM | TAM |
| CADD |  | 27.3 | 26.5 | 24.1 | 29.6 |
| Skeleton, joints and limbs |  |  |  |  |  |
| Scoliosis |  | - | + | + | + |
| Kyphosis |  | + | - | + | - |
| Lordosis |  | + | - | - | - |
| Vertebral malformation |  | Fusion of cervical vertebrae | L2 wedged vertebra | T6 wedged vertebra | Fusion of thoracic vertebrae |
| Other skeletal deformity |  | Hypoplastic scapulae | Brachydactyly | Hypoplastic scapulae, Short stature | Spinal bifida occulta, Clinodactyly |

*Abbreviations: DIM: dimerization domain; HMG: high mobility group domain; TAM:* *transactivation middle domain.*

Two of the four probands RDD2003P0397 (Figure 1B) and SCO2003P0412 (Figure 1C) exhibited deformities consistent with the clinical manifestations of acampomelic CMPD. RDD2003P0397 presented congenital vertebral malformation caused by the fusion of the cervical vertebra (Figure 1B). Exome sequencing identified a *de novo* heterozygous missense variant c.337A>C (p.Met113Leu) located within the HMG-box of *SOX9* (Figure 1A). Two missense variants (Met113Thr and Met113Val) that affect the same amino acid have been reported previously in a CMPD patient (Met113Thr) and an acampomelic CMPD patient (Met113Val), respectively (38, 39). SCO2003P0412 presented congenital kyphosis (Figure 1C). He had a heterozygous missense variant (c.398C>G, p.Ala133Gly) within the HMG-box of *SOX9* with an unknown inheritance origin (Figure 1C). No pathogenic variant of this amino acid has been reported previously. Notably, Ala133Gly is predicted to be highly deleterious (combined annotation dependent depletion [CADD] score = 24.1).

In contrast to these two cases diagnosed as acampomelic CMPD, the third proband SCO2003P0060 presented a milder skeletal dysplasia phenotype characterized by a wedged vertebra at T6 and thoracic scoliosis (Figure 1D). She also presented with short stature and kyphoscoliosis (Table 1). She carried a *de novo* variant c.218T>C (Ile73Thr) located in the DIM domain of *SOX9* (Figure 1A). A variant affecting the same amino acid (Ile73Asn) has been reported previously in a patient with multiple congenital anomalies (40). This c.218T>C (Ile73Thr) variant has not been included in the gnomAD (v2.1.1, https://gnomad.broadinstitute.org, accessed on January 5, 2023), and it is classified as likely pathogenic (PM2_Supporting + PS3_Supporting + PS2 + PM5_Supporting) according to the ACMG guideline (41).

In the fourth proband (SCO2003P1898), we identified a *de novo* variant c.826G>T (Gly276Cys) located at the TAM domain (8) of *SOX9* (Figure 1A). This Gly276 residual is highly conserved among vertebrate orthologues (Figure 1G). To our knowledge, this is the first missense mutation reported within the TAM domain in humans. This proband exhibited fused vertebrae from T6-T11 and spinal bifida occulta of L4-L5 (Figure 1E). She also exhibited thoracic scoliosis and clinodactyly of the distal 5^th^ phalange (Figure 1E). Interestingly, the multiple fusion of thoracic vertebrae phenotype in this proband was distinct from the other three patients and the classical CMPD/ acampomelic CMPD patients, suggesting that variants in the TAM domain of *SOX9* may be associated with a distinct type of skeletal anomalies.

To further characterize the impact of *SOX9* variants, we modeled the potential structural effects of these mutations predicted by Alpha Fold (Supplemental Figure 1, https://alphafold.ebi.ac.uk) (42). The evolutionally conserved *SOX9* Ile73, Met113, and Ala133 residues are predicted to help stabilize alpha-helical regions (Supplemental Figure 1, B-D) within the regions with high confidence of prediction or within areas where structural information was empirically determined (unpublished PDB data: 4EUW and 4S2Q). The highly conserved *SOX9* Gly276 residue (Fig. 1G) is predicted to hydrogen bond interact with Asp274 (Supplemental Figure 1E) in a putative helical region of the protein. Altogether, the *in silico* evidence showed that the affected residues are highly conserved among vertebrate orthologues, and alteration of these residues may lead to compromised protein structure and altered interactions with other residues, which may jeopardize the function and structural stability of the SOX9 protein.

### Effects of the *SOX9* variants on DNA-binding and transcriptional activities of SOX9

Next, we sought to understand how *SOX9* variants affect the DNA-binding and transcriptional activities of the SOX9 protein using electrophoretic mobility shift assay (EMSA) and luciferase reporter assay. We synthesized two sets of EMSA probes derived from the enhancer region of the mouse *Col11a2* gene (30). Each probe contains a canonical SOX9 DNA-binding motif and was named *Col11a2* B/C and *Col11a2* D/E respectively. An equal amount of SOX9 wild-type and variants proteins were incubated with these probe sets for EMSA analyses (Supplemental Figure 2). We discovered that both *SOX9* variants within the HMG-box (Met113Leu and Ala133Gly) display impaired DNA-binding capability characterized by decreased probe binding signal (Figure 2A, black arrows). The *SOX9* variant within the DIM domain (Ile73Thr), which is an essential domain for the dimerization of SOX9 proteins (43), showed reduced DNA-binding capability and loss of dimerization (Figure 2A, black arrows, and red arrows). To define if these variants affected SOX9-dependent gene transcription, we co-transfected various SOX9-expression vectors with a luciferase reporter construct (named *Col11a2*-Luc) which contains 4 tandem copies of the canonical SOX9 DNA-binding motif *Col11*a2 D/E (Supplemental Figure 3 and Supplemental Table 2) (30, 44). We showed that both the HMG-box variants (Met113Leu and Ala133Gly) and the DIM domain variant (Ile73Thr) exhibited decreased transcriptional activity of SOX9 (Figure 2B). In contrast, the variant in the TAM domain (Gly276Cys) did not obviously compromise the transcriptional activity of SOX9 compared with the wild-type protein but showed significantly higher transcriptional activity compared with the Ile73Thr, Met113Leu, and Ala133Gly variants (Figure 2B). Taken together, these data suggest that the variants in HMG and DIM domains are likely affecting the DNA-binding capabilities and homotypic interactions of SOX9, which may lead to compromised SOX9-dependent transcriptional activities. In contrast, variants in the TAM domain may still allow for some transcriptional activity at the canonical SOX9 targets thus permitting partial SOX9-dependent function.

**Figure 2.**
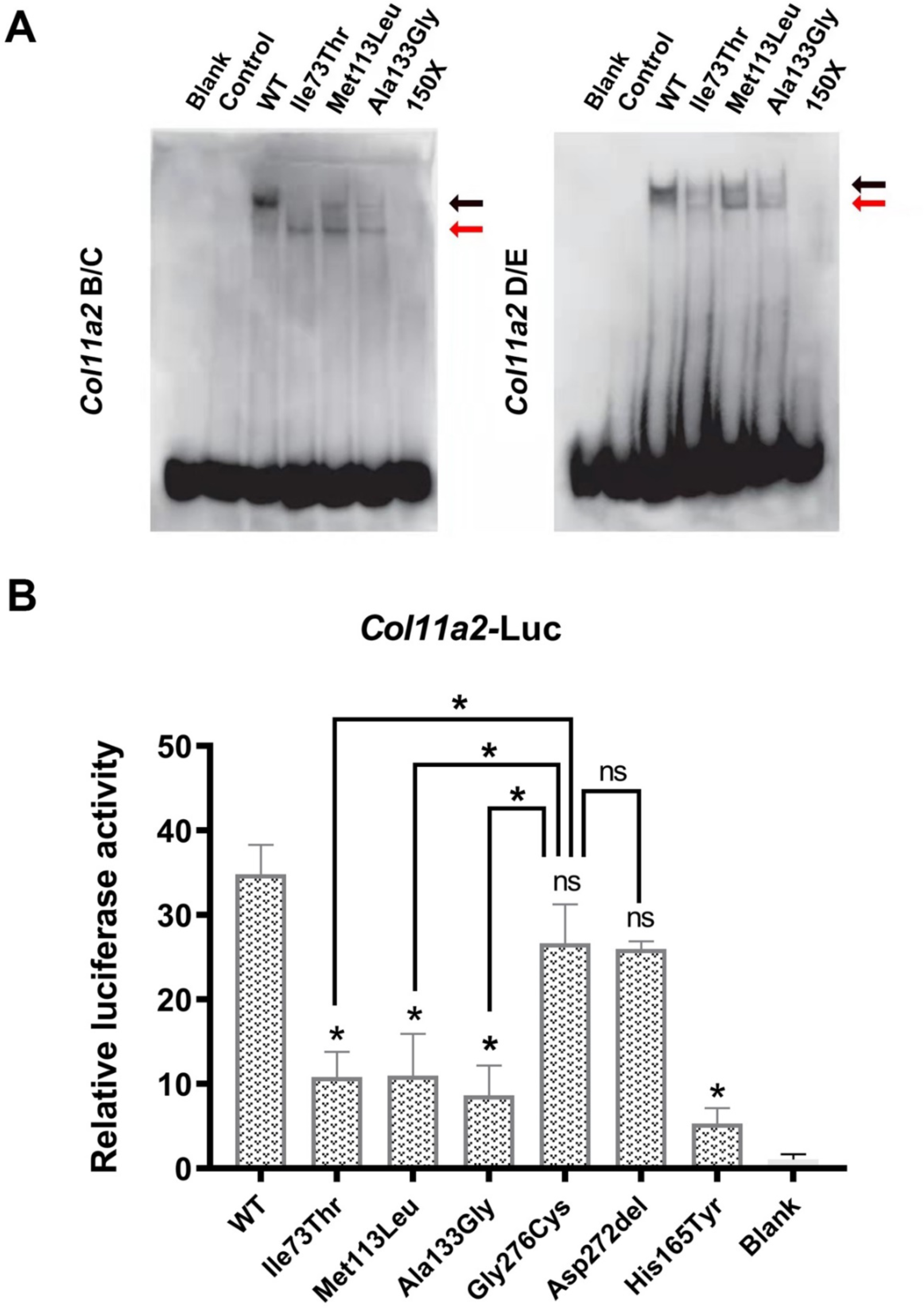
Effects of the *SOX9* variants on DNA-binding and transcriptional activities of SOX9. **(A)** Electrophoretic mobility shift assay (EMSA) was performed on the whole cell lysate extracted from Cos-7 cells. Cell lysate with no transfection was used as a blank control (Blank). Cos-7 cells transfected with a GFP control construct were used as a mock control (Control). The 150-fold unlabeled DNA probes (150X) were incubated with cell lysate of Cos-7 cells transfected with wild-type SOX9 protein. Free probes were detected at the bottom of the gel. The bound dimer and monomer SOX9 proteins are indicated with black and red arrows, respectively. **(B)** Luciferase assay was performed on wild-type (WT) SOX9 protein and various SOX9 variant mutant proteins using a *Col11a2*-Luc construct. His165Tyr variant protein was used as a negative control (31). *The statistical difference is evaluated by one-way ANOVA followed by Tukey’s multiple comparison test. The data are the mean of n = 3 independent experiments. Error bars show one standard deviation. * p<0.05 ** p<0.01 ns: not significant.*

### A TAM domain mutation in mice leads to dose-dependent reductions of SOX9 expression and tail kinking during perinatal development

To further our understanding of the role of the TAM domain variant in skeletal dysplasia, we set out to use a CRISPR-Cas9 gene-editing approach to knock in the human variant Gly276Cys in the mouse (Figure 1E). After multiple injections, we isolated only three CRISPR-injected mice as adults (0.3%; n=123 implanted), likely due to haploinsufficient lethality of *Sox9* in the mouse (23). Serendipitously, we obtained an in-frame microdeletion allele (*Sox9^em1Rgray^*), which deleted Asp272 (Asp272del), four amino acids upstream of Gly276Cys within the TAM domain (Figure 1A, Figure 3, A and N). Like the Gly276 residual, the Asp272 residual is also highly conserved among vertebrate orthologues (Figure 1G) and is predicted to hydrogen bond interact with Asp274 by Alpha Fold structural prediction (Supplemental Figure 1E). Intercross of these *Sox9^Asp272del/+^* mice (hereafter called *Sox9^del^*) generated *Sox9^del/del^* mutant mice at Mendelian ratios that are indistinguishable from the *Sox9^del/+^* or wild-type (*Sox9^+/+^*) littermates at birth. However, by postnatal (P) day 5 *Sox9^del/del^* mutant mice exhibited a fully penetrant kinked tail phenotype (100%, n=12) (Figure 3D, red arrow). In contrast, heterozygous *Sox9^del/+^* and wild-type littermates uniformly displayed normal tails (Figure 3, B and C).

**Figure 3.**
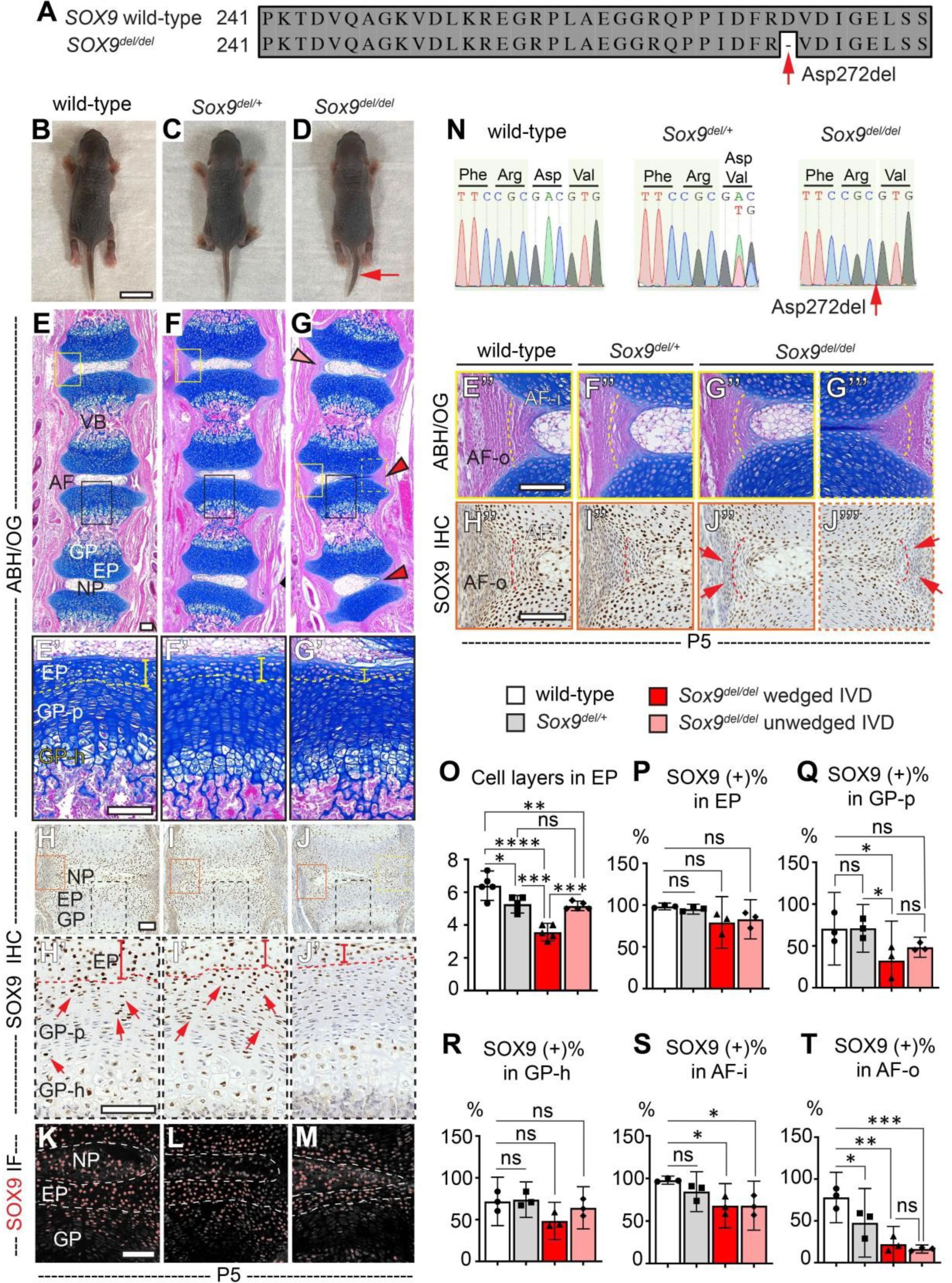
A TAM domain mutation in mice leads to dose-dependent reductions of SOX9 expression and tail kinking during perinatal development. (**A**) Illustration of the in-frame deletion of Asp272 in the *Sox9^del/del^* mutant mouse. The red arrow indicates the Asp272del mutation. (**B-D**) At P5, an obvious kinked tail phenotype was observed in 100% of the *Sox9^del/del^* mutant mice, but not in heterozygous *Sox9^del/+^* or wild-type littermates. (n=12 for *Sox9^del/del^* mice, n=8 for *Sox9^del/+^* and wild-type littermates). (**E-G’’’**) Alcian Blue Hematoxylin /Orange G (ABH/OG) staining performed on P5 mouse tail sections revealed normal segmentation of the vertebral bodies and typical proteoglycan staining of IVD and growth plate in *Sox9^del/del^* mutant mice (**G**) compared with the wildtype (**E**) and *Sox9^del/+^* (**F**) littermates. Red and pink arrowheads indicate wedged and unwedged IVDs in *Sox9^del/del^*mutant mouse, respectively. Higher magnification images of the endplate and growth plate and the annulus fibrosus are shown in (**E’-G’**) and (**E’’-G’’’**). The endplate is indicated with a dotted line and a vertical bar in (**E’G’**). The boundary between inner and outer annulus fibrosus is indicated with a dotted line in (**E’’-G’’’**). (n=4 for each genotype). (**H-J’’**) Immunohistochemistry (IHC) analyses of SOX9 performed on P5 mouse tail sections. Higher magnification images of the endplate and growth plate and the annulus fibrosus are shown in (**H’-J’**) and (**H’’-J’’’**). The endplate is indicated with a dotted line and a vertical bar in (**H’-J’**). SOX9 expression in proliferative growth plates is indicated with red arrows in (**H’**) and (**I’**). The boundary between inner and outer annulus fibrosus is indicted with a dot-line in (**H’’-J’’’**). The absence of SOX9 expression in the outer annulus fibrosus is indicated with red arrows in (**J’’**) and (**J’’’**). (n=4 for each genotype.) (**K-M**) Immunofluorescence (IF) analyses of SOX9 were performed on P5 mouse tail sections. (n=6 for each genotype.) (**N**) Sanger sequencing results of wild-type, *Sox9^del/+,^* and *Sox9^del/del^* mice. The red arrow indicates the Asp272del mutation. (**O-T**) Quantification of the number of cell layers in endplate (**O**), and quantification of the percentile of SOX9 (+) cells in endplate (**P**), proliferative growth plate (**Q**), hypertrophic growth plate (**R**), inner annulus fibrosus (**S**), and outer annulus fibrosus (**T**) in three genotypes of mice. Bars are plotted with mean and 95%CI. Each dot represents one mouse analyzed. The statistical difference is evaluated by one-way ANOVA followed by Tukey’s multiple comparison test. * p<0.05, **: p<0.01, ***p<0.001. ns: not significant. For (**O**), n=5 for each genotype; for (**PT**), n=3 for each genotype. *Scale bars: 10mm in **B**; 100μm in **E-E’’’**, **H-H’’’** and **K**. VB: vertebral body; AF: annulus fibrosus; EP: endplate; GP: growth plate; NP: nucleus pulposus; GP-p: proliferative growth plate; GPh: hypertrophic growth plate; AF-i: inner annulus fibrosus; AF-o: outer annulus fibrosus*

Many congenital vertebral malformations are due to patterning defects of embryonic somites which contribute to the formation of vertebral bodies and intervertebral discs (IVDs) (45). Histological analysis of *Sox9^del/del^* mutant mouse tails at P5 revealed normal segmentation and ossification of the vertebral bodies and typical proteoglycan staining in the IVDs and growth plates (Figure 3, E-G). However, the *Sox9^del/del^* mice showed wedging of the IVDs in regions of tail kinking, a shift of the nucleus pulposus toward the convex side (Figure 3G, red arrowheads), and compression of annulus fibrosus on the concave side of the curvature (Figure 3G’’’). In contrast, unwedged IVDs adjacent to severe curvatures could display comparably normal structures (Figure 3G, pink arrowhead, and Supplemental Figure 4A). Detailed analysis of the tail sections revealed that the thickness of the cartilaginous endplate of the IVDs is reduced in *Sox9^del/+^* and the *Sox9^del/del^* mutant mice compared with that of the wild-type mice (Figure 3, E’ and G’, yellow bars) quantified by the number of cell layers within the endplate (Figure 3O). The number of endplate cell layers in the unwedged IVDs of the *Sox9^del/del^* mutant mice is slightly increased compared with that of the wedged IVDs but is still reduced in comparison with the wild-type mice (Figure 3O and Supplemental Figure 4A’).

We next sought to determine how SOX9 expression was affected in *Sox9^del/del^*mutant tissues. Immunohistochemistry (IHC) and immunofluorescence (IF) analyses of P5 tail sections revealed that SOX9 expression is regionally reduced in the *Sox9^del/+^*and the *Sox9^del/del^* mutant mice (Figure 3, H-J’ and K-M). We observed a decreased SOX9 expression in the proliferative growth plate adjacent to the wedged IVDs but not the unwedged IVDs of the *Sox9^del/del^* mutant mice (Figure 3, H’-J’ and Q, and Supplemental Figure 4, B-B’). No difference in SOX9 expression was observed in the hypertrophic growth plate (Figure 3, H’-J’ and R) nor the endplate (Figure 3P) among the three genotypes. We also observed reduced expression of SOX9 in both inner and outer layers of the annulus fibrosus of the *Sox9^del/del^* mutant mice in both wedged and unwedged IVDs (Figure 3, H’’-H’’’ and 3S-T, and Supplemental Figure 4B’’), with more pronounced effects on the outer layers (Figure 3, J’’-J’’’, red arrows; and Figure 3T), as well as decreased *SOX9* expression in the outer annulus fibrous of the *Sox9^del/+^*mice (Figure 3T). IHC analyses of type II collagen (COLII) and type X collagen (COLX) showed a comparable pattern of staining at this age among all three genotypes (Supplemental Figure 4, C-H). All these results suggest that the mutation in the TAM domain leads to a dose-dependent mild reduction of SOX9 expression in various compartments of the spinal tissues, associated with a tail kinking phenotype during perinatal development. However, the overall patterning and segmentation of the vertebral bodies are not affected.

### *Sox9^Asp272del^* is a hypomorphic loss-of-function allele

The tail kinking phenotype and the reduced SOX9 expression in the *Sox9^del/+^* and the *Sox9^del/del^* mutant mice are dose-dependent and region-specific, suggesting that the Asp272del microdeletion was behaving as a mild hypomorphic allele of *Sox9*. To test the nature of the *Sox9^del^* allele, we assessed how skeletal development would be affected when the *Sox9^del^* allele was *in trans* with an established floxed null allele of *Sox9* (*Sox9^flox^*) (3). Previous studies show that completely inactivating *Sox9* in osteochondral progenitors using the *Col2a1-Cre* deleter strain (*Col2a1-Cre;Sox9^flox/flox^*) resulted in embryonic lethality and severe chondrodysplasia in mutant embryos, while the heterozygous recombination of *Sox9* (*Col2a1-Cre;Sox9^flox/+^*) caused perinatal lethality around P10 displaying dwarfism and kyphosis of the axial skeleton (3). Given these findings, we set out to assess skeletogenesis of the *Col2a1-Cre;Sox9^flox/del^*compound heterozygous mutants in comparison with the *Col2a1-Cre;Sox9^flox/+^*heterozygous mutants at P1.

The *Col2a1-Cre;Sox9^flox/del^* compound heterozygous mice were born at Mendelian ratio (n=8; total 67 pups) with a 63% survival rate at P1. Skeletal preparations showed increased defects of craniofacial and ribcage development and bending of the long bone in the *Col2a1-Cre;Sox9^flox/del^* compound heterozygous mutants compared with the *Col2a1-Cre;Sox9^flox/+^* heterozygous littermates (Supplemental Figure 5, B and C). To better quantify skeletal phenotypes, we analyzed the skull morphology with MicroCT imaging (Figure 4 and Supplemental Figure 5, D-F). We observed decreased skull size in *Col2a1-Cre;Sox9^flox/del^*compound heterozygous mutants (Figure 4, C-D) compared with Cre (-) controls (Figure 4, A and A’) and *Col2a1-Cre;Sox9^flox/+^* heterozygous mutants (Figure 4, B and B’). However, we observed no significant change in the percentage of bone volume when normalized to total tissue volume (BV/TV) (Figure 4E), which is consistent with the previous report that *Sox9* deletion does not obviously affect osteoblast differentiation (3). Taken together, these genetic studies provide functional evidence that the *Sox9^del^* allele, which disturbs the TAM domain of *SOX9*, is acting as a mild hypomorphic allele.

**Figure 4.**
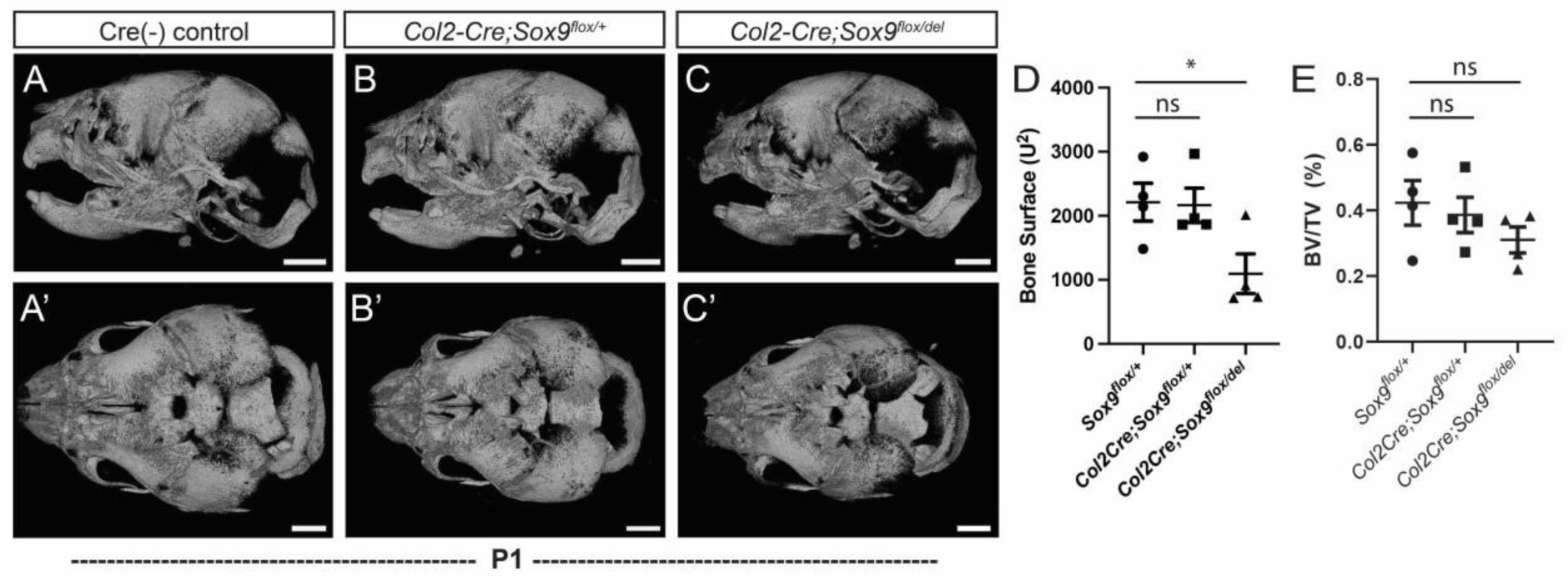
*Sox9^Asp272del^* is a hypomorphic loss-of-function allele. (**A-C’**), 3D MicroCT renderings of mouse skulls from Cre(-) control (**A, A’**), *Col2a1-Cre;Sox9^flox/+^* (**B, B’**), and *Col2a1-Cre;Sox9^flox/del^* mutant (**C, C’**) mice at P1 in lateral (**A-C**) or dorsal views (**A’-C’**). (**D**) Quantification of bone surface area (µm^2^) of the skull shows a reduced skull size of the *Col2a1-Cre;Sox9^flox/del^* compound heterozygous mutant mice (p= 0.0428) compared to wild-type Cre(-) control mice. (**E**) Quantification of the percentage of bone volume vs tissue volume (BV/TV) shows no loss of bone mass density in the skulls of *Col2a1-Cre;Sox9^flox/del^* mutant mice (p=0.2972) compared to wild-type Cre(-) control mice. Bars are plotted with mean and standard deviation. Each dot represents one mouse analyzed. The statistical difference is evaluated by one-way ANOVA followed by Tukey’s multiple comparison test. n=4 for each genotype. *Scale bars 1mm.*

### Disturbance of the TAM domain of SOX9 results in dose-dependent skeletal dysplasia in young adult mice

By postnatal (P) day 60 (P60), *Sox9^del/del^* mutant mice are adult-viable (Figure 5C). Dorsal X-ray imaging showed normal segmentation and growth of the spine irrespective of genotype (Figure 5, C-D). However, *Sox9^del/del^*mutant mice did display shorter femurs compared with *Sox9^del/+^* and the wild-type mice (Figure 5D). We observed loss of the thoracic (T) 13 floating ribs (100%, n=18; 8 male, 10 female) in *Sox9^del/del^* mice (Figure 5C’, red asterisks; and Figure 4E), with the majority of *Sox9^del/del^* mice (94%) displaying bilateral loss of the T13 floating ribs (Figure 5E). Notably, 16% of heterozygous *Sox9^del/+^* mice (n=25, 15 males, 10 females) also displayed loss of either the right or left T13 floating rib (Figure 5C’, red asterisk; and Figure 5E). In contrast, we never observed the loss of the T13 floating ribs in wild-type mice (n=8, 4 males, 4 females) (Figure 5, A’ and E). Sagittal X-ray imaging showed that 43% of *Sox9^del/del^* mutant mice (n=7, 4 males, 3 females) showed ribcage defects at P60, which was not observed in wildtype (n=4, 2 males, 2 females) or *Sox9^del/+^* mice (n=8, 5 males, 3 females) (Supplemental Figure 6). Altogether, these results show that an in-frame microdeletion within the TAM domain of SOX9 causes spatially restrictive disruptions in the development of the axial and appendicular skeleton, some of which are hallmarks of acampomelic CMPD including loss of ribs, ribcage defects, and shortened long bones (14, 16).

**Figure 5.**
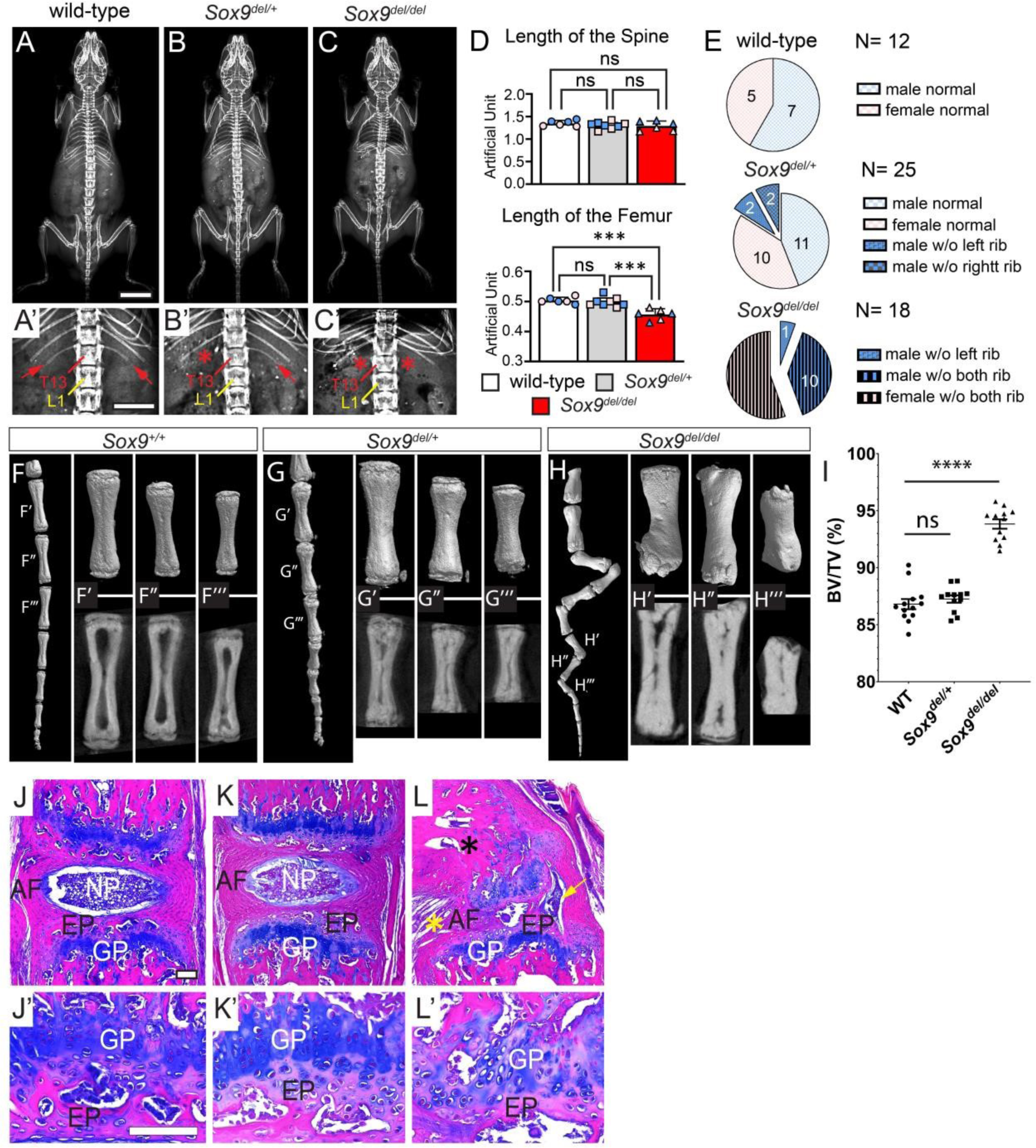
Variants in the TAM domain of SOX9 results in a dose-dependent skeletal dysplasia in adult mouse. (**A-C’**) Dorsal X-ray images of wild-type (**A, A’**), *Sox9^del/+^* (**B, B’**), and *Sox9^del/del^* mutant mice (**C, C’**) at P60. The presence and loss of T13 floating ribs are indicated with red arrows and red asterisks, respectively. (**D**) Measurements of the length of the spine (T1-L6) and the long bone (femur) in P60 mice. Bars are plotted with mean and 95%CI. Each dot represents one mouse analyzed. Male animals are marked with blue dots and female animals are marked with pink dots. The statistical analysis is one-way ANOVA followed by Tukey’s multiple comparison test. ***p<0.001. ns: not significant. (n=6 for wild-type mice; 3 males, 3 females. n=8 for *Sox9^del/+^* mice; 4 males, 4 females. n=6 for *Sox9^del/del^* mice; 3 males, 3 females). (**E**) Quantification of the floating rib phenotype in P60 mice. (n=12 for wild-type mice; 7 males, 5 females. n=25 for *Sox9^del/+^* mice; 15 males, 10 females. n=18 for *Sox9^del/del^* mice; 8 males, 10 females). (**F-H**) MicroCT analyses showing 3D reconstruction of the straight tails in wild-type (**F**) and *Sox9^del/+^* (**G**) mice, but kinked tail in *Sox9^del/del^* mice (**H**) at P60. Demonstrates a dose-dependent increase in attenuating bony matrix deposition in the bone marrow space in the optical sections of *Sox9^del/+^* and *Sox9^del/del^* mutant mice (lower panels: G’-G’’’ and H’-H’’’ respectively). (**I**) Analysis of percent bone volume (BV)/tissue volume (TV) of the 5-7^th^ coccygeal vertebrae counting from the distal-most tip of the tail showing significant increase in BV/TV (%) in *Sox9^del/del^* mutant mice (p=<0.0001) compared with wild-ype control mice. Bars are plotted with mean and SEM. Each dot represents one coccygeal vertebra analyzed. The statistical difference is evaluated by one-way ANOVA followed by Tukey’s multiple comparison test. n=4 mice for each genotype. (**J-L’**) ABH/OG staining performed on tail sections of P60 wild-type (**I, I’**), *Sox9^del/+^* (**J, J’**), and *Sox9^del/del^* (**K, K’**) mice showed severe disruption of IVD in *Sox9^del/del^* mice tails. The yellow arrow indicates nucleus pulposus tissue into the annulus fibrosus layer and a yellow asterisk indicates disorganization of the annulus fibrosus. n=3 for each genotype. *Scale bars: 10mm in **A**; 5mm in **A’**; 2mm in **F**; and 100μm in **I-I’**. AF: annulus fibrosus; EP: endplate; GP: growth plate; NP: nucleus pulposus.*

*Sox9^del/del^* mutant mice also displayed an exacerbated kinked tail phenotype at P60 (Figure 5H). MicroCT imaging showed that tail kinks are associated with dysmorphic coccygeal vertebrae (Figure 5, H-H’’’), including ectopic bone deposition in the marrow space in *Sox9^del/del^* mutant mice, bending of the vertebra, and fusion between vertebrae (Figure 5, H’-H’’’ and I). The bending and fusion of the caudal vertebrae in the *Sox9^del/del^* mice mimicked some phenotypes observed in the human patient with the Gly276Cys mutation (Figure 1E). Histological analysis of these kinked tail regions in *Sox9^del/del^* mice showed severe dysplasia of IVDs and growth plate (Figure 5, L-L’). In some regions, the nucleus pulposus was pushed toward the convex side of the curve, embedded within the annulus fibrosus layers (Figure 5L, yellow arrow). Some *Sox9^del/del^* mice also showed thinning and disruption of IVD endplate, characterized by loss of cellularity and lack of ossification in the endplate (Figure 5L’) compared with the normal structure observed in wild-type and *Sox9^del/+^* mice (Figure 5, J-J’ and K-K’). The vertebral growth plate within the kinked tail region is also disrupted, with disorganized cell columns and reduced proteoglycan staining in some regions of the growth plate (Figure 5L’), consistent with the reported reduction in proteoglycan staining in the IVDs of conditional SOX9 loss-of-function mice (6), further supporting our model that *Sox9^del^* allele behaves as a mild hypomorphic allele. These results demonstrate that a microdeletion variant in the TAM domain of SOX9 is sufficient to cause the progressive onset of skeletal dysplasia characterized by gross histological pathology of the IVDs, bending of the vertebral bodies, and pathological changes of the growth plate.

### Disturbance of the TAM domain of SOX9 leads to late-onset scoliosis in mouse

We consistently observed skeletal dysplasia in the tails of *Sox9^del/del^* mutant mice from P5 to P60 but we never observed obvious dysplasia in the thoracic or lumber spine at this time (Figure 5, A-C). However, when we assessed these mutants at 6 month-of-age, ∼ 43% of *Sox9^del/del^* mutant mice (n=14, 8 males, 6 females) displayed mild lateral curvature of the spine reminiscent of late-onset scoliosis (Figure 6, C and F). The severity of spine curvatures, quantified as Cobb angles (46), ranged from 10.2° to 13.8° (Figure 6I). We observed scoliosis in both male and female *Sox9^del/del^* mutant mice with curvatures in both rightward and leftward directions (Figure 6I). The distribution of the apex of the curvature was enriched in the lower thoracic region (T12-T13) and lumbar region (L1, L4, and L5) (Figure 6J). In comparison, only one female *Sox9^del/+^* mouse (n=15, 12 males, 3 females) showed a mild curve of 10.7° at 6 months (Figure 6H), while the majority of *Sox9^del/+^* mice and all of the wild-type mice (n=8, 4 males and 4 females) had no measurable scoliosis at 6 months (Figure 6G).

**Figure 6.**
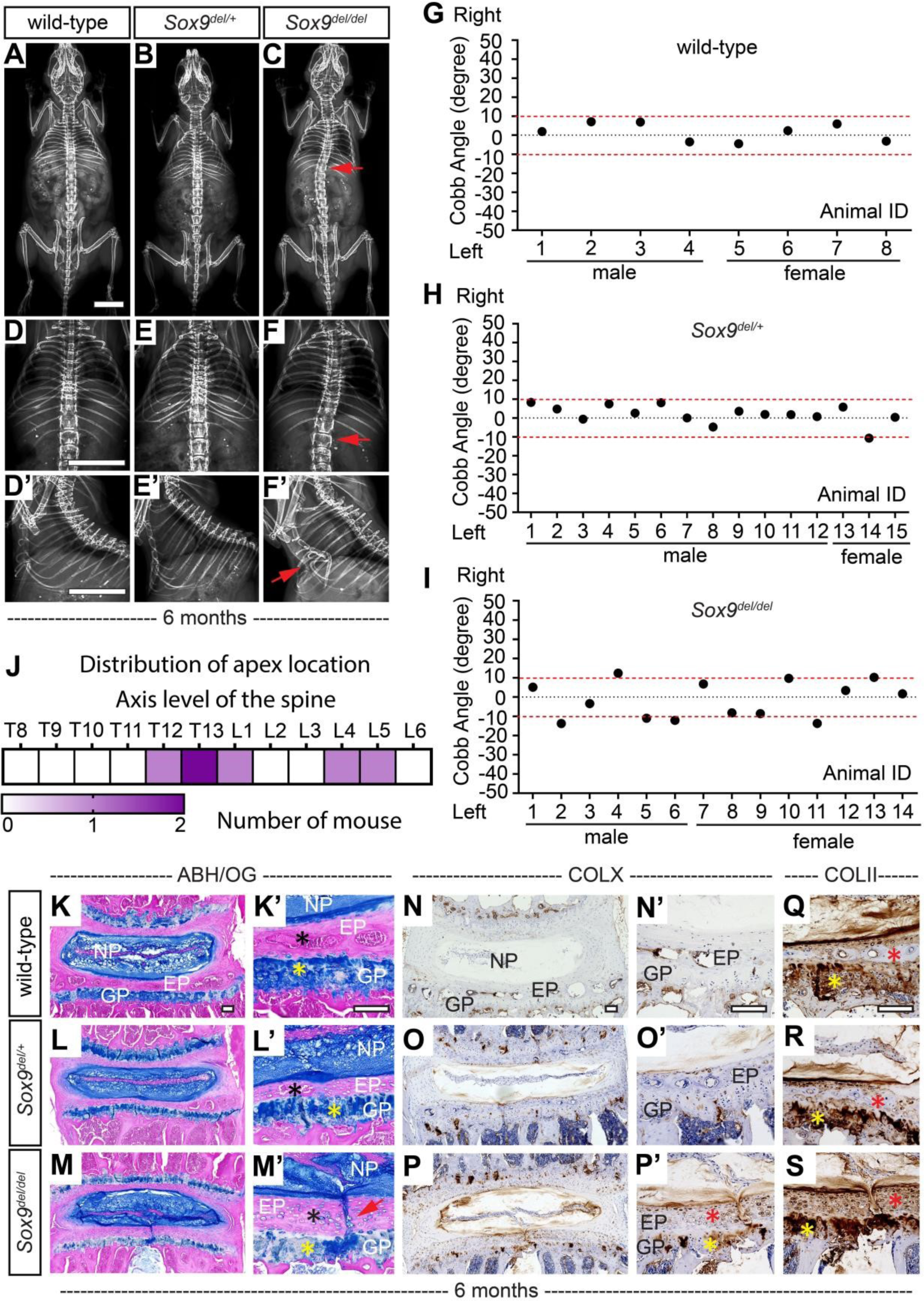
Adult *Sox9^del/del^*mutant mice display IVD defects of the spine. (**A-F’**) Representative dorsal X-ray images of wild-type (**A, D**), *Sox9^del/+^*(**B, E**) and *Sox9^del/del^* mutant mice (**C, F**) at 6 month-of-age. Sagittal X-ray images of the same mouse are shown in (**D’-F’**). Red arrows in (**C**) and (**F**) indicate scoliosis. The red arrow in F’ indicates rib cage deformity. (n=8 for wild-type mice; 4 males, 4 females. n=15 for *Sox9^del/+^* mice; 12 males, 3 females. n=14 for *Sox9^del/del^* mice; 8 males, 6 females.) (**G-I**) Cobb angle measurements of wild-type (**G**), *Sox9^del/+^* (**H**) and *Sox9^del/del^*(**I**) mice at 6 months. Areas of scoliosis (Cobb angle >10 degrees) are indicated with two red dot lines. (**J**) Heat map of the apex distribution of the scoliotic *Sox9^del/del^* mutant mice at 6 months. The heat map is plotted with the axis level of the thoracic spine (T8-L6) and the number of scoliotic mice with apex observed at each level. (n=8 for wild-type mice; 4 males, 4 females. n=15 for *Sox9^del/+^* mice; 12 males, 3 females. n=14 for *Sox9^del/del^*mice; 8 males, 6 females.) (**K-M’**), ABH/OG staining performed on lumbar IVDs (L4/5) of 6-month-old wild-type (**K, K’**), *Sox9^del/+^* (**L, L’**) and *Sox9^del/del^* (**M, M’**) mice. The endplate and growth plate are indicated with black and yellow asterisks, respectively. Endplate clefts observed in *Sox9^del/del^* mice are indicated with a red arrow in (**M’**) (n=3 for each genotype.) (**N-S**), Immunohistochemistry (IHC) analyses of type X collagen (COLX) and type II collagen (COLII) in 6-month-old wild-type (**N**, **N’** and **Q**), *Sox9^del/+^* (**O**, **O’** and **R**) and *Sox9^del/del^* (**P**, **P’** and **S**) mice. Endplate staining and growth plate staining are indicated with red and yellow asterisks, respectively. n=6 for each genotype. *Scale bars: 10mm in **A**, **D**, and **D’**; 100μm in **K-K’**, **N-N’**, and **Q**. EP: endplate; GP: growth plate; NP: nucleus pulposus.*

In addition, we also observed a range of rib cage deformities in 93% of the *Sox9^del/del^* mice at 6 months (n=14, 8 males, 6 females), from obvious dorsal ward bending of the sternums into the rib cage akin to pectus excavatum in humans (47) (Figure 6F’), to milder bending of the sternums (Supplemental Figure 7, C and D) and curvatures of the distal ribs (Supplemental Figure 7E). We only observed 1 male *Sox9^del/+^* mice displaying mild bending of the sternum at 6 months (n=8, 4 males and 4 females) (Supplemental Figure 7B). In contrast, none of the wild-type littermates displayed rib cage deformities (n=8, 4 males and 4 females) (Figure 6D’). The etiology of pectus excavatum is still not clear, but it may be associated with genetic or connective tissue disease that leads to compromised support of the rig cage (48). These results suggest that disturbance of the TAM domain of *SOX9* also affects the stability of the axial skeleton during adult development, modeling late-onset scoliosis and progressive rib cage deformities in a dose-dependent manner, potentially through regulating the homeostasis of cartilaginous and connective tissues in the spine and rib cage.

### Adult *Sox9^del/del^* mutant mice display IVD defects of the spine

SOX9 function is required in adult mice to maintain homeostasis of the cartilaginous tissues (6, 49). To determine if *Sox9^del/del^* mutant mice might accumulate increased pathology of skeletal elements in older mice we looked at 6-month-old cohorts. We observed endplate defects in 50% (n=6) of *Sox9^del/del^* mutant mice (Figure 6, M-M’), characterized by vertical cleft through endplate into growth plate in the lumbar IVDs (Figure 6M’, red arrow), which was not observed in *Sox9^del/+^* or wild-type littermates (n=6 for each genotype) (Figure 6, K-L’). We also observed reduced proteoglycan staining in some growth plates of the lumbar spine in the *Sox9^del/del^*mice (Figure 6M’, yellow asterisk).

The endplates of the lumbar IVDs undergo ossification in wild-type adult mice (Figure 6, K’ and 6L’, black asterisks, and Supplemental Figure 7, F-G’). Interestingly, the *Sox9^del/del^* mice consistently displayed a reduction in endplate ossification in lumbar regions (Figure 6M’ and Supplemental Figure 7J’), which was associated with an accumulation of COLX (+), hypertrophic chondrocyte-like cells in the endplate (Figure 6P’, red asterisk; Figure 6M’, black asterisk, and Supplemental Figure 7, H’ and I’). COLX is a biomarker of IVD degeneration (50, 51). We also observed increased COLX expression enriched around the endplate cleft and in the growth plate and nucleus pulposus of the *Sox9^del/del^* mice (Figure 6, P-P’), which suggests that reduction in SOX9 function is eliciting degenerative changes in the IVD. The ossification of the endplate in wild-type and *Sox9^del/+^* mice was also correlated with a reduction of COLII expression in the endplate (Figure 6, Q and R, red asterisks), however, COLII staining was increased in *Sox9^del/del^*mutant endplate (Figure 6S, red asterisks). The persistent expression of both COLII (Figure 6S) and COLX (Figure 6P’) in large, hypertrophic chondrocyte-like cells in the endplate suggests that the TAM domain of SOX9 is important for pathways involved in chondrocyte maturation in the lumbar spine.

### Variants of the TAM domain decrease SOX9 protein stability and chondrogenesis in cell culture

Next, we sought to determine how disturbance of the TAM domain affects SOX9 function. Consistent with our previous results with the human Gly276Cys *SOX9* TAM domain variant (Figure 2B), the luciferase assay showed that the Asp272del variant of *SOX9* showed no obvious change in transcriptional activity compared with the wild-type protein or the Gly276Cys variant (Figure 2B). Previous studies reported that phosphorylation of the TAM domain helps regulate SOX9 stability in response to DNA damage in cancer cells (52). We next sought to assay SOX9 protein stability by measuring the half-life of different SOX9 variants in Cos-7 cell culture after treatment with cycloheximide (CHX), which blocks *de novo* protein synthesis (53). Quantification of SOX9 revealed that the half-life of wild-type SOX9 was ∼4 hours after CHX treatment, while the half-lives of both the human Gly276Cys and the mouse Asp272del variant of SOX9 proteins were reduced to ∼2.5 hours (Figure 7, A and B, n=3).

**Figure 7.**
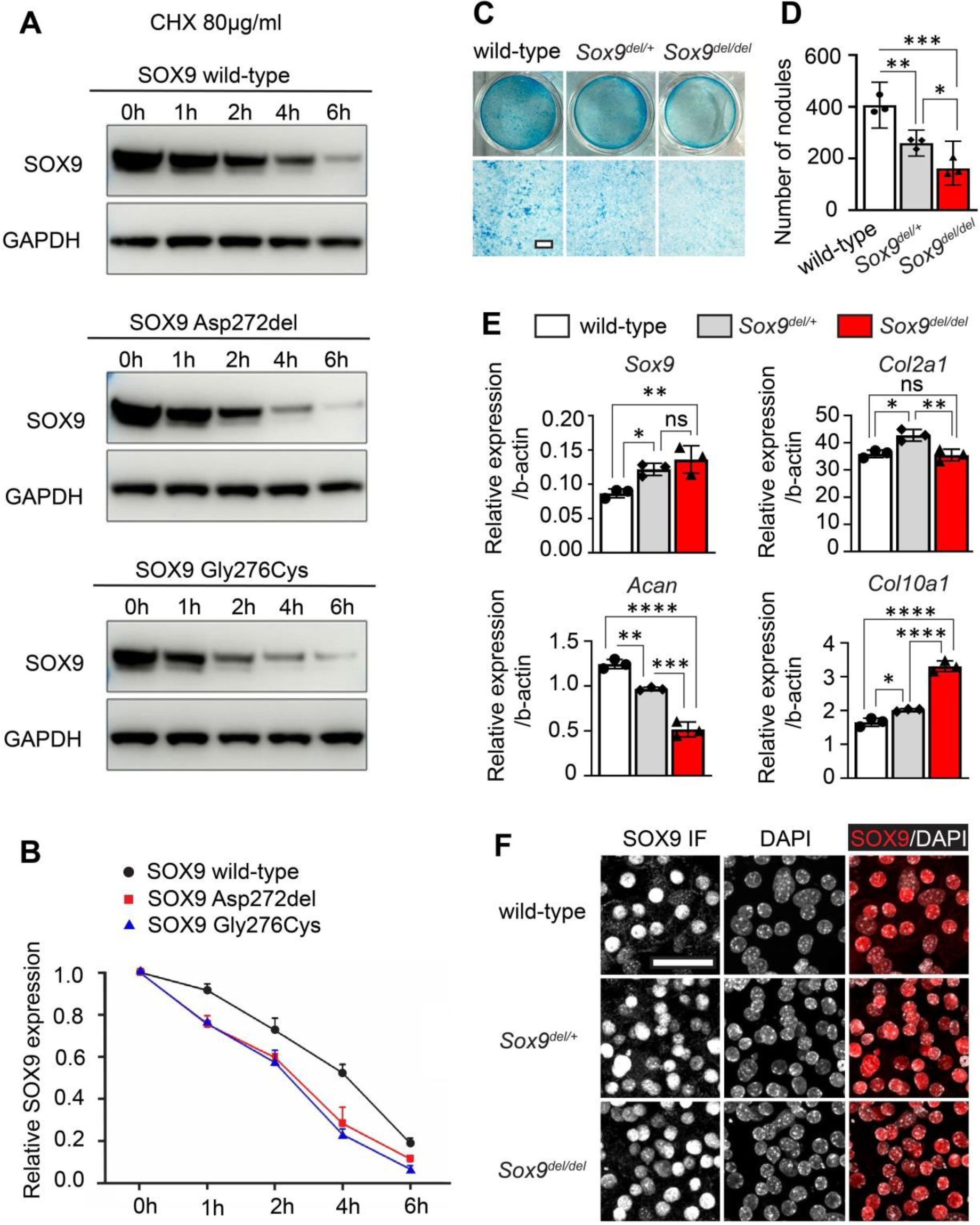
Variants of the TAM domain leads to reduced protein stability of SOX9 and impaired chondrogenesis in cell culture. (**A-B**) Representative Western blot assay of SOX9 half-life measurements (**A**). Cos-7 cells were transfected with plasmids expressing wild-type SOX9, the Asp272del variant, or the Gly276Cys variant for 48 hours, followed by cycloheximide (CHX) treatment (80 μg/ml) for 0h, 1h, 2h, 4h, and 6h. Total protein was harvested at each time point and blotted with SOX9. GAPDH was used as a loading control. Quantification of band intensity was shown in (**B**). (n=3 experiments.) (**C-D**), Alcian Blue staining (**C**) on costal chondrocytes isolated from P5 wild-type, *Sox9^del/+^*, and *Sox9^del/del^* mice that were maturated for 10 days. The number of nodules formed per tissue-culture well was quantified in (**D**). (n=3 experiments.) (**E**), Real-time RT-PCR analyses of *Sox9*, *Col2a1*, *Acan*, and *Col10a1* on RNA isolated from P5 primary costal chondrocytes of three different genotypes. Bars are plotted with mean and 95%CI. Three pups of each genotype were pooled to generate a sample. Three technique replicas were applied for each experiment. The statistical difference is evaluated by one-way ANOVA followed by Tukey’s multiple comparison test. * p<0.05, **: p<0.01, ***p<0.001. ***p<0.0001. ns: not significant. (**F**), Immunofluorescence (IF) analyses of SOX9 performed on P5 primary costal chondrocytes of three different genotypes revealed comparable expression levels and normal nuclei localization of the SOX9 Asp272del variant protein. (n=3 experiments.) *Scale bars: 500μm in **C**; 50μm in **F***.

Given the pathology of cartilaginous tissues in the IVD in *Sox9^del/del^* mutant mice, we next sought to test if primary costal chondrocytes would also model these defects in vitro. Alcian Blue staining on maturated primary costal chondrocytes revealed weaker Alcian Blue staining and a reduction in cartilage nodules formation of chondrocytes derived from both *Sox9^del/+^* and *Sox9^del/del^* mice compared with that derived from wild-type littermates (Figure 7, C and D), suggesting diminished chondrogenesis in vitro (54) with the Asp272del TAM domain variant of *SOX9*. This observation is confirmed by the reduced expression of *Acan* (Figure. 7E), which encodes the major chondroitin sulfate proteoglycan aggrecan and its deposition is considered a hallmark of chondrogenesis (55). In agreement with our observations of increased COLX expression in cartilaginous endplate in *Sox9^del/del^* mutant mice (Figure 6, P-P’), we also observed increased *Col10a1* expression in primary costal chondrocyte culture in both *Sox9^del/+^* and *Sox9^del/del^*mice (Figure 7E). These findings indicate that the Asp272del TAM domain variant of *SOX9* is driving cell autonomous, precocious hypertrophy signaling in chondrocytes. Similar alterations in gene regulation have been previously observed in the *Sox9* haploinsufficiency mice, where the overall markers of chondrogenesis were inhibited, yet are also coupled with increased expression of markers of hypertrophy and mineralization (23, 56).

*Col2a1* is a direct transcriptional target of SOX9 (28), however, *Col2a1* expression was not obviously changed in costal chondrocytes derived from *Sox9^del/+^* or *Sox9^del/del^* mice (Figure 7E), which is consistent with our analysis of COLII in vivo (Supplemental Figure 4, C-E). We observed increased *Sox9* mRNA level in costal chondrocytes of *Sox9^del/+^* and *Sox9^del/del^*mice compared with that in wild-type cells (Figure 7E), likely the result of SOX9 positive feedback on its expression (57). Despite increased *Sox9* transcript levels, we failed to observe any obvious changes in the expression or localization of the SOX9 protein by immunofluorescence or western blotting in primary costal chondrocytes (Figure 7F and Supplemental Figure 8). These results suggest a potential complementation effect driving *Sox9* mRNA overexpression in response to the hypomorphic loss of function of the TAM domain microdeletion in *SOX9*, which was consistent with our previous observations of decreased SOX9 protein stability (Figure 7, A and B).

### Disturbance in of the TAM domain of SOX9 generates global transcriptional changes in chondrocytes and altered protein expression in spinal tissues

Our results show that variants in the TAM domain of *SOX9* lead to the compromised formation of the intervertebral discs (Figure 3G and Figure 5L), altered ossification in the endplates (Figure 6, M, P and S), and decreased chondrogenesis in cell culture (Figure 7, C-E). All of this data suggests that variants in the TAM domain of *SOX9* have a crucial role in the differentiation and maturation of the cartilage lineages. To better understand how the Asp272del variant affects gene regulation, we performed RNA-sequencing analyses on P5 primary costal chondrocytes, a well-accepted source of primary chondrocytes (58, 59). We observed a total of 816 upregulated genes and 452 down-regulated genes in homozygous *Sox9^del/del^* compared to heterozygous *Sox9^del/+^* mutant mice (Figure 8A and Supplemental Table 3). Gene Ontology (GO) enrichment analysis revealed that the differentially expressed genes were strongly related to extracellular matrix (GO:0031012), angiogenesis (GO:0001525), and ossification (GO:0001503) (Supplemental Table 4). Other altered biological processes included connective tissue development (GO:0061448), cell-substrate adhesion (GO:0031589), and epithelial cell migration (GO:0010631) (Supplemental Table 4).

**Figure 8.**
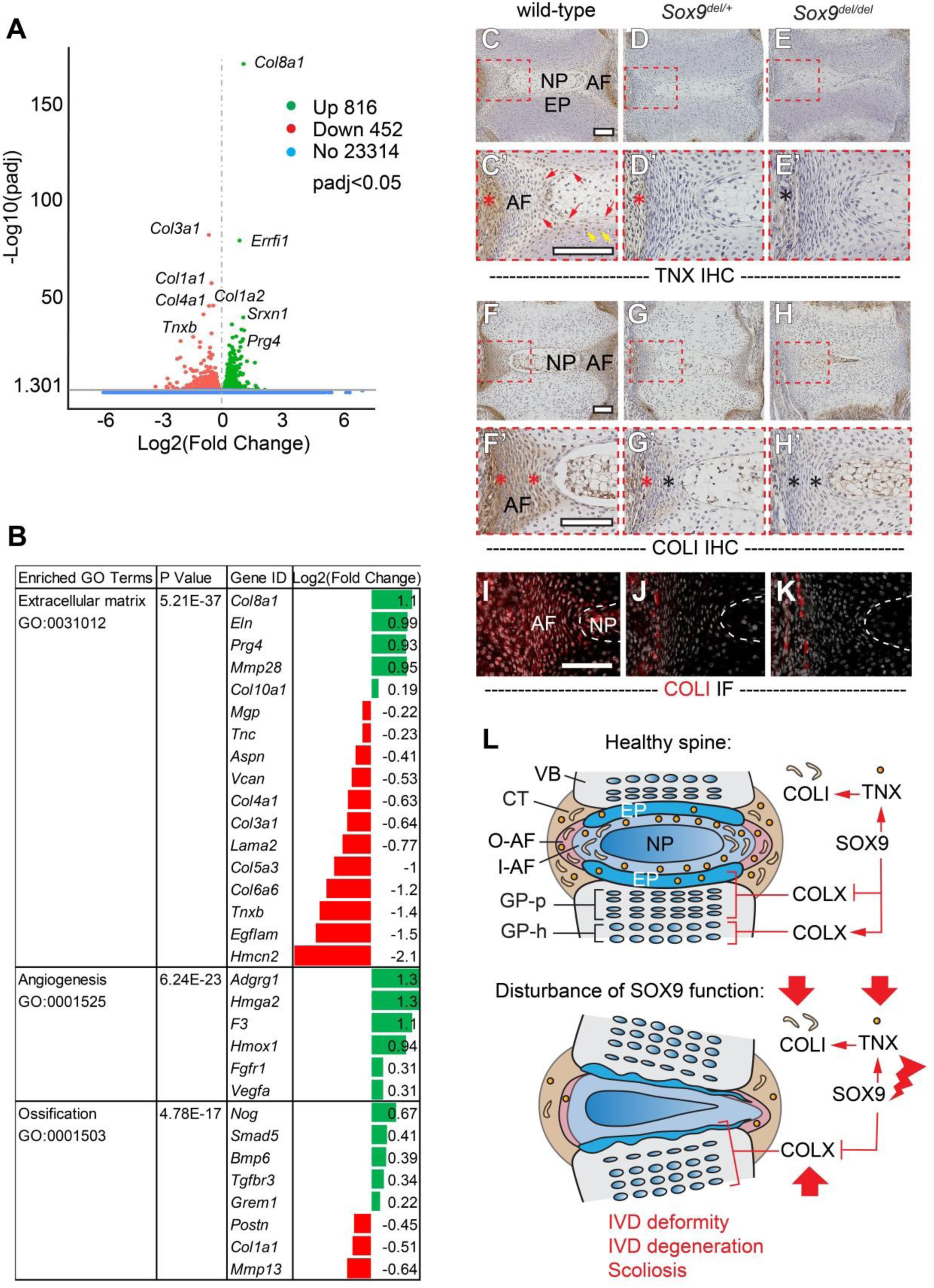
The Asp272del TAM domain variant of SOX9 generates global transcriptional changes in chondrocytes and altered protein expression in spinal tissues of the IVD. (**A**) Volcano plot of RNA-sequencing data generated from P5 primary costal chondrocytes, plotted with Log2 (Fold Change) (X axis) and Log10 (padj)(Y axis, padj<0.05). Several top-changed genes were labeled on the plot. (n=3 replicas for each genotype.) (**B**), A list of candidate genes involved in the top three categories of GO term enrichment analyses. **C-H’,** Immunohistochemistry (IHC) analyses of tenascin X (TNX) and type I collagen (COLI) in P5 tail sections of wild-type (**C, C’** and **F, F’**), *Sox9^del/+^* (**D, D’** and **G, G’**) and *Sox9^del/del^* (**E, E’** and **H, H’**) mice. TNX expression in connective tissues (red asterisks), inner annulus fibrosus (red arrows), and endplate (yellow arrows) are shown in (**C’**). Loss of TNX expression in connective tissues is indicated with a black asterisk in (**E’**). The presence or loss of COLI staining in annulus fibrosus is indicated with red or black asterisks respectively in (**F’-H’**). (n=6 for each genotype.) **I-K,** Immunofluorescence (IF) analyses of COLI were performed on P5 mouse tail sections of three different genotypes. (n=6 for each genotype.) **L,** A model of how SOX9 regulates axial skeleton homeostasis. In a healthy spine, SOX9 maintains normal expression of TNX and COLI in annulus fibrosus, connective tissues and cartilaginous tissues, and precisely controls the expression of COLX in chondrocytes. However, disturbance of SOX9 function results in reduced expression of TNX and COLI in connective tissues and dysregulated expression of COLX in the endplate and growth plate of the spine, which synergistically results in a range of skeletal dysplasia, including intervertebral disc (IVD) deformity, IVD degeneration, and scoliosis. *Scale bars: 100μm. VB: vertebral body; CT: connective tissue; EP: endplate; NP: nucleus pulposus; GP-p: proliferative growth plate; GP-h: hypertrophic growth plate; AF-i: inner annulus fibrosus; AF-o: outer annulus fibrosus*.

We observed global alterations in components of the extracellular matrix, including increased expression of *Col8a1* (type VIII collagen), *Eln* (elastin), and *Prg4* (proteoglycan 4), and mild upregulation of *Col10a1* (type X collagen) (Figure 8B), which is consistent with our observations of increased *Col10a1*/COLX expression in vivo (Figure 6, P-P’) and in primary chondrocyte culture (Figure 7E). On the contrary, the expression of some fibrotic collagens, including *Col3a1* and *Col4a1* was significantly reduced (Figure 8B). Additional extracellular matrix components, such as *Tnxb* (tenascin-X), *Tnc* (tenascin-C) and *Lama2* (laminin subunit alpha 2) were also down-regulated. In addition, several genes involved in angiogenesis such as *Vegfa* were upregulated. Finally, we observed intriguing results in a class of genes associated with ossification. We found reduced expression of some major markers of ossification, including *Col1a1*, *Mmp13,* and *Postn* (periostin, osteoblast specific factor), and increased expression of *Timp3* (tissue inhibitor of metalloproteinase 3) (Figure 8B). These results may explain the progressive, high penetrance (93%) of the rib cage deformities in the *Sox9^del/del^* mice (Figure 6F’), which may be caused by impaired ossification of the rib cage. This dysregulation of genes important for endochondral ossification is also consistent with the loss of endplate ossification observed within the lumbar IVDs of the *Sox9^del/del^* mice (Figure 6, M-M’). We also observed increased expression of the bone morphogenetic protein (BMP) and transforming growth factor-β (TGFβ) pathways, including *Bmp6*, *Smad5,* and *Tgfbr3*, coupled with up-regulation of some BMP signaling inhibitors, such as *Nog* (Noggin) and *Grem1* (gremlin, 1 DAN family BMP antagonist) (Figure 8B). Collectively, these results show large-scale dysregulation of genes that are important for chondrocyte differentiation and ossification in *Sox9^del/del^* mice.

In humans, disruption of the *TNXB* gene (which encodes tenascin-X, TNX, an extracellular matrix protein) is associated with Ehlers-Danlos syndrome, which is a connective tissue disorder caused by defective deposition of type I collagen leading to hypermobility of joints and kyphoscoliosis in some patients (60, 61). Since we observed reduced expression of both *Tnxb* and *Col1a1* in primary chondrocyte cultures prior to the onset of scoliosis, we next set out to assay the expression of TNX and COLI in vivo. IHC analyses on P5 wild-type tail sections showed strong TNX expression in the inner annulus fibrosus (Figure 8, C-C’, red arrows), a portion of cells in the endplate (Figure 8C’, yellow arrows), and the connective tissues surrounding the annulus fibrosus (Figure 8C’, red asterisk). However, the expression of TNX was consistently diminished in both *Sox9^del/+^* and *Sox9^del/del^* mice (Figure 8, D’ and E’), with only weak staining in the connective tissues adjacent to IVD in the *Sox9^del/+^* mice (Figure 8D’, red asterisk). Similarly, IHC and IF analyses revealed robust COLI expression in the entire annulus fibrosus and adjacent connective tissues of the wild-type mice (Figure 8, F-F’, red asterisks; and Figure 8I), which is depleted in both *Sox9^del/+^* and *Sox9^del/del^* mice (Figure 8, G and H’), with limited expression in the outer annulus fibrosus (Figure 8G’, red asterisk; and Figure 8J-K). Interestingly, the diminished expression of TNX and COLI in annulus fibrosus overlaps altered SOX9 expression in annulus fibrosus (Figure 3, H’’-J’’’), suggesting a regulatory role of SOX9 in maintaining the proper expression of these genes in the IVD. Altogether, this finding suggests that disturbance of the TAM domain allows for normal skeletal development to occur but still results in the loss of components, such as TNX and COLI, that are likely necessary for the maturation and homeostasis of the spine (Figure 8L).

## DISCUSSION

Heterozygous mutations in *SOX9* cause CMPD, a rare, dominant semi-lethal skeletal disease characterized by congenital shortening and bending of long bones. Most of the variants that cause CMPD are non-truncating mutations in HMG and DIM domains or truncating mutations in PQA and TAC domains (31–33). Notably, variants associated with acampomelic CMPD, a subpopulation of CMPD patients that exhibit milder skeletal phenotypes without displaying bending of long bones, are more likely to be either missense mutations of *SOX9* or deletions or rearrangements upstream of the *SOX9* coding region (34, 38). These observations suggest that locations and the nature of the variants may underlie the severity of CMPD/ acampomilic CMPD.

We recently reported a non-truncating pathological variant in the TAC domain which is associated with a milder form of axial skeleton dysplasia (35). Here, we reported the first non-truncating pathological variant in the TAM domain of *SOX9*. Patient with this variant exhibit skeletal dysplasia that primarily affects the axial skeleton and is milder than CMPD/acampomelic CMPD. Our studies suggest that non-truncating *SOX9* variants in the middle (TAM) or C-terminal transactivation domains (TAC) of SOX9 may contribute to skeletal dysplasia that is milder than classic CMPD/acampomelic CMPD, in which they affect primarily or exclusively the axial skeleton. These two studies corroborate with in vitro evidence suggest that the two transactivation domains of SOX9 can work synergistically to control the function of SOX9 (8).

Though the function of SOX9 in different skeletal tissues is well studied in mice (23), the molecular, cellular, and developmental abnormalities leading to CMPD/ acampomilic CMPD, especially the milder form of axial skeletal dysplasia, are not well defined, largely due to the lack of human evidence and appropriate animal models (24). In the present study, we analyzed exome sequencing from 424 congenital vertebral malformations cases, a cohort of patients with a broad spectrum of syndromic and non-syndromic vertebral malformations. The inclusion of patients with milder skeletal phenotypes enabled us to identify four novel ultra-rare *SOX9* variants. We demonstrated that these variants were deleterious and provided acampomelic CMPD diagnosis for two of the patients who carry *de novo* variants in the HMG domain of *SOX9*. The identification of these two variants expands the mutational spectrum of *SOX9* in acampomelic CMPD and conforms that the hypomorphic *SOX9* allele that perturbs its DNA binding ability contributes to acampomelic CMPD. Another variant is in the DIM domain and leads to compromised SOX9 dimerization, which is consistent with previous analyses of DIM domain mutations (19).

For the first time, we identified a missense mutation, Gly276Cys, in the TAM domain of *SOX9*. The patient carrying this variant exhibited milder phenotypes than that of classical CMPD/ acampomelic CMPD, with multiple fusions of the thoracic vertebral and clinodactyly of the distal 5^th^ phalange. Unlike the three variants identified in HMG and DIM domains, the mutation in the TAM domain does not obviously affect the transcriptional activity of canonical direct SOX9 targets but leads to reduced protein stability of SOX9. A previous study using cancer cells shows that the S*OX9* TAM domain plays an important role in the cellular response to DNA damage. A conserved phosphodegron (CPD) motif locates within the TAM transactivation domain is essential for the binding of FBW7α to SOX9, which is required for subsequent ubiquitination and proteasomal destruction of the SOX9 protein (52). Our study confirms that TAM domain is also involved in the regulation of SOX9 stability in chondrogenic lineages, however, the mechanisms of how specific amino acid residues in TAM domain regulate the turnover of SOX9 protein warrants further investigation.

Since no non-truncating variants in the TAM domain of SOX9 has been previously reported, we engineered a new mice model (*Sox9^del/del^*) resulting in an in-frame microdeletion of a single amino acid in the TAM domain (Asp272del), which is four residuals upstream of the human variant (Gly276Cys). Though we were not able to generate the exact human mutation, the *Sox9^del^* allele also provides us with valuable information to understand the role of the TAM domain in *SOX9* function and the etiology of a milder form of axial skeleton dysplasia. *In silico* analyses show that the Asp272 and Gly276 residues are highly conserved among vertebrate orthologues, and both are predicted to hydrogen bond interact with Asp274 to stable a putative helical structure of the SOX9 protein. In addition, the Asp272del variant also shows adequate luciferase activity and reduced protein stability that is comparable with the Gly276Cys variant. We further demonstrate that *Sox9^del^* is a hypomorphic allele with the genetic complementary experiment over a conditional *Sox9* null allele (*Sox9^flox^*). These observations suggest that the *Sox9^del^* allele may serve as a good genetic mouse model to study the function of the TAM domain and the pathogenesis of milder forms of axial skeletal dysplasia.

Interestingly, the homozygous *Sox9^del/del^* mutant mice display axial skeleton phenotypes that phenocopy some of the dysplasia observed in the human patient with the Gly276Cys variant, but not in mouse models with typical null or haploinsufficiency of *Sox9* (3, 23). For example, the *Sox9^del/del^* mutant mice display bending and fusion of some caudal vertebrae without displaying bending of the long bones. The *Sox9^del/del^* mutant mice also show loss of T13 floating ribs, rib cage dysmorphologies, pathoses of IVD and growth plates, and late-onset scoliosis without congenital vertebral malformations. In contrast, haploinsufficiency of *Sox9* results in more severe forms of skeletal dysplasia and defective cartilage development (14, 23, 38). On the other hand, the *Sox9^del/+^* mutant mice show much milder skeletal phenotypes compared with *Sox9^del/del^* and *Sox9* haploinsufficiency mice. All these observations confirm that *Sox9^del^*is a mild hypomorphic allele with no dominant-negative impact as reported in other *SOX9* truncating variants (24). Though *Sox9^del^* does not obviously affect the expression of SOX9 protein at the tissue level (Supplemental Figure 8, Western blot of primary chondrocytes) or significantly reduce its transcriptional activity, the function of SOX9 may be compromised by reduced protein stability. Therefore, the hypomorphic *Sox9^del^* allele may permit some transcriptional activity at the canonical SOX9 targets and allows for normal skeletal development to occur, however, it may lead to compromised tissue development and maintenance during neonatal and adult development, such as reduced proliferation of the endplate in P5 IVD, and reduced proteoglycan staining of the vertebral growth plate in adult mice, which is consistent with the previous finding on the crucial role of SOX9 in disc cell survival and phenotype maintenance, and in the regulation of distinct compartment-specific transcriptomic landscapes (6, 7).

Moreover, IHC analysis on tissue sections shows that the expression of SOX9 is reduced in distinct regions of the cartilaginous and connective tissues of the spine (Figure 3, P5 tail). RNA-seq performed on costal chondrocytes at the same age revealed that *Sox9^del^* mutation causes reduced expression of extracellular matrix components, and alterations in angiogenesis and ossification pathways. SOX9 is known to have a direct role in the establishment of tendon and ligament attachments and indirectly for endochondral ossification (2, 5), thus the role of SOX9 TAM variants on the development and homeostasis of bone and dense connective tissues should be further studied.

One possible mechanism of wedged IVD observed in P5 *Sox9^de/dell^* mice is that the altered extracellular matrix components (e.g. abnormally decreased expression of TNX and COLI) lead to altered biomechanical properties of the disc, while the increased expression of type X collagen and dysregulated ossification process within the vertebrae, probably due to the reduced expression of *Col1a1* and *Mmp13* (Figure 8B) and reduced proliferation within the proliferative growth plate (Figure 3Q), synergistically leads to disc deformity (Figure 8L). We also observe signs of disc degeneration in older *Sox9^de/dell^* mice, including reduced proteoglycan staining and endplate abnormalities. Our study adds support to previous studies showing that SOX9 is essential for spine development and maintenance of the structural integrity of the spine (3, 6, 7, 49). Interestingly, we observed a similar endplate phenotype in another genetic mouse model (*Adgrg6* conditional knockout) that was published earlier by our group (62). In *Adgrg6* conditional mutant mice, we suggest that the endplate-oriented phenotype is a secondary effect due to compromised biomechanical properties of the IVD because of altered expression of extracellular matrix components, including reduced expression of SOX9 but increased expression of type X collagen (62, 63). In this study, we observed similar alterations in *Sox9^del/del^* mice, where we showed SOX9 expression is regionally reduced, increased expression of type X collagen in the endplate and growth plate, supporting a potential synergistic interaction between SOX9 and ADGRG6, which is implicated in adolescent idiopathic scoliosis in humans (64).

Taken together, our results demonstrate adult viable skeletal dysplasia phenotypes including scoliosis and malformation of the distal axial skeleton in mice and humans, due to novel pathological mutations localized within the TAM domain of *SOX9*. Disturbance of the *SOX9* TAM domain in mice and in human patient showed axial skeletal malformations without generating more severe skeletal dysplasia and autosomal sex reversal phenotypes, which are commonly associated with known CMPD/ acampomilic CMPD *SOX9* mutations in humans. Our study corroborates previous evidence from human and mouse studies that *SOX9* has critical roles in axial skeleton formation and adult homeostasis and that *SOX9* hypomorphic alleles resulting from variants in the TAM or TAC domain likely underlie skeletal dysplasia that is milder than CMPD/ acampomilic CMPD, which primarily or exclusively affect the axial skeleton. In conclusion, human and mouse variants in *SOX9* contribute to a range of skeletal dysplasia dependent on the domain/residual affected, which is suggestive of a general mechanism to be further explored in skeletal dysplasia in humans.

## MATERIALS AND METHODS

### Human subjects

A total of 424 probands with congenital vertebral malformations admitted into Peking Union Medical College Hospital (PUMCH) were consecutively enrolled under the framework of the <u>D</u>eciphering disorders <u>I</u>nvolving <u>S</u>coliosis and <u>CO</u>morbidities (DISCO) study (http://www.discostudy.org/) from 2009 to 2016. Clinical evaluation and exome sequencing were performed as previously described (65). Raw data were processed using the in-house developed Peking Union Medical College Hospital Pipeline (PUMP) (65, 66). Ultra-rare variants in *SOX9* were selected based on the following criteria: 1) Predicted to alter the protein sequence; 2) Either arising *de novo* or absent from the public databases such as the genome aggregation database (gnomAD, https://gnomad.broadinstitute.org/). Patients carrying candidate variants in *SOX9* are reported in this study.

### Sanger sequencing

For human patients and mouse models, candidate variants in *SOX9*/*Sox9* were validated by Sanger sequencing. For human patients, variant-encoding amplicons were amplified by PCR from genomic DNA obtained from probands and parents from trios (Supplemental Table 1). The amplicons were purified using an Axygen AP-GX-50 kit (lot no. 05915KE1) and sequenced by Sanger sequencing on an ABI3730XL instrument.

### Construction of *SOX9* expression constructs and Cos-7 cell culture

Full-length wild-type *SOX9* and *SOX9* with variants were cloned into the pEGFP-C1 expression vector (Catalog #6084-1, Takara Bio Company). Cos-7 cells were obtained from the Institute of Basic Medical Sciences, Chinese Academy of Medical Sciences (Beijing, China). The cells were cultured in DMEM medium (Gibco) supplemented with 10% fetal bovine serum (Gibco), penicillin (50 U/ml, Life Technology), and streptomycin (50 μg/ml, Life Technology).

### Electrophoretic mobility shift assay

Electrophoretic mobility shift assay (EMSA) was used to detect the binding affinity of SOX9 proteins to the conical SOX9 binging motifs as previously described with modifications (19). Biotin-labeled DNA probes were designed based on two canonical SOX9 binding motifs derived from an enhancer region of the mouse *Col11a2* gene, named *Col11a2* B/C and D/E respectively, according to the original publication (19). The sequences of these motifs are listed in Supplemental Table 2. Cos-7 cells were transfected with the SOX9 expression constructs or GFP control construct for 48 hours. The expression level of wild-type and various SOX9 mutant proteins was quantified by Western blot before the experiment and equal amounts of SOX9 were detected (Supplemental Figure 2). After 48 hours, the whole-cell lysates were extracted and incubated with 0.5 µl of Biotin-labeled probe (1000fM) for 20 min on ice in a 15μl reaction system as described previously (19), using Poly (dI:dC) as the unspecific competitor. The 150-fold unlabeled DNA probes (150X) were incubated with cell lysate of Cos-7 cells transfected with wild-type SOX9 protein. Samples were loaded onto native 6.5% polyacrylamide gel and electrophoresed in 0.5xTBE at 180V for 1 h. The gel was transferred to a nylon membrane at 100V for 30 min. The biotin-labeled DNA probes transferred to the membrane were cross-linked for 10 min with a hand-held UV lamp and detected by Chemiluminescence (BioRad, TY8836).

### Luciferase assay

Luciferase assay was performed with a luciferase reporter (*Col11a2*-Luc) and various *SOX9* expression constructs in Cos-7 cell cultures. To generate the *Col11a2*-Luc reporter, four tandem copies of a canonical SOX9 binding motif derived from an enhancer region of the mouse *Col11a2* gene (named *Col11a2* D/E) (19) was cloned into the luciferase reporter vector pGL3Basic (Promega E1751). The sequence of this motif is listed in Supplemental Table 2. Full-length wild-type *SOX9* and *SOX9* with variants were cloned into the pEGFP-C1 expression vector (Catalog #6084-1, Takara Bio Company). These overexpression constructs were transfected into the Cos-7 cell line for 48 hours and equal amounts of protein were harvested for Western Blotting analysis of SOX9. No obvious difference in SOX9 expression was observed among wild-type and different *SOX9* variants (Supplemental Figure 3, A and B). A standard curve was generated to determine the linear range of the SOX9-mediated reporter transactivation and to optimize the reaction system (Supplemental Figure 3C). For each independent experiment, Cos-7 cells were transfected with 400ng of *Col11a2*-Luc reporter plasmids, 50ng of various SOX9-expression vectors, and 5ng of Renilla luciferase control reporter vectors. Cells were harvested 48h post-transfection for immediate luciferase activity analysis using the Promega Dual-Luciferase Reporter Assay System (Promega E1910). The Firefly luciferase activity was normalized to the Renilla luciferase activity. Three independent experiments were performed with reproducible results.

### Half-life measurement

The half-life of wild-type and variant SOX9 proteins were evaluated using a cycloheximide (CHX) chase assay. Cos-7 cells were seeded into 6-well plates in DMEM containing 10% FBS and transfected with 2 μg of various *SOX9* expression constructs. 48 hours later, Cos-7 cells were treated with 80 μg/ml of CHX and were harvested at 0h, 1h, 2h, 4h, and 6h after treatment. Cells were dissolved in absolute ethanol and harvested in ice-cold phosphate-buffered saline (PBS, pH 7.4) by centrifuging at 2500 × g for 2 min at 4 °C. Cell pellets were lysed in RIPA lysis and extraction buffer (Thermo Scientific) and sonicated for 3 × 5 s on ice. Total protein from the cell lysate was quantitated using the Pierce BCA protein assay kit (Thermo Scientific) and analyzed by the standard Western blot assay. The membrane was blotted with a primary anti-GFP tag antibody (Cell Signaling Technology, 2956T, 1:1000) and detected with a secondary horseradish peroxidase-conjugated antibody (ab7090; Abcam; 1:5000). Quantification of band intensity was conducted using Image Lab (BioRad). Three independent experiments were performed with reproducible results.

### Generation of mouse models

The *Sox9 ^Asp272del^* mutant mouse was fortuitously isolated after pronuclear injection using an oligo-dependent gene editing approach using a single CRISPR synthetic guide RNA 5’-GTTCACCGATGTCCACGTCG-3’ (Synthego) mixed with Alt-R S.p. HiFi Cas9 Nuclease V3 protein (IDT, 1081060). For gene editing, a donor oligonucleotide 5’-TCTGGCAGAGGGGGGCAGACAGCCCCCCATCGACTTtCGtGAtGTaGAtATCtGcG AACTGAGCAGCGACGTCATCTCCAACATTGAGACCTT-3’, with phosphorothioate linkages between the first three and last three nucleotides, was used to induce a specific edit to model the Gly276Cys human mutation and add synonymous changes to include an EcoRV restriction site for genotyping. The pronuclear injection was performed by the Mouse Genetic Engineering Facility (UT-MGEF) at the University of Texas at Austin using standard protocols (https://www.biomedsupport.utexas.edu/transgenics). Of the 123 implanted embryos from two independent sessions, 3 mice were isolated at sexual maturity. Of these, 1 founder male transmitted the *Sox9 ^Asp272del^* allele. Locus-specific genotyping of the *Sox9 ^Asp272del^* mutation was performed using PCR primers 5’-GTCTTTCTCTTTTATGGCCTGC-3’and 5’TGGCAAGTATTGGTCAAACTCA-3’, followed by Sanger sequencing confirmation. Heterozygous *Sox9 ^Asp272del^* mutant mice are maintained in the C57B6/J background and have been outcrossed to the C57BL/6J strain for over 10 generations. At each cross, we observe a 100% correspondence between genotype and phenotype.

### Analysis of mouse tissues

Radiographs of the mouse skeleton were generated using a Kubtec DIGIMUS X-ray system (Kubtec T0081B) with auto exposure under 25 kV. All radiographs were taken immediately after the mice were euthanized to avoid the stiffness of the skeleton. Cobb’s angle was measured on high-resolution X-ray images with the software Surgimap (https://www.surgimap.com), as previously described by (46). Measures of the length of the spine (T1-L6) and long bone (femur) were performed on high-resolution X-ray images with ImageJ (https://imagej.nih.gov/ij/).

Mouse spine or tail tissues were harvested at P5, P60, and 6 months of age. Histological analysis was performed on the distal tail or lumbar (L3-L5) spines fixed in 10% neutral buffered formalin for 3 days at room temperature, followed by 3-5 days of decalcification in Formic Acid Bone Decalcifier (Immunocal, StatLab). After decalcification, bones were embedded in paraffin and sectioned at 5μm thickness. Alcian Blue Hematoxylin/Orange G (ABH/OG) staining was performed following standard protocols (Center for Musculoskeletal Research, University of Rochester). Immunohistochemical (IHC) analyses were performed on paraffin sections with traditional antigen retrieval (COLII and COLX: 4 mg/ml pepsin in 0.01 N HCl solution, 37°C water bath for10 min; SOX9 and Collagen I: 10 µg/ml proteinase K in 1x PBS, room temperature for 10 min; TNX: 10 mM Tris-EDTA with 0.05% Triton-X-100, pH 9.0, 75°C water bath for 5 min) and colorimetric developed development methodologies with the following primary antibodies: anti-SOX9 (EMD Millipore AB5535, 1:200), anti-COLII (Thermo Scientific MS235B, 1:100), anti-COLX (Quarteet, 1-CO097-05, 1:200), anti-COLI (Abcam, ab138492, 1:1000), and anti-TNX (Santa Cruz Biotechnology sc-271594, 1:100). Immunofluorescence (IF) staining was performed on paraffin sections as previously described with modifications (67). Briefly, paraffin sections of P5 mouse tails were de-paraffinized and incubated with 10 µg/ml proteinase K in 1x PBS at room temperature for 10 min. Sections were washed in 1xPBST and incubated with primary antibodies: anti-SOX9 (Chemicon AB5535, 1:200) and anti-TNX (Santa Cruz Biotechnology sc-271594, 1:100), and developed with secondary antibody Alexa Flour 488 Goat anti Rabbit (A11034, 1:1500). Images were taken using a Keyence BZ-X710 all-in-One fluorescence microscope and a Nikon Ti2E/CSU-W1 spinning disc confocal system. Quantification of SOX9 (+) cells was performed on high resolution SOX9 IHC staining images with ImageJ. At least two sections were analyzed for each mouse. At least three mice for each genotype were analyzed.

### MicroCT analysis

MicroCT analysis was performed on the tails of P60 mice and P1 mouse pups. All samples were fixed in 10% neutral-buffered formalin for 3 days at room temperature, thoroughly washed, and scanned on a Bruker SkyScan 1276 (Bruker Corporation, Billerica, Massachusetts). The tails were scanned at 50 kV and 200 μA with a 0.25 mm Al filter, with a 3 μm voxel size. The mouse pups were first proceeded for Alcian Blue/Alizarin Red skeletal stain using standard acid-alcohol conditions and cleared in 1% KOH and Glycerol. These embryos were scanned in glycerol at 50 kV and 100 μA with a 0.5 mm Al filter, with a 3 μm voxel size. 3D Rendering and analysis was done using Bruker CTVox and CTAn software.

### Costal chondrocytes isolation and maturation

Murine costal chondrocytes were isolated as previously described by (67) with modifications. Briefly, anterior rib cages and sterna were isolated from 5-day-old wild-type, *Sox9^del/+^*and *Sox9^del/del^* pups with the soft tissue removed. Rib cages were digested in 2mg/ml Pronase (Roche) solution at 37°C for 1 hour with constant agitation, followed by 1-hour digestion in 3mg/ml collagenase D (Roche) solution in high-glucose Dulbecco’s modified Eagle’s medium (DMEM, Gibco) at 37°C. The rib cages were then transferred into a petri dish and digested in collagenase D (3 mg/ml in high-glucose DMEM) for 4 to 6 hours. Murine costal chondrocyte cell suspensions were filtered through 40µm filters and seeded at a density of 200,000 cells per well in 24-well tissue culture plates. Cells were maturated with maturation medium [highglucose DMEM supplemented with 10% fetal bovine serum, 1% penicillin/streptomycin, and 50µg/ml ascorbic acid (Sigma, A4403)] for 10 days with changing of the medium in every two days. After 10 days of maturation, cells were fixed with 500µl of 10% Formaldehyde at RT for 20min, thoroughly washed with PBS, and incubated with Alcian Blue staining solution (Sigma, B8438) or anti-SOX9 antibody (EMD Millipore AB5535, 1:400) overnight at room temperature or 4°C. The next day Alcian Blue stained cells were washed with deionized water and air-dried overnight. The SOX9 antibody-stained cells were developed with a secondary antibody Alexa Flour 488 Goat anti Rabbit (A11034, 1:1500).

### RNA isolation and real-time RT-PCR

RNA was isolated from primary costal chondrocytes without maturation with a QIAGEN RNeasy mini kit (QIAGEN 74104). Reverse transcription was performed using 500 ng total RNA with an iScript cDNA synthesis kit (Bio-Rad 1708840). Real-time RT-PCR (qPCR) analyses were performed as previously described by (67). Gene expression was normalized to b-actin mRNA and relative expression was calculated using the 2-(ΔΔCt) method. The mouse-specific primers were used to quantify gene expression (Supplemental Table 1). Real-time RT-PCR efficiency was optimized and melting curve analyses of products were performed to ensure reaction specificity.

### Protein extraction and western blot of the primary cells

Total proteins were extracted from primary costal chondrocytes without maturation with protein extraction buffer [50mM HEPES, 1.5mM EDTA (pH 8.0), 150mM NaCl, 10% glycerol, 1% Triton X-100] supplemented with protease and phosphatase inhibitors (Roche). 10mg of protein from each sample was resolved by 4–15% SDS-polyacrylamide gel electrophoresis and transferred to the nitrocellulose membranes. Western blots were then blocked with LI-COR blocking buffer and incubated overnight with primary antibodies anti-SOX9 (EMD Millipore AB5535, 1:1000) and anti-GAPDH (Cell Signaling, #2118, 1:2500) at 4°C with gentle rocking. The next day western blots were detected with the LI-COR Odyssey infrared imaging system. Quantification of band intensity was conducted with ImageJ. Three independent experiments were performed with reproducible results.

### RNA-sequencing and data analysis

RNA-sequencing was performed on costal chondrocytes isolated from P5 *Sox9^del/+^* mice and *Sox9^del/del^* mice. Briefly, anterior rib cages and sterna were isolated from P5 pups, digested in 2mg/ml Pronase solution at 37°C for 1 hour with constant agitation, followed by 1-hour digestion in 3mg/ml collagenase D solution in 1XPBS. Cells were disassociated, thoroughly washed in 1XPBS and filtered through a 40µm cell strainer. Cells were pelleted by centrifuging at 300 × g for 4 min at room temperature. RNA was isolated with a QIAGEN RNeasy mini kit (QIAGEN 74104).

Library preparation, RNA sequencing, and bioinformatics analysis were performed by Novogene Co., Ltd. Messenger RNA was purified from total RNA using poly-T oligo-attached magnetic beads. After fragmentation, the first strand of cDNA was synthesized using random hexamer primers, followed by the second strand of cDNA synthesis. The library was examined with Qubit and real-time PCR for quantification and a bioanalyzer for size distribution detection. Quantified libraries were pooled and sequenced on Illumina platforms and pairedend reads were generated. Raw data in fastq format were firstly processed through in-house Perl scripts and clean reads were obtained. Reference genome and gene model annotation files were downloaded from the genome website directly. Index of the reference genome was built using Hisat2 v2.0.5 and paired-end clean reads were aligned to the reference genome using Hisat2 v2.0.5. featureCounts v1.5.0-p3 was used to count the reads numbers mapped to each gene. FPKM of each gene was calculated based on the length of the gene and the reads count mapped to this gene. Differential expression analysis was performed using the DESeq2 R package (1.20.0). The resulting P-values were adjusted using Benjamini and Hochberg’s approach for controlling the false discovery rate. Genes with an adjusted P-value <=0.05 found by DESeq2 were assigned as differentially expressed. Gene Ontology (GO) enrichment analysis of differentially expressed genes was implemented by the clusterProfiler R package, in which gene length bias was corrected. GO terms with corrected P value less than 0.05 were considered significantly enriched in differential expressed genes.

### Statistics

Statistical analyses were performed in GraphPad Prism 9.1.2 (GraphPad Software Inc, SanDiego, CA) or SPSS version 15.0 (SPSS). For luciferase assay, half-life analysis, Cobb angle, and gene expression analysis to compare two or more experimental groups, 2-tailed Student’s t-test and Welch one-way ANOVA followed by Turkey’s multiple comparison test were applied when appropriate. A p-value of less than 0.05 is considered statistically significant.

### Study approval

This study was approved by the Institutional Review Board of Peking Union Medical College Hospital (JS-2364). Written informed consent was obtained from each patient or legal guardian of the patient. All mouse studies and procedures were approved by the Animal Studies Committee at the University of Texas at Austin.

### Statement

The patient ID is a unique research identifier that cannot be linked back to the original patients outside of the research group. Medical history details have also been removed to protect the patients’ privacy.

## Data Availability

All data produced in the present study are available upon reasonable request to the authors

## Data availability

Data are available from the corresponding author upon reasonable request.

## Author contributions

N.W., R.S.G., L.W., Z.L. and S.Z. conceptualized the project. L.W., Z.L., S.Z. L.Z., Xiaoxin.L., N.W. and R.S.G. designed the study. L.W., S.Z., Z.L., C.Q., B.T., G.H., R.S.G. L.L. Xinyu.L. and L.Z. performed the experiments. S.Z., L.W., Z.L., K.X., C.M.K, Xiaoxin.L., R.S.G., H.G. and L.L. validated the data. N.W., S.Z., L.W., Z.L., H.G., S.Y., V.A., S.M., S.W., Y.N., R.S.G. and T.J.Z. analyzed the data. L.W., S.Z., Z.L., K.X., N.W. and R.S.G. wrote the manuscript. S.Z., Z.L., L.W., K.X., N.W., Z.W., C.Q., Xinyu.L. and R.S.G. critically revised the manuscript. Z.L., R.S.G., N.W. and Z.W. supervised the work. All authors have read and approved the final submitted manuscript. The first authorship is shared by Z.L., L.W., S.Z., and K.X. The order was determined based on their respective contributions and efforts to the study.

## Acknowledgments

We thank Dr. Yunjia Wang at the Department of Spine and Orthopaedics in Xiangya Hospital/Central South University, and Ms. Karagan Day at the Department of Pediatrics in Dell Medical School of the University of Texas at Austin for their supporting work in mouse husbandry and X-ray image taking. We also thank Nanjing Geneseeq Technology Inc. for the technical help in sequencing and Ekitech Technology Inc. for the technical support in database and data management.

## SUPPLEMENTAL FIGURE LEGENDS

**Supplemental Figure 1. The 3D structures of SOX9 are predicted by AlphaFold.** (**A**) A model of human SOX9 was built by AlphaFold (htts://alphafold.ebi.ac.uk/). (**B**) Ile73 is predicted to be important for stabilizing alpha-helical interactions. (**C**) Met113 is predicted to stabilize alpha-helical interactions through a single hydrogen bond with Gln117 and two hydrogen bonds with Aan110. (**D**) Ala133 is predicted to stabilize alpha-helical interactions by a hydrogen bond with Ser136. (**E**) Gly276 is predicted to hydrogen bond with Asp274. Asp272 is also predicted to hydrogen bond with Asp274 by stabilizing an alpha-helical structure.

**Supplemental Figure 2. Equal amounts of SOX9 proteins were used for the EMSA assay.** (**A**) Western blotting of anti-GFP was performed on total protein extracted from blank Cos-7 cells (Blank), Cos-7 cells transfected with empty EGFP expression vectors (Control), wild-type SOX9 (WT), or Ile73Thr, Met113Leu, and Ala133Gly SOX9 variants that were cloned into the EGFP expression vectors at 48 hours post-transfection. GAPDH is used as a loading control. No obvious changes in expression were observed among wild-type and variant SOX9 proteins. (**B**) Quantification analysis shows that comparable amounts of SOX9 were detected among wild-type and various SOX9 variant proteins. Bars are plotted with mean and SD. Each dot represents one experiment analyzed. The statistical difference is evaluated by one-way ANOVA followed by Tukey’s multiple comparison test. ns: not significant. (n=3 experiments.)

**Supplemental Figure 3. Optimization of the luciferase assay. (A)** Western blotting was performed on total protein extracted from blank Cos-7 cells (Blank), Cos-7 cells transfected with wild-type SOX9 construct (Wild type), or Ile73Thr, Gly276Cys, Met113Leu, Ala133Gly, and Asp276Del SOX9 variants at 48 hours post-transfection. GAPDH is used as a loading control. No obvious changes in expression were observed among wild-type and variant SOX9 proteins. **(B)** Quantification analysis shows that comparable amounts of SOX9 were detected among wild-type and various SOX9 variant proteins. Bars are plotted with mean and SD. Each dot represents one experiment analyzed. The statistical difference is evaluated by one-way ANOVA followed by Tukey’s multiple comparison test. ns: not significant. (n=3 experiments.) **(C)** A standard curve was generated to optimize the transfection dose for the luciferase assays. SOX9-expression plasmids were diluted to a gradient of concentration with the final amount from 10^-4^ to 10^6^ ng. Relative luciferase activity was normalized to the Renilla internal control. The linear range of the SOX9-mediated reporter transactivation is between 1ng-100ng. 50ng of SOX9-expression plasmids was selected for luciferase assays shown in Figure 2B. n=3 experiments.

**Supplemental Figure 4. Expression of extracellular matrix components in P5 mouse tail sections. (A-B’’)** Representative Alcian Blue Hematoxylin /Orange G (ABH/OG) staining (**AA’’**) and IHC staining of SOX9 (**B-B’’**) on unwedged IVD in tail sections of P5 Sox9del/del mutant mice. The endplate is indicated with a dot line and a vertical bar in (**A’**) and (**B’**). The boundary between inner and outer annulus fibrosus is indicated with a dot-line in (**A’’**) and (**B’’**). (n=3.) C-H, IHC analysis of COLII **(C-E)** and COLX **(F-H)** on tail sections of P5 wildtype, *Sox9^del/+^,* and *Sox9^del/del^* mice. n=3 for each genotype. Scale bars: 100μm. EP: endplate; GP-p: proliferative growth plate; GP-h: hypertrophic growth plate; AF-i: inner annulus fibrosus; AF-o: outer annulus fibrosus.

**Supplemental Figure 5. Assessment of skeletal development in *Sox9* loss of function mutant mice.** (**A-C**) Representative skeletal preps of Cre(-) wild-type (**A**), heterozygous *Col2a1-Cre; Sox9^flox/+^* mutant (**B**), and compound heterozygous *Col2a1-Cre; Sox9^flox/del^* mutant mice (**C**) at P1. **(D-F)** Representative MicroCT images of Cre(-) wild-type (**D**), heterozygous *Col2a1-Cre; Sox9^flox/+^* mutant (**E**), and compound heterozygous *Col2a1-Cre; Sox9^flox/del^* mutant mice (**F**) at P1. n=4 for each genotype. Scale bar: 1mm.

**Supplemental Figure 6. Young adult *Sox9^del/del^* mice display mild rib cage deformities.** (**A-C**) Representative sagittal X-ray images of wild-type (**A**), *Sox9^del/+^* (**B**), and *Sox9^del/del^* mice (**C**) at P60. Red arrows indicate the curvature of the distal ribs (**C**). n=4 for wild-type mice; 2 males, 2 females. n=8 for *Sox9^del/+^* mice; 5 males, 3 females. n=7 for *Sox9^del/del^* mice; 4 males, 3 females. Scale bar: 10mm.

**Supplemental Figure 7. Mature adult *Sox9^del/del^*mice display rib cage deformities and impaired ossification in the endplate of the intervertebral discs.** (**A-E**) Additional sagittal X-ray images of wild-type (**A**), *Sox9^del/+^* (**B**), and *Sox9^del/del^* mice (**C-E**) at 6 months. Red arrows indicate the bending of the sternums (**B-D**) or the curvature of the distal ribs I. (**F-J’**) Additional ABH/OG staining of lumbar IVDs (L3/4 or L4/5) of wild-type (**F, F’**), *Sox9^del/+^* (**G, G’**) and *Sox9^del/del^*(**H-J’**) mice at 6 months. Wild-type and *Sox9^del/+^* mice consistently showed ossification within the endplate at 6 months (yellow arrows, **F’** and **G’**). On the contrary, *Sox9^del/del^* mice showed reduced endplate ossification (yellow arrow, **J’**) or persistency of hypertrophic chondrocyte-like cells within the endplate (yellow asterisks, **H’**, **I’**). n=3 for each genotype. Scale bars: 10mm in **A**; 100μm in **F-F’**. AF: annulus fibrosus; EP: endplate; GP: growth plate; NP: nucleus pulposus.

**Supplemental Figure 8. SOX9 protein expression in costal chondrocytes.** (**A**) Representative Western blot image of SOX9 performed on P5 costal chondrocytes isolated from three different genotypes of mice. GAPDH is used as a loading control (n=3 experiments). (B) Quantification analysis shows that comparable amounts of SOX9 protein were detected among three genotypes. Bars are plotted with mean and 95%CI. Each dot represents one experiment analyzed. The statistical difference is evaluated by one-way ANOVA followed by Tukey’s multiple comparison test. ns: not significant (n=3 experiments).

**Supplemental Table 1.** Primers used for amplification of the DNA segments containing the candidate *SOX9* variants and for RT-PCR quantification.

**Supplemental Table 2.** Sequences of the SOX9 binding motifs used for EMSA and luciferase assay.

**Supplemental Table 3.** Differential gene expression analysis of primary costal chondrocytes derived from *Sox9^del/+^* and *Sox9^del/del^* mutant mice.

**Supplemental Table 4.** GO-terms associated with differential gene expression analysis of primary costal chondrocytes derived from *Sox9^del/+^* and *Sox9^del/del^* mutant mice.

