## Supplemental Files for "Variants in the SOX9 transactivation middle domain induce axial skeleton dysplasia and scoliosis"

### Supplemental Figure 1

Three-dimensional structure of SOX9 (AlphaFold prediction) with mutated residues marked

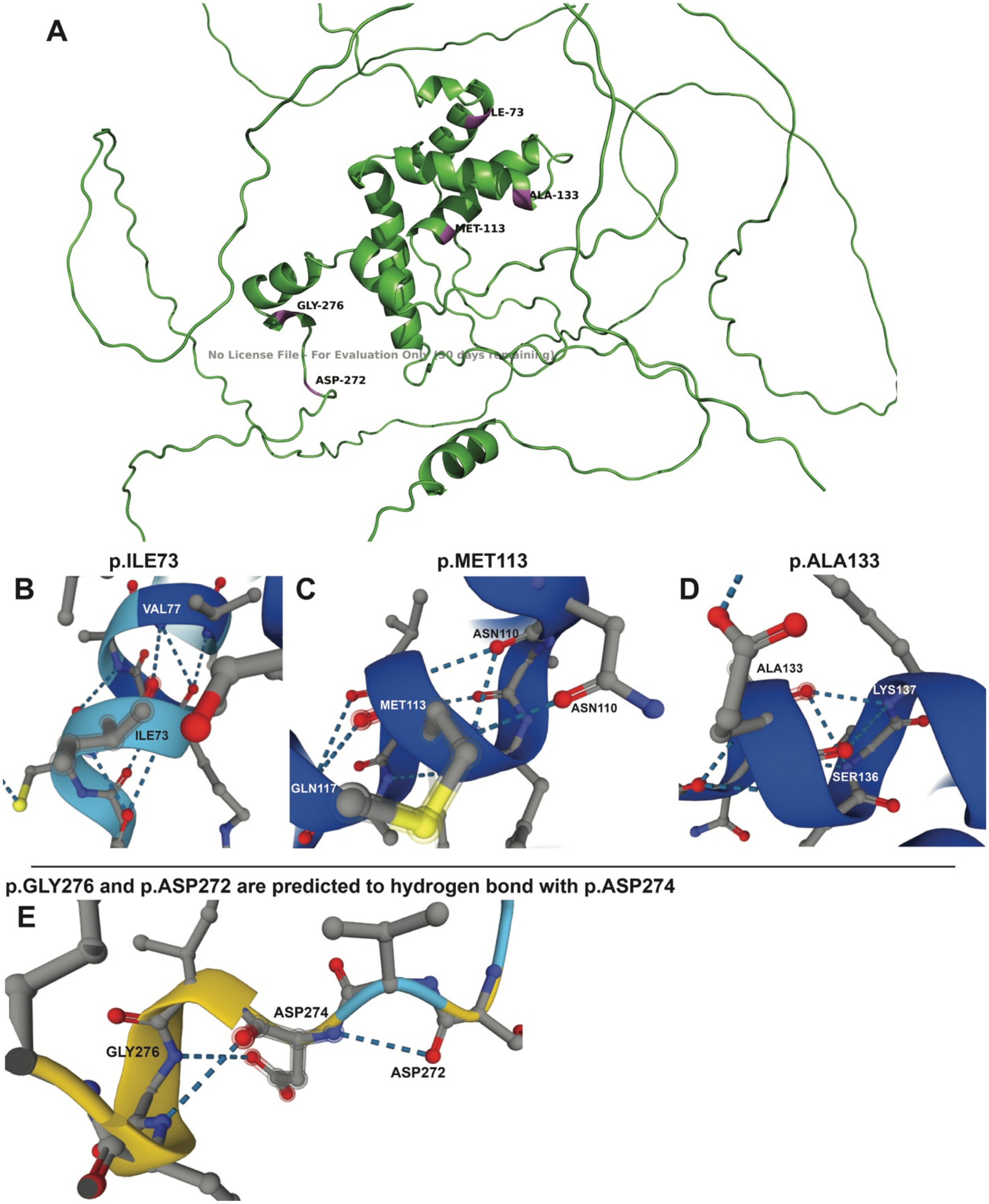

**Supplemental Figure 1. The 3D structures of SOX9 are predicted by AlphaFold.** (**A**) A model of human SOX9 was built by AlphaFold (htts://alphafold.ebi.ac.uk/). (**B**) Ile73 is predicted to be important for stabilizing alpha-helical interactions. (**C**) Met113 is predicted to stabilize alpha-helical interactions through a single hydrogen bond with Gln117 and two hydrogen bonds with Aan110. (**D**) Ala133 is predicted to stabilize alpha-helical interactions by a hydrogen bond with Ser136. (**E**) Gly276 is predicted to hydrogen bond with Asp274. Asp272 is also predicted to hydrogen bond with Asp274 by stabilizing an alpha-helical structure.

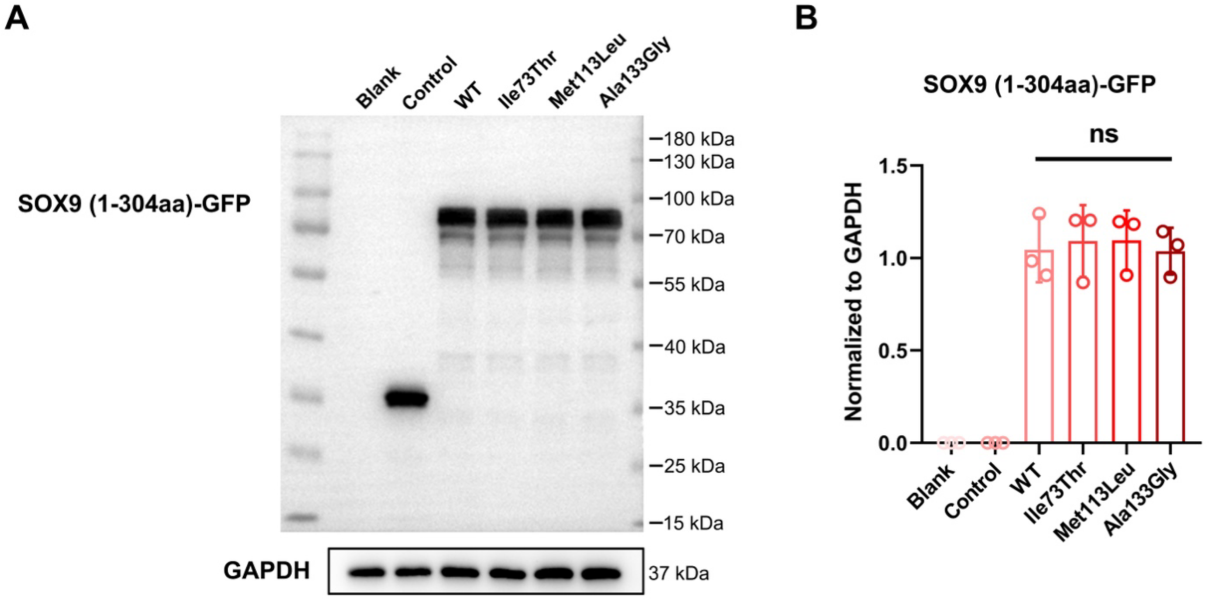


**Supplemental Figure 2. Equal amounts of SOX9 proteins were used for the EMSA assay.**

(A) Western blotting of anti-GFP was performed on total protein extracted from blank Cos-7 cells (Blank), Cos-7 cells transfected with empty EGFP expression vectors (Control), wild-type SOX9 (WT), or Ile73Thr, Met113Leu, and Ala133Gly SOX9 variants that were cloned into the EGFP expression vectors at 48 hours post-transfection. GAPDH is used as a loading control. No obvious changes in expression were observed among wild-type and variant SOX9 proteins. (B) Quantification analysis shows that comparable amounts of SOX9 were detected among wild-type and various SOX9 variant proteins. Bars are plotted with mean and SD. Each dot represents one experiment analyzed. The statistical difference is evaluated by one-way ANOVA followed by Tukey's multiple comparison test. ns: not significant. (n=3 experiments.)

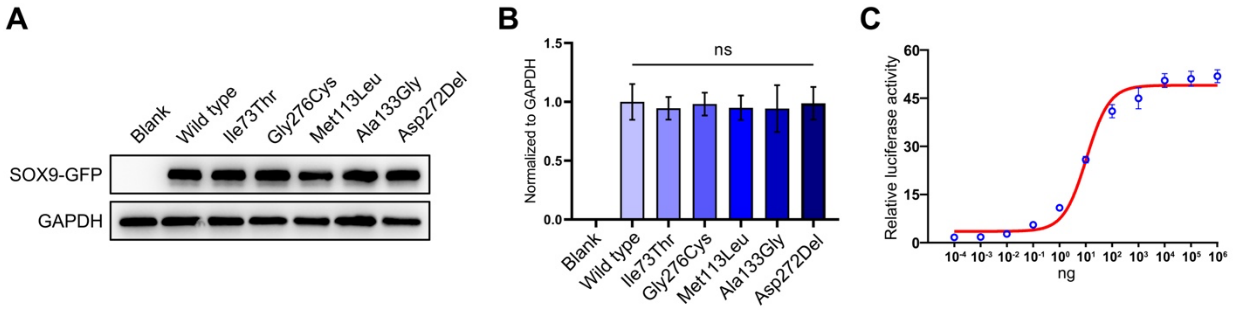


**Supplemental Figure 3. Optimization of the luciferase assay. (A)** Western blotting was performed on total protein extracted from blank Cos-7 cells (Blank), Cos-7 cells transfected with wild-type SOX9 construct (Wild type), or Ile73Thr, Gly276Cys, Met113Leu, Ala133Gly, and Asp276Del SOX9 variants at 48 hours post-transfection. GAPDH is used as a loading control. No obvious changes in expression were observed among wild-type and variant SOX9 proteins. **(B)** Quantification analysis shows that comparable amounts of SOX9 were detected among wild-type and various SOX9 variant proteins. Bars are plotted with mean and SD. Each dot represents one experiment analyzed. The statistical difference is evaluated by one-way ANOVA followed by Tukey's multiple comparison test. ns: not significant. (n=3 experiments.) **(C)** A standard curve was generated to optimize the transfection dose for the luciferase assays. SOX9-expression plasmids were diluted to a gradient of concentration with the final amount from $10^{-4}$ to $10^{6}$ ng. Relative luciferase activity was normalized to the Renilla internal control. The linear range of the SOX9-mediated reporter transactivation is between 1ng-100ng. 50ng of SOX9-expression plasmids was selected for luciferase assays shown in Figure 2B. n=3 experiments.

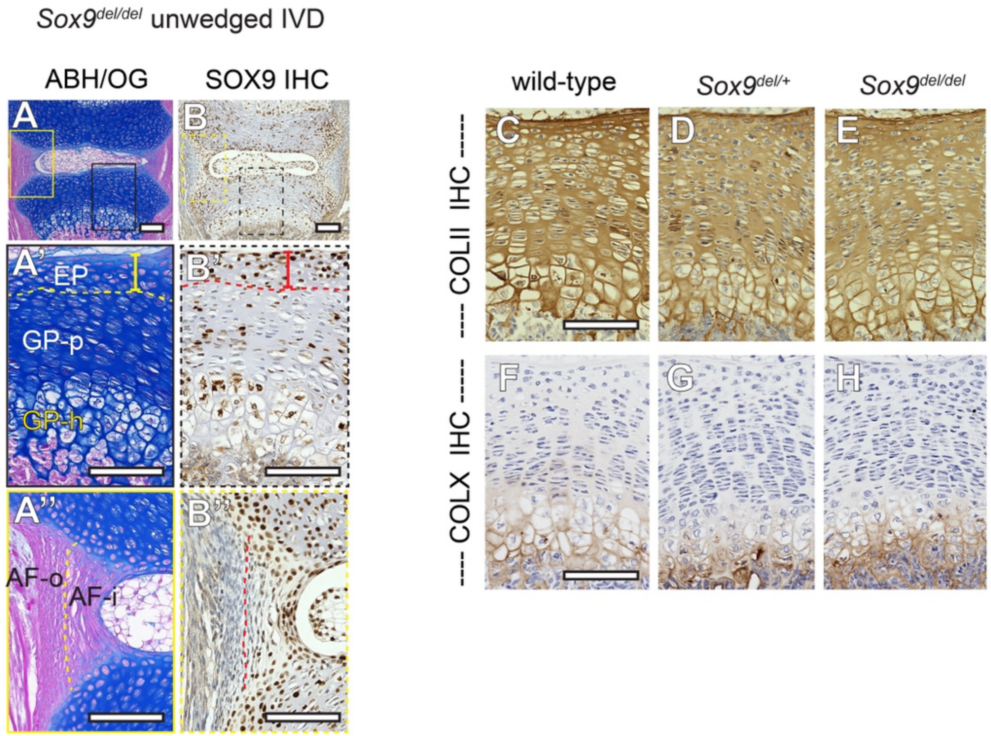


**Supplemental Figure 4. Expression of extracellular matrix components in P5 mouse tail sections. (A-B")** Representative Alcian Blue Hematoxylin /Orange G (ABH/OG) staining (**AA"**) and IHC staining of SOX9 (**B-B"**) on unwedged IVD in tail sections of P5 Sox9del/del mutant mice. The endplate is indicated with a dot line and a vertical bar in (**A'**) and (**B'**). The boundary between inner and outer annulus fibrosus is indicated with a dot-line in (**A"**) and (**B"**). (n=3.) C-H, IHC analysis of COLII **(C-E)** and COLX **(F-H)** on tail sections of P5 wildtype, *Sox9*$^{del/+}$*,* and *Sox9*$^{del/del}$ mice. n=3 for each genotype. Scale bars: 100µm. EP: endplate; GP-p: proliferative growth plate; GP-h: hypertrophic growth plate; AF-i: inner annulus fibrosus; AF-o: outer annulus fibrosus.

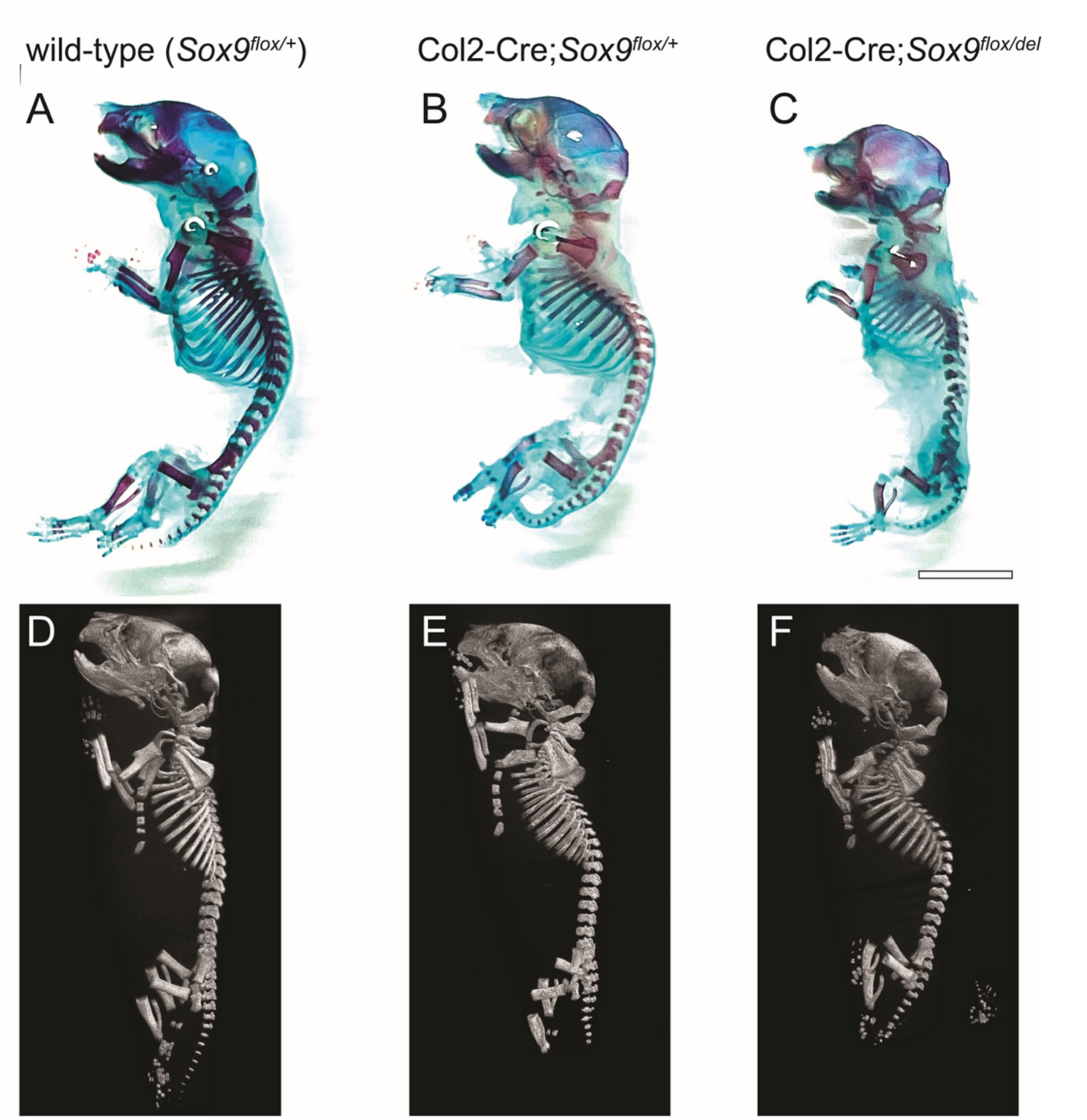


**Supplemental Figure 5. Assessment of skeletal development in *Sox9* loss of function mutant mice.** (**A-C**) Representative skeletal preps of Cre(-) wild-type (**A**), heterozygous *Col2a1-Cre; Sox9*$^{flox/+}$ mutant (**B**), and compound heterozygous *Col2a1-Cre; Sox9*$^{flox/del}$ mutant mice (**C**) at P1. **(D-F)** Representative MicroCT images of Cre(-) wild-type (**D**), heterozygous *Col2a1-Cre; Sox9*$^{flox/+}$ mutant (**E**), and compound heterozygous *Col2a1-Cre; Sox9*$^{flox/del}$ mutant mice (**F**) at P1. n=4 for each genotype. Scale bar: 1mm.

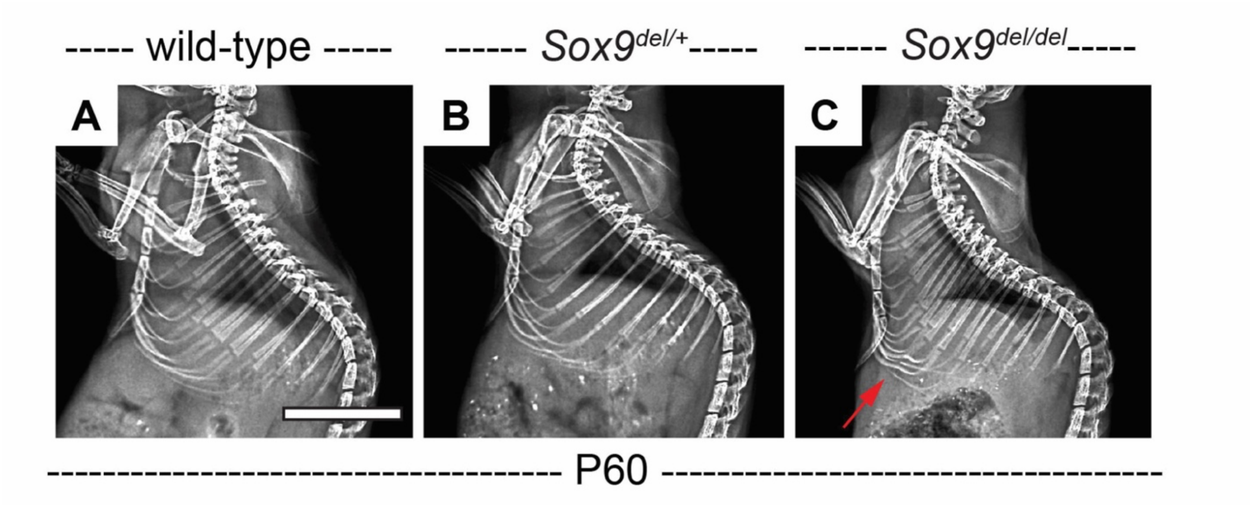


**Supplemental Figure 6. Young adult *Sox9$^{del/del}$* mice display mild rib cage deformities.**

(**A-C**) Representative sagittal X-ray images of wild-type (**A**), *Sox9$^{del/+}$* (**B**), and *Sox9$^{del/del}$* mice (**C**) at P60. Red arrows indicate the curvature of the distal ribs (**C**). n=4 for wild-type mice; 2 males, 2 females. n=8 for *Sox9$^{del/+}$* mice; 5 males, 3 females. n=7 for *Sox9$^{del/del}$* mice; 4 males, 3 females. Scale bar: 10mm.

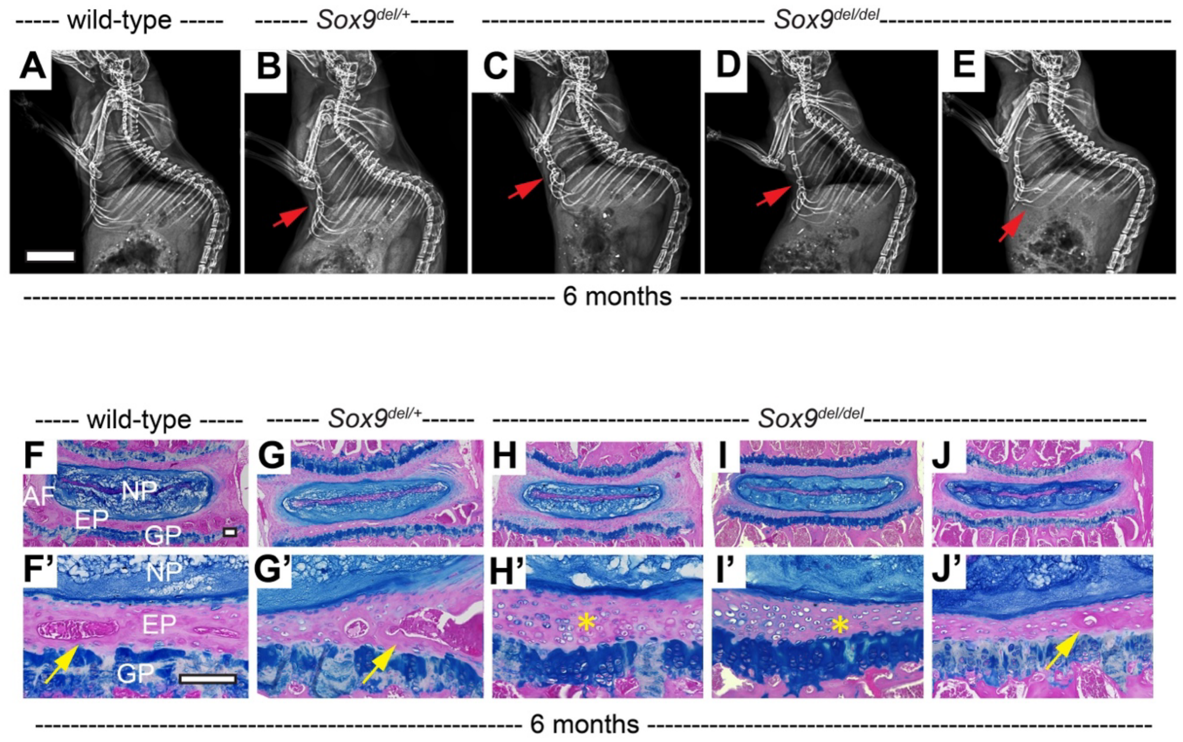


**Supplemental Figure 7. Mature adult *Sox9*$^{del/del}$ mice display rib cage deformities and impaired ossification in the endplate of the intervertebral discs.** (**A-E**) Additional sagittal X-ray images of wild-type (**A**), *Sox9*$^{del/+}$ (**B**), and *Sox9*$^{del/del}$ mice (**C-E**) at 6 months. Red arrows indicate the bending of the sternums (**B-D**) or the curvature of the distal ribs I. (**F-J'**) Additional ABH/OG staining of lumbar IVDs (L3/4 or L4/5) of wild-type (**F, F'**), *Sox9*$^{del/+}$ (**G, G'**) and *Sox9*$^{del/del}$ (**H-J'**) mice at 6 months. Wild-type and *Sox9*$^{del/+}$ mice consistently showed ossification within the endplate at 6 months (yellow arrows, **F'** and **G'**). On the contrary, *Sox9*$^{del/del}$ mice showed reduced endplate ossification (yellow arrow, **J'**) or persistency of hypertrophic chondrocyte-like cells within the endplate (yellow asterisks, **H'**, **I'**). n=3 for each genotype. Scale bars: 10mm in **A**; 100µm in **F-F'**. AF: annulus fibrosus; EP: endplate; GP: growth plate; NP: nucleus pulposus.

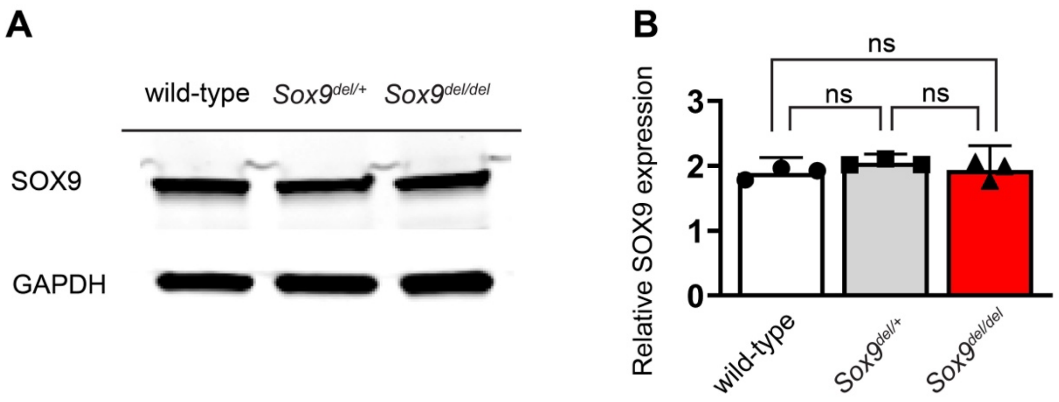


**Supplemental Figure 8. SOX9 protein expression in costal chondrocytes.** (**A**) Representative Western blot image of SOX9 performed on P5 costal chondrocytes isolated from three different genotypes of mice. GAPDH is used as a loading control (n=3 experiments). (**B**) Quantification analysis shows that comparable amounts of SOX9 protein were detected among three genotypes. Bars are plotted with mean and 95%CI. Each dot represents one experiment analyzed. The statistical difference is evaluated by one-way ANOVA followed by Tukey's multiple comparison test. ns: not significant (n=3 experiments).

**Supplementary Table 1. Primers used for amplifications of the DNA segments containing the candidate *SOX9* variants and RT-PCR quantification.**

| Primers | Primer sequence |
|---|---|
| *SOX9*-T218C-F | AAGTTCCCCGTGTGCACCCGCGAGGCGGTCAGCCA |
| *SOX9*-T218C-R | GTGCACACGGGGAACTTGTCCTCCTCGCTCTC |
| *SOX9*-G826T-F | TGGACATCTGCGAGCTGAGCAGCGACGTCATC |
| *SOX9*-G826T-R | CAGCTCGCAGATGTCCACGTCGCGGAAGTCGA |
| *SOX9*-A337C-F | ATGAACGCCTTCCTGGTGTGGGCGCAGGCGGC |
| *SOX9*-A337C-R | ACCAGGAAGGCGTTCATGGGCCGCTTGACGTG |
| *SOX9*-C398G-F | TTGCACAACGGCGAGCTCAGCAAGACGCTGGG |
| *SOX9*-C398G-R | AGCTCGCCGTTGTGCAAGTGCGGGTACTGGTC |
| *SOX9*-G442T-F | TTCTGAACTAGAGCGAGAAGCGGCCCTTCGTG |
| *SOX9*-G442T-R | CTCGCTCTAGTTCAGAAGTCTCCAGAGCTTGC |
| *SOX9*-H165Y-F | CGTGCAGTACAAGAAGGACCACCCGGATTACA |
| *SOX9*-H165Y-R | CCTTCTTGTACTGCACGCGCAGCCGCTCCGCC |
| *SOX9*-D272del-F | TATCGACTTCCGCGTGGACATCGGCGAGCTGAG |
| *SOX9*-D272del-R | TCCACGCGGAAGTCGATAGGGGGCTGTCTGCCC |
| *b-actin*-mouse-F | AGATGTGGATCAGCAAGCAG |
| *b-actin*-mouse-R | GCGCAAGTTAGGTTTTGTCA |
| *Sox9*-mouse-F | AGGAAGCTGGCAGACCAGTA |
| *Sox9*-mouse-R | CGTTCTTCACCGACTTCCTC |
| *Col2a1*-mouse-F | ACTGGTAAGTGGGGCAAGAC |
| *Col2a1*-mouse-R | CCACACCAAATTCCTGTTCA |
| *Acan*-mouse-F | CGTGTTTCCAAGGAAAAGGA |
| *Acan*-mouse-R | TGTGCTGATCAAAGTCCAG |
| *Col10a1*--mouse-F | CTTTGTGTGCCTTTCAATCG |
| *Col10a1*-mouse-R | GTGAGGTACAGCCTACCAGTTTT |

**Supplementary Table 2. Sequences of the SOX9 binding motifs used for EMSA and luciferase assay**

| | **Sequence** |
|---|---|
| B/C | GGGGACACAAGGCGTGCTTTGTCGGCCC |
| D/E | GGGTTTTCAAAGGCGCCTTGTTTGCCCC |

B/C and D/E bindings sites (underlined) of SOX9 were derived from the enhancer region of mouse *Col11a2* gene[1]. Probes for electrophoretic mobility shift assay (EMSA) were synthesized based on the B/C and D/E sequences. 4 tandem copies of the D/E sequence were cloned into a luciferase reporter vector pGL3-Basic to construct the luciferase reporter vector *Col11a2*-Luc.

**Supplemental Table 3.** Differential gene expression analysis of primary costal chondrocytes derived from *Sox9*$^{del/+}$ and *Sox9*$^{del/del}$ mutant mice.

| gene_id | H1 | H2 | H3 | M1 | M2 | M3 | log2FoldCh | pvalue | padj | gene_name | gene_chr | gene_start | gene_end | gene_stran | gene_lengt | gene_bioty | gene_descr | tf_family | H1_readco | H2_readco | H3_readco | M1_readco | M2_readco | M3_readco | H1_fpkm | H2_fpkm | H3_fpkm | M1_fpkm | M2_fpkm | M3_fpkm |
|---|---|---|---|---|---|---|---|---|---|---|---|---|---|---|---|---|---|---|---|---|---|---|---|---|---|---|---|---|---|---|
| ENSMUSG0 | 6446.784 | 6494.04 | 6527.945 | 14182.07 | 13632.11 | 13642.94 | -1.09039 | 5.47E-176 | 7.19E-172 | Col8a1 | 16 | 57624258 | 57754737 | - | 7833 | protein_co | collagen, ty | - | 6470 | 6447 | 7004 | 13614 | 13448 | 14068 | 43.44207 | 43.76051 | 43.98898 | 95.56681 | 91.86083 | 91.93382 |
| ENSMUSG0 | 82287.66 | 83815.12 | 80956.22 | 52326.99 | 54432.13 | 52060.01 | 0.637466 | 1.18E-86 | 7.77E-83 | Col3a1 | 1 | 45311538 | 45349706 | + | 9816 | protein_co | collagen, ty | - | 82584 | 83208 | 86860 | 50231 | 53697 | 53682 | 442.4821 | 450.6956 | 435.3226 | 281.3758 | 292.6957 | 279.9402 |
| ENSMUSG0 | 5666.593 | 5838.29 | 5599.642 | 10109.96 | 10528.19 | 11109.86 | -0.89247 | 1.64E-83 | 7.19E-80 | Errfi1 | 4 | 1.51E+08 | 1.51E+08 | + | 3582 | protein_co | ERBB recep | - | 5687 | 5796 | 6008 | 9705 | 10386 | 11456 | 83.50106 | 86.03112 | 82.51449 | 148.9771 | 155.1399 | 163.7112 |
| ENSMUSG0 | 182751.9 | 190628.8 | 184152.6 | 129664.8 | 130046.3 | 132916.8 | 0.505887 | 3.62E-61 | 1.19E-57 | Col1a1 | 11 | 94936224 | 94953042 | + | 6563 | protein_co | collagen, ty | - | 183410 | 189248 | 197582 | 124471 | 128290 | 137058 | 1469.789 | 1533.14 | 1481.054 | 1042.834 | 1045.903 | 1068.989 |
| ENSMUSG0 | 194497.6 | 196728 | 192962.1 | 144081.3 | 146085.9 | 147188.2 | 0.417619 | 1.90E-49 | 4.99E-46 | Col1a2 | 6 | 4504814 | 4541544 | + | 12426 | protein_co | collagen, ty | - | 195198 | 195303 | 207034 | 138310 | 144113 | 151774 | 826.187 | 835.6614 | 819.6647 | 612.0286 | 620.5441 | 625.2261 |
| ENSMUSG0 | 7872.649 | 7549.686 | 7683.664 | 4992.998 | 5007.63 | 4939.116 | 0.629146 | 3.64E-49 | 7.98E-46 | Col4a1 | 8 | 11198423 | 11312826 | - | 7464 | protein_co | collagen, ty | - | 7901 | 7495 | 8244 | 4793 | 4940 | 5093 | 55.67302 | 53.38913 | 54.33657 | 35.30899 | 35.41246 | 34.92795 |
| ENSMUSG0 | 3541.247 | 3708.865 | 3653.562 | 2093.871 | 1938.176 | 1809.619 | 0.900784 | 1.09E-44 | 2.04E-41 | Hspa1b | 17 | 34956436 | 34959238 | - | 2803 | protein_co | heat shock | - | 3554 | 3682 | 3920 | 2010 | 1912 | 1866 | 66.68508 | 69.84149 | 68.80007 | 39.4296 | 36.49771 | 34.07686 |
| ENSMUSG0 | 954.5624 | 988.1577 | 904.0701 | 2008.45 | 1886.478 | 2113.162 | -1.07808 | 5.07E-43 | 8.33E-40 | Srxn1 | 2 | 1.52E+08 | 1.52E+08 | + | 2820 | protein_co | sulfiredoxi | - | 958 | 981 | 970 | 1928 | 1861 | 2179 | 17.86696 | 18.49578 | 16.92188 | 37.59303 | 35.31003 | 39.55298 |
| ENSMUSG0 | 6788.553 | 6617.937 | 6778.662 | 9616.182 | 9571.264 | 9589.236 | -0.51156 | 2.53E-39 | 3.69E-36 | Gem | 4 | 11704457 | 11714752 | + | 2104 | protein_co | GTP bindin | - | 6813 | 6570 | 7273 | 9231 | 9442 | 9888 | 170.3048 | 166.0246 | 170.0567 | 241.2418 | 240.1149 | 240.5658 |
| ENSMUSG0 | 1149.859 | 1069.749 | 1135.214 | 2420.974 | 2112.531 | 2124.799 | -0.98856 | 1.19E-36 | 1.57E-33 | Aoc2 | 11 | 1.01E+08 | 1.01E+08 | + | 2543 | protein_co | amine oxid | - | 1154 | 1062 | 1218 | 2324 | 2084 | 2191 | 23.86678 | 22.20399 | 23.5628 | 50.25037 | 43.84824 | 44.10289 |
| ENSMUSG0 | 626.7429 | 661.7937 | 607.6842 | 1392.789 | 1302.592 | 1415.886 | -1.1169 | 3.68E-36 | 4.39E-33 | Fosl1 | 19 | 5447703 | 5455945 | + | 1707 | protein_co | fos-like anti | TF_bZIP | 629 | 657 | 652 | 1337 | 1285 | 1460 | 19.3799 | 20.46372 | 18.79057 | 43.06727 | 40.27823 | 43.78148 |
| ENSMUSG0 | 16253.47 | 16467.28 | 16559.4 | 11004.8 | 12129.82 | 11470.62 | 0.509993 | 1.56E-34 | 1.71E-31 | Capn6 | X | 1.44E+08 | 1.44E+08 | - | 7795 | protein_co | calpain 6 [S | - | 16312 | 16348 | 17767 | 10564 | 11966 | 11828 | 110.059 | 111.5068 | 112.1305 | 74.51809 | 82.13603 | 77.67232 |
| ENSMUSG0 | 2478.076 | 2558.533 | 2687.977 | 4367.961 | 3965.556 | 4417.372 | -0.72291 | 6.18E-33 | 6.25E-30 | Spry2 | 14 | 1.06E+08 | 1.06E+08 | - | 2087 | protein_co | sprouty RTI | - | 2487 | 2540 | 2884 | 4193 | 3912 | 4555 | 62.67404 | 64.70891 | 67.98275 | 110.4719 | 100.2945 | 111.7216 |
| ENSMUSG0 | 1169.787 | 1234.945 | 1283.407 | 403.1484 | 417.6404 | 558.5963 | 1.417747 | 1.18E-32 | 1.10E-29 | Tnxb | 17 | 34670535 | 34719815 | + | 12209 | protein_co | tenascin XE | - | 1174 | 1226 | 1377 | 387 | 412 | 576 | 5.057342 | 5.33904 | 5.548553 | 1.742932 | 1.805585 | 2.41498 |
| ENSMUSG0 | 6204.656 | 6168.683 | 6582.935 | 9031.773 | 9590.524 | 9001.546 | -0.54308 | 2.79E-32 | 2.45E-29 | Sulf2 | 2 | 1.66E+08 | 1.66E+08 | - | 5871 | protein_co | sulfatase 2 | - | 6227 | 6124 | 7063 | 8670 | 9461 | 9282 | 55.7829 | 55.45949 | 59.18382 | 81.20008 | 86.22352 | 80.92833 |
| ENSMUSG0 | 726.3841 | 734.3191 | 717.6639 | 1390.706 | 1656.37 | 1482.802 | -1.0564 | 1.71E-31 | 1.41E-28 | F3 | 3 | 1.22E+08 | 1.22E+08 | + | 2527 | protein_co | coagulatioı | - | 729 | 729 | 770 | 1335 | 1634 | 1529 | 15.17248 | 15.33822 | 14.99034 | 29.04862 | 34.59773 | 30.97229 |
| ENSMUSG0 | 1432.84 | 1231.923 | 1372.882 | 2398.056 | 2699.457 | 3075.189 | -1.01745 | 1.98E-30 | 1.53E-27 | Mt1 | 8 | 94179082 | 94180327 | + | 1246 | protein_co | metallothi | - | 1438 | 1223 | 1473 | 2302 | 2663 | 3171 | 60.69811 | 52.18687 | 58.15815 | 101.5867 | 114.3547 | 130.2715 |
| ENSMUSG0 | 862.8925 | 802.8152 | 853.7404 | 207.3037 | 272.6827 | 130.921 | 2.045672 | 2.24E-30 | 1.64E-27 | Acta1 | 8 | 1.24E+08 | 1.24E+08 | - | 1536 | protein_co | actin, alpha | - | 866 | 797 | 916 | 199 | 269 | 135 | 29.65248 | 27.58798 | 29.33798 | 7.123794 | 9.370481 | 4.498976 |
| ENSMUSG0 | 2169.188 | 2174.753 | 2149.264 | 3763.76 | 4848.481 | 3782.162 | -0.93268 | 7.45E-30 | 5.15E-27 | Prg4 | 1 | 1.5E+08 | 1.5E+08 | - | 4280 | protein_co | proteoglyca | - | 2177 | 2159 | 2306 | 3613 | 4783 | 3900 | 26.75155 | 26.82018 | 26.50583 | 46.41664 | 59.79397 | 46.64358 |
| ENSMUSG0 | 1799.519 | 1923.936 | 1782.043 | 963.5975 | 741.0076 | 911.5981 | 1.073372 | 8.24E-30 | 5.41E-27 | Hspa1a | 17 | 34969190 | 34972156 | - | 2967 | protein_co | heat shock | - | 1806 | 1910 | 1912 | 925 | 731 | 940 | 32.0136 | 34.22698 | 31.7027 | 17.14248 | 13.18259 | 16.21741 |
| ENSMUSG0 | 24646.24 | 24928.57 | 25032.49 | 31315.36 | 33330.14 | 32492.65 | -0.38073 | 4.40E-28 | 2.75E-25 | Thbs1 | 2 | 1.18E+08 | 1.18E+08 | + | 6073 | protein_co | thrombosp | - | 24735 | 24748 | 26858 | 30061 | 32880 | 33505 | 214.2116 | 216.6654 | 217.5686 | 272.1759 | 289.6872 | 282.4083 |
| ENSMUSG0 | 791.1509 | 722.2315 | 767.0615 | 1531.339 | 1370.509 | 1465.345 | -0.93719 | 3.23E-27 | 1.93E-24 | Hmox1 | 8 | 75093621 | 75100589 | + | 1751 | protein_co | heme oxyge | - | 794 | 717 | 823 | 1470 | 1352 | 1511 | 23.84892 | 21.77137 | 23.12276 | 46.16158 | 41.31344 | 44.17224 |
| ENSMUSG0 | 3758.465 | 3742.106 | 3649.834 | 5270.097 | 5530.694 | 5231.991 | -0.52399 | 4.23E-27 | 2.42E-24 | Hmgcr | 13 | 96648967 | 96670936 | - | 7275 | protein_co | 3-hydroxy- | - | 3772 | 3715 | 3916 | 5059 | 5456 | 5395 | 27.26924 | 27.15055 | 26.48107 | 38.23677 | 40.12751 | 37.96029 |
| ENSMUSG0 | 29835.56 | 33625.57 | 33820.61 | 16331.15 | 13618.93 | 15988.85 | 1.082456 | 4.44E-27 | 2.43E-24 | Gm26917 | 17 | 39843013 | 39846341 | + | 3329 | lincRNA | predicted g | - | 29943 | 33382 | 36287 | 15677 | 13435 | 16487 | 473.0596 | 533.1524 | 536.2449 | 258.9397 | 215.9358 | 253.5122 |
| ENSMUSG0 | 7132.315 | 7411.687 | 7070.387 | 4719.023 | 5218.478 | 4576.416 | 0.574585 | 8.09E-27 | 4.25E-24 | Col14a1 | 15 | 55307750 | 55520803 | + | 7013 | protein_co | collagen, ty | - | 7158 | 7358 | 7586 | 4530 | 5148 | 4719 | 53.6812 | 55.78388 | 53.2151 | 35.51762 | 39.27675 | 34.44429 |
| ENSMUSG0 | 3359.9 | 3479.202 | 3550.106 | 5096.129 | 5327.956 | 4860.563 | -0.5568 | 9.95E-26 | 5.03E-23 | Ldlr | 9 | 21723483 | 21749919 | + | 9622 | protein_co | low density | - | 3372 | 3454 | 3809 | 4892 | 5256 | 5012 | 18.43132 | 19.08577 | 19.47473 | 27.95571 | 29.22744 | 26.66348 |
| ENSMUSG0 | 2351.532 | 2392.329 | 2463.358 | 3501.245 | 3847.968 | 3630.876 | -0.60729 | 2.35E-25 | 1.14E-22 | Myc | 15 | 61985391 | 61990374 | + | 2717 | protein_co | myelocytor | bHLH | 2360 | 2375 | 2643 | 3361 | 3796 | 3744 | 45.68322 | 46.47579 | 47.85567 | 68.0187 | 74.75451 | 70.53705 |
| ENSMUSG0 | 971.5014 | 1006.289 | 936.6912 | 1604.26 | 1828.697 | 1829.015 | -0.85287 | 4.96E-25 | 2.33E-22 | Tlr2 | 3 | 83836272 | 83841767 | - | 3014 | protein_co | toll-like rec | - | 975 | 999 | 1005 | 1540 | 1804 | 1886 | 17.01358 | 17.62281 | 16.40396 | 28.09486 | 32.02537 | 32.03093 |
| ENSMUSG0 | 3037.063 | 3151.83 | 3010.46 | 4437.757 | 4332.512 | 4719.945 | -0.55255 | 2.24E-24 | 1.02E-21 | Vasn | 16 | 4639941 | 4650802 | + | 3007 | protein_co | vasorin [So | - | 3048 | 3129 | 3230 | 4260 | 4274 | 4867 | 53.31088 | 55.32545 | 52.84392 | 77.89788 | 76.05047 | 82.85124 |
| ENSMUSG0 | 4134.112 | 4055.375 | 3894.026 | 2472.018 | 2678.17 | 2789.102 | 0.60556 | 2.74E-24 | 1.20E-21 | Igfbp5 | 1 | 72857932 | 72874884 | - | 6006 | protein_co | insulin-like | - | 4149 | 4026 | 4178 | 2373 | 2642 | 2876 | 36.33226 | 35.64029 | 34.22229 | 21.7251 | 23.53685 | 24.51177 |
| ENSMUSG0 | 2922.475 | 3158.881 | 2860.403 | 4404.422 | 4310.211 | 4589.993 | -0.5736 | 3.44E-24 | 1.46E-21 | Rgs2 | 1 | 1.44E+08 | 1.44E+08 | - | 3627 | protein_co | regulator o | - | 2933 | 3136 | 3069 | 4228 | 4252 | 4733 | 42.53034 | 45.97072 | 41.62701 | 64.09688 | 62.72584 | 66.79747 |
| ENSMUSG0 | 4765.837 | 4698.03 | 4720.737 | 6407.663 | 6363.948 | 6389.915 | -0.43388 | 5.12E-24 | 2.10E-21 | Atp8b2 | 3 | 89939481 | 89963508 | - | 8561 | protein_co | ATPase, cla | - | 4783 | 4664 | 5065 | 6151 | 6278 | 6589 | 29.38395 | 28.96589 | 29.10589 | 39.50669 | 39.23717 | 39.39727 |
| ENSMUSG0 | 10138.49 | 9795.957 | 10558.05 | 13198.68 | 13659.48 | 13471.29 | -0.40328 | 2.70E-23 | 1.07E-20 | Sox9 | 11 | 1.13E+08 | 1.13E+08 | + | 4135 | protein_co | SRY (sex det | HMG | 10175 | 9725 | 11328 | 12670 | 13475 | 13891 | 129.4176 | 125.0452 | 134.7732 | 168.4809 | 174.3629 | 171.9606 |
| ENSMUSG0 | 2457.151 | 2272.461 | 2304.913 | 3467.909 | 3931.091 | 3542.625 | -0.63736 | 3.41E-23 | 1.32E-20 | Slc7a1 | 5 | 1.48E+08 | 1.48E+08 | - | 11530 | protein_co | solute carri | - | 2466 | 2256 | 2473 | 3329 | 3878 | 3653 | 11.24859 | 10.4031 | 10.55166 | 15.87574 | 17.99614 | 16.21778 |
| ENSMUSG0 | 6009.359 | 6175.734 | 6289.345 | 4504.428 | 4008.131 | 4360.154 | 0.521305 | 4.41E-23 | 1.65E-20 | Tubb2a | 13 | 34074274 | 34078007 | - | 1610 | protein_co | tubulin, be | - | 6031 | 6131 | 6748 | 4324 | 3954 | 4496 | 197.0143 | 202.4689 | 206.1936 | 147.6758 | 131.4049 | 142.9458 |
| ENSMUSG0 | 35189.27 | 36037.04 | 33359.25 | 24864.98 | 27304.76 | 24169.96 | 0.454169 | 7.02E-23 | 2.56E-20 | Postn | 3 | 54361109 | 54391037 | + | 8702 | protein_co | periostin, c | - | 35316 | 35776 | 35792 | 23869 | 26936 | 24923 | 213.4454 | 218.5876 | 202.3451 | 150.822 | 165.6208 | 146.6062 |
| ENSMUSG0 | 31471.66 | 30367.97 | 34490.74 | 68128.95 | 56107.76 | 66855.05 | -0.98819 | 5.34E-22 | 1.90E-19 | Eln | 5 | 1.35E+08 | 1.35E+08 | - | 8098 | protein_co | elastin [Sou | - | 31585 | 30148 | 37006 | 65400 | 55350 | 68938 | 205.1339 | 197.94 | 224.8124 | 444.068 | 365.7132 | 435.7646 |
| ENSMUSG0 | 13369.85 | 14164.6 | 14439.02 | 17809.37 | 19704.11 | 19679.85 | -0.44638 | 8.36E-22 | 2.89E-19 | Dlk1 | 12 | 1.09E+08 | 1.09E+08 | + | 5657 | protein_co | delta like n | - | 13418 | 14062 | 15492 | 17096 | 19438 | 20293 | 124.7487 | 132.1642 | 134.7247 | 166.172 | 183.8511 | 183.6247 |
| ENSMUSG0 | 3434.631 | 3545.683 | 3512.825 | 2407.431 | 2043.6 | 2323.605 | 0.631274 | 1.12E-21 | 3.78E-19 | Mfap4 | 11 | 61485431 | 61488900 | + | 1922 | protein_co | microfibril | - | 3447 | 3520 | 3769 | 2311 | 2016 | 2396 | 94.32401 | 97.37379 | 96.47142 | 66.11439 | 56.12262 | 63.81232 |
| ENSMUSG0 | 1330.21 | 1350.784 | 1277.815 | 2126.165 | 2003.052 | 2152.923 | -0.66648 | 1.48E-21 | 4.86E-19 | Map2k3 | 11 | 60932033 | 60952811 | + | 3314 | protein_co | mitogen-ac | - | 1335 | 1341 | 1371 | 2041 | 1976 | 2220 | 21.18669 | 21.51439 | 20.35218 | 33.86413 | 31.90327 | 34.29032 |
| ENSMUSG0 | 303.9056 | 290.1014 | 413.8218 | 65.6288 | 84.13629 | 92.12959 | 2.057838 | 1.95E-21 | 6.26E-19 | Hmcn2 | 2 | 31314415 | 31460738 | + | 16073 | protein_co | hemicentir | - | 305 | 288 | 444 | 63 | 83 | 95 | 0.998015 | 0.952683 | 1.358976 | 0.215523 | 0.276301 | 0.30255 |
| ENSMUSG0 | 3170.582 | 3300.91 | 3480.204 | 4599.225 | 4748.125 | 4745.159 | -0.50174 | 2.98E-21 | 9.32E-19 | Serpine1 | 5 | 1.37E+08 | 1.37E+08 | - | 3876 | protein_co | serine (or c | - | 3182 | 3277 | 3734 | 4415 | 4684 | 4893 | 43.17683 | 44.95163 | 47.39324 | 62.63202 | 64.65974 | 64.61934 |
| ENSMUSG0 | 3668.788 | 3828.734 | 3490.456 | 2483.477 | 2572.746 | 2500.106 | 0.539963 | 5.43E-21 | 1.66E-18 | Tnmd | X | 1.34E+08 | 1.34E+08 | + | 1353 | protein_co | tenomodul | - | 3682 | 3801 | 3745 | 2384 | 2538 | 2578 | 143.1266 | 149.3664 | 136.1695 | 96.88531 | 100.3679 | 97.53404 |
| ENSMUSG0 | 575.926 | 526.816 | 556.4225 | 1082.354 | 984.2933 | 1067.734 | -0.91761 | 7.67E-21 | 2.29E-18 | Chac1 | 2 | 1.19E+08 | 1.19E+08 | + | 1637 | protein_co | ChaC, catio | - | 578 | 523 | 597 | 1039 | 971 | 1101 | 18.57007 | 16.98657 | 17.9412 | 34.89927 | 31.7374 | 34.42784 |
| ENSMUSG0 | 259.067 | 299.167 | 299.182 | 67.71226 | 77.04046 | 40.73098 | 2.211775 | 1.18E-20 | 3.45E-18 | Myl1 | 1 | 66924295 | 66945404 | - | 2074 | protein_co | myosin, ligl | - | 260 | 297 | 321 | 65 | 76 | 42 | 6.593241 | 7.613783 | 7.614163 | 1.723273 | 1.960675 | 1.036601 |
| ENSMUSG0 | 3193.499 | 3104.487 | 3082.227 | 4319 | 4600.127 | 4278.692 | -0.49263 | 1.41E-20 | 4.02E-18 | Cdon | 9 | 35421128 | 35507652 | + | 7715 | protein_co | cell adhesic | - | 3205 | 3082 | 3307 | 4146 | 4538 | 4412 | 21.84874 | 21.23975 | 21.08745 | 29.549 | 31.47236 | 29.27323 |
| ENSMUSG0 | 986.4476 | 961.968 | 939.4873 | 1556.34 | 1792.204 | 1582.689 | -0.77207 | 2.30E-20 | 6.42E-18 | Fnip2 | 3 | 79455974 | 79567796 | - | 8364 | protein_co | folliculin ir | - | 990 | 955 | 1008 | 1494 | 1768 | 1632 | 6.225232 | 6.070747 | 5.928877 | 9.821686 | 11.31017 | 9.98797 |
| ENSMUSG0 | 1926.064 | 2114.315 | 2049.536 | 2913.711 | 3050.194 | 3148.893 | -0.58161 | 2.54E-20 | 6.96E-18 | Trib1 | 15 | 59648350 | 59657099 | + | 6248 | protein_co | tribbles pse | - | 1933 | 2099 | 2199 | 2797 | 3009 | 3247 | 16.27141 | 17.86176 | 17.31451 | 24.61506 | 25.76808 | 26.60188 |
| ENSMUSG0 | 8673.764 | 8553.961 | 9437.746 | 11959.03 | 11826.72 | 11688.82 | -0.41162 | 2.77E-20 | 7.43E-18 | Klf4 | 4 | 55527143 | 55532466 | - | 3049 | protein_co | Kruppel-lik | zf-C2H2 | 8705 | 8492 | 10126 | 11480 | 11667 | 12053 | 150.157 | 148.0831 | 163.3828 | 207.0303 | 204.7399 | 202.3526 |
| ENSMUSG0 | 1221.601 | 1112.055 | 1112.845 | 575.0333 | 654.8439 | 640.0582 | 0.881672 | 8.07E-20 | 2.12E-17 | Rgs5 | 1 | 1.7E+08 | 1.7E+08 | + | 4546 | protein_co | regulator o | - | 1226 | 1104 | 1194 | 552 | 646 | 660 | 14.18389 | 12.91197 | 12.92114 | 6.676658 | 7.603331 | 7.431656 |
| ENSMUSG0 | 5253.082 | 5191.606 | 4957.473 | 6655.594 | 6919.45 | 7091.069 | -0.42437 | 1.35E-19 | 3.47E-17 | Sgms2 | 3 | 1.31E+08 | 1.31E+08 | - | 6227 | protein_co | sphingomy | - | 5272 | 5154 | 5319 | 6389 | 6826 | 7312 | 44.52776 | 44.00665 | 42.02203 | 56.41616 | 58.65273 | 60.10746 |
| ENSMUSG0 | 5735.346 | 5779.867 | 6103.871 | 8262.979 | 7681.745 | 7747.614 | -0.42706 | 1.96E-19 | 4.95E-17 | Plec | 15 | 76170974 | 76232574 | - | 18113 | protein_co | plectin [Sou | - | 5756 | 5738 | 6549 | 7932 | 7578 | 7989 | 16.7134 | 16.84314 | 17.78732 | 24.07918 | 22.38541 | 22.57736 |
| ENSMUSG0 | 2294.736 | 2132.446 | 2314.233 | 3266.856 | 3174.878 | 3311.816 | -0.53271 | 2.13E-19 | 5.28E-17 | Itga5 | 15 | 1.03E+08 | 1.03E+08 | - | 4933 | protein_co | integrin alp | - | 2303 | 2117 | 2483 | 3136 | 3132 | 3415 | 24.55371 | 22.81721 | 24.76233 | 34.95541 | 33.97124 | 35.43649 |
| ENSMUSG0 | 261.0599 | 237.7219 | 268.4249 | 528.1556 | 658.8987 | 646.8467 | -1.25712 | 2.24E-19 | 5.45E-17 | Hivep3 | 4 | 1.2E+08 | 1.2E+08 | + | 17627 | protein_co | human imr | zf-C2H2 | 262 | 236 | 288 | 507 | 650 | 667 | 0.781731 | 0.711847 | 0.803785 | 1.581536 | 1.973039 | 1.93695 |
| ENSMUSG0 | 3086.883 | 3316.02 | 3100.867 | 4731.524 | 4509.908 | 4286.451 | -0.50932 | 2.63E-19 | 6.28E-17 | Acly | 11 | 1E+08 | 1.01E+08 | - | 5279 | protein_co | ATP citrate | - | 3098 | 3292 | 3327 | 4542 | 4449 | 4420 | 30.86484 | 33.15591 | 31.00466 | 47.30913 | 45.09325 | 42.85897 |
| ENSMUSG0 | 1517.535 | 1549.222 | 1596.569 | 2416.807 | 2283.844 | 2303.24 | -0.58651 | 2.95E-19 | 6.92E-17 | Scube3 | 17 | 28142141 | 28174854 | + | 8844 | protein_co | signal pepti | - | 1523 | 1538 | 1713 | 2320 | 2253 | 2375 | 9.057024 | 9.246139 | 9.528719 | 14.4241 | 13.63055 | 13.7463 |
| ENSMUSG0 | 3044.038 | 3101.466 | 3164.245 | 4216.911 | 4397.388 | 4193.351 | -0.46006 | 3.55E-19 | 8.20E-17 | Hipk1 | 3 | 1.04E+08 | 1.04E+08 | - | 10914 | protein_co | homeodom | - | 3055 | 3079 | 3395 | 4048 | 4338 | 4324 | 14.72182 | 14.99956 | 15.30318 | 20.39417 | 21.26701 | 20.28023 |
| ENSMUSG0 | 1659.025 | 1699.309 | 1645.035 | 2741.826 | 2382.172 | 2535.988 | -0.6144 | 8.31E-19 | 1.88E-16 | Klhl21 | 4 | 1.52E+08 | 1.52E+08 | + | 3986 | protein_co | kelch-like 2 | - | 1665 | 1687 | 1765 | 2632 | 2350 | 2615 | 21.96905 | 22.50249 | 21.78378 | 36.30764 | 31.54506 | 33.58192 |
| ENSMUSG0 | 136.5084 | 160.1601 | 99.72732 | 379.1886 | 459.2017 | 408.2796 | -1.65552 | 1.35E-18 | 3.01E-16 | Plet1 | 9 | 50494525 | 50505482 | + | 1947 | protein_co | placenta ex | - | 137 | 159 | 107 | 364 | 453 | 421 | 3.700743 | 4.341942 | 2.703608 | 10.27981 | 12.44896 | 11.06846 |
| ENSMUSG0 | 947.5875 | 970.0264 | 861.1967 | 472.9441 | 391.2844 | 507.1977 | 1.018121 | 3.96E-18 | 8.67E-16 | Col5a3 | 9 | 20770050 | 20815067 | - | 6809 | protein_co | collagen, ty | - | 951 | 963 | 924 | 454 | 386 | 523 | 7.345672 | 7.519618 | 6.675972 | 3.666249 | 3.033226 | 3.931782 |
| ENSMUSG0 | 21288.34 | 21249.92 | 21281.06 | 25312.93 | 25988.99 | 26395.61 | -0.28391 | 7.87E-18 | 1.69E-15 | Emp1 | 6 | 1.35E+08 | 1.35E+08 | + | 5935 | protein_co | epithelial n | - | 21365 | 21096 | 22833 | 24299 | 25638 | 27218 | 189.3287 | 188.9871 | 189.2641 | 225.1216 | 231.1342 | 234.7505 |
| ENSMUSG0 | 4732.955 | 4754.439 | 4641.515 | 6209.735 | 6149.045 | 6023.336 | -0.37966 | 8.62E-18 | 1.83E-15 | Slc3a2 | 19 | 8706882 | 8723369 | - | 3529 | protein_co | solute carri | - | 4750 | 4720 | 4980 | 5961 | 6066 | 6211 | 70.79071 | 71.11204 | 69.42303 | 92.87887 | 91.97114 | 90.09091 |
| ENSMUSG0 | 504.1843 | 508.6847 | 463.2194 | 975.0565 | 843.3903 | 1006.637 | -0.93709 | 9.37E-18 | 1.96E-15 | Otor | 2 | 1.43E+08 | 1.43E+08 | + | 949 | protein_co | otoraplin [S | - | 506 | 505 | 497 | 936 | 832 | 1038 | 28.04262 | 28.29293 | 25.76417 | 54.23243 | 46.90919 | 55.98894 |
| ENSMUSG0 | 3640.888 | 3647.42 | 3884.705 | 4828.405 | 5315.792 | 5361.942 | -0.4728 | 2.98E-17 | 6.12E-15 | Slc7a5 | 8 | 1.22E+08 | 1.22E+08 | - | 3708 | protein_co | solute carri | - | 3654 | 3621 | 4168 | 4635 | 5244 | 5529 | 51.82785 | 51.92083 | 55.29857 | 68.73208 | 75.67001 | 76.32696 |
| ENSMUSG0 | 777.2011 | 760.5088 | 794.0904 | 1204.236 | 1374.564 | 1310.18 | -0.73794 | 7.85E-17 | 1.59E-14 | Moxd1 | 10 | 24223517 | 24302790 | + | 3198 | protein_co | monooxyge | - | 780 | 755 | 852 | 1156 | 1356 | 1351 | 12.82775 | 12.55224 | 13.10651 | 19.87599 | 22.68726 | 21.62459 |
| ENSMUSG0 | 1110.003 | 1138.245 | 1248.922 | 691.7068 | 682.2136 | 694.3662 | 0.758076 | 1.40E-16 | 2.79E-14 | Cilp | 9 | 65265180 | 65280605 | + | 4252 | protein_co | cartilage in | - | 1114 | 1130 | 1340 | 664 | 673 | 716 | 13.77927 | 14.12986 | 15.50377 | 8.58666 | 8.468814 | 8.619674 |
| ENSMUSG0 | 256.0778 | 222.6125 | 266.5609 | 77.0878 | 73.99939 | 57.21733 | 1.841236 | 1.47E-16 | 2.88E-14 | Abca8a | 11 | 1.1E+08 | 1.1E+08 | - | 6272 | protein_co | ATP-bindin | - | 257 | 221 | 286 | 74 | 73 | 59 | 2.15507 | 1.873437 | 2.243292 | 0.648747 | 0.622756 | 0.481523 |
| ENSMUSG0 | 1170.784 | 1120.114 | 1220.961 | 1796.979 | 1859.108 | 1729.127 | -0.61634 | 1.68E-16 | 3.26E-14 | Stk40 | 4 | 1.26E+08 | 1.26E+08 | + | 4117 | protein_co | serine/thre | - | 1175 | 1112 | 1310 | 1725 | 1834 | 1783 | 15.01037 | 14.36074 | 15.65368 | 23.03869 | 23.83523 | 22.16877 |
| ENSMUSG0 | 599.8398 | 630.5675 | 619.8006 | 1132.357 | 983.2796 | 1122.041 | -0.80729 | 1.77E-16 | 3.36E-14 | Inhba | 13 | 16011851 | 16031621 | + | 6390 | protein_co | inhibin bet | - | 602 | 626 | 665 | 1087 | 970 | 1157 | 4.954844 | 5.208663 | 5.119726 | 9.353586 | 8.122163 | 9.268375 |
| ENSMUSG0 | 2761.057 | 2678.401 | 2692.638 | 1720.933 | 2003.052 | 1764.039 | 0.567319 | 3.28E-16 | 6.15E-14 | Nid1 | 13 | 13437551 | 13512269 | + | 6943 | protein_co | nidogen 1 [ | - | 2771 | 2659 | 2889 | 1652 | 1976 | 1819 | 20.99054 | 20.36217 | 20.4704 | 13.08315 | 15.22792 | 13.41086 |
| ENSMUSG0 | 3908.923 | 3781.391 | 3937.831 | 4966.955 | 5147.519 | 5122.405 | -0.38991 | 3.76E-16 | 6.95E-14 | Cyp51 | 5 | 4081145 | 4104746 | - | 4415 | protein_co | cytochrom | - | 3923 | 3754 | 4225 | 4768 | 5078 | 5282 | 46.73281 | 45.20811 | 47.07842 | 59.38202 | 61.54075 | 61.24051 |
| ENSMUSG0 | 6902.144 | 6839.542 | 7095.552 | 8926.559 | 8939.735 | 8534.11 | -0.34124 | 3.84E-16 | 7.02E-14 | Xylt1 | 7 | 1.17E+08 | 1.18E+08 | + | 9651 | protein_co | xylosyltran | - | 6927 | 6790 | 7613 | 8569 | 8819 | 8800 | 37.74915 | 37.40677 | 38.80694 | 48.82107 | 48.89313 | 46.67469 |
| ENSMUSG0 | 4929.249 | 5403.138 | 5116.85 | 3310.609 | 3949.337 | 3281.753 | 0.551453 | 6.08E-16 | 1.09E-13 | Mest | 6 | 30723547 | 30748465 | + | 3945 | protein_co | mesoderm | - | 4947 | 5364 | 5490 | 3178 | 3896 | 3384 | 65.95219 | 72.29272 | 68.46226 | 44.29517 | 52.84121 | 43.90909 |
| ENSMUSG0 | 15262.04 | 15101.39 | 14968.42 | 12333.01 | 12384.25 | 11480.32 | 0.324708 | 7.19E-16 | 1.28E-13 | Col5a2 | 1 | 45374321 | 45503282 | - | 9591 | protein_co | collagen, ty | - | 15317 | 14992 | 16060 | 11839 | 12217 | 11838 | 83.99321 | 83.10908 | 82.3773 | 67.87355 | 68.15559 | 63.18086 |
| ENSMUSG0 | 2941.407 | 3035.991 | 2999.276 | 4115.864 | 3931.091 | 3947.996 | -0.41809 | 8.28E-16 | 1.45E-13 | Slc20a1 | 2 | 1.29E+08 | 1.29E+08 | + | 5580 | protein_co | solute carri | - | 2952 | 3014 | 3218 | 3951 | 3878 | 4071 | 27.8238 | 28.71851 | 28.3712 | 38.9334 | 37.18557 | 37.34547 |
| ENSMUSG0 | 503.1879 | 476.4512 | 570.403 | 900.0522 | 979.2248 | 930.024 | -0.85714 | 8.80E-16 | 1.52E-13 | Rasgef1b | 5 | 99217426 | 99729065 | - | 13805 | protein_co | RasGEF dor | - | 505 | 473 | 612 | 864 | 966 | 959 | 1.92393 | 1.821703 | 2.180926 | 3.441334 | 3.744049 | 3.555931 |
| ENSMUSG0 | 3213.428 | 3305.947 | 3263.041 | 4242.954 | 4482.538 | 4273.843 | -0.41018 | 9.00E-16 | 1.54E-13 | Piezo1 | 8 | 1.22E+08 | 1.23E+08 | - | 9935 | protein_co | piezo-type | - | 3225 | 3282 | 3501 | 4073 | 4422 | 4407 | 17.07246 | 17.564 | 17.33605 | 22.54218 | 23.81506 | 22.70629 |
| ENSMUSG0 | 3861.095 | 3639.362 | 3675.93 | 4747.15 | 5054.26 | 5183.502 | -0.42324 | 1.48E-15 | 2.49E-13 | Tiam2 | 17 | 3326573 | 3531344 | + | 12741 | protein_co | T cell lympl | - | 3875 | 3613 | 3944 | 4557 | 4986 | 5345 | 15.99567 | 15.07708 | 15.22858 | 19.6664 | 20.93869 | 21.47411 |
| ENSMUSG0 | 9621.351 | 9707.315 | 9410.717 | 11717.35 | 12347.76 | 11671.36 | -0.31441 | 3.55E-15 | 5.90E-13 | Egr1 | 18 | 34859823 | 34864984 | + | 4484 | protein_co | early growt | zf-C2H2 | 9656 | 9637 | 10097 | 11248 | 12181 | 12035 | 113.2572 | 114.2692 | 110.7778 | 137.9301 | 145.351 | 137.3889 |
| ENSMUSG0 | 343.762 | 305.2108 | 287.9976 | 89.58853 | 114.547 | 120.2534 | 1.52855 | 3.76E-15 | 6.18E-13 | Ptp4a1 | 1 | 30940302 | 30949309 | - | 5541 | protein_co | protein tyr | - | 345 | 303 | 309 | 86 | 113 | 124 | 3.274653 | 2.907417 | 2.743445 | 0.853414 | 1.091167 | 1.145525 |
| ENSMUSG0 | 6225.58 | 6312.726 | 5865.271 | 7592.107 | 8440.999 | 8165.591 | -0.39515 | 5.60E-15 | 9.09E-13 | Herpud1 | 8 | 94386438 | 94395377 | + | 2333 | protein_co | homocyste | - | 6248 | 6267 | 6293 | 7288 | 8327 | 8420 | 140.8512 | 142.8229 | 132.6994 | 171.7683 | 190.9742 | 184.7432 |
| ENSMUSG0 | 858.9069 | 849.1508 | 862.1287 | 1269.865 | 1374.564 | 1390.672 | -0.6509 | 6.06E-15 | 9.71E-13 | Zfand2a | 5 | 1.39E+08 | 1.39E+08 | - | 3407 | protein_co | zinc finger, | - | 862 | 843 | 925 | 1219 | 1356 | 1434 | 13.30667 | 13.15553 | 13.35659 | 19.67347 | 21.29553 | 21.54508 |
| ENSMUSG0 | 809.0863 | 915.6324 | 822.0514 | 1294.867 | 1394.838 | 1387.763 | -0.67944 | 7.14E-15 | 1.13E-12 | Fam43a | 16 | 30599723 | 30602797 | + | 3075 | protein_co | family with | - | 812 | 909 | 882 | 1243 | 1376 | 1431 | 13.88818 | 15.71707 | 14.11073 | 22.22672 | 23.94276 | 23.82131 |
| ENSMUSG0 | 38351.89 | 39287.58 | 40398.89 | 46932.93 | 51315.03 | 47732.83 | -0.30651 | 7.45E-15 | 1.16E-12 | Chad | 11 | 94565047 | 94569127 | + | 1680 | protein_co | chondroad | - | 38490 | 39003 | 43345 | 45053 | 50622 | 49220 | 1204.961 | 1234.359 | 1269.275 | 1474.565 | 1612.244 | 1499.697 |

| gene_id | H1 | H2 | H3 | M1 | M2 | M3 | log2FoldCh | pvalue | padj | gene_name | gene_chr | gene_start | gene_end | gene_stran | gene_lengt | gene_bioty | gene_descr | tf_family | H1_readcou | H2_readcou | H3_readcou | M1_readco | M2_readco | M3_readco | H1_fpkm | H2_fpkm | H3_fpkm | M1_fpkm | M2_fpkm | M3_fpkm |
|---|---|---|---|---|---|---|---|---|---|---|---|---|---|---|---|---|---|---|---|---|---|---|---|---|---|---|---|---|---|---|
| ENSMUSG0 | 8840.164 | 9477.652 | 9148.817 | 11293.36 | 11782.12 | 11310.6 | -0.32416 | 7.50E-15 | 1.16E-12 | Serpinf1 | 11 | 75409769 | 75422701 | - | 2359 | protein_co | serine (or c | - | 8872 | 9409 | 9816 | 10841 | 11623 | 11663 | 197.8007 | 212.0647 | 204.7069 | 252.6916 | 263.6277 | 253.0774 |
| ENSMUSG0 | 16421.86 | 16728.17 | 17363.74 | 20809.54 | 22297.13 | 20304.39 | -0.32799 | 7.57E-15 | 1.16E-12 | A2m | 6 | 1.22E+08 | 1.22E+08 | + | 7021 | protein_co | alpha-2-ma | - | 16481 | 16607 | 18630 | 19976 | 21996 | 20937 | 123.4579 | 125.7607 | 130.5388 | 156.444 | 167.6276 | 152.6464 |
| ENSMUSG0 | 3304.101 | 3574.895 | 3176.362 | 4398.172 | 4604.181 | 4652.06 | -0.44174 | 8.46E-15 | 1.28E-12 | Ifrd1 | 12 | 40201567 | 40248504 | - | 5858 | protein_co | interferon- | - | 3316 | 3549 | 3408 | 4222 | 4542 | 4797 | 29.77142 | 32.21139 | 28.62043 | 39.62948 | 41.48571 | 41.91712 |
| ENSMUSG0 | 3390.789 | 3386.53 | 3402.845 | 5253.429 | 4292.978 | 5386.187 | -0.55275 | 9.49E-15 | 1.42E-12 | Ier5 | 1 | 1.55E+08 | 1.55E+08 | - | 3276 | protein_co | immediate | - | 3403 | 3362 | 3651 | 5043 | 4235 | 5554 | 54.63267 | 54.56405 | 54.82692 | 84.64368 | 69.16881 | 86.78268 |
| ENSMUSG0 | 1388.998 | 1527.061 | 1324.416 | 855.2579 | 827.1713 | 939.7219 | 0.692825 | 1.74E-14 | 2.56E-12 | Lamb1 | 12 | 31265234 | 31329644 | + | 9917 | protein_co | laminin B1 | - | 1394 | 1516 | 1421 | 821 | 816 | 969 | 7.392933 | 8.127775 | 7.049197 | 4.552105 | 4.402614 | 5.001664 |
| ENSMUSG0 | 1799.519 | 1740.608 | 1851.946 | 2491.811 | 2732.909 | 2463.254 | -0.51152 | 2.23E-14 | 3.26E-12 | Med13l | 5 | 1.19E+08 | 1.19E+08 | + | 15427 | protein_co | mediator c | - | 1806 | 1728 | 1987 | 2392 | 2696 | 2540 | 6.157021 | 5.955456 | 6.336396 | 8.525683 | 9.350594 | 8.427977 |
| ENSMUSG0 | 1121.96 | 1040.537 | 1142.67 | 2084.496 | 1575.275 | 1836.773 | -0.73346 | 2.32E-14 | 3.35E-12 | Gadd45a | 6 | 67035096 | 67037457 | - | 1764 | protein_co | growth arr | - | 1126 | 1033 | 1226 | 2001 | 1554 | 1894 | 33.57176 | 31.13541 | 34.19147 | 62.37319 | 47.13606 | 54.96073 |
| ENSMUSG0 | 458.3494 | 379.7507 | 423.1421 | 185.4274 | 154.0809 | 192.9873 | 1.243759 | 2.54E-14 | 3.63E-12 | Col6a6 | 9 | 1.06E+08 | 1.06E+08 | - | 9166 | protein_co | collagen, ty | - | 460 | 377 | 454 | 178 | 152 | 199 | 2.639443 | 2.186826 | 2.436699 | 1.067799 | 0.887288 | 1.111333 |
| ENSMUSG0 | 334.7943 | 330.3932 | 368.1523 | 125.0072 | 152.0535 | 138.6793 | 1.313513 | 3.86E-14 | 5.46E-12 | Rgs16 | 1 | 1.54E+08 | 1.54E+08 | + | 2339 | protein_co | regulator o | - | 336 | 328 | 395 | 120 | 150 | 143 | 7.555155 | 7.455837 | 8.307929 | 2.820983 | 3.431325 | 3.129514 |
| ENSMUSG0 | 5799.116 | 5899.735 | 5881.116 | 7210.835 | 7399.939 | 7237.507 | -0.31358 | 4.04E-14 | 5.65E-12 | Fgfr1 | 8 | 25513654 | 25575718 | + | 9875 | protein_co | fibroblast g | - | 5820 | 5857 | 6310 | 6922 | 7300 | 7463 | 30.99704 | 31.53486 | 31.43534 | 38.54286 | 39.55365 | 38.68543 |
| ENSMUSG0 | 1203.665 | 1217.821 | 1111.913 | 718.7917 | 761.2814 | 736.067 | 0.672647 | 4.99E-14 | 6.90E-12 | Igfbp3 | 11 | 7206086 | 7213923 | - | 3130 | protein_co | insulin-like | - | 1208 | 1209 | 1193 | 690 | 751 | 759 | 20.29817 | 20.53689 | 18.7509 | 12.12144 | 12.83797 | 12.41276 |
| ENSMUSG0 | 323.8338 | 302.1889 | 315.0265 | 118.7569 | 95.28689 | 134.8001 | 1.430807 | 5.73E-14 | 7.84E-12 | Mfap5 | 6 | 1.23E+08 | 1.23E+08 | + | 3589 | protein_co | microfibril | - | 325 | 300 | 338 | 114 | 94 | 139 | 4.762601 | 4.444272 | 4.633073 | 1.746549 | 1.401378 | 1.982496 |
| ENSMUSG0 | 6485.644 | 6254.303 | 6165.385 | 7669.194 | 8795.791 | 8345.971 | -0.39231 | 6.34E-14 | 8.59E-12 | Bzw1 | 1 | 58392898 | 58407353 | + | 4033 | protein_co | basic leucir | - | 6509 | 6209 | 6615 | 7362 | 8677 | 8606 | 84.88293 | 81.85519 | 80.69145 | 100.373 | 115.1177 | 109.2306 |
| ENSMUSG0 | 5377.634 | 4527.797 | 4541.787 | 3414.781 | 3386.739 | 3498.015 | 0.488069 | 8.97E-14 | 1.20E-11 | Xist | X | 1.03E+08 | 1.03E+08 | - | 17946 | lincRNA | inactive X s | - | 5397 | 4495 | 4873 | 3278 | 3341 | 3607 | 15.81682 | 13.31726 | 13.35841 | 10.04363 | 9.961153 | 10.28844 |
| ENSMUSG0 | 2662.412 | 2634.08 | 2663.745 | 3511.662 | 3631.039 | 3405.886 | -0.40605 | 9.74E-14 | 1.29E-11 | 2900097C | 2 | 1.56E+08 | 1.56E+08 | - | 4915 | lincRNA | RIKEN cDN/ | - | 2672 | 2615 | 2858 | 3371 | 3582 | 3512 | 28.59218 | 28.28792 | 28.60649 | 37.71245 | 38.99446 | 36.57649 |
| ENSMUSG0 | 2302.707 | 2276.49 | 2341.262 | 3079.345 | 3197.179 | 3010.213 | -0.42414 | 1.22E-13 | 1.61E-11 | Sec14l1 | 11 | 1.17E+08 | 1.17E+08 | + | 7176 | protein_co | SEC14-like | - | 2311 | 2260 | 2512 | 2956 | 3154 | 3104 | 16.9376 | 16.74476 | 17.22119 | 22.65017 | 23.5169 | 22.14167 |
| ENSMUSG0 | 1797.527 | 1774.856 | 1729.85 | 2514.729 | 2454.144 | 2361.427 | -0.46724 | 1.29E-13 | 1.68E-11 | Stard4 | 18 | 33199355 | 33213862 | - | 5586 | protein_co | StAR-relate | - | 1804 | 1762 | 1856 | 2414 | 2421 | 2435 | 16.98517 | 16.77095 | 16.34568 | 23.76215 | 23.18968 | 22.31357 |
| ENSMUSG0 | 1114.985 | 914.6251 | 1042.943 | 559.4074 | 634.5701 | 588.6596 | 0.78545 | 1.63E-13 | 2.10E-11 | Rn7sk | 9 | 78175303 | 78175633 | - | 331 | misc_RNA | RNA, 7SK, r | - | 1119 | 908 | 1119 | 537 | 626 | 607 | 177.8019 | 145.8514 | 166.3137 | 89.20635 | 101.1922 | 93.87107 |
| ENSMUSG0 | 202.2716 | 215.5614 | 189.2023 | 68.75398 | 59.80773 | 47.51947 | 1.787443 | 1.87E-13 | 2.38E-11 | Mylpf | 7 | 1.27E+08 | 1.27E+08 | + | 1062 | protein_co | myosin ligh | - | 203 | 214 | 203 | 66 | 59 | 49 | 10.05324 | 10.71376 | 9.403671 | 3.417188 | 2.972544 | 2.361797 |
| ENSMUSG0 | 722.3985 | 833.0341 | 681.3147 | 403.1484 | 408.5172 | 409.2494 | 0.872923 | 1.97E-13 | 2.49E-11 | Marcks | 10 | 37133375 | 37138920 | - | 4048 | protein_co | myristoylat | - | 725 | 827 | 731 | 387 | 403 | 422 | 9.419587 | 10.8622 | 8.883882 | 5.256781 | 5.326787 | 5.336334 |
| ENSMUSG0 | 2045.633 | 1908.827 | 1739.17 | 2594.942 | 2826.169 | 2745.462 | -0.52084 | 2.90E-13 | 3.63E-11 | Csgalnact2 | 6 | 1.18E+08 | 1.18E+08 | - | 5438 | protein_co | chondroiti | - | 2053 | 1895 | 1866 | 2491 | 2788 | 2831 | 19.85565 | 18.52776 | 16.88101 | 25.18744 | 27.4318 | 26.64844 |
| ENSMUSG0 | 397.5683 | 440.1885 | 379.3366 | 684.4147 | 738.9802 | 879.5952 | -0.92111 | 3.51E-13 | 4.35E-11 | Cd14 | 18 | 36725074 | 36726736 | - | 1572 | protein_co | CD14 antig | - | 399 | 437 | 407 | 657 | 729 | 907 | 13.34918 | 14.78025 | 12.73702 | 22.98065 | 24.8128 | 29.53424 |
| ENSMUSG0 | 12155.23 | 12134.9 | 12898.38 | 15232.13 | 15517.57 | 14766.92 | -0.29141 | 3.60E-13 | 4.43E-11 | Wif1 | 10 | 1.21E+08 | 1.21E+08 | + | 2427 | protein_co | Wnt inhibit | - | 12199 | 12047 | 13839 | 14622 | 15308 | 15227 | 264.3558 | 263.9137 | 280.5181 | 331.2733 | 337.4811 | 321.1557 |
| ENSMUSG0 | 1544.438 | 1545.193 | 1390.59 | 2007.408 | 2326.419 | 2329.424 | -0.57323 | 4.04E-13 | 4.92E-11 | Has2 | 15 | 56665627 | 56694539 | - | 4237 | protein_co | hyaluronar | - | 1550 | 1534 | 1492 | 1927 | 2295 | 2402 | 19.24011 | 19.24951 | 17.32352 | 25.00764 | 28.98178 | 29.01921 |
| ENSMUSG0 | 2845.752 | 2978.575 | 3273.293 | 2125.123 | 1955.409 | 2256.69 | 0.521699 | 4.44E-13 | 5.36E-11 | Pcolce | 5 | 1.38E+08 | 1.38E+08 | - | 3823 | protein_co | procollage | - | 2856 | 2957 | 3512 | 2040 | 1929 | 2327 | 39.29056 | 41.12442 | 45.19351 | 29.34103 | 26.99782 | 31.15754 |
| ENSMUSG0 | 5533.074 | 6093.136 | 5496.187 | 4117.947 | 3926.022 | 4546.353 | 0.443297 | 5.25E-13 | 6.27E-11 | Dusp1 | 17 | 26505590 | 26508519 | - | 2697 | protein_co | dual specifi | - | 5553 | 6049 | 5897 | 3953 | 3873 | 4688 | 108.2882 | 119.2492 | 107.5663 | 80.59263 | 76.83646 | 88.977 |
| ENSMUSG0 | 2331.603 | 2328.869 | 2442.853 | 3424.157 | 3014.715 | 3293.391 | -0.45408 | 6.85E-13 | 8.11E-11 | S100a10 | 3 | 93555080 | 93564643 | + | 1212 | protein_co | S100 calciu | - | 2340 | 2312 | 2621 | 3287 | 2974 | 3396 | 101.5424 | 101.4234 | 106.3874 | 149.1237 | 131.2923 | 143.4287 |
| ENSMUSG0 | 2157.231 | 2087.118 | 1904.139 | 2749.118 | 2838.333 | 2952.026 | -0.47441 | 7.00E-13 | 8.22E-11 | Mapk6 | 9 | 75369062 | 75410005 | - | 6675 | protein_co | mitogen-ac | - | 2165 | 2072 | 2043 | 2639 | 2800 | 3044 | 17.05851 | 16.50408 | 15.05716 | 21.7389 | 22.44438 | 23.34342 |
| ENSMUSG0 | 1542.445 | 1389.062 | 1555.56 | 2025.117 | 2306.145 | 2214.989 | -0.54479 | 7.69E-13 | 8.94E-11 | Crybg3 | 16 | 59490775 | 59600979 | - | 12711 | protein_co | beta-gamm | - | 1548 | 1379 | 1669 | 1944 | 2275 | 2284 | 6.405095 | 5.768161 | 6.459554 | 8.409419 | 9.576405 | 9.197874 |
| ENSMUSG0 | 1635.112 | 1490.799 | 1578.861 | 2544.939 | 2112.531 | 2285.784 | -0.56126 | 7.84E-13 | 9.04E-11 | C4b | 17 | 34728380 | 34743882 | - | 5695 | protein_co | complemer | - | 1641 | 1480 | 1694 | 2443 | 2084 | 2357 | 15.15476 | 13.81722 | 14.63341 | 23.58735 | 19.57964 | 21.18541 |
| ENSMUSG0 | 710.4415 | 752.4504 | 781.974 | 1185.485 | 1284.346 | 1107.495 | -0.67178 | 7.93E-13 | 9.07E-11 | Fem1b | 9 | 62791821 | 62811934 | - | 6785 | protein_co | feminizatio | - | 713 | 747 | 839 | 1138 | 1267 | 1142 | 5.526803 | 5.853607 | 6.083283 | 9.222356 | 9.991428 | 8.615636 |
| ENSMUSG0 | 698.4846 | 739.3555 | 666.4022 | 273.9742 | 417.6404 | 338.455 | 1.029929 | 8.27E-13 | 9.37E-11 | Lpl | 8 | 68880491 | 68907448 | + | 10478 | protein_co | lipoprotein | - | 701 | 734 | 715 | 263 | 412 | 349 | 3.518633 | 3.724521 | 3.357017 | 1.380152 | 2.103874 | 1.704975 |
| ENSMUSG0 | 7092.458 | 6953.367 | 7014.466 | 5254.471 | 5855.075 | 5188.351 | 0.36989 | 9.11E-13 | 1.02E-10 | Thbs2 | 17 | 14665500 | 14694235 | - | 7003 | protein_co | thrombosp | - | 7118 | 6903 | 7526 | 5044 | 5776 | 5350 | 53.45744 | 52.40908 | 52.8696 | 39.60412 | 44.13101 | 39.10576 |
| ENSMUSG0 | 1209.644 | 1192.639 | 1268.494 | 1845.94 | 1714.15 | 2240.204 | -0.66021 | 1.40E-12 | 1.56E-10 | Tnfrsf12a | 17 | 23675445 | 23677468 | - | 1932 | protein_co | tumor necr | - | 1214 | 1184 | 1361 | 1772 | 1691 | 2310 | 33.04805 | 32.58347 | 34.65588 | 50.43198 | 46.83142 | 61.20345 |
| ENSMUSG0 | 258.0706 | 355.5756 | 241.396 | 602.1182 | 598.0773 | 586.72 | -1.06459 | 1.51E-12 | 1.67E-10 | Met | 6 | 17463800 | 17573980 | + | 7679 | protein_co | met proto- | - | 259 | 353 | 259 | 578 | 590 | 605 | 1.773901 | 2.444122 | 1.659285 | 4.138783 | 4.111006 | 4.03294 |
| ENSMUSG0 | 23122.73 | 23189.98 | 23528.19 | 18821.92 | 20247.45 | 18687.76 | 0.274102 | 1.60E-12 | 1.76E-10 | Col12a1 | 9 | 79598991 | 79718831 | - | 15228 | protein_co | collagen, ty | - | 23206 | 23022 | 25244 | 18068 | 19974 | 19270 | 80.14783 | 80.38092 | 81.55324 | 65.24041 | 70.18156 | 64.77537 |
| ENSMUSG0 | 1245.515 | 1385.033 | 1349.581 | 1916.778 | 1829.711 | 2118.011 | -0.55938 | 1.75E-12 | 1.90E-10 | Ywhag | 5 | 1.36E+08 | 1.36E+08 | - | 5500 | protein_co | tyrosine 3- | - | 1250 | 1375 | 1448 | 1840 | 1805 | 2184 | 11.95313 | 13.29208 | 12.95185 | 18.3952 | 17.55963 | 20.32643 |
| ENSMUSG0 | 1947.985 | 1867.527 | 1976.838 | 1414.665 | 1324.893 | 1375.155 | 0.493568 | 1.81E-12 | 1.95E-10 | Cald1 | 6 | 34598500 | 34775473 | + | 12041 | protein_co | caldesmon | - | 1955 | 1854 | 2121 | 1358 | 1307 | 1418 | 8.539227 | 8.186532 | 8.665707 | 6.201356 | 5.807829 | 6.02816 |
| ENSMUSG0 | 4388.197 | 4468.367 | 4417.827 | 5448.232 | 5512.448 | 5552.02 | -0.31499 | 1.87E-12 | 2.00E-10 | Hspa9 | 18 | 34937414 | 34954357 | - | 3932 | protein_co | heat shock | - | 4404 | 4436 | 4740 | 5230 | 5438 | 5725 | 58.90717 | 59.98336 | 59.30492 | 73.13708 | 73.99911 | 74.53033 |
| ENSMUSG0 | 2118.371 | 2186.84 | 2202.389 | 2857.457 | 2851.511 | 2903.537 | -0.40427 | 1.88E-12 | 2.00E-10 | Pacsin2 | 15 | 83375607 | 83464606 | - | 4908 | protein_co | protein kin | - | 2126 | 2171 | 2363 | 2743 | 2813 | 2994 | 22.78206 | 23.51842 | 23.68564 | 30.73058 | 30.66663 | 31.22614 |
| ENSMUSG0 | 6791.542 | 6983.586 | 6932.447 | 8502.576 | 8364.972 | 9311.878 | -0.33839 | 2.09E-12 | 2.20E-10 | Rcan1 | 16 | 92391953 | 92470867 | - | 3085 | protein_co | regulator o | - | 6816 | 6933 | 7438 | 8162 | 8252 | 9602 | 116.2007 | 119.4865 | 118.6115 | 145.4758 | 143.1215 | 159.3227 |
| ENSMUSG0 | 5104.617 | 4991.154 | 4813.008 | 6135.772 | 6996.49 | 6324.939 | -0.38425 | 2.44E-12 | 2.53E-10 | Cd109 | 9 | 78615546 | 78716253 | + | 6900 | protein_co | CD109 anti | - | 5123 | 4955 | 5164 | 5890 | 6902 | 6522 | 39.04897 | 38.18101 | 36.81824 | 46.93704 | 53.5213 | 48.38411 |
| ENSMUSG0 | 7518.923 | 7135.688 | 7665.023 | 5952.428 | 6086.196 | 5908.901 | 0.314658 | 2.44E-12 | 2.53E-10 | Col4a2 | 8 | 11312805 | 11449287 | + | 6619 | protein_co | collagen, ty | - | 7546 | 7084 | 8224 | 5714 | 6004 | 6093 | 59.9596 | 56.9035 | 61.12468 | 47.4676 | 48.53434 | 47.1205 |
| ENSMUSG0 | 999.4009 | 1076.8 | 1069.04 | 1513.629 | 1485.056 | 1674.819 | -0.57151 | 3.44E-12 | 3.53E-10 | Tmem2 | 19 | 21778342 | 21858327 | + | 6667 | protein_co | transmemb | - | 1003 | 1069 | 1147 | 1453 | 1465 | 1727 | 7.912338 | 8.525111 | 8.463673 | 11.98353 | 11.75731 | 13.25968 |
| ENSMUSG0 | 15075.71 | 15674.54 | 14384.97 | 10536.03 | 12663.02 | 11037.13 | 0.398675 | 3.74E-12 | 3.81E-10 | Omd | 13 | 49582462 | 49592822 | + | 2249 | protein_co | osteomodu | - | 15130 | 15561 | 15434 | 10114 | 12492 | 11381 | 353.8212 | 367.8755 | 337.6097 | 247.2766 | 297.1963 | 259.0371 |
| ENSMUSG0 | 2225.984 | 2264.402 | 2204.253 | 2834.539 | 3010.66 | 3028.639 | -0.40678 | 4.34E-12 | 4.39E-10 | Eif1a | 18 | 46597704 | 46610225 | + | 2879 | protein_co | eukaryotic | - | 2234 | 2248 | 2365 | 2721 | 2970 | 3123 | 40.81087 | 41.51523 | 40.41247 | 51.96804 | 55.19702 | 55.52664 |
| ENSMUSG0 | 3855.117 | 4209.492 | 4120.509 | 3113.722 | 3194.138 | 2888.02 | 0.406271 | 5.54E-12 | 5.55E-10 | Cdkn1c | 7 | 1.43E+08 | 1.43E+08 | - | 2039 | protein_co | cyclin-depe | - | 3869 | 4179 | 4421 | 2989 | 3151 | 2978 | 99.79662 | 108.9702 | 106.6668 | 80.60429 | 82.686 | 74.76159 |
| ENSMUSG0 | 252.0922 | 305.2108 | 233.9398 | 570.8664 | 473.3934 | 565.3848 | -1.02556 | 5.65E-12 | 5.62E-10 | Il6ra | 3 | 89864059 | 89913196 | - | 8642 | protein_co | interleukin | - | 253 | 303 | 251 | 548 | 467 | 583 | 1.539716 | 1.864151 | 1.428846 | 3.486709 | 2.891368 | 3.453229 |
| ENSMUSG0 | 818.054 | 821.9538 | 880.7693 | 1310.493 | 1198.182 | 1503.167 | -0.67045 | 6.04E-12 | 5.97E-10 | Efhd2 | 4 | 1.42E+08 | 1.42E+08 | - | 2381 | protein_co | EF hand do | - | 821 | 816 | 945 | 1258 | 1182 | 1550 | 18.13502 | 18.22148 | 19.52533 | 29.05164 | 26.56188 | 33.32295 |
| ENSMUSG0 | 2109.404 | 2206.986 | 2167.904 | 2866.833 | 2769.402 | 2965.603 | -0.4078 | 8.22E-12 | 8.07E-10 | Rin2 | 2 | 1.46E+08 | 1.46E+08 | + | 11271 | protein_co | Ras and Rat | - | 2117 | 2191 | 2326 | 2752 | 2732 | 3058 | 9.87854 | 10.33553 | 10.1525 | 13.42565 | 12.96938 | 13.8882 |
| ENSMUSG0 | 3852.128 | 3791.464 | 3722.532 | 4689.855 | 5073.52 | 4733.522 | -0.35107 | 8.64E-12 | 8.41E-10 | Dnajb9 | 12 | 44205559 | 44210326 | - | 2549 | protein_co | DnaJ heat s | - | 3866 | 3764 | 3994 | 4502 | 5005 | 4881 | 79.76757 | 78.51137 | 77.08398 | 97.11473 | 105.0594 | 98.01895 |
| ENSMUSG0 | 2132.321 | 1982.359 | 2261.107 | 2828.289 | 2843.401 | 3040.277 | -0.4502 | 1.20E-11 | 1.16E-09 | Lgals3 | 14 | 47367751 | 47386160 | + | 1858 | protein_co | lectin, gala | - | 2140 | 1968 | 2426 | 2715 | 2805 | 3135 | 60.57625 | 56.31605 | 64.23489 | 80.34773 | 80.77705 | 86.37 |
| ENSMUSG0 | 346.7513 | 408.9623 | 401.7054 | 712.5413 | 626.4606 | 725.3993 | -0.83469 | 1.33E-11 | 1.28E-09 | Sipa1l2 | 8 | 1.25E+08 | 1.26E+08 | - | 9617 | protein_co | signal-indu | - | 348 | 406 | 431 | 684 | 618 | 748 | 1.903154 | 2.244601 | 2.204771 | 3.910803 | 3.438347 | 3.981375 |
| ENSMUSG0 | 8431.636 | 8317.246 | 8700.51 | 10468.31 | 9931.124 | 10690.91 | -0.2888 | 1.62E-11 | 1.55E-09 | Sqstm1 | 11 | 50199366 | 50210827 | - | 3356 | protein_co | sequestoso | - | 8462 | 8257 | 9335 | 10049 | 9797 | 11024 | 132.6128 | 130.8137 | 136.8416 | 164.6457 | 156.1967 | 168.1467 |
| ENSMUSG0 | 2853.723 | 2863.744 | 2947.082 | 3635.627 | 4253.444 | 3771.495 | -0.42838 | 1.67E-11 | 1.58E-09 | Scd1 | 19 | 44394451 | 44407709 | - | 4836 | protein_co | stearoyl-Co | - | 2864 | 2843 | 3162 | 3490 | 4196 | 3889 | 31.14735 | 31.25672 | 32.16633 | 39.68155 | 46.4248 | 41.16449 |
| ENSMUSG0 | 3084.891 | 3030.955 | 3229.488 | 4008.566 | 3902.708 | 4033.337 | -0.35386 | 1.76E-11 | 1.65E-09 | Ppp1r15b | 1 | 1.33E+08 | 1.33E+08 | + | 5594 | protein_co | protein pho | - | 3096 | 3009 | 3465 | 3848 | 3850 | 4159 | 29.10803 | 28.59911 | 30.4724 | 37.82353 | 36.82469 | 38.05726 |
| ENSMUSG0 | 7517.926 | 7165.906 | 7568.092 | 5415.939 | 6205.812 | 5766.343 | 0.355819 | 1.94E-11 | 1.80E-09 | Itih5l-ps | X | 1.51E+08 | 1.51E+08 | + | 4077 | transcribed | inter-alpha | - | 7545 | 7114 | 8120 | 5199 | 6122 | 5946 | 97.33138 | 92.77393 | 97.98085 | 70.11785 | 80.344 | 74.65438 |
| ENSMUSG0 | 2062.572 | 2200.943 | 2223.826 | 2824.122 | 3059.317 | 2820.135 | -0.42383 | 1.94E-11 | 1.80E-09 | Fdft1 | 14 | 63145150 | 63179578 | - | 4227 | protein_co | farnesyl dip | - | 2070 | 2185 | 2386 | 2711 | 3018 | 2908 | 25.75565 | 27.4835 | 27.76925 | 35.26523 | 38.20215 | 35.21545 |
| ENSMUSG0 | 1348.145 | 1364.887 | 1305.775 | 1800.104 | 1844.916 | 1936.661 | -0.47431 | 2.21E-11 | 2.03E-09 | Pex5 | 6 | 1.24E+08 | 1.24E+08 | - | 3943 | protein_co | peroxisoma | - | 1353 | 1355 | 1401 | 1728 | 1820 | 1997 | 18.04701 | 18.27113 | 17.47983 | 24.09719 | 24.69707 | 25.92522 |
| ENSMUSG0 | 2059.583 | 2137.483 | 2111.982 | 1587.592 | 1541.823 | 1531.291 | 0.437003 | 2.40E-11 | 2.19E-09 | Lamc1 | 1 | 1.53E+08 | 1.53E+08 | - | 8272 | protein_co | laminin, ga | - | 2067 | 2122 | 2266 | 1524 | 1521 | 1579 | 13.14209 | 13.63916 | 13.47644 | 10.13034 | 9.838287 | 9.771083 |
| ENSMUSG0 | 4986.044 | 4702.059 | 5421.625 | 6401.413 | 6374.085 | 6886.445 | -0.37979 | 2.41E-11 | 2.19E-09 | Slc2a1 | 4 | 1.19E+08 | 1.19E+08 | + | 3506 | protein_co | solute carri | - | 5004 | 4668 | 5817 | 6145 | 6288 | 7101 | 75.06539 | 70.78997 | 81.62309 | 96.3739 | 95.96247 | 103.6761 |
| ENSMUSG0 | 12939.4 | 12824.9 | 13089.44 | 15527.98 | 14896.18 | 15897.69 | -0.25365 | 2.53E-11 | 2.28E-09 | Atf4 | 15 | 80255184 | 80257541 | + | 2358 | protein_co | activating t | TF_bZIP | 12986 | 12732 | 14044 | 14906 | 14695 | 16393 | 289.6449 | 287.0818 | 293.0036 | 347.5896 | 333.4468 | 355.8654 |
| ENSMUSG0 | 1040.254 | 1032.479 | 1034.554 | 1466.752 | 1460.728 | 1478.922 | -0.50398 | 2.61E-11 | 2.33E-09 | Ddi2 | 4 | 1.42E+08 | 1.42E+08 | - | 9553 | protein_co | DNA-damag | - | 1044 | 1025 | 1110 | 1408 | 1441 | 1525 | 5.747713 | 5.704754 | 5.716222 | 8.104241 | 8.070957 | 8.171488 |
| ENSMUSG0 | 9181.934 | 9929.928 | 9691.259 | 8085.885 | 7492.185 | 7696.215 | 0.307557 | 2.66E-11 | 2.36E-09 | Jun | 4 | 95049034 | 95052222 | - | 3189 | protein_co | jun proto-c | TF_bZIP | 9215 | 9858 | 10398 | 7762 | 7391 | 7936 | 151.9761 | 164.3566 | 160.4062 | 133.8347 | 124.0079 | 127.385 |
| ENSMUSG0 | 2976.282 | 3082.327 | 2946.15 | 3765.843 | 4052.734 | 3787.011 | -0.36616 | 2.75E-11 | 2.42E-09 | Msmo1 | 8 | 64718139 | 64733792 | - | 6144 | protein_co | methylster | - | 2987 | 3060 | 3161 | 3615 | 3998 | 3905 | 25.56927 | 26.48031 | 25.31041 | 32.3524 | 34.81708 | 32.53426 |
| ENSMUSG0 | 1227.579 | 1180.551 | 1164.107 | 821.9226 | 716.679 | 809.7706 | 0.605058 | 2.79E-11 | 2.45E-09 | Sorbs2 | 8 | 45507788 | 45827906 | + | 27851 | protein_co | sorbin and | - | 1232 | 1172 | 1249 | 789 | 707 | 835 | 2.326506 | 2.237379 | 2.206214 | 1.557707 | 1.358249 | 1.534676 |
| ENSMUSG0 | 2296.729 | 2290.592 | 2488.523 | 3014.758 | 3119.125 | 3213.868 | -0.40156 | 3.59E-11 | 3.13E-09 | Aars | 8 | 1.11E+08 | 1.11E+08 | + | 9282 | protein_co | alanyl-tRN/ | - | 2305 | 2274 | 2670 | 2894 | 3077 | 3314 | 13.06062 | 13.02572 | 14.15128 | 17.14377 | 17.73727 | 18.27604 |
| ENSMUSG0 | 768.2334 | 679.9251 | 694.3631 | 379.1886 | 453.1196 | 408.2796 | 0.78801 | 4.11E-11 | 3.56E-09 | Cdh5 | 8 | 1.04E+08 | 1.04E+08 | + | 4203 | protein_co | cadherin 5 | - | 771 | 675 | 745 | 364 | 447 | 421 | 9.647823 | 8.538807 | 8.720126 | 4.762023 | 5.690481 | 5.12736 |
| ENSMUSG0 | 550.0192 | 511.7066 | 597.4319 | 955.2637 | 892.0475 | 844.6829 | -0.69731 | 5.29E-11 | 4.54E-09 | Aacs | 5 | 1.25E+08 | 1.26E+08 | + | 3854 | protein_co | acetoacety | - | 552 | 508 | 641 | 917 | 880 | 871 | 7.532891 | 7.008173 | 8.18224 | 13.08299 | 12.2172 | 11.56851 |
| ENSMUSG0 | 619.7681 | 630.5675 | 644.0334 | 958.3889 | 957.9373 | 975.6039 | -0.61022 | 5.64E-11 | 4.81E-09 | Eif4ebp1 | 8 | 27260329 | 27276674 | + | 2196 | protein_co | eukaryotic | - | 622 | 626 | 691 | 920 | 945 | 1006 | 14.89678 | 15.15636 | 15.48002 | 23.03589 | 23.02503 | 23.44967 |
| ENSMUSG0 | 5462.329 | 5624.743 | 4929.512 | 3939.812 | 4324.403 | 3814.165 | 0.407031 | 5.69E-11 | 4.83E-09 | Aspn | 13 | 49544443 | 49567565 | + | 2354 | protein_co | asporin [So | - | 5482 | 5584 | 5289 | 3782 | 4266 | 3933 | 122.4805 | 126.1223 | 110.5333 | 88.34145 | 96.96505 | 85.52411 |
| ENSMUSG0 | 774.2119 | 916.6397 | 906.8662 | 1314.66 | 1263.058 | 1356.729 | -0.59879 | 5.73E-11 | 4.83E-09 | Tet3 | 6 | 83362373 | 83459084 | - | 16695 | protein_co | tet methylc | - | 777 | 910 | 973 | 1262 | 1246 | 1399 | 2.447761 | 2.898063 | 2.867163 | 4.156448 | 3.993305 | 4.289458 |
| ENSMUSG0 | 6811.47 | 6494.04 | 6868.137 | 8087.969 | 8113.577 | 8736.795 | -0.30593 | 6.01E-11 | 5.03E-09 | Ahnak | 19 | 8989284 | 9076919 | + | 18543 | protein_co | AHNAK nuc | - | 6836 | 6447 | 7369 | 7764 | 8004 | 9009 | 19.38904 | 18.48547 | 19.55035 | 23.02263 | 23.09553 | 24.86954 |
| ENSMUSG0 | 1294.339 | 1122.128 | 1322.552 | 1666.763 | 1939.19 | 2030.73 | -0.59216 | 6.22E-11 | 5.18E-09 | Efna1 | 3 | 89271733 | 89281142 | - | 1830 | protein_co | ephrin A1 [ | - | 1299 | 1114 | 1419 | 1600 | 1913 | 2094 | 37.33296 | 32.36584 | 38.14672 | 48.0749 | 55.93256 | 58.57289 |
| ENSMUSG0 | 2894.576 | 2843.598 | 2920.985 | 3659.587 | 3583.395 | 3737.552 | -0.34264 | 6.61E-11 | 5.46E-09 | Rtn4 | 11 | 29692947 | 29744331 | + | 6991 | protein_co | reticulon 4 | - | 2905 | 2823 | 3134 | 3513 | 3535 | 3854 | 21.85451 | 21.46962 | 22.05391 | 27.63047 | 27.05521 | 28.21912 |
| ENSMUSG0 | 8197.479 | 8589.216 | 8774.14 | 10273.51 | 10271.72 | 10187.59 | -0.26575 | 7.35E-11 | 6.04E-09 | Atp1a1 | 3 | 1.02E+08 | 1.02E+08 | - | 4568 | protein_co | ATPase, Na- | - | 8227 | 8527 | 9414 | 9862 | 10133 | 10505 | 94.72176 | 99.24827 | 101.3851 | 118.7103 | 118.6896 | 117.7175 |
| ENSMUSG0 | 1389.994 | 1231.923 | 1344.921 | 1881.359 | 1873.3 | 1796.042 | -0.4844 | 7.55E-11 | 6.16E-09 | Larp1 | 11 | 58009064 | 58062034 | + | 6829 | protein_co | La ribonucl | - | 1395 | 1223 | 1443 | 1806 | 1848 | 1852 | 10.74364 | 9.521868 | 10.39525 | 14.54153 | 14.47924 | 13.88209 |
| ENSMUSG0 | 234.1567 | 260.8898 | 217.1632 | 89.58853 | 103.3964 | 81.46196 | 1.375943 | 8.62E-11 | 7.00E-09 | Serping1 | 2 | 84765387 | 84775444 | - | 2325 | protein_co | serine (or c | - | 235 | 259 | 233 | 86 | 102 | 84 | 5.31593 | 5.922835 | 4.930136 | 2.033878 | 2.347351 | 1.849385 |
| ENSMUSG0 | 1907.132 | 2033.731 | 2049.536 | 1480.294 | 1301.578 | 1479.892 | 0.491238 | 8.91E-11 | 7.18E-09 | Lgals1 | 15 | 78926725 | 78930465 | + | 903 | protein_co | lectin, gala | - | 1914 | 2019 | 2199 | 1421 | 1284 | 1526 | 111.4778 | 118.878 | 119.8018 | 86.52783 | 76.08133 | 86.50433 |
| ENSMUSG0 | 15183.32 | 15967.66 | 15946.12 | 13284.1 | 13266.16 | 13377.22 | 0.238272 | 9.10E-11 | 7.29E-09 | Smoc2 | 17 | 14279506 | 14404790 | + | 6951 | protein_co | SPARC relat | - | 15238 | 15852 | 17109 | 12752 | 13087 | 13794 | 115.2962 | 121.2522 | 121.0886 | 100.8743 | 100.7381 | 101.5814 |
| ENSMUSG0 | 678.5564 | 660.7864 | 582.5194 | 351.062 | 305.1208 | 398.5817 | 0.864606 | 1.08E-10 | 8.63E-09 | Akap12 | 10 | 4266380 | 4359470 | + | 6461 | protein_co | A kinase (PF | - | 681 | 656 | 625 | 337 | 301 | 411 | 5.54347 | 5.398299 | 4.758896 | 2.868003 | 2.492686 | 3.256216 |
| ENSMUSG0 | 8247.299 | 8371.64 | 7950.225 | 9869.322 | 11166.81 | 9865.625 | -0.33089 | 1.18E-10 | 9.38E-09 | Plagl1 | 10 | 13060504 | 13131694 | + | 7819 | protein_co | pleiomorpl | zf-C2H2 | 8277 | 8311 | 8530 | 9474 | 11016 | 10173 | 55.67447 | 56.51385 | 53.66903 | 66.62414 | 75.38302 | 66.59918 |
| ENSMUSG0 | 361.6974 | 420.0426 | 424.0741 | 217.721 | 177.3958 | 196.8664 | 1.026962 | 1.29E-10 | 1.01E-08 | Gm37352 | 8 | 84200869 | 84203049 | + | 2181 | TEC | predicted g | - | 363 | 417 | 455 | 209 | 175 | 203 | 8.753572 | 10.1656 | 10.26317 | 5.269144 | 4.29322 | 4.764436 |
| ENSMUSG0 | 860.8997 | 928.7273 | 941.3513 | 1353.203 | 1294.482 | 1300.482 | -0.53146 | 1.57E-10 | 1.23E-08 | Mafg | 11 | 1.21E+08 | 1.21E+08 | - | 5613 | protein_co | v-maf musc | TF_bZIP | 864 | 922 | 1010 | 1299 | 1277 | 1341 | 8.095674 | 8.733507 | 8.852221 | 12.72517 | 12.17297 | 12.22939 |

| gene_id | H1 | H2 | H3 | M1 | M2 | M3 | log2FoldCh | pvalue | padj | gene_name | gene_chr | gene_start | gene_end | gene_stran | gene_lengt | gene_bioty | gene_descr | tf_family | H1_readco | H2_readco | H3_readco | M1_readco | M2_readco | M3_readco | H1_fpkm | H2_fpkm | H3_fpkm | M1_fpkm | M2_fpkm | M3_fpkm |
|---|---|---|---|---|---|---|---|---|---|---|---|---|---|---|---|---|---|---|---|---|---|---|---|---|---|---|---|---|---|---|
| ENSMUSG0 | 886.8064 | 961.968 | 932.031 | 576.0751 | 640.6523 | 559.5661 | 0.646805 | 1.81E-10 | 1.41E-08 | Chrdl1 | X | 1.43E+08 | 1.43E+08 | - | 6758 | protein_co | chordin-lik | - | 890 | 955 | 1000 | 553 | 632 | 577 | 6.926377 | 7.513425 | 7.279603 | 4.499419 | 5.003797 | 4.370476 |
| ENSMUSG0 | 1361.098 | 1332.653 | 1378.474 | 894.8435 | 1008.622 | 912.5679 | 0.532233 | 2.11E-10 | 1.63E-08 | Vcan | 13 | 89655312 | 89742509 | - | 14462 | protein_co | versican [Sc | - | 1366 | 1323 | 1479 | 859 | 995 | 941 | 4.967715 | 4.863896 | 5.031132 | 3.265986 | 3.681252 | 3.330676 |
| ENSMUSG0 | 4418.089 | 4459.301 | 4434.604 | 5320.1 | 5518.53 | 5407.522 | -0.2874 | 2.29E-10 | 1.76E-08 | Iqgap1 | 7 | 80711583 | 80825974 | - | 16538 | protein_co | IQ motif co | - | 4434 | 4427 | 4758 | 5107 | 5444 | 5576 | 14.10091 | 14.23244 | 14.15361 | 16.97979 | 17.61311 | 17.25881 |
| ENSMUSG0 | 1222.597 | 1286.317 | 1253.582 | 1629.261 | 1816.533 | 1808.649 | -0.4821 | 2.38E-10 | 1.82E-08 | Mast4 | 13 | 1.03E+08 | 1.03E+08 | - | 32326 | protein_co | microtubul | - | 1227 | 1277 | 1345 | 1564 | 1792 | 1865 | 1.996305 | 2.10035 | 2.046898 | 2.660322 | 2.966107 | 2.953235 |
| ENSMUSG0 | 814.0683 | 788.7131 | 766.1295 | 1279.241 | 1200.209 | 1085.19 | -0.58931 | 2.45E-10 | 1.86E-08 | Smoc1 | 12 | 81026808 | 81186414 | + | 3496 | protein_co | SPARC relat | - | 817 | 783 | 822 | 1228 | 1184 | 1119 | 12.29094 | 11.90812 | 11.56715 | 19.31419 | 18.12096 | 16.38437 |
| ENSMUSG0 | 769.2298 | 686.9761 | 788.4983 | 405.2318 | 414.5993 | 497.4998 | 0.768395 | 2.53E-10 | 1.91E-08 | Lama2 | 10 | 26980036 | 27619758 | - | 16338 | protein_co | laminin, al | - | 772 | 682 | 846 | 389 | 409 | 513 | 2.485151 | 2.219414 | 2.547401 | 1.309182 | 1.339446 | 1.607273 |
| ENSMUSG0 | 319.8482 | 363.634 | 360.696 | 177.0936 | 157.122 | 171.652 | 1.045758 | 2.54E-10 | 1.91E-08 | Igfbp6 | 15 | 1.02E+08 | 1.02E+08 | + | 1112 | protein_co | insulin-like | - | 321 | 361 | 387 | 170 | 155 | 177 | 15.1822 | 17.26057 | 17.12112 | 8.406082 | 7.458092 | 8.147785 |
| ENSMUSG0 | 688.5205 | 678.9178 | 750.285 | 1018.809 | 1241.771 | 1054.157 | -0.646 | 3.13E-10 | 2.33E-08 | Noct | 3 | 51224447 | 51251644 | + | 10658 | protein_co | nocturnin [ | - | 691 | 674 | 805 | 978 | 1225 | 1087 | 3.409861 | 3.362304 | 3.715746 | 5.045597 | 6.149803 | 5.220654 |
| ENSMUSG0 | 2747.107 | 2620.985 | 2578.93 | 3498.119 | 3237.727 | 3571.719 | -0.3754 | 3.23E-10 | 2.40E-08 | Cdkn1a | 17 | 29090976 | 29100727 | + | 3553 | protein_co | cyclin-depe | - | 2757 | 2602 | 2767 | 3358 | 3194 | 3683 | 40.81087 | 38.93721 | 38.31244 | 51.96787 | 48.09949 | 53.06126 |
| ENSMUSG0 | 1272.418 | 1181.559 | 1468.881 | 1875.109 | 2088.202 | 1816.408 | -0.55845 | 3.58E-10 | 2.64E-08 | Zcchc14 | 8 | 1.22E+08 | 1.22E+08 | - | 7780 | protein_co | zinc finger, | - | 1277 | 1173 | 1576 | 1800 | 2060 | 1873 | 8.63268 | 8.016249 | 9.965579 | 12.72162 | 14.16734 | 12.32336 |
| ENSMUSG0 | 705.4595 | 694.0272 | 718.5959 | 426.0664 | 340.5999 | 456.7688 | 0.791509 | 4.31E-10 | 3.17E-08 | Pecam1 | 11 | 1.07E+08 | 1.07E+08 | - | 6228 | protein_co | platelet/en | - | 708 | 689 | 771 | 409 | 336 | 471 | 5.978868 | 5.881978 | 6.090202 | 3.610972 | 2.886632 | 3.87118 |
| ENSMUSG0 | 3668.788 | 3420.778 | 3793.366 | 4823.196 | 4460.237 | 4583.205 | -0.34927 | 4.75E-10 | 3.46E-08 | Nfe2l1 | 11 | 96817414 | 96829968 | - | 5441 | protein_co | nuclear fac | TF_bZIP | 3682 | 3396 | 4070 | 4630 | 4400 | 4726 | 35.59093 | 33.18499 | 36.79947 | 46.78986 | 43.26879 | 44.4617 |
| ENSMUSG0 | 6759.657 | 7145.76 | 6815.011 | 5557.614 | 5841.897 | 5642.21 | 0.281907 | 5.32E-10 | 3.86E-08 | Mmp14 | 14 | 54431612 | 54445364 | + | 5960 | protein_co | matrix met | - | 6784 | 7094 | 7312 | 5335 | 5763 | 5818 | 59.86513 | 63.28455 | 60.35536 | 49.21955 | 51.73723 | 49.96876 |
| ENSMUSG0 | 2256.872 | 2168.709 | 2068.177 | 2742.867 | 2933.62 | 2804.619 | -0.38546 | 5.49E-10 | 3.96E-08 | Spred1 | 2 | 1.17E+08 | 1.17E+08 | + | 6784 | protein_co | sprouty prc | - | 2265 | 2153 | 2219 | 2633 | 2894 | 2892 | 17.55968 | 16.87373 | 16.09153 | 21.34099 | 22.82514 | 21.82145 |
| ENSMUSG0 | 3536.265 | 3617.201 | 3677.794 | 4745.067 | 4737.988 | 4271.904 | -0.34449 | 6.23E-10 | 4.47E-08 | Srebf2 | 15 | 82147181 | 82205379 | + | 7184 | protein_co | sterol regul | bHLH | 3549 | 3591 | 3946 | 4555 | 4674 | 4405 | 25.98209 | 26.57675 | 27.02195 | 34.86354 | 34.81154 | 31.38706 |
| ENSMUSG0 | 1025.308 | 1118.099 | 1265.698 | 1624.052 | 1595.549 | 1765.009 | -0.54772 | 7.00E-10 | 5.00E-08 | Ier3 | 17 | 35821684 | 35822923 | + | 1131 | protein_co | immediate | - | 1029 | 1110 | 1358 | 1559 | 1574 | 1820 | 47.85057 | 52.18109 | 59.06947 | 75.79368 | 74.46341 | 82.37205 |
| ENSMUSG0 | 8596.044 | 8795.712 | 9502.056 | 10747.5 | 10729.91 | 11295.09 | -0.28513 | 7.83E-10 | 5.55E-08 | Cst3 | 2 | 1.49E+08 | 1.49E+08 | - | 1266 | protein_co | cystatin C [ | - | 8627 | 8732 | 10195 | 10317 | 10585 | 11647 | 358.3938 | 366.7185 | 396.168 | 448.0941 | 447.3608 | 470.9247 |
| ENSMUSG0 | 3081.901 | 2833.525 | 3033.761 | 2348.053 | 2309.186 | 2288.693 | 0.365766 | 7.86E-10 | 5.55E-08 | Igfbp4 | 11 | 99041244 | 99054392 | + | 4416 | protein_co | insulin-like | - | 3093 | 2813 | 3255 | 2254 | 2278 | 2360 | 36.83708 | 33.86831 | 36.26167 | 28.0656 | 27.60104 | 27.35609 |
| ENSMUSG0 | 73.73446 | 99.72234 | 86.67889 | 6.250362 | 20.27381 | 13.57699 | 2.694228 | 7.95E-10 | 5.59E-08 | Ablim3 | 18 | 61799395 | 61911852 | - | 4560 | protein_co | actin bindi | - | 74 | 99 | 93 | 6 | 20 | 14 | 0.853495 | 1.154312 | 1.00333 | 0.07235 | 0.234675 | 0.157157 |
| ENSMUSG0 | 280.9881 | 375.7215 | 349.5116 | 135.4245 | 179.4232 | 117.344 | 1.219989 | 1.02E-09 | 7.14E-08 | Clec3b | 9 | 1.23E+08 | 1.23E+08 | + | 992 | protein_co | C-type lecti | - | 282 | 373 | 375 | 130 | 177 | 121 | 14.95105 | 19.99171 | 18.59711 | 7.205782 | 9.546901 | 6.24374 |
| ENSMUSG0 | 1652.051 | 1741.615 | 1628.258 | 2123.04 | 2372.035 | 2234.385 | -0.42251 | 1.11E-09 | 7.73E-08 | Crispld2 | 8 | 1.2E+08 | 1.2E+08 | + | 4777 | protein_co | cysteine-ric | - | 1658 | 1729 | 1747 | 2038 | 2340 | 2304 | 18.25423 | 19.24388 | 17.99134 | 23.4584 | 26.20966 | 24.68871 |
| ENSMUSG0 | 2316.657 | 2701.569 | 2307.709 | 1826.147 | 1813.492 | 1614.692 | 0.479573 | 1.26E-09 | 8.71E-08 | Tuba1a | 15 | 98949837 | 98953703 | - | 1803 | protein_co | tubulin, alp | - | 2325 | 2682 | 2476 | 1753 | 1789 | 1665 | 67.82059 | 79.08896 | 67.55863 | 53.46082 | 53.09033 | 47.27043 |
| ENSMUSG0 | 1542.445 | 1448.492 | 1491.25 | 2007.408 | 2270.666 | 1925.024 | -0.46865 | 1.31E-09 | 8.99E-08 | Ncoa3 | 2 | 1.66E+08 | 1.66E+08 | + | 7895 | protein_co | nuclear rec | - | 1548 | 1438 | 1600 | 1927 | 2240 | 1985 | 10.31224 | 9.684106 | 9.969968 | 13.42082 | 15.18087 | 12.87003 |
| ENSMUSG0 | 979.4727 | 991.1796 | 1084.884 | 1440.708 | 1381.66 | 1482.802 | -0.49431 | 1.52E-09 | 1.04E-07 | Slc35e4 | 11 | 3907021 | 3914664 | - | 3063 | protein_co | solute carri | - | 983 | 984 | 1164 | 1383 | 1363 | 1529 | 16.87877 | 17.08051 | 18.69528 | 24.82702 | 23.80947 | 25.55239 |
| ENSMUSG0 | 1310.281 | 1292.361 | 1376.61 | 1814.688 | 1917.902 | 1711.671 | -0.45183 | 1.58E-09 | 1.07E-07 | Uck2 | 1 | 1.67E+08 | 1.67E+08 | - | 5464 | protein_co | uridine-cyt | - | 1315 | 1283 | 1477 | 1742 | 1892 | 1765 | 12.65754 | 12.48443 | 13.29829 | 17.5302 | 18.52726 | 16.53503 |
| ENSMUSG0 | 2194.099 | 2037.761 | 2005.731 | 2573.066 | 2931.592 | 2786.193 | -0.41079 | 1.71E-09 | 1.16E-07 | Acsl3 | 1 | 78657825 | 78707743 | + | 4357 | protein_co | acyl-CoA sy | - | 2202 | 2023 | 2152 | 2470 | 2892 | 2873 | 26.58056 | 24.68659 | 24.29856 | 31.17158 | 35.51498 | 33.75353 |
| ENSMUSG0 | 3214.424 | 3231.407 | 3352.516 | 2639.736 | 2562.609 | 2599.994 | 0.328794 | 1.95E-09 | 1.32E-07 | Dnm1 | 2 | 32308471 | 32353338 | - | 5249 | protein_co | dynamin 1 | - | 3226 | 3208 | 3597 | 2534 | 2528 | 2681 | 32.32378 | 32.49455 | 33.71241 | 26.5448 | 25.76922 | 26.14516 |
| ENSMUSG0 | 1021.322 | 923.6908 | 1088.612 | 1490.711 | 1365.441 | 1499.288 | -0.52132 | 2.46E-09 | 1.65E-07 | Lrrfip1 | 1 | 90998737 | 91128944 | + | 6469 | protein_co | leucine rich | - | 1025 | 917 | 1168 | 1431 | 1347 | 1546 | 8.333377 | 7.536765 | 8.882426 | 12.16331 | 11.14118 | 12.23329 |
| ENSMUSG0 | 2743.121 | 2881.875 | 2753.22 | 3599.167 | 3449.588 | 3447.586 | -0.32518 | 2.71E-09 | 1.81E-07 | Eif2ak3 | 6 | 70844515 | 70905245 | + | 6026 | protein_co | eukaryotic | - | 2753 | 2861 | 2954 | 3455 | 3403 | 3555 | 24.02766 | 25.24303 | 24.11611 | 31.52596 | 30.21577 | 30.19823 |
| ENSMUSG0 | 5475.282 | 5988.377 | 5733.855 | 4589.849 | 4805.906 | 4597.752 | 0.297431 | 2.98E-09 | 1.98E-07 | Cyr61 | 3 | 1.46E+08 | 1.46E+08 | - | 2019 | protein_co | cysteine ric | - | 5495 | 5945 | 6152 | 4406 | 4741 | 4741 | 143.1415 | 156.5555 | 149.9015 | 119.9935 | 125.6419 | 120.2001 |
| ENSMUSG0 | 6376.038 | 6307.69 | 6336.879 | 7690.029 | 7912.866 | 7280.178 | -0.26662 | 3.24E-09 | 2.14E-07 | Hmgcs1 | 13 | 1.2E+08 | 1.2E+08 | + | 4594 | protein_co | 3-hydroxy- | - | 6399 | 6262 | 6799 | 7382 | 7806 | 7507 | 73.25807 | 72.47277 | 72.80814 | 88.35528 | 90.91559 | 83.64626 |
| ENSMUSG0 | 1510.56 | 1546.2 | 1733.578 | 1199.028 | 929.554 | 1092.948 | 0.572808 | 3.54E-09 | 2.32E-07 | F13a1 | 13 | 36867178 | 37050244 | - | 6739 | protein_co | coagulatior | - | 1516 | 1535 | 1860 | 1151 | 917 | 1127 | 11.83145 | 12.1106 | 13.57824 | 9.391378 | 7.280726 | 8.560508 |
| ENSMUSG0 | 1759.663 | 1779.893 | 1804.412 | 2476.185 | 2239.242 | 2290.633 | -0.39045 | 3.66E-09 | 2.39E-07 | Golga4 | 9 | 1.19E+08 | 1.19E+08 | + | 13602 | protein_co | golgi autoa | - | 1766 | 1767 | 1936 | 2377 | 2209 | 2362 | 6.828452 | 6.906954 | 7.002103 | 9.608949 | 8.689479 | 8.888904 |
| ENSMUSG0 | 961.5373 | 938.8002 | 867.7209 | 606.2851 | 634.5701 | 613.874 | 0.577323 | 3.69E-09 | 2.40E-07 | Adamts2 | 11 | 50602084 | 50807573 | + | 9336 | protein_co | a disintegri | - | 965 | 932 | 931 | 582 | 626 | 633 | 5.436267 | 5.307718 | 4.905855 | 3.427769 | 3.587685 | 3.470675 |
| ENSMUSG0 | 11905.13 | 11548.65 | 12896.51 | 15196.71 | 14101.45 | 14725.22 | -0.27617 | 3.91E-09 | 2.53E-07 | Lmna | 3 | 88480147 | 88509956 | - | 5838 | protein_co | lamin A [So | - | 11948 | 11465 | 13837 | 14588 | 13911 | 15184 | 107.638 | 104.415 | 116.6014 | 137.3982 | 127.4956 | 133.1353 |
| ENSMUSG0 | 750.298 | 644.6697 | 812.7311 | 450.0261 | 425.7499 | 471.3156 | 0.713011 | 3.92E-09 | 2.53E-07 | Fbln2 | 6 | 91212455 | 91272540 | + | 6370 | protein_co | fibulin 2 [Sc | - | 753 | 640 | 872 | 432 | 420 | 486 | 6.217129 | 5.34187 | 6.734463 | 3.729012 | 3.527855 | 3.905422 |
| ENSMUSG0 | 5094.653 | 5354.788 | 5731.059 | 4437.757 | 4252.431 | 4315.544 | 0.315334 | 3.98E-09 | 2.55E-07 | Tubb4b | 2 | 25222160 | 25224702 | - | 2088 | protein_co | tubulin, be | - | 5113 | 5316 | 6149 | 4260 | 4195 | 4450 | 128.7893 | 135.3653 | 144.8772 | 112.1834 | 107.4985 | 109.0939 |
| ENSMUSG0 | 776.2047 | 877.3551 | 930.167 | 1225.071 | 1272.181 | 1250.053 | -0.53599 | 4.02E-09 | 2.56E-07 | Gtf3c4 | 2 | 28822299 | 28840360 | - | 7230 | protein_co | general trar | - | 779 | 871 | 998 | 1176 | 1255 | 1289 | 5.666743 | 6.405199 | 6.790756 | 8.943726 | 9.287659 | 9.12611 |
| ENSMUSG0 | 615.7824 | 604.3778 | 667.3342 | 910.4694 | 916.376 | 997.909 | -0.58146 | 4.14E-09 | 2.63E-07 | Ddit3 | 10 | 1.27E+08 | 1.27E+08 | + | 1713 | protein_co | DNA-damag | TF_bZIP | 618 | 600 | 716 | 874 | 904 | 1029 | 18.97429 | 18.62287 | 20.56277 | 28.05456 | 28.23657 | 30.74887 |
| ENSMUSG0 | 504.1843 | 655.7499 | 564.8108 | 955.2637 | 826.1576 | 1046.398 | -0.71373 | 4.49E-09 | 2.84E-07 | Hmga1 | 17 | 27556497 | 27563674 | + | 2125 | protein_co | high mobili | HMGI/HMG | 506 | 651 | 606 | 917 | 815 | 1079 | 12.52351 | 16.28827 | 14.02942 | 23.72793 | 20.52105 | 25.99163 |
| ENSMUSG0 | 2442.205 | 2455.789 | 2582.658 | 3465.826 | 2985.318 | 3311.816 | -0.38395 | 4.53E-09 | 2.85E-07 | Phldb1 | 9 | 44686304 | 44735198 | - | 7179 | protein_co | pleckstrin h | - | 2451 | 2438 | 2771 | 3327 | 2945 | 3415 | 17.95617 | 18.05604 | 18.98884 | 25.48228 | 21.94937 | 24.34994 |
| ENSMUSG0 | 1344.159 | 1284.303 | 1292.727 | 1752.185 | 1905.738 | 1678.698 | -0.44454 | 4.91E-09 | 3.07E-07 | Aff1 | 5 | 1.04E+08 | 1.04E+08 | + | 9458 | protein_co | AF4/FMR2 | AF-4 | 1349 | 1275 | 1387 | 1682 | 1880 | 1731 | 7.50148 | 7.167433 | 7.214447 | 9.778588 | 10.63554 | 9.368474 |
| ENSMUSG0 | 5893.775 | 5596.539 | 5441.197 | 6763.934 | 7612.814 | 6774.919 | -0.32113 | 5.12E-09 | 3.19E-07 | Ankrd28 | 14 | 31698768 | 31830651 | - | 12233 | protein_co | ankyrin rep | - | 5915 | 5556 | 5838 | 6493 | 7510 | 6986 | 25.43057 | 24.14805 | 23.47778 | 29.18515 | 32.84791 | 29.23255 |
| ENSMUSG0 | 1632.122 | 1772.842 | 1600.297 | 2215.753 | 2126.722 | 2279.965 | -0.40428 | 5.15E-09 | 3.19E-07 | Ank | 15 | 27466677 | 27594909 | + | 3610 | protein_co | progressive | - | 1638 | 1760 | 1717 | 2127 | 2098 | 2351 | 23.86388 | 25.92139 | 23.39855 | 32.39737 | 31.09561 | 33.33623 |
| ENSMUSG0 | 2618.57 | 2775.102 | 2771.86 | 3452.283 | 3388.767 | 3355.457 | -0.32031 | 5.69E-09 | 3.51E-07 | Sqle | 15 | 59315077 | 59331192 | + | 5546 | protein_co | squalene ep | - | 2628 | 2755 | 2974 | 3314 | 3343 | 3460 | 24.92182 | 26.41159 | 26.38074 | 32.85656 | 32.25205 | 31.93503 |
| ENSMUSG0 | 1675.964 | 1627.791 | 1655.287 | 2420.974 | 2068.942 | 2170.379 | -0.4253 | 6.05E-09 | 3.72E-07 | Mafk | 5 | 1.4E+08 | 1.4E+08 | + | 3901 | protein_co | v-maf musc | TF_bZIP | 1682 | 1616 | 1776 | 2324 | 2041 | 2238 | 22.67694 | 22.02512 | 22.39716 | 32.75742 | 27.99419 | 29.3667 |
| ENSMUSG0 | 12053.59 | 11997.91 | 11580.49 | 13887.26 | 15101.96 | 13636.15 | -0.25856 | 6.30E-09 | 3.85E-07 | Scd2 | 19 | 44293676 | 44306864 | + | 5453 | protein_co | stearoyl-Co | - | 12097 | 11911 | 12425 | 13331 | 14898 | 14061 | 116.6747 | 116.1356 | 112.0951 | 134.424 | 146.1818 | 131.9933 |
| ENSMUSG0 | 1360.102 | 1352.799 | 1367.29 | 990.6824 | 1024.841 | 946.5104 | 0.462279 | 6.61E-09 | 4.02E-07 | Ctsk | 3 | 95499256 | 95509362 | + | 1512 | protein_co | cathepsin K | - | 1365 | 1343 | 1467 | 951 | 1011 | 976 | 47.48049 | 47.22555 | 47.73141 | 34.58424 | 35.77669 | 33.04221 |
| ENSMUSG0 | 566.9582 | 572.1443 | 603.0241 | 888.5932 | 815.007 | 916.447 | -0.58859 | 6.92E-09 | 4.19E-07 | Fxyd5 | 7 | 31032722 | 31042481 | - | 4192 | protein_co | FXYD doma | - | 569 | 568 | 647 | 853 | 804 | 945 | 7.138802 | 7.204102 | 7.592921 | 11.18864 | 10.26208 | 11.53936 |
| ENSMUSG0 | 752.2908 | 853.18 | 876.1092 | 1279.241 | 1201.223 | 1137.558 | -0.54336 | 7.00E-09 | 4.22E-07 | Cep170b | 12 | 1.13E+08 | 1.13E+08 | + | 8002 | protein_co | centrosom | - | 755 | 847 | 940 | 1228 | 1185 | 1173 | 4.962297 | 5.627787 | 5.779033 | 8.43819 | 7.923565 | 7.503615 |
| ENSMUSG0 | 3236.345 | 3318.034 | 3387.933 | 4109.613 | 4006.104 | 4059.521 | -0.29216 | 7.19E-09 | 4.31E-07 | Rpn1 | 6 | 88084482 | 88105304 | + | 4512 | protein_co | ribophorin | - | 3248 | 3294 | 3635 | 3945 | 3952 | 4186 | 37.86006 | 38.81569 | 39.63339 | 48.0759 | 46.865 | 47.4899 |
| ENSMUSG0 | 2472.097 | 2418.519 | 2392.524 | 1959.489 | 1814.506 | 1885.262 | 0.363981 | 7.35E-09 | 4.39E-07 | Tpm1 | 9 | 67022590 | 67049406 | - | 6485 | protein_co | tropomyos | - | 2481 | 2401 | 2567 | 1881 | 1790 | 1944 | 20.12107 | 19.68498 | 19.4734 | 15.94881 | 14.76875 | 15.34466 |
| ENSMUSG0 | 753.2872 | 708.1294 | 589.9756 | 368.7714 | 445.01 | 400.5213 | 0.755738 | 7.64E-09 | 4.54E-07 | Dpt | 1 | 1.65E+08 | 1.65E+08 | + | 1724 | protein_co | dermatopo | - | 756 | 703 | 633 | 354 | 439 | 413 | 23.06316 | 21.68058 | 18.0631 | 11.29056 | 13.62474 | 12.26264 |
| ENSMUSG0 | 1379.034 | 1385.033 | 1338.397 | 1745.935 | 1873.3 | 1814.468 | -0.40567 | 8.79E-09 | 5.20E-07 | Smad5 | 13 | 56703010 | 56742377 | + | 7420 | protein_co | SMAD famil | MH1 | 1384 | 1375 | 1436 | 1676 | 1848 | 1871 | 9.809944 | 9.852617 | 9.520866 | 12.41994 | 13.32597 | 12.90747 |
| ENSMUSG0 | 983.4583 | 1015.355 | 841.624 | 580.242 | 648.7618 | 628.4208 | 0.612078 | 9.75E-09 | 5.75E-07 | Cdh11 | 8 | 1.03E+08 | 1.03E+08 | - | 8904 | protein_co | cadherin 1 | - | 987 | 1008 | 903 | 557 | 640 | 648 | 5.82997 | 6.019053 | 4.989172 | 3.439692 | 3.845879 | 3.725297 |
| ENSMUSG0 | 490.2345 | 607.3997 | 433.3944 | 247.931 | 272.6827 | 300.6334 | 0.897293 | 1.02E-08 | 5.99E-07 | Cd93 | 2 | 1.48E+08 | 1.48E+08 | - | 6677 | protein_co | CD93 antig | - | 492 | 603 | 465 | 238 | 269 | 310 | 3.875414 | 4.80163 | 3.42608 | 1.95995 | 2.155618 | 2.376574 |
| ENSMUSG0 | 1144.877 | 1072.771 | 1152.922 | 750.0435 | 826.1576 | 785.526 | 0.513193 | 1.04E-08 | 6.06E-07 | Col18a1 | 10 | 77052178 | 77166548 | - | 7021 | protein_co | collagen, ty | - | 1149 | 1065 | 1237 | 720 | 815 | 810 | 8.607071 | 8.064983 | 8.667555 | 5.638752 | 6.21097 | 5.905506 |
| ENSMUSG0 | 472.2991 | 584.2319 | 525.6655 | 306.2677 | 259.5047 | 326.8176 | 0.825408 | 1.05E-08 | 6.09E-07 | Tpm2 | 4 | 43514711 | 43523765 | - | 2909 | protein_co | tropomyos | - | 474 | 580 | 564 | 294 | 256 | 337 | 8.569766 | 10.60076 | 9.538087 | 5.557162 | 4.708657 | 5.930035 |
| ENSMUSG0 | 1662.015 | 1447.485 | 1586.317 | 2045.952 | 2210.859 | 2044.307 | -0.42408 | 1.07E-08 | 6.19E-07 | Abl2 | 1 | 1.57E+08 | 1.57E+08 | + | 10800 | protein_co | v-abl Abels | - | 1668 | 1437 | 1702 | 1964 | 2181 | 2108 | 8.122816 | 7.074338 | 7.752856 | 9.999245 | 10.8052 | 9.991206 |
| ENSMUSG0 | 8419.679 | 8485.465 | 8574.685 | 6938.944 | 7400.953 | 7226.839 | 0.240513 | 1.08E-08 | 6.23E-07 | Fos | 12 | 85473890 | 85477273 | + | 2352 | protein_co | FBJ osteosa | TF_bZIP | 8450 | 8424 | 9200 | 6661 | 7301 | 7452 | 188.953 | 190.4293 | 192.4316 | 155.7226 | 166.0909 | 162.1835 |
| ENSMUSG0 | 509.1664 | 469.4001 | 423.1421 | 668.7888 | 811.9659 | 782.6167 | -0.69217 | 1.10E-08 | 6.32E-07 | Fgl2 | 5 | 21372642 | 21378374 | + | 3788 | protein_co | fibrinogen- | - | 511 | 466 | 454 | 642 | 801 | 807 | 7.094883 | 6.540768 | 5.896194 | 9.319112 | 11.31419 | 10.90523 |
| ENSMUSG0 | 3901.948 | 3693.756 | 3715.076 | 4613.809 | 4604.181 | 4527.927 | -0.28131 | 1.11E-08 | 6.36E-07 | Fgfr2 | 7 | 1.3E+08 | 1.33E+08 | - | 10328 | protein_co | fibroblast g | - | 3916 | 3667 | 3986 | 4429 | 4542 | 4669 | 19.94164 | 18.87763 | 18.98659 | 23.57973 | 23.53053 | 23.14082 |
| ENSMUSG0 | 743.3231 | 760.5088 | 740.9647 | 1033.393 | 1117.087 | 1047.368 | -0.51061 | 1.18E-08 | 6.72E-07 | Epb41l1 | 2 | 1.56E+08 | 1.57E+08 | + | 11530 | protein_co | erythrocyte | - | 746 | 755 | 795 | 992 | 1102 | 1080 | 3.402858 | 3.481532 | 3.392062 | 4.73077 | 5.11391 | 4.794745 |
| ENSMUSG0 | 644.6784 | 704.1002 | 702.7514 | 950.0551 | 994.4302 | 1049.308 | -0.54539 | 1.19E-08 | 6.72E-07 | Dixdc1 | 9 | 50662752 | 50739517 | - | 9971 | protein_co | DIX domair | - | 647 | 699 | 754 | 912 | 981 | 1082 | 3.412714 | 3.727273 | 3.720134 | 5.029277 | 5.264184 | 5.554687 |
| ENSMUSG0 | 421.4821 | 452.2761 | 385.8608 | 621.911 | 776.4868 | 671.0914 | -0.71697 | 1.21E-08 | 6.84E-07 | Cmtm4 | 8 | 1.04E+08 | 1.04E+08 | - | 7734 | protein_co | CKLF-like M | - | 423 | 449 | 414 | 597 | 766 | 692 | 2.876541 | 3.086704 | 2.633432 | 4.244432 | 5.299384 | 4.580079 |
| ENSMUSG0 | 339.7764 | 319.313 | 328.0749 | 179.1771 | 180.4369 | 135.7699 | 0.996001 | 1.52E-08 | 8.53E-07 | Mpz | 1 | 1.71E+08 | 1.71E+08 | + | 2111 | protein_co | myelin pro | - | 341 | 317 | 352 | 172 | 178 | 140 | 8.495726 | 7.984061 | 8.203144 | 4.48012 | 4.511621 | 3.394774 |
| ENSMUSG0 | 1993.82 | 1966.243 | 1905.071 | 2404.306 | 2513.952 | 2555.384 | -0.34995 | 1.65E-08 | 9.24E-07 | Gars | 6 | 55038007 | 55079500 | + | 3119 | protein_co | glycyl-tRNA | - | 2001 | 1952 | 2044 | 2308 | 2480 | 2635 | 33.74163 | 33.27494 | 32.23973 | 40.68833 | 42.54388 | 43.24504 |
| ENSMUSG0 | 16941.99 | 16507.57 | 15934.93 | 19335.5 | 19495.29 | 18543.26 | -0.21636 | 1.67E-08 | 9.29E-07 | Slc38a2 | 15 | 96687392 | 96699730 | - | 8838 | protein_co | solute carri | - | 17003 | 16388 | 17097 | 18561 | 19232 | 19121 | 101.1826 | 98.58816 | 95.16819 | 115.4774 | 116.4317 | 110.7459 |
| ENSMUSG0 | 3974.686 | 4189.346 | 3913.598 | 4873.199 | 5163.738 | 4782.011 | -0.29519 | 1.69E-08 | 9.37E-07 | Atp2a2 | 5 | 1.22E+08 | 1.23E+08 | - | 6081 | protein_co | ATPase, Ca | - | 3989 | 4159 | 4199 | 4678 | 5094 | 4931 | 34.50034 | 36.36359 | 33.9701 | 42.29945 | 44.82133 | 41.50793 |
| ENSMUSG0 | 4693.099 | 4472.396 | 4509.166 | 3381.446 | 3921.968 | 3518.381 | 0.337511 | 1.71E-08 | 9.44E-07 | Nrk | X | 1.39E+08 | 1.39E+08 | + | 9210 | protein_co | Nik related | - | 4710 | 4440 | 4838 | 3246 | 3869 | 3628 | 26.89649 | 25.63162 | 25.84236 | 19.37931 | 22.47708 | 20.16409 |
| ENSMUSG0 | 3348.94 | 3284.793 | 3352.516 | 4040.859 | 4247.362 | 3956.724 | -0.29406 | 1.74E-08 | 9.55E-07 | Chpf | 1 | 75474569 | 75479307 | - | 3856 | protein_co | chondroiti | - | 3361 | 3261 | 3597 | 3879 | 4190 | 4080 | 45.84224 | 44.96417 | 45.89119 | 55.31364 | 58.14037 | 54.16194 |
| ENSMUSG0 | 823.036 | 854.1873 | 822.9834 | 590.6592 | 537.2559 | 557.6265 | 0.568943 | 1.83E-08 | 1.00E-06 | Selenop | 15 | 3268547 | 3280508 | + | 2842 | protein_co | selenoprot | - | 826 | 848 | 883 | 567 | 530 | 575 | 15.28588 | 15.86443 | 15.2849 | 10.97005 | 9.978209 | 10.35654 |
| ENSMUSG0 | 333.7979 | 335.4297 | 294.5218 | 102.0892 | 152.0535 | 179.4103 | 1.150196 | 1.96E-08 | 1.07E-06 | Cavin2 | 1 | 51289126 | 51302960 | + | 3055 | protein_co | caveolae as | - | 335 | 333 | 316 | 98 | 150 | 185 | 5.767239 | 5.795432 | 5.08864 | 1.763861 | 2.627126 | 3.099785 |
| ENSMUSG0 | 232.1639 | 209.5176 | 181.7461 | 82.29644 | 58.79404 | 99.88788 | 1.369553 | 2.23E-08 | 1.21E-06 | Zfp648 | 1 | 1.54E+08 | 1.54E+08 | + | 1892 | protein_co | zinc finger | zf-C2H2 | 233 | 208 | 195 | 79 | 58 | 103 | 6.476929 | 5.845141 | 5.070367 | 2.295913 | 1.640241 | 2.78668 |
| ENSMUSG0 | 765.2442 | 761.5161 | 709.2756 | 1088.605 | 990.3754 | 1181.198 | -0.54457 | 2.32E-08 | 1.25E-06 | Adgrg1 | 8 | 94974751 | 95014217 | + | 5421 | protein_co | adhesion G | - | 768 | 756 | 761 | 1045 | 977 | 1218 | 7.451027 | 7.414727 | 6.906073 | 10.59952 | 9.643084 | 11.50109 |
| ENSMUSG0 | 1969.906 | 2081.074 | 1933.032 | 2448.059 | 2570.719 | 2633.937 | -0.35523 | 2.44E-08 | 1.31E-06 | Sars | 3 | 1.08E+08 | 1.08E+08 | - | 5142 | protein_co | seryl-aminc | - | 1977 | 2066 | 2074 | 2350 | 2536 | 2716 | 20.22129 | 21.36245 | 19.84278 | 25.12958 | 26.3887 | 27.03764 |
| ENSMUSG0 | 560.9798 | 592.2903 | 544.3061 | 876.0924 | 776.4868 | 902.87 | -0.59054 | 2.45E-08 | 1.32E-06 | Ngef | 1 | 87476834 | 87573870 | - | 3905 | protein_co | neuronal gu | - | 563 | 588 | 584 | 841 | 766 | 931 | 7.582662 | 8.00588 | 7.357287 | 11.84198 | 10.49563 | 12.20393 |
| ENSMUSG0 | 1394.976 | 1513.966 | 1423.211 | 2200.128 | 1897.628 | 1833.864 | -0.45311 | 2.73E-08 | 1.46E-06 | Itpr3 | 17 | 27057304 | 27122223 | + | 8990 | protein_co | inositol 1,4 | - | 1400 | 1503 | 1527 | 2112 | 1872 | 1891 | 8.190354 | 8.888983 | 8.356131 | 12.91766 | 11.14158 | 10.7672 |
| ENSMUSG0 | 3492.423 | 3341.202 | 3880.977 | 4634.644 | 4459.224 | 4414.462 | -0.33388 | 2.79E-08 | 1.49E-06 | Sdk2 | 11 | 1.14E+08 | 1.14E+08 | - | 15469 | protein_co | sidekick cel | - | 3505 | 3317 | 4164 | 4449 | 4399 | 4552 | 11.91681 | 11.40082 | 13.24263 | 15.81429 | 15.21572 | 15.06299 |
| ENSMUSG0 | 1003.387 | 1103.997 | 989.817 | 1462.585 | 1392.81 | 1384.853 | -0.45337 | 3.06E-08 | 1.62E-06 | Rxra | 2 | 27676440 | 27762957 | + | 5462 | protein_co | retinoid X r | RXR-like | 1007 | 1096 | 1062 | 1404 | 1374 | 1428 | 9.696435 | 10.6687 | 9.565303 | 14.13399 | 13.45971 | 13.38282 |
| ENSMUSG0 | 1309.285 | 1049.603 | 1206.98 | 1688.64 | 1783.081 | 1570.082 | -0.49938 | 3.18E-08 | 1.68E-06 | Sertad2 | 11 | 20543253 | 20653021 | + | 6352 | protein_co | SERTA dom | - | 1314 | 1042 | 1295 | 1621 | 1759 | 1619 | 10.87976 | 8.721878 | 10.02964 | 14.03208 | 14.81686 | 13.0469 |
| ENSMUSG0 | 2321.639 | 2454.781 | 2291.864 | 3077.262 | 3131.289 | 2808.498 | -0.35125 | 3.23E-08 | 1.70E-06 | Nars | 18 | 64499665 | 64516557 | - | 2632 | protein_co | asparaginyl | - | 2330 | 2437 | 2459 | 2954 | 3089 | 2896 | 46.55908 | 49.22916 | 45.96196 | 61.71263 | 62.79612 | 56.32274 |
| ENSMUSG0 | 3722.594 | 3659.508 | 3698.299 | 4485.677 | 4704.537 | 4315.544 | -0.28546 | 3.34E-08 | 1.75E-06 | Sfr1 | 19 | 47731756 | 47735588 | + | 1583 | protein_co | SWI5 depe | - | 3736 | 3633 | 3968 | 4306 | 4641 | 4450 | 124.1253 | 122.0218 | 123.3152 | 149.5693 | 156.867 | 143.8965 |
| ENSMUSG0 | 1401.951 | 1498.857 | 1586.317 | 1102.147 | 972.129 | 1138.528 | 0.482034 | 3.67E-08 | 1.92E-06 | Socs3 | 11 | 1.18E+08 | 1.18E+08 | - | 2891 | protein_co | suppressor | - | 1407 | 1488 | 1702 | 1058 | 959 | 1174 | 25.59649 | 27.36577 | 28.96259 | 20.12274 | 17.7489 | 20.78697 |

| gene_id | H1 | H2 | H3 | M1 | M2 | M3 | log2FoldCh | pvalue | padj | gene_name | gene_chr | gene_start | gene_end | gene_stran | gene_lengt | gene_bioty | gene_descr | tf_family | H1_readcou | H2_readcou | H3_readcou | M1_readco | M2_readco | M3_readco | H1_fpkm | H2_fpkm | H3_fpkm | M1_fpkm | M2_fpkm | M3_fpkm |
|---|---|---|---|---|---|---|---|---|---|---|---|---|---|---|---|---|---|---|---|---|---|---|---|---|---|---|---|---|---|---|
| ENSMUSG0 | 607.8111 | 645.677 | 573.1991 | 879.2176 | 881.9105 | 900.9305 | -0.54387 | 3.82E-08 | 1.98E-06 | Zswim4 | 8 | 84210678 | 84237055 | - | 4625 | protein_co | zinc finger | - | 610 | 641 | 615 | 844 | 870 | 929 | 6.936692 | 7.368839 | 6.54168 | 10.03414 | 10.06487 | 10.28194 |
| ENSMUSG0 | 1262.454 | 1281.281 | 1288.067 | 1699.057 | 1718.205 | 1620.511 | -0.39456 | 3.95E-08 | 2.05E-06 | Hyou1 | 9 | 44379490 | 44392369 | + | 7230 | protein_co | hypoxia up | - | 1267 | 1272 | 1382 | 1631 | 1695 | 1671 | 9.216641 | 9.354091 | 9.403632 | 12.4041 | 12.54389 | 11.83067 |
| ENSMUSG0 | 1176.762 | 1141.267 | 1326.28 | 908.386 | 827.1713 | 819.4685 | 0.513042 | 3.99E-08 | 2.05E-06 | Megf6 | 4 | 1.54E+08 | 1.54E+08 | + | 7877 | protein_co | multiple EC | - | 1181 | 1133 | 1423 | 872 | 816 | 845 | 7.885394 | 7.647542 | 8.887302 | 6.087025 | 5.542812 | 5.491196 |
| ENSMUSG0 | 1813.469 | 1909.834 | 1710.277 | 1281.324 | 1438.427 | 1325.696 | 0.425288 | 3.99E-08 | 2.05E-06 | Olfml2b | 1 | 1.71E+08 | 1.71E+08 | + | 3095 | protein_co | olfactomec | - | 1820 | 1896 | 1835 | 1230 | 1419 | 1367 | 30.92752 | 32.57095 | 29.16764 | 21.85214 | 24.53141 | 22.60887 |
| ENSMUSG0 | 8807.283 | 9063.653 | 9346.407 | 10533.94 | 10431.89 | 10732.61 | -0.21983 | 4.15E-08 | 2.12E-06 | Ckap4 | 10 | 84526305 | 84534062 | - | 3089 | protein_co | cytoskeletc | - | 8839 | 8998 | 10028 | 10112 | 10291 | 11067 | 150.4941 | 154.8748 | 159.7064 | 179.9984 | 178.2545 | 183.3931 |
| ENSMUSG0 | 876.8423 | 850.1581 | 896.6139 | 1208.403 | 1222.51 | 1168.591 | -0.45593 | 4.92E-08 | 2.50E-06 | Maml1 | 11 | 50255634 | 50292311 | - | 5528 | protein_co | mastermin | - | 880 | 844 | 962 | 1160 | 1206 | 1205 | 8.372381 | 8.117592 | 8.561166 | 11.53824 | 11.67294 | 11.1581 |
| ENSMUSG0 | 1219.608 | 1210.77 | 1339.329 | 1696.973 | 1648.26 | 1665.121 | -0.40999 | 5.03E-08 | 2.55E-06 | Sesn2 | 4 | 1.32E+08 | 1.33E+08 | - | 3475 | protein_co | sestrin 2 [So | - | 1224 | 1202 | 1437 | 1629 | 1626 | 1717 | 18.52512 | 18.39088 | 20.3436 | 25.77601 | 25.03609 | 25.2922 |
| ENSMUSG0 | 1047.229 | 1028.45 | 1140.806 | 1364.662 | 1545.878 | 1512.865 | -0.45949 | 5.25E-08 | 2.65E-06 | Anxa8 | 14 | 34085981 | 34100571 | + | 1880 | protein_co | annexin A8 | - | 1051 | 1021 | 1224 | 1310 | 1525 | 1560 | 29.40216 | 28.87491 | 32.02945 | 38.31448 | 43.40231 | 42.47543 |
| ENSMUSG0 | 758.2693 | 722.2315 | 770.7897 | 1049.019 | 1020.786 | 1082.28 | -0.48543 | 6.30E-08 | 3.17E-06 | S100a6 | 3 | 90612882 | 90624181 | + | 1367 | protein_co | S100 calciu | - | 761 | 717 | 827 | 1007 | 1007 | 1116 | 29.27861 | 27.88711 | 29.76205 | 40.50517 | 39.41502 | 41.78946 |
| ENSMUSG0 | 2180.149 | 2107.264 | 2235.942 | 1788.645 | 1570.206 | 1647.665 | 0.382143 | 6.59E-08 | 3.31E-06 | Rtl3 | X | 1.07E+08 | 1.07E+08 | - | 2948 | protein_co | retrotransp | - | 2188 | 2092 | 2399 | 1717 | 1549 | 1699 | 39.035 | 37.73002 | 40.03397 | 32.02523 | 28.11414 | 29.50102 |
| ENSMUSG0 | 520.1269 | 539.9109 | 524.7335 | 793.796 | 750.1308 | 784.5562 | -0.55518 | 6.69E-08 | 3.34E-06 | Sdad1 | 5 | 92284010 | 92310479 | - | 6537 | protein_co | SDA1 doma | - | 522 | 536 | 563 | 762 | 740 | 809 | 4.199778 | 4.359524 | 4.236974 | 6.409527 | 6.056951 | 6.33492 |
| ENSMUSG0 | 2729.172 | 2539.394 | 2690.774 | 3199.144 | 3280.302 | 3379.701 | -0.30883 | 6.89E-08 | 3.42E-06 | Pcsk6 | 7 | 65861734 | 66050386 | + | 5190 | protein_co | proprotein | - | 2739 | 2521 | 2887 | 3071 | 3236 | 3485 | 27.75614 | 25.82607 | 27.36562 | 32.53583 | 33.36122 | 34.37213 |
| ENSMUSG0 | 66.75958 | 86.62749 | 96.93123 | 231.2634 | 199.697 | 175.5311 | -1.27476 | 6.91E-08 | 3.42E-06 | Hmga2 | 10 | 1.2E+08 | 1.2E+08 | - | 6772 | protein_co | high mobili | HMGI/HMG | 67 | 86 | 104 | 222 | 197 | 181 | 0.520346 | 0.675203 | 0.755514 | 1.802542 | 1.556503 | 1.368147 |
| ENSMUSG0 | 6845.348 | 6608.872 | 6913.806 | 7791.077 | 7967.606 | 8067.643 | -0.22623 | 6.92E-08 | 3.42E-06 | Insig1 | 5 | 28071363 | 28078662 | + | 3093 | protein_co | insulin indu | - | 6870 | 6561 | 7418 | 7479 | 7860 | 8319 | 116.8184 | 112.7828 | 117.9866 | 132.9576 | 135.9701 | 137.6773 |
| ENSMUSG0 | 545.0372 | 564.086 | 623.5288 | 1067.77 | 778.5141 | 891.2326 | -0.65919 | 7.08E-08 | 3.48E-06 | Nab2 | 10 | 1.28E+08 | 1.28E+08 | - | 2941 | protein_co | Ngfi-A bind | - | 547 | 560 | 669 | 1025 | 768 | 919 | 9.781977 | 10.12385 | 11.19069 | 19.16365 | 13.97227 | 15.99527 |
| ENSMUSG0 | 2614.584 | 2612.927 | 2754.152 | 3401.239 | 3356.329 | 3146.953 | -0.31104 | 8.55E-08 | 4.19E-06 | Map7d1 | 4 | 1.26E+08 | 1.26E+08 | - | 3652 | protein_co | MAP7 dom | - | 2624 | 2594 | 2955 | 3265 | 3311 | 3245 | 37.78917 | 37.76522 | 39.80637 | 49.15887 | 48.50977 | 45.48362 |
| ENSMUSG0 | 1853.326 | 1629.806 | 1798.82 | 2164.709 | 2460.226 | 2327.484 | -0.3964 | 8.71E-08 | 4.26E-06 | Lysmd3 | 13 | 81657663 | 81672871 | + | 4250 | protein_co | LysM, puta | - | 1860 | 1618 | 1930 | 2078 | 2427 | 2400 | 23.01751 | 20.24149 | 22.34057 | 26.88475 | 30.55496 | 28.90636 |
| ENSMUSG0 | 450.3781 | 406.9477 | 381.2007 | 617.7441 | 689.3094 | 630.3604 | -0.64603 | 8.87E-08 | 4.32E-06 | Cldn12 | 5 | 5489537 | 5514958 | - | 4228 | protein_co | claudin 12 | - | 452 | 404 | 409 | 593 | 680 | 650 | 5.622608 | 5.080415 | 4.758984 | 7.712038 | 8.605473 | 7.869542 |
| ENSMUSG0 | 1805.498 | 1816.155 | 1738.238 | 2197.002 | 2459.213 | 2241.174 | -0.36401 | 9.72E-08 | 4.71E-06 | Tars | 15 | 11382301 | 11399665 | - | 5901 | protein_co | threonyl-tR | - | 1812 | 1803 | 1865 | 2109 | 2426 | 2311 | 16.14979 | 16.24512 | 15.54816 | 19.65171 | 21.99713 | 20.04682 |
| ENSMUSG0 | 3329.011 | 3294.866 | 3474.612 | 4036.692 | 4049.693 | 4083.765 | -0.26906 | 1.11E-07 | 5.35E-06 | Sgk1 | 10 | 21882184 | 21999903 | + | 4903 | protein_co | serum/gluc | - | 3341 | 3271 | 3728 | 3875 | 3995 | 4211 | 35.83843 | 35.47084 | 37.40588 | 43.45695 | 43.59691 | 43.96372 |
| ENSMUSG0 | 685.5312 | 810.8736 | 684.1108 | 372.9383 | 477.4481 | 493.6207 | 0.697088 | 1.14E-07 | 5.48E-06 | Itih5 | 2 | 10153571 | 10256529 | + | 7900 | protein_co | inter-alpha | - | 688 | 805 | 734 | 358 | 471 | 509 | 4.580319 | 5.417783 | 4.570828 | 2.491755 | 3.190029 | 3.298084 |
| ENSMUSG0 | 94.65911 | 68.49615 | 97.86326 | 21.87627 | 25.34226 | 8.728067 | 2.22749 | 1.27E-07 | 6.08E-06 | Ckm | 7 | 19404776 | 19422841 | + | 2913 | protein_co | creatine kir | - | 95 | 68 | 105 | 21 | 25 | 9 | 1.715211 | 1.241141 | 1.773269 | 0.396395 | 0.459198 | 0.158151 |
| ENSMUSG0 | 5511.153 | 5131.168 | 5205.393 | 6128.48 | 6270.688 | 6451.011 | -0.25043 | 1.38E-07 | 6.60E-06 | Klf6 | 13 | 5861482 | 5870394 | + | 4391 | protein_co | Kruppel-lik | zf-C2H2 | 5531 | 5094 | 5585 | 5883 | 6186 | 6652 | 66.24827 | 61.68056 | 62.57281 | 73.66902 | 75.37847 | 77.54609 |
| ENSMUSG0 | 203.268 | 219.5906 | 219.0273 | 92.71371 | 63.86249 | 113.4649 | 1.247914 | 1.49E-07 | 7.09E-06 | Meox2 | 12 | 37108540 | 37179534 | + | 2360 | protein_co | mesenchyn | Homeobox | 204 | 218 | 235 | 89 | 63 | 117 | 4.546242 | 4.911309 | 4.89871 | 2.073612 | 1.428333 | 2.537727 |
| ENSMUSG0 | 14233.74 | 14096.11 | 14623.57 | 12938.25 | 11676.7 | 12050.55 | 0.228452 | 1.55E-07 | 7.37E-06 | Wwp2 | 8 | 1.07E+08 | 1.08E+08 | + | 8871 | protein_co | WW domai | - | 14285 | 13994 | 15690 | 12420 | 11519 | 12426 | 84.69192 | 83.87298 | 87.01142 | 76.98365 | 69.47731 | 71.70177 |
| ENSMUSG0 | 408.5288 | 445.225 | 464.1515 | 279.1828 | 221.9982 | 265.7211 | 0.781476 | 1.60E-07 | 7.55E-06 | Cd34 | 1 | 1.95E+08 | 1.95E+08 | + | 7290 | protein_co | CD34 antig | - | 410 | 442 | 498 | 268 | 219 | 274 | 2.957949 | 3.223647 | 3.360684 | 2.021421 | 1.607376 | 1.923951 |
| ENSMUSG0 | 1254.482 | 1266.172 | 1306.708 | 1631.345 | 1615.822 | 1777.616 | -0.39281 | 1.65E-07 | 7.79E-06 | Bmp6 | 13 | 38345107 | 38500302 | + | 3900 | protein_co | bone morp | - | 1259 | 1257 | 1402 | 1566 | 1594 | 1833 | 16.97835 | 17.13655 | 17.68517 | 22.07886 | 21.86878 | 24.05852 |
| ENSMUSG0 | 2344.557 | 2368.154 | 2301.185 | 2756.41 | 3063.372 | 2928.751 | -0.31902 | 1.78E-07 | 8.36E-06 | Pde4b | 4 | 1.02E+08 | 1.03E+08 | + | 10560 | protein_co | phosphodi | - | 2353 | 2351 | 2469 | 2646 | 3022 | 3020 | 11.71905 | 11.837 | 11.50226 | 13.77766 | 15.31198 | 14.63909 |
| ENSMUSG0 | 514.1484 | 464.3636 | 517.2772 | 751.0852 | 726.8159 | 730.2483 | -0.5615 | 1.83E-07 | 8.54E-06 | Rnf41 | 10 | 1.28E+08 | 1.28E+08 | + | 4457 | protein_co | ring finger | - | 516 | 461 | 555 | 721 | 717 | 753 | 6.088936 | 5.499347 | 6.12599 | 8.894921 | 8.607506 | 8.648154 |
| ENSMUSG0 | 749.3016 | 763.5307 | 691.567 | 939.6378 | 1198.182 | 1086.159 | -0.54902 | 1.88E-07 | 8.75E-06 | Hsd17b7 | 1 | 1.7E+08 | 1.7E+08 | - | 5104 | protein_co | hydroxyste | - | 752 | 758 | 742 | 902 | 1182 | 1120 | 7.748926 | 7.896076 | 7.151862 | 9.717294 | 12.39104 | 11.23255 |
| ENSMUSG0 | 1102.031 | 1086.873 | 1078.36 | 1353.203 | 1493.166 | 1519.653 | -0.41858 | 1.89E-07 | 8.77E-06 | Arfgef2 | 2 | 1.67E+08 | 1.67E+08 | + | 8770 | protein_co | ADP-ribosy | - | 1106 | 1079 | 1157 | 1299 | 1473 | 1567 | 6.632693 | 6.541459 | 6.490224 | 8.144399 | 8.986777 | 9.146196 |
| ENSMUSG0 | 290.9522 | 265.9262 | 262.8328 | 146.8835 | 135.8345 | 145.4678 | 0.936696 | 1.96E-07 | 9.06E-06 | Mylk | 16 | 34745210 | 35002420 | + | 11263 | protein_co | myosin, lig | - | 292 | 264 | 282 | 141 | 134 | 150 | 1.363525 | 1.246243 | 1.231745 | 0.688358 | 0.636578 | 0.681723 |
| ENSMUSG0 | 1484.653 | 1528.069 | 1534.123 | 1949.071 | 1989.874 | 1867.806 | -0.35267 | 2.07E-07 | 9.55E-06 | Xpot | 10 | 1.22E+08 | 1.22E+08 | - | 6033 | protein_co | exportin, tl | - | 1490 | 1517 | 1646 | 1871 | 1963 | 1926 | 12.98935 | 13.36919 | 13.42216 | 17.05257 | 17.40956 | 16.34158 |
| ENSMUSG0 | 31438.78 | 31259.43 | 32351.73 | 35017.65 | 36121.84 | 35658.03 | -0.1681 | 2.08E-07 | 9.55E-06 | Col6a2 | 10 | 76595762 | 76623630 | - | 8326 | protein_co | collagen, ty | - | 31552 | 31033 | 34711 | 33615 | 35634 | 36769 | 199.308 | 198.171 | 205.0957 | 221.9965 | 228.9966 | 226.0562 |
| ENSMUSG0 | 1022.318 | 1015.355 | 1083.952 | 749.0017 | 700.46 | 785.526 | 0.482031 | 2.09E-07 | 9.56E-06 | Rflna | 5 | 1.25E+08 | 1.25E+08 | + | 1940 | protein_co | refilin A [So | - | 1026 | 1008 | 1163 | 719 | 691 | 810 | 27.81506 | 27.62559 | 29.49198 | 20.37871 | 19.05799 | 21.37245 |
| ENSMUSG0 | 737.3446 | 749.4285 | 781.042 | 982.3486 | 1071.471 | 1115.253 | -0.48287 | 2.10E-07 | 9.58E-06 | Cfap43 | 19 | 47737561 | 47919299 | - | 9955 | protein_co | cilia and fla | - | 740 | 744 | 838 | 943 | 1057 | 1150 | 3.909532 | 3.973603 | 4.141223 | 5.208587 | 5.681127 | 5.913269 |
| ENSMUSG0 | 1472.696 | 1445.47 | 1346.785 | 1943.863 | 1973.655 | 1723.308 | -0.40337 | 2.19E-07 | 9.94E-06 | Usp36 | 11 | 1.18E+08 | 1.18E+08 | - | 7689 | protein_co | ubiquitin s | - | 1478 | 1435 | 1445 | 1866 | 1947 | 1777 | 10.10971 | 9.922814 | 9.245361 | 13.34416 | 13.54868 | 11.83011 |
| ENSMUSG0 | 2351.532 | 2433.628 | 2582.658 | 1991.782 | 1708.068 | 1959.936 | 0.380655 | 2.24E-07 | 1.02E-05 | Synpo | 18 | 60593973 | 60660142 | - | 9533 | protein_co | synaptopoc | - | 2360 | 2416 | 2771 | 1912 | 1685 | 2021 | 13.02017 | 13.47473 | 14.29989 | 11.02828 | 9.457386 | 10.85195 |
| ENSMUSG0 | 265.0455 | 207.5031 | 230.2117 | 127.0907 | 66.90356 | 101.8274 | 1.249025 | 2.27E-07 | 1.02E-05 | Myh11 | 16 | 14194535 | 14291372 | - | 7599 | protein_co | myosin, he | - | 266 | 206 | 247 | 122 | 66 | 105 | 1.841024 | 1.44133 | 1.599066 | 0.882781 | 0.464717 | 0.7073 |
| ENSMUSG0 | 4947.184 | 4816.891 | 5132.695 | 5722.207 | 5878.39 | 6138.74 | -0.25194 | 2.28E-07 | 1.03E-05 | Ctdsp2 | 10 | 1.27E+08 | 1.27E+08 | + | 5524 | protein_co | CTD (carbo | - | 4965 | 4782 | 5507 | 5493 | 5799 | 6330 | 47.27156 | 46.02658 | 49.04416 | 54.67709 | 56.16946 | 58.65717 |
| ENSMUSG0 | 1375.048 | 1394.098 | 1454.9 | 1711.558 | 1919.929 | 1880.414 | -0.38393 | 2.31E-07 | 1.04E-05 | Ibtk | 9 | 85687360 | 85749334 | - | 8628 | protein_co | inhibitor o | - | 1380 | 1384 | 1561 | 1643 | 1894 | 1939 | 8.412078 | 8.52862 | 8.900587 | 10.47073 | 11.74548 | 11.50373 |
| ENSMUSG0 | 873.853 | 747.4139 | 833.2357 | 1092.772 | 1198.182 | 1138.528 | -0.48244 | 2.36E-07 | 1.05E-05 | Golt1b | 6 | 1.42E+08 | 1.42E+08 | + | 2818 | protein_co | golgi transp | - | 877 | 742 | 894 | 1049 | 1182 | 1174 | 16.3679 | 13.9996 | 15.60711 | 20.4684 | 22.44281 | 21.32545 |
| ENSMUSG0 | 7517.926 | 7280.738 | 7757.294 | 8771.342 | 8713.682 | 8683.457 | -0.21418 | 2.40E-07 | 1.07E-05 | Angptl2 | 2 | 33216069 | 33247717 | + | 6286 | protein_co | angiopoieti | - | 7545 | 7228 | 8323 | 8420 | 8596 | 8954 | 63.12759 | 61.13594 | 65.13755 | 73.65245 | 73.16828 | 72.91448 |
| ENSMUSG0 | 230.1711 | 227.649 | 232.0757 | 331.2692 | 436.9005 | 484.8926 | -0.86171 | 2.48E-07 | 1.10E-05 | Cx3cl1 | 8 | 94772009 | 94782427 | + | 3930 | protein_co | chemokine | - | 231 | 226 | 249 | 318 | 431 | 500 | 3.09139 | 3.057516 | 3.116971 | 4.449222 | 5.867938 | 6.512512 |
| ENSMUSG0 | 1673.972 | 1678.156 | 1717.733 | 2064.703 | 2217.954 | 2122.86 | -0.33732 | 2.63E-07 | 1.16E-05 | Rmnd5a | 6 | 71388634 | 71440637 | - | 6783 | protein_co | required fo | - | 1680 | 1666 | 1843 | 1982 | 2188 | 2189 | 13.02632 | 13.05888 | 13.36686 | 16.06687 | 17.25942 | 16.51943 |
| ENSMUSG0 | 864.8853 | 1046.581 | 1038.283 | 1334.452 | 1314.756 | 1483.771 | -0.48661 | 2.87E-07 | 1.27E-05 | Psat1 | 19 | 15904678 | 15947337 | - | 4254 | protein_co | phosphose | - | 868 | 1039 | 1114 | 1281 | 1297 | 1530 | 10.7314 | 12.98586 | 12.8829 | 16.55774 | 16.31336 | 18.41048 |
| ENSMUSG0 | 6367.071 | 6284.522 | 6256.724 | 7210.835 | 7745.607 | 7232.658 | -0.23084 | 2.90E-07 | 1.28E-05 | Fndc3b | 3 | 27416162 | 27711307 | - | 11485 | protein_co | fibronectin | - | 6390 | 6239 | 6713 | 6922 | 7641 | 7458 | 29.26201 | 28.88263 | 28.75488 | 33.13981 | 35.59754 | 33.24011 |
| ENSMUSG0 | 54.80264 | 77.56182 | 85.74685 | 144.8001 | 193.6148 | 206.5642 | -1.32046 | 3.53E-07 | 1.55E-05 | Gprc5a | 6 | 1.35E+08 | 1.35E+08 | + | 2083 | protein_co | G protein-c | - | 55 | 77 | 92 | 139 | 191 | 213 | 1.388698 | 1.965415 | 2.172824 | 3.66923 | 4.906196 | 5.234334 |
| ENSMUSG0 | 2021.719 | 1970.272 | 1899.479 | 2324.093 | 2513.952 | 2625.209 | -0.34159 | 3.72E-07 | 1.62E-05 | Tgfbr3 | 5 | 1.07E+08 | 1.07E+08 | - | 6545 | protein_co | transformi | - | 2029 | 1956 | 2038 | 2231 | 2480 | 2707 | 16.30447 | 15.88956 | 15.31865 | 18.74301 | 20.27416 | 21.17141 |
| ENSMUSG0 | 10529.08 | 10687.41 | 10764.96 | 9054.691 | 9449.621 | 9414.675 | 0.195954 | 4.09E-07 | 1.78E-05 | Chst11 | 10 | 82985498 | 83195900 | + | 5527 | protein_co | carbohydra | - | 10567 | 10610 | 11550 | 8692 | 9322 | 9708 | 100.5534 | 102.0654 | 102.806 | 86.47284 | 90.24444 | 89.9107 |
| ENSMUSG0 | 2179.152 | 2334.913 | 2146.467 | 1629.261 | 1873.3 | 1591.418 | 0.386811 | 4.13E-07 | 1.79E-05 | Abi3bp | 16 | 56477878 | 56690135 | + | 7416 | protein_co | ABI gene fa | - | 2187 | 2318 | 2303 | 1564 | 1848 | 1641 | 15.51006 | 16.61868 | 15.27742 | 11.59622 | 13.33316 | 11.32687 |
| ENSMUSG0 | 187.3254 | 256.8606 | 172.4257 | 68.75398 | 88.19105 | 101.8274 | 1.250321 | 4.15E-07 | 1.79E-05 | Thy1 | 9 | 44043384 | 44048579 | + | 2995 | protein_co | thymus cell | - | 188 | 255 | 185 | 66 | 87 | 105 | 3.301379 | 4.52685 | 3.038791 | 1.211704 | 1.554258 | 1.794583 |
| ENSMUSG0 | 995.4153 | 1006.289 | 941.3513 | 1337.578 | 1266.099 | 1306.301 | -0.41009 | 4.32E-07 | 1.86E-05 | Timp1 | X | 20870166 | 20874735 | + | 888 | protein_co | tissue inhib | - | 999 | 999 | 1010 | 1284 | 1249 | 1347 | 59.168 | 59.81434 | 55.95441 | 79.5063 | 75.25759 | 77.64718 |
| ENSMUSG0 | 455.3601 | 429.1083 | 449.239 | 241.6807 | 288.9017 | 281.2377 | 0.715745 | 4.51E-07 | 1.94E-05 | Egfr | 11 | 16752203 | 16918158 | + | 11326 | protein_co | epidermal g | - | 457 | 426 | 482 | 232 | 285 | 290 | 2.12214 | 1.999797 | 2.093613 | 1.126318 | 1.346385 | 1.310667 |
| ENSMUSG0 | 912.7131 | 903.5448 | 987.0209 | 1206.32 | 1345.167 | 1244.234 | -0.43693 | 4.63E-07 | 1.98E-05 | Trp53bp2 | 1 | 1.82E+08 | 1.82E+08 | + | 4418 | protein_co | transforma | - | 916 | 897 | 1059 | 1158 | 1327 | 1283 | 10.90446 | 10.79492 | 11.79224 | 14.41227 | 16.07112 | 14.86524 |
| ENSMUSG0 | 3301.112 | 3306.954 | 3297.526 | 4089.82 | 4084.158 | 3775.374 | -0.27045 | 4.82E-07 | 2.05E-05 | Slc39a14 | 14 | 70303469 | 70351425 | - | 10550 | protein_co | solute carri | - | 3313 | 3283 | 3538 | 3926 | 4029 | 3893 | 16.51594 | 16.54517 | 16.498 | 20.46196 | 20.43363 | 18.88874 |
| ENSMUSG0 | 2111.396 | 2150.578 | 2121.303 | 2626.194 | 2601.129 | 2588.357 | -0.29206 | 4.83E-07 | 2.05E-05 | Scarb2 | 5 | 92441314 | 92506833 | - | 6285 | protein_co | scavenger r | - | 2119 | 2135 | 2276 | 2521 | 2566 | 2669 | 17.73209 | 18.06115 | 17.81529 | 22.05551 | 21.84501 | 21.73774 |
| ENSMUSG0 | 7358.5 | 7911.306 | 7922.264 | 9329.707 | 9346.224 | 8680.547 | -0.23811 | 4.84E-07 | 2.05E-05 | P3h1 | 4 | 1.19E+08 | 1.19E+08 | + | 3846 | protein_co | prolyl 3-hy | - | 7385 | 7854 | 8500 | 8956 | 9220 | 8951 | 100.9893 | 108.5761 | 108.7265 | 128.0425 | 128.2692 | 119.1334 |
| ENSMUSG0 | 3081.901 | 3118.59 | 3027.237 | 3596.042 | 3839.859 | 3677.425 | -0.26836 | 4.86E-07 | 2.06E-05 | Stat3 | 11 | 1.01E+08 | 1.01E+08 | - | 9527 | protein_co | signal trans | STAT | 3093 | 3096 | 3248 | 3452 | 3788 | 3792 | 17.0749 | 17.27816 | 16.77203 | 19.92343 | 21.27427 | 20.37433 |
| ENSMUSG0 | 1566.359 | 1590.521 | 1528.531 | 1979.281 | 2096.312 | 1895.93 | -0.34992 | 5.06E-07 | 2.13E-05 | Mybbp1a | 11 | 72441355 | 72451768 | + | 5803 | protein_co | MYB bindir | - | 1572 | 1579 | 1640 | 1900 | 2068 | 1955 | 14.24736 | 14.46713 | 13.90328 | 18.00323 | 19.06772 | 17.24508 |
| ENSMUSG0 | 894.7777 | 753.4577 | 783.8381 | 1052.144 | 1194.127 | 1172.47 | -0.49161 | 5.17E-07 | 2.17E-05 | Cebpg | 7 | 35046422 | 35056573 | - | 5621 | protein_co | CCAAT/enh | C/EBP | 898 | 748 | 841 | 1010 | 1178 | 1209 | 8.402279 | 7.075234 | 7.360517 | 9.880007 | 11.21328 | 11.00991 |
| ENSMUSG0 | 1895.175 | 1888.681 | 1814.664 | 2478.269 | 2421.706 | 2196.563 | -0.34194 | 5.18E-07 | 2.17E-05 | Tnks2 | 19 | 36834232 | 36893477 | + | 10406 | protein_co | tankyrase, | - | 1902 | 1875 | 1947 | 2379 | 2389 | 2265 | 9.613046 | 9.580105 | 9.204666 | 12.57072 | 12.28381 | 11.1418 |
| ENSMUSG0 | 192.3075 | 183.3279 | 191.0664 | 377.1052 | 379.1202 | 282.2075 | -0.87291 | 5.23E-07 | 2.18E-05 | Glis3 | 19 | 28258851 | 28680077 | - | 8842 | protein_co | GLIS family | zf-C2H2 | 193 | 182 | 205 | 362 | 374 | 291 | 1.147998 | 1.094394 | 1.140589 | 2.251166 | 2.263195 | 1.684665 |
| ENSMUSG0 | 847.9463 | 808.859 | 855.6045 | 1096.939 | 1187.031 | 1117.193 | -0.4368 | 5.27E-07 | 2.19E-05 | Atrn | 2 | 1.31E+08 | 1.31E+08 | + | 13208 | protein_co | attractin [S | - | 851 | 803 | 918 | 1053 | 1171 | 1152 | 3.388651 | 3.232446 | 3.419255 | 4.383699 | 4.743737 | 4.464641 |
| ENSMUSG0 | 2061.576 | 2063.95 | 2156.72 | 2593.9 | 2868.744 | 2489.439 | -0.33979 | 5.60E-07 | 2.32E-05 | Mef2d | 3 | 88142372 | 88172086 | + | 5985 | protein_co | myocyte en | SRF | 2069 | 2049 | 2314 | 2490 | 2830 | 2567 | 18.18154 | 18.20248 | 19.02064 | 22.87624 | 25.30015 | 21.95497 |
| ENSMUSG0 | 5957.545 | 5970.246 | 6636.993 | 7435.848 | 7430.35 | 7230.718 | -0.25102 | 5.88E-07 | 2.43E-05 | Fasn | 11 | 1.21E+08 | 1.21E+08 | - | 10947 | protein_co | fatty acid s | - | 5979 | 5927 | 7121 | 7138 | 7330 | 7456 | 28.72551 | 28.78675 | 32.00161 | 35.85344 | 35.82694 | 34.86437 |
| ENSMUSG0 | 5904.736 | 5585.458 | 5615.487 | 4783.611 | 4927.548 | 4852.805 | 0.232006 | 5.96E-07 | 2.46E-05 | Tnc | 4 | 63959785 | 64047015 | - | 7772 | protein_co | tenascin C [ | - | 5926 | 5545 | 6025 | 4592 | 4861 | 5004 | 40.10174 | 37.93338 | 38.13732 | 32.48767 | 33.46522 | 32.9576 |
| ENSMUSG0 | 5655.633 | 5977.297 | 5430.013 | 4932.578 | 4724.81 | 4322.333 | 0.287613 | 6.04E-07 | 2.48E-05 | Thbs4 | 13 | 92751590 | 92794818 | - | 3289 | protein_co | thrombosp | - | 5676 | 5934 | 5826 | 4735 | 4661 | 4457 | 90.76384 | 95.92603 | 87.143 | 79.15996 | 75.82563 | 69.36651 |
| ENSMUSG0 | 624.7501 | 590.2757 | 682.2467 | 844.8406 | 1025.855 | 893.1722 | -0.54232 | 6.10E-07 | 2.50E-05 | Slc5a3 | 16 | 92058322 | 92087473 | + | 10913 | protein_co | solute carri | - | 627 | 586 | 732 | 811 | 1012 | 921 | 3.021744 | 2.855001 | 3.29984 | 4.086262 | 4.961776 | 4.320028 |
| ENSMUSG0 | 3798.321 | 4205.462 | 4147.538 | 5184.675 | 4912.343 | 4698.609 | -0.28385 | 6.22E-07 | 2.54E-05 | P3h4 | 11 | 1E+08 | 1E+08 | - | 2178 | protein_co | prolyl 3-hy | - | 3812 | 4175 | 4450 | 4977 | 4846 | 4845 | 92.05118 | 101.9181 | 100.5143 | 125.6491 | 119.0492 | 113.8694 |
| ENSMUSG0 | 1965.92 | 1970.272 | 2050.468 | 2310.551 | 2737.977 | 2618.42 | -0.35697 | 6.25E-07 | 2.54E-05 | Mef2a | 7 | 67231163 | 67372858 | - | 14681 | protein_co | myocyte en | SRF | 1973 | 1956 | 2200 | 2218 | 2701 | 2700 | 7.06815 | 7.083795 | 7.372129 | 8.307213 | 9.843958 | 9.414109 |
| ENSMUSG0 | 192.3075 | 162.1747 | 174.2898 | 76.04607 | 56.76666 | 90.19002 | 1.244431 | 6.33E-07 | 2.57E-05 | Steap4 | 5 | 7960457 | 7982213 | + | 3155 | protein_co | STEAP famil | - | 193 | 161 | 187 | 73 | 56 | 93 | 3.217305 | 2.713184 | 2.91587 | 1.272251 | 0.949707 | 1.50888 |
| ENSMUSG0 | 38.86006 | 61.44508 | 74.56248 | 8.333816 | 14.19166 | 6.788496 | 2.579538 | 6.62E-07 | 2.68E-05 | Galnt17 | 5 | 1.31E+08 | 1.31E+08 | - | 9408 | protein_co | polypeptid | - | 39 | 61 | 80 | 8 | 14 | 7 | 0.218023 | 0.344735 | 0.41833 | 0.046757 | 0.079622 | 0.038087 |
| ENSMUSG0 | 748.3052 | 651.7208 | 576.9272 | 904.2191 | 1116.073 | 940.6916 | -0.58338 | 7.10E-07 | 2.86E-05 | Eaf1 | 14 | 31494399 | 31509858 | + | 5490 | protein_co | ELL associa | - | 751 | 647 | 619 | 868 | 1101 | 970 | 7.194522 | 6.265918 | 5.546822 | 8.693544 | 10.7304 | 9.044207 |
| ENSMUSG0 | 735.3518 | 648.6989 | 626.3249 | 959.4306 | 881.9105 | 1045.428 | -0.52258 | 7.87E-07 | 3.17E-05 | Wbp1l | 19 | 46599084 | 46657389 | + | 4463 | protein_co | WW domai | - | 738 | 644 | 672 | 921 | 870 | 1078 | 8.696887 | 7.672056 | 7.407442 | 11.34703 | 10.43021 | 12.36411 |
| ENSMUSG0 | 271.024 | 310.2473 | 276.8132 | 456.2764 | 488.5987 | 434.4638 | -0.68496 | 8.05E-07 | 3.23E-05 | B4galt5 | 2 | 1.67E+08 | 1.67E+08 | - | 4308 | protein_co | UDP-Gal:be | - | 272 | 308 | 297 | 438 | 482 | 448 | 3.320684 | 3.801262 | 3.391616 | 5.590464 | 5.986489 | 5.323208 |
| ENSMUSG0 | 2333.596 | 2277.497 | 2333.806 | 2786.62 | 2911.318 | 2763.888 | -0.28492 | 8.52E-07 | 3.40E-05 | Fbxw11 | 11 | 32642724 | 32746816 | + | 4327 | protein_co | F-box and V | - | 2342 | 2261 | 2504 | 2675 | 2872 | 2850 | 28.46652 | 27.78219 | 28.46907 | 33.99275 | 35.5139 | 33.71546 |
| ENSMUSG0 | 1273.414 | 1359.85 | 1424.143 | 1820.939 | 1690.835 | 1733.976 | -0.3702 | 8.56E-07 | 3.40E-05 | Lzts2 | 19 | 45015176 | 45027104 | + | 2921 | protein_co | leucine zip | - | 1278 | 1350 | 1528 | 1748 | 1668 | 1788 | 23.0109 | 24.57282 | 25.73462 | 32.90481 | 30.5538 | 31.33337 |
| ENSMUSG0 | 23347.92 | 23644.27 | 23633.51 | 27918.28 | 25862.28 | 26358.76 | -0.18228 | 8.57E-07 | 3.40E-05 | Fkbp9 | 6 | 56832059 | 56879358 | + | 3314 | protein_co | FK506 binc | - | 23432 | 23473 | 25357 | 26800 | 25513 | 27180 | 371.87 | 376.5901 | 376.4188 | 444.6638 | 411.9171 | 419.8247 |
| ENSMUSG0 | 858.9069 | 899.5157 | 942.2834 | 1162.567 | 1187.031 | 1291.754 | -0.43115 | 8.90E-07 | 3.52E-05 | Tiam1 | 16 | 89787111 | 90143769 | - | 10048 | protein_co | T cell lympl | - | 862 | 893 | 1011 | 1116 | 1171 | 1332 | 4.511926 | 4.725248 | 4.949911 | 6.107086 | 6.235598 | 6.785716 |
| ENSMUSG0 | 158.4295 | 164.1893 | 166.8336 | 76.04607 | 39.53392 | 74.67346 | 1.363034 | 8.92E-07 | 3.52E-05 | Tnni2 | 7 | 1.42E+08 | 1.42E+08 | + | 1497 | protein_co | troponin I, | - | 159 | 163 | 179 | 73 | 39 | 77 | 5.586112 | 5.789201 | 5.882435 | 2.681332 | 1.393939 | 2.632934 |
| ENSMUSG0 | 3473.491 | 3474.165 | 3295.662 | 3930.436 | 4446.046 | 4110.919 | -0.28598 | 8.97E-07 | 3.53E-05 | Aff4 | 11 | 53350833 | 53421830 | + | 13450 | protein_co | AF4/FMR2 | AF-4 | 3486 | 3449 | 3536 | 3773 | 4386 | 4239 | 13.63137 | 13.63401 | 12.93349 | 15.4246 | 17.44806 | 16.13289 |
| ENSMUSG0 | 2446.191 | 2664.299 | 2734.579 | 2093.871 | 2171.325 | 1888.172 | 0.350806 | 9.00E-07 | 3.53E-05 | Amotl2 | 9 | 1.03E+08 | 1.03E+08 | + | 6289 | protein_co | angiomotir | - | 2455 | 2645 | 2934 | 2010 | 2142 | 1947 | 20.53072 | 22.36129 | 22.95115 | 17.57373 | 18.22379 | 15.84731 |
| ENSMUSG0 | 3378.832 | 3564.822 | 3523.077 | 2775.161 | 2899.154 | 2981.12 | 0.273972 | 9.14E-07 | 3.57E-05 | Ldlrad3 | 2 | 1.02E+08 | 1.02E+08 | - | 6023 | protein_co | low density | - | 3391 | 3539 | 3780 | 2664 | 2860 | 3074 | 29.61074 | 31.24068 | 30.87485 | 24.32041 | 25.40703 | 26.12535 |

| gene_id | H1 | H2 | H3 | M1 | M2 | M3 | log2FoldCh | pvalue | padj | gene_name | gene_chr | gene_start | gene_end | gene_stran | gene_lengt | gene_bioty | gene_descr | tf_family | H1_readco | H2_readco | H3_readco | M1_readco | M2_readco | M3_readco | H1_fpkm | H2_fpkm | H3_fpkm | M1_fpkm | M2_fpkm | M3_fpkm |
|---|---|---|---|---|---|---|---|---|---|---|---|---|---|---|---|---|---|---|---|---|---|---|---|---|---|---|---|---|---|---|
| ENSMUSG0 | 1107.013 | 1173.5 | 1265.698 | 1515.713 | 1597.576 | 1536.14 | -0.39038 | 9.16E-07 | 3.57E-05 | Bcl6 | 16 | 23965052 | 23988852 | - | 6700 | protein_co | B cell leuke | ZBTB | 1111 | 1165 | 1358 | 1455 | 1576 | 1584 | 8.721147 | 9.244936 | 9.971279 | 11.94092 | 12.58584 | 12.10184 |
| ENSMUSG0 | 1844.358 | 2191.877 | 2080.293 | 2431.391 | 2718.717 | 2782.314 | -0.37522 | 9.42E-07 | 3.66E-05 | Mthfd2 | 6 | 83305691 | 83325908 | - | 3487 | protein_co | methylenet | - | 1851 | 2176 | 2232 | 2334 | 2682 | 2869 | 27.91829 | 33.17873 | 31.48967 | 36.80428 | 41.15358 | 42.11625 |
| ENSMUSG0 | 3891.984 | 4356.557 | 4486.797 | 5260.722 | 5085.684 | 5113.677 | -0.27951 | 9.60E-07 | 3.72E-05 | Txndc5 | 13 | 38500079 | 38528824 | - | 4492 | protein_co | thioredoxi | - | 3906 | 4325 | 4814 | 5050 | 5017 | 5273 | 45.7327 | 51.19165 | 52.72204 | 61.81603 | 59.75925 | 60.08819 |
| ENSMUSG0 | 5634.708 | 6033.705 | 5799.097 | 7012.906 | 7451.637 | 6504.349 | -0.2635 | 1.07E-06 | 4.14E-05 | Ctgf | 10 | 24595442 | 24598683 | + | 2861 | protein_co | connective | - | 5655 | 5990 | 6222 | 6732 | 7351 | 6707 | 103.9559 | 111.3171 | 106.9887 | 129.3826 | 137.4768 | 120.0001 |
| ENSMUSG0 | 121.5622 | 127.9266 | 95.9992 | 30.21008 | 45.61606 | 43.64033 | 1.529575 | 1.13E-06 | 4.34E-05 | AW551984 | 9 | 39587396 | 39604403 | - | 6082 | protein_co | expressed s | - | 122 | 127 | 103 | 29 | 45 | 45 | 1.054989 | 1.110223 | 0.833138 | 0.262181 | 0.395883 | 0.378737 |
| ENSMUSG0 | 552.0121 | 568.1152 | 444.5788 | 335.4361 | 340.5999 | 251.1744 | 0.755258 | 1.23E-06 | 4.73E-05 | Kera | 10 | 97606879 | 97613692 | + | 2280 | protein_co | keratocan [ | - | 554 | 564 | 477 | 322 | 336 | 259 | 12.77936 | 13.15216 | 10.29223 | 7.765519 | 7.885064 | 5.814816 |
| ENSMUSG0 | 2567.753 | 2637.102 | 2708.482 | 3209.561 | 3991.912 | 3134.346 | -0.38516 | 1.24E-06 | 4.74E-05 | Usp53 | 3 | 1.23E+08 | 1.23E+08 | - | 12572 | protein_co | ubiquitin s | - | 2577 | 2618 | 2906 | 3081 | 3938 | 3232 | 10.78064 | 11.0718 | 11.37148 | 13.47525 | 16.75993 | 13.15946 |
| ENSMUSG0 | 487.2453 | 524.8014 | 509.821 | 782.337 | 668.0219 | 765.1605 | -0.5418 | 1.27E-06 | 4.85E-05 | Cdca4 | 12 | 1.13E+08 | 1.13E+08 | - | 4198 | protein_co | cell divisio | - | 489 | 521 | 547 | 751 | 659 | 789 | 6.126335 | 6.598544 | 6.410188 | 9.836644 | 8.399313 | 9.620677 |
| ENSMUSG0 | 225.189 | 219.5906 | 238.5999 | 373.98 | 414.5993 | 352.032 | -0.73841 | 1.28E-06 | 4.87E-05 | Lrp8 | 4 | 1.08E+08 | 1.08E+08 | + | 10514 | protein_co | low density | - | 226 | 218 | 256 | 359 | 409 | 363 | 1.130511 | 1.102405 | 1.197837 | 1.877483 | 2.081403 | 1.767298 |
| ENSMUSG0 | 1019.329 | 997.2234 | 971.1763 | 1246.947 | 1313.743 | 1369.337 | -0.39591 | 1.32E-06 | 5.03E-05 | Dhcr24 | 4 | 1.07E+08 | 1.07E+08 | + | 4028 | protein_co | 24-dehydro | - | 1023 | 990 | 1042 | 1197 | 1296 | 1412 | 13.35736 | 13.06768 | 12.72636 | 16.34008 | 17.21537 | 17.94388 |
| ENSMUSG0 | 1224.59 | 1185.588 | 1267.562 | 1477.169 | 1670.562 | 1648.635 | -0.38314 | 1.34E-06 | 5.09E-05 | Klf10 | 15 | 38291463 | 38300706 | - | 4179 | protein_co | Kruppel-lik | zf-C2H2 | 1229 | 1177 | 1360 | 1418 | 1648 | 1700 | 15.46728 | 14.97466 | 16.01004 | 18.65749 | 21.10016 | 20.82321 |
| ENSMUSG0 | 1759.663 | 1627.791 | 1675.792 | 2186.585 | 2086.175 | 2065.642 | -0.32392 | 1.38E-06 | 5.22E-05 | Mtmr3 | 11 | 4480868 | 4594863 | - | 7588 | protein_co | myotubula | - | 1766 | 1616 | 1798 | 2099 | 2058 | 2130 | 12.24046 | 11.32314 | 11.65704 | 15.21019 | 14.51172 | 14.36889 |
| ENSMUSG0 | 2147.267 | 2175.76 | 2021.575 | 2624.11 | 2899.154 | 2487.499 | -0.33648 | 1.49E-06 | 5.59E-05 | P3h2 | 16 | 25959288 | 26105784 | - | 3721 | protein_co | prolyl 3-hy | - | 2155 | 2160 | 2169 | 2519 | 2860 | 2565 | 30.45944 | 30.86362 | 28.67648 | 37.22356 | 41.12512 | 35.2857 |
| ENSMUSG0 | 1678.954 | 1584.477 | 1674.86 | 2112.622 | 2214.913 | 1946.359 | -0.34502 | 1.56E-06 | 5.85E-05 | Raph1 | 1 | 60482292 | 60567104 | - | 20498 | protein_co | Ras associa | - | 1685 | 1573 | 1797 | 2028 | 2185 | 2007 | 4.323373 | 4.080093 | 4.312831 | 5.440088 | 5.703491 | 5.011953 |
| ENSMUSG0 | 146.4725 | 201.4593 | 202.2507 | 67.71226 | 64.87618 | 99.88788 | 1.241214 | 1.56E-06 | 5.85E-05 | Ly6a | 15 | 74994877 | 74998031 | - | 1205 | protein_co | lymphocyt | - | 147 | 200 | 217 | 65 | 64 | 103 | 6.416004 | 8.824615 | 8.859283 | 2.966032 | 2.841801 | 4.375435 |
| ENSMUSG0 | 1973.892 | 1959.191 | 2242.467 | 2575.149 | 2629.513 | 2543.747 | -0.32687 | 1.59E-06 | 5.94E-05 | Uap1 | 1 | 1.7E+08 | 1.7E+08 | - | 4011 | protein_co | UDP-N-ace | - | 1981 | 1945 | 2406 | 2472 | 2594 | 2623 | 25.97563 | 25.78219 | 29.50998 | 33.88795 | 34.60335 | 33.4747 |
| ENSMUSG0 | 2157.231 | 2295.628 | 2312.369 | 2820.997 | 2672.088 | 2782.314 | -0.29056 | 1.60E-06 | 5.95E-05 | Slc35e1 | 8 | 72480641 | 72492614 | - | 4470 | protein_co | solute carri | - | 2165 | 2279 | 2481 | 2708 | 2636 | 2869 | 25.47327 | 27.10751 | 27.30519 | 33.31122 | 31.55286 | 32.85444 |
| ENSMUSG0 | 1819.448 | 1900.768 | 1896.683 | 1511.546 | 1188.045 | 1486.681 | 0.424106 | 1.73E-06 | 6.42E-05 | Tubb2b | 13 | 34126748 | 34130466 | - | 2294 | protein_co | tubulin, be | - | 1826 | 1887 | 2035 | 1451 | 1172 | 1533 | 41.86409 | 43.73522 | 43.64122 | 34.77951 | 27.336 | 34.20738 |
| ENSMUSG0 | 1291.35 | 1390.069 | 1440.92 | 1781.353 | 1784.095 | 1701.973 | -0.35325 | 1.76E-06 | 6.50E-05 | Gab1 | 8 | 80764438 | 80880519 | - | 5082 | protein_co | growth fact | - | 1296 | 1380 | 1546 | 1710 | 1760 | 1755 | 13.41234 | 14.43767 | 14.96583 | 18.50167 | 18.53015 | 17.6772 |
| ENSMUSG0 | 4169.983 | 4356.557 | 4296.663 | 4826.321 | 5078.588 | 5257.206 | -0.24186 | 1.84E-06 | 6.79E-05 | Ppp2ca | 11 | 52098681 | 52127778 | + | 7490 | protein_co | protein ph | - | 4185 | 4325 | 4610 | 4633 | 5010 | 5421 | 29.38651 | 30.70132 | 30.27924 | 34.01183 | 35.78959 | 37.04833 |
| ENSMUSG0 | 188.3218 | 195.4155 | 207.8429 | 306.2677 | 331.4767 | 386.9443 | -0.79277 | 1.86E-06 | 6.83E-05 | Trib3 | 2 | 1.52E+08 | 1.52E+08 | - | 2125 | protein_co | tribbles pse | - | 189 | 194 | 223 | 294 | 327 | 399 | 4.677752 | 4.853953 | 5.16264 | 7.607428 | 8.233597 | 9.611364 |
| ENSMUSG0 | 5642.679 | 5587.473 | 5309.781 | 4574.223 | 4903.22 | 4319.423 | 0.261631 | 1.92E-06 | 7.07E-05 | Prelp | 1 | 1.34E+08 | 1.34E+08 | - | 3615 | protein_co | proline arg | - | 5663 | 5547 | 5697 | 4391 | 4837 | 4454 | 82.38964 | 81.58356 | 77.52893 | 66.78895 | 71.59268 | 63.06857 |
| ENSMUSG0 | 3789.354 | 3984.864 | 3844.628 | 4404.422 | 5488.119 | 4695.7 | -0.3284 | 1.94E-06 | 7.10E-05 | Xbp1 | 11 | 5520659 | 5525893 | + | 3248 | protein_co | X-box bind | TF_bZIP | 3803 | 3956 | 4125 | 4228 | 5414 | 4842 | 61.5807 | 64.75795 | 62.47897 | 71.57616 | 89.18731 | 76.30972 |
| ENSMUSG0 | 3092.862 | 2897.992 | 3218.303 | 3612.709 | 4176.404 | 3667.728 | -0.31488 | 1.97E-06 | 7.19E-05 | Il6st | 13 | 1.12E+08 | 1.13E+08 | + | 11099 | protein_co | interleukin | - | 3104 | 2877 | 3453 | 3468 | 4120 | 3782 | 14.70863 | 13.78189 | 15.30519 | 17.18085 | 19.8616 | 17.4425 |
| ENSMUSG0 | 233.1603 | 283.0503 | 260.9687 | 105.2144 | 100.3553 | 166.8031 | 1.05949 | 1.98E-06 | 7.22E-05 | Car3 | 3 | 14863512 | 14872523 | + | 2865 | protein_co | carbonic ar | - | 234 | 281 | 280 | 101 | 99 | 172 | 4.295617 | 5.214762 | 4.807943 | 1.938413 | 1.848891 | 3.073087 |
| ENSMUSG0 | 737.3446 | 704.1002 | 790.3623 | 1069.854 | 969.0879 | 1011.486 | -0.45022 | 1.99E-06 | 7.23E-05 | Smcr8 | 11 | 60777524 | 60788287 | + | 8078 | protein_co | Smith-Mag | - | 740 | 699 | 848 | 1027 | 956 | 1043 | 4.817949 | 4.600723 | 5.164377 | 6.990626 | 6.332203 | 6.60924 |
| ENSMUSG0 | 182.3433 | 143.0361 | 148.1929 | 279.1828 | 272.6827 | 311.3011 | -0.86674 | 2.00E-06 | 7.24E-05 | 6430548M | 8 | 1.2E+08 | 1.2E+08 | + | 6626 | protein_co | RIKEN cDN | - | 183 | 142 | 159 | 268 | 269 | 321 | 1.45256 | 1.139435 | 1.180515 | 2.22399 | 2.172209 | 2.479846 |
| ENSMUSG0 | 2700.276 | 2762.007 | 2741.103 | 3185.601 | 3540.82 | 3238.113 | -0.28058 | 2.16E-06 | 7.81E-05 | Gorasp2 | 2 | 70661576 | 70712636 | + | 4145 | protein_co | golgi reasse | - | 2710 | 2742 | 2941 | 3058 | 3493 | 3339 | 34.3858 | 35.17189 | 34.9057 | 40.56602 | 45.08944 | 41.23471 |
| ENSMUSG0 | 4517.731 | 4383.754 | 4235.149 | 5141.965 | 5442.503 | 4986.636 | -0.24532 | 2.17E-06 | 7.82E-05 | Sec24d | 3 | 1.23E+08 | 1.23E+08 | + | 6325 | protein_co | Sec24 relat | - | 4534 | 4352 | 4544 | 4936 | 5369 | 5142 | 37.70121 | 36.58315 | 35.34302 | 42.91055 | 45.41859 | 41.6143 |
| ENSMUSG0 | 359.7046 | 418.028 | 355.1038 | 241.6807 | 211.8613 | 226.9297 | 0.735153 | 2.22E-06 | 7.99E-05 | Car8 | 4 | 8143362 | 8239041 | - | 3853 | protein_co | carbonic ar | - | 361 | 415 | 381 | 232 | 209 | 234 | 4.92768 | 5.726666 | 4.864653 | 3.310842 | 2.902339 | 3.108765 |
| ENSMUSG0 | 511.1592 | 523.7941 | 540.578 | 307.3095 | 332.4904 | 381.1256 | 0.62525 | 2.35E-06 | 8.42E-05 | Arsi | 18 | 60911780 | 60918561 | + | 2975 | protein_co | arylsulfatas | - | 513 | 520 | 580 | 295 | 328 | 393 | 9.069111 | 9.293283 | 9.591067 | 5.45236 | 5.899126 | 6.762023 |
| ENSMUSG0 | 1325.228 | 1562.317 | 1331.872 | 1105.272 | 989.3617 | 1044.459 | 0.426547 | 2.50E-06 | 8.92E-05 | Hsph1 | 5 | 1.5E+08 | 1.5E+08 | - | 7266 | protein_co | heat shock | - | 1330 | 1551 | 1429 | 1061 | 976 | 1077 | 9.626991 | 11.3493 | 9.675262 | 8.029148 | 7.187126 | 7.587372 |
| ENSMUSG0 | 1514.546 | 1499.864 | 1622.666 | 1860.524 | 1968.587 | 2011.335 | -0.33275 | 2.52E-06 | 8.96E-05 | Btg2 | 1 | 1.34E+08 | 1.34E+08 | - | 2743 | protein_co | B cell trans | - | 1520 | 1489 | 1741 | 1786 | 1942 | 2074 | 29.1442 | 28.86169 | 31.22474 | 35.80182 | 37.88124 | 38.70384 |
| ENSMUSG0 | 1396.969 | 1486.769 | 1323.484 | 1046.936 | 1135.333 | 1005.667 | 0.400122 | 2.53E-06 | 9.00E-05 | Agtr2 | X | 21484544 | 21489164 | + | 3326 | protein_co | angiotensir | - | 1402 | 1476 | 1420 | 1005 | 1120 | 1037 | 22.16971 | 23.59483 | 21.00352 | 16.61473 | 18.01759 | 15.95981 |
| ENSMUSG0 | 967.5157 | 1113.062 | 963.7201 | 1355.287 | 1355.304 | 1315.999 | -0.40375 | 2.62E-06 | 9.28E-05 | Pcnx | 12 | 81860023 | 82000924 | + | 15613 | protein_co | pecanex ho | - | 971 | 1105 | 1034 | 1301 | 1337 | 1357 | 3.2709 | 3.762952 | 3.258068 | 4.581845 | 4.581903 | 4.449022 |
| ENSMUSG0 | 2723.193 | 2877.846 | 2914.461 | 3283.524 | 3418.164 | 3608.571 | -0.27601 | 2.67E-06 | 9.44E-05 | Ndel1 | 11 | 68821434 | 68871858 | - | 3253 | protein_co | nudE neurc | - | 2733 | 2857 | 3127 | 3152 | 3372 | 3721 | 44.18653 | 46.69592 | 47.29004 | 53.27845 | 55.46312 | 58.55267 |
| ENSMUSG0 | 9396.162 | 9324.543 | 9317.514 | 10298.51 | 11229.66 | 10640.48 | -0.19829 | 2.80E-06 | 9.85E-05 | Clic4 | 4 | 1.35E+08 | 1.35E+08 | - | 6572 | protein_co | chloride in | - | 9430 | 9257 | 9997 | 9886 | 11078 | 10972 | 75.4655 | 74.89029 | 74.83384 | 82.71276 | 90.19129 | 85.4593 |
| ENSMUSG0 | 12267.82 | 12902.46 | 13088.51 | 14391.46 | 14296.07 | 14687.4 | -0.18104 | 2.89E-06 | 0.000102 | Nr4a1 | 15 | 1.01E+08 | 1.01E+08 | + | 3509 | protein_co | nuclear rec | NGFIB-like | 12312 | 12809 | 14043 | 13815 | 14103 | 15145 | 184.5354 | 194.0817 | 196.8804 | 216.4796 | 215.0448 | 220.9312 |
| ENSMUSG0 | 1609.205 | 1643.908 | 1708.413 | 2078.245 | 2027.381 | 2032.67 | -0.3068 | 2.93E-06 | 0.000103 | Zmiz1 | 14 | 25459185 | 25666743 | + | 12523 | protein_co | zinc finger, | zf-MIZ | 1615 | 1632 | 1833 | 1995 | 2000 | 2096 | 6.782636 | 6.928904 | 7.200787 | 8.759594 | 8.545204 | 8.567497 |
| ENSMUSG0 | 20497.18 | 19658.4 | 19962.24 | 17591.64 | 18364.01 | 17401.83 | 0.172119 | 2.94E-06 | 0.000103 | C1qtnf3 | 15 | 10952332 | 10980150 | + | 2557 | protein_co | C1q and tu | - | 20571 | 19516 | 21418 | 16887 | 18116 | 17944 | 423.1156 | 405.8008 | 412.0729 | 363.1376 | 379.0813 | 359.2193 |
| ENSMUSG0 | 2646.469 | 2501.117 | 3071.974 | 3231.437 | 3658.408 | 3609.541 | -0.35292 | 3.03E-06 | 0.000105 | Ubr4 | 4 | 1.39E+08 | 1.39E+08 | + | 17997 | protein_co | ubiquitin p | - | 2656 | 2483 | 3296 | 3102 | 3609 | 3722 | 7.761797 | 7.335495 | 9.009755 | 9.477442 | 10.7297 | 10.58638 |
| ENSMUSG0 | 1310.281 | 1282.288 | 1498.706 | 1757.394 | 1871.272 | 1695.185 | -0.37931 | 3.10E-06 | 0.000108 | Klf9 | 19 | 23141226 | 23166911 | + | 3263 | protein_co | Kruppel-lik | zf-C2H2 | 1315 | 1273 | 1608 | 1687 | 1846 | 1748 | 21.19547 | 20.74264 | 24.24347 | 28.42808 | 30.27021 | 27.42177 |
| ENSMUSG0 | 1090.074 | 1068.741 | 1028.03 | 1342.786 | 1359.359 | 1400.37 | -0.36472 | 3.10E-06 | 0.000108 | Slc39a6 | 18 | 24579881 | 24603817 | - | 4073 | protein_co | solute carri | - | 1094 | 1061 | 1103 | 1289 | 1341 | 1444 | 14.12659 | 13.85013 | 13.32254 | 17.40155 | 17.61632 | 18.14779 |
| ENSMUSG0 | 681.5456 | 748.4212 | 740.0326 | 975.0565 | 987.3343 | 964.9363 | -0.43173 | 3.12E-06 | 0.000108 | Trib2 | 12 | 15791727 | 15816922 | - | 4335 | protein_co | tribbles pse | - | 684 | 743 | 794 | 936 | 974 | 995 | 8.298533 | 9.112814 | 9.010674 | 11.87234 | 12.02183 | 11.74911 |
| ENSMUSG0 | 452.3709 | 396.8748 | 437.1226 | 612.5355 | 612.2689 | 652.6654 | -0.54546 | 3.15E-06 | 0.000109 | Gfod1 | 13 | 43195245 | 43304172 | - | 7208 | protein_co | glucose-fru | - | 454 | 394 | 469 | 588 | 604 | 673 | 3.312649 | 2.906259 | 3.200987 | 4.485512 | 4.48356 | 4.779378 |
| ENSMUSG0 | 2067.554 | 1973.294 | 1781.111 | 2482.436 | 2603.157 | 2302.27 | -0.34384 | 3.21E-06 | 0.000111 | Jak2 | 19 | 29251828 | 29313080 | + | 5030 | protein_co | Janus kinas | - | 2075 | 1959 | 1911 | 2383 | 2568 | 2374 | 21.69624 | 20.7071 | 18.6904 | 26.04987 | 27.31668 | 24.15927 |
| ENSMUSG0 | 1310.281 | 1235.953 | 1187.408 | 932.3457 | 995.4439 | 897.0513 | 0.402368 | 3.29E-06 | 0.000113 | Sik1 | 17 | 31844248 | 31855792 | - | 4511 | protein_co | salt inducit | - | 1315 | 1227 | 1274 | 895 | 982 | 925 | 15.33159 | 14.46187 | 13.89385 | 10.90937 | 11.64768 | 10.49639 |
| ENSMUSG0 | 1423.872 | 1374.96 | 1440.92 | 1124.023 | 1130.265 | 1011.486 | 0.377037 | 3.32E-06 | 0.000114 | Fbln7 | 2 | 1.29E+08 | 1.29E+08 | + | 3228 | protein_co | fibulin 7 [S | - | 1429 | 1365 | 1546 | 1079 | 1115 | 1043 | 23.28268 | 22.48288 | 23.56144 | 18.37966 | 18.48171 | 16.53948 |
| ENSMUSG0 | 307.8912 | 317.2984 | 324.3468 | 145.8418 | 114.547 | 218.2017 | 0.98687 | 3.50E-06 | 0.00012 | Olfml2a | 2 | 38931978 | 38963753 | + | 5579 | protein_co | olfactomec | - | 309 | 315 | 348 | 140 | 113 | 225 | 2.912973 | 3.001974 | 3.06866 | 1.379816 | 1.083734 | 2.064416 |
| ENSMUSG0 | 1035.272 | 1004.274 | 959.0599 | 1236.53 | 1415.112 | 1298.542 | -0.39794 | 3.52E-06 | 0.00012 | Phtf2 | 5 | 20758663 | 20882124 | - | 6929 | protein_co | putative hc | - | 1039 | 997 | 1029 | 1187 | 1396 | 1339 | 7.886409 | 7.650281 | 7.305849 | 9.419538 | 10.77992 | 9.891931 |
| ENSMUSG0 | 344.7584 | 352.5537 | 327.1429 | 206.262 | 168.2726 | 224.0204 | 0.774566 | 3.70E-06 | 0.000125 | Nrp1 | 8 | 1.28E+08 | 1.29E+08 | + | 9785 | protein_co | neuropilin | - | 346 | 350 | 351 | 198 | 166 | 231 | 1.859729 | 1.901779 | 1.764705 | 1.112638 | 0.907712 | 1.208432 |
| ENSMUSG0 | 2456.155 | 2564.577 | 2450.31 | 2080.329 | 2023.326 | 2053.035 | 0.279084 | 3.70E-06 | 0.000125 | Pxdn | 12 | 29937608 | 30017658 | + | 9648 | protein_co | peroxidasir | - | 2465 | 2546 | 2629 | 1997 | 1996 | 2117 | 13.43736 | 14.03052 | 13.40538 | 11.38126 | 11.0694 | 11.23194 |
| ENSMUSG0 | 1072.139 | 1128.172 | 1161.311 | 1437.583 | 1477.96 | 1399.4 | -0.35987 | 3.81E-06 | 0.000129 | Yars | 4 | 1.29E+08 | 1.29E+08 | + | 5943 | protein_co | tyrosyl-tRN | - | 1076 | 1120 | 1246 | 1380 | 1458 | 1443 | 9.522278 | 10.01994 | 10.31426 | 12.768 | 13.12661 | 12.42887 |
| ENSMUSG0 | 1617.176 | 1653.981 | 1629.19 | 1955.322 | 2190.585 | 1989.029 | -0.32419 | 3.92E-06 | 0.000132 | Tram2 | 1 | 20996299 | 21079229 | - | 6661 | protein_co | translocati | - | 1623 | 1642 | 1748 | 1877 | 2161 | 2051 | 12.81485 | 13.10649 | 12.91005 | 15.49438 | 17.35866 | 15.76149 |
| ENSMUSG0 | 2381.424 | 2597.817 | 2509.028 | 3003.299 | 3206.302 | 2920.993 | -0.286 | 4.01E-06 | 0.000135 | Sec23a | 12 | 58958383 | 59012017 | - | 4840 | protein_co | SEC23 hom | - | 2390 | 2579 | 2692 | 2883 | 3163 | 3012 | 25.97089 | 28.3308 | 27.36249 | 32.75282 | 34.9667 | 31.85523 |
| ENSMUSG0 | 826.0253 | 854.1873 | 647.7616 | 1060.478 | 1098.84 | 1122.041 | -0.49618 | 4.02E-06 | 0.000135 | Asns | 6 | 7675169 | 7693254 | - | 2735 | protein_co | asparagine | - | 829 | 848 | 695 | 1018 | 1084 | 1157 | 15.94159 | 16.48509 | 12.50125 | 20.46633 | 21.20668 | 21.65445 |
| ENSMUSG0 | 710.4415 | 819.9392 | 820.1873 | 934.4292 | 1183.99 | 1283.026 | -0.53346 | 4.11E-06 | 0.000137 | Nab1 | 1 | 52457294 | 52500679 | - | 4100 | protein_co | Ngfi-A bind | - | 713 | 814 | 880 | 897 | 1168 | 1323 | 9.146186 | 10.55585 | 10.55905 | 12.02979 | 15.24263 | 16.5176 |
| ENSMUSG0 | 1280.389 | 1228.902 | 1267.562 | 1510.504 | 1801.328 | 1600.146 | -0.37914 | 4.13E-06 | 0.000138 | Arrdc3 | 13 | 80883384 | 80896042 | + | 6266 | protein_co | arrestin do | - | 1285 | 1220 | 1360 | 1450 | 1777 | 1650 | 10.78567 | 10.35195 | 10.67762 | 12.7241 | 15.17392 | 13.47922 |
| ENSMUSG0 | 1809.484 | 1780.9 | 1944.217 | 2343.886 | 2181.461 | 2334.273 | -0.30936 | 4.21E-06 | 0.00014 | Armcx4 | X | 1.35E+08 | 1.35E+08 | + | 7996 | protein_co | armadillo r | - | 1816 | 1768 | 2086 | 2250 | 2152 | 2407 | 11.94476 | 11.75607 | 12.83416 | 15.47245 | 14.40026 | 15.409 |
| ENSMUSG0 | 1736.746 | 1795.002 | 1631.986 | 2231.379 | 2297.022 | 2003.576 | -0.33913 | 4.23E-06 | 0.00014 | Npc1 | 18 | 12189693 | 12236400 | - | 10725 | protein_co | NPC intrace | - | 1743 | 1782 | 1751 | 2142 | 2266 | 2066 | 8.547408 | 8.834118 | 8.031834 | 10.98175 | 11.30481 | 9.860617 |
| ENSMUSG0 | 4263.646 | 4053.361 | 4313.44 | 4846.114 | 5368.504 | 4847.956 | -0.25394 | 4.31E-06 | 0.000143 | Neo1 | 9 | 58874687 | 59036441 | - | 8716 | protein_co | neogenin [S | - | 4279 | 4024 | 4628 | 4652 | 5296 | 4999 | 25.82019 | 24.54672 | 26.12174 | 29.34756 | 32.51109 | 29.35871 |
| ENSMUSG0 | 137.5048 | 164.1893 | 132.3484 | 61.4619 | 64.87618 | 64.97561 | 1.181093 | 4.39E-06 | 0.000145 | Filip1l | 16 | 57353093 | 57573126 | + | 5335 | protein_co | filamin A in | - | 138 | 163 | 142 | 59 | 64 | 67 | 1.360439 | 1.624449 | 1.309423 | 0.608089 | 0.641869 | 0.642853 |
| ENSMUSG0 | 555.9977 | 446.2323 | 452.9671 | 252.0979 | 331.4767 | 311.3011 | 0.70067 | 4.41E-06 | 0.000145 | Notch3 | 17 | 32120820 | 32166880 | - | 8044 | protein_co | notch 3 [So | - | 558 | 443 | 486 | 242 | 327 | 321 | 3.64835 | 2.92809 | 2.972283 | 1.654218 | 2.175086 | 2.042697 |
| ENSMUSG0 | 14288.54 | 14502.05 | 14540.62 | 12662.19 | 12917.46 | 13118.28 | 0.163113 | 4.42E-06 | 0.000145 | Maged1 | X | 94535474 | 94542143 | - | 4351 | protein_co | melanoma | - | 14340 | 14397 | 15601 | 12155 | 12743 | 13527 | 173.3382 | 175.9283 | 176.3962 | 153.6085 | 156.7052 | 159.1415 |
| ENSMUSG0 | 2280.786 | 2453.774 | 2363.631 | 2881.417 | 2765.347 | 2939.419 | -0.27467 | 4.44E-06 | 0.000145 | Adgra3 | 5 | 49959956 | 50059006 | - | 8113 | protein_co | adhesion G | - | 2289 | 2436 | 2536 | 2766 | 2728 | 3031 | 14.8388 | 15.96425 | 15.37778 | 18.7465 | 17.99135 | 19.12386 |
| ENSMUSG0 | 1908.128 | 1803.061 | 1909.732 | 2441.808 | 2167.27 | 2384.702 | -0.31512 | 4.52E-06 | 0.000148 | Prrc2b | 2 | 32151082 | 32234537 | + | 11262 | protein_co | proline-ricl | - | 1915 | 1790 | 2049 | 2344 | 2138 | 2459 | 8.94309 | 8.450654 | 8.950604 | 11.44436 | 10.15764 | 11.17671 |
| ENSMUSG0 | 4090.27 | 4178.265 | 4059.927 | 4897.159 | 5170.834 | 4582.235 | -0.24885 | 4.57E-06 | 0.000149 | Ski | 4 | 1.55E+08 | 1.55E+08 | - | 5562 | protein_co | ski sarcoma | - | 4105 | 4148 | 4356 | 4701 | 5101 | 4725 | 38.81651 | 39.65159 | 38.52856 | 46.47386 | 49.07103 | 43.48524 |
| ENSMUSG0 | 386.6077 | 390.831 | 384.9288 | 548.9901 | 565.6392 | 577.992 | -0.54241 | 4.62E-06 | 0.00015 | Dusp5 | 19 | 53529109 | 53542431 | + | 2473 | protein_co | dual specifi | - | 388 | 388 | 413 | 527 | 558 | 596 | 8.251672 | 8.341812 | 8.215837 | 11.71753 | 12.07288 | 12.33654 |
| ENSMUSG0 | 98.64476 | 77.56182 | 100.6594 | 41.66908 | 19.26012 | 23.27484 | 1.720819 | 4.72E-06 | 0.000153 | Ushbp1 | 8 | 71384272 | 71395802 | - | 6461 | protein_co | USH1 prote | - | 99 | 77 | 108 | 40 | 19 | 24 | 0.805879 | 0.633642 | 0.822337 | 0.340416 | 0.157346 | 0.190144 |
| ENSMUSG0 | 6667.987 | 6311.719 | 6491.596 | 5849.297 | 5324.915 | 5484.135 | 0.225217 | 4.72E-06 | 0.000153 | Hnrnpa2b1 | 6 | 51460932 | 51469894 | - | 6584 | protein_co | heterogene | - | 6692 | 6266 | 6965 | 5615 | 5253 | 5655 | 53.45649 | 50.60033 | 52.04239 | 46.89315 | 42.68924 | 43.96569 |
| ENSMUSG0 | 2354.521 | 2177.775 | 2128.759 | 2616.818 | 2947.811 | 2693.094 | -0.31018 | 4.77E-06 | 0.000154 | Tiparp | 3 | 65528410 | 65555518 | + | 4358 | protein_co | TCDD-indu | - | 2363 | 2162 | 2284 | 2512 | 2908 | 2777 | 28.51746 | 26.37675 | 25.78308 | 31.69435 | 35.70327 | 32.61818 |
| ENSMUSG0 | 228.1783 | 230.6709 | 173.3578 | 361.4793 | 450.0785 | 314.2104 | -0.83324 | 4.86E-06 | 0.000157 | Pi15 | 1 | 17601901 | 17630939 | + | 6923 | protein_co | peptidase i | - | 229 | 229 | 186 | 347 | 444 | 324 | 1.739705 | 1.758709 | 1.321735 | 2.756034 | 3.431543 | 2.395641 |
| ENSMUSG0 | 6387.995 | 6680.39 | 6941.767 | 5849.297 | 5462.777 | 5816.772 | 0.224406 | 5.11E-06 | 0.000164 | Islr | 9 | 58156268 | 58204306 | - | 2686 | protein_co | immunoglc | - | 6411 | 6632 | 7448 | 5615 | 5389 | 5998 | 125.5319 | 131.2778 | 136.4142 | 114.9458 | 107.3502 | 114.3067 |
| ENSMUSG0 | 2531.882 | 2466.869 | 2528.6 | 2910.585 | 3114.057 | 2992.757 | -0.26056 | 5.14E-06 | 0.000165 | Fosl2 | 5 | 32135801 | 32157842 | + | 9680 | protein_co | fos-like ant | TF_bZIP | 2541 | 2449 | 2713 | 2794 | 3072 | 3086 | 13.80587 | 13.45136 | 13.78797 | 15.87086 | 16.98035 | 16.31893 |
| ENSMUSG0 | 516.1413 | 546.9619 | 449.239 | 770.878 | 671.063 | 746.7346 | -0.53398 | 5.37E-06 | 0.000172 | Kcnk6 | 7 | 29221926 | 29232515 | - | 4151 | protein_co | potassium i | - | 518 | 543 | 482 | 740 | 662 | 770 | 6.563135 | 6.955044 | 5.712421 | 9.80231 | 8.533085 | 9.495308 |
| ENSMUSG0 | 20847.92 | 20907.44 | 20845.81 | 17286.42 | 19489.21 | 17720.89 | 0.200019 | 5.44E-06 | 0.000174 | Matn3 | 12 | 8947929 | 8972028 | + | 3014 | protein_co | matrilin 3 [ | - | 20923 | 20756 | 22366 | 16594 | 19226 | 18273 | 365.1027 | 366.1451 | 365.0657 | 302.7313 | 341.308 | 310.34 |
| ENSMUSG0 | 1912.114 | 1844.36 | 2038.352 | 1533.422 | 1398.893 | 1616.632 | 0.349284 | 5.48E-06 | 0.000174 | Cytl1 | 5 | 37735519 | 37739820 | + | 1056 | protein_co | cytokine-lil | - | 1919 | 1831 | 2187 | 1472 | 1380 | 1667 | 95.57524 | 92.1886 | 101.8851 | 76.64668 | 69.92234 | 80.80584 |
| ENSMUSG0 | 2468.112 | 2405.424 | 2408.368 | 3096.013 | 2896.113 | 2809.468 | -0.27326 | 5.56E-06 | 0.000177 | Rab2a | 4 | 8535644 | 8607778 | + | 2133 | protein_co | RAB2A, me | - | 2477 | 2388 | 2584 | 2972 | 2857 | 2897 | 61.07585 | 59.52457 | 59.59743 | 76.61387 | 71.66716 | 69.52303 |
| ENSMUSG0 | 399.5611 | 301.1816 | 339.2593 | 194.803 | 207.8065 | 211.4132 | 0.759918 | 6.03E-06 | 0.000191 | Vipr2 | 12 | 1.16E+08 | 1.16E+08 | + | 4463 | protein_co | vasoactive i | - | 401 | 299 | 364 | 187 | 205 | 218 | 4.725544 | 3.562026 | 4.012365 | 2.303903 | 2.457694 | 2.500349 |
| ENSMUSG0 | 2622.556 | 2575.657 | 2587.318 | 3084.554 | 2991.4 | 3220.657 | -0.25604 | 6.46E-06 | 0.000204 | Ankrd11 | 8 | 1.23E+08 | 1.23E+08 | - | 16580 | protein_co | ankyrin rep | - | 2632 | 2557 | 2776 | 2961 | 2951 | 3321 | 8.349024 | 8.19972 | 8.236844 | 9.819816 | 9.523257 | 10.25311 |
| ENSMUSG0 | 1554.402 | 1504.901 | 1303.911 | 1138.608 | 1056.265 | 1121.072 | 0.395557 | 6.56E-06 | 0.000207 | Srsf7 | 17 | 80200089 | 80207305 | - | 2279 | protein_co | serine/argi | - | 1560 | 1494 | 1399 | 1093 | 1042 | 1156 | 36.001 | 34.85452 | 30.19947 | 26.37092 | 24.46382 | 25.96478 |
| ENSMUSG0 | 469.3099 | 453.2834 | 406.3655 | 213.554 | 331.4767 | 233.7182 | 0.770721 | 6.66E-06 | 0.000209 | Adgrf5 | 17 | 43360451 | 43459557 | + | 9010 | protein_co | adhesion G | - | 471 | 450 | 436 | 205 | 327 | 241 | 2.749353 | 2.655465 | 2.380606 | 1.251061 | 1.941886 | 1.369189 |
| ENSMUSG0 | 1442.804 | 1454.536 | 1374.746 | 1779.27 | 1838.834 | 1714.58 | -0.32003 | 6.72E-06 | 0.000211 | Gas6 | 8 | 13465374 | 13494490 | - | 2548 | protein_co | growth arre | - | 1448 | 1444 | 1475 | 1708 | 1814 | 1768 | 29.88846 | 30.13149 | 28.47859 | 36.85852 | 38.09243 | 35.51844 |
| ENSMUSG0 | 674.5707 | 767.5598 | 676.6545 | 874.009 | 1133.306 | 989.1809 | -0.50048 | 6.82E-06 | 0.000213 | Sfrp2 | 3 | 83766321 | 83774316 | + | 2153 | protein_co | secreted fri | - | 677 | 762 | 726 | 839 | 1118 | 1020 | 16.53785 | 18.81758 | 16.58893 | 21.4273 | 27.78424 | 24.25086 |

| gene_id | H1 | H2 | H3 | M1 | M2 | M3 | log2FoldCh | pvalue | padj | gene_name | gene_chr | gene_start | gene_end | gene_stran | gene_lengtl | gene_bioty | gene_descr | tf_family | H1_readco | H2_readco | H3_readco | M1_readco | M2_readco | M3_readco | H1_fpkm | H2_fpkm | H3_fpkm | M1_fpkm | M2_fpkm | M3_fpkm |
|---|---|---|---|---|---|---|---|---|---|---|---|---|---|---|---|---|---|---|---|---|---|---|---|---|---|---|---|---|---|---|
| ENSMUSG0 | 234.1567 | 269.9554 | 241.396 | 363.5627 | 446.0237 | 390.8234 | -0.68765 | 6.83E-06 | 0.000213 | Ado | 10 | 67544511 | 67548955 | - | 4445 | protein_co | 2-aminoeth | - | 235 | 268 | 259 | 349 | 440 | 403 | 2.780548 | 3.205648 | 2.866513 | 4.31721 | 5.296412 | 4.640923 |
| ENSMUSG0 | 3870.063 | 4191.36 | 3750.493 | 4487.76 | 5189.081 | 4682.123 | -0.2819 | 7.05E-06 | 0.000219 | Dlc1 | 8 | 36567751 | 36953143 | - | 14694 | protein_co | deleted in l | - | 3884 | 4161 | 4024 | 4308 | 5119 | 4828 | 13.90188 | 15.05603 | 13.47236 | 16.12075 | 18.64 | 16.81893 |
| ENSMUSG0 | 35400.51 | 34875.62 | 38636.41 | 30770.53 | 32341.79 | 32390.83 | 0.189577 | 7.44E-06 | 0.000231 | Hspg2 | 4 | 1.37E+08 | 1.38E+08 | + | 14307 | protein_co | perlecan (h | - | 35528 | 34623 | 41454 | 29538 | 31905 | 33400 | 130.604 | 128.6675 | 142.5423 | 113.5225 | 119.3194 | 119.5003 |
| ENSMUSG0 | 1228.576 | 1195.661 | 1343.057 | 1511.546 | 1669.548 | 1646.695 | -0.3576 | 7.74E-06 | 0.00024 | Csgalnact1 | 8 | 68356781 | 68735146 | - | 5093 | protein_co | chondroiti | - | 1233 | 1187 | 1441 | 1451 | 1647 | 1698 | 12.73279 | 12.39167 | 13.91926 | 15.66546 | 17.30297 | 17.06613 |
| ENSMUSG0 | 471.3027 | 454.2907 | 443.6468 | 302.1008 | 291.9428 | 313.2406 | 0.593404 | 8.11E-06 | 0.000251 | Sema3b | 9 | 1.08E+08 | 1.08E+08 | - | 4777 | protein_co | sema doma | - | 473 | 451 | 476 | 290 | 288 | 323 | 5.207631 | 5.019658 | 4.902049 | 3.338045 | 3.225805 | 3.461134 |
| ENSMUSG0 | 301.9127 | 291.1087 | 370.0163 | 183.344 | 208.8202 | 161.9541 | 0.798701 | 8.33E-06 | 0.000257 | Kcnq1ot1 | 7 | 1.43E+08 | 1.43E+08 | - | 93092 | antisense | KCNQ1 ove | - | 303 | 289 | 397 | 176 | 206 | 167 | 0.171185 | 0.165059 | 0.209799 | 0.103956 | 0.118401 | 0.091828 |
| ENSMUSG0 | 787.1652 | 800.8006 | 750.285 | 377.1052 | 379.1202 | 646.8467 | 0.735617 | 8.53E-06 | 0.000263 | Cxcl12 | 6 | 1.17E+08 | 1.17E+08 | + | 8176 | protein_co | chemokine | - | 790 | 795 | 805 | 362 | 374 | 667 | 5.081835 | 5.169863 | 4.843741 | 2.434541 | 2.44755 | 4.175957 |
| ENSMUSG0 | 522.1197 | 561.0641 | 626.3249 | 760.4607 | 812.9796 | 815.5894 | -0.48237 | 8.70E-06 | 0.000267 | Ralgds | 2 | 28513125 | 28553081 | + | 6159 | protein_co | ral guanine | - | 524 | 557 | 672 | 730 | 802 | 841 | 4.474612 | 4.808369 | 5.36766 | 6.517216 | 6.967307 | 6.989673 |
| ENSMUSG0 | 2681.344 | 2534.358 | 2361.767 | 2971.006 | 3042.085 | 3219.687 | -0.28548 | 8.72E-06 | 0.000267 | Azin1 | 15 | 38487427 | 38519266 | - | 6256 | protein_co | antizyme ir | - | 2691 | 2516 | 2534 | 2852 | 3001 | 3320 | 22.62306 | 21.3829 | 19.92672 | 25.06699 | 25.6667 | 27.16517 |
| ENSMUSG0 | 3350.932 | 3046.064 | 3228.555 | 3672.088 | 3867.228 | 3930.539 | -0.25297 | 8.76E-06 | 0.000268 | Pxylp1 | 9 | 96823336 | 96892669 | - | 6148 | protein_co | 2-phospho | - | 3363 | 3024 | 3464 | 3525 | 3815 | 4053 | 28.76917 | 26.15175 | 27.71851 | 31.52642 | 33.20179 | 33.74534 |
| ENSMUSG0 | 1387.005 | 1212.785 | 1173.427 | 1595.926 | 1913.847 | 1544.868 | -0.42194 | 9.01E-06 | 0.000275 | Iars | 13 | 49682100 | 49734267 | + | 5272 | protein_co | isoleucine- | - | 1392 | 1204 | 1259 | 1532 | 1888 | 1593 | 13.88667 | 12.14238 | 11.74833 | 15.97838 | 19.16141 | 15.46719 |
| ENSMUSG0 | 1463.729 | 1525.047 | 1433.464 | 1779.27 | 1800.314 | 1922.114 | -0.31551 | 9.05E-06 | 0.000275 | Hk1 | 10 | 62268855 | 62379908 | - | 8532 | protein_co | hexokinase | - | 1469 | 1514 | 1538 | 1708 | 1776 | 1982 | 9.055351 | 9.434694 | 8.868116 | 11.00744 | 11.13763 | 11.89115 |
| ENSMUSG0 | 556.9941 | 479.4731 | 479.064 | 361.4793 | 305.1208 | 329.727 | 0.605213 | 9.06E-06 | 0.000275 | Fabp4 | 3 | 10204088 | 10208576 | - | 4489 | protein_co | fatty acid b | - | 559 | 476 | 514 | 347 | 301 | 340 | 6.549325 | 5.637806 | 5.632996 | 4.250395 | 3.587713 | 3.877041 |
| ENSMUSG0 | 803.1078 | 814.9028 | 814.5951 | 1189.652 | 1053.224 | 1007.607 | -0.41772 | 9.15E-06 | 0.000277 | Ehd4 | 2 | 1.2E+08 | 1.2E+08 | - | 4007 | protein_co | EH-domain | - | 806 | 809 | 874 | 1142 | 1039 | 1039 | 10.57913 | 10.7345 | 10.73045 | 15.67098 | 13.87385 | 13.27294 |
| ENSMUSG0 | 421.4821 | 402.9185 | 527.5296 | 629.2031 | 653.8302 | 740.9159 | -0.5815 | 9.23E-06 | 0.000279 | Fbxo45 | 16 | 32230112 | 32247158 | - | 4179 | protein_co | F-box prote | - | 423 | 400 | 566 | 604 | 645 | 764 | 5.323562 | 5.089093 | 6.663002 | 7.947198 | 8.258252 | 9.358195 |
| ENSMUSG0 | 2911.515 | 3046.064 | 2948.946 | 3702.298 | 3703.011 | 3309.877 | -0.26654 | 9.34E-06 | 0.000281 | Bhlhe40 | 6 | 1.09E+08 | 1.09E+08 | + | 4705 | protein_co | basic helix- | bHLH | 2922 | 3024 | 3164 | 3554 | 3653 | 3413 | 32.66291 | 34.17236 | 33.08284 | 41.53433 | 41.54233 | 37.13195 |
| ENSMUSG0 | 1374.052 | 1236.96 | 1259.174 | 964.6392 | 963.0058 | 1048.338 | 0.378663 | 9.54E-06 | 0.000286 | Ehmt2 | 17 | 34898469 | 34914052 | + | 5489 | protein_co | euchromat | - | 1379 | 1228 | 1351 | 926 | 950 | 1081 | 13.21312 | 11.89482 | 12.10844 | 9.276138 | 9.260431 | 10.081 |
| ENSMUSG0 | 1427.858 | 1353.806 | 1391.522 | 1832.398 | 1671.575 | 1715.55 | -0.32259 | 9.54E-06 | 0.000286 | Tns1 | 1 | 73910231 | 74124449 | - | 16010 | protein_co | tensin 1 [Sc | - | 1433 | 1344 | 1493 | 1759 | 1649 | 1769 | 4.707488 | 4.463348 | 4.587693 | 6.041211 | 5.510996 | 5.655976 |
| ENSMUSG0 | 17.93541 | 39.28456 | 28.89296 | 116.6734 | 90.21844 | 70.79432 | -1.68868 | 9.55E-06 | 0.000286 | Ren1 | 1 | 1.33E+08 | 1.33E+08 | + | 1657 | protein_co | renin 1 stru | - | 18 | 39 | 31 | 112 | 89 | 73 | 0.571326 | 1.251396 | 0.920376 | 3.716593 | 2.873878 | 2.255129 |
| ENSMUSG0 | 2269.826 | 2422.548 | 2273.224 | 2638.695 | 2882.935 | 2984.999 | -0.2887 | 9.58E-06 | 0.000286 | Lnpep | 17 | 17521410 | 17625050 | - | 14144 | protein_co | leucyl/cysti | - | 2278 | 2405 | 2439 | 2533 | 2844 | 3078 | 8.470632 | 9.040566 | 8.483312 | 9.847192 | 10.75866 | 11.13954 |
| ENSMUSG0 | 2494.018 | 2424.562 | 2594.774 | 2999.132 | 3192.111 | 2883.171 | -0.27205 | 9.60E-06 | 0.000286 | Nr1d1 | 11 | 98767932 | 98775333 | - | 3077 | protein_co | nuclear rec | THR-like | 2503 | 2407 | 2784 | 2879 | 3149 | 2973 | 42.78265 | 41.59119 | 44.51103 | 51.44742 | 54.7578 | 49.45822 |
| ENSMUSG0 | 1753.685 | 1551.236 | 1618.938 | 1266.74 | 1350.235 | 1259.751 | 0.344977 | 9.65E-06 | 0.000287 | Tardbp | 4 | 1.49E+08 | 1.49E+08 | - | 7796 | protein_co | TAR DNA bi | - | 1760 | 1540 | 1737 | 1216 | 1332 | 1299 | 11.8734 | 10.50272 | 10.96109 | 8.576522 | 9.141831 | 8.529202 |
| ENSMUSG0 | 535.0731 | 539.9109 | 571.335 | 401.0649 | 365.9422 | 363.6695 | 0.542672 | 9.88E-06 | 0.000293 | Ighm | 12 | 1.13E+08 | 1.13E+08 | - | 1886 | IG_C_gene | immunoglo | - | 537 | 536 | 613 | 385 | 361 | 375 | 14.975 | 15.1104 | 15.98986 | 11.22454 | 10.24156 | 10.17796 |
| ENSMUSG0 | 3576.122 | 3525.537 | 3310.574 | 2999.132 | 2937.674 | 2592.236 | 0.287975 | 1.01E-05 | 0.000299 | Fndc1 | 17 | 7738569 | 7827302 | - | 9903 | protein_co | fibronectin | - | 3589 | 3500 | 3552 | 2879 | 2898 | 2673 | 19.0608 | 18.79118 | 17.64542 | 15.98543 | 15.65786 | 13.81667 |
| ENSMUSG0 | 488.2417 | 536.889 | 506.0929 | 320.8519 | 352.7642 | 359.7903 | 0.56675 | 1.04E-05 | 0.000309 | Egfl6 | X | 1.67E+08 | 1.67E+08 | - | 2704 | protein_co | EGF-like-do | - | 490 | 533 | 543 | 308 | 348 | 371 | 9.530677 | 10.48029 | 9.879138 | 6.26316 | 6.886101 | 7.023253 |
| ENSMUSG0 | 2032.68 | 2045.819 | 2075.633 | 2379.305 | 2605.184 | 2468.103 | -0.27618 | 1.06E-05 | 0.000312 | Enpp2 | 15 | 54838901 | 54952892 | - | 5779 | protein_co | ectonucleo | - | 2040 | 2031 | 2227 | 2284 | 2570 | 2545 | 18.56572 | 18.68573 | 18.95804 | 21.73166 | 23.79476 | 22.54271 |
| ENSMUSG0 | 59352.26 | 59865.64 | 63756.51 | 69335.27 | 67925.36 | 66825.96 | -0.1575 | 1.09E-05 | 0.000322 | Comp | 8 | 70373558 | 70382066 | + | 2967 | protein_co | cartilage ol | - | 59566 | 59432 | 68406 | 66558 | 67008 | 68908 | 1055.882 | 1065.015 | 1134.234 | 1233.48 | 1208.398 | 1188.839 |
| ENSMUSG0 | 786.1688 | 882.3916 | 740.0326 | 1022.976 | 1095.799 | 1109.434 | -0.42327 | 1.11E-05 | 0.000325 | Secisbp2l | 2 | 1.26E+08 | 1.26E+08 | - | 8222 | protein_co | SECIS bindi | - | 789 | 876 | 794 | 982 | 1081 | 1144 | 5.047007 | 5.664733 | 4.750824 | 6.567249 | 7.034756 | 7.12229 |
| ENSMUSG0 | 544.0408 | 590.2757 | 690.635 | 855.2579 | 871.7736 | 828.1966 | -0.48471 | 1.13E-05 | 0.000331 | 5031439G | 15 | 84943936 | 84988551 | - | 9604 | protein_co | RIKEN cDN | - | 546 | 586 | 741 | 821 | 860 | 854 | 2.990025 | 3.24413 | 3.795701 | 4.700461 | 4.791231 | 4.551733 |
| ENSMUSG0 | 2390.392 | 2437.657 | 2329.146 | 2857.457 | 3050.194 | 2734.794 | -0.27205 | 1.13E-05 | 0.000331 | Pum2 | 12 | 8674134 | 8752581 | + | 7260 | protein_co | pumilio RN | - | 2399 | 2420 | 2499 | 2743 | 3009 | 2820 | 17.37913 | 17.72277 | 16.93384 | 20.77489 | 22.17616 | 19.88308 |
| ENSMUSG0 | 700.4774 | 643.6624 | 731.6444 | 955.2637 | 981.2522 | 874.7463 | -0.43688 | 1.15E-05 | 0.000334 | Entpd7 | 19 | 43689672 | 43733697 | + | 8963 | protein_co | ectonucleo | - | 703 | 639 | 785 | 917 | 968 | 902 | 4.125117 | 3.790533 | 4.308659 | 5.625555 | 5.778602 | 5.151388 |
| ENSMUSG0 | 941.609 | 936.7856 | 979.5646 | 777.1284 | 651.8029 | 682.7288 | 0.437145 | 1.17E-05 | 0.00034 | Pfkm | 15 | 98092589 | 98132451 | + | 3340 | protein_co | phosphofru | - | 945 | 930 | 1051 | 746 | 643 | 704 | 14.88057 | 14.80435 | 15.4804 | 12.28123 | 10.30067 | 10.7894 |
| ENSMUSG0 | 98.64476 | 91.66397 | 110.9117 | 175.0101 | 211.8613 | 196.8664 | -0.95374 | 1.19E-05 | 0.000346 | D430019H | 12 | 1.05E+08 | 1.05E+08 | + | 5786 | protein_co | RIKEN cDN | - | 99 | 91 | 119 | 168 | 209 | 203 | 0.899893 | 0.836211 | 1.011799 | 1.596542 | 1.932719 | 1.795927 |
| ENSMUSG0 | 1784.573 | 1648.944 | 1662.743 | 2101.163 | 2072.997 | 2058.854 | -0.29053 | 1.24E-05 | 0.000358 | Pmepa1 | 2 | 1.73E+08 | 1.73E+08 | - | 4913 | protein_co | prostate tra | - | 1791 | 1637 | 1784 | 2017 | 2045 | 2123 | 19.17269 | 17.71555 | 17.86381 | 22.574 | 22.27139 | 22.11945 |
| ENSMUSG0 | 593.8614 | 655.7499 | 569.471 | 437.5254 | 428.791 | 396.6421 | 0.526408 | 1.24E-05 | 0.000359 | Ltbp1 | 17 | 75005568 | 75392512 | + | 10480 | protein_co | latent trans | - | 596 | 651 | 611 | 420 | 423 | 409 | 2.99102 | 3.302726 | 2.868176 | 2.203624 | 2.159633 | 1.997713 |
| ENSMUSG0 | 899.7597 | 1061.69 | 1027.098 | 1345.911 | 1227.579 | 1349.941 | -0.39261 | 1.36E-05 | 0.00039 | Polr3d | 14 | 70438757 | 70443447 | - | 2854 | protein_co | polymerase | - | 903 | 1054 | 1102 | 1292 | 1211 | 1392 | 16.64057 | 19.63539 | 18.99562 | 24.8919 | 22.7034 | 24.96642 |
| ENSMUSG0 | 616.7788 | 635.604 | 610.4803 | 445.8592 | 401.4214 | 462.5875 | 0.507769 | 1.36E-05 | 0.00039 | Cd36 | 5 | 17781690 | 17888801 | - | 3590 | protein_co | CD36 mole | - | 619 | 631 | 655 | 428 | 396 | 477 | 9.068397 | 9.345181 | 8.975791 | 6.555394 | 5.902032 | 6.801348 |
| ENSMUSG0 | 2319.646 | 2343.979 | 2332.874 | 1992.824 | 1812.478 | 1957.027 | 0.279996 | 1.40E-05 | 0.000401 | Lmcd1 | 6 | 1.12E+08 | 1.12E+08 | + | 1730 | protein_co | LIM and cys | - | 2328 | 2327 | 2503 | 1913 | 1788 | 2018 | 70.77359 | 71.51598 | 71.17716 | 60.80207 | 55.29963 | 59.70987 |
| ENSMUSG0 | 2618.57 | 2488.022 | 2639.512 | 2202.211 | 2113.544 | 2142.256 | 0.262584 | 1.41E-05 | 0.000403 | S100b | 10 | 76253853 | 76261159 | + | 1484 | protein_co | S100 prote | - | 2628 | 2470 | 2832 | 2114 | 2085 | 2209 | 93.13776 | 88.49441 | 93.88263 | 78.32863 | 75.17492 | 76.19613 |
| ENSMUSG0 | 11615.17 | 11129.62 | 10483.49 | 9985.995 | 9614.852 | 9142.165 | 0.209204 | 1.42E-05 | 0.000405 | Rian | 12 | 1.1E+08 | 1.1E+08 | + | 18234 | processed_ | RNA imprin | - | 11657 | 11049 | 11248 | 9586 | 9485 | 9427 | 33.62321 | 32.21765 | 30.34724 | 28.90713 | 27.83275 | 26.46444 |
| ENSMUSG0 | 1284.375 | 1122.128 | 1189.272 | 1708.432 | 1440.454 | 1528.381 | -0.37915 | 1.42E-05 | 0.000405 | Cgnl1 | 9 | 71626509 | 71771602 | - | 7844 | protein_co | cingulin-lik | - | 1289 | 1114 | 1276 | 1640 | 1421 | 1576 | 8.642705 | 7.550929 | 8.002745 | 11.49624 | 9.69298 | 10.28465 |
| ENSMUSG0 | 203.268 | 215.5614 | 235.8039 | 362.521 | 338.5726 | 344.2737 | -0.67447 | 1.44E-05 | 0.000409 | Nog | 11 | 89300638 | 89302332 | - | 1695 | protein_co | noggin [Sou | - | 204 | 214 | 253 | 348 | 334 | 355 | 6.32987 | 6.712694 | 7.343054 | 11.28909 | 10.54332 | 10.72086 |
| ENSMUSG0 | 927.6593 | 896.4938 | 975.8365 | 1146.941 | 1255.962 | 1219.99 | -0.37154 | 1.44E-05 | 0.000409 | Lrrc1 | 9 | 77430823 | 77544870 | - | 8195 | protein_co | leucine rich | - | 931 | 890 | 1047 | 1101 | 1239 | 1258 | 5.974961 | 5.774227 | 6.285265 | 7.387336 | 8.089528 | 7.857832 |
| ENSMUSG0 | 599.8398 | 598.334 | 538.7139 | 418.7743 | 412.5719 | 367.5486 | 0.534933 | 1.45E-05 | 0.00041 | Gm42047 | 8 | 1.22E+08 | 1.22E+08 | + | 5647 | lincRNA | predicted g | - | 602 | 594 | 578 | 402 | 407 | 379 | 5.606774 | 5.592699 | 5.035423 | 3.914333 | 3.856359 | 3.43552 |
| ENSMUSG0 | 1424.869 | 1425.324 | 1274.086 | 1120.898 | 1021.8 | 1059.975 | 0.364653 | 1.53E-05 | 0.000431 | Prdx6 | 1 | 1.61E+08 | 1.61E+08 | - | 4804 | protein_co | peroxiredo | - | 1430 | 1415 | 1367 | 1076 | 1008 | 1093 | 15.65552 | 15.66052 | 13.99882 | 12.31569 | 11.22686 | 11.64631 |
| ENSMUSG0 | 62.77394 | 50.36482 | 85.74685 | 148.967 | 134.8208 | 166.8031 | -1.17749 | 1.55E-05 | 0.000437 | Gpha2 | 19 | 6226380 | 6227768 | + | 770 | protein_co | glycoprotei | - | 63 | 50 | 92 | 143 | 133 | 172 | 4.303127 | 3.452487 | 5.877911 | 10.21162 | 9.24191 | 11.43428 |
| ENSMUSG0 | 1491.628 | 1466.624 | 1575.132 | 1809.48 | 1834.779 | 1972.543 | -0.30913 | 1.62E-05 | 0.000455 | Tnfrsf1a | 6 | 1.25E+08 | 1.25E+08 | + | 2591 | protein_co | tumor necr | - | 1497 | 1456 | 1690 | 1737 | 1810 | 2034 | 30.38706 | 29.87767 | 32.08819 | 36.86225 | 37.37765 | 40.18413 |
| ENSMUSG0 | 936.627 | 957.9389 | 928.3029 | 1183.402 | 1162.703 | 1293.693 | -0.36708 | 1.63E-05 | 0.000457 | Cdc73 | 1 | 1.44E+08 | 1.44E+08 | - | 17613 | protein_co | cell divisio | - | 940 | 951 | 996 | 1136 | 1147 | 1334 | 2.806912 | 2.87078 | 2.781967 | 3.546455 | 3.484423 | 3.87698 |
| ENSMUSG0 | 914.7059 | 952.9024 | 849.0803 | 1134.441 | 1230.62 | 1165.682 | -0.3786 | 1.64E-05 | 0.000459 | Tgfbr1 | 4 | 47353222 | 47414931 | + | 6534 | protein_co | transformi | - | 918 | 946 | 911 | 1089 | 1214 | 1202 | 7.389208 | 7.697768 | 6.859069 | 9.164277 | 9.941236 | 9.41665 |
| ENSMUSG0 | 590.8721 | 568.1152 | 554.5585 | 789.6291 | 749.1171 | 780.6771 | -0.43697 | 1.75E-05 | 0.000488 | Gmppb | 9 | 1.08E+08 | 1.08E+08 | + | 2785 | protein_co | GDP-mann | - | 593 | 564 | 595 | 758 | 739 | 805 | 11.1986 | 10.7673 | 10.51036 | 14.96558 | 14.19777 | 14.79591 |
| ENSMUSG0 | 566.9582 | 610.4216 | 466.9475 | 755.2521 | 808.9248 | 767.1001 | -0.50457 | 1.81E-05 | 0.000504 | Ctdp1 | 18 | 80407959 | 80469695 | - | 3910 | protein_co | CTD (carbo | - | 569 | 606 | 501 | 725 | 798 | 791 | 7.653672 | 8.240407 | 6.303574 | 10.19555 | 10.92011 | 10.35549 |
| ENSMUSG0 | 711.4379 | 712.1585 | 684.1108 | 511.488 | 507.8588 | 520.7747 | 0.452383 | 1.81E-05 | 0.000504 | Camk4 | 18 | 32939041 | 33195767 | + | 12331 | protein_co | calcium/ca | - | 714 | 707 | 734 | 491 | 501 | 537 | 3.045329 | 3.048414 | 2.928355 | 2.189438 | 2.173903 | 2.22919 |
| ENSMUSG0 | 21767.61 | 22473.79 | 22431.19 | 23696.16 | 25950.47 | 25403.52 | -0.17079 | 1.81E-05 | 0.000504 | Vim | 2 | 13573927 | 13582826 | + | 4743 | protein_co | vimentin [S | - | 21846 | 22311 | 24067 | 22747 | 25600 | 26195 | 242.2441 | 250.1029 | 249.6288 | 263.7063 | 288.7937 | 282.7069 |
| ENSMUSG0 | 80.70935 | 100.7296 | 61.51405 | 22.91799 | 34.46547 | 16.48635 | 1.718466 | 1.90E-05 | 0.000527 | Gm45153 | 7 | 1.17E+08 | 1.17E+08 | + | 1901 | TEC | predicted g | - | 81 | 100 | 66 | 22 | 34 | 17 | 2.240976 | 2.79686 | 1.707999 | 0.636341 | 0.956968 | 0.45776 |
| ENSMUSG0 | 188.3218 | 186.3498 | 164.0375 | 331.2692 | 270.6553 | 298.6938 | -0.74188 | 1.93E-05 | 0.000533 | Slc6a17 | 3 | 1.07E+08 | 1.08E+08 | - | 10283 | protein_co | solute carri | - | 189 | 185 | 176 | 318 | 267 | 308 | 0.966666 | 0.956543 | 0.842013 | 1.700422 | 1.389288 | 1.533211 |
| ENSMUSG0 | 646.6712 | 757.4869 | 693.4311 | 968.8061 | 975.1701 | 882.5045 | -0.43015 | 1.93E-05 | 0.000533 | Slc30a1 | 1 | 1.92E+08 | 1.92E+08 | + | 5244 | protein_co | solute carri | - | 649 | 752 | 744 | 930 | 962 | 910 | 6.509031 | 7.62444 | 6.97969 | 9.751462 | 9.815517 | 8.882798 |
| ENSMUSG0 | 802.1114 | 843.1071 | 812.7311 | 1124.023 | 1147.497 | 979.4831 | -0.40313 | 1.94E-05 | 0.000533 | Bag5 | 12 | 1.12E+08 | 1.12E+08 | - | 2508 | protein_co | BCL2-assoc | - | 805 | 837 | 872 | 1079 | 1132 | 1010 | 16.88118 | 17.74397 | 17.10468 | 23.65611 | 24.15014 | 20.61413 |
| ENSMUSG0 | 32867.64 | 31622.06 | 34698.58 | 36632.33 | 37246.02 | 37098.16 | -0.16197 | 1.98E-05 | 0.000544 | Col6a3 | 1 | 90765923 | 90843971 | - | 11963 | protein_co | collagen, ty | - | 32986 | 31393 | 37229 | 35165 | 36743 | 38254 | 145.0187 | 139.5229 | 153.0972 | 161.6293 | 164.337 | 163.6846 |
| ENSMUSG0 | 5583.891 | 5454.51 | 5694.71 | 6772.267 | 6321.373 | 6209.535 | -0.20592 | 2.00E-05 | 0.000548 | Eif4g1 | 16 | 20668313 | 20692884 | + | 7301 | protein_co | eukaryotic | - | 5604 | 5415 | 6110 | 6501 | 6236 | 6403 | 40.3692 | 39.43383 | 41.17037 | 48.96066 | 45.70088 | 44.89234 |
| ENSMUSG0 | 831.0073 | 840.0852 | 790.3623 | 970.8896 | 1145.47 | 1160.833 | -0.41356 | 2.00E-05 | 0.000548 | Ndrg1 | 15 | 66929318 | 66969640 | - | 6596 | protein_co | N-myc dow | - | 834 | 834 | 848 | 932 | 1130 | 1197 | 6.649971 | 6.722614 | 6.324717 | 7.769351 | 9.166396 | 9.289334 |
| ENSMUSG0 | 1396.969 | 1387.047 | 1556.492 | 1720.933 | 1849.985 | 1863.927 | -0.32413 | 2.01E-05 | 0.000549 | Tacc2 | 7 | 1.31E+08 | 1.31E+08 | + | 15408 | protein_co | transformi | - | 1402 | 1377 | 1670 | 1652 | 1825 | 1922 | 4.785597 | 4.751606 | 5.332073 | 5.895399 | 6.337492 | 6.385254 |
| ENSMUSG0 | 5721.396 | 5716.407 | 6255.792 | 6862.898 | 6774.492 | 6759.403 | -0.20489 | 2.04E-05 | 0.000557 | Fam129b | 2 | 32876114 | 32925254 | + | 4672 | protein_co | family with | - | 5742 | 5675 | 6712 | 6588 | 6683 | 6970 | 64.63901 | 64.58265 | 70.6765 | 77.53543 | 76.53665 | 76.36617 |
| ENSMUSG0 | 596.8506 | 699.0637 | 651.4897 | 372.9383 | 485.5576 | 467.4365 | 0.553911 | 2.05E-05 | 0.000557 | Angptl4 | 17 | 33773750 | 33781575 | - | 3286 | protein_co | angiopoieti | - | 599 | 694 | 699 | 358 | 479 | 482 | 9.58724 | 11.22909 | 10.46491 | 5.990526 | 7.799536 | 7.508455 |
| ENSMUSG0 | 1882.222 | 1990.418 | 1803.48 | 2350.136 | 2319.323 | 2226.627 | -0.28102 | 2.05E-05 | 0.000558 | Arl6ip5 | 6 | 97210689 | 97233315 | + | 1442 | protein_co | ADP-ribosy | - | 1889 | 1976 | 1935 | 2256 | 2288 | 2296 | 68.89711 | 72.85754 | 66.01485 | 86.02473 | 84.89685 | 81.50377 |
| ENSMUSG0 | 1348.145 | 1313.514 | 1184.611 | 1556.34 | 1775.985 | 1583.659 | -0.3544 | 2.11E-05 | 0.000569 | Tcp11l2 | 10 | 84576626 | 84614359 | + | 3438 | protein_co | t-complex 1 | - | 1353 | 1304 | 1271 | 1494 | 1752 | 1633 | 20.6979 | 20.16622 | 18.18719 | 23.89429 | 27.26648 | 24.31372 |
| ENSMUSG0 | 658.6281 | 705.1075 | 639.3733 | 839.632 | 926.5129 | 913.5377 | -0.42037 | 2.11E-05 | 0.000569 | Dhx8 | 11 | 1.02E+08 | 1.02E+08 | + | 5032 | protein_co | DEAH (Asp- | - | 661 | 700 | 686 | 806 | 914 | 942 | 6.908681 | 7.396227 | 6.706708 | 8.807322 | 9.71866 | 9.582556 |
| ENSMUSG0 | 1393.98 | 1315.529 | 1416.687 | 1758.435 | 1809.437 | 1604.025 | -0.32544 | 2.11E-05 | 0.000569 | Igf1r | 7 | 67952827 | 68233668 | + | 16791 | protein_co | insulin-like | - | 1399 | 1306 | 1520 | 1688 | 1785 | 1654 | 4.382031 | 4.135418 | 4.453413 | 5.527711 | 5.688038 | 5.042316 |
| ENSMUSG0 | 613.7896 | 532.8598 | 616.0725 | 872.9673 | 750.1308 | 813.6498 | -0.46663 | 2.12E-05 | 0.000571 | Kifc3 | 8 | 95099828 | 95202812 | - | 5298 | protein_co | kinesin fam | - | 616 | 529 | 661 | 838 | 740 | 839 | 6.115093 | 5.308802 | 6.137838 | 8.697241 | 7.47344 | 8.10627 |
| ENSMUSG0 | 2978.275 | 3051.101 | 3131.624 | 3667.921 | 3549.943 | 3504.804 | -0.22686 | 2.12E-05 | 0.000571 | Furin | 7 | 80388585 | 80405436 | - | 9178 | protein_co | furin (paire | - | 2989 | 3029 | 3360 | 3521 | 3502 | 3614 | 17.12822 | 17.54705 | 18.01014 | 21.09441 | 20.41592 | 20.15632 |
| ENSMUSG0 | 3469.505 | 3466.107 | 3526.805 | 3910.643 | 4212.897 | 4063.4 | -0.22013 | 2.19E-05 | 0.000588 | Kdelr2 | 5 | 1.43E+08 | 1.43E+08 | + | 1829 | protein_co | KDEL (Lys-A | - | 3482 | 3441 | 3784 | 3754 | 4156 | 4190 | 100.1266 | 100.0285 | 101.7802 | 112.8574 | 121.5801 | 117.2658 |
| ENSMUSG0 | 1411.915 | 1433.383 | 1371.018 | 1778.228 | 1721.246 | 1685.487 | -0.29837 | 2.20E-05 | 0.000588 | Nolc1 | 19 | 46075863 | 46085530 | + | 4483 | protein_co | nucleolar a | - | 1417 | 1423 | 1471 | 1707 | 1698 | 1738 | 16.624 | 16.87676 | 16.14246 | 20.93699 | 20.26608 | 19.84505 |
| ENSMUSG0 | 77.72011 | 71.51804 | 91.33904 | 131.2576 | 216.9297 | 167.7728 | -1.09992 | 2.21E-05 | 0.000589 | Fzd10 | 5 | 1.29E+08 | 1.29E+08 | + | 3250 | protein_co | frizzled clas | - | 78 | 71 | 98 | 126 | 214 | 173 | 1.262251 | 1.161523 | 1.483435 | 2.131752 | 3.523151 | 2.724795 |
| ENSMUSG0 | 555.9977 | 618.48 | 645.8975 | 807.3385 | 841.3629 | 817.5289 | -0.43769 | 2.22E-05 | 0.000592 | Cdt1 | 8 | 1.23E+08 | 1.23E+08 | + | 3140 | protein_co | chromatin | - | 558 | 614 | 693 | 775 | 830 | 843 | 9.346282 | 10.3966 | 10.85749 | 13.5713 | 14.14325 | 13.7426 |
| ENSMUSG0 | 682.542 | 636.6113 | 638.4413 | 911.5112 | 853.5272 | 844.6829 | -0.41472 | 2.23E-05 | 0.000592 | Nt5dc3 | 10 | 86779005 | 86838389 | + | 6004 | protein_co | 5'-nucleoti | - | 685 | 632 | 685 | 875 | 842 | 871 | 6.000456 | 5.596663 | 5.612751 | 8.0134 | 7.503644 | 7.425891 |
| ENSMUSG0 | 997.4081 | 1020.391 | 1211.64 | 1450.084 | 1302.592 | 1524.502 | -0.40493 | 2.24E-05 | 0.000595 | Cebpb | 2 | 1.68E+08 | 1.68E+08 | + | 1504 | protein_co | CCAAT/enh | C/EBP | 1001 | 1013 | 1300 | 1392 | 1285 | 1572 | 35.00423 | 35.81083 | 42.52276 | 50.89099 | 45.71472 | 53.50271 |
| ENSMUSG0 | 2233.955 | 2316.782 | 2261.107 | 1859.483 | 1966.559 | 1811.559 | 0.273038 | 2.25E-05 | 0.000595 | Tuba1b | 15 | 98931425 | 98934565 | - | 1875 | protein_co | tubulin, alp | - | 2242 | 2300 | 2426 | 1785 | 1940 | 1868 | 62.88813 | 65.21978 | 63.65249 | 52.34635 | 55.36066 | 50.99724 |
| ENSMUSG0 | 1079.114 | 986.1431 | 1058.787 | 1231.321 | 1461.741 | 1358.669 | -0.37517 | 2.30E-05 | 0.000608 | Slc35d1 | 4 | 1.03E+08 | 1.03E+08 | - | 8528 | protein_co | solute carri | - | 1083 | 979 | 1136 | 1182 | 1442 | 1401 | 6.679064 | 6.103631 | 6.553255 | 7.621137 | 9.047298 | 8.409342 |
| ENSMUSG0 | 221.2034 | 252.8314 | 226.4835 | 132.2993 | 114.547 | 145.4678 | 0.83562 | 2.36E-05 | 0.000622 | Anpep | 7 | 79821803 | 79861059 | - | 4135 | protein_co | alanyl (mer | - | 222 | 251 | 243 | 127 | 113 | 150 | 2.823656 | 3.227387 | 2.891057 | 1.688798 | 1.46219 | 1.856893 |
| ENSMUSG0 | 1166.798 | 1106.011 | 1205.116 | 1660.513 | 1370.509 | 1467.285 | -0.3707 | 2.37E-05 | 0.000624 | Tnks1bp1 | 2 | 85048022 | 85073048 | + | 6184 | protein_co | tankyrase 1 | - | 1171 | 1098 | 1293 | 1594 | 1352 | 1513 | 9.959138 | 9.440297 | 10.2862 | 14.17321 | 11.6979 | 12.52393 |
| ENSMUSG0 | 751.2944 | 801.8079 | 681.3147 | 1083.396 | 917.3897 | 1015.365 | -0.43323 | 2.38E-05 | 0.000625 | Arid5a | 1 | 36307733 | 36324029 | + | 12540 | protein_co | AT rich inte | ARID | 754 | 796 | 731 | 1040 | 905 | 1047 | 3.162337 | 3.374958 | 2.867779 | 4.560214 | 3.861463 | 4.273859 |
| ENSMUSG0 | 310.8804 | 303.1962 | 332.7351 | 464.6103 | 458.188 | 468.4063 | -0.55475 | 2.38E-05 | 0.000625 | Zfp212 | 6 | 47920476 | 47932639 | + | 3002 | protein_co | Zinc finger | zf-C2H2 | 312 | 301 | 357 | 446 | 452 | 483 | 5.466108 | 5.330999 | 5.850371 | 8.169089 | 8.056169 | 8.235833 |
| ENSMUSG0 | 2136.307 | 2218.067 | 2474.542 | 2779.328 | 2640.663 | 3034.458 | -0.3079 | 2.39E-05 | 0.000625 | Cxcl14 | 13 | 56288643 | 56296551 | - | 1827 | protein_co | chemokine | - | 2144 | 2202 | 2655 | 2668 | 2605 | 3129 | 61.71924 | 64.08134 | 71.49108 | 80.29652 | 76.29042 | 87.66739 |
| ENSMUSG0 | 507.1735 | 545.9546 | 466.9475 | 745.8766 | 633.5564 | 778.7375 | -0.50637 | 2.42E-05 | 0.000633 | Sap30bp | 11 | 1.16E+08 | 1.16E+08 | + | 3708 | protein_co | SAP30 binc | - | 509 | 542 | 501 | 716 | 625 | 803 | 7.219588 | 7.771634 | 6.646973 | 10.61751 | 9.018641 | 11.08529 |
| ENSMUSG0 | 472.2991 | 497.6044 | 480.928 | 614.619 | 702.4874 | 704.0641 | -0.47873 | 2.44E-05 | 0.000635 | Gosr1 | 11 | 76726602 | 76763579 | - | 4289 | protein_co | golgi SNAP | - | 474 | 494 | 516 | 590 | 693 | 726 | 5.812415 | 6.123838 | 5.918608 | 7.563894 | 8.645259 | 8.664663 |

| gene_id | H1 | H2 | H3 | M1 | M2 | M3 | log2FoldCh | pvalue | padj | gene_name | gene_chr | gene_start | gene_end | gene_stran | gene_lengt | gene_bioty | gene_descr | tf_family | H1_readco | H2_readco | H3_readco | M1_readco | M2_readco | M3_readco | H1_fpkm | H2_fpkm | H3_fpkm | M1_fpkm | M2_fpkm | M3_fpkm |
|---|---|---|---|---|---|---|---|---|---|---|---|---|---|---|---|---|---|---|---|---|---|---|---|---|---|---|---|---|---|---|
| ENSMUSG0 | 362.6939 | 325.3567 | 301.9781 | 222.9296 | 184.4916 | 200.7455 | 0.702955 | 2.44E-05 | 0.000635 | Sorcs2 | 5 | 36017180 | 36398139 | - | 8802 | protein_co | sortilin-rela | - | 364 | 323 | 324 | 214 | 182 | 207 | 2.174975 | 1.951075 | 1.810879 | 1.336848 | 1.106346 | 1.203816 |
| ENSMUSG0 | 1264.446 | 1293.369 | 1202.32 | 1526.13 | 1805.382 | 1505.107 | -0.36334 | 2.55E-05 | 0.000661 | Etnk1 | 6 | 1.43E+08 | 1.43E+08 | + | 14151 | protein_co | ethanolami | - | 1269 | 1284 | 1290 | 1465 | 1781 | 1552 | 4.71638 | 4.82426 | 4.484649 | 5.69246 | 6.73407 | 5.614042 |
| ENSMUSG0 | 467.3171 | 379.7507 | 416.6179 | 552.1153 | 630.5154 | 647.8165 | -0.5349 | 2.74E-05 | 0.00071 | Steap3 | 1 | 1.2E+08 | 1.2E+08 | - | 5715 | protein_co | STEAP famil | - | 469 | 377 | 447 | 530 | 622 | 668 | 4.316095 | 3.50734 | 3.847842 | 5.099283 | 5.823378 | 5.98317 |
| ENSMUSG0 | 1331.206 | 1350.784 | 1341.193 | 890.6766 | 1117.087 | 1064.824 | 0.388479 | 2.75E-05 | 0.000711 | Igf1 | 10 | 87858265 | 87937042 | + | 9904 | protein_co | insulin-like | - | 1336 | 1341 | 1439 | 855 | 1102 | 1098 | 7.094637 | 7.19898 | 7.14786 | 4.746844 | 5.953492 | 5.67496 |
| ENSMUSG0 | 682.542 | 701.0783 | 749.3529 | 909.4277 | 943.7457 | 951.3593 | -0.39464 | 2.75E-05 | 0.000711 | Erc1 | 6 | 1.2E+08 | 1.2E+08 | - | 16271 | protein_co | ELKS/RAB6 | - | 685 | 696 | 804 | 873 | 931 | 981 | 2.214168 | 2.2743 | 2.430903 | 2.950186 | 3.061514 | 3.086213 |
| ENSMUSG0 | 853.9248 | 855.1946 | 732.5764 | 1213.612 | 1023.827 | 1047.368 | -0.42817 | 2.78E-05 | 0.000714 | Rnf103 | 6 | 71493894 | 71510881 | + | 6602 | protein_co | ring finger p | - | 857 | 849 | 786 | 1165 | 1010 | 1080 | 6.827153 | 6.837305 | 5.856969 | 9.702862 | 8.185527 | 8.373737 |
| ENSMUSG0 | 1938.021 | 1721.47 | 1891.091 | 2167.834 | 2428.802 | 2246.992 | -0.30201 | 2.78E-05 | 0.000714 | Slc6a6 | 6 | 91684053 | 91759066 | + | 8457 | protein_co | solute carri | - | 1945 | 1709 | 2029 | 2081 | 2396 | 2317 | 12.09588 | 10.74431 | 11.80298 | 13.53023 | 15.15903 | 14.02429 |
| ENSMUSG0 | 84.69499 | 72.52534 | 68.03827 | 27.0849 | 23.31488 | 26.1842 | 1.556134 | 2.78E-05 | 0.000714 | Flt4 | 11 | 49609263 | 49652739 | + | 8232 | protein_co | FMS-like ty | - | 85 | 72 | 73 | 26 | 23 | 27 | 0.54306 | 0.465029 | 0.436258 | 0.173667 | 0.149494 | 0.167892 |
| ENSMUSG0 | 277.0024 | 263.9116 | 244.1921 | 430.2333 | 446.0237 | 350.0925 | -0.64331 | 2.81E-05 | 0.000721 | Cnot11 | 1 | 39534992 | 39546889 | + | 3508 | protein_co | CCR4-NOT t | - | 278 | 262 | 262 | 413 | 440 | 361 | 4.167922 | 3.970951 | 3.674241 | 6.473512 | 6.711103 | 5.267672 |
| ENSMUSG0 | 1075.128 | 1023.413 | 1055.991 | 752.1269 | 853.5272 | 816.5591 | 0.380912 | 2.98E-05 | 0.000763 | Zmym2 | 14 | 56886653 | 56962701 | + | 7860 | protein_co | zinc finger, | - | 1079 | 1016 | 1133 | 722 | 842 | 842 | 7.219934 | 6.872646 | 7.091421 | 5.050846 | 5.731791 | 5.483535 |
| ENSMUSG0 | 287.963 | 288.0868 | 236.7359 | 143.7583 | 183.4779 | 138.6793 | 0.802479 | 3.12E-05 | 0.000796 | Rtl1 | 12 | 1.1E+08 | 1.1E+08 | - | 6358 | protein_co | retrotransp | - | 289 | 286 | 254 | 138 | 181 | 143 | 2.390626 | 2.391654 | 1.965346 | 1.19346 | 1.523207 | 1.151295 |
| ENSMUSG0 | 1441.808 | 1671.105 | 1583.521 | 1878.234 | 1911.82 | 2067.582 | -0.31897 | 3.17E-05 | 0.000809 | Abtb2 | 2 | 1.04E+08 | 1.04E+08 | + | 6032 | protein_co | ankyrin rep | - | 1447 | 1659 | 1699 | 1803 | 1886 | 2132 | 12.61658 | 14.62305 | 13.85664 | 16.43554 | 16.72943 | 18.09243 |
| ENSMUSG0 | 1200.676 | 1176.522 | 1213.504 | 989.6407 | 925.4992 | 906.7492 | 0.347994 | 3.21E-05 | 0.000816 | Klhdc10 | 6 | 30401868 | 30455179 | + | 8443 | protein_co | kelch doma | - | 1205 | 1168 | 1302 | 950 | 913 | 935 | 7.506277 | 7.355274 | 7.586476 | 6.186945 | 5.785952 | 5.668732 |
| ENSMUSG0 | 1440.811 | 1499.864 | 1455.832 | 1178.193 | 1147.497 | 1213.201 | 0.312878 | 3.24E-05 | 0.000823 | Lpcat1 | 13 | 73467197 | 73516422 | + | 4632 | protein_co | lysophosph | - | 1446 | 1489 | 1562 | 1131 | 1132 | 1251 | 16.41852 | 17.09145 | 16.58969 | 13.4259 | 13.07611 | 13.82483 |
| ENSMUSG0 | 6296.325 | 6293.588 | 6656.566 | 6916.026 | 8072.016 | 7583.72 | -0.22991 | 3.30E-05 | 0.000836 | Fam180a | 6 | 35312668 | 35326141 | - | 1606 | protein_co | family with | - | 6319 | 6248 | 7142 | 6639 | 7963 | 7820 | 206.9365 | 206.8465 | 218.7763 | 227.3037 | 265.2968 | 249.2484 |
| ENSMUSG0 | 48.82417 | 48.35023 | 34.48515 | 6.250362 | 14.19166 | 3.879141 | 2.439048 | 3.33E-05 | 0.000842 | Ldb3 | 14 | 34526603 | 34588682 | - | 6105 | protein_co | LIM domain | - | 49 | 48 | 37 | 6 | 14 | 4 | 0.422129 | 0.418031 | 0.298155 | 0.05404 | 0.1227 | 0.033539 |
| ENSMUSG0 | 4491.824 | 4487.505 | 4948.153 | 5396.146 | 5273.217 | 5531.655 | -0.21796 | 3.43E-05 | 0.000865 | Cavin1 | 11 | 1.01E+08 | 1.01E+08 | - | 3587 | protein_co | caveolae as | - | 4508 | 4455 | 5309 | 5180 | 5202 | 5704 | 66.09778 | 66.03423 | 72.81272 | 79.405 | 77.59608 | 81.39903 |
| ENSMUSG0 | 37.86364 | 28.2043 | 32.62109 | 2.083454 | 8.109522 | 0 | 3.283621 | 3.46E-05 | 0.000871 | Tceal7 | X | | 1.36E+08 | 1.36E+08 | + | 1217 | protein_co | transcriptio | - | 38 | 28 | 35 | 2 | 8 | 0 | 1.642205 | 1.223264 | 1.414827 | 0.090363 | 0.351723 | 0 |
| ENSMUSG0 | 1209.644 | 1169.471 | 1183.679 | 937.5543 | 968.0742 | 902.87 | 0.343369 | 3.48E-05 | 0.000875 | Tmpo | 10 | 91147571 | 91181315 | - | 10998 | protein_co | thymopoie | - | 1214 | 1161 | 1270 | 900 | 955 | 931 | 5.805496 | 5.612693 | 5.680884 | 4.499645 | 4.64612 | 4.333183 |
| ENSMUSG0 | 33.878 | 56.4086 | 24.23281 | 5.208635 | 9.123213 | 2.909356 | 2.73433 | 3.50E-05 | 0.000878 | Actn2 | 13 | 12269426 | 12340760 | - | 3795 | protein_co | actinin alph | - | 34 | 56 | 26 | 5 | 9 | 3 | 0.471196 | 0.784565 | 0.337045 | 0.072445 | 0.126891 | 0.040465 |
| ENSMUSG0 | 1020.326 | 983.1213 | 1052.263 | 1329.244 | 1240.757 | 1287.875 | -0.33598 | 3.51E-05 | 0.000879 | Tsr1 | 11 | 74898071 | 74909342 | + | 3837 | protein_co | TSR1 20S rF | - | 1024 | 976 | 1129 | 1276 | 1224 | 1328 | 14.03597 | 13.52418 | 14.47532 | 18.28556 | 17.06831 | 17.71648 |
| ENSMUSG0 | 13983.64 | 14552.41 | 15294.63 | 13029.92 | 12315.32 | 13259.87 | 0.183195 | 3.53E-05 | 0.000881 | Chadl | 15 | 81678296 | 81697287 | - | 5257 | protein_co | chondroad | - | 14034 | 14447 | 16410 | 12508 | 12149 | 13673 | 140.4035 | 146.1142 | 153.5665 | 130.8276 | 123.6526 | 133.1364 |
| ENSMUSG0 | 231.1675 | 297.1524 | 230.2117 | 355.2289 | 416.6267 | 429.6148 | -0.6647 | 3.62E-05 | 0.000903 | Natd1 | 11 | 60902246 | 60914750 | - | 4247 | protein_co | N-acetyltra | - | 232 | 295 | 247 | 341 | 411 | 443 | 2.873029 | 3.693113 | 2.86115 | 4.414907 | 5.17798 | 5.339401 |
| ENSMUSG0 | 848.9428 | 788.7131 | 739.1006 | 1008.392 | 1215.415 | 985.3018 | -0.43339 | 3.63E-05 | 0.000903 | Lrp12 | 15 | 39870589 | 39943994 | - | 4588 | protein_co | low density | - | 852 | 783 | 793 | 968 | 1199 | 1016 | 9.76676 | 9.073841 | 8.503068 | 11.60116 | 13.98288 | 11.33552 |
| ENSMUSG0 | 1045.236 | 893.4719 | 880.7693 | 757.3356 | 669.0356 | 648.7863 | 0.442424 | 3.69E-05 | 0.000916 | Slc38a3 | 9 | 1.08E+08 | 1.08E+08 | - | 5277 | protein_co | solute carri | - | 1049 | 887 | 945 | 727 | 660 | 669 | 10.45497 | 8.93695 | 8.809893 | 7.575247 | 6.692027 | 6.489483 |
| ENSMUSG0 | 1545.435 | 1483.748 | 1590.045 | 1835.523 | 1895.601 | 1885.262 | -0.28183 | 3.70E-05 | 0.000918 | Bcat1 | 6 | 1.45E+08 | 1.45E+08 | - | 9322 | protein_co | branched c | - | 1551 | 1473 | 1706 | 1762 | 1870 | 1944 | 8.750584 | 8.401299 | 9.003178 | 10.39313 | 10.7333 | 10.67476 |
| ENSMUSG0 | 2629.53 | 2793.233 | 2921.917 | 3268.939 | 3341.123 | 3249.75 | -0.24046 | 3.71E-05 | 0.000918 | Plod3 | 5 | 1.37E+08 | 1.37E+08 | + | 4519 | protein_co | procollagen | - | 2639 | 2773 | 3135 | 3138 | 3296 | 3351 | 30.71365 | 32.62574 | 34.12881 | 38.18212 | 39.02525 | 37.95799 |
| ENSMUSG0 | 1204.662 | 1208.756 | 1281.543 | 1024.018 | 887.9927 | 967.8456 | 0.3599 | 3.81E-05 | 0.000942 | Colgalt2 | 1 | 1.52E+08 | 1.53E+08 | + | 4245 | protein_co | collagen be | - | 1209 | 1200 | 1375 | 983 | 876 | 998 | 14.979 | 15.02991 | 15.93496 | 12.73284 | 11.04148 | 12.03439 |
| ENSMUSG0 | 1671.979 | 1730.535 | 1644.103 | 1325.077 | 1435.385 | 1336.364 | 0.300686 | 3.83E-05 | 0.000944 | Pls3 | X | 75785654 | 75875182 | - | 4238 | protein_co | plastin 3 (T | - | 1678 | 1718 | 1764 | 1272 | 1416 | 1378 | 20.82406 | 21.55336 | 20.47687 | 16.50348 | 17.87735 | 16.64406 |
| ENSMUSG0 | 643.6819 | 706.1148 | 712.0717 | 910.4694 | 898.1296 | 900.9305 | -0.39385 | 3.84E-05 | 0.000945 | Klf13 | 7 | 63886351 | 63938915 | - | 7531 | protein_co | Kruppel-lik | zf-C2H2 | 646 | 701 | 764 | 874 | 886 | 929 | 4.51143 | 4.949008 | 4.990759 | 6.381287 | 6.294799 | 6.31443 |
| ENSMUSG0 | 1286.367 | 1249.048 | 1339.329 | 1550.09 | 1623.932 | 1604.995 | -0.30244 | 3.85E-05 | 0.000945 | Ccnt1 | 15 | 98538689 | 98570923 | - | 9157 | protein_co | cyclin T1 [S | - | 1291 | 1240 | 1437 | 1488 | 1602 | 1655 | 7.414935 | 7.199814 | 7.720216 | 8.935095 | 9.360738 | 9.251579 |
| ENSMUSG0 | 3443.599 | 3107.509 | 2984.363 | 2594.942 | 2735.95 | 2595.145 | 0.266534 | 3.93E-05 | 0.000963 | Srsf11 | 3 | 1.58E+08 | 1.58E+08 | - | 8446 | protein_co | serine/argi | - | 3456 | 3085 | 3202 | 2491 | 2699 | 2676 | 21.52073 | 19.42034 | 18.65074 | 16.21706 | 17.09829 | 16.21833 |
| ENSMUSG0 | 2539.853 | 2609.905 | 2771.86 | 3034.551 | 3380.657 | 3074.219 | -0.26031 | 3.99E-05 | 0.000976 | Mlec | 5 | 1.15E+08 | 1.15E+08 | - | 6007 | protein_co | malectin [S | - | 2549 | 2591 | 2974 | 2913 | 3335 | 3170 | 22.31755 | 22.93309 | 24.35618 | 26.66443 | 29.70565 | 27.01299 |
| ENSMUSG0 | 978.4763 | 1042.552 | 926.4388 | 1190.694 | 1256.976 | 1325.696 | -0.35704 | 4.00E-05 | 0.000976 | Cd44 | 2 | 1.03E+08 | 1.03E+08 | - | 6776 | protein_co | CD44 antig | - | 982 | 1035 | 994 | 1143 | 1240 | 1367 | 7.622061 | 8.121191 | 7.216704 | 9.275179 | 9.791497 | 10.32681 |
| ENSMUSG0 | 7171.175 | 7283.76 | 7274.502 | 8087.969 | 7947.332 | 8513.744 | -0.1761 | 4.06E-05 | 0.00099 | Surf4 | 2 | 26920040 | 26933928 | - | 3281 | protein_co | surfeit gene | - | 7197 | 7231 | 7805 | 7764 | 7840 | 8779 | 115.3665 | 117.1777 | 117.0288 | 130.1154 | 127.8529 | 136.9651 |
| ENSMUSG0 | 3592.064 | 3558.778 | 3399.117 | 2709.532 | 3064.386 | 3069.37 | 0.254231 | 4.11E-05 | 0.000999 | Lox | 18 | 52516069 | 52529867 | - | 4810 | protein_co | lysyl oxidas | - | 3605 | 3533 | 3647 | 2601 | 3023 | 3165 | 39.418 | 39.05273 | 37.30067 | 29.73341 | 33.62745 | 33.68214 |
| ENSMUSG0 | 488.2417 | 610.4216 | 582.5194 | 763.5859 | 768.3772 | 781.6469 | -0.46058 | 4.11E-05 | 0.000999 | Tlnrd1 | 7 | 83879873 | 83884305 | - | 4433 | protein_co | talin rod do | - | 490 | 606 | 625 | 733 | 758 | 806 | 5.813433 | 7.268214 | 6.935986 | 9.091922 | 9.148972 | 9.306972 |
| ENSMUSG0 | 2678.355 | 2550.474 | 2858.539 | 2944.962 | 3544.875 | 3468.922 | -0.30028 | 4.16E-05 | 0.001009 | Hectd1 | 12 | 51743722 | 51829536 | - | 14950 | protein_co | HECT doma | - | 2688 | 2532 | 3067 | 2827 | 3497 | 3577 | 9.456325 | 9.004826 | 10.09249 | 10.39762 | 12.5157 | 12.24754 |
| ENSMUSG0 | 3961.733 | 3759.23 | 4001.209 | 3461.659 | 3306.658 | 3317.635 | 0.217111 | 4.27E-05 | 0.001033 | Maged2 | X | 1.51E+08 | 1.51E+08 | - | 3239 | protein_co | melanoma | - | 3976 | 3732 | 4293 | 3323 | 3262 | 3421 | 64.56093 | 61.26092 | 65.20424 | 56.41166 | 53.88574 | 54.06463 |
| ENSMUSG0 | 1651.054 | 1743.63 | 1629.19 | 1279.241 | 1462.755 | 1254.902 | 0.329892 | 4.30E-05 | 0.001038 | Arhgap29 | 3 | 1.22E+08 | 1.22E+08 | + | 8815 | protein_co | Rho GTPase | - | 1657 | 1731 | 1748 | 1228 | 1443 | 1294 | 9.886317 | 10.44065 | 9.755398 | 7.659943 | 8.758804 | 7.514205 |
| ENSMUSG0 | 2163.21 | 2552.489 | 2235.01 | 1920.945 | 1932.094 | 1570.082 | 0.358267 | 4.32E-05 | 0.001041 | Ssc5d | 7 | 4925785 | 4944826 | + | 4590 | protein_co | scavenger r | - | 2171 | 2534 | 2398 | 1844 | 1906 | 1619 | 24.87605 | 29.35261 | 25.70173 | 22.0901 | 22.21831 | 18.05532 |
| ENSMUSG0 | 760.2621 | 784.6839 | 827.6436 | 999.0162 | 1107.963 | 987.2413 | -0.38275 | 4.34E-05 | 0.001044 | Larp4b | 13 | 9093881 | 9174451 | + | 8464 | protein_co | La ribonucl | - | 763 | 779 | 888 | 959 | 1093 | 1018 | 4.741145 | 4.893444 | 5.161349 | 6.230063 | 6.909479 | 6.156632 |
| ENSMUSG0 | 446.3924 | 400.904 | 411.0257 | 307.3095 | 277.7511 | 243.4161 | 0.603796 | 4.39E-05 | 0.001056 | Robo1 | 16 | 72308306 | 73046095 | + | 16511 | protein_co | roundabou | - | 448 | 398 | 441 | 295 | 274 | 251 | 1.427049 | 1.281629 | 1.313987 | 0.982422 | 0.887929 | 0.778165 |
| ENSMUSG0 | 590.8721 | 689.998 | 629.1209 | 433.3584 | 489.6124 | 438.3429 | 0.488465 | 4.41E-05 | 0.001057 | Pik3r2 | 8 | 70768176 | 70776713 | - | 4176 | protein_co | phosphoin | - | 593 | 685 | 675 | 416 | 483 | 452 | 7.468417 | 8.721333 | 7.951868 | 5.477499 | 6.188529 | 5.540501 |
| ENSMUSG0 | 587.8829 | 759.5015 | 594.6358 | 403.1484 | 468.3249 | 460.648 | 0.543056 | 4.42E-05 | 0.001058 | Marcksl1 | 4 | 1.3E+08 | 1.3E+08 | + | 1605 | protein_co | MARCKS-lik | - | 590 | 754 | 638 | 387 | 462 | 475 | 19.33354 | 24.97751 | 19.55562 | 13.25823 | 15.40167 | 15.1492 |
| ENSMUSG0 | 1235.55 | 1344.741 | 1353.309 | 1563.632 | 1629 | 1681.608 | -0.30936 | 4.44E-05 | 0.00106 | Tmem184b | 15 | 79360684 | 79403569 | - | 4702 | protein_co | transmemb | - | 1240 | 1335 | 1452 | 1501 | 1607 | 1734 | 13.8699 | 15.09564 | 15.19182 | 17.55285 | 18.28665 | 18.8772 |
| ENSMUSG0 | 3930.844 | 3619.216 | 3843.696 | 4230.453 | 4511.935 | 4541.504 | -0.22148 | 4.48E-05 | 0.001066 | Zbtb20 | 16 | 42875881 | 43642602 | + | 30730 | protein_co | zinc finger a | ZBTB | 3945 | 3593 | 4124 | 4061 | 4451 | 4683 | 6.751788 | 6.216522 | 6.602098 | 7.26641 | 7.749895 | 7.800684 |
| ENSMUSG0 | 643.6819 | 670.8594 | 654.2858 | 473.9858 | 516.982 | 419.917 | 0.481193 | 4.48E-05 | 0.001066 | Fbln5 | 12 | 1.02E+08 | 1.02E+08 | - | 6911 | protein_co | fibulin 5 [So | - | 646 | 666 | 702 | 455 | 510 | 433 | 4.916159 | 5.123729 | 4.997147 | 3.620095 | 3.948481 | 3.207141 |
| ENSMUSG0 | 1625.147 | 1749.674 | 1604.025 | 1949.071 | 2158.147 | 1989.029 | -0.29234 | 4.60E-05 | 0.001094 | Ddx21 | 10 | 62580251 | 62602281 | - | 5177 | protein_co | DEAD (Asp- | - | 1631 | 1737 | 1721 | 1871 | 2129 | 2051 | 16.56953 | 17.83916 | 16.35417 | 19.87216 | 22.00383 | 20.27956 |
| ENSMUSG0 | 622.7573 | 659.7791 | 609.5483 | 847.9658 | 802.8427 | 849.5318 | -0.40246 | 4.65E-05 | 0.001102 | Dgkd | 1 | 87853287 | 87945180 | + | 6850 | protein_co | diacylglyce | - | 625 | 655 | 654 | 814 | 792 | 876 | 4.798702 | 5.083976 | 4.696919 | 6.534063 | 6.186363 | 6.54613 |
| ENSMUSG0 | 931.6449 | 921.6762 | 1107.253 | 1231.321 | 1282.318 | 1332.485 | -0.37704 | 4.79E-05 | 0.001134 | Irf2bp2 | 8 | 1.27E+08 | 1.27E+08 | - | 5080 | protein_co | interferon r | - | 935 | 915 | 1188 | 1182 | 1265 | 1374 | 9.680153 | 9.576574 | 11.50479 | 12.79391 | 13.32379 | 13.84504 |
| ENSMUSG0 | 577.9188 | 543.94 | 508.8889 | 797.9629 | 745.0624 | 686.6079 | -0.45127 | 4.86E-05 | 0.00115 | Rabgef1 | 5 | 1.3E+08 | 1.3E+08 | + | 4168 | protein_co | RAB guanin | - | 580 | 540 | 546 | 766 | 735 | 708 | 7.318711 | 6.888408 | 6.444524 | 10.10533 | 9.435403 | 8.695141 |
| ENSMUSG0 | 1627.14 | 1670.097 | 1768.063 | 2005.325 | 2083.134 | 2029.76 | -0.27221 | 4.99E-05 | 0.001178 | Fuca1 | 4 | 1.36E+08 | 1.36E+08 | + | 3276 | protein_co | fucosidase, | - | 1633 | 1658 | 1897 | 1925 | 2055 | 2093 | 26.21662 | 26.90874 | 28.48717 | 32.30995 | 33.56362 | 32.70366 |
| ENSMUSG0 | 1602.23 | 1718.448 | 1443.716 | 1216.737 | 1251.908 | 1302.422 | 0.336713 | 5.07E-05 | 0.001194 | Top2a | 11 | 98992943 | 99024189 | - | 9013 | protein_co | topoisomer | - | 1608 | 1706 | 1549 | 1168 | 1235 | 1343 | 9.383201 | 10.06381 | 8.45489 | 7.125625 | 7.331594 | 7.627421 |
| ENSMUSG0 | 1462.732 | 1222.858 | 1464.221 | 1758.435 | 1825.656 | 1671.91 | -0.34047 | 5.16E-05 | 0.001213 | Slc1a4 | 11 | 20302180 | 20332713 | - | 3926 | protein_co | solute carri | - | 1468 | 1214 | 1571 | 1688 | 1801 | 1724 | 19.66573 | 16.44073 | 19.68574 | 23.64131 | 24.54507 | 22.47802 |
| ENSMUSG0 | 71.74164 | 100.7296 | 84.81482 | 166.6763 | 189.5601 | 150.3167 | -0.97734 | 5.22E-05 | 0.001226 | 5430430B1 | 8 | 27279441 | 27281070 | + | 1630 | TEC | RIKEN cDNA | - | 72 | 100 | 91 | 160 | 187 | 155 | 2.323161 | 3.261859 | 2.746501 | 5.397366 | 6.138395 | 4.867603 |
| ENSMUSG0 | 803.1078 | 758.4942 | 792.2264 | 1026.101 | 950.8415 | 1075.492 | -0.37503 | 5.27E-05 | 0.001234 | Retnlg | 16 | 48872621 | 48874496 | + | 593 | protein_co | resistin like | - | 806 | 753 | 850 | 985 | 938 | 1109 | 71.48496 | 67.51388 | 70.5164 | 91.33369 | 84.6348 | 95.72997 |
| ENSMUSG0 | 165.4043 | 201.4593 | 178.95 | 102.0892 | 79.06784 | 105.7066 | 0.927531 | 5.33E-05 | 0.001247 | Zfp385c | 11 | 1.01E+08 | 1.01E+08 | - | 3101 | protein_co | zinc finger p | - | 166 | 200 | 192 | 98 | 78 | 109 | 2.815404 | 3.429107 | 3.045968 | 1.737696 | 1.345841 | 1.799268 |
| ENSMUSG0 | 389.597 | 389.8237 | 438.0546 | 569.8247 | 575.7761 | 559.5661 | -0.48534 | 5.40E-05 | 0.001261 | Kctd15 | 7 | 34639014 | 34656732 | - | 4238 | protein_co | potassium c | - | 391 | 387 | 470 | 547 | 568 | 577 | 4.852328 | 4.855152 | 5.455855 | 7.097017 | 7.17114 | 6.969248 |
| ENSMUSG0 | 4757.866 | 5031.445 | 4925.784 | 5390.937 | 6320.359 | 5629.603 | -0.23691 | 5.44E-05 | 0.001267 | Twsg1 | 17 | 65921972 | 65951226 | - | 5153 | protein_co | twisted gas | - | 4775 | 4995 | 5285 | 5175 | 6235 | 5805 | 48.73574 | 51.53807 | 50.45576 | 55.22042 | 64.74066 | 57.66511 |
| ENSMUSG0 | 8264.238 | 8505.611 | 8968.003 | 7676.487 | 7065.421 | 7707.853 | 0.197268 | 5.46E-05 | 0.001269 | Hist1h1c | 13 | 23738808 | 23740367 | + | 1560 | protein_co | histone clu | - | 8294 | 8444 | 9622 | 7369 | 6970 | 7948 | 279.6236 | 287.7905 | 303.4357 | 259.7368 | 239.0611 | 260.7981 |
| ENSMUSG0 | 3354.918 | 3413.727 | 3321.759 | 4010.649 | 4041.583 | 3702.64 | -0.22016 | 5.49E-05 | 0.001274 | Boc | 16 | 44485049 | 44558897 | - | 4451 | protein_co | biregional c | - | 3367 | 3389 | 3564 | 3850 | 3987 | 3818 | 39.78505 | 40.48245 | 39.39182 | 47.56118 | 47.92802 | 43.90858 |
| ENSMUSG0 | 459.3458 | 518.7576 | 470.6757 | 698.9988 | 619.3648 | 677.8799 | -0.46265 | 5.54E-05 | 0.001285 | Hs6st1 | 1 | 36068400 | 36106446 | + | 5542 | protein_co | heparan sul | - | 461 | 515 | 505 | 671 | 611 | 699 | 4.374906 | 4.940757 | 4.482814 | 6.657412 | 5.898961 | 6.456271 |
| ENSMUSG0 | 950.5767 | 745.3993 | 887.2935 | 1263.615 | 1207.305 | 1029.912 | -0.43791 | 5.60E-05 | 0.001296 | Hr | 14 | 70552212 | 70573548 | + | 6919 | protein_co | hairless [So | - | 954 | 740 | 952 | 1213 | 1191 | 1062 | 7.251693 | 5.686449 | 6.768922 | 9.639776 | 9.210204 | 7.856919 |
| ENSMUSG0 | 254.085 | 303.1962 | 197.5906 | 373.98 | 419.6678 | 415.0681 | -0.68082 | 5.76E-05 | 0.00133 | Ptpn3 | 4 | 57190841 | 57301837 | - | 7238 | protein_co | protein tyr | - | 255 | 301 | 212 | 359 | 414 | 428 | 1.852917 | 2.211061 | 1.440931 | 2.727252 | 3.060431 | 3.026887 |
| ENSMUSG0 | 1304.303 | 1374.96 | 1293.659 | 1743.851 | 1517.494 | 1685.487 | -0.31642 | 5.80E-05 | 0.001338 | Ctsf | 19 | 4855129 | 4860912 | + | 1975 | protein_co | cathepsin F | - | 1309 | 1365 | 1388 | 1674 | 1497 | 1738 | 34.85835 | 36.7467 | 34.57389 | 46.60557 | 40.55604 | 45.04574 |
| ENSMUSG0 | 1043.243 | 1042.552 | 1132.418 | 1404.248 | 1373.55 | 1275.268 | -0.33214 | 5.87E-05 | 0.001352 | Rela | 19 | 5637483 | 5648130 | + | 2716 | protein_co | v-rel reticul | RHD | 1047 | 1035 | 1215 | 1348 | 1355 | 1315 | 20.27455 | 20.26112 | 22.00759 | 27.29038 | 26.6938 | 24.78375 |
| ENSMUSG0 | 1666.997 | 1528.069 | 1389.658 | 1817.814 | 1984.806 | 1918.235 | -0.31988 | 5.91E-05 | 0.001359 | Arl5b | 2 | 15049395 | 15082456 | + | 10428 | protein_co | ADP-ribosy | - | 1673 | 1517 | 1491 | 1745 | 1958 | 1978 | 8.437801 | 7.734591 | 7.034003 | 9.201187 | 10.04645 | 9.709488 |
| ENSMUSG0 | 830.0109 | 689.998 | 787.5662 | 1040.685 | 930.5677 | 1118.162 | -0.42096 | 5.96E-05 | 0.001367 | Plaur | 7 | 24462484 | 24475968 | + | 2617 | protein_co | plasminoge | - | 833 | 685 | 845 | 999 | 918 | 1153 | 16.74078 | 13.91681 | 15.8847 | 20.98994 | 18.76894 | 22.5526 |
| ENSMUSG0 | 562.9726 | 569.1225 | 552.6944 | 727.1255 | 766.3499 | 744.795 | -0.41 | 5.97E-05 | 0.001368 | Icmt | 4 | 1.52E+08 | 1.52E+08 | + | 5287 | protein_co | isoprenylcy | - | 565 | 565 | 593 | 698 | 756 | 768 | 5.620481 | 5.681878 | 5.517868 | 7.259314 | 7.650914 | 7.435719 |
| ENSMUSG0 | 191.311 | 162.1747 | 149.125 | 76.04607 | 68.93094 | 104.7368 | 1.007074 | 6.31E-05 | 0.001441 | Npr1 | 3 | 90450591 | 90465866 | - | 4783 | protein_co | natriuretic | - | 192 | 161 | 160 | 73 | 68 | 108 | 2.111228 | 1.789692 | 1.64568 | 0.839212 | 0.760693 | 1.155831 |
| ENSMUSG0 | 116.5802 | 157.1382 | 137.9406 | 66.67053 | 73.99939 | 54.30797 | 1.079093 | 6.49E-05 | 0.00148 | Foxp2 | 6 | 14901349 | 15441977 | + | 16346 | protein_co | forkhead bo | Fork | 117 | 156 | 148 | 64 | 73 | 56 | 0.376451 | 0.507418 | 0.445427 | 0.215287 | 0.238953 | 0.175367 |
| ENSMUSG0 | 8578.108 | 8983.069 | 8723.81 | 9952.66 | 9580.387 | 9829.743 | -0.15976 | 6.56E-05 | 0.001495 | Brd2 | 17 | 34112023 | 34122634 | - | 7285 | protein_co | bromodom | - | 8609 | 8918 | 9360 | 9554 | 9451 | 10136 | 62.15234 | 65.08647 | 63.20802 | 72.1116 | 69.41431 | 71.22101 |
| ENSMUSG0 | 1033.279 | 1074.785 | 1083.952 | 1253.198 | 1425.249 | 1335.394 | -0.33054 | 6.79E-05 | 0.001545 | Phlda1 | 10 | 1.12E+08 | 1.12E+08 | + | 2205 | protein_co | pleckstrin h | - | 1037 | 1067 | 1163 | 1203 | 1406 | 1377 | 24.73458 | 25.72815 | 25.94759 | 29.99898 | 34.11753 | 31.9666 |
| ENSMUSG0 | 1542.445 | 1366.901 | 1555.56 | 1933.445 | 1751.657 | 1841.622 | -0.30743 | 6.82E-05 | 0.001549 | Vegfa | 17 | 46016993 | 46032369 | - | 4285 | protein_co | vascular en | - | 1548 | 1357 | 1669 | 1856 | 1728 | 1899 | 19.00004 | 16.83766 | 19.16158 | 23.81643 | 21.57713 | 22.68534 |
| ENSMUSG0 | 673.5743 | 658.7718 | 693.4311 | 820.8809 | 984.2933 | 882.5045 | -0.40782 | 6.89E-05 | 0.001562 | Nrip1 | 16 | 76287400 | 76373827 | - | 11457 | protein_co | nuclear rec | - | 676 | 654 | 744 | 788 | 971 | 910 | 3.103203 | 3.035007 | 3.194684 | 3.781854 | 4.534706 | 4.065758 |
| ENSMUSG0 | 426.4642 | 381.7653 | 351.3757 | 579.2002 | 527.1189 | 542.1099 | -0.50789 | 6.97E-05 | 0.001573 | Agfg2 | 5 | 1.38E+08 | 1.38E+08 | - | 3822 | protein_co | ArfGAP wit | - | 428 | 379 | 377 | 556 | 520 | 559 | 5.889622 | 5.272315 | 4.852623 | 7.998961 | 7.2797 | 7.486731 |
| ENSMUSG0 | 7065.555 | 7370.388 | 7194.348 | 8024.423 | 7971.66 | 8237.356 | -0.16402 | 6.98E-05 | 0.001573 | Eif1 | 11 | 1E+08 | 1E+08 | + | 2098 | protein_co | eukaryotic | - | 7091 | 7317 | 7719 | 7703 | 7864 | 8494 | 177.7609 | 185.4302 | 181.0012 | 201.8849 | 200.5575 | 207.242 |
| ENSMUSG0 | 259.067 | 186.3498 | 274.0171 | 356.2706 | 432.8458 | 371.4277 | -0.68885 | 6.98E-05 | 0.001573 | Serpini1 | 3 | 75557547 | 75643495 | + | 7286 | protein_co | serine (or c | - | 260 | 185 | 294 | 342 | 427 | 383 | 1.876802 | 1.350005 | 1.985108 | 2.58099 | 3.135736 | 2.690795 |
| ENSMUSG0 | 1039.257 | 1025.428 | 961.856 | 1236.53 | 1395.852 | 1218.05 | -0.34754 | 7.11E-05 | 0.0016 | Slc2a10 | 2 | 1.66E+08 | 1.66E+08 | + | 3570 | protein_co | solute carri | - | 1043 | 1018 | 1032 | 1187 | 1377 | 1256 | 15.36563 | 15.16116 | 14.22124 | 18.28235 | 20.63795 | 18.00912 |
| ENSMUSG0 | 544.0408 | 505.6628 | 510.753 | 682.3312 | 705.5284 | 703.0943 | -0.42237 | 7.24E-05 | 0.001627 | Ackr3 | 1 | 90203980 | 90216751 | + | 3348 | protein_co | atypical ch | - | 546 | 502 | 548 | 655 | 696 | 725 | 8.577121 | 7.972069 | 8.05232 | 10.75735 | 11.12307 | 11.08469 |
| ENSMUSG0 | 867.8746 | 772.5963 | 763.3334 | 1031.31 | 1031.937 | 1032.821 | -0.36538 | 7.35E-05 | 0.001649 | Tfe3 | X | 7762560 | 7775202 | + | 9261 | protein_co | transcriptio | bHLH | 871 | 767 | 819 | 990 | 1018 | 1065 | 4.946461 | 4.403422 | 4.350628 | 5.877962 | 5.881535 | 5.886577 |
| ENSMUSG0 | 94.65911 | 49.35752 | 96.93123 | 34.37699 | 16.21904 | 26.1842 | 1.651994 | 7.69E-05 | 0.001723 | Igll1 | 16 | 16860671 | 16863985 | - | 2164 | protein_co | immunoglo | - | 95 | 49 | 104 | 33 | 16 | 27 | 2.308876 | 1.203903 | 2.364297 | 0.838506 | 0.395607 | 0.638672 |
| ENSMUSG0 | 1563.37 | 1557.28 | 1476.337 | 1871.983 | 1912.834 | 1780.526 | -0.27585 | 7.88E-05 | 0.001761 | 6-Mar | 15 | 31455891 | 31531053 | - | 9174 | protein_co | membrane | - | 1569 | 1546 | 1584 | 1797 | 1887 | 1836 | 8.994946 | 8.959908 | 8.494197 | 10.77057 | 11.00561 | 10.24436 |

| gene_id | H1 | H2 | H3 | M1 | M2 | M3 | log2FoldCh | pvalue | padj | gene_name | gene_chr | gene_start | gene_end | gene_stran | gene_lengtl | gene_bioty | gene_descr | tf_family | H1_readcou | H2_readcou | H3_readcou | M1_readco | M2_readco | M3_readco | H1_fpkm | H2_fpkm | H3_fpkm | M1_fpkm | M2_fpkm | M3_fpkm |
|---|---|---|---|---|---|---|---|---|---|---|---|---|---|---|---|---|---|---|---|---|---|---|---|---|---|---|---|---|---|---|
| ENSMUSG0 | 411.518 | 429.1083 | 383.0648 | 563.5743 | 637.6112 | 526.5934 | -0.49789 | 8.22E-05 | 0.001834 | Fbxo28 | 1 | 1.82E+08 | 1.82E+08 | - | 5318 | protein_co | F-box prote | - | 413 | 426 | 411 | 541 | 629 | 543 | 4.084473 | 4.259063 | 3.802064 | 5.59369 | 6.328534 | 5.22664 |
| ENSMUSG0 | 1025.308 | 1067.734 | 961.856 | 1244.864 | 1405.988 | 1232.597 | -0.34649 | 8.39E-05 | 0.001847 | Tomm70a | 16 | 57121703 | 57156705 | + | 9777 | protein_co | translocase | - | 1029 | 1060 | 1032 | 1195 | 1387 | 1271 | 5.535337 | 5.764386 | 5.192781 | 6.720657 | 7.590522 | 6.654432 |
| ENSMUSG0 | 355.719 | 386.8018 | 372.8124 | 495.8621 | 503.8041 | 586.72 | -0.50881 | 8.44E-05 | 0.001876 | Rnf149 | 1 | 39551296 | 39577405 | - | 2838 | protein_co | ring finger | - | 357 | 384 | 400 | 476 | 497 | 605 | 6.615919 | 7.19402 | 6.933835 | 9.222401 | 9.370113 | 10.91224 |
| ENSMUSG0 | 1373.055 | 1376.974 | 1412.959 | 1682.389 | 1865.19 | 1600.146 | -0.30606 | 8.45E-05 | 0.001876 | Tmf1 | 6 | 97152997 | 97179122 | - | 5732 | protein_co | TATA eleme | - | 1378 | 1367 | 1516 | 1615 | 1840 | 1650 | 12.64379 | 12.67988 | 13.01125 | 15.4923 | 17.17562 | 14.73496 |
| ENSMUSG0 | 2935.429 | 3087.363 | 2953.606 | 3556.456 | 3443.506 | 3400.067 | -0.21236 | 8.48E-05 | 0.001881 | Itgb5 | 16 | 33829665 | 33949338 | + | 3553 | protein_co | integrin be | - | 2946 | 3065 | 3169 | 3414 | 3397 | 3506 | 43.60857 | 45.8657 | 43.87862 | 52.83452 | 51.15654 | 50.51121 |
| ENSMUSG0 | 423.475 | 438.1739 | 507.0249 | 681.2895 | 617.3374 | 607.0855 | -0.47652 | 8.50E-05 | 0.001882 | Inf2 | 12 | 1.13E+08 | 1.13E+08 | + | 4871 | protein_co | inverted fo | - | 425 | 435 | 544 | 654 | 609 | 626 | 4.588863 | 4.748144 | 5.494228 | 7.382596 | 6.689597 | 6.578506 |
| ENSMUSG0 | 95.65552 | 79.57641 | 79.22264 | 30.21008 | 32.43809 | 37.82162 | 1.339071 | 8.67E-05 | 0.001915 | Pi16 | 17 | 29318877 | 29329413 | + | 3987 | protein_co | peptidase i | - | 96 | 79 | 85 | 29 | 32 | 39 | 1.266366 | 1.053498 | 1.048814 | 0.399946 | 0.429442 | 0.500714 |
| ENSMUSG0 | 307.8912 | 261.8971 | 221.8234 | 390.6476 | 407.5035 | 412.1587 | -0.61366 | 8.75E-05 | 0.001929 | Dusp10 | 1 | 1.84E+08 | 1.84E+08 | + | 3042 | protein_co | dual specifi | - | 309 | 260 | 238 | 375 | 402 | 425 | 5.342366 | 4.544299 | 3.848962 | 6.778311 | 7.070785 | 7.15156 |
| ENSMUSG0 | 6553.4 | 6605.85 | 6771.205 | 5682.621 | 6079.101 | 5871.08 | 0.176729 | 8.79E-05 | 0.001936 | Nfatc1 | 18 | 80606205 | 80713071 | - | 7433 | protein_co | nuclear fac | RHD | 6577 | 6558 | 7265 | 5455 | 5997 | 6054 | 46.53696 | 46.90942 | 48.08365 | 40.3534 | 43.16887 | 41.69168 |
| ENSMUSG0 | 1003.387 | 1153.354 | 993.5451 | 771.9197 | 829.1987 | 826.257 | 0.37557 | 8.97E-05 | 0.001972 | Smarcc2 | 10 | 1.28E+08 | 1.28E+08 | + | 5552 | protein_co | SWI/SNF re | MYB | 1007 | 1145 | 1066 | 741 | 818 | 852 | 9.539253 | 10.965 | 9.445689 | 7.338684 | 7.883238 | 7.855272 |
| ENSMUSG0 | 661.6174 | 730.2899 | 735.3725 | 929.2205 | 1016.731 | 860.1995 | -0.39916 | 9.00E-05 | 0.001975 | Gpx3 | 11 | 54902453 | 54910377 | + | 1813 | protein_co | glutathione | - | 664 | 725 | 789 | 892 | 1003 | 887 | 19.26214 | 21.26146 | 21.40943 | 27.05307 | 29.60083 | 25.04361 |
| ENSMUSG0 | 743.3231 | 733.3118 | 709.2756 | 852.1327 | 1033.964 | 990.1507 | -0.39642 | 9.12E-05 | 0.001997 | Senp5 | 16 | 31959672 | 32003287 | - | 7260 | protein_co | SUMO/sent | - | 746 | 728 | 761 | 818 | 1020 | 1021 | 5.404264 | 5.331477 | 5.156725 | 6.195354 | 7.517343 | 7.198802 |
| ENSMUSG0 | 82.70217 | 101.7369 | 71.76639 | 166.6763 | 180.4369 | 147.4074 | -0.94991 | 9.15E-05 | 0.002 | Mmp28 | 11 | 83440768 | 83463071 | - | 3874 | protein_co | matrix met | - | 83 | 101 | 77 | 160 | 178 | 152 | 1.126816 | 1.386164 | 0.977816 | 2.270962 | 2.458449 | 2.008422 |
| ENSMUSG0 | 5129.527 | 5037.489 | 4970.521 | 5570.114 | 5853.048 | 5697.488 | -0.1777 | 9.24E-05 | 0.002017 | Copb2 | 9 | 98563721 | 98588382 | + | 6351 | protein_co | coatomer p | - | 5148 | 5001 | 5333 | 5347 | 5774 | 5875 | 42.63151 | 41.86659 | 41.31002 | 46.29324 | 48.64469 | 47.35184 |
| ENSMUSG0 | 1852.329 | 1693.265 | 1767.131 | 2033.451 | 2262.557 | 2124.799 | -0.27335 | 9.52E-05 | 0.002075 | Lats1 | 10 | 7681214 | 7716460 | + | 7297 | protein_co | large tumo | - | 1859 | 1681 | 1896 | 1952 | 2232 | 2191 | 13.39891 | 12.24831 | 12.78262 | 14.70906 | 16.36631 | 15.36983 |
| ENSMUSG0 | 72.73805 | 92.67127 | 68.03827 | 20.83454 | 15.20535 | 39.76119 | 1.618063 | 9.55E-05 | 0.002078 | Ppfibp2 | 7 | 1.08E+08 | 1.08E+08 | + | 10553 | protein_co | PTPRF inter | - | 73 | 92 | 73 | 20 | 15 | 41 | 0.363816 | 0.463516 | 0.340309 | 0.104209 | 0.076053 | 0.198874 |
| ENSMUSG0 | 968.5122 | 1018.377 | 1003.797 | 777.1284 | 744.0487 | 816.5591 | 0.355162 | 9.59E-05 | 0.002083 | Axl | 7 | 25757273 | 25788705 | - | 5033 | protein_co | AXL recepto | - | 972 | 1011 | 1077 | 746 | 734 | 842 | 10.15719 | 10.68014 | 10.52724 | 8.15007 | 7.80315 | 8.563597 |
| ENSMUSG0 | 705.4595 | 631.5748 | 647.7616 | 893.8018 | 825.1439 | 877.6556 | -0.3877 | 9.61E-05 | 0.002084 | Chsy3 | 18 | 59175401 | 59410446 | + | 2931 | protein_co | chondroiti | - | 708 | 627 | 695 | 858 | 814 | 905 | 12.70433 | 11.37377 | 11.66527 | 16.09611 | 14.85968 | 15.80534 |
| ENSMUSG0 | 200.2787 | 218.5833 | 217.1632 | 346.8951 | 342.6273 | 294.8147 | -0.62943 | 9.63E-05 | 0.002086 | Zfp217 | 2 | 1.7E+08 | 1.7E+08 | - | 6139 | protein_co | zinc finger p | zf-C2H2 | 201 | 217 | 233 | 333 | 338 | 304 | 1.721998 | 1.879381 | 1.867171 | 2.982607 | 2.945913 | 2.534819 |
| ENSMUSG0 | 253.0886 | 199.4447 | 202.2507 | 125.0072 | 127.725 | 125.1023 | 0.792976 | 9.80E-05 | 0.002119 | Cped1 | 6 | 21985916 | 22256404 | + | 8279 | protein_co | cadherin-li | - | 254 | 198 | 217 | 120 | 126 | 129 | 1.613579 | 1.27157 | 1.28946 | 0.79699 | 0.814317 | 0.797596 |
| ENSMUSG0 | 1068.153 | 1253.077 | 1163.175 | 1411.54 | 1460.728 | 1480.862 | -0.32132 | 0.000102 | 0.002206 | Nus1 | 10 | 52417547 | 52440183 | + | 4867 | protein_co | NUS1 dehy | - | 1072 | 1244 | 1248 | 1355 | 1441 | 1527 | 11.58425 | 13.58976 | 12.61476 | 15.30832 | 15.84176 | 16.06012 |
| ENSMUSG0 | 833.0002 | 886.4208 | 804.3428 | 637.5369 | 638.6249 | 659.4539 | 0.382361 | 0.000103 | 0.002213 | Acta2 | 19 | 34241090 | 34255336 | - | 1781 | protein_co | actin, alpha | - | 836 | 880 | 863 | 612 | 630 | 680 | 24.68748 | 26.27069 | 23.83816 | 18.89457 | 18.92681 | 19.54412 |
| ENSMUSG0 | 254.085 | 279.0211 | 210.639 | 168.7598 | 120.6291 | 138.6793 | 0.796889 | 0.000103 | 0.00222 | Ddah2 | 17 | 35059035 | 35062095 | + | 1593 | protein_co | dimethylar | - | 255 | 277 | 226 | 162 | 119 | 143 | 8.418966 | 9.24521 | 6.979408 | 5.591762 | 3.99698 | 4.595062 |
| ENSMUSG0 | 334.7943 | 347.5172 | 340.1913 | 227.0965 | 163.2041 | 245.3557 | 0.685282 | 0.000105 | 0.002263 | Ebf3 | 7 | 1.37E+08 | 1.37E+08 | - | 9227 | protein_co | early B cell | COE | 336 | 345 | 365 | 218 | 161 | 253 | 1.915195 | 1.987977 | 1.946069 | 1.299109 | 0.933611 | 1.40356 |
| ENSMUSG0 | 11865.27 | 11965.67 | 12163.94 | 10994.39 | 10962.05 | 10619.15 | 0.144096 | 0.000107 | 0.002284 | Eno1 | 4 | 1.5E+08 | 1.5E+08 | + | 5379 | protein_co | enolase 1, a | - | 11908 | 11879 | 13051 | 10554 | 10814 | 10950 | 116.4318 | 117.417 | 119.3626 | 107.886 | 107.5686 | 104.2038 |
| ENSMUSG0 | 1046.232 | 1043.559 | 1041.079 | 894.8435 | 787.6374 | 724.4295 | 0.380005 | 0.000107 | 0.002284 | Ints6l | X | 56454857 | 56507843 | + | 7173 | protein_co | integrator c | - | 1050 | 1036 | 1117 | 859 | 777 | 747 | 7.698796 | 7.679125 | 7.660873 | 6.584788 | 5.795902 | 5.330781 |
| ENSMUSG0 | 1672.975 | 1612.681 | 1684.18 | 1895.943 | 1980.751 | 2121.89 | -0.2716 | 0.000107 | 0.002284 | Smg1 | 7 | 1.18E+08 | 1.18E+08 | - | 23984 | protein_co | SMG1 hom | - | 1679 | 1601 | 1807 | 1820 | 1954 | 2188 | 3.681827 | 3.549135 | 3.706487 | 4.172528 | 4.359169 | 4.669784 |
| ENSMUSG0 | 69.74882 | 41.29915 | 59.64999 | 106.2562 | 122.6565 | 149.3469 | -1.14799 | 0.000109 | 0.002324 | Fam83g | 11 | 61684091 | 61709951 | + | 6659 | protein_co | family with | - | 70 | 41 | 64 | 102 | 121 | 154 | 0.55287 | 0.327362 | 0.472821 | 0.842249 | 0.972248 | 1.183812 |
| ENSMUSG0 | 659.6245 | 710.1439 | 729.7803 | 507.3211 | 563.6118 | 480.0437 | 0.437361 | 0.00011 | 0.002349 | Plxnd1 | 6 | 1.16E+08 | 1.16E+08 | - | 7301 | protein_co | plexin D1 [S | - | 662 | 705 | 783 | 487 | 556 | 495 | 4.76881 | 5.134044 | 5.276006 | 3.667719 | 4.074678 | 3.470515 |
| ENSMUSG0 | 247.1101 | 251.8241 | 262.8328 | 348.9786 | 357.8327 | 459.6782 | -0.61528 | 0.000111 | 0.002352 | Has1 | 17 | 17843323 | 17855205 | - | 4518 | protein_co | hyaluronar | - | 248 | 250 | 282 | 335 | 353 | 474 | 2.886954 | 2.942026 | 3.070639 | 4.077069 | 4.18051 | 5.370357 |
| ENSMUSG0 | 1572.338 | 1603.616 | 1452.104 | 1855.316 | 1784.095 | 2045.277 | -0.29723 | 0.000111 | 0.002352 | Ube2z | 11 | 96047432 | 96065388 | - | 3997 | protein_co | ubiquitin-c | - | 1578 | 1592 | 1558 | 1781 | 1760 | 2109 | 20.76382 | 21.17687 | 19.17605 | 24.50074 | 23.56022 | 27.00931 |
| ENSMUSG0 | 1485.65 | 1477.704 | 1571.404 | 1785.52 | 1792.204 | 1894.96 | -0.27115 | 0.000111 | 0.002352 | Golga3 | 5 | 1.1E+08 | 1.1E+08 | + | 12671 | protein_co | golgi autoa | - | 1491 | 1467 | 1686 | 1714 | 1768 | 1954 | 6.188724 | 6.155623 | 6.545948 | 7.437884 | 7.465728 | 7.893775 |
| ENSMUSG0 | 256.0778 | 284.0576 | 288.9296 | 159.3842 | 188.5464 | 169.7124 | 0.67958 | 0.000111 | 0.002352 | Svep1 | 4 | 58042442 | 58206859 | - | 16117 | protein_co | sushi, von \ | - | 257 | 282 | 310 | 153 | 186 | 175 | 0.838655 | 0.930289 | 0.946245 | 0.521983 | 0.617489 | 0.555808 |
| ENSMUSG0 | 2175.167 | 1995.454 | 2207.049 | 2549.106 | 2493.678 | 2515.623 | -0.24478 | 0.000113 | 0.002386 | Pafah1b1 | 11 | 74673949 | 74724670 | - | 9620 | protein_co | platelet-act | - | 2183 | 1981 | 2368 | 2447 | 2460 | 2594 | 11.93474 | 10.94869 | 12.10968 | 13.98648 | 13.68235 | 13.80276 |
| ENSMUSG0 | 924.67 | 851.1654 | 806.2068 | 1077.146 | 1069.443 | 1148.226 | -0.35224 | 0.000113 | 0.002386 | Phf2 | 13 | 48801750 | 48871119 | - | 5494 | protein_co | PHD finger | - | 928 | 845 | 865 | 1034 | 1055 | 1184 | 8.883695 | 8.177506 | 7.745569 | 10.34859 | 10.27459 | 11.03149 |
| ENSMUSG0 | 1013.351 | 1096.946 | 1130.554 | 1303.201 | 1363.413 | 1361.578 | -0.31361 | 0.000115 | 0.002418 | Sbno2 | 10 | 80056992 | 80105571 | - | 10715 | protein_co | strawberry | - | 1017 | 1089 | 1213 | 1251 | 1345 | 1404 | 4.991868 | 5.403666 | 5.569222 | 6.419698 | 6.716313 | 6.707273 |
| ENSMUSG0 | 565.9618 | 599.3413 | 562.0147 | 378.1469 | 389.2571 | 464.5271 | 0.486676 | 0.000118 | 0.00249 | Notch1 | 2 | 26457903 | 26516663 | - | 14232 | protein_co | notch 1 [So | - | 568 | 595 | 603 | 363 | 384 | 479 | 2.099021 | 2.222818 | 2.084382 | 1.402459 | 1.443664 | 1.722823 |
| ENSMUSG0 | 274.0132 | 342.4808 | 286.1335 | 403.1484 | 442.9827 | 496.53 | -0.57387 | 0.000119 | 0.00249 | Urgcp | 11 | 5713417 | 5762376 | - | 5871 | protein_co | upregulato | - | 275 | 340 | 307 | 387 | 437 | 512 | 2.463514 | 3.079071 | 2.572481 | 3.624502 | 3.982632 | 4.464049 |
| ENSMUSG0 | 1299.321 | 1428.346 | 1261.97 | 1535.506 | 1693.876 | 1711.671 | -0.30911 | 0.000119 | 0.002497 | Lrrc58 | 16 | 37868389 | 37888858 | + | 8604 | protein_co | leucine rich | - | 1304 | 1418 | 1354 | 1474 | 1671 | 1765 | 7.970976 | 8.762512 | 7.741839 | 9.419905 | 10.39147 | 10.50063 |
| ENSMUSG0 | 3477.477 | 3335.158 | 3332.011 | 3851.265 | 4223.034 | 3761.797 | -0.22245 | 0.000122 | 0.00255 | Pdia4 | 6 | 47796141 | 47813430 | - | 2399 | protein_co | protein dis | - | 3490 | 3311 | 3575 | 3697 | 4166 | 3879 | 76.51199 | 73.38068 | 73.31143 | 84.73614 | 92.91586 | 82.76765 |
| ENSMUSG0 | 282.9809 | 248.8022 | 215.2992 | 135.4245 | 166.2452 | 141.5886 | 0.752424 | 0.000126 | 0.002627 | Gm973 | 1 | 59516264 | 59636417 | + | 5484 | protein_co | predicted g | - | 284 | 247 | 231 | 130 | 164 | 146 | 2.723675 | 2.394707 | 2.072242 | 1.303453 | 1.6001 | 1.362782 |
| ENSMUSG0 | 426.4642 | 430.1156 | 395.1812 | 564.6161 | 761.2814 | 518.8351 | -0.55949 | 0.000127 | 0.002635 | Slc25a32 | 15 | 39091231 | 39112716 | - | 6081 | protein_co | solute carri | - | 428 | 427 | 424 | 542 | 751 | 535 | 3.701716 | 3.73341 | 3.430179 | 4.900876 | 6.607935 | 4.503497 |
| ENSMUSG0 | 538.0623 | 431.1228 | 469.7436 | 682.3312 | 631.529 | 663.3331 | -0.45843 | 0.000127 | 0.002635 | Fam126a | 5 | 23915276 | 24030690 | - | 10117 | protein_co | family with | - | 540 | 428 | 504 | 655 | 623 | 684 | 2.807219 | 2.249287 | 2.450782 | 3.55991 | 3.294861 | 3.460791 |
| ENSMUSG0 | 1436.826 | 1370.93 | 1458.629 | 1614.677 | 1841.875 | 1765.009 | -0.29148 | 0.000127 | 0.002635 | Cic | 7 | 25267704 | 25294159 | + | 10294 | protein_co | capicua tra | HMG | 1442 | 1361 | 1565 | 1550 | 1817 | 1820 | 7.36742 | 7.029538 | 7.479216 | 8.279364 | 9.444339 | 9.050203 |
| ENSMUSG0 | 526.1054 | 568.1152 | 583.4514 | 730.2507 | 724.7886 | 762.2512 | -0.4022 | 0.000127 | 0.002635 | Zfp568 | 7 | 29983955 | 30028282 | + | 7354 | protein_co | zinc finger p | zf-C2H2 | 528 | 564 | 626 | 701 | 715 | 786 | 3.77611 | 4.077634 | 4.18771 | 5.241358 | 5.202154 | 5.471041 |
| ENSMUSG0 | 967.5157 | 1006.289 | 943.2154 | 1176.11 | 1234.675 | 1217.08 | -0.31494 | 0.000128 | 0.002642 | Zfx | X | 94074631 | 94123702 | - | 7755 | protein_co | zinc finger p | zf-C2H2 | 971 | 999 | 1012 | 1129 | 1218 | 1255 | 6.585242 | 6.849147 | 6.419846 | 8.005005 | 8.403618 | 8.283865 |
| ENSMUSG0 | 16596.23 | 16903.44 | 15926.55 | 13941.43 | 15586.5 | 14238.39 | 0.175404 | 0.000128 | 0.002642 | Nedd4 | 9 | 72662346 | 72749852 | + | 10973 | protein_co | neural prec | - | 16656 | 16781 | 17088 | 13383 | 15376 | 14682 | 79.83249 | 81.31024 | 76.61111 | 67.06216 | 74.9754 | 68.49059 |
| ENSMUSG0 | 1493.621 | 1398.127 | 1307.64 | 1675.097 | 1798.287 | 1677.728 | -0.29502 | 0.000128 | 0.002642 | Fbxl3 | 14 | 1.03E+08 | 1.03E+08 | - | 5103 | protein_co | F-box and l | - | 1499 | 1388 | 1403 | 1608 | 1774 | 1730 | 15.44936 | 14.46161 | 13.52565 | 17.32646 | 18.60068 | 17.35368 |
| ENSMUSG0 | 2311.675 | 2300.665 | 2332.874 | 2681.405 | 2841.374 | 2618.42 | -0.22909 | 0.00013 | 0.002683 | Ctnna1 | 18 | 35118888 | 35254773 | + | 3698 | protein_co | catenin (ca | - | 2320 | 2284 | 2503 | 2574 | 2803 | 2700 | 32.99955 | 32.8384 | 33.29813 | 38.27288 | 40.55618 | 37.37386 |
| ENSMUSG0 | 126.5443 | 150.0872 | 138.8726 | 86.46334 | 62.8488 | 50.42883 | 1.059004 | 0.000132 | 0.002721 | Esam | 9 | 37528078 | 37538319 | + | 2867 | protein_co | endothelia | - | 127 | 149 | 149 | 83 | 62 | 52 | 2.329756 | 2.763194 | 2.556728 | 1.591842 | 1.157084 | 0.928425 |
| ENSMUSG0 | 883.8172 | 903.5448 | 842.5561 | 1063.603 | 1140.402 | 1103.616 | -0.33114 | 0.000132 | 0.002722 | Uhrf1bp1l | 10 | 89744991 | 89819871 | + | 9235 | protein_co | UHRF1 (ICB | - | 887 | 897 | 904 | 1021 | 1125 | 1138 | 5.051508 | 5.164263 | 4.815678 | 6.079086 | 6.518031 | 6.307779 |
| ENSMUSG0 | 2876.641 | 3106.502 | 3137.216 | 3459.575 | 3741.531 | 3466.012 | -0.22587 | 0.000134 | 0.002753 | Plxnb2 | 15 | 89155549 | 89180788 | - | 7720 | protein_co | plexin B2 [S | - | 2887 | 3084 | 3366 | 3321 | 3691 | 3574 | 19.66816 | 21.23977 | 21.44977 | 23.65381 | 25.58159 | 23.69782 |
| ENSMUSG0 | 3000.196 | 3411.713 | 3112.052 | 2609.526 | 2752.169 | 2705.701 | 0.239277 | 0.000134 | 0.002755 | Scarf2 | 16 | 17797282 | 17808293 | + | 3839 | protein_co | scavenger r | - | 3011 | 3387 | 3339 | 2505 | 2715 | 2790 | 41.25029 | 46.90832 | 42.78822 | 35.8789 | 37.84012 | 37.20122 |
| ENSMUSG0 | 1056.196 | 981.1067 | 994.4771 | 1167.776 | 1349.222 | 1300.482 | -0.33275 | 0.000137 | 0.002795 | Dcaf10 | 4 | 45342101 | 45379759 | + | 7305 | protein_co | DDB1 and C | - | 1060 | 974 | 1067 | 1121 | 1331 | 1341 | 7.631677 | 7.089107 | 7.185717 | 8.437909 | 9.748968 | 9.396793 |
| ENSMUSG0 | 686.5276 | 624.5238 | 685.0428 | 876.0924 | 894.0748 | 823.3476 | -0.37727 | 0.000137 | 0.002804 | Man2a1 | 17 | 64600736 | 64755110 | + | 7223 | protein_co | mannosida | - | 689 | 620 | 735 | 841 | 882 | 849 | 5.016906 | 4.563803 | 5.006055 | 6.402179 | 6.533589 | 6.016739 |
| ENSMUSG0 | 94.65911 | 119.8683 | 82.01873 | 50.0029 | 29.39702 | 42.67055 | 1.280223 | 0.000138 | 0.002815 | Robo2 | 16 | 73891839 | 74411825 | - | 16983 | protein_co | roundabou | - | 95 | 119 | 88 | 48 | 29 | 44 | 0.294201 | 0.372551 | 0.254914 | 0.155409 | 0.091366 | 0.13262 |
| ENSMUSG0 | 179.3541 | 269.9554 | 288.9296 | 408.357 | 339.5862 | 462.5875 | -0.71301 | 0.000142 | 0.002903 | Krt16 | 11 | 1E+08 | 1E+08 | - | 1565 | protein_co | keratin 16 [ | - | 180 | 268 | 310 | 392 | 335 | 477 | 6.049124 | 9.10486 | 9.744807 | 13.77277 | 11.45332 | 15.60181 |
| ENSMUSG0 | 656.6353 | 524.8014 | 541.51 | 722.9586 | 791.6921 | 829.1663 | -0.44452 | 0.000143 | 0.002913 | Pappa2 | 1 | 1.59E+08 | 1.59E+08 | - | 10294 | protein_co | pappalysin | - | 659 | 521 | 581 | 694 | 781 | 855 | 3.366942 | 2.690955 | 2.776629 | 3.707019 | 4.059455 | 4.251606 |
| ENSMUSG0 | 271.024 | 260.8898 | 211.571 | 161.4677 | 133.8071 | 150.3167 | 0.738039 | 0.000145 | 0.002944 | Ccnd1 | 7 | 1.45E+08 | 1.45E+08 | - | 4709 | protein_co | cyclin D1 [S | - | 272 | 259 | 227 | 155 | 132 | 155 | 3.037908 | 2.924313 | 2.3715 | 1.809891 | 1.499844 | 1.6849 |
| ENSMUSG0 | 9371.252 | 9158.339 | 9585.939 | 8760.924 | 7232.68 | 8153.954 | 0.219574 | 0.000145 | 0.002947 | Mgp | 6 | 1.37E+08 | 1.37E+08 | + | 1187 | protein_co | matrix Gla | - | 9405 | 9092 | 10285 | 8410 | 7135 | 8408 | 416.7182 | 407.2504 | 426.2648 | 389.5783 | 321.6208 | 362.5877 |
| ENSMUSG0 | 1855.319 | 1768.812 | 2037.42 | 2272.007 | 2173.352 | 2412.826 | -0.27639 | 0.000148 | 0.00299 | Tgm2 | 2 | 1.58E+08 | 1.58E+08 | + | 4608 | protein_co | transglutar | - | 1862 | 1756 | 2186 | 2181 | 2144 | 2488 | 21.25208 | 20.26118 | 23.338 | 26.02512 | 24.89506 | 27.63815 |
| ENSMUSG0 | 521.1233 | 433.1374 | 522.8694 | 719.8334 | 671.063 | 633.2697 | -0.45372 | 0.00015 | 0.003036 | Nop2 | 6 | 1.25E+08 | 1.25E+08 | + | 2772 | protein_co | NOP2 nucle | - | 523 | 430 | 561 | 691 | 662 | 653 | 9.922996 | 8.247608 | 9.956244 | 13.70674 | 12.77808 | 12.05844 |
| ENSMUSG0 | 1255.479 | 1274.23 | 1126.826 | 1431.333 | 1591.494 | 1510.925 | -0.31076 | 0.000151 | 0.003052 | Brd1 | 15 | 88687034 | 88734233 | - | 8404 | protein_co | bromodom | - | 1260 | 1265 | 1209 | 1374 | 1570 | 1558 | 7.885311 | 8.003082 | 7.077276 | 8.989803 | 9.995728 | 9.4897 |
| ENSMUSG0 | 234.1567 | 254.846 | 247.9203 | 343.7699 | 349.7231 | 406.34 | -0.5782 | 0.000153 | 0.003077 | Asprv1 | 6 | 86628164 | 86629710 | + | 1547 | protein_co | aspartic pe | - | 235 | 253 | 266 | 330 | 345 | 419 | 7.989358 | 8.695269 | 8.458965 | 11.72933 | 11.93245 | 13.8642 |
| ENSMUSG0 | 651.6532 | 583.2246 | 595.5678 | 322.9354 | 323.3672 | 530.4725 | 0.636112 | 0.000154 | 0.003108 | Mmp13 | 9 | 7272514 | 7283331 | + | 2673 | protein_co | matrix met | - | 654 | 579 | 639 | 310 | 319 | 547 | 12.86806 | 11.51682 | 11.76055 | 6.376938 | 6.385465 | 10.47513 |
| ENSMUSG0 | 1773.613 | 1817.163 | 1793.228 | 1471.96 | 1151.552 | 1563.294 | 0.362628 | 0.000156 | 0.00313 | Tmsb10 | 6 | 72957347 | 72958748 | - | 1276 | protein_co | thymosin, l | - | 1780 | 1804 | 1924 | 1413 | 1136 | 1612 | 73.36749 | 75.16898 | 74.17888 | 60.8893 | 47.63525 | 64.66741 |
| ENSMUSG0 | 66.75958 | 66.48156 | 63.37811 | 29.16836 | 17.23273 | 22.30506 | 1.518131 | 0.000156 | 0.003138 | E130317F2 | 10 | 79851381 | 79854971 | - | 3591 | bidirection | RIKEN cDNA | - | 67 | 66 | 68 | 28 | 17 | 23 | 0.981282 | 0.977195 | 0.931578 | 0.428738 | 0.253299 | 0.327856 |
| ENSMUSG0 | 1048.225 | 1115.077 | 1196.728 | 1379.247 | 1541.823 | 1316.968 | -0.33444 | 0.000157 | 0.003154 | Heatr5a | 12 | 51875871 | 51971321 | - | 20676 | protein_co | HEAT repea | - | 1052 | 1107 | 1284 | 1324 | 1521 | 1358 | 2.675984 | 2.846649 | 3.055093 | 3.52104 | 3.936076 | 3.362051 |
| ENSMUSG0 | 1591.269 | 1633.835 | 1879.907 | 1990.74 | 2190.585 | 2079.219 | -0.29397 | 0.000159 | 0.003183 | Birc6 | 17 | 74528295 | 74703356 | + | 28830 | protein_co | baculoviral | - | 1597 | 1622 | 2017 | 1911 | 2161 | 2144 | 2.913363 | 2.991293 | 3.441812 | 3.644731 | 4.010615 | 3.806722 |
| ENSMUSG0 | 384.6149 | 384.7872 | 388.6569 | 500.029 | 526.1053 | 575.0826 | -0.46784 | 0.000159 | 0.003183 | Pvr | 7 | 19903578 | 19921160 | - | 6122 | protein_co | poliovirus r | - | 386 | 382 | 417 | 480 | 519 | 593 | 3.316105 | 3.317591 | 3.350955 | 4.311192 | 4.536019 | 4.958296 |
| ENSMUSG0 | 2933.436 | 3010.809 | 2982.499 | 3267.898 | 3550.957 | 3514.502 | -0.21125 | 0.00016 | 0.003186 | Eif3c | 7 | 1.27E+08 | 1.27E+08 | - | 3511 | protein_co | eukaryotic | - | 2944 | 2989 | 3200 | 3137 | 3503 | 3624 | 44.10028 | 45.26347 | 44.83788 | 49.12846 | 53.38388 | 52.83582 |
| ENSMUSG0 | 80.70935 | 86.62749 | 64.31014 | 31.25181 | 34.46547 | 22.30506 | 1.39699 | 0.00016 | 0.003187 | Kcp | 6 | 29473162 | 29507952 | - | 7807 | protein_co | kielin/chor | - | 81 | 86 | 69 | 30 | 34 | 23 | 0.545676 | 0.585689 | 0.434801 | 0.211294 | 0.233021 | 0.150805 |
| ENSMUSG0 | 313.8697 | 311.2546 | 321.5507 | 480.2362 | 454.1332 | 414.0983 | -0.50977 | 0.000162 | 0.003225 | Tjp2 | 19 | 24094505 | 24225030 | - | 5335 | protein_co | tight juncti | - | 315 | 309 | 345 | 461 | 448 | 427 | 3.105349 | 3.079476 | 3.181344 | 4.751338 | 4.493083 | 4.096986 |
| ENSMUSG0 | 242.128 | 219.5906 | 251.6484 | 134.3828 | 102.3827 | 169.7124 | 0.810606 | 0.000164 | 0.003255 | Cldn5 | 16 | 18776847 | 18778262 | + | 1416 | protein_co | claudin 5 [S | - | 243 | 218 | 270 | 129 | 101 | 175 | 9.025627 | 8.185516 | 9.380509 | 5.009287 | 3.816444 | 6.326243 |
| ENSMUSG0 | 2398.363 | 2300.665 | 2742.035 | 2929.336 | 2830.223 | 3424.312 | -0.3035 | 0.000167 | 0.003311 | Syne1 | 10 | 5020917 | 5551482 | - | 40491 | protein_co | spectrin rep | - | 2407 | 2284 | 2942 | 2812 | 2792 | 3531 | 3.126453 | 2.999096 | 3.574457 | 3.818619 | 3.689417 | 4.463857 |
| ENSMUSG0 | 95.65552 | 119.8683 | 89.47498 | 43.75254 | 46.62975 | 46.54969 | 1.15431 | 0.000168 | 0.003317 | Rasgrp3 | 17 | 75435905 | 75529043 | + | 4921 | protein_co | RAS, guany | - | 96 | 119 | 96 | 42 | 46 | 48 | 1.026011 | 1.28572 | 0.959718 | 0.469294 | 0.500156 | 0.499297 |
| ENSMUSG0 | 103.6268 | 106.7734 | 102.5234 | 166.6763 | 172.3273 | 216.2621 | -0.82833 | 0.00018 | 0.00356 | Eid2 | 7 | 28267881 | 28269272 | + | 1392 | protein_co | EP300 inte | - | 104 | 106 | 110 | 160 | 170 | 223 | 3.92942 | 4.048736 | 3.88758 | 6.320191 | 6.534472 | 8.200432 |
| ENSMUSG0 | 302.9092 | 259.8825 | 285.2015 | 403.1484 | 385.2023 | 446.1012 | -0.54193 | 0.000184 | 0.003626 | Extl3 | 14 | 65052060 | 65149855 | - | 6063 | protein_co | exostoses ( | - | 304 | 258 | 306 | 387 | 380 | 460 | 2.637062 | 2.262481 | 2.482903 | 3.509723 | 3.353489 | 3.883661 |
| ENSMUSG0 | 217.2177 | 192.3936 | 256.3085 | 142.7166 | 126.7113 | 122.1929 | 0.767378 | 0.000185 | 0.003646 | Plin3 | 17 | 56277476 | 56292873 | - | 3786 | protein_co | perilipin 3 | - | 218 | 191 | 275 | 137 | 125 | 126 | 3.028379 | 2.682289 | 3.57337 | 1.989708 | 1.766567 | 1.703574 |
| ENSMUSG0 | 860.8997 | 818.932 | 819.2553 | 1005.267 | 1059.306 | 1073.552 | -0.32878 | 0.000188 | 0.003698 | Galnt18 | 7 | 1.11E+08 | 1.12E+08 | - | 2575 | protein_co | polypeptid | - | 864 | 813 | 879 | 965 | 1045 | 1107 | 17.647 | 16.78673 | 16.79336 | 20.60628 | 21.714 | 22.00602 |
| ENSMUSG0 | 245.1173 | 197.4301 | 248.8523 | 155.2173 | 122.6565 | 131.8908 | 0.755825 | 0.00019 | 0.003736 | Cd248 | 19 | 5068078 | 5070637 | + | 2560 | protein_co | CD248 anti | - | 246 | 196 | 267 | 149 | 121 | 136 | 5.053933 | 4.070698 | 5.130943 | 3.200337 | 2.528985 | 2.719381 |
| ENSMUSG0 | 260.0634 | 238.7292 | 309.4343 | 456.2764 | 412.5719 | 354.9414 | -0.59691 | 0.00019 | 0.003736 | Ptx3 | 3 | 66219910 | 66225805 | + | 2682 | protein_co | pentraxin r | - | 261 | 237 | 332 | 438 | 407 | 366 | 5.118186 | 4.698318 | 6.089831 | 8.979761 | 8.119634 | 6.985434 |
| ENSMUSG0 | 140.494 | 101.7369 | 122.0961 | 61.4619 | 43.58868 | 65.94539 | 1.090812 | 0.000193 | 0.003789 | Sfrp4 | 13 | 19623104 | 19632994 | + | 4179 | protein_co | secreted fri | - | 141 | 101 | 131 | 59 | 43 | 68 | 1.774521 | 1.284996 | 1.542144 | 0.776299 | 0.55055 | 0.832928 |
| ENSMUSG0 | 4637.3 | 5071.737 | 5356.382 | 5624.284 | 5813.514 | 5975.816 | -0.20888 | 0.000197 | 0.00384 | Crtap | 9 | 1.14E+08 | 1.14E+08 | - | 1772 | protein_co | cartilage as | - | 4654 | 5035 | 5747 | 5399 | 5735 | 6162 | 138.1329 | 151.0736 | 159.5524 | 167.5325 | 173.1691 | 178.0037 |

| gene_id | H1 | H2 | H3 | M1 | M2 | M3 | log2FoldCh | pvalue | padj | gene_name | gene_chr | gene_start | gene_end | gene_stran | gene_lengtl | gene_bioty | gene_descr | tf_family | H1_readcoı | H2_readcoı | H3_readcoı | M1_readco | M2_readco | M3_readco | H1_fpkm | H2_fpkm | H3_fpkm | M1_fpkm | M2_fpkm | M3_fpkm |
|---|---|---|---|---|---|---|---|---|---|---|---|---|---|---|---|---|---|---|---|---|---|---|---|---|---|---|---|---|---|---|
| ENSMUSG0 | 64.76676 | 84.6129 | 55.92186 | 13.54245 | 26.35595 | 29.09356 | 1.568971 | 0.000197 | 0.00384 | Arl5c | 11 | 97989578 | 97996181 | - | 2073 | protein_co | ADP-ribosy | - | 65 | 84 | 60 | 13 | 26 | 30 | 1.649105 | 2.154432 | 1.423895 | 0.344821 | 0.671081 | 0.740786 |
| ENSMUSG0 | 1744.717 | 1769.82 | 1846.353 | 2010.533 | 2247.351 | 2143.225 | -0.25579 | 0.000197 | 0.00384 | Hipk3 | 2 | 1.04E+08 | 1.04E+08 | - | 7987 | protein_co | homeodom | - | 1751 | 1757 | 1981 | 1930 | 2217 | 2210 | 11.5302 | 11.69609 | 12.20188 | 13.28688 | 14.85193 | 14.1638 |
| ENSMUSG0 | 4666.196 | 4629.534 | 4662.019 | 5440.94 | 4971.137 | 5561.718 | -0.19469 | 0.000197 | 0.003846 | Actn4 | 7 | 28893248 | 28962340 | - | 6127 | protein_co | actinin alpl | - | 4683 | 4596 | 5002 | 5223 | 4904 | 5735 | 40.19857 | 39.88274 | 40.16259 | 46.87288 | 42.82559 | 47.91336 |
| ENSMUSG0 | 2068.551 | 2125.395 | 2043.012 | 1826.147 | 1742.534 | 1694.215 | 0.245112 | 0.000199 | 0.003864 | Slc25a37 | 14 | 69241848 | 69285112 | - | 6021 | protein_co | solute carri | - | 2076 | 2110 | 2192 | 1753 | 1719 | 1747 | 18.13398 | 18.63231 | 17.91009 | 16.00895 | 15.27594 | 14.85236 |
| ENSMUSG0 | 2911.515 | 2490.037 | 2758.812 | 2230.338 | 2164.229 | 2400.218 | 0.264119 | 0.000201 | 0.003905 | C1qtnf6 | 15 | 78523346 | 78531416 | - | 2702 | protein_co | C1q and tu | - | 2922 | 2472 | 2960 | 2141 | 2135 | 2475 | 56.87602 | 48.6425 | 53.89299 | 43.56932 | 42.27789 | 46.88792 |
| ENSMUSG0 | 2012.752 | 1945.089 | 2107.322 | 2409.515 | 2778.525 | 2240.204 | -0.29216 | 0.000202 | 0.003924 | Supt6 | 11 | 78206746 | 78245987 | - | 7095 | protein_co | suppressor | - | 2020 | 1931 | 2261 | 2313 | 2741 | 2310 | 14.97384 | 14.47047 | 15.6774 | 17.92556 | 20.67081 | 16.66597 |
| ENSMUSG0 | 854.9212 | 891.4573 | 756.8092 | 1087.563 | 1049.169 | 1047.368 | -0.34763 | 0.000203 | 0.003933 | Timm29 | 9 | 21592722 | 21595970 | + | 3170 | protein_co | translocase | - | 858 | 885 | 812 | 1044 | 1035 | 1080 | 14.23516 | 14.84352 | 12.60151 | 18.10884 | 17.46956 | 17.43956 |
| ENSMUSG0 | 36.86723 | 36.26267 | 17.70859 | 6.250362 | 6.082142 | 1.93957 | 2.676171 | 0.000204 | 0.003937 | Anxa11os | 14 | 25856777 | 25870266 | - | 2479 | antisense | annexin A1 | - | 37 | 36 | 19 | 6 | 6 | 2 | 0.784982 | 0.772109 | 0.377053 | 0.133083 | 0.129502 | 0.041298 |
| ENSMUSG0 | 76.7237 | 50.36482 | 73.63045 | 33.33527 | 8.109522 | 17.45613 | 1.772517 | 0.000211 | 0.004074 | Vash1 | 12 | 86678700 | 86692091 | + | 2599 | protein_co | vasohibin 1 | - | 77 | 50 | 79 | 32 | 8 | 18 | 1.558184 | 1.022861 | 1.495363 | 0.677007 | 0.164697 | 0.354517 |
| ENSMUSG0 | 281.9845 | 274.9919 | 244.1921 | 397.9397 | 390.2708 | 374.3371 | -0.53759 | 0.000214 | 0.004132 | Mir22hg | 11 | 75461539 | 75466676 | + | 1924 | processed_ | Mir22 host | - | 283 | 273 | 262 | 382 | 385 | 386 | 7.735987 | 7.544151 | 6.699187 | 10.91711 | 10.70672 | 10.2696 |
| ENSMUSG0 | 1188.719 | 1199.69 | 1201.388 | 1405.29 | 1443.495 | 1505.107 | -0.27859 | 0.000216 | 0.004163 | Fmnl3 | 15 | 99317225 | 99370482 | - | 5021 | protein_co | formin-like | - | 1193 | 1191 | 1289 | 1349 | 1424 | 1552 | 12.49639 | 12.61172 | 12.62957 | 14.77308 | 15.17472 | 15.82241 |
| ENSMUSG0 | 5126.538 | 5273.197 | 5426.285 | 4570.057 | 4731.906 | 4720.914 | 0.174555 | 0.00022 | 0.004234 | Gnai2 | 9 | 1.08E+08 | 1.08E+08 | - | 3779 | protein_co | guanine nu | - | 5145 | 5235 | 5822 | 4387 | 4668 | 4868 | 71.60491 | 73.65336 | 75.79162 | 63.83226 | 66.09289 | 65.93936 |
| ENSMUSG0 | 1796.53 | 1849.396 | 2103.594 | 2320.968 | 2272.649 | 2313.907 | -0.26429 | 0.000224 | 0.004289 | Kdm6b | 11 | 69398508 | 69413675 | - | 6954 | protein_co | KDM1 lysin | - | 1803 | 1836 | 2257 | 2228 | 2242 | 2386 | 13.63626 | 14.03753 | 15.96698 | 17.61692 | 17.2505 | 17.56333 |
| ENSMUSG0 | 3131.722 | 3287.815 | 3376.748 | 2704.323 | 2978.222 | 2695.033 | 0.22586 | 0.000226 | 0.004322 | Gtf2i | 5 | 1.34E+08 | 1.34E+08 | - | 10199 | protein_co | general trai | GTF2I | 3143 | 3264 | 3623 | 2596 | 2938 | 2779 | 16.20769 | 17.01553 | 17.47578 | 13.99576 | 15.41328 | 13.94768 |
| ENSMUSG0 | 1508.567 | 1602.609 | 1675.792 | 1929.278 | 2157.133 | 1796.042 | -0.29693 | 0.000226 | 0.004322 | Abhd2 | 7 | 79273199 | 79365508 | + | 10573 | protein_co | abhydrolas | - | 1514 | 1591 | 1798 | 1852 | 2128 | 1852 | 7.531162 | 8.00064 | 8.36599 | 9.631462 | 10.76897 | 8.966312 |
| ENSMUSG0 | 560.9798 | 577.1808 | 558.2866 | 712.5413 | 724.7886 | 772.9188 | -0.38207 | 0.000226 | 0.004322 | Tent4a | 13 | 69497959 | 69534617 | - | 8918 | protein_co | terminal nu | - | 563 | 573 | 599 | 684 | 715 | 797 | 3.320284 | 3.416174 | 3.304344 | 4.217335 | 4.289823 | 4.574693 |
| ENSMUSG0 | 487.2453 | 375.7215 | 401.7054 | 596.9096 | 565.6392 | 596.4179 | -0.47616 | 0.000229 | 0.004364 | Sh2b3 | 5 | 1.22E+08 | 1.22E+08 | - | 4807 | protein_co | SH2B adapt | - | 489 | 373 | 431 | 573 | 558 | 615 | 5.350188 | 4.125604 | 4.410919 | 6.554355 | 6.210991 | 6.548956 |
| ENSMUSG0 | 1049.222 | 1131.194 | 1157.583 | 1318.826 | 1536.754 | 1324.727 | -0.3244 | 0.00023 | 0.004386 | Trim35 | 14 | 66297031 | 66311424 | + | 4618 | protein_co | tripartite m | - | 1053 | 1123 | 1242 | 1266 | 1516 | 1366 | 11.99247 | 12.92941 | 13.23103 | 15.07403 | 17.56492 | 15.14146 |
| ENSMUSG0 | 1585.291 | 1827.236 | 1651.559 | 2055.327 | 1898.642 | 2296.451 | -0.30401 | 0.000238 | 0.004535 | Oaf | 9 | 43221235 | 43239911 | - | 2629 | protein_co | out at first | - | 1591 | 1814 | 1772 | 1973 | 1873 | 2368 | 31.82834 | 36.68593 | 33.15882 | 41.26539 | 38.11957 | 46.1065 |
| ENSMUSG0 | 1624.151 | 1640.886 | 1743.83 | 2077.204 | 1902.697 | 1998.727 | -0.25501 | 0.00024 | 0.004567 | Amotl1 | 9 | 14541966 | 14645056 | - | 10064 | protein_co | angiomotir | - | 1630 | 1629 | 1871 | 1994 | 1877 | 2061 | 8.518268 | 8.606038 | 9.145955 | 10.89442 | 9.979171 | 10.48283 |
| ENSMUSG0 | 467.3171 | 452.2761 | 394.2491 | 302.1008 | 262.5458 | 335.5457 | 0.544508 | 0.000245 | 0.004635 | Abca9 | 11 | 1.1E+08 | 1.1E+08 | - | 8048 | protein_co | ATP-bindin | - | 469 | 449 | 423 | 290 | 259 | 346 | 3.06492 | 2.966273 | 2.585701 | 1.981342 | 1.721919 | 2.200692 |
| ENSMUSG0 | 1237.543 | 1107.019 | 1216.3 | 799.0046 | 1059.306 | 858.2599 | 0.390495 | 0.000245 | 0.004635 | Sh3bgrl | X | | 1.09E+08 | 1.09E+08 | + | 3298 | protein_co | SH3-bindin | - | 1242 | 1099 | 1305 | 767 | 1045 | 885 | 19.80639 | 17.71739 | 19.46641 | 12.78775 | 16.95378 | 13.73611 |
| ENSMUSG0 | 93.6627 | 63.45967 | 96.93123 | 130.2159 | 200.7107 | 165.8333 | -0.96649 | 0.000245 | 0.004635 | Nptx2 | 5 | 1.45E+08 | 1.45E+08 | + | 2536 | protein_co | neuronal p | - | 94 | 63 | 104 | 125 | 198 | 171 | 1.949454 | 1.320821 | 2.017483 | 2.710255 | 4.177503 | 3.45158 |
| ENSMUSG0 | 4102.227 | 3825.712 | 3978.84 | 4850.281 | 4451.114 | 4398.946 | -0.20224 | 0.000251 | 0.004744 | Ugdh | 5 | 65413221 | 65435949 | - | 6752 | protein_co | UDP-glucos | - | 4117 | 3798 | 4269 | 4656 | 4391 | 4536 | 32.0688 | 29.90717 | 31.10424 | 37.91665 | 34.7962 | 34.38838 |
| ENSMUSG0 | 1162.812 | 1189.617 | 1191.136 | 1552.173 | 1307.66 | 1543.898 | -0.31353 | 0.000252 | 0.004753 | Tbc1d20 | 2 | 1.52E+08 | 1.52E+08 | + | 4171 | protein_co | TBC1 doma | - | 1167 | 1181 | 1278 | 1490 | 1290 | 1592 | 14.71516 | 15.05437 | 15.07359 | 19.64244 | 16.54818 | 19.53772 |
| ENSMUSG0 | 11819.44 | 11736.01 | 11319.52 | 10390.19 | 10668.08 | 10604.6 | 0.1393 | 0.000254 | 0.004777 | Fstl1 | 16 | 37776873 | 37836514 | + | 4914 | protein_co | follistatin-l | - | 11862 | 11651 | 12145 | 9974 | 10524 | 10935 | 126.9571 | 126.061 | 121.5873 | 111.605 | 114.5899 | 113.9081 |
| ENSMUSG0 | 802.1114 | 791.7349 | 804.3428 | 659.4132 | 626.4606 | 551.8078 | 0.384719 | 0.000264 | 0.004969 | Cpz | 5 | 35502218 | 35525698 | - | 2245 | protein_co | carboxypep | - | 805 | 786 | 863 | 633 | 618 | 569 | 18.85879 | 18.61483 | 18.91125 | 15.50375 | 14.72899 | 12.97379 |
| ENSMUSG0 | 2152.249 | 2163.673 | 2158.584 | 2500.145 | 2445.021 | 2577.689 | -0.21661 | 0.000267 | 0.00501 | Myh9 | 15 | 77760587 | 77842175 | - | 12928 | protein_co | myosin, he | - | 2160 | 2148 | 2316 | 2400 | 2412 | 2658 | 8.787326 | 8.833966 | 8.81319 | 10.20773 | 9.982671 | 10.52434 |
| ENSMUSG0 | 146.4725 | 128.9339 | 170.5617 | 62.50362 | 75.01308 | 93.09938 | 0.950485 | 0.000267 | 0.00501 | Rasd1 | 11 | 59963181 | 59964944 | - | 1611 | protein_co | RAS, dexam | - | 147 | 128 | 183 | 60 | 74 | 96 | 4.799059 | 4.224421 | 5.588322 | 2.047883 | 2.457746 | 3.05033 |
| ENSMUSG0 | 344.7584 | 375.7215 | 342.0554 | 475.0275 | 568.6802 | 456.7688 | -0.49801 | 0.000271 | 0.005071 | Hipk2 | 6 | 38694390 | 38876165 | - | 8836 | protein_co | homeodom | - | 346 | 373 | 367 | 456 | 561 | 471 | 2.059466 | 2.244429 | 2.043319 | 2.837648 | 3.397097 | 2.728577 |
| ENSMUSG0 | 3314.065 | 3373.436 | 3138.148 | 3773.135 | 3764.846 | 3694.882 | -0.19327 | 0.000273 | 0.005105 | Cdv3 | 9 | 1.03E+08 | 1.03E+08 | - | 7403 | protein_co | carnitine d | - | 3326 | 3349 | 3367 | 3622 | 3714 | 3810 | 23.62919 | 24.0525 | 22.37491 | 26.90235 | 26.84324 | 26.3444 |
| ENSMUSG0 | 87.68423 | 94.68586 | 167.7656 | 242.7224 | 200.7107 | 215.2923 | -0.90891 | 0.000276 | 0.00515 | Sfn | 4 | 1.34E+08 | 1.34E+08 | - | 1613 | protein_co | stratifin [Sc | - | 88 | 94 | 180 | 233 | 198 | 222 | 2.869344 | 3.098463 | 5.489895 | 7.942752 | 6.567977 | 7.045141 |
| ENSMUSG0 | 1889.197 | 1788.958 | 1613.346 | 2006.366 | 2196.667 | 2196.563 | -0.27484 | 0.000277 | 0.00515 | Cab39 | 1 | 85793441 | 85851576 | + | 7077 | protein_co | calcium bir | - | 1896 | 1776 | 1731 | 1926 | 2167 | 2265 | 14.0904 | 13.34279 | 12.033 | 14.9643 | 16.38364 | 16.38287 |
| ENSMUSG0 | 116.5802 | 57.41589 | 117.4359 | 42.71081 | 35.47916 | 39.76119 | 1.306239 | 0.000277 | 0.00515 | Ncald | 15 | 37366175 | 37792570 | - | 8581 | protein_co | neurocalcir | - | 117 | 57 | 126 | 41 | 35 | 41 | 0.717104 | 0.353175 | 0.722368 | 0.262721 | 0.218238 | 0.244578 |
| ENSMUSG0 | 852.9284 | 803.8225 | 902.206 | 1161.526 | 972.129 | 1146.286 | -0.35779 | 0.000277 | 0.005151 | Epha2 | 4 | 1.41E+08 | 1.41E+08 | + | 4674 | protein_co | Eph recept | - | 856 | 798 | 968 | 1115 | 959 | 1182 | 9.632065 | 9.077515 | 10.18855 | 13.11703 | 10.97819 | 12.94493 |
| ENSMUSG0 | 392.5862 | 357.5902 | 331.803 | 506.2793 | 502.7904 | 487.802 | -0.46871 | 0.00028 | 0.005198 | Samd8 | 14 | 21750531 | 21798726 | + | 7639 | protein_co | sterile alph | - | 394 | 355 | 356 | 486 | 496 | 503 | 2.712652 | 2.47084 | 2.292659 | 3.498237 | 3.474129 | 3.370564 |
| ENSMUSG0 | 204.2644 | 166.2039 | 233.0078 | 84.37989 | 145.9714 | 90.19002 | 0.913552 | 0.00028 | 0.005198 | Ift81 | 5 | 1.23E+08 | 1.23E+08 | - | 5840 | protein_co | intraflagell | - | 205 | 165 | 250 | 81 | 144 | 93 | 1.846186 | 1.502187 | 2.105974 | 0.762644 | 1.319321 | 0.815157 |
| ENSMUSG0 | 1397.966 | 1385.033 | 1512.686 | 1832.398 | 1627.987 | 1748.523 | -0.2777 | 0.000282 | 0.005221 | G3bp1 | 11 | 55469685 | 55504838 | + | 7064 | protein_co | GTPase acti | - | 1403 | 1375 | 1623 | 1759 | 1606 | 1803 | 10.44579 | 10.34915 | 11.303 | 13.69193 | 12.16454 | 13.0652 |
| ENSMUSG0 | 2325.625 | 2383.263 | 2019.711 | 2610.568 | 2713.649 | 2696.973 | -0.25399 | 0.000283 | 0.005231 | Timp3 | 10 | 86300372 | 86349506 | + | 8144 | protein_co | tissue inhib | - | 2334 | 2366 | 2167 | 2506 | 2677 | 2781 | 15.07292 | 15.44649 | 13.09022 | 16.9197 | 17.5878 | 17.47971 |
| ENSMUSG0 | 263.0527 | 224.6271 | 230.2117 | 453.1513 | 317.2851 | 339.4248 | -0.62809 | 0.000285 | 0.005254 | Zbtb7b | 3 | 89377644 | 89394776 | - | 5591 | protein_co | zinc finger | ZBTB | 264 | 223 | 247 | 435 | 313 | 350 | 2.483412 | 2.120646 | 2.173368 | 4.278083 | 2.995406 | 3.204421 |
| ENSMUSG0 | 276.006 | 317.2984 | 340.1913 | 414.6074 | 466.2975 | 451.9199 | -0.51337 | 0.000288 | 0.005311 | Tfam | 10 | 71225464 | 71238280 | - | 4438 | protein_co | transcripti | HMG | 277 | 315 | 365 | 398 | 460 | 466 | 3.282667 | 3.773775 | 4.046052 | 4.931116 | 5.545891 | 5.374891 |
| ENSMUSG0 | 1647.069 | 1618.725 | 1686.976 | 1974.073 | 2082.12 | 1854.229 | -0.25471 | 0.000295 | 0.005423 | Spire1 | 18 | 67488209 | 67610790 | - | 7766 | protein_co | spire type a | - | 1653 | 1607 | 1810 | 1895 | 2054 | 1912 | 11.19463 | 11.00199 | 11.46587 | 13.41718 | 14.15155 | 12.60264 |
| ENSMUSG0 | 270.0276 | 186.3498 | 258.1726 | 153.1339 | 99.34165 | 151.2865 | 0.823819 | 0.000296 | 0.005434 | Gimap6 | 6 | 48701582 | 48708225 | - | 3496 | protein_co | GTPase, IM | - | 271 | 185 | 277 | 147 | 98 | 156 | 4.07692 | 2.81354 | 3.897932 | 2.31204 | 1.499876 | 2.284148 |
| ENSMUSG0 | 709.4451 | 718.2023 | 732.5764 | 487.5283 | 611.2552 | 520.7747 | 0.41556 | 0.0003 | 0.005502 | Snx2 | 18 | 53176365 | 53220860 | + | 2013 | protein_co | sorting nex | - | 712 | 713 | 786 | 468 | 603 | 537 | 18.60247 | 18.83209 | 19.209 | 12.78355 | 16.02782 | 13.65531 |
| ENSMUSG0 | 324.8302 | 393.8529 | 371.8804 | 511.488 | 529.1463 | 477.1343 | -0.4766 | 0.000303 | 0.005551 | Bicral | 17 | 46798120 | 46858482 | - | 6851 | protein_co | BRD4 inter | - | 326 | 391 | 399 | 491 | 522 | 492 | 2.502638 | 3.034419 | 2.865133 | 3.940733 | 4.076781 | 3.676057 |
| ENSMUSG0 | 166.4008 | 149.0799 | 142.6007 | 228.1382 | 238.2172 | 263.7816 | -0.6735 | 0.000303 | 0.005551 | Fhl3 | 4 | 1.25E+08 | 1.25E+08 | + | 1683 | protein_co | four and a l | - | 167 | 148 | 153 | 219 | 235 | 272 | 5.218752 | 4.675525 | 4.472324 | 7.154997 | 7.4711 | 8.272864 |
| ENSMUSG0 | 422.4786 | 326.364 | 422.2101 | 553.1571 | 515.9684 | 577.0222 | -0.49079 | 0.000309 | 0.005638 | Bdkrb2 | 12 | 1.06E+08 | 1.06E+08 | + | 3792 | protein_co | bradykinin | - | 424 | 324 | 453 | 531 | 509 | 595 | 5.880738 | 4.542861 | 5.877001 | 7.699733 | 7.18208 | 8.031926 |
| ENSMUSG0 | 802.1114 | 731.2972 | 830.4396 | 1077.106 | 928.5403 | 1015.365 | -0.35343 | 0.000309 | 0.005638 | Smim3 | 18 | 60474193 | 60501983 | - | 1984 | protein_co | small integ | - | 805 | 726 | 891 | 1034 | 916 | 1047 | 21.33971 | 19.45574 | 22.09337 | 28.65684 | 24.70328 | 27.0132 |
| ENSMUSG0 | 2414.306 | 2422.548 | 2523.94 | 2997.049 | 2923.483 | 2684.365 | -0.22491 | 0.00031 | 0.005647 | Clstn1 | 4 | 1.5E+08 | 1.5E+08 | + | 4662 | protein_co | calsyntenir | - | 2423 | 2405 | 2708 | 2877 | 2884 | 2768 | 27.33477 | 27.42809 | 28.57606 | 33.93259 | 33.09968 | 30.39239 |
| ENSMUSG0 | 11.95694 | 35.25537 | 45.66952 | 2.083454 | 7.095832 | 4.848926 | 2.725029 | 0.00031 | 0.005647 | Foxd3 | 4 | 99656299 | 99658622 | + | 2324 | protein_co | forkhead b | Fork | 12 | 35 | 49 | 2 | 7 | 5 | 0.271569 | 0.800727 | 1.037256 | 0.04732 | 0.161162 | 0.11013 |
| ENSMUSG0 | 1006.376 | 1004.274 | 968.3802 | 1255.281 | 1169.799 | 1229.688 | -0.29507 | 0.000314 | 0.005714 | Mgat1 | 11 | 49244191 | 49263030 | + | 4733 | protein_co | mannoside | - | 1010 | 997 | 1039 | 1205 | 1154 | 1268 | 11.22326 | 11.19983 | 10.79953 | 13.9991 | 13.04578 | 13.71367 |
| ENSMUSG0 | 319.8482 | 363.634 | 356.0359 | 521.9052 | 520.0231 | 426.7055 | -0.49789 | 0.000321 | 0.005825 | Rasa2 | 9 | 96539300 | 96631617 | - | 15856 | protein_co | RAS p21 pr | - | 321 | 361 | 382 | 501 | 513 | 440 | 1.064745 | 1.210504 | 1.185211 | 1.737375 | 1.731109 | 1.420464 |
| ENSMUSG0 | 63.77035 | 36.26267 | 35.41718 | 15.62591 | 11.15059 | 9.697852 | 1.894964 | 0.000325 | 0.005899 | Exoc3l4 | 12 | 1.11E+08 | 1.11E+08 | + | 5650 | protein_co | exocyst cor | - | 64 | 36 | 38 | 15 | 11 | 10 | 0.595752 | 0.338771 | 0.330873 | 0.14598 | 0.104171 | 0.090599 |
| ENSMUSG0 | 1631.126 | 1555.266 | 1502.434 | 2027.201 | 1693.876 | 2029.76 | -0.29475 | 0.000327 | 0.005913 | Sik3 | 9 | 46012820 | 46224194 | + | 9318 | protein_co | SIK family k | - | 1637 | 1544 | 1612 | 1946 | 1671 | 2093 | 9.239752 | 8.81003 | 8.510758 | 11.48338 | 9.595212 | 11.49788 |
| ENSMUSG0 | 161.4187 | 133.9704 | 139.8047 | 264.5987 | 211.8613 | 230.8089 | -0.70044 | 0.000331 | 0.005975 | Eno2 | 6 | 1.25E+08 | 1.25E+08 | - | 4705 | protein_co | enolase 2, | - | 162 | 133 | 150 | 254 | 209 | 238 | 1.81088 | 1.502951 | 1.568403 | 2.968408 | 2.376772 | 2.589336 |
| ENSMUSG0 | 4513.745 | 4629.534 | 4651.767 | 3716.882 | 4239.253 | 4044.004 | 0.201001 | 0.000331 | 0.005983 | Adgrg6 | 10 | 14402583 | 14545659 | - | 12228 | protein_co | adhesion G | - | 4530 | 4596 | 4991 | 3568 | 4182 | 4170 | 19.48395 | 19.98377 | 20.07974 | 16.04423 | 18.29909 | 17.45628 |
| ENSMUSG0 | 174.372 | 157.1382 | 164.0375 | 79.17125 | 94.2732 | 104.7368 | 0.831773 | 0.000333 | 0.005995 | Tfpi | 2 | 84432855 | 84476775 | - | 5463 | protein_co | tissue facto | - | 175 | 156 | 176 | 76 | 93 | 108 | 1.684772 | 1.51826 | 1.58492 | 0.764948 | 0.910862 | 1.011961 |
| ENSMUSG0 | 1998.802 | 1905.805 | 1828.645 | 2262.631 | 2391.295 | 2146.135 | -0.24625 | 0.000335 | 0.006039 | Tmem263 | 10 | 85102627 | 85120618 | + | 4014 | protein_co | transmemb | - | 2006 | 1892 | 1962 | 2172 | 2359 | 2213 | 26.28378 | 25.06089 | 24.04626 | 29.75308 | 31.44498 | 28.22118 |
| ENSMUSG0 | 765.2442 | 759.5015 | 810.867 | 950.0551 | 1009.636 | 967.8456 | -0.32562 | 0.000342 | 0.006141 | Ino80 | 2 | 1.19E+08 | 1.19E+08 | - | 6803 | protein_co | INO80 com | - | 768 | 754 | 870 | 912 | 996 | 998 | 5.937383 | 5.892827 | 6.291362 | 7.371296 | 7.833569 | 7.509329 |
| ENSMUSG0 | 1050.218 | 949.8805 | 986.0888 | 1120.898 | 1256.976 | 1432.373 | -0.35208 | 0.000343 | 0.006152 | Sc5d | 9 | 42251595 | 42264256 | - | 4866 | protein_co | sterol-C5-d | - | 1054 | 943 | 1058 | 1076 | 1240 | 1477 | 11.39208 | 10.30368 | 10.69645 | 12.15877 | 13.63485 | 15.53744 |
| ENSMUSG0 | 1282.382 | 1250.055 | 1287.135 | 1437.583 | 1715.164 | 1533.23 | -0.29496 | 0.000347 | 0.006223 | D630045J1 | 6 | 38123174 | 38254009 | - | 13450 | protein_co | RIKEN cDN | - | 1287 | 1241 | 1381 | 1380 | 1692 | 1581 | 5.032579 | 4.905715 | 5.051232 | 5.641652 | 6.730989 | 6.017009 |
| ENSMUSG0 | 476.2848 | 452.2761 | 407.2976 | 345.8534 | 290.9291 | 304.5126 | 0.505032 | 0.000353 | 0.006313 | Acadl | 1 | 66830839 | 66863277 | - | 2005 | protein_co | acyl-Coenz | - | 478 | 449 | 437 | 332 | 287 | 314 | 12.53857 | 11.90652 | 10.72242 | 9.104858 | 7.658934 | 8.016529 |
| ENSMUSG0 | 6337.178 | 6533.324 | 6335.015 | 6817.062 | 7209.365 | 7908.598 | -0.19192 | 0.000353 | 0.006313 | Col10a1 | 10 | 34389981 | 34397085 | + | 3130 | protein_co | collagen, ty | - | 6360 | 6486 | 6797 | 6544 | 7112 | 8155 | 106.8679 | 110.1756 | 106.8314 | 114.9604 | 121.5761 | 133.3677 |
| ENSMUSG0 | 528.0982 | 510.6993 | 524.7335 | 702.124 | 631.529 | 718.6108 | -0.39246 | 0.000357 | 0.006368 | Retreg1 | 15 | 25843180 | 25973687 | + | 4142 | protein_co | reticulopha | - | 530 | 507 | 563 | 674 | 623 | 741 | 6.729768 | 6.508047 | 6.68689 | 8.94745 | 8.047829 | 9.157548 |
| ENSMUSG0 | 2056.594 | 2090.14 | 1928.372 | 2289.716 | 2499.76 | 2358.518 | -0.23493 | 0.000358 | 0.00638 | Ppp3ca | 3 | 1.37E+08 | 1.37E+08 | + | 9981 | protein_co | protein ph | - | 2064 | 2075 | 2069 | 2198 | 2466 | 2432 | 10.87602 | 11.05342 | 10.19794 | 12.10886 | 13.21965 | 12.4727 |
| ENSMUSG0 | 15.94259 | 15.10945 | 11.18437 | 56.25326 | 46.62975 | 37.82162 | -1.73761 | 0.000358 | 0.006383 | Gm20036 | 12 | 1.02E+08 | 1.02E+08 | + | 844 | processed_ | predicted | - | 16 | 15 | 12 | 54 | 46 | 39 | 0.997038 | 0.944934 | 0.699463 | 3.51804 | 2.916193 | 2.365338 |
| ENSMUSG0 | 77.72011 | 84.6129 | 95.06717 | 28.12663 | 46.62975 | 34.91227 | 1.23076 | 0.00036 | 0.00639 | Syt8 | 7 | 1.42E+08 | 1.42E+08 | + | 1894 | protein_co | synaptotag | - | 78 | 84 | 102 | 27 | 46 | 36 | 2.165953 | 2.358045 | 2.649391 | 0.783851 | 1.299507 | 0.972957 |
| ENSMUSG0 | 13245.3 | 13541.09 | 13158.41 | 11210.02 | 12858.66 | 11063.31 | 0.185231 | 0.00036 | 0.00639 | Lum | 10 | 97565128 | 97572703 | + | 2448 | protein_co | lumican [Sc | - | 13293 | 13443 | 14118 | 10761 | 12685 | 11408 | 285.5919 | 291.9696 | 283.7185 | 241.7078 | 277.2553 | 238.5444 |
| ENSMUSG0 | 398.5647 | 468.3928 | 454.8311 | 327.1023 | 329.4493 | 256.9931 | 0.533889 | 0.000367 | 0.006507 | Ptprd | 4 | 75941238 | 78211961 | - | 22822 | protein_co | protein tyr | - | 400 | 465 | 488 | 314 | 325 | 265 | 0.921808 | 1.083308 | 1.051943 | 0.756529 | 0.761957 | 0.594379 |
| ENSMUSG0 | 110.6017 | 109.7953 | 119.3 | 63.54535 | 52.71189 | 54.30797 | 0.994967 | 0.000369 | 0.006532 | Hic1 | 11 | 75164565 | 75169519 | - | 3750 | protein_co | hypermeth | ZBTB | 111 | 109 | 128 | 61 | 52 | 56 | 1.556776 | 1.545425 | 1.679208 | 0.894433 | 0.741947 | 0.764413 |
| ENSMUSG0 | 1998.802 | 1917.892 | 2081.225 | 2305.342 | 2273.707 | 2489.439 | -0.23691 | 0.00037 | 0.006532 | Tmem43 | 6 | 91473703 | 91488463 | + | 3973 | protein_co | transmemb | - | 2006 | 1904 | 2233 | 2213 | 2243 | 2567 | 26.55502 | 25.4801 | 27.65006 | 30.62755 | 30.20727 | 33.07337 |
| ENSMUSG0 | 3181.543 | 3207.232 | 3271.429 | 3499.161 | 3833.777 | 3734.643 | -0.19626 | 0.00037 | 0.006532 | Tgoln1 | 6 | 72608432 | 72617000 | - | 5013 | protein_co | trans-golgi | - | 3193 | 3184 | 3510 | 3359 | 3782 | 3851 | 33.49929 | 33.76977 | 34.44572 | 36.84357 | 40.36683 | 39.32302 |
| ENSMUSG0 | 1353.127 | 1269.193 | 1423.211 | 1681.347 | 1569.193 | 1632.148 | -0.27093 | 0.000374 | 0.006598 | Mdfi | 17 | 47815328 | 47834691 | - | 3092 | protein_co | MyoD famil | - | 1358 | 1260 | 1527 | 1614 | 1548 | 1683 | 23.09908 | 21.66626 | 24.29548 | 28.70209 | 26.78751 | 27.86222 |
| ENSMUSG0 | 786.1688 | 622.5092 | 728.8483 | 908.386 | 961.9921 | 903.8398 | -0.37587 | 0.000375 | 0.006598 | Ero1l | 14 | 45283087 | 45318771 | - | 5285 | protein_co | ERO1-like ( | - | 789 | 618 | 782 | 872 | 949 | 932 | 7.851748 | 6.217221 | 7.279267 | 9.072374 | 9.607757 | 9.02697 |
| ENSMUSG0 | 11261.44 | 12418.96 | 12482.69 | 10741.25 | 10964.07 | 10623.03 | 0.161787 | 0.000375 | 0.006598 | Cilp2 | 8 | 69880369 | 69887687 | - | 4314 | protein_co | cartilage in | - | 11302 | 12329 | 13393 | 10311 | 10816 | 10954 | 137.7874 | 151.9499 | 152.7297 | 131.4226 | 134.149 | 129.9762 |
| ENSMUSG0 | 876.8423 | 942.8294 | 959.0599 | 1085.48 | 1242.784 | 1140.467 | -0.31988 | 0.000378 | 0.006644 | Idi1 | 13 | 8885501 | 8892451 | + | 3505 | protein_co | isopenteny | - | 880 | 936 | 1029 | 1042 | 1226 | 1176 | 13.20471 | 14.19844 | 14.44286 | 16.34666 | 18.71558 | 17.17475 |
| ENSMUSG0 | 638.6999 | 603.3705 | 619.8006 | 751.0852 | 961.9921 | 763.221 | -0.41137 | 0.000381 | 0.006694 | Foxn2 | 17 | 88440711 | 88490533 | + | 7357 | protein_co | forkhead b | Fork | 641 | 599 | 665 | 721 | 949 | 787 | 4.582385 | 4.328913 | 4.446792 | 5.3887 | 6.901862 | 5.475768 |
| ENSMUSG0 | 669.5887 | 691.0053 | 797.8186 | 904.2191 | 1003.553 | 884.4441 | -0.37072 | 0.000382 | 0.006694 | Crebrf | 17 | 26715650 | 26776635 | + | 8564 | protein_co | CREB3 regu | - | 672 | 686 | 856 | 868 | 990 | 912 | 4.126929 | 4.258928 | 4.917258 | 5.573045 | 6.18528 | 5.451164 |
| ENSMUSG0 | 568.9511 | 527.8233 | 518.2093 | 400.0232 | 399.394 | 400.5213 | 0.428358 | 0.000382 | 0.006694 | Mllt10 | 2 | 18055237 | 18212388 | + | 5907 | protein_co | myeloid/ly | - | 571 | 524 | 556 | 384 | 394 | 413 | 5.083976 | 4.71647 | 4.630562 | 3.574487 | 3.568865 | 3.578938 |
| ENSMUSG0 | 3886.006 | 4130.922 | 4006.801 | 4831.53 | 4389.279 | 4508.531 | -0.19129 | 0.000384 | 0.006706 | P4ha2 | 11 | 54100095 | 54131665 | + | 6239 | protein_co | procollage | - | 3900 | 4101 | 4299 | 4638 | 4330 | 4649 | 32.87638 | 34.94842 | 33.89833 | 40.8757 | 37.13417 | 38.14307 |
| ENSMUSG0 | 342.7656 | 269.9554 | 256.3085 | 167.7181 | 155.0946 | 215.2923 | 0.690085 | 0.000386 | 0.006742 | Mad1l1 | 5 | 1.4E+08 | 1.4E+08 | - | 3649 | protein_co | MAD1 mitc | - | 344 | 268 | 275 | 161 | 153 | 222 | 4.958141 | 3.904934 | 3.70753 | 2.42606 | 2.24346 | 3.114226 |
| ENSMUSG0 | 5819.044 | 5954.129 | 5767.408 | 6154.523 | 6928.573 | 6828.258 | -0.18307 | 0.000389 | 0.006774 | Ssr1 | 13 | 37966605 | 37994217 | - | 10431 | protein_co | signal sequ | - | 5840 | 5911 | 6188 | 5908 | 6835 | 7041 | 29.44566 | 30.12921 | 29.18437 | 31.14326 | 35.06012 | 34.5525 |
| ENSMUSG0 | 1601.234 | 1534.112 | 1727.054 | 2111.581 | 1745.575 | 2143.225 | -0.30319 | 0.000393 | 0.00684 | Tnrc18 | 5 | 1.43E+08 | 1.43E+08 | - | 12266 | protein_co | trinucleoti | - | 1607 | 1523 | 1853 | 2027 | 1722 | 2210 | 6.890445 | 6.601608 | 7.431874 | 9.086576 | 7.511575 | 9.222749 |
| ENSMUSG0 | 300.9163 | 319.313 | 382.1327 | 453.1513 | 460.2154 | 503.3185 | -0.49845 | 0.000395 | 0.006876 | Dgat2 | 7 | 99153658 | 99182719 | - | 3083 | protein_co | diacylglyce | - | 302 | 317 | 410 | 435 | 454 | 519 | 5.151904 | 5.466867 | 6.542387 | 7.758275 | 7.879218 | 8.617175 |

| gene_id | H1 | H2 | H3 | M1 | M2 | M3 | log2FoldCh | pvalue | padj | gene_name | gene_chr | gene_start | gene_end | gene_stran | gene_lengt | gene_bioty | gene_descr | tf_family | H1_readcou | H2_readcou | H3_readcou | M1_readco | M2_readco | M3_readco | H1_fpkm | H2_fpkm | H3_fpkm | M1_fpkm | M2_fpkm | M3_fpkm |
|---|---|---|---|---|---|---|---|---|---|---|---|---|---|---|---|---|---|---|---|---|---|---|---|---|---|---|---|---|---|---|
| ENSMUSG0 | 657.6317 | 741.3701 | 631.917 | 817.7557 | 861.6367 | 958.1478 | -0.37792 | 0.000396 | 0.006877 | Lrrc41 | 4 | 1.16E+08 | 1.16E+08 | + | 3713 | protein_co | leucine rich | - | 660 | 736 | 678 | 785 | 850 | 988 | 9.348745 | 10.53915 | 8.983191 | 11.62503 | 12.24884 | 13.62081 |
| ENSMUSG0 | 900.7562 | 958.9461 | 848.1482 | 1120.898 | 1135.333 | 1099.736 | -0.30995 | 0.000399 | 0.006924 | Impact | 18 | 12972252 | 12992948 | + | 3432 | protein_co | impact, RW | - | 904 | 952 | 910 | 1076 | 1120 | 1134 | 13.85337 | 14.74832 | 13.04428 | 17.23909 | 17.4611 | 16.91363 |
| ENSMUSG0 | 289.9558 | 343.4881 | 298.2499 | 201.0533 | 224.0256 | 193.957 | 0.589749 | 0.0004 | 0.006936 | Sdccag8 | 1 | 1.77E+08 | 1.77E+08 | + | 4129 | protein_co | serological | - | 291 | 341 | 320 | 193 | 221 | 200 | 3.706657 | 4.390988 | 3.812686 | 2.57017 | 2.863836 | 2.479455 |
| ENSMUSG0 | 786.1688 | 758.4942 | 843.4881 | 717.7499 | 511.9136 | 431.5544 | 0.524789 | 0.000405 | 0.007006 | Col22a1 | 15 | 71795795 | 72034227 | - | 9203 | protein_co | collagen, ty | - | 789 | 753 | 905 | 689 | 505 | 445 | 4.509017 | 4.350291 | 4.837768 | 4.116606 | 2.936045 | 2.475151 |
| ENSMUSG0 | 1813.469 | 1715.426 | 1830.509 | 2068.87 | 2134.832 | 2082.129 | -0.22986 | 0.000406 | 0.007009 | Edem3 | 1 | 1.52E+08 | 1.52E+08 | + | 6381 | protein_co | ER degrada | - | 1820 | 1703 | 1964 | 1986 | 2106 | 2147 | 15.00089 | 14.18988 | 15.14184 | 17.11354 | 17.65918 | 17.22322 |
| ENSMUSG0 | 5874.843 | 6025.647 | 6155.133 | 6541.004 | 6662.986 | 6982.453 | -0.16098 | 0.000411 | 0.007093 | Bsg | 10 | 79704491 | 79711969 | + | 1814 | protein_co | basigin [Sou | - | 5896 | 5982 | 6604 | 6279 | 6573 | 7200 | 170.9443 | 175.3323 | 179.1 | 190.328 | 193.8774 | 203.1731 |
| ENSMUSG0 | 334.7943 | 312.2619 | 311.2984 | 204.1785 | 232.1351 | 213.3527 | 0.560629 | 0.000415 | 0.007156 | Ifitm3 | 7 | 1.41E+08 | 1.41E+08 | - | 678 | protein_co | interferon i | - | 336 | 310 | 334 | 196 | 229 | 220 | 26.06417 | 24.30999 | 24.23498 | 15.89556 | 18.07202 | 16.60979 |
| ENSMUSG0 | 34.87441 | 31.22619 | 27.96093 | 5.208635 | 9.123213 | 5.818711 | 2.222264 | 0.00042 | 0.007218 | Gm22068 | 11 | 87426716 | 87426879 | + | 164 | snRNA | predicted g | - | 35 | 31 | 30 | 5 | 9 | 6 | 11.22428 | 10.05011 | 8.999188 | 1.676392 | 2.936294 | 1.872744 |
| ENSMUSG0 | 924.67 | 1003.267 | 1030.826 | 1140.691 | 1224.538 | 1316.968 | -0.3157 | 0.00042 | 0.007218 | Elovl5 | 9 | 77917364 | 77984519 | + | 3522 | protein_co | ELOVL fami | - | 928 | 996 | 1106 | 1095 | 1208 | 1358 | 13.85776 | 15.03567 | 15.44869 | 17.0952 | 18.35179 | 19.73702 |
| ENSMUSG0 | 427.4606 | 515.7357 | 468.8116 | 675.0391 | 578.8172 | 648.7863 | -0.43032 | 0.000423 | 0.007252 | Nup62 | 7 | 44816088 | 44830812 | + | 2854 | protein_co | nucleopori | - | 429 | 512 | 503 | 648 | 571 | 669 | 7.905651 | 9.538252 | 8.670416 | 12.48448 | 10.70491 | 11.99895 |
| ENSMUSG0 | 1146.87 | 1127.165 | 1145.466 | 1371.955 | 1357.331 | 1382.914 | -0.26611 | 0.000423 | 0.007252 | Trrap | 5 | 1.45E+08 | 1.45E+08 | + | 20859 | protein_co | transforma | - | 1151 | 1119 | 1229 | 1317 | 1339 | 1426 | 2.902125 | 2.852262 | 2.898573 | 3.471697 | 3.434693 | 3.499428 |
| ENSMUSG0 | 17.93541 | 10.07296 | 12.1164 | 46.87772 | 57.78035 | 33.94248 | -1.78817 | 0.000424 | 0.007252 | Zfp213 | 17 | 23556767 | 23564238 | - | 2686 | protein_co | zinc finger | zf-C2H2 | 18 | 10 | 13 | 45 | 57 | 35 | 0.352453 | 0.197946 | 0.238102 | 0.921204 | 1.135454 | 0.667011 |
| ENSMUSG0 | 984.4547 | 1049.603 | 1128.69 | 715.6665 | 874.8147 | 867.9578 | 0.363254 | 0.000426 | 0.007285 | Slc16a2 | X | 1.04E+08 | 1.04E+08 | - | 4263 | protein_co | solute carri | - | 988 | 1042 | 1211 | 687 | 863 | 895 | 12.18922 | 12.99586 | 13.97509 | 8.861166 | 10.83169 | 10.74679 |
| ENSMUSG0 | 247.1101 | 243.7657 | 187.3382 | 328.144 | 335.5315 | 351.0622 | -0.58273 | 0.000428 | 0.007299 | Thap7 | 16 | 17527982 | 17531136 | - | 1882 | protein_co | THAP doma | THAP | 248 | 242 | 201 | 315 | 331 | 362 | 6.930529 | 6.836732 | 5.254148 | 9.203233 | 9.410424 | 9.846005 |
| ENSMUSG0 | 1524.51 | 1622.754 | 1538.783 | 1856.358 | 1861.135 | 1815.438 | -0.23973 | 0.000429 | 0.007306 | Plekhh2 | 17 | 84511895 | 84622142 | + | 6886 | protein_co | pleckstrin h | - | 1530 | 1611 | 1651 | 1782 | 1836 | 1872 | 11.68581 | 12.43888 | 11.79522 | 14.22952 | 14.26614 | 13.91586 |
| ENSMUSG0 | 78.71652 | 66.48156 | 73.63045 | 33.33527 | 33.45178 | 21.33527 | 1.314684 | 0.000429 | 0.007306 | Nkd2 | 13 | 73818534 | 73847631 | - | 4826 | protein_co | naked cutic | - | 79 | 66 | 79 | 32 | 33 | 22 | 0.860942 | 0.727126 | 0.805315 | 0.364596 | 0.365871 | 0.233349 |
| ENSMUSG0 | 615.7824 | 572.1443 | 598.3639 | 767.7528 | 748.1034 | 769.0397 | -0.3551 | 0.000438 | 0.007443 | Nop14 | 5 | 34638536 | 34660148 | - | 2730 | protein_co | NOP14 nuc | - | 618 | 568 | 642 | 737 | 738 | 793 | 11.90584 | 11.06212 | 11.56907 | 14.84411 | 14.4642 | 14.869 |
| ENSMUSG0 | 616.7788 | 586.2465 | 658.0139 | 471.9023 | 461.2291 | 478.1041 | 0.39947 | 0.000442 | 0.007501 | Ptp4a3 | 15 | 73723145 | 73758766 | + | 9351 | protein_co | protein tyr | - | 619 | 582 | 706 | 453 | 455 | 493 | 3.481504 | 3.30916 | 3.714262 | 2.663726 | 2.603479 | 2.698733 |
| ENSMUSG0 | 79.71293 | 63.45967 | 67.10623 | 177.0936 | 107.4512 | 129.9512 | -0.97856 | 0.000445 | 0.007544 | Tmem51 | 4 | 1.42E+08 | 1.42E+08 | - | 1875 | protein_co | transmemb | - | 80 | 63 | 72 | 170 | 106 | 134 | 2.244001 | 1.786455 | 1.889109 | 4.985367 | 3.024861 | 3.65826 |
| ENSMUSG0 | 207.2536 | 169.2258 | 219.9593 | 111.4648 | 122.6565 | 122.1929 | 0.743541 | 0.000446 | 0.00755 | Rbms3 | 9 | 1.17E+08 | 1.18E+08 | - | 10413 | protein_co | RNA bindin | - | 208 | 168 | 236 | 107 | 121 | 126 | 1.050562 | 0.8578 | 1.114967 | 0.565012 | 0.621742 | 0.619392 |
| ENSMUSG0 | 875.8459 | 940.8148 | 902.206 | 693.7902 | 780.5415 | 646.8467 | 0.358375 | 0.000449 | 0.0076 | Spats2l | 1 | 57774162 | 57948394 | + | 8041 | protein_co | spermatoge | - | 879 | 934 | 968 | 666 | 770 | 667 | 5.749276 | 6.175749 | 5.922311 | 4.554216 | 5.123674 | 4.246067 |
| ENSMUSG0 | 2356.514 | 2227.132 | 2417.688 | 1946.988 | 2090.229 | 1924.054 | 0.23226 | 0.00045 | 0.007606 | Nedd9 | 13 | 41309581 | 41487362 | - | 6376 | protein_co | neural prec | - | 2365 | 2211 | 2594 | 1869 | 2062 | 1984 | 19.5082 | 18.43713 | 20.01463 | 16.11797 | 17.30379 | 15.92812 |
| ENSMUSG0 | 834.993 | 793.7495 | 904.0701 | 1043.81 | 1149.525 | 1001.788 | -0.33454 | 0.000455 | 0.007674 | Prkca | 11 | 1.08E+08 | 1.08E+08 | - | 8992 | protein_co | protein kin | - | 838 | 788 | 970 | 1002 | 1134 | 1033 | 4.901422 | 4.659322 | 5.306905 | 6.127183 | 6.747728 | 5.880512 |
| ENSMUSG0 | 3372.854 | 3338.18 | 3343.195 | 3179.351 | 2611.266 | 2683.396 | 0.24695 | 0.000457 | 0.007702 | Srsf5 | 12 | 80945504 | 80950507 | + | 3988 | protein_co | serine/argi | - | 3385 | 3314 | 3587 | 3052 | 2576 | 2767 | 44.64141 | 44.18249 | 44.24886 | 42.0803 | 34.56142 | 35.51609 |
| ENSMUSG0 | 1164.805 | 1273.223 | 1274.086 | 1491.753 | 1436.399 | 1563.294 | -0.27495 | 0.000461 | 0.007752 | Adipor2 | 6 | 1.19E+08 | 1.19E+08 | - | 5979 | protein_co | adiponecti | - | 1169 | 1264 | 1367 | 1432 | 1417 | 1612 | 10.28301 | 11.24013 | 11.24776 | 13.16934 | 12.68067 | 13.8009 |
| ENSMUSG0 | 4091.266 | 4337.418 | 4136.354 | 3601.25 | 3833.777 | 3623.118 | 0.184244 | 0.000461 | 0.007752 | Adamts1 | 16 | 85793827 | 85803113 | - | 5302 | protein_co | a disintegri | - | 4106 | 4306 | 4438 | 3457 | 3782 | 3736 | 40.72992 | 43.18044 | 41.17878 | 35.85165 | 38.16652 | 36.06934 |
| ENSMUSG0 | 2261.855 | 2252.315 | 1969.382 | 2649.112 | 2545.376 | 2476.831 | -0.24295 | 0.000462 | 0.00776 | Sat1 | X | 1.55E+08 | 1.55E+08 | - | 2774 | protein_co | spermidine | - | 2270 | 2236 | 2113 | 2543 | 2511 | 2554 | 43.03816 | 42.85664 | 37.47304 | 50.40683 | 48.43297 | 47.12871 |
| ENSMUSG0 | 779.1939 | 816.9174 | 872.381 | 1047.977 | 1139.388 | 948.4499 | -0.34461 | 0.000471 | 0.007893 | Mast2 | 4 | 1.16E+08 | 1.16E+08 | - | 7645 | protein_co | microtubul | - | 782 | 811 | 936 | 1006 | 1124 | 978 | 5.379769 | 5.640222 | 6.023158 | 7.235523 | 7.866646 | 6.548358 |
| ENSMUSG0 | 826.0253 | 764.5379 | 772.6537 | 890.6766 | 1131.278 | 1017.305 | -0.36326 | 0.000473 | 0.007896 | Kcnma1 | 14 | 23289431 | 24014491 | - | 16976 | protein_co | potassium | - | 829 | 759 | 829 | 855 | 1116 | 1049 | 2.568346 | 2.377164 | 2.402399 | 2.769365 | 3.517464 | 3.163087 |
| ENSMUSG0 | 1306.296 | 1258.113 | 1246.125 | 1512.588 | 1627.987 | 1454.678 | -0.27011 | 0.000473 | 0.007896 | Pprc1 | 19 | 46044886 | 46072915 | + | 7516 | protein_co | peroxisome | - | 1311 | 1249 | 1337 | 1452 | 1606 | 1500 | 9.173821 | 8.835446 | 8.751259 | 10.62256 | 11.43298 | 10.21587 |
| ENSMUSG0 | 693.5025 | 790.7277 | 801.5467 | 992.7659 | 932.5951 | 963.9665 | -0.33771 | 0.000473 | 0.007896 | Nifk | 1 | 1.18E+08 | 1.18E+08 | + | 1965 | protein_co | nucleolar p | - | 696 | 785 | 860 | 953 | 920 | 994 | 18.62863 | 21.24026 | 21.53088 | 26.66735 | 25.05106 | 25.89375 |
| ENSMUSG0 | 414.5073 | 440.1885 | 426.8702 | 276.0577 | 275.8238 | 347.1831 | 0.510619 | 0.000484 | 0.00808 | Kif23 | 9 | 61915905 | 61946774 | - | 7901 | protein_co | kinesin fam | - | 416 | 437 | 458 | 265 | 272 | 358 | 2.769144 | 2.94071 | 2.851736 | 1.844222 | 1.841992 | 2.319381 |
| ENSMUSG0 | 6713.822 | 6607.864 | 6842.04 | 7463.974 | 7337.09 | 7465.406 | -0.14304 | 0.000486 | 0.0081 | Kank1 | 19 | 25236975 | 25434496 | + | 6166 | protein_co | KN motif ar | - | 6738 | 6560 | 7341 | 7165 | 7238 | 7698 | 57.47273 | 56.56569 | 58.57032 | 63.8943 | 62.80813 | 63.90656 |
| ENSMUSG0 | 1470.704 | 1462.594 | 1461.425 | 1244.864 | 1150.538 | 1258.781 | 0.266132 | 0.000488 | 0.008118 | Bag1 | 4 | 40936398 | 40948294 | - | 1349 | protein_co | BCL2-assoc | - | 1476 | 1452 | 1568 | 1195 | 1135 | 1298 | 57.54515 | 57.22785 | 57.18209 | 48.70857 | 45.01785 | 49.25313 |
| ENSMUSG0 | 903.7454 | 932.7564 | 931.099 | 701.0823 | 735.9391 | 770.9792 | 0.325589 | 0.00049 | 0.008145 | Plxdc2 | 2 | 16356304 | 16755839 | + | 6947 | protein_co | plexin dom | - | 907 | 926 | 999 | 673 | 726 | 795 | 6.866641 | 7.087066 | 7.074473 | 5.32681 | 5.591652 | 5.857886 |
| ENSMUSG0 | 427.4606 | 379.7507 | 410.0937 | 276.0577 | 324.3809 | 235.6578 | 0.542671 | 0.0005 | 0.008299 | Itgbl1 | 14 | 1.24E+08 | 1.24E+08 | + | 4229 | protein_co | integrin, be | - | 429 | 377 | 440 | 265 | 320 | 243 | 5.33524 | 4.739761 | 5.118479 | 3.445543 | 4.048677 | 2.941302 |
| ENSMUSG0 | 875.8459 | 901.5303 | 935.7592 | 1135.482 | 1093.772 | 1099.736 | -0.29481 | 0.000509 | 0.008429 | Hbegf | 18 | 36504929 | 36515805 | - | 2373 | protein_co | heparin-bir | - | 879 | 895 | 1004 | 1090 | 1079 | 1134 | 19.48164 | 20.05294 | 20.8143 | 25.25679 | 24.32901 | 24.46169 |
| ENSMUSG0 | 380.6293 | 394.8602 | 409.1616 | 517.7383 | 546.3791 | 521.7444 | -0.42054 | 0.00051 | 0.008429 | Zbtb38 | 9 | 96682770 | 96752831 | - | 11733 | protein_co | zinc finger | ZBTB | 382 | 392 | 439 | 497 | 539 | 538 | 1.712335 | 1.776355 | 1.840693 | 2.329146 | 2.457992 | 2.347169 |
| ENSMUSG0 | 1189.716 | 1137.238 | 1074.632 | 864.6334 | 899.1433 | 974.6341 | 0.312218 | 0.00051 | 0.008429 | Pbx1 | 1 | 1.68E+08 | 1.68E+08 | - | 13844 | protein_co | pre B cell le | Homeobox | 1194 | 1129 | 1153 | 830 | 887 | 1005 | 4.536042 | 4.335959 | 4.097261 | 3.296598 | 3.428174 | 3.715999 |
| ENSMUSG0 | 1002.39 | 927.72 | 1065.311 | 1237.572 | 1286.373 | 1175.38 | -0.30391 | 0.000511 | 0.008442 | Kif13b | 14 | 64647265 | 64809617 | + | 13017 | protein_co | kinesin fam | - | 1006 | 921 | 1143 | 1188 | 1269 | 1212 | 4.064634 | 3.76185 | 4.319776 | 5.018281 | 5.216167 | 4.766096 |
| ENSMUSG0 | 3652.845 | 3915.361 | 3868.861 | 3326.234 | 3493.177 | 3109.131 | 0.204259 | 0.000519 | 0.008552 | Lamb2 | 9 | 1.08E+08 | 1.08E+08 | + | 6082 | protein_co | laminin, be | - | 3666 | 3887 | 4151 | 3193 | 3446 | 3206 | 31.70154 | 33.97981 | 33.57625 | 28.86702 | 30.31584 | 26.98287 |
| ENSMUSG0 | 605.8183 | 627.5456 | 640.3053 | 829.2147 | 816.0207 | 746.7346 | -0.35185 | 0.00052 | 0.008561 | Id4 | 13 | 48261228 | 48266026 | + | 3849 | protein_co | inhibitor o | bHLH | 608 | 623 | 687 | 796 | 805 | 770 | 8.307876 | 8.605833 | 8.780813 | 11.37142 | 11.19048 | 10.24033 |
| ENSMUSG0 | 9571.531 | 9540.104 | 9488.076 | 10112.04 | 11543.9 | 10488.23 | -0.16858 | 0.000522 | 0.008594 | Epyc | 10 | 97644068 | 97682454 | + | 2561 | protein_co | epiphycan | - | 9606 | 9471 | 10180 | 9707 | 11388 | 10815 | 197.2729 | 196.6251 | 195.5528 | 208.4131 | 237.9242 | 216.1663 |
| ENSMUSG0 | 1164.805 | 1141.267 | 1179.951 | 1010.475 | 949.8278 | 784.5562 | 0.345583 | 0.000527 | 0.008663 | Cercam | 2 | 29869164 | 29882840 | + | 3383 | protein_co | cerebral en | - | 1169 | 1133 | 1266 | 970 | 937 | 809 | 18.17385 | 17.80659 | 18.41016 | 15.76592 | 14.81967 | 12.24102 |
| ENSMUSG0 | 1542.445 | 1538.142 | 1443.716 | 1794.896 | 1772.944 | 1773.737 | -0.23981 | 0.000529 | 0.008675 | Klhl9 | 4 | 88718292 | 88722465 | - | 4174 | protein_co | kelch-like 9 | - | 1548 | 1527 | 1549 | 1723 | 1749 | 1829 | 19.50531 | 19.45089 | 18.25681 | 22.69772 | 22.42013 | 22.43016 |
| ENSMUSG0 | 2772.017 | 2888.926 | 2756.948 | 3057.469 | 3401.945 | 3246.841 | -0.20565 | 0.000531 | 0.008695 | Eif2s2 | 2 | 1.55E+08 | 1.55E+08 | - | 6755 | protein_co | eukaryotic | - | 2782 | 2868 | 2958 | 2935 | 3356 | 3348 | 21.66038 | 22.5739 | 21.54263 | 23.89088 | 26.5826 | 25.37063 |
| ENSMUSG0 | 1183.737 | 1114.07 | 1136.146 | 1328.202 | 1531.686 | 1344.122 | -0.29189 | 0.000532 | 0.008705 | Desi2 | 1 | 1.78E+08 | 1.78E+08 | + | 9156 | protein_co | desumoyla | - | 1188 | 1106 | 1219 | 1275 | 1511 | 1386 | 6.824094 | 6.422471 | 6.549736 | 7.656916 | 8.829975 | 7.748694 |
| ENSMUSG0 | 209.2465 | 241.7511 | 280.5413 | 129.1742 | 150.0262 | 171.652 | 0.697909 | 0.000533 | 0.008708 | Nkd1 | 8 | 88521354 | 88594884 | + | 4558 | protein_co | naked cutic | - | 210 | 240 | 301 | 124 | 148 | 177 | 2.423144 | 2.79956 | 3.248763 | 1.49588 | 1.737354 | 1.987788 |
| ENSMUSG0 | 592.865 | 577.1808 | 631.917 | 758.3773 | 758.2403 | 782.6167 | -0.35129 | 0.000542 | 0.008852 | Lss | 10 | 76531588 | 76557138 | + | 10787 | protein_co | lanosterol | - | 595 | 573 | 678 | 728 | 748 | 807 | 2.901019 | 2.824273 | 3.09211 | 3.710908 | 3.710238 | 3.829516 |
| ENSMUSG0 | 1387.005 | 1345.748 | 1286.203 | 1118.815 | 1128.237 | 1080.341 | 0.272345 | 0.000547 | 0.008924 | Zfp521 | 18 | 13687013 | 13972733 | - | 6152 | protein_co | zinc finger | zf-C2H2 | 1392 | 1336 | 1380 | 1074 | 1113 | 1114 | 11.90028 | 11.5463 | 11.03541 | 9.599253 | 9.680096 | 9.269151 |
| ENSMUSG0 | 308.8876 | 380.758 | 358.8319 | 430.2333 | 577.8035 | 485.8624 | -0.51089 | 0.000551 | 0.008979 | Vldlr | 19 | 27216484 | 27254231 | + | 10068 | protein_co | very low de | - | 310 | 378 | 385 | 413 | 570 | 501 | 1.619395 | 1.996188 | 1.881237 | 2.25557 | 3.029231 | 2.547215 |
| ENSMUSG0 | 623.7537 | 641.6478 | 534.0538 | 819.8392 | 750.1308 | 768.0699 | -0.3783 | 0.000552 | 0.008982 | Csnk1g3 | 18 | 53862113 | 53955684 | + | 4394 | protein_co | casein kina | - | 626 | 637 | 573 | 787 | 740 | 792 | 7.492877 | 7.707831 | 6.415352 | 9.848365 | 9.010989 | 9.226483 |
| ENSMUSG0 | 498.2058 | 493.5752 | 458.5593 | 633.37 | 616.3237 | 641.9978 | -0.38373 | 0.000563 | 0.009142 | Crtc3 | 7 | 80586627 | 80688877 | - | 9850 | protein_co | CREB regula | - | 500 | 490 | 492 | 608 | 608 | 662 | 2.669735 | 2.644921 | 2.457281 | 3.394039 | 3.302693 | 3.440272 |
| ENSMUSG0 | 231.1675 | 225.6344 | 260.9687 | 319.8102 | 385.2023 | 342.3342 | -0.54456 | 0.000565 | 0.00916 | Zbtb39 | 10 | 1.28E+08 | 1.28E+08 | + | 5960 | protein_co | zinc finger | ZBTB | 232 | 224 | 280 | 307 | 380 | 353 | 2.047274 | 1.998272 | 2.311201 | 2.832315 | 3.411443 | 3.031793 |
| ENSMUSG0 | 14579.5 | 14398.29 | 15359.87 | 13962.27 | 13219.53 | 12814.74 | 0.148807 | 0.000565 | 0.00916 | Fosb | 7 | 19302696 | 19310051 | - | 6243 | protein_co | FBJ osteosa | TF_bZIP | 14632 | 14294 | 16480 | 13403 | 13041 | 13214 | 123.2664 | 121.7344 | 129.8643 | 118.0479 | 111.7682 | 108.3458 |
| ENSMUSG0 | 655.6389 | 704.1002 | 671.9944 | 881.3011 | 821.0891 | 857.2901 | -0.33329 | 0.000572 | 0.009257 | Phgdh | 3 | 98313170 | 98339990 | - | 2486 | protein_co | 3-phospho | - | 658 | 699 | 721 | 846 | 810 | 884 | 13.92064 | 14.94958 | 14.2679 | 18.71194 | 17.4335 | 18.20213 |
| ENSMUSG0 | 82.70217 | 74.53993 | 75.49451 | 146.8835 | 165.2315 | 116.3742 | -0.88004 | 0.000577 | 0.009332 | Crhr1 | 11 | 1.04E+08 | 1.04E+08 | + | 2460 | protein_co | corticotrop | - | 83 | 74 | 81 | 141 | 163 | 120 | 1.774505 | 1.599372 | 1.619854 | 3.151617 | 3.545303 | 2.496992 |
| ENSMUSG0 | 544.0408 | 601.3559 | 602.092 | 740.6679 | 704.5147 | 822.3779 | -0.37602 | 0.000582 | 0.009402 | Hdac4 | 1 | 91928779 | 92195699 | - | 15813 | protein_co | histone dea | - | 546 | 597 | 646 | 711 | 695 | 848 | 1.815987 | 2.007303 | 2.00976 | 2.472321 | 2.351643 | 2.745065 |
| ENSMUSG0 | 1498.603 | 1435.397 | 1519.211 | 1687.598 | 1740.506 | 1866.837 | -0.24988 | 0.000589 | 0.009501 | Map3k2 | 18 | 32163089 | 32236751 | + | 10791 | protein_co | mitogen-ac | - | 1504 | 1425 | 1630 | 1620 | 1717 | 1925 | 7.330279 | 7.021113 | 7.431078 | 8.254728 | 8.513525 | 9.131458 |
| ENSMUSG0 | 47.82776 | 37.26997 | 65.24217 | 18.75109 | 5.068451 | 18.42592 | 1.831888 | 0.000591 | 0.009517 | Tpsb2 | 17 | 25366550 | 25368094 | + | 1081 | protein_co | tryptase be | - | 48 | 37 | 70 | 18 | 5 | 19 | 2.335339 | 1.819822 | 3.185651 | 0.91558 | 0.247483 | 0.899703 |
| ENSMUSG0 | 205.2608 | 221.6052 | 208.775 | 310.4347 | 293.9702 | 324.878 | -0.54826 | 0.000599 | 0.009637 | Gan | 8 | 1.17E+08 | 1.17E+08 | + | 13463 | protein_co | giant axona | - | 206 | 220 | 224 | 298 | 290 | 335 | 0.804748 | 0.868828 | 0.818525 | 1.217093 | 1.152543 | 1.27372 |
| ENSMUSG0 | 291.9486 | 302.1889 | 221.8234 | 203.1368 | 151.0399 | 153.2261 | 0.685454 | 0.0006 | 0.009637 | 4933421O | 4 | 33027142 | 33031323 | - | 3847 | processed_ | RIKEN cDN | - | 293 | 300 | 238 | 195 | 149 | 158 | 4.005712 | 4.146215 | 3.043552 | 2.78716 | 2.072359 | 2.102355 |
| ENSMUSG0 | 444.3996 | 484.5096 | 508.8889 | 578.1585 | 696.4052 | 640.0582 | -0.41304 | 0.000602 | 0.009666 | Pdia5 | 16 | 35397312 | 35490873 | - | 2771 | protein_co | protein dis | - | 446 | 481 | 546 | 555 | 687 | 660 | 8.465111 | 9.229142 | 9.693531 | 11.01301 | 13.26542 | 12.1921 |
| ENSMUSG0 | 234.1567 | 250.8168 | 245.1242 | 182.3022 | 141.9166 | 139.6491 | 0.65543 | 0.00061 | 0.009781 | Epha4 | 1 | 77367185 | 77515088 | - | 8054 | protein_co | Eph recepto | - | 235 | 249 | 263 | 175 | 140 | 144 | 1.534584 | 1.643768 | 1.60646 | 1.194747 | 0.930073 | 0.915213 |
| ENSMUSG0 | 1455.757 | 1398.127 | 1640.375 | 1687.598 | 1974.669 | 1823.196 | -0.28721 | 0.000611 | 0.009781 | Actr1b | 1 | 36698114 | 36714422 | - | 4274 | protein_co | ARP1 actin | - | 1461 | 1388 | 1760 | 1620 | 1948 | 1880 | 17.97836 | 17.26664 | 20.25835 | 20.84155 | 24.38682 | 22.51616 |
| ENSMUSG0 | 787.1652 | 741.3701 | 728.8483 | 884.4263 | 1047.142 | 927.1147 | -0.34092 | 0.000612 | 0.009784 | Gtpbp4 | 13 | 8966331 | 8996083 | - | 9566 | protein_co | GTP bindin | - | 790 | 736 | 782 | 849 | 1033 | 956 | 4.343412 | 4.090725 | 4.021632 | 4.880078 | 5.77791 | 5.115624 |
| ENSMUSG0 | 669.5887 | 609.4143 | 629.1209 | 782.337 | 830.2123 | 805.8915 | -0.34202 | 0.000614 | 0.0098 | Lemd3 | 10 | 1.21E+08 | 1.21E+08 | - | 4809 | protein_co | LEM domai | - | 672 | 605 | 675 | 751 | 819 | 831 | 7.349348 | 6.68888 | 6.905178 | 8.586865 | 9.11234 | 8.845397 |
| ENSMUSG0 | 1133.916 | 1053.632 | 1042.011 | 922.9702 | 870.76 | 799.103 | 0.317132 | 0.000617 | 0.009837 | Pycr1 | 11 | 1.21E+08 | 1.21E+08 | - | 3935 | protein_co | pyrroline-5 | - | 1138 | 1046 | 1118 | 886 | 859 | 824 | 15.21009 | 14.13318 | 13.97729 | 12.38051 | 11.68017 | 10.71898 |
| ENSMUSG0 | 381.6257 | 352.5537 | 389.589 | 518.7801 | 551.4475 | 461.6178 | -0.44623 | 0.000617 | 0.009837 | Ndrg4 | 8 | 95676980 | 95715119 | + | 4893 | protein_co | N-myc dow | - | 383 | 350 | 418 | 498 | 544 | 476 | 4.116782 | 3.803169 | 4.202686 | 5.596334 | 5.948733 | 4.979696 |
| ENSMUSG0 | 130.5299 | 105.7661 | 150.057 | 60.42017 | 78.05415 | 63.03604 | 0.939986 | 0.000621 | 0.00987 | Tek | 4 | 94739289 | 94874976 | + | 4712 | protein_co | TEK recepto | - | 131 | 105 | 161 | 58 | 77 | 65 | 1.462178 | 1.184778 | 1.680918 | 0.676818 | 0.874352 | 0.706121 |
| ENSMUSG0 | 443.4032 | 411.9842 | 401.7054 | 598.993 | 502.7904 | 591.569 | -0.43007 | 0.000621 | 0.00987 | Hif1an | 19 | 44562850 | 44576274 | + | 6606 | protein_co | hypoxia-inc | - | 445 | 409 | 431 | 575 | 496 | 610 | 3.542875 | 3.291831 | 3.209701 | 4.786067 | 4.017389 | 4.726747 |
| ENSMUSG0 | 218.2142 | 229.6636 | 182.6781 | 141.6749 | 118.6018 | 127.0419 | 0.702749 | 0.000627 | 0.009949 | B230118H | 2 | 1.02E+08 | 1.02E+08 | - | 4289 | protein_co | RIKEN cDN | - | 219 | 228 | 196 | 136 | 117 | 131 | 2.685483 | 2.826387 | 2.248153 | 1.743542 | 1.459589 | 1.563458 |
| ENSMUSG0 | 213.2321 | 293.1232 | 229.2796 | 421.8994 | 371.0106 | 317.1198 | -0.59346 | 0.000629 | 0.009966 | Champ1 | 8 | 13869641 | 13881639 | + | 4040 | protein_co | chromosor | - | 214 | 291 | 246 | 405 | 366 | 327 | 2.785908 | 3.829697 | 2.995571 | 5.512177 | 4.847307 | 4.143215 |
| ENSMUSG0 | 2756.075 | 2536.372 | 2512.756 | 3089.762 | 2890.031 | 3038.337 | -0.20851 | 0.000643 | 0.01018 | Stk38l | 6 | 1.47E+08 | 1.47E+08 | + | 4880 | protein_co | serine/thre | - | 2766 | 2518 | 2696 | 2966 | 2851 | 3133 | 29.81032 | 27.43397 | 27.17853 | 33.41956 | 31.25922 | 32.86334 |
| ENSMUSG0 | 3388.796 | 3180.035 | 3489.524 | 3889.809 | 3694.901 | 3895.627 | -0.19059 | 0.000644 | 0.01018 | Prnp | 2 | 1.32E+08 | 1.32E+08 | + | 2343 | protein_co | prion prote | - | 3401 | 3157 | 3744 | 3734 | 3645 | 4017 | 76.3429 | 71.63992 | 78.61211 | 87.62973 | 83.23885 | 87.76081 |
| ENSMUSG0 | 2101.432 | 2006.534 | 2181.885 | 2529.313 | 2277.762 | 2603.873 | -0.23653 | 0.000647 | 0.01022 | Rcc2 | 4 | 1.41E+08 | 1.41E+08 | + | 4155 | protein_co | regulator o | - | 2109 | 1992 | 2341 | 2428 | 2247 | 2685 | 26.69561 | 25.49007 | 27.71764 | 32.13121 | 28.93562 | 33.07839 |
| ENSMUSG0 | 70.74523 | 68.49615 | 50.32968 | 23.95972 | 17.23273 | 30.06334 | 1.408974 | 0.000654 | 0.010316 | 2810006K | 5 | 1.24E+08 | 1.24E+08 | + | 2873 | protein_co | RIKEN cDN | - | 71 | 68 | 54 | 23 | 17 | 31 | 1.299742 | 1.258421 | 0.924664 | 0.440192 | 0.316602 | 0.552328 |
| ENSMUSG0 | 16.939 | 45.32834 | 37.28124 | 3.125181 | 6.082142 | 10.66764 | 2.317036 | 0.000655 | 0.010319 | Scarna2 | 3 | 1.09E+08 | 1.09E+08 | - | 414 | scaRNA | small Cajal | - | 17 | 45 | 40 | 3 | 6 | 11 | 2.159648 | 5.779163 | 4.753194 | 0.398447 | 0.775446 | 1.360077 |
| ENSMUSG0 | 1262.454 | 1299.412 | 1111.913 | 1463.626 | 1509.385 | 1481.832 | -0.27866 | 0.000658 | 0.010352 | Wdr43 | 17 | 71616215 | 71659031 | + | 3450 | protein_co | WD repeat | - | 1267 | 1290 | 1193 | 1405 | 1489 | 1528 | 19.31487 | 19.88032 | 17.01168 | 22.39271 | 23.09279 | 22.67124 |
| ENSMUSG0 | 236.1496 | 282.043 | 273.0851 | 386.4807 | 336.5452 | 418.9472 | -0.52942 | 0.000661 | 0.010399 | Itpkc | 7 | 27207172 | 27228661 | - | 5285 | protein_co | inositol 1,4 | - | 237 | 280 | 293 | 371 | 332 | 432 | 2.35851 | 2.816864 | 2.727398 | 3.859921 | 3.361197 | 4.184175 |
| ENSMUSG0 | 374.6508 | 365.6486 | 412.8897 | 285.4332 | 178.4095 | 287.0564 | 0.619025 | 0.000667 | 0.01047 | Flnc | 6 | 29433256 | 29461883 | + | 9402 | protein_co | filamin C, g | - | 376 | 363 | 443 | 274 | 176 | 296 | 2.103303 | 2.052765 | 2.317978 | 1.602433 | 1.001597 | 1.611545 |
| ENSMUSG0 | 355.719 | 527.8233 | 428.7343 | 279.1828 | 248.3541 | 343.304 | 0.590731 | 0.000667 | 0.01047 | Apold1 | 6 | 1.35E+08 | 1.35E+08 | + | 3567 | protein_co | apolipopro | - | 357 | 524 | 460 | 268 | 245 | 354 | 5.263801 | 7.810538 | 6.344255 | 4.131247 | 3.675055 | 5.080088 |
| ENSMUSG0 | 83.69858 | 127.9266 | 128.6203 | 217.721 | 176.3821 | 192.9873 | -0.78599 | 0.000669 | 0.01048 | Nptx1 | 11 | 1.2E+08 | 1.2E+08 | - | 5217 | protein_co | neuronal p | - | 84 | 127 | 138 | 209 | 174 | 199 | 0.846823 | 1.294302 | 1.30132 | 2.202799 | 1.784552 | 1.952555 |
| ENSMUSG0 | 1837.383 | 2014.593 | 1981.498 | 2380.346 | 2181.461 | 2277.056 | -0.22925 | 0.00067 | 0.010487 | Ubn1 | 16 | 5050057 | 5086289 | + | 8272 | protein_co | ubinuclein | - | 1844 | 2000 | 2126 | 2285 | 2152 | 2348 | 11.72424 | 12.85501 | 12.64383 | 15.18886 | 13.91979 | 14.52977 |

| gene_id | H1 | H2 | H3 | M1 | M2 | M3 | log2FoldCh | pvalue | padj | gene_name | gene_chr | gene_start | gene_end | gene_stran | gene_lengtl | gene_bioty | gene_descr | tf_family | H1_readco | H2_readcou | H3_readco | M1_readco | M2_readco | M3_readco | H1_fpkm | H2_fpkm | H3_fpkm | M1_fpkm | M2_fpkm | M3_fpkm |
|---|---|---|---|---|---|---|---|---|---|---|---|---|---|---|---|---|---|---|---|---|---|---|---|---|---|---|---|---|---|---|
| ENSMUSG0 | 128.5371 | 143.0361 | 202.2507 | 229.1799 | 264.5732 | 280.2679 | -0.70665 | 0.000676 | 0.010569 | Csdc2 | 15 | 81936759 | 81950941 | + | 4623 | protein_co | cold shock | CSD | 129 | 142 | 217 | 220 | 261 | 289 | 1.467574 | 1.633117 | 2.309201 | 2.616666 | 3.020768 | 3.199964 |
| ENSMUSG0 | 600.8362 | 630.5675 | 581.5874 | 782.337 | 773.4457 | 744.795 | -0.34377 | 0.000685 | 0.010686 | Vps4b | 1 | 1.07E+08 | 1.07E+08 | - | 8905 | protein_co | vacuolar pr | - | 603 | 626 | 624 | 751 | 763 | 768 | 3.561375 | 3.737603 | 3.44728 | 4.637196 | 4.584494 | 4.414671 |
| ENSMUSG0 | 2802.906 | 2767.043 | 2632.988 | 2464.726 | 2379.131 | 2246.023 | 0.210463 | 0.000686 | 0.010686 | Tnn | 1 | 1.6E+08 | 1.6E+08 | - | 5817 | protein_co | tenascin N | - | 2813 | 2747 | 2825 | 2366 | 2347 | 2316 | 25.43343 | 25.10802 | 23.8916 | 22.36481 | 21.58812 | 20.3803 |
| ENSMUSG0 | 669.5887 | 708.1294 | 693.4311 | 457.3182 | 608.2142 | 477.1343 | 0.425014 | 0.000686 | 0.010686 | Klhl13 | X | 23219271 | 23365082 | - | 4253 | protein_co | kelch-like 1 | - | 672 | 703 | 744 | 439 | 600 | 492 | 8.310138 | 8.788459 | 8.606042 | 5.675689 | 7.548431 | 5.921624 |
| ENSMUSG0 | 757.2729 | 753.4577 | 671.0623 | 906.3025 | 1000.512 | 863.1088 | -0.34464 | 0.000689 | 0.010724 | Sptlc2 | 12 | 87305058 | 87388355 | - | 8115 | protein_co | serine palm | - | 760 | 748 | 720 | 870 | 987 | 890 | 4.925603 | 4.900787 | 4.364856 | 5.894951 | 6.507729 | 5.614002 |
| ENSMUSG0 | 940.6126 | 825.983 | 900.342 | 1061.52 | 1168.785 | 1076.462 | -0.3101 | 0.000695 | 0.010798 | Kmt5b | 19 | 3767421 | 3818303 | + | 14116 | protein_co | lysine meth | - | 944 | 820 | 966 | 1019 | 1153 | 1110 | 3.517181 | 3.088553 | 3.366599 | 3.969282 | 4.370373 | 4.025153 |
| ENSMUSG0 | 1504.582 | 1419.281 | 1495.91 | 1200.07 | 1241.771 | 1261.691 | 0.25502 | 0.000696 | 0.010798 | Rnf144a | 12 | 26300964 | 26415254 | - | 7340 | protein_co | ring finger p | - | 1510 | 1409 | 1605 | 1152 | 1225 | 1301 | 10.8197 | 10.20629 | 10.75734 | 8.629902 | 8.929782 | 9.073029 |
| ENSMUSG0 | 2286.765 | 2150.578 | 2201.457 | 2513.687 | 2653.841 | 2500.106 | -0.20783 | 0.000698 | 0.010812 | Sec16a | 2 | 26409431 | 26445216 | - | 9189 | protein_co | SEC16 hom | - | 2295 | 2135 | 2362 | 2413 | 2618 | 2578 | 13.13557 | 12.35328 | 12.64554 | 14.43905 | 15.24411 | 14.36104 |
| ENSMUSG0 | 594.8578 | 618.48 | 672.9264 | 805.255 | 777.5004 | 812.68 | -0.34433 | 0.000703 | 0.010883 | Mthfr | 4 | 1.48E+08 | 1.48E+08 | + | 7315 | protein_co | methylenet | - | 597 | 614 | 722 | 773 | 767 | 838 | 4.292342 | 4.462794 | 4.855666 | 5.810515 | 5.610245 | 5.864092 |
| ENSMUSG0 | 983.4583 | 1028.45 | 969.3123 | 1208.403 | 1187.031 | 1211.262 | -0.27499 | 0.000706 | 0.010913 | Elk3 | 10 | 93247414 | 93311135 | - | 4550 | protein_co | ELK3, mem | ETS | 987 | 1021 | 1040 | 1160 | 1171 | 1249 | 11.4088 | 11.93073 | 11.2447 | 14.01832 | 13.77039 | 14.05148 |
| ENSMUSG0 | 1008.369 | 905.5594 | 1059.719 | 811.5054 | 683.2273 | 828.1966 | 0.356491 | 0.000707 | 0.010913 | Fubp1 | 3 | 1.52E+08 | 1.52E+08 | + | 15500 | protein_co | far upstrea | - | 1012 | 899 | 1137 | 779 | 674 | 854 | 3.433864 | 3.083762 | 3.608732 | 2.763473 | 2.326639 | 2.820313 |
| ENSMUSG0 | 778.1975 | 706.1148 | 843.4881 | 1009.433 | 937.6635 | 984.332 | -0.33201 | 0.00071 | 0.010949 | Eif4e2 | 1 | 87213914 | 87240488 | + | 7497 | protein_co | eukaryotic | - | 781 | 701 | 905 | 969 | 925 | 1015 | 5.478957 | 4.971453 | 5.93864 | 7.10699 | 6.601689 | 6.930261 |
| ENSMUSG0 | 42.8457 | 48.35023 | 43.80546 | 104.1727 | 91.23213 | 84.37131 | -1.05106 | 0.000714 | 0.011008 | Pdzd7 | 19 | 45026906 | 45046614 | - | 4089 | protein_co | PDZ domai | - | 43 | 48 | 47 | 100 | 90 | 87 | 0.553077 | 0.624133 | 0.565466 | 1.344722 | 1.177677 | 1.089114 |
| ENSMUSG0 | 766.2406 | 702.0856 | 602.092 | 943.8047 | 803.8564 | 958.1478 | -0.38684 | 0.000716 | 0.011018 | Plekha1 | 7 | 1.31E+08 | 1.31E+08 | + | 8527 | protein_co | pleckstrin h | - | 769 | 697 | 646 | 906 | 793 | 988 | 4.743123 | 4.345996 | 3.727024 | 5.842267 | 4.97597 | 5.931052 |
| ENSMUSG0 | 569.9475 | 559.0495 | 624.4608 | 794.8377 | 768.3772 | 696.3058 | -0.36497 | 0.000728 | 0.011187 | Nacc1 | 8 | 84670483 | 84687862 | - | 4300 | protein_co | nucleus acc | - | 572 | 555 | 670 | 763 | 758 | 718 | 6.996195 | 6.86242 | 7.665355 | 9.756758 | 9.431952 | 8.547263 |
| ENSMUSG0 | 4880.424 | 5267.153 | 5072.113 | 5502.402 | 5793.24 | 5758.585 | -0.16429 | 0.000736 | 0.011297 | Lrrc59 | 11 | 94629767 | 94645216 | + | 3073 | protein_co | leucine rich | - | 4898 | 5229 | 5442 | 5282 | 5715 | 5938 | 83.82828 | 90.47089 | 87.1208 | 94.51164 | 99.5072 | 98.91194 |
| ENSMUSG0 | 109.6053 | 90.65667 | 93.2031 | 52.08635 | 48.65713 | 43.64033 | 1.023989 | 0.000742 | 0.011379 | Zfp945 | 17 | 22846697 | 22867134 | - | 10173 | protein_co | zinc finger p | zf-C2H2 | 110 | 90 | 100 | 50 | 48 | 45 | 0.568693 | 0.470377 | 0.483589 | 0.270253 | 0.25246 | 0.22643 |
| ENSMUSG0 | 1586.287 | 1695.28 | 1457.697 | 1826.147 | 1965.545 | 1863.927 | -0.25547 | 0.000745 | 0.011416 | Fam91a1 | 15 | 58415468 | 58457740 | + | 6627 | protein_co | family with | - | 1592 | 1683 | 1564 | 1753 | 1939 | 1922 | 12.63457 | 13.50268 | 11.61036 | 14.54502 | 15.65531 | 14.84593 |
| ENSMUSG0 | 1009.365 | 967.0045 | 908.7303 | 1135.482 | 1377.605 | 1118.162 | -0.33201 | 0.000758 | 0.011593 | Fem1c | 18 | 46501746 | 46525971 | - | 5726 | protein_co | fem-1 hom | - | 1013 | 960 | 975 | 1090 | 1359 | 1153 | 9.304487 | 8.914001 | 8.37682 | 10.46706 | 12.69898 | 10.3074 |
| ENSMUSG0 | 138.5012 | 115.8391 | 89.47498 | 50.0029 | 66.90356 | 54.30797 | 1.004663 | 0.000761 | 0.011625 | Ebf2 | 14 | 67233292 | 67430918 | + | 6928 | protein_co | early B cell | COE | 139 | 115 | 96 | 48 | 66 | 56 | 1.055216 | 0.882557 | 0.681694 | 0.380963 | 0.509726 | 0.413763 |
| ENSMUSG0 | 1888.2 | 1754.71 | 1686.044 | 2031.368 | 2065.901 | 2170.379 | -0.23448 | 0.000768 | 0.011721 | Degs1 | 1 | 1.82E+08 | 1.82E+08 | - | 2591 | protein_co | delta(4)-de | - | 1895 | 1742 | 1809 | 1950 | 2038 | 2238 | 38.46592 | 35.7465 | 34.34765 | 41.38249 | 42.08599 | 44.2144 |
| ENSMUSG0 | 157.433 | 232.6855 | 212.5031 | 286.4749 | 305.1208 | 330.6968 | -0.61413 | 0.000779 | 0.011883 | Amigo3 | 9 | 1.08E+08 | 1.08E+08 | + | 2543 | protein_co | adhesion m | - | 158 | 231 | 228 | 275 | 301 | 341 | 3.267722 | 4.829681 | 4.41077 | 5.946149 | 6.333167 | 6.864029 |
| ENSMUSG0 | 1647.069 | 1716.433 | 1697.229 | 1460.501 | 1346.181 | 1467.285 | 0.243731 | 0.000788 | 0.011999 | Id3 | 4 | 1.36E+08 | 1.36E+08 | + | 1791 | protein_co | inhibitor o | bHLH | 1653 | 1704 | 1821 | 1402 | 1328 | 1513 | 48.54132 | 50.58559 | 50.01961 | 43.04294 | 39.67375 | 43.24286 |
| ENSMUSG0 | 12895.56 | 12995.13 | 12957.1 | 13846.64 | 13982.84 | 14363.49 | -0.11922 | 0.00079 | 0.012 | Calr | 8 | 84841850 | 84846934 | - | 2880 | protein_co | calreticulin | - | 12942 | 12901 | 13902 | 13292 | 13794 | 14811 | 236.3433 | 238.1682 | 237.4711 | 253.7741 | 256.2705 | 263.2467 |
| ENSMUSG0 | 266.0419 | 254.846 | 315.0265 | 205.2202 | 188.5464 | 129.9512 | 0.676606 | 0.00079 | 0.012 | Nov | 15 | 54745702 | 54754039 | + | 2869 | protein_co | nephroblas | - | 267 | 253 | 338 | 197 | 186 | 134 | 4.894576 | 4.688596 | 5.795782 | 3.775593 | 3.468832 | 2.390811 |
| ENSMUSG0 | 101.634 | 82.5983 | 69.90233 | 50.0029 | 23.31488 | 33.94248 | 1.245454 | 0.000791 | 0.012013 | Lgr5 | 10 | 1.15E+08 | 1.16E+08 | - | 10264 | protein_co | leucine rich | - | 102 | 82 | 75 | 48 | 23 | 35 | 0.522658 | 0.424766 | 0.359477 | 0.257143 | 0.119898 | 0.174551 |
| ENSMUSG0 | 1763.649 | 1772.842 | 1769.927 | 2113.664 | 2457.185 | 1893.991 | -0.28467 | 0.000801 | 0.012136 | Mia3 | 1 | 1.83E+08 | 1.83E+08 | - | 4431 | protein_co | melanoma | - | 1770 | 1760 | 1899 | 2029 | 2424 | 1953 | 21.00902 | 21.11853 | 21.08381 | 25.17849 | 29.2706 | 22.56169 |
| ENSMUSG0 | 204.2644 | 229.6636 | 178.0179 | 118.7569 | 122.6565 | 135.7699 | 0.696779 | 0.000802 | 0.012149 | Stox2 | 8 | 47180048 | 47446362 | - | 18735 | protein_co | storkhead b | - | 205 | 228 | 191 | 114 | 121 | 140 | 0.575486 | 0.647044 | 0.50154 | 0.33458 | 0.345567 | 0.382512 |
| ENSMUSG0 | 439.4176 | 434.1447 | 417.5499 | 541.6981 | 587.9404 | 564.415 | -0.39219 | 0.000813 | 0.012293 | Slc14a1 | 18 | 78100091 | 78142119 | - | 3995 | protein_co | solute carri | - | 441 | 431 | 448 | 520 | 580 | 582 | 5.805721 | 5.736055 | 5.516799 | 7.157082 | 7.76805 | 7.457226 |
| ENSMUSG0 | 286.9666 | 364.6413 | 250.7163 | 386.4807 | 453.1196 | 474.225 | -0.54354 | 0.000814 | 0.012305 | Slc22a4 | 11 | 53983123 | 54028090 | - | 2553 | protein_co | solute carri | - | 288 | 362 | 269 | 371 | 447 | 489 | 5.933023 | 7.538945 | 5.183551 | 7.990474 | 9.368229 | 9.804583 |
| ENSMUSG0 | 1552.409 | 1461.587 | 1457.697 | 1667.805 | 2029.408 | 1727.187 | -0.27873 | 0.000834 | 0.012581 | Zfp9 | 6 | 1.18E+08 | 1.18E+08 | - | 5389 | protein_co | zinc finger p | zf-C2H2 | 1558 | 1451 | 1564 | 1601 | 2002 | 1781 | 15.20525 | 14.31568 | 14.27758 | 16.33551 | 19.87727 | 16.91714 |
| ENSMUSG0 | 1496.61 | 1576.419 | 1493.114 | 1841.773 | 1803.355 | 1724.278 | -0.23374 | 0.00084 | 0.012651 | 1810055G | 19 | 3708333 | 3717881 | + | 1882 | protein_co | RIKEN cDNA | - | 1502 | 1565 | 1602 | 1768 | 1779 | 1778 | 41.97441 | 44.21275 | 41.87635 | 51.65497 | 50.57747 | 48.35966 |
| ENSMUSG0 | 825.0289 | 826.9903 | 925.5068 | 1082.354 | 1165.744 | 982.3924 | -0.32506 | 0.000841 | 0.012651 | Zfp142 | 1 | 74565127 | 74588246 | - | 11355 | protein_co | zinc finger p | zf-C2H2 | 828 | 821 | 993 | 1039 | 1150 | 1013 | 3.835107 | 3.844225 | 4.302174 | 5.031273 | 5.418905 | 4.566604 |
| ENSMUSG0 | 2867.673 | 3019.875 | 2685.181 | 2509.52 | 2554.5 | 2251.841 | 0.228742 | 0.000841 | 0.012651 | Adamts10 | 17 | 33524204 | 33553782 | + | 5453 | protein_co | a disintegri | - | 2878 | 2998 | 2881 | 2409 | 2520 | 2322 | 27.75809 | 29.23135 | 25.99164 | 24.2913 | 24.72668 | 21.79705 |
| ENSMUSG0 | 3737.54 | 3523.523 | 3624.669 | 3908.56 | 4409.553 | 4109.95 | -0.19121 | 0.000849 | 0.012748 | Tent5a | 9 | 85320439 | 85327348 | - | 5703 | protein_co | terminal nu | - | 3751 | 3498 | 3889 | 3752 | 4350 | 4238 | 34.59219 | 32.61138 | 33.54752 | 36.17503 | 40.81189 | 38.03896 |
| ENSMUSG0 | 1364.088 | 1179.544 | 1248.922 | 1074.021 | 1008.622 | 1024.093 | 0.287895 | 0.000854 | 0.012811 | Gramd1a | 7 | 31130127 | 31155896 | - | 5239 | protein_co | GRAM dom | - | 1369 | 1171 | 1340 | 1031 | 995 | 1056 | 13.74325 | 11.88396 | 12.58295 | 10.82081 | 10.16191 | 10.31779 |
| ENSMUSG0 | 1030.29 | 1081.836 | 1032.69 | 1397.998 | 1274.209 | 1179.259 | -0.29212 | 0.000866 | 0.012973 | Cnksr3 | 10 | 7119063 | 7212237 | - | 3837 | protein_co | Cnksr famil | - | 1034 | 1074 | 1108 | 1342 | 1257 | 1216 | 14.17304 | 14.88214 | 14.20607 | 19.23137 | 17.52848 | 16.22232 |
| ENSMUSG0 | 2198.084 | 2144.534 | 1988.022 | 1731.35 | 1901.683 | 1739.795 | 0.236445 | 0.00087 | 0.013023 | Dab2 | 15 | 6299788 | 6440712 | + | 7342 | protein_co | disabled 2, | - | 2206 | 2129 | 2133 | 1662 | 1876 | 1794 | 15.80249 | 15.4175 | 14.29231 | 12.44704 | 13.6716 | 12.50775 |
| ENSMUSG0 | 2249.898 | 2144.534 | 2173.496 | 2381.388 | 2586.938 | 2646.544 | -0.21363 | 0.000873 | 0.013055 | Cct6a | 5 | 1.3E+08 | 1.3E+08 | + | 3620 | processed_ | chaperonin | - | 2258 | 2129 | 2332 | 2286 | 2552 | 2729 | 32.80573 | 31.26942 | 31.69172 | 34.72299 | 37.72011 | 38.58922 |
| ENSMUSG0 | 883.8172 | 968.0118 | 1064.379 | 772.9615 | 717.6927 | 814.6196 | 0.339368 | 0.000877 | 0.013101 | Stmn1 | 4 | 1.34E+08 | 1.34E+08 | + | 1922 | protein_co | stathmin 1 | - | 887 | 961 | 1142 | 742 | 708 | 840 | 24.27194 | 26.58415 | 29.23066 | 21.22756 | 19.70973 | 22.3716 |
| ENSMUSG0 | 433.4391 | 463.3563 | 403.5694 | 553.1571 | 579.8308 | 577.0222 | -0.39576 | 0.000886 | 0.013218 | Atxn1l | 8 | 1.1E+08 | 1.1E+08 | - | 7526 | protein_co | ataxin 1-lik | - | 435 | 460 | 433 | 531 | 572 | 595 | 3.039901 | 3.249724 | 2.830412 | 3.879536 | 4.066611 | 4.046913 |
| ENSMUSG0 | 359.7046 | 348.5245 | 350.4437 | 491.6952 | 477.4481 | 450.9501 | -0.42348 | 0.000897 | 0.013369 | Tmem168 | 6 | 13580687 | 13608100 | - | 5825 | protein_co | transmemb | - | 361 | 346 | 376 | 472 | 471 | 465 | 3.25946 | 3.158152 | 3.175542 | 4.455491 | 4.326391 | 4.08628 |
| ENSMUSG0 | 1091.071 | 1000.245 | 1121.233 | 1225.071 | 1516.481 | 1265.57 | -0.3186 | 0.000899 | 0.013383 | Coq10b | 1 | 55052770 | 55072702 | + | 4805 | protein_co | coenzyme ( | - | 1095 | 993 | 1203 | 1176 | 1496 | 1305 | 11.98547 | 10.98775 | 12.31681 | 13.45747 | 16.65862 | 13.90235 |
| ENSMUSG0 | 2188.12 | 2199.935 | 2284.408 | 2597.026 | 2428.802 | 2692.124 | -0.20998 | 0.000905 | 0.013456 | Derl1 | 15 | 57869502 | 57892448 | - | 3328 | protein_co | Der1-like d | - | 2196 | 2184 | 2451 | 2493 | 2396 | 2776 | 34.70431 | 34.8917 | 36.23146 | 41.18968 | 38.5216 | 42.69797 |
| ENSMUSG0 | 9729.96 | 9942.015 | 9872.073 | 10289.14 | 11614.86 | 10968.27 | -0.15405 | 0.000913 | 0.013555 | Clu | 14 | 65968483 | 65981547 | + | 3437 | protein_co | clusterin [S | - | 9765 | 9870 | 10592 | 9877 | 11458 | 11310 | 149.4263 | 152.6829 | 151.6088 | 158.0138 | 178.3734 | 168.4435 |
| ENSMUSG0 | 118.573 | 126.9193 | 134.2125 | 183.344 | 196.6559 | 228.8693 | -0.68147 | 0.000918 | 0.013616 | Pappa | 4 | 65124174 | 65357509 | + | 11027 | protein_co | pregnancy- | - | 119 | 126 | 144 | 176 | 194 | 236 | 0.567576 | 0.607528 | 0.642438 | 0.877616 | 0.941337 | 1.095534 |
| ENSMUSG0 | 2058.587 | 2059.921 | 2014.119 | 2153.25 | 2697.43 | 2478.771 | -0.25744 | 0.000919 | 0.013626 | Ptgs2 | 1 | 1.5E+08 | 1.5E+08 | + | 4580 | protein_co | prostaglanc | - | 2066 | 2045 | 2161 | 2067 | 2661 | 2556 | 23.72462 | 23.74 | 23.21214 | 24.81558 | 31.0871 | 28.56712 |
| ENSMUSG0 | 453.3673 | 475.4439 | 438.0546 | 282.308 | 356.819 | 349.1227 | 0.467045 | 0.000927 | 0.013725 | Lhfpl2 | 13 | 94057796 | 94195409 | + | 8415 | protein_co | lipoma HM | - | 455 | 472 | 470 | 271 | 352 | 360 | 2.843751 | 2.982227 | 2.747702 | 1.77078 | 2.238151 | 2.189876 |
| ENSMUSG0 | 183.3397 | 182.3206 | 172.4257 | 105.2144 | 121.6428 | 101.8274 | 0.711157 | 0.000933 | 0.0138 | Arhgap18 | 10 | 26753421 | 26918648 | + | 6602 | protein_co | Rho GTPase | - | 184 | 181 | 185 | 101 | 120 | 105 | 1.465806 | 1.457659 | 1.378549 | 0.841192 | 0.972538 | 0.814113 |
| ENSMUSG0 | 4425.064 | 4384.761 | 4385.206 | 4043.984 | 3800.325 | 3942.177 | 0.162897 | 0.000935 | 0.0138 | Ddx17 | 15 | 79527736 | 79546741 | - | 7643 | protein_co | DEAD (Asp- | - | 4441 | 4353 | 4705 | 3882 | 3749 | 4065 | 30.55985 | 30.28152 | 30.28459 | 27.92808 | 26.24535 | 27.22499 |
| ENSMUSG0 | 2114.386 | 2251.307 | 2217.302 | 2370.971 | 2610.252 | 2687.275 | -0.22043 | 0.000935 | 0.0138 | Arih1 | 9 | 59388258 | 59486618 | - | 7707 | protein_co | ariadne RBI | - | 2122 | 2235 | 2379 | 2276 | 2575 | 2771 | 14.48086 | 15.4186 | 15.18571 | 16.23814 | 17.87692 | 18.40442 |
| ENSMUSG0 | 935.6306 | 950.8878 | 982.3607 | 1229.238 | 1223.524 | 1068.703 | -0.29515 | 0.000939 | 0.01384 | Akap11 | 14 | 78492246 | 78536808 | - | 11015 | protein_co | A kinase (PF | - | 939 | 944 | 1054 | 1180 | 1207 | 1102 | 4.483482 | 4.556594 | 4.70741 | 5.89043 | 5.86305 | 5.121158 |
| ENSMUSG0 | 559.9834 | 568.1152 | 582.5194 | 427.1081 | 422.7088 | 459.6782 | 0.385313 | 0.000947 | 0.01394 | Copg2 | 6 | 30747554 | 30896794 | - | 5121 | protein_co | coatomer p | - | 562 | 564 | 625 | 410 | 417 | 474 | 5.771861 | 5.855677 | 6.004145 | 4.402289 | 4.356945 | 4.737996 |
| ENSMUSG0 | 782.1832 | 730.2899 | 715.7998 | 868.8003 | 955.9099 | 949.4197 | -0.31655 | 0.000952 | 0.013988 | Paip2b | 6 | 83805401 | 83831741 | - | 6335 | protein_co | poly(A) bin | - | 785 | 725 | 768 | 834 | 943 | 979 | 6.517145 | 6.08477 | 5.964039 | 7.238839 | 7.964635 | 7.910559 |
| ENSMUSG0 | 70.74523 | 86.62749 | 97.86326 | 51.04462 | 21.2875 | 35.88205 | 1.239948 | 0.000952 | 0.013988 | Tmem44 | 16 | 30511855 | 30550842 | - | 8893 | protein_co | transmemb | - | 71 | 86 | 105 | 49 | 21 | 37 | 0.419899 | 0.514166 | 0.580854 | 0.302968 | 0.126349 | 0.212973 |
| ENSMUSG0 | 249.1029 | 285.0649 | 244.1921 | 167.7181 | 177.3958 | 173.5916 | 0.584985 | 0.000961 | 0.0141 | Mettl7a1 | 15 | 1E+08 | 1E+08 | + | 2595 | protein_co | methyltran | - | 250 | 283 | 262 | 161 | 175 | 179 | 5.066838 | 5.798316 | 4.96695 | 3.411442 | 3.608291 | 3.530911 |
| ENSMUSG0 | 280.9881 | 237.7219 | 324.3468 | 170.8432 | 200.7107 | 182.3196 | 0.606768 | 0.000962 | 0.014101 | Lgals9 | 11 | 78962974 | 78984946 | - | 2137 | protein_co | lectin, gala | - | 282 | 236 | 348 | 164 | 198 | 188 | 6.940311 | 5.871652 | 8.011256 | 4.21977 | 4.957486 | 4.503233 |
| ENSMUSG0 | 3280.187 | 3447.975 | 3213.643 | 3727.299 | 3822.626 | 3680.335 | -0.17594 | 0.000966 | 0.014144 | Man1a | 10 | 53904785 | 54076609 | - | 9647 | protein_co | mannosida | - | 3292 | 3423 | 3448 | 3578 | 3771 | 3795 | 17.94741 | 18.86546 | 17.58332 | 20.39377 | 20.91535 | 20.13681 |
| ENSMUSG0 | 2013.748 | 1987.396 | 2008.527 | 2267.84 | 2263.57 | 2387.611 | -0.20341 | 0.000977 | 0.014288 | Rapgef2 | 3 | 79062516 | 79286517 | - | 14051 | protein_co | Rap guanin | - | 2021 | 1973 | 2155 | 2177 | 2233 | 2462 | 7.56473 | 7.465736 | 7.545116 | 8.519236 | 8.503198 | 8.969163 |
| ENSMUSG0 | 1508.567 | 1430.361 | 1618.006 | 802.1298 | 721.7475 | 1176.349 | 0.754791 | 0.000983 | 0.014354 | Matn1 | 4 | 1.31E+08 | 1.31E+08 | + | 2001 | protein_co | matrilin 1, | - | 1514 | 1420 | 1736 | 770 | 712 | 1213 | 39.79359 | 37.73063 | 42.6804 | 21.1589 | 19.03854 | 31.03022 |
| ENSMUSG0 | 1609.205 | 1660.024 | 1752.218 | 1424.041 | 1477.96 | 1318.908 | 0.251016 | 0.000994 | 0.014504 | Fto | 8 | 91313525 | 91668439 | + | 13031 | protein_co | fat mass an | - | 1615 | 1648 | 1880 | 1367 | 1458 | 1360 | 6.518222 | 6.724071 | 7.09751 | 5.768199 | 5.986605 | 5.342349 |
| ENSMUSG0 | 46.83135 | 79.57641 | 63.37811 | 30.21008 | 26.35595 | 13.57699 | 1.439467 | 0.000997 | 0.014523 | Cap2 | 13 | 46501848 | 46650281 | + | 4159 | protein_co | CAP, adeny | - | 47 | 79 | 68 | 29 | 26 | 14 | 0.594351 | 1.009929 | 0.804352 | 0.383406 | 0.334492 | 0.17231 |
| ENSMUSG0 | 934.6342 | 970.0264 | 945.0795 | 1228.196 | 1075.525 | 1173.44 | -0.28705 | 0.001004 | 0.014611 | N4bp2 | 5 | 65763521 | 65830108 | + | 14946 | protein_co | NEDD4 bin | - | 938 | 963 | 1014 | 1179 | 1061 | 1210 | 3.300747 | 3.425738 | 3.337635 | 4.337488 | 3.798317 | 4.144112 |
| ENSMUSG0 | 88.68064 | 114.8318 | 97.86326 | 167.7181 | 147.9988 | 196.8664 | -0.76714 | 0.001008 | 0.014652 | Lrrc8a | 2 | 30237715 | 30263790 | + | 4308 | protein_co | leucine rich | - | 89 | 114 | 105 | 161 | 146 | 203 | 1.086547 | 1.406961 | 1.199056 | 2.054942 | 1.813335 | 2.412078 |
| ENSMUSG0 | 1053.207 | 940.8148 | 932.031 | 1160.484 | 1210.346 | 1191.866 | -0.2843 | 0.001012 | 0.014692 | Dido1 | 2 | 1.81E+08 | 1.81E+08 | - | 13307 | protein_co | death indu | - | 1057 | 934 | 1000 | 1114 | 1194 | 1229 | 4.177622 | 3.73181 | 3.696968 | 4.603143 | 4.800926 | 4.727623 |
| ENSMUSG0 | 2514.943 | 2564.577 | 2509.028 | 2847.04 | 3015.729 | 2798.8 | -0.19079 | 0.001023 | 0.014842 | Huwe1 | X | 1.52E+08 | 1.52E+08 | + | 23507 | protein_co | HECT, UBA | - | 2524 | 2546 | 2692 | 2733 | 2975 | 2886 | 5.647113 | 5.758561 | 5.63383 | 6.392811 | 6.771589 | 6.284492 |
| ENSMUSG0 | 3651.849 | 3861.974 | 3773.794 | 3490.827 | 3277.261 | 3164.409 | 0.184726 | 0.001033 | 0.014974 | Col15a1 | 4 | 47208161 | 47313167 | + | 6349 | protein_co | collagen, ty | - | 3665 | 3834 | 4049 | 3351 | 3233 | 3263 | 30.36009 | 32.10699 | 31.37389 | 29.02141 | 27.2459 | 26.3077 |
| ENSMUSG0 | 936.627 | 971.0337 | 996.3412 | 1253.198 | 1079.58 | 1225.808 | -0.2931 | 0.001036 | 0.01499 | Vat1 | 11 | 1.01E+08 | 1.01E+08 | - | 2766 | protein_co | vesicle ami | - | 940 | 964 | 1069 | 1203 | 1065 | 1264 | 17.87352 | 18.5301 | 19.01303 | 23.91459 | 20.60147 | 23.39193 |
| ENSMUSG0 | 81.70576 | 125.912 | 104.3875 | 40.62735 | 41.5613 | 65.94539 | 1.072206 | 0.001037 | 0.015001 | Ackr4 | 9 | 1.04E+08 | 1.04E+08 | - | 5380 | protein_co | atypical ch | - | 82 | 125 | 112 | 39 | 41 | 68 | 0.801615 | 1.235323 | 1.024145 | 0.398595 | 0.407758 | 0.64699 |
| ENSMUSG0 | 129.5335 | 151.0945 | 196.6585 | 78.12953 | 84.13629 | 104.7368 | 0.837855 | 0.00104 | 0.015026 | Tmem132a | 19 | 10857822 | 10869940 | - | 4069 | protein_co | transmemb | - | 130 | 150 | 211 | 75 | 83 | 108 | 1.680312 | 1.960001 | 2.55106 | 1.013498 | 1.091418 | 1.358649 |
| ENSMUSG0 | 2095.454 | 1967.25 | 2087.75 | 2195.961 | 2556.527 | 2474.892 | -0.23288 | 0.001042 | 0.015036 | Med13 | 11 | 86267033 | 86357602 | - | 10840 | protein_co | mediator c | - | 2103 | 1953 | 2240 | 2108 | 2522 | 2552 | 10.20339 | 9.579123 | 10.16587 | 10.69278 | 12.44849 | 12.05098 |
| ENSMUSG0 | 447.3888 | 532.8598 | 570.403 | 702.124 | 664.9808 | 666.2424 | -0.39029 | 0.001057 | 0.015237 | Lbp | 2 | 1.58E+08 | 1.58E+08 | + | 4831 | protein_co | lipopolysac | - | 449 | 529 | 612 | 674 | 656 | 687 | 4.88814 | 5.82199 | 6.232184 | 7.671359 | 7.265535 | 7.279319 |
| ENSMUSG0 | 498.2058 | 530.8452 | 475.3358 | 408.357 | 365.9422 | 345.2435 | 0.426549 | 0.001062 | 0.015275 | Firre | X | 50555744 | 50635321 | - | 8247 | processed_ | functional | - | 500 | 527 | 510 | 392 | 361 | 356 | 3.188661 | 3.397562 | 3.042286 | 2.613602 | 2.342136 | 2.209658 |
| ENSMUSG0 | 973.4942 | 865.2676 | 952.5357 | 767.7528 | 779.5278 | 674.9705 | 0.32951 | 0.001063 | 0.015275 | Odf2 | 2 | 29889221 | 29931746 | + | 9919 | protein_co | outer dens | - | 977 | 859 | 1022 | 737 | 769 | 696 | 5.180373 | 4.604453 | 5.068844 | 4.085536 | 4.148196 | 3.591802 |
| ENSMUSG0 | 958.548 | 976.0702 | 952.5357 | 1066.728 | 1211.36 | 1261.691 | -0.29449 | 0.001064 | 0.015275 | Rab21 | 10 | 1.15E+08 | 1.15E+08 | - | 5047 | protein_co | RAB21, me | - | 962 | 969 | 1022 | 1024 | 1195 | 1301 | 10.02481 | 10.20806 | 9.96193 | 11.1562 | 12.6688 | 13.19517 |
| ENSMUSG0 | 927.6593 | 912.6105 | 1012.186 | 1088.605 | 1209.333 | 1190.896 | -0.29035 | 0.001065 | 0.015275 | Dusp7 | 9 | 1.06E+08 | 1.06E+08 | + | 3356 | protein_co | dual specifi | - | 931 | 906 | 1086 | 1045 | 1193 | 1228 | 14.59023 | 14.35354 | 15.91966 | 17.12158 | 19.02038 | 18.73042 |
| ENSMUSG0 | 213.2321 | 190.379 | 178.0179 | 105.2144 | 115.5607 | 136.7397 | 0.700558 | 0.001066 | 0.015275 | Chodl | 16 | 78930948 | 78951733 | + | 5491 | protein_co | chondrolec | - | 214 | 189 | 191 | 101 | 114 | 141 | 2.04973 | 1.830051 | 1.711228 | 1.011392 | 1.110847 | 1.314434 |
| ENSMUSG0 | 545.0372 | 492.5679 | 458.5593 | 615.6607 | 747.0897 | 618.723 | -0.40568 | 0.00107 | 0.015315 | Rrp12 | 19 | 41862852 | 41896153 | - | 4319 | protein_co | ribosomal I | - | 547 | 489 | 492 | 591 | 737 | 638 | 6.660985 | 6.01975 | 5.604125 | 7.524086 | 9.130301 | 7.56151 |
| ENSMUSG0 | 260671.3 | 248277.4 | 264721 | 241796.3 | 237439.7 | 239475.8 | 0.10631 | 0.001078 | 0.015417 | Col11a1 | 3 | 1.14E+08 | 1.14E+08 | + | 12744 | protein_co | collagen, ty | - | 261610 | 246479 | 284026 | 232111 | 234233 | 246937 | 1079.65 | 1028.317 | 1096.423 | 1001.473 | 983.4293 | 991.8625 |
| ENSMUSG0 | 2070.543 | 1922.929 | 2134.351 | 2403.264 | 2305.132 | 2393.43 | -0.21255 | 0.001086 | 0.015518 | Dst | 1 | 33908225 | 34308661 | + | 41155 | protein_co | dystonin [S | - | 2078 | 1909 | 2290 | 2307 | 2274 | 2468 | 2.655567 | 2.466244 | 2.737403 | 3.082297 | 2.956437 | 3.069684 |
| ENSMUSG0 | 260.0634 | 227.649 | 260.0367 | 331.2692 | 382.1612 | 345.2435 | -0.50128 | 0.001112 | 0.015861 | Rabif | 1 | 1.34E+08 | 1.35E+08 | + | 2861 | protein_co | RAB interac | - | 261 | 226 | 279 | 318 | 377 | 356 | 4.797964 | 4.199943 | 4.79747 | 6.111654 | 7.050571 | 6.369469 |
| ENSMUSG0 | 1650.058 | 1583.47 | 1686.976 | 1913.692 | 1911.82 | 1894.96 | -0.21709 | 0.001113 | 0.015861 | Ywhah | 5 | 33018816 | 33027966 | + | 3709 | protein_co | tyrosine 3-r | - | 1656 | 1572 | 1810 | 1837 | 1886 | 1954 | 23.48215 | 22.53453 | 24.00754 | 27.2334 | 27.20731 | 26.96738 |
| ENSMUSG0 | 61.77752 | 60.43778 | 81.0867 | 26.04318 | 9.123213 | 36.85184 | 1.495326 | 0.001118 | 0.015919 | Egflam | 15 | 7206120 | 7398395 | - | 6121 | protein_co | EGF-like, fil | - | 62 | 60 | 87 | 25 | 9 | 38 | 0.532726 | 0.521173 | 0.699234 | 0.224578 | 0.078672 | 0.317784 |
| ENSMUSG0 | 413.5108 | 375.7215 | 347.6476 | 301.0591 | 218.9571 | 268.6305 | 0.527552 | 0.001133 | 0.016113 | Man2c1 | 9 | 57130690 | 57142722 | + | 7052 | protein_co | mannosida | - | 415 | 373 | 373 | 289 | 216 | 277 | 3.095068 | 2.81222 | 2.602091 | 2.253383 | 1.638862 | 2.01066 |

| gene_id | H1 | H2 | H3 | M1 | M2 | M3 | log2FoldCh | pvalue | padj | gene_name | gene_chr | gene_start | gene_end | gene_stran | gene_lengt | gene_bioty | gene_descr | tf_family | H1_readco | H2_readco | H3_readco | M1_readco | M2_readco | M3_readco | H1_fpkm | H2_fpkm | H3_fpkm | M1_fpkm | M2_fpkm | M3_fpkm |
|---|---|---|---|---|---|---|---|---|---|---|---|---|---|---|---|---|---|---|---|---|---|---|---|---|---|---|---|---|---|---|
| ENSMUSG0 | 225.189 | 217.576 | 171.4937 | 117.7152 | 80.08153 | 153.2261 | 0.805458 | 0.001143 | 0.016244 | Cryab | 9 | 50751325 | 50756636 | + | 1479 | protein_co | crystallin, a | - | 226 | 216 | 184 | 113 | 79 | 158 | 8.036642 | 7.764945 | 6.12034 | 4.201068 | 2.857984 | 5.468396 |
| ENSMUSG0 | 2647.466 | 2634.08 | 2798.889 | 3086.637 | 3180.96 | 2952.996 | -0.19011 | 0.001145 | 0.016251 | Dag1 | 9 | 1.08E+08 | 1.08E+08 | - | 6357 | protein_co | dystroglyca | - | 2657 | 2615 | 3003 | 2963 | 3138 | 3045 | 21.98233 | 21.87118 | 23.23962 | 25.62884 | 26.41202 | 24.51919 |
| ENSMUSG0 | 394.579 | 412.9915 | 329.007 | 507.3211 | 521.0368 | 501.379 | -0.42948 | 0.001151 | 0.016325 | Mnt | 11 | 74830920 | 74845725 | + | 4627 | protein_co | max bindin | bHLH | 396 | 410 | 353 | 487 | 514 | 517 | 4.501218 | 4.711261 | 3.753195 | 5.787339 | 5.943803 | 5.719553 |
| ENSMUSG0 | 217.2177 | 170.2331 | 163.1054 | 258.3483 | 282.8196 | 287.0564 | -0.59008 | 0.001167 | 0.016536 | Gpnmb | 6 | 49036546 | 49070929 | + | 6514 | protein_co | glycoprotei | - | 218 | 169 | 175 | 248 | 279 | 296 | 1.760123 | 1.379405 | 1.321649 | 2.093406 | 2.291697 | 2.326029 |
| ENSMUSG0 | 451.3745 | 360.6121 | 357.8999 | 492.7369 | 525.0916 | 572.1733 | -0.44344 | 0.001183 | 0.016735 | Cav2 | 6 | 17281185 | 17289115 | + | 3335 | protein_co | caveolin 2 [ | - | 453 | 358 | 384 | 473 | 518 | 590 | 7.143922 | 5.707422 | 5.664496 | 7.798566 | 8.310645 | 9.055809 |
| ENSMUSG0 | 1592.266 | 1673.119 | 1830.509 | 2003.241 | 1952.367 | 2037.519 | -0.23364 | 0.001188 | 0.016793 | Nomo1 | 7 | 46033698 | 46084212 | + | 5936 | protein_co | nodal mod | - | 1598 | 1661 | 1964 | 1923 | 1926 | 2101 | 14.1585 | 14.87745 | 16.27697 | 17.81291 | 17.36054 | 18.11771 |
| ENSMUSG0 | 1166.798 | 1231.923 | 1222.825 | 1529.255 | 1410.043 | 1382.914 | -0.25485 | 0.00119 | 0.016793 | Pik3ca | 3 | 32397671 | 32468486 | + | 9088 | protein_co | phosphatic | - | 1171 | 1223 | 1312 | 1468 | 1391 | 1426 | 6.776773 | 7.155021 | 7.102176 | 8.881927 | 8.189542 | 8.031974 |
| ENSMUSG0 | 799.1222 | 777.6328 | 662.6741 | 914.6363 | 925.4992 | 968.8154 | -0.32772 | 0.001191 | 0.016793 | Creb3l1 | 2 | 91982328 | 92024502 | - | 2603 | protein_co | cAMP respc | TF_bZIP | 802 | 772 | 711 | 878 | 913 | 999 | 16.20446 | 15.7687 | 13.43759 | 18.54684 | 18.76711 | 19.64547 |
| ENSMUSG0 | 199.2823 | 198.4374 | 200.3867 | 279.1828 | 309.1755 | 277.3586 | -0.53338 | 0.001204 | 0.016957 | Cdyl | 13 | 35659833 | 35874063 | + | 4392 | protein_co | chromodor | - | 200 | 197 | 215 | 268 | 305 | 286 | 2.394981 | 2.384826 | 2.408253 | 3.355227 | 3.71568 | 3.333303 |
| ENSMUSG0 | 187.3254 | 208.5103 | 200.3867 | 106.2562 | 154.0809 | 98.91809 | 0.731317 | 0.001211 | 0.017042 | Slc30a6 | 17 | 74395621 | 74424221 | + | 3062 | protein_co | solute carri | - | 188 | 207 | 215 | 102 | 152 | 102 | 3.229141 | 3.59433 | 3.454293 | 1.831658 | 2.656068 | 1.705164 |
| ENSMUSG0 | 163.4115 | 169.2258 | 185.4742 | 92.71371 | 119.6155 | 101.8274 | 0.721951 | 0.001236 | 0.01738 | Sox7 | 14 | 63943673 | 63950732 | + | 3299 | protein_co | SRY (sex det | HMG | 164 | 168 | 199 | 89 | 118 | 105 | 2.614543 | 2.70757 | 2.967541 | 1.483396 | 1.913817 | 1.629214 |
| ENSMUSG0 | 5631.719 | 5026.409 | 5624.807 | 6284.739 | 5828.719 | 6328.818 | -0.17956 | 0.001239 | 0.017402 | Rps23 | 13 | 90922958 | 90924950 | + | 1268 | protein_co | ribosomal p | - | 5652 | 4990 | 6035 | 6033 | 5750 | 6526 | 234.4322 | 209.2349 | 234.1445 | 261.6155 | 242.6327 | 263.4504 |
| ENSMUSG0 | 846.9499 | 803.8225 | 959.992 | 1055.269 | 1082.621 | 1084.22 | -0.30296 | 0.001242 | 0.017418 | Smim7 | 8 | 72565188 | 72571088 | - | 3266 | protein_co | small integ | - | 850 | 798 | 1030 | 1013 | 1068 | 1118 | 13.68791 | 12.99091 | 15.51483 | 17.05465 | 17.49669 | 17.52253 |
| ENSMUSG0 | 1794.537 | 1825.221 | 1939.557 | 2038.66 | 2321.351 | 2154.863 | -0.22869 | 0.00126 | 0.017663 | Camsap2 | 1 | 1.36E+08 | 1.36E+08 | - | 9195 | protein_co | calmodulir | - | 1801 | 1812 | 2081 | 1957 | 2290 | 2222 | 10.3014 | 10.47754 | 11.13387 | 11.70277 | 13.32553 | 12.36982 |
| ENSMUSG0 | 191.311 | 161.1674 | 291.7257 | 320.8519 | 314.244 | 385.0047 | -0.66162 | 0.001274 | 0.017827 | Ccdc85b | 19 | 5456789 | 5457397 | - | 609 | protein_co | coiled-coil | - | 192 | 160 | 313 | 308 | 310 | 397 | 16.58129 | 13.96868 | 25.28442 | 27.80884 | 27.23612 | 33.36908 |
| ENSMUSG0 | 775.2083 | 904.5521 | 765.1975 | 1019.851 | 1066.402 | 961.0571 | -0.31798 | 0.001283 | 0.017936 | Gas2l1 | 11 | 5054132 | 5065327 | - | 6100 | protein_co | growth arre | - | 778 | 898 | 821 | 979 | 1052 | 991 | 6.707862 | 7.827071 | 6.621238 | 8.824748 | 9.227557 | 8.316008 |
| ENSMUSG0 | 205.2608 | 218.5833 | 161.2414 | 286.4749 | 342.6273 | 257.9629 | -0.60127 | 0.001286 | 0.017956 | Zfp334 | 2 | 1.65E+08 | 1.65E+08 | - | 8205 | protein_co | zinc finger p | zf-C2H2 | 206 | 217 | 173 | 275 | 338 | 266 | 1.320453 | 1.406157 | 1.037274 | 1.842908 | 2.204139 | 1.659488 |
| ENSMUSG0 | 2354.521 | 2201.95 | 2507.163 | 2685.572 | 2631.54 | 2890.93 | -0.21649 | 0.001289 | 0.017956 | Dync1h1 | 12 | 1.11E+08 | 1.11E+08 | + | 15658 | protein_co | dynein cyto | - | 2363 | 2186 | 2690 | 2578 | 2596 | 2981 | 7.937099 | 7.422781 | 8.451658 | 9.053074 | 8.870931 | 9.745335 |
| ENSMUSG0 | 155.4402 | 202.4666 | 170.5617 | 238.5555 | 293.9702 | 266.6909 | -0.59738 | 0.001291 | 0.017956 | Arl4c | 1 | 88673125 | 88702221 | - | 5492 | protein_co | ADP-ribosy | - | 156 | 201 | 183 | 229 | 290 | 275 | 1.493924 | 1.94589 | 1.639255 | 2.292738 | 2.825324 | 2.563145 |
| ENSMUSG0 | 215957.3 | 207766 | 224540.3 | 238278.4 | 227469.1 | 236176.6 | -0.11473 | 0.001291 | 0.017956 | Col9a1 | 1 | 24177610 | 24252684 | + | 5068 | protein_co | collagen, ty | - | 216735 | 206261 | 240915 | 228734 | 224397 | 243535 | 2249.193 | 2163.881 | 2338.585 | 2481.667 | 2369.088 | 2459.778 |
| ENSMUSG0 | 1047.229 | 1023.413 | 1067.176 | 1149.025 | 1302.592 | 1368.367 | -0.28409 | 0.001291 | 0.017956 | Baiap2l1 | 5 | 1.44E+08 | 1.44E+08 | - | 3366 | protein_co | BAI1-associ | - | 1051 | 1016 | 1145 | 1103 | 1285 | 1411 | 16.42188 | 16.04842 | 16.73467 | 18.01818 | 20.42631 | 21.45774 |
| ENSMUSG0 | 733.359 | 731.2972 | 695.2951 | 865.6752 | 938.6772 | 866.0182 | -0.30625 | 0.001304 | 0.018118 | Nfkbiz | 16 | 55811375 | 55838899 | - | 5018 | protein_co | nuclear fac | - | 736 | 726 | 746 | 831 | 926 | 893 | 7.714033 | 7.692345 | 7.313648 | 9.105836 | 9.873728 | 9.109444 |
| ENSMUSG0 | 1108.01 | 1003.267 | 1015.914 | 1267.782 | 1179.935 | 1348.971 | -0.28018 | 0.001314 | 0.018242 | Lpin2 | 17 | 71182560 | 71249817 | + | 12233 | protein_co | lipin 2 [Sou | - | 1112 | 996 | 1090 | 1217 | 1164 | 1391 | 4.780861 | 4.328916 | 4.383484 | 5.470249 | 5.091208 | 5.820566 |
| ENSMUSG0 | 309.884 | 320.3202 | 319.6866 | 186.4691 | 208.8202 | 255.0535 | 0.545397 | 0.001316 | 0.018253 | Csrp2 | 10 | 1.11E+08 | 1.11E+08 | + | 2171 | protein_co | cysteine an | - | 311 | 318 | 343 | 179 | 206 | 263 | 7.534161 | 7.787895 | 7.77249 | 4.533594 | 5.077012 | 6.201075 |
| ENSMUSG0 | 436.4283 | 467.3855 | 411.9577 | 517.7383 | 621.3921 | 596.4179 | -0.40025 | 0.001322 | 0.018307 | Krr1 | 10 | 1.12E+08 | 1.12E+08 | + | 5958 | protein_co | KRR1, smal | - | 438 | 464 | 442 | 497 | 613 | 615 | 3.86641 | 4.140667 | 3.64962 | 4.586753 | 5.505044 | 5.283792 |
| ENSMUSG0 | 350.7369 | 302.1889 | 258.1726 | 216.6792 | 168.2726 | 219.1715 | 0.59195 | 0.001337 | 0.018505 | Ldb2 | 5 | 44472132 | 44799680 | - | 3336 | protein_co | LIM domair | - | 352 | 300 | 277 | 208 | 166 | 226 | 5.549463 | 4.781322 | 4.084883 | 3.428363 | 2.662459 | 3.467795 |
| ENSMUSG0 | 812.0755 | 770.5817 | 758.6733 | 950.0551 | 1018.759 | 913.5377 | -0.29993 | 0.001342 | 0.018542 | Selenoi | 5 | 30232581 | 30272427 | + | 13135 | protein_co | selenoprot | - | 815 | 765 | 814 | 912 | 1005 | 942 | 3.263337 | 3.096593 | 3.048739 | 3.817809 | 4.093896 | 3.671064 |
| ENSMUSG0 | 1025.308 | 1055.647 | 1069.04 | 1359.454 | 1212.374 | 1223.869 | -0.26865 | 0.001349 | 0.018628 | Eml1 | 12 | 1.08E+08 | 1.09E+08 | + | 7436 | protein_co | echinoderr | - | 1029 | 1048 | 1147 | 1305 | 1196 | 1262 | 7.277971 | 7.493327 | 7.588395 | 9.649851 | 8.605827 | 8.687425 |
| ENSMUSG0 | 111.5981 | 105.7661 | 105.3195 | 72.92089 | 31.4244 | 51.39862 | 1.05218 | 0.001358 | 0.018737 | Gm38190 | 1 | 1.65E+08 | 1.65E+08 | - | 4755 | sense_intro | predicted g | - | 112 | 105 | 113 | 70 | 31 | 53 | 1.238802 | 1.174063 | 1.169106 | 0.809463 | 0.348829 | 0.570554 |
| ENSMUSG0 | 266.0419 | 222.6125 | 349.5116 | 167.7181 | 167.2589 | 201.7153 | 0.64361 | 0.001409 | 0.019378 | Dbn1 | 13 | 55473429 | 55488111 | - | 4514 | protein_co | drebrin 1 [S | - | 267 | 221 | 375 | 161 | 165 | 208 | 3.110886 | 2.603056 | 4.086915 | 1.961163 | 1.955794 | 2.358701 |
| ENSMUSG0 | 332.8015 | 318.3057 | 397.0452 | 522.947 | 452.1059 | 456.7688 | -0.44875 | 0.001409 | 0.019378 | Zfp64 | 2 | 1.69E+08 | 1.69E+08 | - | 5918 | protein_co | zinc finger p | zf-C2H2 | 334 | 316 | 426 | 502 | 446 | 471 | 2.968287 | 2.838997 | 3.541282 | 4.664211 | 4.032373 | 4.073962 |
| ENSMUSG0 | 136.5084 | 132.9631 | 73.63045 | 67.71226 | 46.62975 | 53.33819 | 1.032296 | 0.001409 | 0.019378 | Fst | 13 | 1.14E+08 | 1.14E+08 | - | 4589 | protein_co | follistatin [S | - | 137 | 132 | 79 | 65 | 46 | 55 | 1.570134 | 1.529356 | 0.846905 | 0.778834 | 0.53634 | 0.613502 |
| ENSMUSG0 | 5957.545 | 5535.094 | 5807.485 | 6723.306 | 6289.948 | 6265.782 | -0.1561 | 0.001418 | 0.019474 | Pdia6 | 12 | 17266545 | 17284770 | + | 3745 | protein_co | protein dis | - | 5979 | 5495 | 6231 | 6454 | 6205 | 6461 | 83.96747 | 78.0133 | 81.85248 | 94.76034 | 88.65246 | 88.31185 |
| ENSMUSG0 | 829.0145 | 685.9688 | 651.4897 | 857.3414 | 963.0058 | 941.6614 | -0.35098 | 0.001423 | 0.019528 | Zbtb1 | 12 | 76370266 | 76396950 | + | 12190 | protein_co | zinc finger a | ZBTB | 832 | 681 | 699 | 823 | 950 | 971 | 3.589665 | 2.970272 | 2.820976 | 3.712322 | 4.169853 | 4.077431 |
| ENSMUSG0 | 370.6651 | 407.955 | 402.6374 | 498.9873 | 500.763 | 558.5963 | -0.39995 | 0.001426 | 0.019547 | Adra2a | 19 | 54045182 | 54048982 | + | 3801 | protein_co | adrenergic | - | 372 | 405 | 432 | 479 | 494 | 576 | 5.147299 | 5.665131 | 5.591287 | 6.929264 | 6.953923 | 7.757034 |
| ENSMUSG0 | 218.2142 | 198.4374 | 195.7265 | 153.1339 | 115.5607 | 117.344 | 0.666531 | 0.001437 | 0.01968 | Atp1a2 | 1 | 1.72E+08 | 1.72E+08 | - | 7038 | protein_co | ATPase, Na- | - | 219 | 197 | 210 | 147 | 114 | 121 | 1.63655 | 1.488229 | 1.467898 | 1.148464 | 0.866675 | 0.88005 |
| ENSMUSG0 | 604.8219 | 536.889 | 647.7616 | 488.57 | 367.9696 | 468.4063 | 0.434033 | 0.00144 | 0.019683 | Cnn2 | 10 | 79988584 | 79996062 | + | 2460 | protein_co | calponin 2 | - | 607 | 533 | 695 | 469 | 363 | 483 | 12.97741 | 11.5198 | 13.89875 | 10.48304 | 7.895367 | 10.05039 |
| ENSMUSG0 | 32.88159 | 25.18241 | 59.64999 | 12.50072 | 14.19166 | 3.879141 | 1.952193 | 0.001441 | 0.019683 | Myoz2 | 3 | 1.23E+08 | 1.23E+08 | - | 3423 | protein_co | myozenin 2 | - | 33 | 25 | 64 | 12 | 14 | 4 | 0.507039 | 0.388317 | 0.919812 | 0.192763 | 0.218838 | 0.059817 |
| ENSMUSG0 | 1183.737 | 1272.215 | 1371.95 | 1077.146 | 1073.498 | 1016.335 | 0.273985 | 0.001444 | 0.019692 | Fzd9 | 5 | 1.35E+08 | 1.35E+08 | - | 2293 | protein_co | frizzled clas | - | 1188 | 1263 | 1472 | 1034 | 1059 | 1048 | 27.24876 | 29.28546 | 31.58127 | 24.7951 | 24.71114 | 23.39528 |
| ENSMUSG0 | 1291.35 | 1290.347 | 1376.61 | 1542.798 | 1469.851 | 1734.946 | -0.2624 | 0.001444 | 0.019692 | Ptpn14 | 1 | 1.9E+08 | 1.9E+08 | + | 13413 | protein_co | protein tyr | - | 1296 | 1281 | 1477 | 1481 | 1450 | 1789 | 5.081751 | 5.077805 | 5.41727 | 6.071257 | 5.784195 | 6.827402 |
| ENSMUSG0 | 3257.27 | 2958.429 | 3210.847 | 3500.203 | 3460.739 | 3837.44 | -0.19608 | 0.001446 | 0.019693 | Rplp2 | 7 | 1.41E+08 | 1.41E+08 | + | 2072 | protein_co | ribosomal p | - | 3269 | 2937 | 3445 | 3360 | 3414 | 3957 | 82.97734 | 75.36453 | 81.79474 | 89.16594 | 88.16061 | 97.75689 |
| ENSMUSG0 | 2089.475 | 2042.797 | 2073.769 | 2402.223 | 2825.155 | 2199.473 | -0.2589 | 0.001466 | 0.019948 | Fnip1 | 11 | 54438199 | 54518235 | + | 7119 | protein_co | folliculin ir | - | 2097 | 2028 | 2225 | 2306 | 2787 | 2268 | 15.49222 | 15.14613 | 15.37577 | 17.81106 | 20.94685 | 16.30779 |
| ENSMUSG0 | 179.3541 | 157.1382 | 169.6296 | 101.0475 | 107.4512 | 103.767 | 0.696721 | 0.00147 | 0.019981 | Mis18bp1 | 12 | 65132734 | 65172604 | - | 7990 | protein_co | MIS18 binc | - | 180 | 156 | 182 | 97 | 106 | 107 | 1.184841 | 1.03808 | 1.1206 | 0.667536 | 0.709839 | 0.685501 |
| ENSMUSG0 | 5756.27 | 5561.283 | 5677.001 | 5937.844 | 6630.548 | 6412.22 | -0.15954 | 0.001476 | 0.020039 | Cltc | 11 | 86694351 | 86757565 | - | 8499 | protein_co | clathrin, he | - | 5777 | 5521 | 6091 | 5700 | 6541 | 6612 | 35.74941 | 34.53844 | 35.25711 | 36.87708 | 41.17913 | 39.82319 |
| ENSMUSG0 | 137.5048 | 132.9631 | 137.0086 | 89.58853 | 57.78035 | 81.46196 | 0.832715 | 0.001501 | 0.020339 | Rab24 | 13 | 55319743 | 55321946 | - | 2015 | protein_co | RAB24, me | - | 138 | 132 | 147 | 86 | 57 | 84 | 3.601956 | 3.482986 | 3.588956 | 2.346783 | 1.513563 | 2.133906 |
| ENSMUSG0 | 46.83135 | 40.29185 | 39.1453 | 13.54245 | 18.24643 | 9.697852 | 1.607591 | 0.001501 | 0.020339 | Gstt1 | 10 | 75783813 | 75798584 | - | 1528 | protein_co | glutathione | - | 47 | 40 | 42 | 13 | 18 | 10 | 1.61774 | 1.39184 | 1.352234 | 0.46781 | 0.630304 | 0.335002 |
| ENSMUSG0 | 2486.047 | 2262.388 | 2435.397 | 2789.745 | 2615.321 | 2863.776 | -0.2029 | 0.001503 | 0.020339 | Rpl27 | 11 | 1.01E+08 | 1.01E+08 | + | 1535 | protein_co | ribosomal p | - | 2495 | 2246 | 2613 | 2678 | 2580 | 2953 | 85.4863 | 77.79544 | 83.74462 | 95.92938 | 89.93156 | 98.47504 |
| ENSMUSG0 | 289.9558 | 207.5031 | 312.2304 | 186.4691 | 156.1083 | 180.38 | 0.631531 | 0.001506 | 0.02036 | Tie1 | 4 | 1.18E+08 | 1.18E+08 | - | 4917 | protein_co | tyrosine kir | - | 291 | 206 | 335 | 179 | 154 | 186 | 3.112627 | 2.227511 | 3.351741 | 2.001715 | 1.675797 | 1.93635 |
| ENSMUSG0 | 1461.736 | 1533.105 | 1739.17 | 1882.401 | 1801.328 | 1950.238 | -0.25072 | 0.001509 | 0.020389 | Osbp | 19 | 11965844 | 11994105 | + | 4578 | protein_co | oxysterol b | - | 1467 | 1522 | 1866 | 1807 | 1777 | 2011 | 16.85344 | 17.67631 | 20.05219 | 21.7036 | 20.76885 | 22.48575 |
| ENSMUSG0 | 981.4655 | 1048.596 | 1057.855 | 1170.901 | 1331.989 | 1227.748 | -0.27273 | 0.001514 | 0.020432 | Acvr1 | 2 | 58388644 | 58567157 | - | 4829 | protein_co | activin A re | - | 985 | 1041 | 1135 | 1124 | 1314 | 1266 | 10.72787 | 11.46163 | 11.56284 | 12.79849 | 14.55925 | 13.41985 |
| ENSMUSG0 | 1336.188 | 1168.464 | 1286.203 | 1611.552 | 1466.81 | 1463.406 | -0.26036 | 0.001517 | 0.020455 | Rab18 | 18 | 6765205 | 6790231 | + | 2257 | protein_co | RAB18, me | - | 1341 | 1160 | 1380 | 1547 | 1447 | 1509 | 31.24867 | 27.3262 | 30.07969 | 37.68844 | 34.30345 | 34.22384 |
| ENSMUSG0 | 1352.131 | 1368.916 | 1549.968 | 1592.801 | 1734.424 | 1770.828 | -0.25515 | 0.001522 | 0.020494 | Chd9 | 8 | 90828352 | 91054516 | + | 23148 | protein_co | chromodor | - | 1357 | 1359 | 1663 | 1529 | 1711 | 1826 | 3.083193 | 3.121467 | 3.53431 | 3.63198 | 3.954917 | 4.037926 |
| ENSMUSG0 | 569.9475 | 559.0495 | 592.7717 | 443.7757 | 475.4207 | 403.4306 | 0.381085 | 0.001527 | 0.020545 | Fam129a | 1 | 1.52E+08 | 1.52E+08 | + | 7380 | protein_co | family with | - | 572 | 555 | 636 | 426 | 469 | 416 | 4.076374 | 3.998429 | 4.239617 | 3.173969 | 3.4003 | 2.885413 |
| ENSMUSG0 | 235.1532 | 298.1597 | 307.5702 | 200.0116 | 176.3821 | 190.0779 | 0.570472 | 0.001534 | 0.020594 | Ace | 11 | 1.06E+08 | 1.06E+08 | + | 5287 | protein_co | angiotensir | - | 236 | 296 | 330 | 192 | 174 | 196 | 2.34767 | 2.976701 | 3.070651 | 1.996831 | 1.760925 | 1.897658 |
| ENSMUSG0 | 2381.424 | 2535.365 | 2419.553 | 2145.958 | 2064.887 | 2190.745 | 0.196471 | 0.001534 | 0.020594 | Enah | 1 | 1.82E+08 | 1.82E+08 | - | 21452 | protein_co | ENAH actin | - | 2390 | 2517 | 2596 | 2060 | 2037 | 2259 | 5.859553 | 6.238328 | 5.953369 | 5.280182 | 5.080706 | 5.390382 |
| ENSMUSG0 | 730.3698 | 756.4796 | 734.4405 | 900.0522 | 846.4314 | 1028.942 | -0.32178 | 0.001538 | 0.020633 | Atf5 | 7 | 44812256 | 44816658 | - | 1993 | protein_co | activating t | TF_bZIP | 733 | 751 | 788 | 864 | 835 | 1061 | 19.34332 | 20.03482 | 19.45113 | 23.83724 | 22.41713 | 27.25079 |
| ENSMUSG0 | 1864.286 | 1768.812 | 1604.025 | 1900.11 | 2239.242 | 2103.464 | -0.25389 | 0.001543 | 0.020662 | Tbc1d15 | 10 | 1.15E+08 | 1.15E+08 | - | 4261 | protein_co | TBC1 doma | - | 1871 | 1756 | 1721 | 1824 | 2209 | 2169 | 23.09386 | 21.91118 | 19.86988 | 23.53763 | 27.73863 | 26.05668 |
| ENSMUSG0 | 987.444 | 799.7933 | 953.4677 | 1028.185 | 1148.511 | 1289.814 | -0.33909 | 0.001544 | 0.020662 | Rest | 5 | 77265491 | 77286432 | + | 7222 | protein_co | RE1-silenci | zf-C2H2 | 991 | 794 | 1023 | 987 | 1133 | 1330 | 7.216897 | 5.845421 | 6.968576 | 7.514656 | 8.394083 | 9.42682 |
| ENSMUSG0 | 841.9679 | 803.8225 | 756.8092 | 907.3442 | 1085.662 | 992.0903 | -0.31364 | 0.001548 | 0.020699 | Atp13a3 | 16 | 30312423 | 30405975 | - | 9938 | protein_co | ATPase type | - | 845 | 798 | 812 | 871 | 1071 | 1023 | 4.4719 | 4.2693 | 4.019601 | 4.81913 | 5.766221 | 5.269237 |
| ENSMUSG0 | 72.73805 | 81.59101 | 92.27107 | 42.71081 | 42.57499 | 32.9727 | 1.062184 | 0.001559 | 0.020827 | Lipe | 7 | 25379527 | 25398710 | - | 5886 | protein_co | lipase, horr | - | 73 | 81 | 99 | 41 | 42 | 34 | 0.652284 | 0.731674 | 0.827448 | 0.383013 | 0.381795 | 0.295685 |
| ENSMUSG0 | 7764.04 | 7395.57 | 7130.969 | 7918.167 | 8653.874 | 8216.02 | -0.15334 | 0.001561 | 0.020829 | Ssr3 | 3 | 65379655 | 65392623 | - | 4172 | protein_co | signal sequ | - | 7792 | 7342 | 7651 | 7601 | 8537 | 8472 | 98.22883 | 93.56704 | 90.21937 | 100.1788 | 109.4868 | 103.9472 |
| ENSMUSG0 | 1159.823 | 1229.909 | 1262.902 | 974.0148 | 1065.388 | 996.9392 | 0.266748 | 0.001567 | 0.020892 | Taldo1 | 7 | 1.41E+08 | 1.41E+08 | + | 2171 | protein_co | transaldola | - | 1164 | 1221 | 1355 | 935 | 1051 | 1028 | 28.1986 | 29.90258 | 30.70474 | 23.68107 | 25.90262 | 24.23842 |
| ENSMUSG0 | 108.6089 | 111.8099 | 118.3679 | 72.92089 | 46.62975 | 61.09647 | 0.908105 | 0.001577 | 0.021008 | Rasip1 | 7 | 45627488 | 45639093 | + | 3202 | protein_co | Ras interac | - | 109 | 111 | 127 | 70 | 46 | 63 | 1.790356 | 1.843124 | 1.951229 | 1.20206 | 0.768665 | 1.007141 |
| ENSMUSG0 | 1571.341 | 1447.485 | 1598.433 | 1735.517 | 1855.053 | 1820.287 | -0.22867 | 0.00158 | 0.02102 | Tacc1 | 8 | 25154552 | 25256588 | - | 11920 | protein_co | transformir | - | 1577 | 1437 | 1715 | 1666 | 1830 | 1877 | 6.958086 | 6.409635 | 7.078052 | 7.685077 | 8.214396 | 8.060446 |
| ENSMUSG0 | 897.7669 | 919.6616 | 932.9631 | 694.8319 | 816.0207 | 701.1547 | 0.314414 | 0.001582 | 0.021024 | Prpf39 | 12 | 65036333 | 65063386 | + | 11967 | protein_co | pre-mRNA p | - | 901 | 913 | 1001 | 667 | 805 | 723 | 3.959805 | 4.056377 | 4.115046 | 3.064714 | 3.599245 | 3.092602 |
| ENSMUSG0 | 179.3541 | 194.4082 | 193.8625 | 311.4764 | 233.1488 | 297.7241 | -0.56932 | 0.001584 | 0.021024 | Cth | 3 | 1.58E+08 | 1.58E+08 | - | 2791 | protein_co | cystathiona | - | 180 | 193 | 208 | 299 | 230 | 307 | 3.391931 | 3.676633 | 3.666312 | 5.890617 | 4.409291 | 5.630534 |
| ENSMUSG0 | 1490.632 | 1514.974 | 1519.211 | 1357.37 | 1281.305 | 1151.135 | 0.256236 | 0.001588 | 0.021036 | Ptn | 6 | 36714929 | 36810220 | - | 3433 | protein_co | pleiotrophi | - | 1496 | 1504 | 1630 | 1303 | 1264 | 1187 | 22.91881 | 23.29308 | 23.35822 | 20.86989 | 19.70036 | 17.69897 |
| ENSMUSG0 | 1417.894 | 1485.762 | 1627.326 | 1753.227 | 1725.301 | 1886.232 | -0.2435 | 0.001588 | 0.021036 | Rnf19b | 4 | 1.29E+08 | 1.29E+08 | + | 2872 | protein_co | ring finger p | - | 1423 | 1475 | 1746 | 1683 | 1702 | 1945 | 26.05882 | 27.30614 | 29.90789 | 32.22175 | 31.70852 | 34.6662 |
| ENSMUSG0 | 1316.26 | 1213.792 | 1361.697 | 1575.091 | 1790.177 | 1402.309 | -0.29236 | 0.001597 | 0.021136 | Foxo1 | 3 | 52268336 | 52353221 | + | 8665 | protein_co | forkhead bo | Fork | 1321 | 1205 | 1461 | 1512 | 1766 | 1446 | 8.018047 | 7.393861 | 8.294831 | 9.594728 | 10.90493 | 8.542221 |
| ENSMUSG0 | 4189.911 | 4132.937 | 4234.217 | 4400.255 | 5104.944 | 4732.552 | -0.18126 | 0.001605 | 0.021205 | Loxl2 | 14 | 69609068 | 69695834 | + | 5405 | protein_co | lysyl oxidas | - | 4205 | 4103 | 4543 | 4224 | 5036 | 4880 | 40.91708 | 40.36069 | 41.34975 | 42.97122 | 49.85295 | 46.2163 |
| ENSMUSG0 | 7888.591 | 7927.422 | 7675.276 | 8816.136 | 8281.85 | 8666.97 | -0.13334 | 0.001605 | 0.021205 | Thbs3 | 3 | 89215180 | 89226837 | + | 4823 | protein_co | thrombosp | - | 7917 | 7870 | 8235 | 8463 | 8170 | 8937 | 86.33318 | 86.75815 | 83.99864 | 96.48428 | 90.63702 | 94.8518 |
| ENSMUSG0 | 1766.638 | 1689.236 | 1560.22 | 1940.737 | 1869.245 | 2111.222 | -0.23984 | 0.00162 | 0.021374 | Dse | 10 | 34151393 | 34207715 | - | 4980 | protein_co | dermatan s | - | 1773 | 1677 | 1674 | 1863 | 1844 | 2177 | 18.72465 | 17.90427 | 16.53682 | 20.56994 | 19.81219 | 22.37691 |
| ENSMUSG0 | 129.5335 | 125.912 | 158.4453 | 238.5555 | 215.916 | 192.0175 | -0.64159 | 0.001641 | 0.021626 | Celsr2 | 3 | 1.08E+08 | 1.08E+08 | - | 11513 | protein_co | cadherin, E | - | 130 | 125 | 170 | 229 | 213 | 198 | 0.593867 | 0.577264 | 0.726418 | 1.093696 | 0.989901 | 0.880335 |
| ENSMUSG0 | 353.7261 | 360.6121 | 312.2304 | 467.7354 | 471.366 | 434.4638 | -0.42052 | 0.001642 | 0.021626 | Gclc | 9 | 77754535 | 77794485 | + | 3766 | protein_co | glutamate- | - | 355 | 358 | 335 | 449 | 465 | 448 | 4.957724 | 5.054236 | 4.376132 | 6.555646 | 6.606531 | 6.08932 |
| ENSMUSG0 | 305.8984 | 393.8529 | 299.182 | 451.0678 | 443.9963 | 476.1645 | -0.45795 | 0.001655 | 0.021772 | Gramd4 | 15 | 86057695 | 86137634 | + | 5567 | protein_co | GRAM dom | - | 307 | 391 | 321 | 433 | 438 | 491 | 2.900357 | 3.734293 | 2.836676 | 4.276772 | 4.209725 | 4.514726 |
| ENSMUSG0 | 130.5299 | 80.58371 | 111.8437 | 171.885 | 192.6012 | 174.5613 | -0.7385 | 0.001667 | 0.021883 | Nos1 | 5 | 1.18E+08 | 1.18E+08 | + | 11151 | protein_co | nitric oxide | - | 131 | 80 | 120 | 165 | 190 | 180 | 0.617862 | 0.381442 | 0.529411 | 0.813616 | 0.911676 | 0.826285 |
| ENSMUSG0 | 270.0276 | 354.5683 | 398.9093 | 244.8059 | 200.7107 | 245.3557 | 0.567581 | 0.001667 | 0.021883 | Exoc7 | 11 | 1.16E+08 | 1.16E+08 | - | 4367 | protein_co | exocyst cor | - | 271 | 352 | 428 | 235 | 198 | 253 | 3.263777 | 4.285606 | 4.821548 | 2.958926 | 2.425955 | 2.965572 |
| ENSMUSG0 | 960.5409 | 978.0848 | 952.5357 | 1118.815 | 1323.88 | 1100.706 | -0.29346 | 0.001669 | 0.021891 | Nfe2l2 | 2 | 75675513 | 75704641 | - | 3796 | protein_co | nuclear fac | TF_bZIP | 964 | 971 | 1022 | 1074 | 1306 | 1135 | 13.35627 | 13.60022 | 13.24496 | 15.55706 | 18.40847 | 15.30526 |
| ENSMUSG0 | 321.841 | 332.4078 | 305.7062 | 396.898 | 484.544 | 421.8566 | -0.44156 | 0.001675 | 0.021946 | Cep104 | 4 | 1.54E+08 | 1.54E+08 | + | 5279 | protein_co | centrosom | - | 323 | 330 | 328 | 381 | 478 | 435 | 3.217994 | 3.323648 | 3.056667 | 3.968467 | 4.844813 | 4.21802 |
| ENSMUSG0 | 102.6304 | 74.53993 | 78.29061 | 120.8403 | 172.3273 | 154.1958 | -0.80926 | 0.001686 | 0.022076 | Slc1a1 | 19 | 28835049 | 28913960 | + | 6170 | protein_co | solute carri | - | 103 | 74 | 84 | 116 | 170 | 159 | 0.877984 | 0.637675 | 0.669761 | 1.033766 | 1.474228 | 1.319116 |
| ENSMUSG0 | 258.0706 | 366.6559 | 307.5702 | 188.5526 | 229.094 | 211.4132 | 0.567116 | 0.001695 | 0.022172 | Plvap | 8 | 71497765 | 71511752 | - | 1987 | protein_co | plasmalem | - | 259 | 364 | 330 | 181 | 226 | 218 | 6.855454 | 9.739941 | 8.170375 | 5.008759 | 6.085712 | 5.616034 |
| ENSMUSG0 | 1630.13 | 1639.878 | 1958.197 | 1505.296 | 1364.427 | 1471.164 | 0.268806 | 0.001699 | 0.022203 | Cope | 8 | 70302518 | 70312993 | + | 3353 | protein_co | coatomer p | - | 1636 | 1628 | 2101 | 1445 | 1346 | 1517 | 25.66162 | 25.81509 | 30.82609 | 23.69648 | 21.47891 | 23.15918 |
| ENSMUSG0 | 495.2166 | 543.94 | 511.685 | 380.2304 | 397.3666 | 409.2494 | 0.385528 | 0.00171 | 0.022315 | Phactr2 | 10 | 13207717 | 13474412 | - | 11692 | protein_co | phosphatas | - | 497 | 540 | 549 | 365 | 392 | 422 | 2.23564 | 2.455601 | 2.309986 | 1.716538 | 1.793899 | 1.847544 |
| ENSMUSG0 | 75.72729 | 49.35752 | 48.46561 | 83.33816 | 141.9166 | 119.2836 | -0.99086 | 0.001715 | 0.02234 | Mamld1 | X | 71050256 | 71156056 | + | 4796 | protein_co | mastermin | - | 76 | 49 | 52 | 80 | 140 | 123 | 0.833429 | 0.543212 | 0.533396 | 0.917192 | 1.561887 | 1.312795 |

| gene_id | H1 | H2 | H3 | M1 | M2 | M3 | log2FoldCh | pvalue | padj | gene_name | gene_chr | gene_start | gene_end | gene_stran | gene_lengtl | gene_bioty | gene_descr | tf_family | H1_readco | H2_readco | H3_readco | M1_readco | M2_readco | M3_readco | H1_fpkm | H2_fpkm | H3_fpkm | M1_fpkm | M2_fpkm | M3_fpkm |
|---|---|---|---|---|---|---|---|---|---|---|---|---|---|---|---|---|---|---|---|---|---|---|---|---|---|---|---|---|---|---|
| ENSMUSG0 | 101.634 | 110.8026 | 93.2031 | 184.3857 | 157.122 | 157.1052 | -0.70643 | 0.001716 | 0.02234 | Aen | 7 | 78895854 | 78911209 | + | 5801 | protein_co | apoptosis e | - | 102 | 110 | 100 | 177 | 155 | 162 | 0.924766 | 1.008191 | 0.848053 | 1.677721 | 1.42965 | 1.429497 |
| ENSMUSG0 | 278.9953 | 305.2108 | 337.3952 | 210.4289 | 170.3 | 242.4463 | 0.564279 | 0.001717 | 0.02234 | AC149090. | JH584304. | 52190 | 59690 | - | 4238 | protein_coding | | - | 280 | 303 | 362 | 202 | 168 | 250 | 3.474813 | 3.80132 | 4.202169 | 2.620836 | 2.121041 | 3.019605 |
| ENSMUSG0 | 351.7333 | 265.9262 | 300.114 | 443.7757 | 428.791 | 395.6724 | -0.46629 | 0.001721 | 0.022369 | Trp53inp1 | 4 | 11156431 | 11174379 | + | 6475 | protein_co | transforma | - | 353 | 264 | 322 | 426 | 423 | 408 | 2.867274 | 2.167789 | 2.446482 | 3.61759 | 3.495436 | 3.225459 |
| ENSMUSG0 | 778.1975 | 605.3851 | 660.81 | 428.1498 | 574.7624 | 520.7747 | 0.423678 | 0.001722 | 0.022369 | Itga11 | 9 | 62677826 | 62783982 | + | 5131 | protein_co | integrin alp | - | 781 | 601 | 709 | 411 | 567 | 537 | 8.005406 | 6.227665 | 6.797827 | 4.404426 | 5.912646 | 5.357268 |
| ENSMUSG0 | 704.4631 | 668.8448 | 680.3827 | 785.4622 | 893.0611 | 869.8973 | -0.31165 | 0.001724 | 0.022373 | Psmd12 | 11 | 1.07E+08 | 1.08E+08 | + | 2649 | protein_co | proteasom | - | 707 | 664 | 730 | 754 | 881 | 897 | 14.03692 | 13.3272 | 13.5571 | 15.65088 | 17.79487 | 17.33331 |
| ENSMUSG0 | 918.6916 | 856.2019 | 819.2553 | 722.9586 | 728.8433 | 603.2064 | 0.336456 | 0.001732 | 0.022454 | Srek1 | 13 | 1.04E+08 | 1.04E+08 | - | 9293 | protein_co | splicing reg | - | 922 | 850 | 879 | 694 | 719 | 622 | 5.218063 | 4.863129 | 4.653276 | 4.106322 | 4.139747 | 3.426143 |
| ENSMUSG0 | 95.65552 | 111.8099 | 136.0765 | 63.54535 | 65.88987 | 56.24754 | 0.889208 | 0.001737 | 0.022494 | Myh14 | 7 | 44605803 | 44670843 | - | 7273 | protein_co | myosin, he | - | 96 | 111 | 146 | 61 | 65 | 58 | 0.694212 | 0.811451 | 0.987564 | 0.461175 | 0.47819 | 0.408212 |
| ENSMUSG0 | 930.6485 | 880.377 | 964.6521 | 612.5355 | 865.6915 | 648.7863 | 0.384167 | 0.00174 | 0.022508 | Ly6e | 15 | 74955051 | 74959905 | + | 3784 | protein_co | lymphocyt | - | 934 | 874 | 1035 | 588 | 854 | 669 | 12.98166 | 12.28042 | 13.45597 | 8.544284 | 12.07557 | 9.049948 |
| ENSMUSG0 | 157.433 | 169.2258 | 168.6976 | 115.6317 | 104.4101 | 75.64325 | 0.746201 | 0.001753 | 0.02266 | Amot | X | 1.45E+08 | 1.46E+08 | - | 8402 | protein_co | angiomotir | - | 158 | 168 | 181 | 111 | 103 | 78 | 0.989028 | 1.063113 | 1.059795 | 0.726423 | 0.655927 | 0.475207 |
| ENSMUSG0 | 222.1998 | 258.8752 | 222.7554 | 345.8534 | 313.2303 | 330.6968 | -0.49227 | 0.001759 | 0.022714 | Sgsm3 | 15 | 80977765 | 81012290 | + | 4477 | protein_co | small G pro | - | 223 | 257 | 239 | 332 | 309 | 341 | 2.619703 | 3.0521 | 2.626254 | 4.077561 | 3.69294 | 3.898867 |
| ENSMUSG0 | 2535.868 | 2577.671 | 2563.085 | 2267.84 | 2190.585 | 2301.3 | 0.183445 | 0.00177 | 0.022825 | Tob2 | 15 | 81848272 | 81858796 | - | 6664 | protein_co | transducer | - | 2545 | 2559 | 2750 | 2177 | 2161 | 2373 | 20.08571 | 20.41682 | 20.30129 | 17.96275 | 17.35084 | 18.22778 |
| ENSMUSG0 | 1338.181 | 1302.434 | 1316.028 | 1059.436 | 1197.168 | 1043.489 | 0.261875 | 0.001781 | 0.022945 | Zfp395 | 14 | 65358389 | 65398930 | + | 4733 | protein_co | zinc finger p | - | 1343 | 1293 | 1412 | 1017 | 1181 | 1076 | 14.92361 | 14.52496 | 14.67655 | 11.81501 | 13.35101 | 11.63716 |
| ENSMUSG0 | 4889.392 | 5132.175 | 5042.288 | 4513.803 | 4585.935 | 4520.169 | 0.145378 | 0.001782 | 0.022945 | Enpp1 | 10 | 24637914 | 24712159 | - | 10257 | protein_co | ectonucleo | - | 4907 | 5095 | 5410 | 4333 | 4524 | 4661 | 25.16112 | 26.4105 | 25.94794 | 23.22832 | 23.59951 | 23.26108 |
| ENSMUSG0 | 604.8219 | 641.6478 | 588.1116 | 818.7975 | 782.5689 | 706.9734 | -0.33127 | 0.001796 | 0.023091 | Fbxo38 | 18 | 62504069 | 62548743 | - | 4388 | protein_co | F-box prote | - | 607 | 637 | 631 | 786 | 772 | 729 | 7.275392 | 7.71837 | 7.074384 | 9.849301 | 9.413508 | 8.504171 |
| ENSMUSG0 | 902.749 | 862.2457 | 934.8271 | 662.5384 | 832.2397 | 566.3546 | 0.389983 | 0.001798 | 0.023091 | Phyh | 2 | 4919019 | 4938730 | + | 1893 | protein_co | phytanoyl- | - | 906 | 856 | 1003 | 636 | 821 | 584 | 25.17166 | 24.0423 | 26.06611 | 18.47379 | 23.20563 | 15.79186 |
| ENSMUSG0 | 1736.746 | 1729.528 | 1652.491 | 1924.07 | 1939.19 | 2056.914 | -0.21022 | 0.001799 | 0.023091 | Tmem248 | 5 | 1.3E+08 | 1.3E+08 | + | 4086 | protein_co | transmemb | - | 1743 | 1717 | 1773 | 1847 | 1913 | 2121 | 22.43538 | 22.34214 | 21.34697 | 24.85524 | 25.05056 | 26.57134 |
| ENSMUSG0 | 201.2752 | 186.3498 | 256.3085 | 155.2173 | 128.7387 | 123.1627 | 0.663311 | 0.001804 | 0.023129 | Pola2 | 19 | 5940543 | 5964206 | - | 2872 | protein_co | polymerase | - | 202 | 185 | 275 | 149 | 127 | 127 | 3.699144 | 3.424838 | 4.710578 | 2.852668 | 2.366029 | 2.263551 |
| ENSMUSG0 | 937.6234 | 907.574 | 1145.466 | 1207.362 | 1264.072 | 1222.899 | -0.30412 | 0.001812 | 0.02321 | Zfp146 | 7 | 30161269 | 30169750 | - | 3270 | protein_co | zinc finger p | zf-C2H2 | 941 | 901 | 1229 | 1159 | 1247 | 1261 | 15.13478 | 14.64974 | 18.48971 | 19.4888 | 20.4042 | 19.7396 |
| ENSMUSG0 | 2611.595 | 2941.305 | 2560.289 | 2078.245 | 2371.022 | 2439.98 | 0.235458 | 0.001819 | 0.023274 | Tmsb4x | X | 1.67E+08 | 1.67E+08 | - | 1371 | protein_co | thymosin, l | - | 2621 | 2920 | 2747 | 1995 | 2339 | 2516 | 100.5458 | 113.2396 | 98.57053 | 80.01196 | 91.28377 | 93.93864 |
| ENSMUSG0 | 98411.6 | 97040.92 | 101503.8 | 93046.02 | 91618.34 | 93212.84 | 0.095815 | 0.001822 | 0.023295 | Fmod | 1 | 1.34E+08 | 1.34E+08 | + | 3306 | protein_co | fibromodul | - | 98766 | 96338 | 108906 | 89319 | 90381 | 96117 | 1571.227 | 1549.343 | 1620.596 | 1485.561 | 1462.767 | 1488.224 |
| ENSMUSG0 | 525.109 | 472.422 | 447.3749 | 328.144 | 402.435 | 340.3946 | 0.431707 | 0.001826 | 0.023315 | Zeb2 | 2 | 44983632 | 45117395 | - | 19942 | protein_co | zinc finger I | zf-C2H2 | 527 | 469 | 480 | 315 | 397 | 351 | 1.389877 | 1.250423 | 1.184127 | 0.868543 | 1.065179 | 0.900968 |
| ENSMUSG0 | 601.8327 | 576.1735 | 603.0241 | 750.0435 | 719.7201 | 747.7044 | -0.31607 | 0.001828 | 0.023315 | Ythdf2 | 4 | 1.32E+08 | 1.32E+08 | - | 4572 | protein_co | YTH domai | - | 604 | 572 | 647 | 720 | 710 | 771 | 6.948084 | 6.651852 | 6.961839 | 8.659159 | 8.30908 | 8.632155 |
| ENSMUSG0 | 80.70935 | 75.54723 | 48.46561 | 37.50217 | 27.36964 | 20.36549 | 1.265623 | 0.001829 | 0.023315 | Rdh5 | 10 | 1.29E+08 | 1.29E+08 | - | 2809 | protein_co | retinol deh | - | 81 | 75 | 52 | 36 | 27 | 21 | 1.516588 | 1.419588 | 0.910705 | 0.704693 | 0.514296 | 0.382683 |
| ENSMUSG0 | 298.9235 | 315.2838 | 284.2695 | 433.3584 | 363.9148 | 428.6451 | -0.44875 | 0.001832 | 0.023339 | Fam110b | 4 | 5644090 | 6108223 | + | 8164 | protein_co | family with | - | 300 | 313 | 305 | 416 | 359 | 442 | 1.932647 | 2.038422 | 1.837904 | 2.801817 | 2.352839 | 2.771343 |
| ENSMUSG0 | 65.76317 | 73.53264 | 106.2515 | 33.33527 | 37.50654 | 43.64033 | 1.101044 | 0.001843 | 0.023447 | Tspan11 | 6 | 1.28E+08 | 1.28E+08 | + | 5326 | protein_co | tetraspanir | - | 66 | 73 | 114 | 32 | 37 | 45 | 0.651744 | 0.728743 | 1.053003 | 0.330368 | 0.371708 | 0.432496 |
| ENSMUSG0 | 1654.043 | 1572.39 | 1684.18 | 1849.065 | 1873.3 | 1974.483 | -0.21425 | 0.00187 | 0.023767 | Ccnd2 | 6 | 1.27E+08 | 1.27E+08 | - | 9409 | protein_co | cyclin D2 [S | - | 1660 | 1561 | 1807 | 1775 | 1848 | 2036 | 9.278953 | 8.820886 | 9.448015 | 10.373 | 10.50895 | 11.07657 |
| ENSMUSG0 | 241.1316 | 219.5906 | 195.7265 | 171.885 | 123.6702 | 119.2836 | 0.662958 | 0.001878 | 0.023847 | Pdlim1 | 19 | 40221173 | 40271842 | - | 5260 | protein_co | PDZ and LIN | - | 242 | 218 | 210 | 165 | 122 | 123 | 2.419714 | 2.203553 | 1.964081 | 1.724835 | 1.241009 | 1.19699 |
| ENSMUSG0 | 1653.047 | 1590.521 | 1682.316 | 1914.694 | 1890.532 | 1879.444 | -0.20647 | 0.001883 | 0.023896 | Sfrp1 | 8 | 23411502 | 23449632 | + | 4372 | protein_co | secreted fri | - | 1659 | 1579 | 1805 | 1838 | 1865 | 1938 | 19.95724 | 19.20237 | 20.31061 | 23.11611 | 22.82441 | 22.69053 |
| ENSMUSG0 | 997.4081 | 972.041 | 1062.515 | 1191.736 | 1167.771 | 1282.056 | -0.2642 | 0.001893 | 0.023991 | Eif4ebp2 | 10 | 61428491 | 61452661 | - | 5784 | protein_co | eukaryotic | - | 1001 | 965 | 1140 | 1144 | 1152 | 1322 | 9.102069 | 8.870576 | 9.69622 | 10.87545 | 10.65676 | 11.69969 |
| ENSMUSG0 | 1729.771 | 1570.375 | 1549.968 | 1902.194 | 1803.355 | 1977.392 | -0.22892 | 0.001895 | 0.024001 | Pcyt1a | 16 | 32430921 | 32475070 | + | 7412 | protein_co | phosphate | - | 1736 | 1559 | 1663 | 1826 | 1779 | 2039 | 12.31824 | 11.18313 | 11.03781 | 13.54612 | 12.84226 | 14.08163 |
| ENSMUSG0 | 1281.385 | 1069.749 | 1149.194 | 1002.141 | 942.732 | 921.2959 | 0.288595 | 0.0019 | 0.02404 | Vill | 9 | 1.19E+08 | 1.19E+08 | + | 4409 | protein_co | villin-like [S | - | 1286 | 1062 | 1233 | 962 | 930 | 950 | 15.34035 | 12.8067 | 13.7578 | 11.99733 | 11.28609 | 11.02947 |
| ENSMUSG0 | 1517.535 | 1639.878 | 1640.375 | 1394.873 | 1252.921 | 1405.219 | 0.243402 | 0.001902 | 0.024043 | Ucp2 | 7 | 1E+08 | 1.01E+08 | + | 6989 | protein_co | uncoupling | - | 1523 | 1628 | 1760 | 1339 | 1236 | 1449 | 11.46091 | 12.38489 | 12.38864 | 10.53453 | 9.462464 | 10.61266 |
| ENSMUSG0 | 80.70935 | 100.7296 | 129.5523 | 56.25326 | 59.80773 | 41.70076 | 0.98158 | 0.001917 | 0.024176 | Atl1 | 12 | 69892614 | 69966417 | + | 9175 | protein_co | atlastin GTI | - | 81 | 100 | 139 | 54 | 59 | 43 | 0.464316 | 0.579491 | 0.745306 | 0.323621 | 0.34407 | 0.239902 |
| ENSMUSG0 | 370.6651 | 345.5027 | 383.9968 | 476.0693 | 452.1059 | 531.4423 | -0.40787 | 0.001918 | 0.024176 | Zkscan1 | 5 | 1.38E+08 | 1.38E+08 | + | 8541 | protein_co | zinc finger v | zf-C2H2 | 372 | 343 | 412 | 457 | 446 | 548 | 2.290702 | 2.135198 | 2.373091 | 2.942097 | 2.794004 | 3.284301 |
| ENSMUSG0 | 928.6557 | 823.9684 | 908.7303 | 686.4981 | 741.0076 | 728.3087 | 0.303941 | 0.00192 | 0.024176 | Cat | 2 | 1.03E+08 | 1.03E+08 | - | 2613 | protein_co | catalase [Sc | - | 932 | 818 | 975 | 659 | 731 | 751 | 18.75905 | 16.64434 | 18.35655 | 13.86741 | 14.96852 | 14.712 |
| ENSMUSG0 | 293.9414 | 275.9992 | 253.5124 | 338.5613 | 395.3392 | 403.4306 | -0.46688 | 0.001923 | 0.024176 | Mrgprf | 7 | 1.45E+08 | 1.45E+08 | + | 2214 | protein_co | MAS-relate | - | 295 | 274 | 272 | 325 | 390 | 416 | 7.007752 | 6.579998 | 6.043899 | 8.071518 | 9.42514 | 9.618045 |
| ENSMUSG0 | 79.71293 | 68.49615 | 66.1742 | 123.9655 | 112.5196 | 140.6189 | -0.81578 | 0.001924 | 0.024176 | Cd33 | 7 | 43524216 | 43544428 | - | 7771 | protein_co | CD33 antig | - | 80 | 68 | 71 | 119 | 111 | 145 | 0.541436 | 0.465248 | 0.449477 | 0.842014 | 0.76427 | 0.955129 |
| ENSMUSG0 | 538.0623 | 506.6701 | 585.3155 | 736.501 | 734.9255 | 623.5719 | -0.36108 | 0.001924 | 0.024176 | Rnf144b | 13 | 47122656 | 47247991 | + | 4936 | protein_co | ring finger p | - | 540 | 503 | 628 | 707 | 725 | 643 | 5.753776 | 5.418083 | 6.259078 | 7.875783 | 7.858934 | 6.668174 |
| ENSMUSG0 | 76.7237 | 88.64208 | 102.5234 | 59.37844 | 18.24643 | 37.82162 | 1.216583 | 0.001955 | 0.024539 | Dpm1 | 2 | 1.68E+08 | 1.68E+08 | - | 7489 | protein_co | dolichol-pl | - | 77 | 88 | 110 | 57 | 18 | 39 | 0.540756 | 0.624758 | 0.722595 | 0.418505 | 0.128603 | 0.26657 |
| ENSMUSG0 | 1089.078 | 955.9243 | 976.7685 | 1212.57 | 1170.812 | 1256.842 | -0.26887 | 0.001964 | 0.024631 | Cd82 | 2 | 93419111 | 93463140 | - | 3005 | protein_co | CD82 antig | - | 1093 | 949 | 1048 | 1164 | 1155 | 1296 | 19.12978 | 16.79092 | 17.15705 | 21.29894 | 20.56545 | 22.07657 |
| ENSMUSG0 | 901.7526 | 938.8002 | 867.7209 | 1141.733 | 1015.718 | 1125.921 | -0.27808 | 0.00197 | 0.02468 | Mras | 9 | 99385420 | 99437381 | - | 5134 | protein_co | muscle and | - | 905 | 932 | 931 | 1096 | 1002 | 1161 | 9.27101 | 9.651901 | 8.921127 | 11.73827 | 10.4427 | 11.5757 |
| ENSMUSG0 | 388.6006 | 370.6851 | 293.5898 | 272.9325 | 249.3678 | 203.6549 | 0.536591 | 0.001976 | 0.024737 | Hes1 | 16 | 30064384 | 30067796 | + | 2037 | protein_co | hes family b | bHLH | 390 | 368 | 315 | 262 | 246 | 210 | 10.0695 | 9.60527 | 7.607561 | 7.072285 | 6.461672 | 5.277149 |
| ENSMUSG0 | 1658.029 | 1750.681 | 1707.481 | 2197.002 | 1799.3 | 2065.642 | -0.24465 | 0.001979 | 0.024748 | Arf6 | 12 | 69372150 | 69373783 | + | 1534 | protein_co | ADP-ribosy | - | 1664 | 1738 | 1832 | 2109 | 1775 | 2130 | 57.05087 | 60.23892 | 58.75245 | 75.59632 | 61.91185 | 71.07638 |
| ENSMUSG0 | 9475.875 | 9758.687 | 9536.542 | 10639.16 | 10011.21 | 10866.44 | -0.13156 | 0.001994 | 0.024912 | Ldha | 7 | 46841475 | 46855627 | + | 3310 | protein_co | lactate deh | - | 9510 | 9688 | 10232 | 10213 | 9876 | 11205 | 151.1078 | 155.6177 | 152.0752 | 169.6582 | 159.6445 | 173.2826 |
| ENSMUSG0 | 128.5371 | 123.8975 | 105.3195 | 69.79571 | 76.02677 | 49.45905 | 0.874341 | 0.002 | 0.024966 | Dync2li1 | 17 | 84626496 | 84655558 | + | 1324 | protein_co | dynein cytc | - | 129 | 123 | 113 | 67 | 75 | 51 | 5.124318 | 4.939351 | 4.198715 | 2.782507 | 3.030917 | 1.971756 |
| ENSMUSG0 | 215.2249 | 202.4666 | 224.6195 | 146.8835 | 136.8482 | 142.5584 | 0.591868 | 0.002014 | 0.025113 | Shank3 | 15 | 89499623 | 89560261 | + | 9189 | protein_co | SH3 and mu | - | 216 | 201 | 241 | 141 | 135 | 147 | 1.236289 | 1.163002 | 1.290252 | 0.843724 | 0.786079 | 0.81888 |
| ENSMUSG0 | 1564.366 | 1652.973 | 1813.732 | 1967.822 | 1903.71 | 2022.002 | -0.22795 | 0.002016 | 0.025118 | Chd2 | 7 | 73426638 | 73541830 | - | 14300 | protein_co | chromodoı | - | 1570 | 1641 | 1946 | 1889 | 1878 | 2085 | 5.774282 | 6.101342 | 6.694724 | 7.263491 | 7.026845 | 7.463475 |
| ENSMUSG0 | 203.268 | 173.255 | 152.8531 | 218.7627 | 304.1071 | 280.2679 | -0.60265 | 0.002057 | 0.025609 | Twnk | 19 | 45006558 | 45012762 | + | 3488 | protein_co | twinkle mt | - | 204 | 172 | 164 | 210 | 300 | 289 | 3.076012 | 2.621831 | 2.313094 | 3.31049 | 4.601989 | 4.241236 |
| ENSMUSG0 | 880.8279 | 922.6835 | 868.6529 | 711.4996 | 616.3237 | 793.2843 | 0.332648 | 0.002091 | 0.026006 | Uvrag | 7 | 98885021 | 99141141 | - | 9035 | protein_co | UV radiatio | - | 884 | 916 | 932 | 683 | 608 | 818 | 5.145866 | 5.390389 | 5.074738 | 4.156636 | 3.600611 | 4.634429 |
| ENSMUSG0 | 864.8853 | 832.0268 | 795.0225 | 584.4089 | 713.638 | 684.6684 | 0.329174 | 0.002101 | 0.0261 | Cfh | 1 | 1.4E+08 | 1.4E+08 | - | 8383 | protein_co | complemer | - | 868 | 826 | 853 | 561 | 704 | 706 | 5.445711 | 5.238819 | 5.005823 | 3.679704 | 4.493389 | 4.310983 |
| ENSMUSG0 | 47.82776 | 51.37212 | 48.46561 | 14.58418 | 24.32857 | 17.45613 | 1.388776 | 0.002123 | 0.026346 | Pycard | 7 | 1.28E+08 | 1.28E+08 | - | 3800 | protein_co | PYD and CA | - | 48 | 51 | 52 | 14 | 24 | 18 | 0.664342 | 0.713575 | 0.673202 | 0.202579 | 0.337931 | 0.242471 |
| ENSMUSG0 | 1671.979 | 1584.477 | 1738.238 | 1860.524 | 1918.916 | 2027.821 | -0.21742 | 0.002125 | 0.026349 | Pim3 | 15 | 88862186 | 88865726 | + | 2427 | protein_co | proviral int | - | 1678 | 1573 | 1865 | 1786 | 1893 | 2091 | 36.36273 | 34.45972 | 37.80376 | 40.46329 | 41.7332 | 44.1017 |
| ENSMUSG0 | 43847.1 | 44441.92 | 45921.17 | 48605.94 | 48142.18 | 47114.1 | -0.10015 | 0.00213 | 0.026386 | Col6a1 | 10 | 76708792 | 76726168 | - | 4380 | protein_co | collagen, ty | - | 44005 | 44120 | 49270 | 46659 | 47492 | 48582 | 528.3993 | 535.5675 | 553.3939 | 585.748 | 580.1592 | 567.7699 |
| ENSMUSG0 | 1994.816 | 1892.71 | 1864.994 | 2104.289 | 2317.296 | 2210.14 | -0.20541 | 0.002134 | 0.026412 | Sec24a | 11 | 51692264 | 51763634 | - | 10683 | protein_co | Sec24 relat | - | 2002 | 1879 | 2001 | 2020 | 2286 | 2279 | 9.856102 | 9.351609 | 9.214669 | 10.39699 | 11.44943 | 10.91999 |
| ENSMUSG0 | 2628.534 | 2513.204 | 2599.435 | 2140.749 | 2225.05 | 2392.46 | 0.195652 | 0.002145 | 0.026525 | Alas2 | X | 1.51E+08 | 1.51E+08 | + | 2403 | protein_co | aminolevul | - | 2638 | 2495 | 2789 | 2055 | 2195 | 2467 | 57.73715 | 55.20388 | 57.09797 | 47.0227 | 48.87441 | 52.55166 |
| ENSMUSG0 | 922.6772 | 947.8659 | 889.1576 | 1053.186 | 1100.868 | 1166.652 | -0.26751 | 0.002155 | 0.026618 | Rragc | 4 | 1.24E+08 | 1.24E+08 | + | 3341 | protein_co | Ras-related | - | 926 | 941 | 954 | 1011 | 1086 | 1203 | 14.57702 | 14.97497 | 14.04746 | 16.63888 | 17.39219 | 18.43148 |
| ENSMUSG0 | 2141.289 | 2062.943 | 2172.564 | 2272.007 | 2545.376 | 2526.29 | -0.20376 | 0.002158 | 0.026628 | Pttg1ip | 10 | 77581720 | 77598732 | + | 3877 | protein_co | pituitary tu | - | 2149 | 2048 | 2331 | 2181 | 2511 | 2605 | 29.15244 | 28.08581 | 29.57824 | 30.93209 | 34.65387 | 34.39403 |
| ENSMUSG0 | 35.87082 | 51.37212 | 43.80546 | 20.83454 | 13.17797 | 11.63742 | 1.524559 | 0.002193 | 0.02704 | Ptch2 | 4 | 1.17E+08 | 1.17E+08 | + | 5974 | protein_co | patched 2 [ | - | 36 | 51 | 47 | 20 | 13 | 12 | 0.316936 | 0.453897 | 0.387042 | 0.184083 | 0.116434 | 0.102822 |
| ENSMUSG0 | 155.4402 | 208.5103 | 189.2023 | 267.7238 | 280.7927 | 261.842 | -0.55076 | 0.00221 | 0.027218 | Mief1 | 15 | 80234018 | 80253371 | + | 8106 | protein_co | mitochond | - | 156 | 207 | 203 | 257 | 277 | 270 | 1.012167 | 1.35774 | 1.232013 | 1.743315 | 1.828412 | 1.705015 |
| ENSMUSG0 | 914.7059 | 984.1286 | 1042.943 | 908.386 | 712.6243 | 696.3058 | 0.345072 | 0.002224 | 0.027355 | Ampd2 | 3 | 1.08E+08 | 1.08E+08 | - | 5224 | protein_co | adenosine | - | 918 | 977 | 1119 | 872 | 703 | 718 | 9.242168 | 9.943613 | 10.53787 | 9.178311 | 7.200339 | 7.035458 |
| ENSMUSG0 | 12804.89 | 13080.75 | 12993.44 | 12304.88 | 11843.96 | 11796.47 | 0.113251 | 0.002225 | 0.027355 | App | 16 | 84949685 | 85173766 | - | 12996 | protein_co | amyloid be | - | 12851 | 12986 | 13941 | 11812 | 11684 | 12164 | 52.00697 | 53.12739 | 52.7728 | 49.9762 | 48.10416 | 47.91128 |
| ENSMUSG0 | 933.6377 | 1076.8 | 1058.787 | 1269.865 | 1273.195 | 1163.742 | -0.27193 | 0.002228 | 0.027368 | Bach1 | 16 | 87698945 | 87733346 | + | 6179 | protein_co | BTB and CN | TF_bZIP | 937 | 1069 | 1136 | 1219 | 1256 | 1200 | 7.97546 | 9.1984 | 9.044531 | 10.84763 | 10.87608 | 9.941093 |
| ENSMUSG0 | 176.3649 | 188.3644 | 164.9695 | 228.1382 | 275.7238 | 271.5399 | -0.55078 | 0.00224 | 0.027495 | 1520401A( | 17 | 23704488 | 23722783 | - | 4111 | protein_co | RIKEN cDNA | - | 177 | 187 | 177 | 219 | 272 | 280 | 2.264436 | 2.418505 | 2.118126 | 2.92918 | 3.540155 | 3.486435 |
| ENSMUSG0 | 122.5586 | 98.71504 | 137.0086 | 43.75254 | 79.06784 | 66.91518 | 0.916628 | 0.00225 | 0.027591 | Spock2 | 10 | 60106219 | 60135198 | + | 5533 | protein_co | sparc/oste | - | 123 | 98 | 147 | 42 | 78 | 69 | 1.169173 | 0.941712 | 1.307021 | 0.417386 | 0.754284 | 0.638351 |
| ENSMUSG0 | 1234.554 | 1229.909 | 1218.165 | 1387.58 | 1441.468 | 1486.681 | -0.22912 | 0.002272 | 0.027828 | Cpox | 16 | 58670292 | 58717636 | + | 6661 | protein_co | coproporpl | - | 1239 | 1221 | 1307 | 1332 | 1422 | 1533 | 9.782868 | 9.746059 | 9.652994 | 10.99548 | 11.42249 | 11.78077 |
| ENSMUSG0 | 332.8015 | 350.5391 | 410.0937 | 529.1973 | 428.791 | 518.8351 | -0.43289 | 0.002274 | 0.027828 | Klf16 | 10 | 80567124 | 80577321 | - | 2674 | protein_co | Kruppel-lik | zf-C2H2 | 334 | 348 | 440 | 508 | 423 | 535 | 6.569305 | 6.919435 | 8.095006 | 10.44604 | 8.46408 | 10.2415 |
| ENSMUSG0 | 622.7573 | 561.0641 | 521.9374 | 738.5845 | 728.8433 | 686.6079 | -0.33691 | 0.002299 | 0.02811 | Asap2 | 12 | 20990459 | 21270171 | + | 6962 | protein_co | ArfGAP wit | - | 625 | 557 | 560 | 709 | 719 | 708 | 4.721504 | 4.25377 | 3.957126 | 5.59966 | 5.525807 | 5.205594 |
| ENSMUSG0 | 567.9547 | 498.6117 | 524.7335 | 755.2521 | 588.9541 | 717.6411 | -0.37368 | 0.002347 | 0.028671 | Bet1l | 7 | 1.41E+08 | 1.41E+08 | - | 3953 | protein_co | Bet1 golgi v | - | 570 | 495 | 563 | 725 | 581 | 740 | 7.583722 | 6.657807 | 7.006603 | 10.08465 | 7.86412 | 9.582437 |
| ENSMUSG0 | 631.725 | 625.531 | 603.9561 | 742.7514 | 790.6784 | 764.1907 | -0.30415 | 0.002351 | 0.028698 | Lpgat1 | 1 | 1.92E+08 | 1.92E+08 | + | 7610 | protein_co | lysophosph | - | 634 | 621 | 648 | 713 | 780 | 788 | 4.381663 | 4.338701 | 4.189057 | 5.151745 | 5.484168 | 5.300449 |
| ENSMUSG0 | 174.372 | 172.2477 | 172.4257 | 86.46334 | 120.6291 | 115.4044 | 0.685553 | 0.00237 | 0.028894 | Acads | 5 | 1.15E+08 | 1.15E+08 | - | 2269 | protein_co | acyl-Coenz | - | 175 | 171 | 185 | 83 | 119 | 119 | 4.056373 | 4.006955 | 4.011097 | 2.011375 | 2.806165 | 2.684624 |
| ENSMUSG0 | 1712.832 | 1731.542 | 1480.997 | 1819.897 | 2288.913 | 1870.716 | -0.28012 | 0.002375 | 0.028937 | Pank3 | 11 | 35769484 | 35791285 | + | 7376 | protein_co | pantothena | - | 1719 | 1719 | 1589 | 1747 | 2258 | 1929 | 12.25714 | 12.39104 | 10.59812 | 13.02331 | 16.37962 | 13.38697 |
| ENSMUSG0 | 3850.135 | 3717.931 | 3987.229 | 4181.492 | 4313.252 | 4378.58 | -0.15575 | 0.002391 | 0.0291 | Serbp1 | 6 | 67238176 | 67297736 | + | 20117 | protein_co | serpine1 m | - | 3864 | 3691 | 4278 | 4014 | 4255 | 4515 | 10.10202 | 9.755143 | 10.46173 | 10.97144 | 11.31715 | 11.48856 |
| ENSMUSG0 | 806.0971 | 758.4942 | 830.4396 | 1022.976 | 976.1837 | 918.3866 | -0.28411 | 0.002393 | 0.0291 | Zzef1 | 11 | 72796226 | 72927120 | + | 16450 | protein_co | zinc finger, | - | 809 | 753 | 891 | 982 | 963 | 947 | 2.586527 | 2.433783 | 2.664635 | 3.282427 | 3.132284 | 2.946831 |
| ENSMUSG0 | 352.7297 | 292.1159 | 387.7249 | 469.8189 | 479.4755 | 444.1616 | -0.43134 | 0.002398 | 0.029131 | Zfp507 | 7 | 35772343 | 35803003 | - | 8231 | protein_co | zinc finger p | zf-C2H2 | 354 | 290 | 416 | 451 | 473 | 458 | 2.26196 | 1.873261 | 2.486375 | 3.012822 | 3.074747 | 2.848288 |
| ENSMUSG0 | 297.9271 | 267.9408 | 270.289 | 197.9281 | 215.916 | 159.0448 | 0.546274 | 0.002409 | 0.029242 | Zfp185 | X | 72987339 | 73031543 | + | 3707 | protein_co | zinc finger p | - | 299 | 266 | 290 | 190 | 213 | 164 | 4.24212 | 3.815152 | 3.848587 | 2.818256 | 3.074382 | 2.264604 |
| ENSMUSG0 | 672.5779 | 663.8083 | 634.7131 | 553.1571 | 501.7767 | 511.0768 | 0.331974 | 0.002422 | 0.029373 | Dars | 1 | 1.28E+08 | 1.28E+08 | - | 4358 | protein_co | aspartyl-tR | - | 675 | 659 | 681 | 531 | 495 | 527 | 8.146122 | 8.039906 | 7.687512 | 6.699722 | 6.077413 | 6.190054 |
| ENSMUSG0 | 527.1018 | 559.0495 | 583.4514 | 675.0391 | 731.8844 | 685.6381 | -0.32546 | 0.002425 | 0.029381 | Gtf2f1 | 17 | 57003405 | 57011288 | - | 1717 | protein_co | general tra | - | 529 | 555 | 626 | 648 | 722 | 707 | 16.20391 | 17.18603 | 17.93618 | 20.75172 | 22.49923 | 21.07755 |
| ENSMUSG0 | 1384.016 | 1343.733 | 1419.483 | 1517.796 | 1671.575 | 1665.121 | -0.22722 | 0.002443 | 0.029557 | Kat6a | 8 | 22859535 | 22943259 | + | 9180 | protein_co | K(lysine) ac | - | 1389 | 1334 | 1523 | 1457 | 1649 | 1717 | 7.957816 | 7.7262 | 8.161747 | 8.727027 | 9.611226 | 9.574116 |
| ENSMUSG0 | 49.82058 | 49.35752 | 39.1453 | 25.00145 | 14.19166 | 5.818711 | 1.625456 | 0.002444 | 0.029557 | Gm17971 | 1 | 55066264 | 55067069 | + | 806 | processed_ | predicted g | - | 50 | 49 | 42 | 24 | 14 | 6 | 3.262641 | 3.232316 | 2.56354 | 1.63729 | 0.929381 | 0.381055 |
| ENSMUSG0 | 89.67705 | 106.7734 | 115.5718 | 41.66908 | 64.87618 | 58.18711 | 0.920889 | 0.002451 | 0.029608 | Hhex | 19 | 37434810 | 37440731 | + | 1918 | protein_co | hematopoi | Homeobox | 90 | 106 | 124 | 40 | 64 | 60 | 2.467904 | 2.938394 | 3.180526 | 1.146729 | 1.785386 | 1.601304 |
| ENSMUSG0 | 1506.574 | 1385.033 | 1341.193 | 1546.965 | 1769.903 | 1689.366 | -0.24249 | 0.00246 | 0.029692 | Nr3c1 | 18 | 39410545 | 39491301 | - | 7425 | protein_co | nuclear rec | ESR-like | 1512 | 1375 | 1439 | 1485 | 1746 | 1742 | 10.71 | 9.845982 | 9.534331 | 10.99713 | 12.58197 | 12.00944 |
| ENSMUSG0 | 676.5635 | 827.9976 | 677.5866 | 870.8838 | 983.2796 | 884.4441 | -0.32814 | 0.002495 | 0.030084 | Stxbp1 | 2 | 32787602 | 32847245 | - | 4997 | protein_co | syntaxin bi | - | 679 | 822 | 727 | 836 | 970 | 912 | 7.146522 | 8.746117 | 7.157329 | 9.199122 | 10.38636 | 9.342359 |
| ENSMUSG0 | 1063.171 | 1067.734 | 1065.311 | 1322.993 | 1288.4 | 1185.078 | -0.24797 | 0.002511 | 0.030229 | Rapgef1 | 2 | 29619720 | 29740978 | + | 7385 | protein_co | Rap guanin | - | 1067 | 1060 | 1143 | 1270 | 1271 | 1222 | 7.598857 | 7.631469 | 7.614154 | 9.455896 | 9.208648 | 8.470163 |
| ENSMUSG0 | 1528.496 | 1481.733 | 1413.891 | 1783.437 | 1778.013 | 1612.753 | -0.2259 | 0.002511 | 0.030229 | Samd4 | 14 | 46882854 | 47105815 | + | 8658 | protein_co | sterile alph | - | 1534 | 1471 | 1517 | 1712 | 1754 | 1663 | 9.318416 | 9.03333 | 8.619735 | 10.87266 | 10.83959 | 9.832088 |

| gene_id | H1 | H2 | H3 | M1 | M2 | M3 | log2FoldCh | pvalue | padj | gene_name | gene_chr | gene_start | gene_end | gene_stran | gene_lengt | gene_bioty | gene_descr | tf_family | H1_readco | H2_readco | H3_readco | M1_readco | M2_readco | M3_readco | H1_fpkm | H2_fpkm | H3_fpkm | M1_fpkm | M2_fpkm | M3_fpkm |
|---|---|---|---|---|---|---|---|---|---|---|---|---|---|---|---|---|---|---|---|---|---|---|---|---|---|---|---|---|---|---|
| ENSMUSG0 | 1148.863 | 1031.471 | 1118.437 | 1296.95 | 1307.66 | 1301.452 | -0.24363 | 0.002528 | 0.030403 | Siah2 | 3 | 58674938 | 58692400 | - | 2999 | protein_co | siah E3 ubi | - | 1153 | 1024 | 1200 | 1245 | 1290 | 1342 | 20.22028 | 18.15417 | 19.68479 | 22.82666 | 23.01516 | 22.90589 |
| ENSMUSG0 | 803.1078 | 780.6547 | 685.9748 | 602.1182 | 634.5701 | 570.2337 | 0.32872 | 0.00254 | 0.030523 | Ints6 | 14 | 62676330 | 62761169 | - | 16805 | protein_co | integrator | - | 806 | 775 | 736 | 578 | 626 | 588 | 2.522498 | 2.451975 | 2.154593 | 1.891206 | 1.993135 | 1.791059 |
| ENSMUSG0 | 757.2729 | 829.0049 | 742.8287 | 962.5558 | 886.979 | 1000.818 | -0.29183 | 0.002565 | 0.030792 | Brix1 | 15 | 10474779 | 10485947 | - | 2731 | protein_co | BRX1, biog | - | 760 | 823 | 797 | 924 | 875 | 1032 | 14.63613 | 16.02252 | 14.35696 | 18.60372 | 17.14301 | 19.34323 |
| ENSMUSG0 | 404.5431 | 376.7288 | 375.6085 | 280.2246 | 288.9017 | 293.8449 | 0.422536 | 0.00263 | 0.031539 | Galnt5 | 2 | 57997884 | 58045860 | + | 4462 | protein_co | polypeptid | - | 406 | 374 | 403 | 269 | 285 | 303 | 4.785538 | 4.456509 | 4.443256 | 3.314913 | 3.41756 | 3.476035 |
| ENSMUSG0 | 58.78829 | 53.38671 | 102.5234 | 32.29354 | 32.43809 | 30.06334 | 1.181793 | 0.002636 | 0.03158 | Nmi | 2 | 51948487 | 51973494 | - | 2001 | protein_co | N-myc (and | - | 59 | 53 | 110 | 31 | 32 | 31 | 1.550741 | 1.408256 | 2.704404 | 0.851852 | 0.855665 | 0.793023 |
| ENSMUSG0 | 581.9044 | 549.9838 | 539.646 | 652.1211 | 761.2814 | 689.5173 | -0.3315 | 0.002652 | 0.031742 | Dennd3 | 15 | 73512560 | 73572242 | + | 5709 | protein_co | DENN/MAD | - | 584 | 546 | 579 | 626 | 751 | 711 | 5.38006 | 5.084935 | 4.989355 | 6.029256 | 7.03851 | 6.375007 |
| ENSMUSG0 | 5443.397 | 5638.845 | 5498.051 | 4855.49 | 5090.753 | 5107.859 | 0.139177 | 0.002657 | 0.031782 | Tubb5 | 17 | 35833921 | 35838306 | - | 3327 | protein_co | tubulin, be | - | 5463 | 5598 | 5899 | 4661 | 5022 | 5267 | 86.36002 | 89.46082 | 87.22711 | 77.03281 | 80.76528 | 81.03667 |
| ENSMUSG0 | 1219.608 | 1286.317 | 1292.727 | 1013.6 | 1105.936 | 1085.19 | 0.245186 | 0.002673 | 0.031944 | Fhl1 | X | 56731787 | 56793346 | + | 3714 | protein_co | four and a h | - | 1224 | 1277 | 1387 | 973 | 1091 | 1119 | 17.333 | 18.28108 | 18.37217 | 14.40524 | 15.71751 | 15.42266 |
| ENSMUSG0 | 1083.099 | 1110.041 | 1065.311 | 1257.365 | 1251.908 | 1334.424 | -0.23865 | 0.002684 | 0.032046 | Psmd7 | 8 | 1.08E+08 | 1.08E+08 | - | 2261 | protein_co | proteasom | - | 1087 | 1102 | 1143 | 1207 | 1235 | 1376 | 25.28502 | 25.91396 | 24.86976 | 29.35325 | 29.22585 | 31.15222 |
| ENSMUSG0 | 604.8219 | 641.6478 | 676.6545 | 695.8737 | 913.3349 | 840.8038 | -0.3494 | 0.002694 | 0.03213 | Gpt2 | 8 | 85492576 | 85527560 | + | 5643 | protein_co | glutamic py | - | 607 | 637 | 726 | 668 | 901 | 867 | 5.657349 | 6.001809 | 6.329253 | 6.509024 | 8.543101 | 7.864663 |
| ENSMUSG0 | 101.634 | 128.9339 | 121.164 | 75.00435 | 71.97201 | 36.85184 | 0.938701 | 0.002696 | 0.03213 | Haghl | 17 | 25779843 | 25785673 | - | 2643 | protein_co | hydroxyacy | - | 102 | 128 | 130 | 72 | 71 | 38 | 2.029726 | 2.574931 | 2.419759 | 1.497907 | 1.437348 | 0.735966 |
| ENSMUSG0 | 2425.266 | 2575.657 | 2717.802 | 3127.265 | 3252.932 | 2657.211 | -0.22714 | 0.002718 | 0.03236 | Mbtps1 | 8 | 1.2E+08 | 1.2E+08 | - | 10539 | protein_co | membrane | - | 2434 | 2557 | 2916 | 3002 | 3209 | 2740 | 12.14662 | 12.89983 | 13.61175 | 15.66249 | 16.29188 | 13.30829 |
| ENSMUSG0 | 118.573 | 132.9631 | 143.5328 | 208.3454 | 242.272 | 167.7728 | -0.64519 | 0.002729 | 0.03246 | Mfap1a | 2 | 1.21E+08 | 1.22E+08 | - | 4198 | protein_co | microfibril | - | 119 | 132 | 154 | 200 | 239 | 173 | 1.490867 | 1.6718 | 1.804697 | 2.619612 | 3.046185 | 2.109477 |
| ENSMUSG0 | 1405.937 | 1408.2 | 1581.657 | 1593.842 | 1826.67 | 1780.526 | -0.2425 | 0.002747 | 0.032645 | Slc41a1 | 1 | 1.32E+08 | 1.32E+08 | + | 4912 | protein_co | solute carri | - | 1411 | 1398 | 1697 | 1530 | 1802 | 1836 | 15.10786 | 15.13218 | 16.9961 | 17.12705 | 19.62896 | 19.1331 |
| ENSMUSG0 | 36.86723 | 29.21159 | 31.68906 | 91.67198 | 60.82142 | 60.12668 | -1.11985 | 0.002766 | 0.032838 | Rasl10b | 11 | 83409137 | 83421039 | + | 4010 | protein_co | RAS-like, fa | - | 37 | 29 | 34 | 88 | 60 | 62 | 0.485279 | 0.384509 | 0.417119 | 1.206668 | 0.800585 | 0.791441 |
| ENSMUSG0 | 513.152 | 630.5675 | 546.1702 | 694.8319 | 762.2951 | 687.5777 | -0.34407 | 0.002784 | 0.033029 | Mecp2 | X | 74026592 | 74135363 | - | 15067 | protein_co | methyl CpG | MBD | 515 | 626 | 586 | 667 | 752 | 709 | 1.79769 | 2.209024 | 1.91336 | 2.434157 | 2.670496 | 2.408743 |
| ENSMUSG0 | 745.3159 | 759.5015 | 810.867 | 1081.313 | 945.773 | 854.3808 | -0.31457 | 0.002793 | 0.033103 | Foxo4 | X | 1.01E+08 | 1.01E+08 | + | 3485 | protein_co | forkhead bo | Fork | 748 | 754 | 870 | 1038 | 933 | 881 | 11.28842 | 11.50327 | 12.28124 | 16.37737 | 14.32451 | 12.9403 |
| ENSMUSG0 | 553.0085 | 559.0495 | 541.51 | 655.2463 | 680.1862 | 726.3691 | -0.31879 | 0.002805 | 0.033187 | Atf1 | 15 | 1E+08 | 1E+08 | + | 3060 | protein_co | activating t | TF_bZIP | 555 | 555 | 581 | 629 | 671 | 749 | 9.539067 | 9.643271 | 9.340725 | 11.30261 | 11.73281 | 12.52943 |
| ENSMUSG0 | 3977.675 | 4113.798 | 3880.977 | 3625.21 | 3431.342 | 3667.728 | 0.158662 | 0.002805 | 0.033187 | Cnn3 | 3 | 1.21E+08 | 1.21E+08 | + | 3037 | protein_co | calponin 3, | - | 3992 | 4084 | 4164 | 3480 | 3385 | 3782 | 69.13215 | 71.49797 | 67.45153 | 63.00629 | 59.63685 | 63.74525 |
| ENSMUSG0 | 67.75599 | 69.50345 | 83.88279 | 46.87772 | 25.34226 | 30.06334 | 1.11511 | 0.002828 | 0.03343 | Cyyr1 | 16 | 85421533 | 85553397 | - | 8894 | protein_co | cysteine an | - | 68 | 69 | 90 | 45 | 25 | 31 | 0.402111 | 0.412482 | 0.497819 | 0.278205 | 0.150399 | 0.178417 |
| ENSMUSG0 | 291.9486 | 288.0868 | 338.3273 | 369.8131 | 404.4624 | 484.8926 | -0.45536 | 0.002847 | 0.033624 | Rwdd4a | 8 | 47533664 | 47552955 | + | 3352 | protein_co | RWD doma | - | 293 | 286 | 363 | 355 | 399 | 500 | 4.597248 | 4.536436 | 5.327562 | 5.823362 | 6.368977 | 7.635493 |
| ENSMUSG0 | 954.5624 | 906.5667 | 906.8662 | 1011.517 | 1157.634 | 1172.47 | -0.27214 | 0.002869 | 0.03385 | Itpr2 | 6 | 1.46E+08 | 1.47E+08 | - | 14802 | protein_co | inositol 1,4 | - | 958 | 900 | 973 | 971 | 1142 | 1209 | 3.403921 | 3.232771 | 3.233839 | 3.607018 | 4.128065 | 4.180969 |
| ENSMUSG0 | 5211.233 | 5309.459 | 5114.054 | 5555.53 | 5619.899 | 6410.28 | -0.1699 | 0.00288 | 0.033945 | S100a9 | 3 | 90692632 | 90695721 | - | 522 | protein_co | S100 calciu | - | 5230 | 5271 | 5487 | 5333 | 5544 | 6610 | 526.9453 | 536.8776 | 517.1188 | 561.7597 | 568.2684 | 648.1896 |
| ENSMUSG0 | 1847.347 | 2038.768 | 1919.984 | 1719.891 | 1712.123 | 1571.052 | 0.214944 | 0.002883 | 0.033953 | Atxn10 | 15 | 85336245 | 85463212 | + | 1937 | protein_co | ataxin 10 [S | - | 1854 | 2024 | 2060 | 1651 | 1689 | 1620 | 50.34014 | 55.55635 | 52.31949 | 46.86697 | 46.65528 | 42.81111 |
| ENSMUSG0 | 3151.65 | 3100.458 | 3239.74 | 2750.159 | 2884.963 | 2838.561 | 0.163751 | 0.002897 | 0.034088 | Mki67 | 7 | 1.36E+08 | 1.36E+08 | - | 10061 | protein_co | antigen ide | - | 3163 | 3078 | 3476 | 2640 | 2846 | 2927 | 16.53455 | 16.26598 | 16.9967 | 14.4282 | 15.13542 | 14.89199 |
| ENSMUSG0 | 2988.239 | 2980.59 | 3035.625 | 3333.527 | 3305.644 | 3389.399 | -0.15539 | 0.002904 | 0.034133 | Atp2b1 | 10 | 98914406 | 99026143 | + | 15218 | protein_co | ATPase, Ca | - | 2999 | 2959 | 3257 | 3200 | 3261 | 3495 | 10.36462 | 10.33809 | 10.52897 | 11.56224 | 11.46553 | 11.75603 |
| ENSMUSG0 | 868.871 | 779.6474 | 897.5459 | 737.5427 | 683.2273 | 627.451 | 0.314726 | 0.002906 | 0.034133 | Utp14b | 1 | 78658038 | 78671512 | + | 7679 | protein_co | UTP14B sm | - | 872 | 774 | 963 | 708 | 674 | 647 | 5.972362 | 5.359066 | 6.169465 | 5.069651 | 4.696302 | 4.312913 |
| ENSMUSG0 | 114.5873 | 112.8172 | 91.33904 | 40.62735 | 72.9857 | 54.30797 | 0.9232 | 0.002911 | 0.034168 | Cys1 | 12 | 24665833 | 24681813 | - | 2952 | protein_co | cystin 1 [So | - | 115 | 112 | 98 | 39 | 72 | 56 | 2.048877 | 2.017226 | 1.633186 | 0.726437 | 1.305019 | 0.971053 |
| ENSMUSG0 | 166.4008 | 200.452 | 184.5421 | 255.2231 | 244.2994 | 299.6636 | -0.53606 | 0.002932 | 0.034378 | Fkrp | 7 | 16809246 | 16816732 | - | 5337 | protein_co | fukutin rela | - | 167 | 199 | 198 | 245 | 241 | 309 | 1.645711 | 1.982479 | 1.82513 | 2.524169 | 2.416132 | 2.963687 |
| ENSMUSG0 | 1586.287 | 1611.674 | 1599.365 | 1828.231 | 1779.026 | 1910.477 | -0.202 | 0.002953 | 0.034591 | Ndfip1 | 18 | 38418975 | 38464399 | + | 930 | protein_co | Nedd4 fami | - | 1592 | 1600 | 1716 | 1755 | 1755 | 1970 | 90.03149 | 91.47235 | 90.77374 | 103.7633 | 100.9706 | 108.4312 |
| ENSMUSG0 | 301.9127 | 325.3567 | 331.803 | 205.2202 | 252.4089 | 230.8089 | 0.478156 | 0.002965 | 0.034709 | Kdr | 5 | 75932827 | 75978458 | - | 9030 | protein_co | kinase inse | - | 303 | 323 | 356 | 197 | 249 | 238 | 1.764774 | 1.901812 | 1.939493 | 1.199576 | 1.475409 | 1.34915 |
| ENSMUSG0 | 1045.236 | 928.7273 | 1048.535 | 1221.946 | 1140.402 | 1262.66 | -0.26207 | 0.00299 | 0.034965 | Lemd2 | 17 | 27189601 | 27204438 | - | 2595 | protein_co | LEM domai | - | 1049 | 922 | 1125 | 1173 | 1125 | 1302 | 21.26045 | 18.89063 | 21.32755 | 24.85479 | 23.19615 | 25.68294 |
| ENSMUSG0 | 879.8315 | 899.5157 | 950.6717 | 1163.609 | 1002.54 | 1133.679 | -0.2732 | 0.003018 | 0.035261 | Zfp703 | 8 | 26977325 | 26981461 | + | 3392 | protein_co | zinc finger | - | 883 | 893 | 1020 | 1117 | 989 | 1169 | 13.69113 | 13.99743 | 14.79348 | 18.10701 | 15.6006 | 17.64127 |
| ENSMUSG0 | 35.87082 | 32.23348 | 36.34921 | 16.66763 | 3.041071 | 9.697852 | 1.831989 | 0.003024 | 0.035268 | Vgll3 | 16 | 65815257 | 65866368 | + | 7104 | protein_co | vestigial lik | - | 36 | 32 | 39 | 16 | 3 | 10 | 0.266523 | 0.239497 | 0.270077 | 0.123842 | 0.022595 | 0.072056 |
| ENSMUSG0 | 1339.177 | 1357.836 | 1529.463 | 1704.265 | 1570.206 | 1701.973 | -0.23524 | 0.003025 | 0.035268 | Loxl4 | 19 | 42593982 | 42612813 | - | 4213 | protein_co | lysyl oxidas | - | 1344 | 1348 | 1641 | 1636 | 1549 | 1755 | 16.77808 | 17.01184 | 19.1621 | 21.35213 | 19.67256 | 21.32341 |
| ENSMUSG0 | 1747.706 | 1622.754 | 1615.21 | 1840.732 | 2020.285 | 1907.567 | -0.21061 | 0.003027 | 0.035268 | Ghitm | 14 | 37120092 | 37135272 | - | 3756 | protein_co | growth hor | - | 1754 | 1611 | 1733 | 1767 | 1993 | 1967 | 24.56056 | 22.80462 | 22.69859 | 25.86786 | 28.39112 | 26.8071 |
| ENSMUSG0 | 3504.38 | 3306.954 | 3330.147 | 3209.561 | 2941.729 | 2720.247 | 0.193289 | 0.003048 | 0.03549 | Nisch | 14 | 31170930 | 31216946 | - | 13198 | protein_co | nischarin [S | - | 3517 | 3283 | 3573 | 3081 | 2902 | 2805 | 14.01518 | 13.22561 | 13.31836 | 12.8361 | 11.76495 | 10.87917 |
| ENSMUSG0 | 77.72011 | 105.7661 | 132.3484 | 57.29499 | 54.73928 | 56.24754 | 0.909535 | 0.003057 | 0.035536 | C130074G | 1 | 1.85E+08 | 1.85E+08 | - | 3035 | protein_co | RIKEN cDN | - | 78 | 105 | 142 | 55 | 54 | 58 | 1.351669 | 1.839431 | 2.301736 | 0.996445 | 0.951998 | 0.978229 |
| ENSMUSG0 | 1239.536 | 1352.799 | 1303.911 | 1732.392 | 1430.317 | 1492.499 | -0.25649 | 0.003058 | 0.035536 | Bcl9l | 9 | 44482825 | 44511896 | + | 7998 | protein_co | B cell CLL/l | - | 1244 | 1343 | 1399 | 1663 | 1411 | 1539 | 8.180377 | 8.927861 | 8.605225 | 11.433 | 9.439444 | 9.84982 |
| ENSMUSG0 | 397.5683 | 349.5318 | 392.3851 | 275.0159 | 304.1071 | 263.7816 | 0.435418 | 0.003063 | 0.035567 | Prcp | 7 | 92874470 | 92934583 | + | 5497 | protein_co | prolylcarbo | - | 399 | 347 | 421 | 264 | 300 | 272 | 3.817522 | 3.356267 | 3.767752 | 2.640752 | 2.920091 | 2.532878 |
| ENSMUSG0 | 767.237 | 755.4723 | 716.7319 | 796.9212 | 1087.69 | 942.6312 | -0.33668 | 0.003097 | 0.035933 | Ube3a | 7 | 59228750 | 59311536 | + | 20626 | protein_co | ubiquitin p | - | 770 | 750 | 769 | 765 | 1073 | 972 | 1.963406 | 1.933299 | 1.83416 | 2.039369 | 2.783463 | 2.41225 |
| ENSMUSG0 | 448.3853 | 525.8087 | 452.0351 | 298.9757 | 418.6541 | 326.8176 | 0.449096 | 0.003108 | 0.036027 | Gstz1 | 12 | 87147165 | 87164723 | + | 5088 | protein_co | glutathione | - | 450 | 522 | 485 | 287 | 413 | 337 | 4.651572 | 5.454767 | 4.689435 | 3.101589 | 4.343139 | 3.390423 |
| ENSMUSG0 | 3077.916 | 3134.706 | 3011.392 | 3302.275 | 3432.355 | 3610.51 | -0.16579 | 0.003114 | 0.036032 | Glud1 | 14 | 34310727 | 34345265 | + | 4210 | protein_co | glutamate | - | 3089 | 3112 | 3231 | 3170 | 3386 | 3723 | 38.58959 | 39.30161 | 37.75555 | 41.40251 | 43.0334 | 45.26703 |
| ENSMUSG0 | 328.8159 | 215.5614 | 253.5124 | 321.8937 | 469.3386 | 376.2767 | -0.54941 | 0.003114 | 0.036032 | Orai2 | 5 | 1.36E+08 | 1.36E+08 | - | 6077 | protein_co | ORAI calciu | - | 330 | 214 | 272 | 309 | 463 | 388 | 2.856006 | 1.872308 | 2.20194 | 2.795881 | 4.076548 | 3.268237 |
| ENSMUSG0 | 34.87441 | 26.18971 | 33.55312 | 15.62591 | 0 | 2.909356 | 2.361881 | 0.003124 | 0.036112 | Gm29340 | 2 | 1.17E+08 | 1.17E+08 | - | 3854 | lincRNA | predicted g | - | 35 | 26 | 36 | 15 | 0 | 3 | 0.477629 | 0.358686 | 0.459533 | 0.214008 | 0 | 0.039846 |
| ENSMUSG0 | 173.3756 | 180.3061 | 193.8625 | 132.2993 | 117.5881 | 103.767 | 0.63201 | 0.003139 | 0.036251 | Fbxo5 | 10 | 5799160 | 5805600 | - | 2003 | protein_co | F-box prote | - | 174 | 179 | 208 | 127 | 116 | 107 | 4.568805 | 4.751436 | 5.108675 | 3.48636 | 3.098688 | 2.734475 |
| ENSMUSG0 | 114.5873 | 113.8245 | 128.6203 | 68.75398 | 59.80773 | 77.58282 | 0.792147 | 0.003145 | 0.036266 | Flywch1 | 17 | 23752793 | 23771602 | - | 4266 | protein_co | FLYWCH-ty | - | 115 | 113 | 138 | 66 | 59 | 80 | 1.417788 | 1.408349 | 1.591418 | 0.850692 | 0.74 | 0.959931 |
| ENSMUSG0 | 2754.082 | 2793.233 | 2961.995 | 2423.057 | 2101.38 | 2705.701 | 0.234904 | 0.003147 | 0.036266 | Gnas | 2 | 1.74E+08 | 1.74E+08 | + | 11186 | protein_co | GNAS (guar | - | 2764 | 2773 | 3178 | 2326 | 2073 | 2790 | 12.99564 | 13.18038 | 13.97671 | 11.43364 | 9.915744 | 12.76734 |
| ENSMUSG0 | 887.8028 | 815.9101 | 835.0998 | 982.3486 | 1038.019 | 1026.033 | -0.26311 | 0.003148 | 0.036266 | Ppp2cb | 8 | 33599625 | 33619441 | + | 1469 | protein_co | protein ph | - | 891 | 810 | 896 | 943 | 1024 | 1058 | 31.89997 | 29.31676 | 30.00628 | 35.29713 | 37.29744 | 36.86676 |
| ENSMUSG0 | 2342.564 | 2291.599 | 2218.234 | 2108.456 | 2020.285 | 1610.813 | 0.256047 | 0.00317 | 0.036487 | Ccnl2 | 4 | 1.56E+08 | 1.56E+08 | + | 5278 | protein_co | cyclin L2 [S | - | 2351 | 2275 | 2380 | 2024 | 1993 | 1661 | 23.42705 | 22.91737 | 22.18367 | 21.08582 | 20.20406 | 16.1091 |
| ENSMUSG0 | 1890.193 | 2153.6 | 2072.837 | 1883.442 | 1558.042 | 1738.825 | 0.239851 | 0.003176 | 0.036517 | Mtch1 | 17 | 29332072 | 29347904 | - | 2878 | protein_co | mitochond | - | 1897 | 2138 | 2224 | 1808 | 1537 | 1793 | 34.66657 | 39.49751 | 38.0163 | 34.54277 | 28.57485 | 31.89045 |
| ENSMUSG0 | 1174.769 | 1051.617 | 1104.457 | 941.7212 | 883.9379 | 955.2384 | 0.260273 | 0.003191 | 0.036661 | Vit | 17 | 78508064 | 78627408 | + | 3227 | protein_co | vitrin [Sour | - | 1179 | 1044 | 1185 | 904 | 872 | 985 | 19.21539 | 17.20103 | 18.0653 | 15.40348 | 14.45834 | 15.62458 |
| ENSMUSG0 | 6844.352 | 6433.602 | 6792.642 | 7211.876 | 7575.307 | 7223.93 | -0.13307 | 0.003202 | 0.036752 | Nr4a2 | 2 | 57106830 | 57124003 | - | 4059 | protein_co | nuclear rec | NGFIB-like | 6869 | 6387 | 7288 | 6923 | 7473 | 7449 | 89.00385 | 83.66247 | 88.33142 | 93.78313 | 98.50919 | 93.93988 |
| ENSMUSG0 | 490.2345 | 566.1006 | 458.5593 | 648.9959 | 652.8165 | 624.5417 | -0.34722 | 0.003238 | 0.037139 | Slc7a6 | 8 | 1.06E+08 | 1.06E+08 | + | 4570 | protein_co | solute carri | - | 492 | 562 | 492 | 623 | 644 | 644 | 5.662174 | 6.538421 | 5.296327 | 7.495857 | 7.539985 | 7.213412 |
| ENSMUSG0 | 752.2908 | 754.465 | 696.2272 | 864.6334 | 933.6088 | 877.6556 | -0.28094 | 0.003244 | 0.037165 | Klhl24 | 16 | 20097542 | 20129221 | + | 10183 | protein_co | kelch-like 2 | - | 755 | 749 | 747 | 830 | 921 | 905 | 3.89947 | 3.910739 | 3.608866 | 4.481793 | 4.839324 | 4.549293 |
| ENSMUSG0 | 505.1807 | 495.5898 | 539.646 | 593.7844 | 686.2683 | 663.3331 | -0.33516 | 0.003246 | 0.037165 | Usp31 | 7 | 1.22E+08 | 1.22E+08 | - | 10304 | protein_co | ubiquitin s | - | 507 | 492 | 579 | 570 | 677 | 684 | 2.587834 | 2.538704 | 2.764386 | 3.041715 | 3.515472 | 3.397984 |
| ENSMUSG0 | 588.8793 | 564.086 | 588.1116 | 637.5369 | 774.4594 | 782.6167 | -0.33436 | 0.003256 | 0.037241 | Tnfrsf10b | 14 | 69767472 | 69784411 | + | 5451 | protein_co | tumor necr | - | 591 | 560 | 631 | 612 | 764 | 807 | 5.702242 | 5.462163 | 5.694808 | 6.173404 | 7.499253 | 7.578241 |
| ENSMUSG0 | 3411.714 | 3210.254 | 3389.797 | 3569.999 | 3865.201 | 3773.434 | -0.16292 | 0.003262 | 0.037285 | Etf1 | 18 | 34902785 | 34932007 | - | 3709 | protein_co | eukaryotic | - | 3424 | 3187 | 3637 | 3427 | 3813 | 3891 | 48.55246 | 45.68546 | 48.24056 | 50.80503 | 55.00609 | 53.70015 |
| ENSMUSG0 | 2549.817 | 2659.262 | 2541.649 | 2784.536 | 3380.657 | 2834.682 | -0.21562 | 0.003268 | 0.037316 | Grem1 | 2 | 1.14E+08 | 1.14E+08 | - | 4148 | protein_co | gremlin 1, I | - | 2559 | 2640 | 2727 | 2673 | 3335 | 2923 | 32.44635 | 33.83904 | 32.3424 | 35.43314 | 43.01876 | 36.07125 |
| ENSMUSG0 | 14.94618 | 38.27726 | 37.28124 | 7.292089 | 1.01369 | 11.63742 | 2.177828 | 0.00328 | 0.037424 | Oxld1 | 11 | 1.2E+08 | 1.2E+08 | - | 813 | protein_co | oxidoreduc | - | 15 | 38 | 40 | 7 | 1 | 12 | 0.970365 | 2.485111 | 2.420446 | 0.473431 | 0.065813 | 0.755548 |
| ENSMUSG0 | 701.4738 | 781.662 | 705.5475 | 606.2851 | 548.4064 | 603.2064 | 0.315944 | 0.003285 | 0.037454 | Panx3 | 9 | 37659902 | 37669235 | - | 3434 | protein_co | pannexin 3 | - | 704 | 776 | 757 | 582 | 541 | 622 | 10.78218 | 12.01474 | 10.8448 | 9.319061 | 8.429422 | 9.271738 |
| ENSMUSG0 | 0.996412 | 10.07296 | 10.25234 | 17.70936 | 50.68451 | 26.1842 | -2.14566 | 0.003295 | 0.03752 | Epha8 | 4 | 1.37E+08 | 1.37E+08 | - | 4713 | protein_co | Eph recept | - | 1 | 10 | 11 | 17 | 50 | 27 | 0.011159 | 0.112812 | 0.114821 | 0.198336 | 0.567641 | 0.29325 |
| ENSMUSG0 | 620.7645 | 576.1735 | 564.8108 | 753.1686 | 717.6927 | 707.9432 | -0.30658 | 0.003302 | 0.03752 | Rasa3 | 8 | 13566948 | 13677603 | - | 5095 | protein_co | RAS p21 pr | - | 623 | 572 | 606 | 723 | 708 | 730 | 6.430995 | 5.969042 | 5.851327 | 7.802676 | 7.435152 | 7.334149 |
| ENSMUSG0 | 253.0886 | 221.6052 | 235.8039 | 135.4245 | 199.697 | 127.0419 | 0.620864 | 0.003302 | 0.03752 | Cryz | 3 | 1.55E+08 | 1.55E+08 | + | 6195 | protein_co | crystallin, z | - | 254 | 220 | 253 | 130 | 197 | 131 | 2.156387 | 1.88814 | 2.009116 | 1.153856 | 1.701476 | 1.082433 |
| ENSMUSG0 | 138.5012 | 166.2039 | 126.7562 | 239.5972 | 179.4232 | 242.4463 | -0.61753 | 0.003303 | 0.03752 | Gm4524 | 16 | 25956475 | 25960342 | + | 2152 | antisense | predicted g | - | 139 | 165 | 136 | 230 | 177 | 250 | 3.397089 | 4.076566 | 3.109013 | 5.87672 | 4.400802 | 5.946601 |
| ENSMUSG0 | 290.9522 | 278.0138 | 194.7945 | 151.0504 | 179.4232 | 174.5613 | 0.595247 | 0.003312 | 0.037582 | Pde7b | 10 | 20398004 | 20725078 | - | 12444 | protein_co | phosphodi | - | 292 | 276 | 209 | 145 | 177 | 180 | 1.234119 | 1.179239 | 0.826251 | 0.640704 | 0.761052 | 0.740429 |
| ENSMUSG0 | 799.1222 | 820.9465 | 731.6444 | 945.8881 | 987.3343 | 915.4772 | -0.27693 | 0.003314 | 0.037582 | Osbpl5 | 7 | 1.44E+08 | 1.44E+08 | - | 4915 | protein_co | oxysterol b | - | 802 | 815 | 785 | 908 | 974 | 944 | 8.581934 | 8.816311 | 7.857276 | 10.15808 | 10.60318 | 9.831495 |
| ENSMUSG0 | 558.987 | 516.743 | 488.3843 | 644.829 | 628.488 | 693.3964 | -0.33108 | 0.003323 | 0.037653 | Prkaa1 | 15 | 5143861 | 5181899 | + | 4655 | protein_co | protein kin | - | 561 | 513 | 524 | 619 | 620 | 715 | 6.338369 | 5.859364 | 5.537803 | 7.311735 | 7.126443 | 7.862442 |
| ENSMUSG0 | 641.6891 | 591.283 | 662.6741 | 786.5039 | 764.3225 | 778.7375 | -0.29696 | 0.003353 | 0.037959 | Dhx38 | 8 | 1.1E+08 | 1.1E+08 | - | 4967 | protein_co | DEAH (Asp- | - | 644 | 587 | 711 | 755 | 754 | 803 | 6.819084 | 6.283429 | 7.042086 | 8.357998 | 8.122281 | 8.275466 |
| ENSMUSG0 | 76.7237 | 74.53993 | 27.96093 | 16.66763 | 15.20535 | 32.9727 | 1.46148 | 0.00336 | 0.038009 | B930059L( | 12 | 1.11E+08 | 1.11E+08 | + | 2189 | lincRNA | RIKEN cDN | - | 77 | 74 | 30 | 16 | 15 | 34 | 1.850032 | 1.797375 | 0.67422 | 0.401905 | 0.366645 | 0.795068 |
| ENSMUSG0 | 32.88159 | 29.21159 | 52.19374 | 17.70936 | 12.16428 | 5.818711 | 1.686059 | 0.003366 | 0.038047 | Ttn | 2 | 76703980 | 76982547 | - | 116825 | protein_co | titin [Sourc | - | 33 | 29 | 56 | 17 | 12 | 6 | 0.014856 | 0.013198 | 0.023582 | 0.008001 | 0.005496 | 0.002629 |
| ENSMUSG0 | 1324.231 | 1279.266 | 1529.463 | 1539.673 | 1613.795 | 1790.223 | -0.25822 | 0.003373 | 0.038087 | Baiap2 | 11 | 1.2E+08 | 1.2E+08 | + | 4992 | protein_co | brain-speci | - | 1329 | 1270 | 1641 | 1478 | 1592 | 1846 | 14.00183 | 13.52639 | 16.17186 | 16.27981 | 17.06355 | 18.92902 |
| ENSMUSG0 | 44.83853 | 38.27726 | 60.58202 | 19.79281 | 5.068451 | 23.27484 | 1.577122 | 0.00338 | 0.038131 | Snora31 | 14 | 75847905 | 75848034 | + | 130 | snoRNA | small nucle | - | 45 | 38 | 65 | 19 | 5 | 24 | 18.20554 | 15.5415 | 24.59778 | 8.036366 | 2.057915 | 9.450156 |
| ENSMUSG0 | 208.25 | 192.3936 | 248.8523 | 223.9713 | 408.5172 | 361.7299 | -0.61411 | 0.003394 | 0.038265 | Gm10180 | 9 | 25101404 | 25101481 | - | 78 | processed_ | predicted g | - | 209 | 191 | 267 | 215 | 403 | 373 | 140.9243 | 130.1942 | 168.4002 | 151.563 | 276.4466 | 244.7853 |
| ENSMUSG0 | 1715.821 | 1707.367 | 1706.549 | 1857.399 | 1965.545 | 2076.31 | -0.20195 | 0.003398 | 0.038277 | Ube2j1 | 4 | 33031416 | 33052363 | + | 3530 | protein_co | ubiquitin-c | - | 1722 | 1695 | 1831 | 1783 | 1939 | 2141 | 25.65623 | 25.52982 | 25.51758 | 27.77321 | 29.39029 | 31.04653 |
| ENSMUSG0 | 594.8578 | 564.086 | 525.6655 | 769.8363 | 652.8165 | 694.3662 | -0.32976 | 0.00341 | 0.038372 | Arl8a | 1 | 1.35E+08 | 1.35E+08 | + | 2299 | protein_co | ADP-ribosy | - | 597 | 560 | 564 | 739 | 644 | 716 | 13.65745 | 12.95096 | 12.06885 | 17.67482 | 14.98814 | 15.94209 |
| ENSMUSG0 | 100.6376 | 106.7734 | 113.7078 | 187.5109 | 225.0392 | 123.1627 | -0.73704 | 0.003435 | 0.038618 | Rnf219 | 14 | 1.04E+08 | 1.05E+08 | - | 4435 | protein_co | ring finger | - | 101 | 106 | 122 | 180 | 222 | 127 | 1.197739 | 1.270764 | 1.353294 | 2.231662 | 2.678306 | 1.465822 |
| ENSMUSG0 | 45.83494 | 44.32104 | 48.46561 | 27.0849 | 11.15059 | 12.60721 | 1.451323 | 0.003441 | 0.038634 | Slc24a3 | 2 | 1.45E+08 | 1.46E+08 | + | 4564 | protein_co | solute carri | - | 46 | 44 | 52 | 26 | 11 | 13 | 0.530086 | 0.512578 | 0.56051 | 0.31324 | 0.128958 | 0.145804 |
| ENSMUSG0 | 845.9535 | 777.6328 | 746.5569 | 1089.646 | 870.76 | 971.7248 | -0.30701 | 0.003442 | 0.038634 | Sirpa | 2 | 1.3E+08 | 1.3E+08 | + | 4911 | protein_co | signal-regu | - | 849 | 772 | 801 | 1046 | 859 | 1002 | 9.092265 | 8.357958 | 8.023955 | 11.71146 | 9.358883 | 10.44405 |
| ENSMUSG0 | 174.372 | 151.0945 | 170.5617 | 257.3066 | 213.8886 | 252.1442 | -0.54393 | 0.003446 | 0.038647 | Bbc3 | 7 | 16308393 | 16318205 | + | 2124 | protein_co | BCL2 bindi | - | 175 | 150 | 183 | 247 | 211 | 260 | 4.333291 | 3.754824 | 4.2386 | 6.394284 | 5.315312 | 6.265993 |
| ENSMUSG0 | 1415.901 | 1364.887 | 1546.239 | 1885.526 | 1620.891 | 1622.451 | -0.24463 | 0.003471 | 0.038881 | Csnk1g2 | 10 | 80622838 | 80640749 | + | 4835 | protein_co | casein kina | - | 1421 | 1355 | 1659 | 1810 | 1599 | 1673 | 15.45724 | 14.90032 | 16.88013 | 20.58408 | 17.69509 | 17.71212 |
| ENSMUSG0 | 917.6952 | 964.9899 | 848.1482 | 770.878 | 790.6784 | 611.9345 | 0.329646 | 0.003473 | 0.038881 | Pamr1 | 2 | 1.03E+08 | 1.03E+08 | + | 6046 | protein_co | peptidase c | - | 921 | 958 | 910 | 740 | 780 | 631 | 8.011721 | 8.424617 | 7.404558 | 6.729969 | 6.902831 | 5.34235 |
| ENSMUSG0 | 14.94618 | 21.15322 | 23.30078 | 50.0029 | 63.86249 | 36.85184 | -1.34093 | 0.003492 | 0.039059 | Zfp763 | 17 | 33016863 | 33033402 | - | 3557 | protein_co | zinc finger | zf-C2H2 | 15 | 21 | 25 | 48 | 63 | 38 | 0.22179 | 0.313898 | 0.345766 | 0.742005 | 0.947671 | 0.546853 |
| ENSMUSG0 | 558.987 | 642.6551 | 625.3928 | 783.3787 | 758.2403 | 722.49 | -0.30909 | 0.003495 | 0.039059 | Usp4 | 9 | 1.08E+08 | 1.08E+08 | + | 8628 | protein_co | ubiquitin s | - | 561 | 638 | 671 | 752 | 748 | 745 | 3.419693 | 3.931546 | 3.825941 | 4.792445 | 4.638657 | 4.419949 |

| gene_id | H1 | H2 | H3 | M1 | M2 | M3 | log2FoldCh | pvalue | padj | gene_name | gene_chr | gene_start | gene_end | gene_stran | gene_lengt | gene_bioty | gene_descr | tf_family | H1_readco | H2_readco | H3_readco | M1_readco | M2_readco | M3_readco | H1_fpkm | H2_fpkm | H3_fpkm | M1_fpkm | M2_fpkm | M3_fpkm |
|---|---|---|---|---|---|---|---|---|---|---|---|---|---|---|---|---|---|---|---|---|---|---|---|---|---|---|---|---|---|---|
| ENSMUSG0 | 210.2429 | 261.8971 | 284.2695 | 341.6865 | 342.6273 | 364.6392 | -0.47113 | 0.003498 | 0.039063 | Hilpda | 6 | 29272488 | 29275446 | + | 1103 | protein_co | hypoxia inc | - | 211 | 260 | 305 | 328 | 338 | 376 | 10.061 | 12.53287 | 13.60349 | 16.35113 | 16.39615 | 17.44952 |
| ENSMUSG0 | 510.1628 | 491.5606 | 518.2093 | 670.8722 | 604.1594 | 628.4208 | -0.32425 | 0.003507 | 0.039129 | Fut11 | 14 | 20694968 | 20701016 | + | 4252 | protein_co | fucosyltran | - | 512 | 488 | 556 | 644 | 596 | 648 | 6.333023 | 6.102101 | 6.432909 | 8.328026 | 7.499871 | 7.801046 |
| ENSMUSG0 | 837.9822 | 827.9976 | 747.4889 | 936.5126 | 1199.196 | 902.87 | -0.33246 | 0.003515 | 0.039191 | Btbd7 | 12 | 1.03E+08 | 1.03E+08 | - | 16456 | protein_co | BTB (POZ) d | - | 841 | 822 | 802 | 899 | 1183 | 931 | 2.687856 | 2.65583 | 2.397596 | 3.003896 | 3.84646 | 2.895986 |
| ENSMUSG0 | 1139.895 | 1105.004 | 1126.826 | 1358.412 | 1367.468 | 1241.325 | -0.23433 | 0.003526 | 0.039274 | Ncoa2 | 1 | 13139105 | 13374083 | - | 15642 | protein_co | nuclear rec | - | 1144 | 1097 | 1209 | 1304 | 1349 | 1280 | 3.846521 | 3.728783 | 3.802419 | 4.583896 | 4.614456 | 4.188792 |
| ENSMUSG0 | 79.71293 | 117.8537 | 102.5234 | 52.08635 | 33.45178 | 66.91518 | 0.975539 | 0.003551 | 0.039523 | Notch4 | 17 | 34564268 | 34588503 | + | 8061 | protein_co | notch 4 [So | - | 80 | 117 | 110 | 50 | 33 | 69 | 0.521958 | 0.771702 | 0.67132 | 0.34106 | 0.219041 | 0.438159 |
| ENSMUSG0 | 462.335 | 408.9623 | 452.9671 | 353.1455 | 272.6827 | 356.881 | 0.43037 | 0.003586 | 0.039872 | Limch1 | 5 | 66745827 | 67057158 | + | 10263 | protein_co | LIM and cal | - | 464 | 406 | 486 | 339 | 269 | 368 | 2.377815 | 2.103316 | 2.329635 | 1.816247 | 1.402422 | 1.835459 |
| ENSMUSG0 | 490.2345 | 492.5679 | 513.5491 | 637.5369 | 582.8719 | 658.4842 | -0.32837 | 0.003593 | 0.039918 | Tsc1 | 2 | 28641228 | 28691167 | + | 11833 | protein_co | tuberous sc | - | 492 | 489 | 551 | 612 | 575 | 679 | 2.186777 | 2.197186 | 2.290776 | 2.843846 | 2.600003 | 2.937284 |
| ENSMUSG0 | 1265.443 | 1458.565 | 1412.959 | 1807.396 | 1482.015 | 1653.484 | -0.25659 | 0.003639 | 0.040399 | Midn | 10 | 80148272 | 80158368 | + | 4307 | protein_co | midnolin [S | - | 1270 | 1448 | 1516 | 1735 | 1462 | 1705 | 15.50826 | 17.87502 | 17.31611 | 22.15002 | 18.1624 | 20.26379 |
| ENSMUSG0 | 228.1783 | 237.7219 | 321.5507 | 398.9815 | 346.6821 | 359.7903 | -0.48779 | 0.00366 | 0.040601 | Bahcc1 | 11 | 1.2E+08 | 1.2E+08 | + | 11118 | protein_co | BAH domai | - | 229 | 236 | 345 | 383 | 342 | 371 | 1.083286 | 1.128595 | 1.526576 | 1.894181 | 1.645888 | 1.70812 |
| ENSMUSG0 | 528.0982 | 559.0495 | 466.0155 | 435.4419 | 362.9011 | 389.8537 | 0.386209 | 0.003665 | 0.04062 | Sox8 | 17 | 25565892 | 25570686 | - | 3118 | protein_co | SRY (sex det | HMG | 530 | 555 | 500 | 418 | 358 | 402 | 8.93993 | 9.46389 | 7.888961 | 7.371394 | 6.143385 | 6.599652 |
| ENSMUSG0 | 1460.74 | 1616.711 | 1400.843 | 1692.806 | 1768.89 | 1755.311 | -0.22073 | 0.003671 | 0.040647 | Rsl1d1 | 16 | 11192970 | 11203331 | - | 2258 | protein_co | ribosomal l | - | 1466 | 1605 | 1503 | 1625 | 1745 | 1810 | 34.14636 | 37.79235 | 32.7462 | 39.57117 | 41.34969 | 41.03229 |
| ENSMUSG0 | 1423.872 | 1412.23 | 1345.853 | 1611.552 | 1645.219 | 1573.961 | -0.2082 | 0.003691 | 0.040832 | Syvn1 | 19 | 6046576 | 6053712 | + | 4772 | protein_co | synovial ap | - | 1429 | 1402 | 1444 | 1547 | 1623 | 1623 | 15.74948 | 15.6207 | 14.8865 | 17.8254 | 18.1978 | 17.40961 |
| ENSMUSG0 | 19.92823 | 21.15322 | 35.41718 | 62.50362 | 35.47916 | 94.06916 | -1.326 | 0.003706 | 0.040971 | Adgra1 | 7 | 1.4E+08 | 1.4E+08 | + | 5386 | protein_co | adhesion G | - | 20 | 21 | 38 | 60 | 35 | 97 | 0.195298 | 0.207303 | 0.347091 | 0.61254 | 0.347698 | 0.921884 |
| ENSMUSG0 | 688.5205 | 664.8156 | 695.2951 | 523.9887 | 577.8035 | 551.8078 | 0.309031 | 0.003751 | 0.041427 | Tpx2 | 2 | 1.53E+08 | 1.53E+08 | + | 4677 | protein_co | TPX2, micr | - | 691 | 660 | 746 | 503 | 570 | 569 | 7.770429 | 7.502904 | 7.846886 | 5.913575 | 6.520911 | 6.227532 |
| ENSMUSG0 | 12112.38 | 12087.56 | 12854.57 | 13345.57 | 13752.74 | 13095.98 | -0.11722 | 0.003762 | 0.041519 | Psap | 10 | 60277627 | 60302597 | + | 2744 | protein_co | prosaposin | - | 12156 | 12000 | 13792 | 12811 | 13567 | 13504 | 232.992 | 232.5144 | 247.2686 | 256.7133 | 264.5456 | 251.9123 |
| ENSMUSG0 | 838.9786 | 697.0491 | 760.5373 | 883.3845 | 937.6635 | 995.9694 | -0.29495 | 0.003782 | 0.041706 | Zfp275 | X | 73342621 | 73359080 | + | 6745 | protein_co | zinc finger p | zf-C2H2 | 842 | 692 | 816 | 848 | 925 | 1027 | 6.56545 | 5.454776 | 5.951605 | 6.912949 | 7.337711 | 7.793985 |
| ENSMUSG0 | 448.3853 | 547.9692 | 592.7717 | 632.3283 | 698.4326 | 709.8828 | -0.36046 | 0.003796 | 0.041826 | Nip7 | 8 | 1.07E+08 | 1.07E+08 | + | 3130 | protein_co | NIP7, nucle | - | 450 | 544 | 636 | 607 | 689 | 732 | 7.561405 | 9.240753 | 9.996286 | 10.66335 | 11.77811 | 11.9712 |
| ENSMUSG0 | 621.7609 | 577.1808 | 574.1311 | 494.8203 | 419.6678 | 480.0437 | 0.346376 | 0.003809 | 0.04192 | Npepl1 | 2 | 1.74E+08 | 1.74E+08 | + | 2998 | protein_co | aminopept | - | 624 | 573 | 616 | 475 | 414 | 495 | 10.9468 | 10.16192 | 10.10823 | 8.711871 | 7.388725 | 8.451711 |
| ENSMUSG0 | 740.3339 | 726.2607 | 770.7897 | 938.5961 | 838.3219 | 937.7823 | -0.27879 | 0.003811 | 0.04192 | Axin1 | 17 | 26138688 | 26195811 | + | 3828 | protein_co | axin 1 [Sou | - | 743 | 721 | 827 | 901 | 827 | 967 | 10.20825 | 10.0142 | 10.62819 | 12.94203 | 11.55938 | 12.93081 |
| ENSMUSG0 | 310.8804 | 286.0722 | 356.0359 | 244.8059 | 191.5875 | 239.5369 | 0.496202 | 0.003823 | 0.042013 | Acadm | 3 | 1.54E+08 | 1.54E+08 | - | 3913 | protein_co | acyl-Coenzy | - | 312 | 284 | 382 | 235 | 189 | 247 | 4.193524 | 3.85888 | 4.802633 | 3.302231 | 2.584359 | 3.231158 |
| ENSMUSG0 | 483.2597 | 425.0791 | 391.453 | 516.6966 | 561.5844 | 603.2064 | -0.37241 | 0.003832 | 0.042074 | Naa30 | 14 | 49172226 | 49191031 | + | 6970 | protein_co | N(alpha)-ac | - | 485 | 422 | 420 | 496 | 554 | 622 | 3.659682 | 3.219085 | 2.964438 | 3.912897 | 4.252828 | 4.568027 |
| ENSMUSG0 | 216.2213 | 209.5176 | 183.6101 | 138.5497 | 110.4922 | 151.2865 | 0.605045 | 0.00392 | 0.043009 | Fes | 7 | 80377756 | 80387946 | - | 5074 | protein_co | feline sarco | - | 217 | 208 | 197 | 133 | 109 | 156 | 2.24928 | 2.179544 | 1.910036 | 1.441288 | 1.149415 | 1.573784 |
| ENSMUSG0 | 41.84929 | 54.394 | 49.39764 | 72.92089 | 87.17736 | 119.2836 | -0.94116 | 0.003926 | 0.043014 | Ip6k3 | 17 | 27143969 | 27167764 | - | 2307 | protein_co | inositol he | - | 42 | 54 | 53 | 70 | 86 | 123 | 0.957494 | 1.244512 | 1.130197 | 1.668399 | 1.994581 | 2.729157 |
| ENSMUSG0 | 5714.421 | 5359.824 | 5406.712 | 5674.287 | 6517.015 | 6217.293 | -0.1597 | 0.003928 | 0.043014 | Arcn1 | 9 | 44741564 | 44767845 | - | 14915 | protein_co | archain 1 [S | - | 5735 | 5321 | 5801 | 5447 | 6429 | 6411 | 20.22295 | 18.96805 | 19.13399 | 20.08092 | 23.06328 | 22.00258 |
| ENSMUSG0 | 621.7609 | 620.4946 | 671.9944 | 513.5714 | 512.9273 | 512.0466 | 0.315558 | 0.00393 | 0.043014 | Brip1os | 11 | 86201370 | 86304443 | + | 6498 | antisense | BRCA1 inte | - | 624 | 616 | 721 | 493 | 506 | 528 | 5.050556 | 5.04027 | 5.458602 | 4.171735 | 4.166502 | 4.159348 |
| ENSMUSG0 | 13.94976 | 28.2043 | 13.04843 | 52.08635 | 45.61606 | 45.5799 | -1.37931 | 0.003946 | 0.043154 | Pak6 | 2 | 1.19E+08 | 1.19E+08 | + | 5211 | protein_co | p21 (RAC1) | - | 14 | 28 | 14 | 50 | 45 | 47 | 0.1413 | 0.285687 | 0.13217 | 0.527592 | 0.462053 | 0.461687 |
| ENSMUSG0 | 5786.163 | 5750.655 | 5498.983 | 6239.945 | 6001.046 | 6503.38 | -0.13808 | 0.00395 | 0.043162 | Atf3 | 1 | 1.91E+08 | 1.91E+08 | - | 2154 | protein_co | activating t | TF_bZIP | 5807 | 5709 | 5900 | 5990 | 5920 | 6706 | 141.7883 | 140.9182 | 134.7511 | 152.9081 | 147.054 | 159.3635 |
| ENSMUSG0 | 22.91747 | 48.35023 | 20.50468 | 8.333816 | 12.16428 | 2.909356 | 1.974344 | 0.003966 | 0.043296 | Gap43 | 16 | 42248442 | 42340651 | - | 1523 | protein_co | growth ass | - | 23 | 48 | 22 | 8 | 12 | 3 | 0.794259 | 1.675692 | 0.710638 | 0.288828 | 0.421582 | 0.100831 |
| ENSMUSG0 | 916.6987 | 934.771 | 929.2349 | 746.9183 | 725.8022 | 827.2268 | 0.273457 | 0.003973 | 0.043334 | Chil3 | 3 | 1.06E+08 | 1.06E+08 | - | 1632 | protein_co | chitinase-li | - | 920 | 928 | 997 | 717 | 716 | 853 | 29.64845 | 30.23296 | 30.0539 | 24.1573 | 23.47436 | 26.75469 |
| ENSMUSG0 | 831.0073 | 859.2238 | 870.517 | 709.4161 | 714.6517 | 699.2151 | 0.270441 | 0.003993 | 0.043499 | Pja1 | X | 99465734 | 99471273 | - | 3324 | protein_co | praja ring fi | - | 834 | 853 | 934 | 681 | 705 | 721 | 13.19591 | 13.64397 | 13.8233 | 11.26511 | 11.34825 | 11.10313 |
| ENSMUSG0 | 1851.333 | 1666.068 | 1702.821 | 2027.201 | 2040.559 | 1931.812 | -0.20069 | 0.003994 | 0.043499 | Ykt6 | 11 | 5955693 | 5967780 | + | 2984 | protein_co | YKT6 v-SNA | - | 1858 | 1654 | 1827 | 1946 | 2013 | 1992 | 32.74773 | 29.47063 | 30.12074 | 35.85861 | 36.09489 | 34.17131 |
| ENSMUSG0 | 536.0695 | 418.028 | 472.5397 | 655.2463 | 612.2689 | 569.2639 | -0.36413 | 0.004001 | 0.043535 | Rundc1 | 11 | 1.01E+08 | 1.01E+08 | + | 3223 | protein_co | RUN domai | - | 538 | 415 | 507 | 629 | 604 | 587 | 8.779228 | 6.846058 | 7.738799 | 10.73099 | 10.02715 | 9.322854 |
| ENSMUSG0 | 30.88876 | 41.29915 | 26.09687 | 12.50072 | 9.123213 | 9.697852 | 1.650118 | 0.004009 | 0.043587 | Sema5b | 16 | 35541145 | 35664732 | + | 7080 | protein_co | sema doma | - | 31 | 41 | 28 | 12 | 9 | 10 | 0.230283 | 0.307896 | 0.194559 | 0.093196 | 0.068016 | 0.0723 |
| ENSMUSG0 | 48.82417 | 27.197 | 39.1453 | 14.58418 | 16.21904 | 1.93957 | 1.820982 | 0.00402 | 0.04364 | Actc1 | 2 | 1.14E+08 | 1.14E+08 | - | 1618 | protein_co | actin, alpha | - | 49 | 27 | 42 | 14 | 16 | 2 | 1.592766 | 0.887234 | 1.277017 | 0.475772 | 0.529106 | 0.063274 |
| ENSMUSG0 | 844.9571 | 806.8444 | 785.7022 | 1003.183 | 948.8141 | 964.9363 | -0.25915 | 0.004021 | 0.04364 | Rab3gap2 | 1 | 1.85E+08 | 1.85E+08 | + | 10800 | protein_co | RAB3 GTPa | - | 848 | 801 | 843 | 963 | 936 | 995 | 4.129585 | 3.943316 | 3.839987 | 4.902888 | 4.637169 | 4.715963 |
| ENSMUSG0 | 90.67346 | 92.67127 | 99.72732 | 46.87772 | 64.87618 | 38.79141 | 0.912283 | 0.004025 | 0.043655 | Ndnf | 6 | 65671590 | 65712326 | + | 10114 | protein_co | neuron-der | - | 91 | 92 | 107 | 45 | 64 | 40 | 0.473209 | 0.483635 | 0.520459 | 0.244647 | 0.338577 | 0.202445 |
| ENSMUSG0 | 6526.497 | 6711.616 | 6680.798 | 7463.974 | 7078.599 | 7157.015 | -0.12345 | 0.004039 | 0.043767 | Anxa5 | 3 | 36448923 | 36475894 | - | 1777 | protein_co | annexin A5 | - | 6550 | 6663 | 7168 | 7165 | 6983 | 7380 | 193.86 | 199.3587 | 198.4433 | 221.7064 | 210.2594 | 212.5886 |
| ENSMUSG0 | 691.5097 | 620.4946 | 626.3249 | 528.1556 | 509.8862 | 516.8955 | 0.317889 | 0.004043 | 0.043768 | Fundc2 | X | 75382399 | 75396459 | + | 2785 | protein_co | FUN14 don | - | 694 | 616 | 672 | 507 | 503 | 533 | 13.10595 | 11.76003 | 11.87053 | 10.00996 | 9.663703 | 9.796548 |
| ENSMUSG0 | 40.85288 | 55.4013 | 52.19374 | 21.87627 | 11.15059 | 25.21442 | 1.348618 | 0.004057 | 0.043894 | Hist1h4n | 13 | 21831767 | 21832196 | - | 430 | protein_co | histone clu | - | 41 | 55 | 56 | 21 | 11 | 26 | 5.014755 | 6.800597 | 6.406863 | 2.685346 | 1.368753 | 3.095109 |
| ENSMUSG0 | 2911.515 | 3033.977 | 3503.505 | 3710.632 | 3371.534 | 3828.712 | -0.20729 | 0.004086 | 0.044129 | Trim47 | 11 | 1.16E+08 | 1.16E+08 | - | 2735 | protein_co | tripartite m | - | 2922 | 3012 | 3759 | 3562 | 3326 | 3948 | 56.18977 | 58.55317 | 67.61466 | 71.61204 | 65.06774 | 73.89089 |
| ENSMUSG0 | 89.67705 | 112.8172 | 106.2515 | 165.6346 | 144.9577 | 176.5009 | -0.65792 | 0.004086 | 0.044129 | Riox1 | 12 | 83950608 | 83952951 | + | 2344 | protein_co | ribosomal c | - | 90 | 112 | 114 | 159 | 143 | 182 | 2.019386 | 2.540465 | 2.392617 | 3.72983 | 3.264219 | 3.974522 |
| ENSMUSG0 | 352.7297 | 367.6632 | 386.7929 | 388.5642 | 621.3921 | 512.0466 | -0.45922 | 0.004091 | 0.044145 | Mob4 | 1 | 55131231 | 55154899 | + | 4855 | protein_co | MOB family | - | 354 | 365 | 415 | 373 | 613 | 528 | 3.83485 | 3.997205 | 4.205182 | 4.224439 | 6.755726 | 5.56693 |
| ENSMUSG0 | 294.9379 | 344.4954 | 349.5116 | 231.2634 | 216.9297 | 266.6909 | 0.467828 | 0.004095 | 0.044159 | Pcdh18 | 3 | 49743296 | 49757325 | - | 5366 | protein_co | protocadhe | - | 296 | 342 | 375 | 222 | 214 | 275 | 2.901185 | 3.388662 | 3.438005 | 2.274845 | 2.13385 | 2.623331 |
| ENSMUSG0 | 523.1161 | 615.4581 | 471.6077 | 686.4981 | 694.3778 | 663.3331 | -0.34502 | 0.004117 | 0.044354 | Kctd3 | 1 | 1.89E+08 | 1.89E+08 | - | 8129 | protein_co | potassium c | - | 525 | 611 | 506 | 659 | 685 | 684 | 3.396695 | 3.996289 | 3.06224 | 4.457566 | 4.50873 | 4.307151 |
| ENSMUSG0 | 980.4691 | 946.8586 | 980.4966 | 728.1672 | 868.7326 | 802.9821 | 0.276818 | 0.004121 | 0.044363 | Mdm4 | 1 | 1.33E+08 | 1.33E+08 | - | 14798 | protein_co | transforme | - | 984 | 940 | 1052 | 699 | 857 | 828 | 3.497248 | 3.377362 | 3.497346 | 2.597309 | 3.098693 | 2.864167 |
| ENSMUSG0 | 962.5337 | 964.9899 | 909.6623 | 1191.736 | 1199.196 | 1020.214 | -0.26561 | 0.00413 | 0.04442 | Cars | 7 | 1.44E+08 | 1.44E+08 | - | 6583 | protein_co | cysteinyl-tR | - | 966 | 958 | 976 | 1144 | 1183 | 1052 | 7.717695 | 7.737389 | 7.293767 | 9.555461 | 9.615275 | 8.180182 |
| ENSMUSG0 | 942.6055 | 839.0779 | 787.5662 | 996.9328 | 1019.772 | 1097.797 | -0.27831 | 0.00414 | 0.044493 | Tmem41b | 7 | 1.1E+08 | 1.1E+08 | - | 5872 | protein_co | transmemb | - | 946 | 833 | 845 | 957 | 1006 | 1132 | 8.473043 | 7.542438 | 7.079402 | 8.961389 | 9.166694 | 9.868053 |
| ENSMUSG0 | 1589.277 | 1501.879 | 1407.367 | 1650.096 | 2038.531 | 1678.698 | -0.25495 | 0.004171 | 0.04479 | Sppl2a | 2 | 1.27E+08 | 1.27E+08 | - | 8665 | protein_co | signal pepti | - | 1595 | 1491 | 1510 | 1584 | 2011 | 1731 | 9.681139 | 9.148752 | 8.573029 | 10.05162 | 12.41779 | 10.22585 |
| ENSMUSG0 | 190.3146 | 190.379 | 181.7461 | 273.9742 | 264.5732 | 250.2046 | -0.48783 | 0.004176 | 0.04481 | Cog7 | 7 | 1.22E+08 | 1.22E+08 | - | 6483 | protein_co | componen | - | 191 | 189 | 195 | 263 | 261 | 258 | 1.5495 | 1.550025 | 1.479737 | 2.230638 | 2.154097 | 2.037111 |
| ENSMUSG0 | 17.93541 | 52.37941 | 13.04843 | 6.250362 | 8.109522 | 4.848926 | 2.117029 | 0.004201 | 0.045035 | Tulp2 | 7 | 45481877 | 45523632 | + | 3150 | protein_co | tubby-like p | Tub | 18 | 52 | 14 | 6 | 8 | 5 | 0.300536 | 0.877699 | 0.218647 | 0.104735 | 0.135888 | 0.081251 |
| ENSMUSG0 | 324.8302 | 304.2035 | 332.7351 | 206.262 | 255.45 | 237.5974 | 0.459576 | 0.004223 | 0.045239 | Cdk14 | 5 | 4803391 | 5420312 | - | 18877 | protein_co | cyclin-depe | - | 326 | 302 | 357 | 198 | 252 | 245 | 0.908278 | 0.850603 | 0.930382 | 0.576742 | 0.71428 | 0.664361 |
| ENSMUSG0 | 200.2787 | 230.6709 | 228.3476 | 172.9267 | 147.9988 | 116.3742 | 0.593894 | 0.004227 | 0.045248 | Myl9 | 2 | 1.57E+08 | 1.57E+08 | + | 1162 | protein_co | myosin, ligl | - | 201 | 229 | 245 | 166 | 146 | 120 | 9.097546 | 10.47809 | 10.37256 | 7.855095 | 6.722759 | 5.286232 |
| ENSMUSG0 | 65.76317 | 70.51075 | 56.85389 | 44.79426 | 16.21904 | 20.36549 | 1.249809 | 0.004237 | 0.045317 | Gm43980 | 6 | 49254851 | 49256462 | + | 1612 | TEC | predicted g | - | 66 | 70 | 61 | 43 | 16 | 21 | 2.153343 | 2.308797 | 1.861619 | 1.466739 | 0.531075 | 0.666846 |
| ENSMUSG0 | 872.8566 | 821.9538 | 783.8381 | 976.0982 | 1031.937 | 963.9665 | -0.26213 | 0.004248 | 0.045398 | Snx4 | 16 | 33251442 | 33300269 | + | 3988 | protein_co | sorting nex | - | 876 | 816 | 841 | 937 | 1018 | 994 | 11.55269 | 10.87897 | 10.37449 | 12.91915 | 13.6582 | 12.75858 |
| ENSMUSG0 | 1222.597 | 1147.311 | 1247.99 | 1111.523 | 953.8826 | 960.0874 | 0.258567 | 0.004268 | 0.045567 | Srgap2 | 1 | 1.31E+08 | 1.32E+08 | - | 12560 | protein_co | SLIT-ROBO | - | 1227 | 1139 | 1339 | 1067 | 941 | 990 | 5.137943 | 4.821552 | 5.244654 | 4.671155 | 4.008675 | 4.03475 |
| ENSMUSG0 | 387.6041 | 368.6705 | 445.5108 | 593.7844 | 438.9279 | 567.3243 | -0.41227 | 0.004272 | 0.045573 | Zmynd19 | 2 | 24949792 | 24962075 | + | 4168 | protein_co | zinc finger, | - | 389 | 366 | 478 | 570 | 433 | 585 | 4.908584 | 4.66881 | 5.641909 | 7.519632 | 5.558543 | 7.184544 |
| ENSMUSG0 | 11.95694 | 9.065667 | 17.70859 | 32.29354 | 34.46547 | 45.5799 | -1.53285 | 0.004275 | 0.045573 | Epcam | 17 | 87635979 | 87651106 | + | 2040 | protein_co | epithelial c | - | 12 | 9 | 19 | 31 | 34 | 47 | 0.309375 | 0.234566 | 0.458194 | 0.835566 | 0.891763 | 1.179339 |
| ENSMUSG0 | 411.518 | 330.3932 | 440.8507 | 550.0319 | 486.5713 | 519.8049 | -0.39514 | 0.004291 | 0.04571 | Pdcd11 | 19 | 47090766 | 47131143 | + | 6041 | protein_co | programme | - | 413 | 328 | 473 | 528 | 480 | 536 | 3.595635 | 2.886807 | 3.851928 | 4.805898 | 4.251412 | 4.54179 |
| ENSMUSG0 | 227.1819 | 269.9554 | 238.5999 | 180.2188 | 166.2452 | 167.7728 | 0.516782 | 0.004301 | 0.045778 | Hdac7 | 15 | 97792664 | 97844502 | - | 6755 | protein_co | histone dea | - | 228 | 268 | 256 | 173 | 164 | 173 | 1.775186 | 2.109416 | 1.864406 | 1.408219 | 1.29903 | 1.310967 |
| ENSMUSG0 | 304.902 | 302.1889 | 278.6773 | 339.603 | 411.5583 | 442.2221 | -0.43111 | 0.004314 | 0.045877 | Vps18 | 2 | 1.19E+08 | 1.19E+08 | + | 4786 | protein_co | VPS18 COR | - | 306 | 300 | 299 | 326 | 406 | 456 | 3.362661 | 3.332739 | 3.073438 | 3.745367 | 4.538937 | 4.877118 |
| ENSMUSG0 | 796.1329 | 815.9101 | 863.0607 | 1039.644 | 915.3623 | 1027.972 | -0.269 | 0.004323 | 0.045905 | Snx9 | 17 | 5841329 | 5931954 | + | 3617 | protein_co | sorting nex | - | 799 | 810 | 926 | 998 | 903 | 1060 | 11.61803 | 11.90664 | 12.59472 | 15.1716 | 13.35796 | 15.00128 |
| ENSMUSG0 | 1148.863 | 1055.647 | 1102.593 | 1325.077 | 1309.688 | 1244.234 | -0.22988 | 0.004324 | 0.045905 | Hcfc1 | X | 73942792 | 73966357 | - | 8360 | protein_co | host cell fac | - | 1153 | 1048 | 1183 | 1272 | 1292 | 1283 | 7.253663 | 6.665117 | 6.961525 | 8.36624 | 8.269077 | 7.855819 |
| ENSMUSG0 | 1579.313 | 1497.85 | 1587.249 | 1327.16 | 1430.317 | 1223.869 | 0.228821 | 0.004351 | 0.046147 | Cybrd1 | 2 | 71117923 | 71142926 | + | 5339 | protein_co | cytochrom | - | 1585 | 1487 | 1703 | 1274 | 1411 | 1262 | 15.61362 | 14.80825 | 15.69208 | 13.12076 | 14.1406 | 12.09959 |
| ENSMUSG0 | 573.9331 | 615.4581 | 554.5585 | 466.6937 | 465.2838 | 456.7688 | 0.32834 | 0.004353 | 0.046147 | Fzd2 | 11 | 1.03E+08 | 1.03E+08 | + | 3663 | protein_co | frizzled clas | - | 576 | 611 | 595 | 448 | 459 | 471 | 8.270274 | 8.868641 | 7.991089 | 6.724973 | 6.704657 | 6.581957 |
| ENSMUSG0 | 669.5887 | 745.3993 | 621.6647 | 913.5946 | 846.4314 | 768.0699 | -0.31196 | 0.004382 | 0.046415 | Ank1 | 8 | 22974844 | 23150497 | + | 13527 | protein_co | ankyrin 1, e | - | 672 | 740 | 667 | 877 | 835 | 792 | 2.612776 | 2.908593 | 2.425773 | 3.564902 | 3.302827 | 2.997055 |
| ENSMUSG0 | 156.4366 | 180.3061 | 168.6976 | 89.58853 | 134.8208 | 90.19002 | 0.684202 | 0.004396 | 0.046521 | Zmym6 | 4 | 1.27E+08 | 1.27E+08 | + | 7692 | protein_co | zinc finger, | - | 157 | 179 | 181 | 86 | 133 | 93 | 1.073482 | 1.237276 | 1.157618 | 0.614764 | 0.925152 | 0.618892 |
| ENSMUSG0 | 69.74882 | 80.58371 | 61.51405 | 36.46045 | 21.2875 | 41.70076 | 1.0892 | 0.004403 | 0.04656 | Pitpnm3 | 11 | 72047528 | 72135778 | - | 9815 | protein_co | PITPNM far | - | 70 | 80 | 66 | 35 | 21 | 43 | 0.375096 | 0.433364 | 0.330811 | 0.196077 | 0.11448 | 0.224259 |
| ENSMUSG0 | 180.3505 | 187.3571 | 227.4156 | 151.0504 | 127.725 | 110.5555 | 0.614129 | 0.004415 | 0.046648 | Ptprv | 1 | 1.35E+08 | 1.35E+08 | - | 6987 | processed_ | protein tyr | - | 181 | 186 | 244 | 145 | 126 | 114 | 1.362455 | 1.415386 | 1.718007 | 1.141108 | 0.964896 | 0.83519 |
| ENSMUSG0 | 105.6196 | 136.9923 | 136.0765 | 67.71226 | 76.02677 | 81.46196 | 0.749344 | 0.004423 | 0.046697 | Slfn5 | 11 | 82951349 | 82964840 | + | 6326 | protein_co | schlafen 5 [ | - | 106 | 136 | 146 | 65 | 75 | 84 | 0.881274 | 1.143043 | 1.135402 | 0.564981 | 0.634356 | 0.679706 |
| ENSMUSG0 | 186.329 | 144.0434 | 195.7265 | 108.3396 | 129.7524 | 94.06916 | 0.664779 | 0.004429 | 0.046716 | Itga2 | 13 | 1.15E+08 | 1.15E+08 | - | 7133 | protein_co | integrin alp | - | 187 | 143 | 210 | 104 | 128 | 97 | 1.378808 | 1.0659 | 1.448348 | 0.801698 | 0.960149 | 0.696098 |
| ENSMUSG0 | 417.4965 | 405.9404 | 363.4921 | 310.4347 | 315.2577 | 261.842 | 0.419566 | 0.004449 | 0.046892 | Tmem258 | 19 | 10204014 | 10207824 | + | 1611 | protein_co | transmemb | - | 419 | 403 | 390 | 298 | 311 | 270 | 13.67895 | 13.30033 | 11.90954 | 10.17115 | 10.32918 | 8.579052 |
| ENSMUSG0 | 464.3278 | 401.9113 | 436.1905 | 543.7815 | 562.5981 | 544.0495 | -0.34151 | 0.004453 | 0.046895 | Nkapd1 | 9 | 50605240 | 50617464 | - | 5321 | protein_co | NKAP doma | - | 466 | 399 | 468 | 522 | 555 | 561 | 4.606032 | 3.986873 | 4.326916 | 5.394196 | 5.580852 | 5.396854 |
| ENSMUSG0 | 980.4691 | 878.3624 | 912.4584 | 1080.271 | 1188.045 | 1049.308 | -0.2595 | 0.004456 | 0.046895 | Rbm27 | 18 | 42275353 | 42341542 | + | 6431 | protein_co | RNA bindin | - | 984 | 872 | 979 | 1037 | 1172 | 1082 | 8.047314 | 7.209261 | 7.489108 | 8.866449 | 9.751017 | 8.612314 |
| ENSMUSG0 | 682.542 | 745.3993 | 715.7998 | 744.8348 | 1134.319 | 889.293 | -0.36913 | 0.004474 | 0.047049 | Emb | 13 | 1.17E+08 | 1.17E+08 | + | 6906 | protein_co | embigin [Sc | - | 685 | 740 | 768 | 715 | 1119 | 917 | 5.21673 | 5.697154 | 5.470922 | 5.692839 | 8.669705 | 6.796946 |
| ENSMUSG0 | 102.6304 | 50.36482 | 56.85389 | 29.16836 | 42.57499 | 11.63742 | 1.333905 | 0.00451 | 0.04739 | Cma1 | 14 | 55941451 | 55944675 | - | 1090 | protein_co | chymase 1, | - | 103 | 50 | 61 | 28 | 42 | 12 | 4.96987 | 2.438913 | 2.753146 | 1.412476 | 2.061691 | 0.563541 |
| ENSMUSG0 | 630.7286 | 610.4216 | 617.9366 | 535.4477 | 489.6124 | 460.648 | 0.323982 | 0.004531 | 0.047569 | Skiv2l | 17 | 34839228 | 34850210 | - | 4977 | protein_co | superkiller | - | 633 | 606 | 663 | 514 | 483 | 475 | 6.689142 | 6.473778 | 6.553477 | 5.678648 | 5.192545 | 4.885365 |
| ENSMUSG0 | 874.8495 | 924.6981 | 789.4303 | 690.665 | 744.0487 | 683.6986 | 0.288992 | 0.004557 | 0.047801 | Pik3c2a | 7 | 1.16E+08 | 1.16E+08 | - | 11133 | protein_co | phosphatic | - | 878 | 918 | 847 | 663 | 734 | 705 | 4.147789 | 4.384128 | 3.742804 | 3.274544 | 3.527643 | 3.241515 |
| ENSMUSG0 | 52.80982 | 95.69316 | 74.56248 | 151.0504 | 107.4512 | 133.8304 | -0.8147 | 0.004576 | 0.047967 | Cbarp | 10 | 80130451 | 80140835 | - | 4996 | protein_co | calcium ch | - | 53 | 95 | 80 | 145 | 106 | 138 | 0.55794 | 1.011007 | 0.787759 | 1.595861 | 1.135231 | 1.413929 |
| ENSMUSG0 | 384.6149 | 453.2834 | 413.8218 | 266.6821 | 358.8464 | 301.6032 | 0.432714 | 0.004584 | 0.048013 | Rhoj | 12 | 75308322 | 75401456 | + | 6484 | protein_co | ras homolo | - | 386 | 450 | 444 | 256 | 354 | 311 | 3.130968 | 3.689965 | 3.368727 | 2.170933 | 2.921198 | 2.455209 |
| ENSMUSG0 | 738.3411 | 813.8955 | 834.1678 | 553.1571 | 553.4749 | 751.5835 | 0.360261 | 0.004593 | 0.048069 | Acaa1a | 9 | 1.19E+08 | 1.19E+08 | + | 4414 | protein_co | acetyl-Coer | - | 741 | 808 | 895 | 531 | 546 | 775 | 8.829177 | 9.732666 | 9.975085 | 6.614723 | 6.618524 | 8.987532 |
| ENSMUSG0 | 6077.115 | 5525.021 | 5749.699 | 6409.746 | 6501.809 | 6188.199 | -0.1384 | 0.004622 | 0.048326 | Tm9sf3 | 19 | 41210842 | 41264004 | - | 6144 | protein_co | transmemb | - | 6099 | 5485 | 6169 | 6153 | 6414 | 6381 | 52.20857 | 47.46552 | 49.39574 | 55.06621 | 55.85712 | 53.16289 |
| ENSMUSG0 | 502.1915 | 464.3636 | 451.103 | 379.1886 | 337.5589 | 380.1558 | 0.369764 | 0.004653 | 0.048621 | Ssbp2 | 13 | 91460283 | 91703429 | + | 7985 | protein_co | single-strar | - | 504 | 461 | 484 | 364 | 333 | 392 | 3.319632 | 3.069579 | 2.981922 | 2.506547 | 2.231362 | 2.512941 |
| ENSMUSG0 | 813.0719 | 653.7353 | 670.1303 | 820.8809 | 1004.567 | 855.3505 | -0.32734 | 0.004686 | 0.04892 | Apc | 18 | 34220924 | 34322189 | + | 15753 | protein_co | adenomatc | - | 816 | 649 | 719 | 788 | 991 | 882 | 2.724339 | 2.190454 | 2.245389 | 2.750505 | 3.365977 | 2.866001 |
| ENSMUSG0 | 2175.167 | 1865.513 | 2070.041 | 2229.296 | 2465.295 | 2349.79 | -0.20509 | 0.004695 | 0.048981 | Myo1e | 9 | 70207350 | 70399766 | + | 5704 | protein_co | myosin IE [S | - | 2183 | 1852 | 2221 | 2140 | 2432 | 2423 | 20.12837 | 17.26292 | 19.15556 | 20.62926 | 22.81313 | 21.74428 |

| gene_id | H1 | H2 | H3 | M1 | M2 | M3 | log2FoldCh | pvalue | padj | gene_name | gene_chr | gene_start | gene_end | gene_strand | gene_length | gene_bioty | gene_descr | tf_family | H1_readco | H2_readco | H3_readco | M1_readco | M2_readco | M3_readco | H1_fpkm | H2_fpkm | H3_fpkm | M1_fpkm | M2_fpkm | M3_fpkm |
|---|---|---|---|---|---|---|---|---|---|---|---|---|---|---|---|---|---|---|---|---|---|---|---|---|---|---|---|---|---|---|
| ENSMUSG0 | 85.6914 | 56.4086 | 70.83436 | 34.37699 | 39.53392 | 30.06334 | 1.035092 | 0.004721 | 0.049205 | Plcl1 | 1 | 55405921 | 55754285 | + | 6687 | protein_co | phospholip | - | 86 | 56 | 76 | 33 | 39 | 31 | 0.676397 | 0.445256 | 0.559124 | 0.271351 | 0.312057 | 0.237302 |
| ENSMUSG0 | 1600.237 | 1484.755 | 1569.54 | 1762.602 | 2155.106 | 1650.574 | -0.25838 | 0.004773 | 0.049642 | Myo5a | 9 | 75071015 | 75223688 | + | 14803 | protein_co | myosin VA | - | 1606 | 1474 | 1684 | 1692 | 2126 | 1702 | 5.705979 | 5.294202 | 5.596522 | 6.284925 | 7.684477 | 5.885467 |
| ENSMUSG0 | 116.5802 | 145.0507 | 117.4359 | 83.33816 | 55.75297 | 82.43174 | 0.774359 | 0.004774 | 0.049642 | Gal3st1 | 11 | 3983636 | 3999326 | + | 2041 | protein_co | galactose-3 | - | 117 | 144 | 126 | 80 | 55 | 85 | 3.01493 | 3.751218 | 3.03706 | 2.155244 | 1.441851 | 2.131803 |
| ENSMUSG0 | 24.91029 | 25.18241 | 33.55312 | 4.166908 | 6.082142 | 12.60721 | 1.865442 | 0.004776 | 0.049642 | Gm38329 | 1 | 1.61E+08 | 1.61E+08 | - | 427 | TEC | predicted g | - | 25 | 25 | 36 | 4 | 6 | 13 | 3.079261 | 3.112898 | 4.147635 | 0.515088 | 0.751838 | 1.558427 |
| ENSMUSG0 | 555.9977 | 574.1589 | 595.5678 | 730.2507 | 802.8427 | 632.3 | -0.32682 | 0.004779 | 0.049642 | Hgsnat | 8 | 25944453 | 25976753 | - | 3305 | protein_co | heparan-al | - | 558 | 570 | 639 | 701 | 792 | 652 | 8.879675 | 9.169722 | 9.511638 | 11.66262 | 12.82196 | 10.09828 |
| ENSMUSG0 | 196.2931 | 148.0726 | 150.989 | 200.0116 | 252.4089 | 287.0564 | -0.57928 | 0.004785 | 0.049642 | Rnf128 | X | 1.4E+08 | 1.4E+08 | + | 3464 | protein_co | ring finger | - | 197 | 147 | 162 | 192 | 249 | 296 | 2.991043 | 2.256276 | 2.300716 | 3.047704 | 3.846115 | 4.374062 |
| ENSMUSG0 | 565.9618 | 609.4143 | 530.3257 | 687.5398 | 715.6653 | 700.1849 | -0.30291 | 0.004785 | 0.049642 | Tbrg4 | 11 | 6615598 | 6626067 | - | 3696 | protein_co | transformi | - | 568 | 605 | 569 | 660 | 706 | 722 | 8.082593 | 8.703145 | 7.573667 | 9.818868 | 10.22053 | 9.999455 |
| ENSMUSG0 | 1264.446 | 1353.806 | 1265.698 | 1511.546 | 1523.576 | 1459.527 | -0.21075 | 0.004797 | 0.049729 | Zfp260 | 7 | 30094777 | 30107622 | + | 5790 | protein_co | zinc finger | zf-C2H2 | 1269 | 1344 | 1358 | 1451 | 1503 | 1505 | 11.52703 | 12.34166 | 11.53844 | 13.77965 | 13.88933 | 13.30543 |

**Supplemental Table 4.** GO-terms associated with differential gene expression analysis of primary costal chondrocytes derived from *Sox9*$^{del/+}$ and *Sox9*$^{del/del}$ mutant mice.

| Category | GOID | Description | GeneRatio | BgRatio | pvalue | padj | geneID | geneName |
|---|---|---|---|---|---|---|---|---|
| CC | GO:003101 | extracellular matrix | 117\|1232 | 421\|1631 | 5.21E-37 | 3.12E-34 | ENSMUS | Col8a1/Col3a1/Col1a1/Col1a2/Col4a1/Tnxb/F3/Thbs1/Col14a1 |
| CC | GO:006202 | collagen-containing extracellular matrix | 100\|1232 | 320\|1631 | 1.81E-36 | 5.40E-34 | ENSMUS | Col8a1/Col3a1/Col1a1/Col1a2/Col4a1/Tnxb/F3/Thbs1/Col14a1 |
| BP | GO:003019 | extracellular matrix organization | 78\|1209 | 254\|1609 | 3.18E-28 | 1.72E-24 | ENSMUS | Col8a1/Col3a1/Col1a1/Col1a2/Col4a1/Tnxb/Sulf2/Col14a1/Sox |
| BP | GO:004306 | extracellular structure organization | 82\|1209 | 294\|1609 | 1.98E-26 | 5.36E-23 | ENSMUS | Col8a1/Col3a1/Col1a1/Col1a2/Col4a1/Tnxb/Sulf2/Col14a1/Sox |
| BP | GO:000152 | angiogenesis | 104\|1209 | 489\|1609 | 6.24E-23 | 1.12E-19 | ENSMUS | Col8a1/Col4a1/Spry2/F3/Thbs1/Hmox1/Serpine1/Tnmd/Klf4/It |
| MF | GO:000520 | extracellular matrix structural constituent | 49\|1190 | 131\|1591 | 1.88E-22 | 1.71E-19 | ENSMUS | Col8a1/Col3a1/Col1a1/Col1a2/Col4a1/Tnxb/Thbs1/Col14a1/Pc |
| BP | GO:000150 | ossification | 76\|1209 | 357\|1609 | 4.78E-17 | 6.46E-14 | ENSMUS | Col1a1/Col1a2/Igfbp5/Sox9/Dlk1/Xylt1/Fgfr1/Igfbp3/Omd/Aspr |
| BP | GO:006144 | connective tissue development | 62\|1209 | 264\|1609 | 3.01E-16 | 3.26E-13 | ENSMUS | Col1a1/Sulf2/Thbs1/Col14a1/Sox9/Otor/Egr1/Fgfr1/Col12a1/Il |
| BP | GO:003158 | cell-substrate adhesion | 70\|1209 | 332\|1609 | 1.44E-15 | 1.30E-12 | ENSMUS | Col8a1/Col3a1/Col1a1/Tnxb/Thbs1/Postn/Serpine1/Itga5/Plet1 |
| BP | GO:001063 | epithelial cell migration | 59\|1209 | 266\|1609 | 2.45E-14 | 1.90E-11 | ENSMUS | Thbs1/Hmox1/Sox9/Map2k3/Serpine1/Klf4/Serpinf1/Fgfr1/Has |
| BP | GO:009013 | epithelium migration | 59\|1209 | 268\|1609 | 3.49E-14 | 2.36E-11 | ENSMUS | Thbs1/Hmox1/Sox9/Map2k3/Serpine1/Klf4/Serpinf1/Fgfr1/Has |
| MF | GO:000553 | glycosaminoglycan binding | 47\|1190 | 184\|1591 | 3.76E-14 | 1.70E-11 | ENSMUS | Tnxb/Sulf2/Thbs1/Tlr2/Postn/Col5a3/Fgfr1/Pcolce/Lpl/Thbs2/S |
| BP | GO:009013 | tissue migration | 59\|1209 | 270\|1609 | 4.93E-14 | 2.96E-11 | ENSMUS | Thbs1/Hmox1/Sox9/Map2k3/Serpine1/Klf4/Serpinf1/Fgfr1/Has |
| MF | GO:001983 | growth factor binding | 39\|1190 | 136\|1591 | 1.02E-13 | 3.07E-11 | ENSMUS | Col3a1/Col1a1/Col1a2/Col4a1/Thbs1/Vasn/Igfbp5/A2m/Fgfr1/I |
| BP | GO:000150 | skeletal system development | 86\|1209 | 493\|1609 | 1.02E-13 | 5.49E-11 | ENSMUS | Col1a1/Col1a2/Sulf2/Thbs1/Col14a1/Myc/Sox9/Dlk1/Otor/Xylt |
| BP | GO:003019 | collagen fibril organization | 24\|1209 | 54\|16097 | 1.30E-13 | 6.41E-11 | ENSMUS | Col3a1/Col1a1/Col1a2/Tnxb/Col14a1/Col5a3/Col5a2/Adamts2 |
| BP | GO:000166 | ameboidal-type cell migration | 73\|1209 | 387\|1609 | 1.49E-13 | 6.73E-11 | ENSMUS | Thbs1/Hmox1/Sox9/Map2k3/Serpine1/Klf4/Serpinf1/Fgfr1/Has |
| BP | GO:006053 | muscle tissue development | 78\|1209 | 439\|1609 | 5.63E-13 | 2.34E-10 | ENSMUS | Col3a1/Acta1/Hmgcr/Col14a1/Myc/Igfbp5/Rgs2/Sox9/Eln/Cdor |
| MF | GO:000820 | heparin binding | 38\|1190 | 140\|1591 | 1.39E-12 | 3.15E-10 | ENSMUS | Tnxb/Thbs1/Postn/Col5a3/Fgfr1/Pcolce/Lpl/Thbs2/Smoc2/Clec |
| BP | GO:004354 | endothelial cell migration | 45\|1209 | 187\|1609 | 1.50E-12 | 5.65E-10 | ENSMUS | Thbs1/Hmox1/Map2k3/Klf4/Serpinf1/Fgfr1/Met/Efna1/Smoc2/ |
| BP | GO:190134 | regulation of vasculature development | 63\|1209 | 323\|1609 | 1.57E-12 | 5.65E-10 | ENSMUS | Spry2/F3/Thbs1/Hmox1/Serpine1/Tnmd/Klf4/Itga5/Egr1/Serpin |
| BP | GO:004353 | blood vessel endothelial cell migration | 33\|1209 | 109\|1609 | 1.72E-12 | 5.80E-10 | ENSMUS | Thbs1/Hmox1/Map2k3/Klf4/Fgfr1/Efna1/Spred1/Meox2/Nr4a1/ |
| CC | GO:004442 | extracellular matrix component | 22\|1232 | 50\|16313 | 1.97E-12 | 3.93E-10 | ENSMUS | Col3a1/Col1a1/Col1a2/Col4a1/Eln/Mfap4/Col5a3/Col5a2/Laml |
| BP | GO:005127 | negative regulation of cellular component movement | 60\|1209 | 302\|1609 | 2.38E-12 | 7.56E-10 | ENSMUS | Col3a1/Thbs1/Igfbp5/Serpine1/Trib1/Klf4/Serpinf1/Igfbp3/Dus |
| MF | GO:003002 | extracellular matrix structural constituent conferring tensile st | 19\|1190 | 38\|15912 | 3.13E-12 | 5.66E-10 | ENSMUS | Col8a1/Col3a1/Col1a1/Col1a2/Col4a1/Col14a1/Col5a3/Col5a2 |
| BP | GO:005121 | cartilage development | 46\|1209 | 198\|1609 | 3.22E-12 | 9.68E-10 | ENSMUS | Col1a1/Sulf2/Thbs1/Col14a1/Sox9/Otor/Fgfr1/Col12a1/Eif2ak3 |
| BP | GO:004576 | regulation of angiogenesis | 58\|1209 | 291\|1609 | 4.84E-12 | 1.32E-09 | ENSMUS | Spry2/F3/Thbs1/Hmox1/Serpine1/Tnmd/Klf4/Itga5/Serpinf1/Th |
| BP | GO:000193 | negative regulation of protein phosphorylation | 70\|1209 | 389\|1609 | 4.89E-12 | 1.32E-09 | ENSMUS | Errfi1/Spry2/Hmgcr/Rgs2/Trib1/Klf4/Gadd45a/Igfbp3/Dusp1/M |
| BP | GO:004232 | negative regulation of phosphorylation | 75\|1209 | 432\|1609 | 5.20E-12 | 1.34E-09 | ENSMUS | Errfi1/Spry2/Hmgcr/Rgs2/Trib1/Klf4/Gadd45a/Igfbp3/Dusp1/M |
| BP | GO:004001 | negative regulation of locomotion | 61\|1209 | 316\|1609 | 5.66E-12 | 1.39E-09 | ENSMUS | Col3a1/Thbs1/Igfbp5/Serpine1/Trib1/Klf4/Serpinf1/Igfbp3/Dus |
| BP | GO:003027 | regulation of ossification | 48\|1209 | 216\|1609 | 6.08E-12 | 1.43E-09 | ENSMUS | Igfbp5/Sox9/Dlk1/Fgfr1/Omd/Smoc1/Noct/Cyr61/Lmna/Ank/Sn |
| BP | GO:003033 | negative regulation of cell migration | 53\|1209 | 257\|1609 | 1.04E-11 | 2.35E-09 | ENSMUS | Col3a1/Thbs1/Igfbp5/Serpine1/Trib1/Klf4/Serpinf1/Igfbp3/Dus |
| BP | GO:006034 | bone development | 47\|1209 | 214\|1609 | 1.57E-11 | 3.39E-09 | ENSMUS | Col1a1/Sulf2/Thbs1/Col14a1/Sox9/Xylt1/Chad/Has2/Col12a1/ |
| BP | GO:001470 | striated muscle tissue development | 72\|1209 | 417\|1609 | 1.83E-11 | 3.80E-09 | ENSMUS | Acta1/Hmgcr/Col14a1/Myc/Rgs2/Eln/Cdon/Hivep3/Egr1/Ifrd1/ |
| BP | GO:200014 | negative regulation of cell motility | 54\|1209 | 269\|1609 | 1.99E-11 | 3.86E-09 | ENSMUS | Col3a1/Thbs1/Igfbp5/Serpine1/Trib1/Klf4/Serpinf1/Igfbp3/Dus |
| BP | GO:001081 | regulation of cell-substrate adhesion | 46\|1209 | 208\|1609 | 2.00E-11 | 3.86E-09 | ENSMUS | Col8a1/Col1a1/Thbs1/Postn/Serpine1/Itga5/Plet1/Nid1/Has2/S |
| BP | GO:006034 | bone morphogenesis | 31\|1209 | 106\|1609 | 2.20E-11 | 4.10E-09 | ENSMUS | Col1a1/Thbs1/Col14a1/Sox9/Has2/Col12a1/Mmp14/Fgfr2/Insi |
| BP | GO:000961 | response to wounding | 75\|1209 | 447\|1609 | 2.85E-11 | 5.13E-09 | ENSMUS | Col3a1/Col1a1/Sulf2/F3/Thbs1/Hmox1/Hmgcr/Serpine1/Klf4/P |
| CC | GO:000560 | basement membrane | 30\|1232 | 102\|1631 | 4.54E-11 | 6.78E-09 | ENSMUS | Col8a1/Col4a1/Hmcn2/Nid1/Serpinf1/Lamb1/Thbs2/Col4a2/La |
| BP | GO:004353 | regulation of blood vessel endothelial cell migration | 27\|1209 | 85\|16097 | 5.21E-11 | 9.09E-09 | ENSMUS | Thbs1/Hmox1/Map2k3/Klf4/Fgfr1/Efna1/Spred1/Meox2/Nrp1/ |
| BP | GO:006035 | endochondral bone morphogenesis | 24\|1209 | 69\|16097 | 7.15E-11 | 1.21E-08 | ENSMUS | Col1a1/Thbs1/Col14a1/Sox9/Col12a1/Mmp14/Nab2/Bmp6/Co |
| MF | GO:190168 | sulfur compound binding | 47\|1190 | 226\|1591 | 9.89E-11 | 1.49E-08 | ENSMUS | Tnxb/Thbs1/Postn/Col5a3/Fgfr1/Pcolce/Lpl/Thbs2/Smoc2/Clec |
| BP | GO:000206 | chondrocyte differentiation | 31\|1209 | 112\|1609 | 1.04E-10 | 1.71E-08 | ENSMUS | Sulf2/Col14a1/Sox9/Fgfr1/Col12a1/Eif2ak3/Bmp6/Col6a2/Rflna |
| BP | GO:000164 | osteoblast differentiation | 41\|1209 | 181\|1609 | 1.13E-10 | 1.80E-08 | ENSMUS | Col1a1/Igfbp5/Dlk1/Fgfr1/Igfbp3/Smoc1/Noct/Cyr61/Lmna/Sm |
| BP | GO:000751 | muscle organ development | 67\|1209 | 391\|1609 | 1.31E-10 | 2.02E-08 | ENSMUS | Col3a1/Acta1/Hmgcr/Col14a1/Myc/Rgs2/Eln/Cdon/Hivep3/Egr |
| BP | GO:003121 | biomineral tissue development | 36\|1209 | 149\|1609 | 2.28E-10 | 3.43E-08 | ENSMUS | Col1a1/Col1a2/Sox9/Dlk1/Slc20a1/Omd/Atf4/Aspn/Clec3b/Eif2 |
| BP | GO:009028 | regulation of cellular response to growth factor stimulus | 50\|1209 | 256\|1609 | 3.06E-10 | 4.47E-08 | ENSMUS | Spry2/Sulf2/Thbs1/Vasn/Tnmd/Fgfr1/Cd109/Cdh5/Aspn/Smoc2 |
| BP | GO:004870 | skeletal system morphogenesis | 48\|1209 | 241\|1609 | 3.47E-10 | 4.94E-08 | ENSMUS | Col1a1/Thbs1/Col14a1/Myc/Sox9/Otor/Chad/Has2/Col12a1/M |
| BP | GO:004873 | gland development | 67\|1209 | 402\|1609 | 4.45E-10 | 6.17E-08 | ENSMUS | Sulf2/Hmox1/Igfbp5/Sox9/Serpinf1/Fgfr1/Fem1b/Met/Cdkn1c/ |
| BP | GO:001063 | regulation of epithelial cell migration | 44\|1209 | 212\|1609 | 4.82E-10 | 6.51E-08 | ENSMUS | Thbs1/Hmox1/Sox9/Map2k3/Serpine1/Klf4/Serpinf1/Fgfr1/Has |
| BP | GO:001081 | positive regulation of cell-substrate adhesion | 32\|1209 | 126\|1609 | 5.87E-10 | 7.74E-08 | ENSMUS | Col8a1/Thbs1/Itga5/Nid1/Has2/S100a10/Smoc2/Iqgap1/Smoc1 |

| Count | Up | Up_Gene_id | Down | Down_Gene_id |
|---|---|---|---|---|
| 117 | 75 | ENSMUSG0 | 42 | ENSMUSG00000068196/ENSMUSG00000028128/ENSMUSG00000040152/ENSMUSG00000039646/ENSMUSG00000029675/ENSMUSG00000037411/ENSMUSG0 |
| 100 | 67 | ENSMUSG0 | 33 | ENSMUSG00000068196/ENSMUSG00000028128/ENSMUSG00000040152/ENSMUSG00000029675/ENSMUSG00000037411/ENSMUSG00000000753/ENSMUSG0 |
| 78 | 44 | ENSMUSG0 | 34 | ENSMUSG00000068196/ENSMUSG00000006800/ENSMUSG00000000567/ENSMUSG00000029675/ENSMUSG00000022367/ENSMUSG00000050335/ENSMUSG0 |
| 82 | 47 | ENSMUSG0 | 35 | ENSMUSG00000068196/ENSMUSG00000006800/ENSMUSG00000000567/ENSMUSG00000029675/ENSMUSG00000022367/ENSMUSG00000050335/ENSMUSG0 |
| 104 | 46 | ENSMUSG0 | 58 | ENSMUSG00000068196/ENSMUSG00000022114/ENSMUSG00000028128/ENSMUSG00000040152/ENSMUSG00000005413/ENSMUSG00000037411/ENSMUSG0 |
| 49 | 40 | ENSMUSG0 | 9 | ENSMUSG00000068196/ENSMUSG00000040152/ENSMUSG00000029675/ENSMUSG00000020241/ENSMUSG00000031849/ENSMUSG00000048126/ENSMUSG0 |
| 76 | 30 | ENSMUSG0 | 46 | ENSMUSG00000000567/ENSMUSG00000040856/ENSMUSG00000030657/ENSMUSG00000031565/ENSMUSG00000021136/ENSMUSG00000023087/ENSMUSG0 |
| 62 | 24 | ENSMUSG0 | 38 | ENSMUSG00000006800/ENSMUSG00000040152/ENSMUSG00000000567/ENSMUSG00000027416/ENSMUSG00000038418/ENSMUSG00000031565/ENSMUSG0 |
| 70 | 40 | ENSMUSG0 | 30 | ENSMUSG00000068196/ENSMUSG00000040152/ENSMUSG00000037411/ENSMUSG00000000555/ENSMUSG00000032068/ENSMUSG00000022367/ENSMUSG0 |
| 59 | 23 | ENSMUSG0 | 36 | ENSMUSG00000040152/ENSMUSG00000005413/ENSMUSG00000000567/ENSMUSG00000018932/ENSMUSG00000037411/ENSMUSG00000003032/ENSMUSG0 |
| 59 | 23 | ENSMUSG0 | 36 | ENSMUSG00000040152/ENSMUSG00000005413/ENSMUSG00000000567/ENSMUSG00000018932/ENSMUSG00000037411/ENSMUSG00000003032/ENSMUSG0 |
| 47 | 27 | ENSMUSG0 | 20 | ENSMUSG00000006800/ENSMUSG00000040152/ENSMUSG00000027995/ENSMUSG00000031565/ENSMUSG00000031825/ENSMUSG00000030849/ENSMUSG0 |
| 59 | 23 | ENSMUSG0 | 36 | ENSMUSG00000040152/ENSMUSG00000005413/ENSMUSG00000000567/ENSMUSG00000018932/ENSMUSG00000037411/ENSMUSG00000003032/ENSMUSG0 |
| 39 | 20 | ENSMUSG0 | 19 | ENSMUSG00000040152/ENSMUSG00000039646/ENSMUSG00000030111/ENSMUSG00000031565/ENSMUSG00000024359/ENSMUSG00000046186/ENSMUSG0 |
| 86 | 31 | ENSMUSG0 | 55 | ENSMUSG00000006800/ENSMUSG00000040152/ENSMUSG00000022346/ENSMUSG00000000567/ENSMUSG00000040856/ENSMUSG00000027416/ENSMUSG0 |
| 24 | 14 | ENSMUSG0 | 10 | ENSMUSG00000028641/ENSMUSG00000006931/ENSMUSG00000027996/ENSMUSG00000031849/ENSMUSG00000007613/ENSMUSG00000004846/ENSMUSG0 |
| 73 | 31 | ENSMUSG0 | 42 | ENSMUSG00000040152/ENSMUSG00000005413/ENSMUSG00000000567/ENSMUSG00000018932/ENSMUSG00000037411/ENSMUSG00000003032/ENSMUSG0 |
| 78 | 33 | ENSMUSG0 | 45 | ENSMUSG00000021670/ENSMUSG00000022346/ENSMUSG00000026360/ENSMUSG00000000567/ENSMUSG00000029675/ENSMUSG00000038119/ENSMUSG0 |
| 38 | 23 | ENSMUSG0 | 15 | ENSMUSG00000040152/ENSMUSG00000031565/ENSMUSG00000031825/ENSMUSG00000030849/ENSMUSG00000031785/ENSMUSG00000030513/ENSMUSG0 |
| 45 | 20 | ENSMUSG0 | 25 | ENSMUSG00000040152/ENSMUSG00000005413/ENSMUSG00000018932/ENSMUSG00000003032/ENSMUSG00000000753/ENSMUSG00000031565/ENSMUSG0 |
| 63 | 27 | ENSMUSG0 | 36 | ENSMUSG00000022114/ENSMUSG00000028128/ENSMUSG00000040152/ENSMUSG00000005413/ENSMUSG00000037411/ENSMUSG00000003032/ENSMUSG0 |
| 33 | 13 | ENSMUSG0 | 20 | ENSMUSG00000040152/ENSMUSG00000005413/ENSMUSG00000018932/ENSMUSG00000003032/ENSMUSG00000031565/ENSMUSG00000027954/ENSMUSG0 |
| 22 | 20 | ENSMUSG0 | 2 | ENSMUSG00000029675/ENSMUSG00000068479 |
| 60 | 24 | ENSMUSG0 | 36 | ENSMUSG00000040152/ENSMUSG00000037411/ENSMUSG00000032501/ENSMUSG00000003032/ENSMUSG00000000753/ENSMUSG00000027351/ENSMUSG0 |
| 19 | 14 | ENSMUSG0 | 5 | ENSMUSG00000068196/ENSMUSG00000020241/ENSMUSG00000048126/ENSMUSG00000039462/ENSMUSG00000001119 |
| 46 | 18 | ENSMUSG0 | 28 | ENSMUSG00000006800/ENSMUSG00000040152/ENSMUSG00000000567/ENSMUSG00000027416/ENSMUSG00000031565/ENSMUSG00000031668/ENSMUSG0 |
| 58 | 26 | ENSMUSG0 | 32 | ENSMUSG00000022114/ENSMUSG00000028128/ENSMUSG00000040152/ENSMUSG00000005413/ENSMUSG00000037411/ENSMUSG00000003032/ENSMUSG0 |
| 70 | 19 | ENSMUSG0 | 51 | ENSMUSG00000028967/ENSMUSG00000022114/ENSMUSG00000021670/ENSMUSG00000026360/ENSMUSG00000032501/ENSMUSG00000003032/ENSMUSG0 |
| 75 | 20 | ENSMUSG0 | 55 | ENSMUSG00000028967/ENSMUSG00000022114/ENSMUSG00000021670/ENSMUSG00000026360/ENSMUSG00000032501/ENSMUSG00000003032/ENSMUSG0 |
| 61 | 25 | ENSMUSG0 | 36 | ENSMUSG00000040152/ENSMUSG00000037411/ENSMUSG00000032501/ENSMUSG00000003032/ENSMUSG00000000753/ENSMUSG00000027351/ENSMUSG0 |
| 48 | 19 | ENSMUSG0 | 29 | ENSMUSG00000000567/ENSMUSG00000040856/ENSMUSG00000031565/ENSMUSG00000021136/ENSMUSG00000023087/ENSMUSG00000028063/ENSMUSG0 |
| 53 | 20 | ENSMUSG0 | 33 | ENSMUSG00000040152/ENSMUSG00000037411/ENSMUSG00000032501/ENSMUSG00000003032/ENSMUSG00000000753/ENSMUSG00000027351/ENSMUSG0 |
| 47 | 16 | ENSMUSG0 | 31 | ENSMUSG00000006800/ENSMUSG00000040152/ENSMUSG00000000567/ENSMUSG00000030657/ENSMUSG00000039084/ENSMUSG00000022367/ENSMUSG0 |
| 72 | 29 | ENSMUSG0 | 43 | ENSMUSG00000021670/ENSMUSG00000022346/ENSMUSG00000026360/ENSMUSG00000029675/ENSMUSG00000038119/ENSMUSG00000028634/ENSMUSG0 |
| 54 | 20 | ENSMUSG0 | 34 | ENSMUSG00000040152/ENSMUSG00000037411/ENSMUSG00000032501/ENSMUSG00000003032/ENSMUSG00000000753/ENSMUSG00000027351/ENSMUSG0 |
| 46 | 25 | ENSMUSG0 | 21 | ENSMUSG00000068196/ENSMUSG00000040152/ENSMUSG00000037411/ENSMUSG00000000555/ENSMUSG00000032068/ENSMUSG00000022367/ENSMUSG0 |
| 31 | 11 | ENSMUSG0 | 20 | ENSMUSG00000040152/ENSMUSG00000000567/ENSMUSG00000022367/ENSMUSG00000030849/ENSMUSG00000045294/ENSMUSG00000025402/ENSMUSG0 |
| 75 | 31 | ENSMUSG0 | 44 | ENSMUSG00000006800/ENSMUSG00000028128/ENSMUSG00000040152/ENSMUSG00000005413/ENSMUSG00000021670/ENSMUSG00000037411/ENSMUSG0 |
| 30 | 19 | ENSMUSG0 | 11 | ENSMUSG00000068196/ENSMUSG00000000753/ENSMUSG00000021136/ENSMUSG00000027447/ENSMUSG00000001131/ENSMUSG00000028641/ENSMUSG0 |
| 27 | 12 | ENSMUSG0 | 15 | ENSMUSG00000040152/ENSMUSG00000005413/ENSMUSG00000018932/ENSMUSG00000003032/ENSMUSG00000031565/ENSMUSG00000027954/ENSMUSG0 |
| 24 | 10 | ENSMUSG0 | 14 | ENSMUSG00000040152/ENSMUSG00000000567/ENSMUSG00000025402/ENSMUSG00000039004/ENSMUSG00000020241/ENSMUSG00000001419/ENSMUSG0 |
| 47 | 28 | ENSMUSG0 | 19 | ENSMUSG00000040152/ENSMUSG00000031565/ENSMUSG00000031825/ENSMUSG00000030849/ENSMUSG00000031785/ENSMUSG00000030513/ENSMUSG0 |
| 31 | 12 | ENSMUSG0 | 19 | ENSMUSG00000006800/ENSMUSG00000000567/ENSMUSG00000031565/ENSMUSG00000031668/ENSMUSG00000039004/ENSMUSG00000020241/ENSMUSG0 |
| 41 | 14 | ENSMUSG0 | 27 | ENSMUSG00000040856/ENSMUSG00000031565/ENSMUSG00000021136/ENSMUSG00000023087/ENSMUSG00000028063/ENSMUSG00000021540/ENSMUSG0 |
| 67 | 26 | ENSMUSG0 | 41 | ENSMUSG00000021670/ENSMUSG00000022346/ENSMUSG00000026360/ENSMUSG00000029675/ENSMUSG00000038119/ENSMUSG00000028634/ENSMUSG0 |
| 36 | 17 | ENSMUSG0 | 19 | ENSMUSG00000000567/ENSMUSG00000040856/ENSMUSG00000027397/ENSMUSG00000042406/ENSMUSG00000031668/ENSMUSG00000022265/ENSMUSG0 |
| 50 | 23 | ENSMUSG0 | 27 | ENSMUSG00000022114/ENSMUSG00000006800/ENSMUSG00000040152/ENSMUSG00000039646/ENSMUSG00000031565/ENSMUSG00000046186/ENSMUSG0 |
| 48 | 14 | ENSMUSG0 | 34 | ENSMUSG00000040152/ENSMUSG00000022346/ENSMUSG00000000567/ENSMUSG00000027416/ENSMUSG00000039084/ENSMUSG00000022367/ENSMUSG0 |
| 67 | 20 | ENSMUSG0 | 47 | ENSMUSG00000006800/ENSMUSG00000005413/ENSMUSG00000000567/ENSMUSG00000000753/ENSMUSG00000031565/ENSMUSG00000032244/ENSMUSG0 |
| 44 | 17 | ENSMUSG0 | 27 | ENSMUSG00000040152/ENSMUSG00000005413/ENSMUSG00000000567/ENSMUSG00000018932/ENSMUSG00000037411/ENSMUSG00000003032/ENSMUSG0 |
| 32 | 19 | ENSMUSG0 | 13 | ENSMUSG00000068196/ENSMUSG00000040152/ENSMUSG00000000555/ENSMUSG00000022367/ENSMUSG00000041959/ENSMUSG00000030536/ENSMUSG0 |

0000039084/ENSMUSG00000000753/ENSMUSG00000041959/ENSMUSG00000050335/ENSMUSG00000021136/ENSMUSG00000027447/ENSMUSG00000031825/ENSMUSG00000030849/ENSMUSG0000000
0000041959/ENSMUSG00000050335/ENSMUSG00000021136/ENSMUSG00000027447/ENSMUSG00000030849/ENSMUSG00000001025/ENSMUSG00000030513/ENSMUSG00000020241/ENSMUSG0000000
0000021136/ENSMUSG00000031825/ENSMUSG00000048537/ENSMUSG00000026596/ENSMUSG00000028641/ENSMUSG00000006931/ENSMUSG00000019997/ENSMUSG00000027996/ENSMUSG0000003
0000021136/ENSMUSG00000031825/ENSMUSG00000032883/ENSMUSG00000048537/ENSMUSG00000026596/ENSMUSG00000028641/ENSMUSG00000006931/ENSMUSG00000019997/ENSMUSG0000002
0000003032/ENSMUSG00000000555/ENSMUSG00000000753/ENSMUSG00000031565/ENSMUSG00000023905/ENSMUSG00000024754/ENSMUSG00000050335/ENSMUSG00000027954/ENSMUSG0000002
0000039462/ENSMUSG00000026147/ENSMUSG00000001119
0000031668/ENSMUSG00000028063/ENSMUSG00000022265/ENSMUSG00000021540/ENSMUSG00000030849/ENSMUSG00000025402/ENSMUSG00000039004/ENSMUSG00000039286/ENSMUSG0000002
0000027947/ENSMUSG00000031668/ENSMUSG00000028063/ENSMUSG00000021540/ENSMUSG00000056758/ENSMUSG00000039004/ENSMUSG00000020241/ENSMUSG00000001419/ENSMUSG0000001
0000041959/ENSMUSG00000023905/ENSMUSG00000027954/ENSMUSG00000030536/ENSMUSG00000021136/ENSMUSG00000039286/ENSMUSG00000002489/ENSMUSG00000022508/ENSMUSG0000001
0000000753/ENSMUSG00000031565/ENSMUSG00000022367/ENSMUSG00000009376/ENSMUSG00000027954/ENSMUSG00000020458/ENSMUSG00000027351/ENSMUSG00000029287/ENSMUSG0000002
0000000753/ENSMUSG00000031565/ENSMUSG00000022367/ENSMUSG00000009376/ENSMUSG00000027954/ENSMUSG00000020458/ENSMUSG00000027351/ENSMUSG00000029287/ENSMUSG0000002
0000031785/ENSMUSG00000030513/ENSMUSG00000029287/ENSMUSG00000019997/ENSMUSG00000037012/ENSMUSG00000031849/ENSMUSG00000005087/ENSMUSG00000023951/ENSMUSG0000002
0000000753/ENSMUSG00000031565/ENSMUSG00000022367/ENSMUSG00000009376/ENSMUSG00000027954/ENSMUSG00000020458/ENSMUSG00000027351/ENSMUSG00000029287/ENSMUSG0000002
0000027947/ENSMUSG00000030849/ENSMUSG00000030513/ENSMUSG00000029287/ENSMUSG00000019997/ENSMUSG00000021756/ENSMUSG00000007613/ENSMUSG00000005533/ENSMUSG0000003
0000030657/ENSMUSG00000039084/ENSMUSG00000031565/ENSMUSG00000022367/ENSMUSG00000050335/ENSMUSG00000031668/ENSMUSG00000021540/ENSMUSG00000030849/ENSMUSG0000004
0000056050/ENSMUSG00000034205/ENSMUSG00000025185/ENSMUSG00000074934
0000000753/ENSMUSG00000031565/ENSMUSG00000022367/ENSMUSG00000009376/ENSMUSG00000027954/ENSMUSG00000020458/ENSMUSG00000027351/ENSMUSG00000029287/ENSMUSG0000000
0000028634/ENSMUSG00000038418/ENSMUSG00000001627/ENSMUSG00000031565/ENSMUSG00000009376/ENSMUSG00000022951/ENSMUSG00000019817/ENSMUSG00000028063/ENSMUSG0000003
0000029287/ENSMUSG00000019997/ENSMUSG00000031849/ENSMUSG00000023951/ENSMUSG00000024486/ENSMUSG00000029816/ENSMUSG00000028047/ENSMUSG00000031548/ENSMUSG0000002
0000009376/ENSMUSG00000027954/ENSMUSG00000027351/ENSMUSG00000023034/ENSMUSG00000007613/ENSMUSG00000023951/ENSMUSG00000023068/ENSMUSG00000013076/ENSMUSG0000002
0000000555/ENSMUSG00000038418/ENSMUSG00000000753/ENSMUSG00000023905/ENSMUSG00000027947/ENSMUSG00000050335/ENSMUSG00000027954/ENSMUSG00000020458/ENSMUSG0000002
0000027351/ENSMUSG00000023034/ENSMUSG00000023951/ENSMUSG00000023068/ENSMUSG00000013076/ENSMUSG00000022443/ENSMUSG00000006445/ENSMUSG00000050965/ENSMUSG0000004

0000031785/ENSMUSG00000025373/ENSMUSG00000031778/ENSMUSG00000029287/ENSMUSG00000001131/ENSMUSG00000004040/ENSMUSG00000037242/ENSMUSG00000027996/ENSMUSG0000003

0000021540/ENSMUSG00000039004/ENSMUSG00000020241/ENSMUSG00000001419/ENSMUSG00000019997/ENSMUSG00000027996/ENSMUSG00000036356/ENSMUSG00000031849/ENSMUSG0000004
0000000555/ENSMUSG00000000753/ENSMUSG00000023905/ENSMUSG00000050335/ENSMUSG00000027954/ENSMUSG00000020458/ENSMUSG00000027351/ENSMUSG00000068739/ENSMUSG0000005
0000036390/ENSMUSG00000009376/ENSMUSG00000046186/ENSMUSG00000046062/ENSMUSG00000027954/ENSMUSG00000023067/ENSMUSG00000027351/ENSMUSG00000078429/ENSMUSG0000003
0000036390/ENSMUSG00000009376/ENSMUSG00000046186/ENSMUSG00000046062/ENSMUSG00000027954/ENSMUSG00000023067/ENSMUSG00000027351/ENSMUSG00000003541/ENSMUSG0000007
0000031785/ENSMUSG00000025373/ENSMUSG00000031778/ENSMUSG00000029287/ENSMUSG00000001131/ENSMUSG00000004040/ENSMUSG00000037242/ENSMUSG00000074794/ENSMUSG0000002
0000022265/ENSMUSG00000021540/ENSMUSG00000039004/ENSMUSG00000039286/ENSMUSG00000028641/ENSMUSG00000021756/ENSMUSG00000029050/ENSMUSG00000027996/ENSMUSG0000003
0000031785/ENSMUSG00000025373/ENSMUSG00000031778/ENSMUSG00000029287/ENSMUSG00000001131/ENSMUSG00000004040/ENSMUSG00000037242/ENSMUSG00000027996/ENSMUSG0000003
0000021540/ENSMUSG00000030849/ENSMUSG00000045294/ENSMUSG00000025402/ENSMUSG00000039004/ENSMUSG00000020241/ENSMUSG00000028641/ENSMUSG00000001419/ENSMUSG0000000
0000038418/ENSMUSG00000001627/ENSMUSG00000031565/ENSMUSG00000009376/ENSMUSG00000022951/ENSMUSG00000019817/ENSMUSG00000028063/ENSMUSG00000030849/ENSMUSG0000001
0000031785/ENSMUSG00000025373/ENSMUSG00000031778/ENSMUSG00000029287/ENSMUSG00000001131/ENSMUSG00000004040/ENSMUSG00000037242/ENSMUSG00000027996/ENSMUSG0000003
0000041959/ENSMUSG00000030536/ENSMUSG00000021136/ENSMUSG00000022508/ENSMUSG00000024789/ENSMUSG00000031523/ENSMUSG00000022425/ENSMUSG00000023951/ENSMUSG0000005
0000039004/ENSMUSG00000020241/ENSMUSG00000001419/ENSMUSG00000002881/ENSMUSG00000029050/ENSMUSG00000027996/ENSMUSG00000036356/ENSMUSG00000031849/ENSMUSG0000004
0000003032/ENSMUSG00000022565/ENSMUSG00000032068/ENSMUSG00000030657/ENSMUSG00000001627/ENSMUSG00000023905/ENSMUSG00000046186/ENSMUSG00000020458/ENSMUSG0000002
0000038168/ENSMUSG00000023951/ENSMUSG00000020044/ENSMUSG00000039952/ENSMUSG00000034205
0000027351/ENSMUSG00000023951/ENSMUSG00000023068/ENSMUSG00000013076/ENSMUSG00000006445/ENSMUSG00000050965/ENSMUSG00000032487/ENSMUSG00000015839/ENSMUSG0000003
0000002881/ENSMUSG00000036356/ENSMUSG00000031849/ENSMUSG00000048126/ENSMUSG00000034135/ENSMUSG00000026147/ENSMUSG00000028047/ENSMUSG00000001119
0000029287/ENSMUSG00000019997/ENSMUSG00000031849/ENSMUSG00000030096/ENSMUSG00000023951/ENSMUSG00000024486/ENSMUSG00000029816/ENSMUSG00000028047/ENSMUSG0000003
0000001419/ENSMUSG00000019997/ENSMUSG00000027996/ENSMUSG00000031849/ENSMUSG00000007613/ENSMUSG00000048126/ENSMUSG00000037447/ENSMUSG00000024927/ENSMUSG0000005
0000039004/ENSMUSG00000039286/ENSMUSG00000029287/ENSMUSG00000001419/ENSMUSG00000021756/ENSMUSG00000029050/ENSMUSG00000027996/ENSMUSG00000048616/ENSMUSG0000005
0000038418/ENSMUSG00000031565/ENSMUSG00000009376/ENSMUSG00000022951/ENSMUSG00000019817/ENSMUSG00000028063/ENSMUSG00000030849/ENSMUSG00000015846/ENSMUSG0000002
0000030849/ENSMUSG00000039004/ENSMUSG00000037465/ENSMUSG00000031451/ENSMUSG00000031849/ENSMUSG00000056501/ENSMUSG00000039662/ENSMUSG00000035673/ENSMUSG0000005
0000029287/ENSMUSG00000020484/ENSMUSG00000032340/ENSMUSG00000029050/ENSMUSG00000027996/ENSMUSG00000038400/ENSMUSG00000048616/ENSMUSG00000030530/ENSMUSG0000002
0000030849/ENSMUSG00000045294/ENSMUSG00000025402/ENSMUSG00000039004/ENSMUSG00000020241/ENSMUSG00000001419/ENSMUSG00000019997/ENSMUSG00000002881/ENSMUSG0000002
0000009376/ENSMUSG00000037071/ENSMUSG00000029482/ENSMUSG00000020458/ENSMUSG00000027678/ENSMUSG00000031668/ENSMUSG00000030849/ENSMUSG00000015846/ENSMUSG0000005
0000000753/ENSMUSG00000031565/ENSMUSG00000022367/ENSMUSG00000009376/ENSMUSG00000027954/ENSMUSG00000020458/ENSMUSG00000027351/ENSMUSG00000029287/ENSMUSG0000002
0000021136/ENSMUSG00000024789/ENSMUSG00000022425/ENSMUSG00000023951/ENSMUSG00000003814/ENSMUSG00000039952/ENSMUSG00000026812

01025/ENSMUSG00000030513/ENSMUSG00000020241/ENSMUSG00000029287/ENSMUSG00000001131/ENSMUSG00000028641/ENSMUSG00000019997/ENSMUSG00000038168/ENSMUSG00000031849/E
01131/ENSMUSG00000028641/ENSMUSG00000038168/ENSMUSG00000031849/ENSMUSG00000048126/ENSMUSG00000004846/ENSMUSG00000023951/ENSMUSG00000036606/ENSMUSG00000037820/E
36356/ENSMUSG00000031849/ENSMUSG00000030341/ENSMUSG00000007613/ENSMUSG00000004846/ENSMUSG00000020682/ENSMUSG00000003665/ENSMUSG00000027832/ENSMUSG00000039462/E
27996/ENSMUSG00000036356/ENSMUSG00000031849/ENSMUSG00000030341/ENSMUSG00000007613/ENSMUSG00000004846/ENSMUSG00000020682/ENSMUSG00000003665/ENSMUSG00000027832/E
20458/ENSMUSG00000027351/ENSMUSG00000031668/ENSMUSG00000021540/ENSMUSG00000030849/ENSMUSG00000031785/ENSMUSG00000068739/ENSMUSG00000015846/ENSMUSG00000056758/E

29287/ENSMUSG00000028641/ENSMUSG00000001419/ENSMUSG00000019997/ENSMUSG00000037465/ENSMUSG00000021756/ENSMUSG00000002881/ENSMUSG00000029050/ENSMUSG00000027996/E
19997/ENSMUSG00000020484/ENSMUSG00000074794/ENSMUSG00000027996/ENSMUSG00000036356/ENSMUSG00000031849/ENSMUSG00000048616/ENSMUSG00000007613/ENSMUSG00000048126/E
19997/ENSMUSG00000024789/ENSMUSG00000031451/ENSMUSG00000031523/ENSMUSG00000022425/ENSMUSG00000023951/ENSMUSG00000022817/ENSMUSG00000054808/ENSMUSG00000032702/E
23034/ENSMUSG00000022425/ENSMUSG00000007613/ENSMUSG00000023951/ENSMUSG00000039384/ENSMUSG00000023068/ENSMUSG00000053797/ENSMUSG00000013076/ENSMUSG00000022443/E
23034/ENSMUSG00000022425/ENSMUSG00000007613/ENSMUSG00000023951/ENSMUSG00000039384/ENSMUSG00000023068/ENSMUSG00000053797/ENSMUSG00000013076/ENSMUSG00000022443/E
24486/ENSMUSG00000079037/ENSMUSG00000029816/ENSMUSG00000028047/ENSMUSG00000031548/ENSMUSG00000027712
23034/ENSMUSG00000022425/ENSMUSG00000007613/ENSMUSG00000023951/ENSMUSG00000039384/ENSMUSG00000023068/ENSMUSG00000053797/ENSMUSG00000013076/ENSMUSG00000022443/E
30530/ENSMUSG00000024098/ENSMUSG00000023951/ENSMUSG00000026836/ENSMUSG00000001119
45294/ENSMUSG00000025402/ENSMUSG00000039004/ENSMUSG00000026675/ENSMUSG00000020241/ENSMUSG00000028641/ENSMUSG00000001419/ENSMUSG00000019997/ENSMUSG00000002881/E

01131/ENSMUSG00000023034/ENSMUSG00000055322/ENSMUSG00000022425/ENSMUSG00000007613/ENSMUSG00000023951/ENSMUSG00000039384/ENSMUSG00000023068/ENSMUSG00000003665/E
30849/ENSMUSG00000015846/ENSMUSG00000050567/ENSMUSG00000029287/ENSMUSG00000001419/ENSMUSG00000030557/ENSMUSG00000020423/ENSMUSG00000023034/ENSMUSG00000029050/E
27712
22443/ENSMUSG00000006445/ENSMUSG00000050965/ENSMUSG00000048661/ENSMUSG00000003814/ENSMUSG00000056050/ENSMUSG00000032487/ENSMUSG00000034205/ENSMUSG00000015839/E
27351/ENSMUSG00000068739/ENSMUSG00000056758/ENSMUSG00000031778/ENSMUSG00000004040/ENSMUSG00000031714/ENSMUSG00000020484/ENSMUSG00000027996/ENSMUSG00000022425/E
48661/ENSMUSG00000056050/ENSMUSG00000032487/ENSMUSG00000015839/ENSMUSG00000031393/ENSMUSG00000074934

31523/ENSMUSG00000048616/ENSMUSG00000025340/ENSMUSG00000039384/ENSMUSG00000020682/ENSMUSG00000003665/ENSMUSG00000037815/ENSMUSG00000053797/ENSMUSG00000054808/E

48616/ENSMUSG00000007613/ENSMUSG00000048126/ENSMUSG00000037447/ENSMUSG00000024927/ENSMUSG00000034135/ENSMUSG00000050965/ENSMUSG00000019936/ENSMUSG00000056050/E
56758/ENSMUSG00000031778/ENSMUSG00000004040/ENSMUSG00000031714/ENSMUSG00000020484/ENSMUSG00000027996/ENSMUSG00000022425/ENSMUSG00000030341/ENSMUSG00000023951/E
35941/ENSMUSG00000046733/ENSMUSG00000020349/ENSMUSG00000032715/ENSMUSG00000020484/ENSMUSG00000049323/ENSMUSG00000034765/ENSMUSG00000027996/ENSMUSG00000038400/E
78429/ENSMUSG00000035941/ENSMUSG00000046733/ENSMUSG00000004040/ENSMUSG00000020349/ENSMUSG00000032715/ENSMUSG00000020484/ENSMUSG00000049323/ENSMUSG00000034765/E
27996/ENSMUSG00000031523/ENSMUSG00000048616/ENSMUSG00000025340/ENSMUSG00000039384/ENSMUSG00000020682/ENSMUSG00000003665/ENSMUSG00000037815/ENSMUSG00000053797/E
31849/ENSMUSG00000048616/ENSMUSG00000056501/ENSMUSG00000024098/ENSMUSG00000023951/ENSMUSG00000026313/ENSMUSG00000056050/ENSMUSG00000027230/ENSMUSG00000026836/E
31523/ENSMUSG00000048616/ENSMUSG00000025340/ENSMUSG00000039384/ENSMUSG00000020682/ENSMUSG00000003665/ENSMUSG00000053797/ENSMUSG00000039202/ENSMUSG00000032702/E
02881/ENSMUSG00000029050/ENSMUSG00000035569/ENSMUSG00000027996/ENSMUSG00000036356/ENSMUSG00000031849/ENSMUSG00000048126/ENSMUSG00000020745/ENSMUSG00000073530/E
15846/ENSMUSG00000050567/ENSMUSG00000029287/ENSMUSG00000001419/ENSMUSG00000030557/ENSMUSG00000020423/ENSMUSG00000023034/ENSMUSG00000029050/ENSMUSG00000048616/E
31523/ENSMUSG00000048616/ENSMUSG00000025340/ENSMUSG00000039384/ENSMUSG00000020682/ENSMUSG00000003665/ENSMUSG00000037815/ENSMUSG00000053797/ENSMUSG00000039202/E
54808/ENSMUSG00000032702/ENSMUSG00000040945/ENSMUSG00000003814/ENSMUSG00000039952/ENSMUSG00000074934/ENSMUSG00000026812
48126/ENSMUSG00000073530/ENSMUSG00000034135/ENSMUSG00000026147/ENSMUSG00000028047/ENSMUSG00000001119
23067/ENSMUSG00000021950/ENSMUSG00000031778/ENSMUSG00000001131/ENSMUSG00000020484/ENSMUSG00000018736/ENSMUSG00000024789/ENSMUSG00000032340/ENSMUSG00000031451/E

31393

39523/ENSMUSG00000031548/ENSMUSG00000018846/ENSMUSG00000074934/ENSMUSG00000027712
50965/ENSMUSG00000056050/ENSMUSG00000034205/ENSMUSG00000001119/ENSMUSG00000074934
56501/ENSMUSG00000024098/ENSMUSG00000023951/ENSMUSG00000006445/ENSMUSG00000021379/ENSMUSG00000026313/ENSMUSG00000027230/ENSMUSG00000026836/ENSMUSG00000029249/E
29287/ENSMUSG00000001419/ENSMUSG00000020484/ENSMUSG00000020423/ENSMUSG00000023034/ENSMUSG00000029050/ENSMUSG00000048616/ENSMUSG00000007613/ENSMUSG00000033323/E
56050/ENSMUSG00000032487/ENSMUSG00000026836/ENSMUSG00000019943/ENSMUSG00000074934
24098/ENSMUSG00000023951/ENSMUSG00000046058/ENSMUSG00000061436/ENSMUSG00000048661/ENSMUSG00000000058/ENSMUSG00000027230/ENSMUSG00000031548/ENSMUSG00000044857/E
29050/ENSMUSG00000034640/ENSMUSG00000035569/ENSMUSG00000027996/ENSMUSG00000036356/ENSMUSG00000031849/ENSMUSG00000048616/ENSMUSG00000007613/ENSMUSG00000048126/E
56758/ENSMUSG00000029287/ENSMUSG00000025153/ENSMUSG00000020484/ENSMUSG00000024789/ENSMUSG00000048616/ENSMUSG00000007613/ENSMUSG00000005533/ENSMUSG00000056501/E
22425/ENSMUSG00000023951/ENSMUSG00000039384/ENSMUSG00000023068/ENSMUSG00000013076/ENSMUSG00000006445/ENSMUSG00000050965/ENSMUSG00000024486/ENSMUSG00000003814/E

ENSMUSG00000048126/ENSMUSG00000004846/ENSMUSG00000023951/ENSMUSG00000036606/ENSMUSG00000037820/ENSMUSG00000027832/ENSMUSG00000020044/ENSMUSG00000039462/ENSMUS

ENSMUSG00000020044/ENSMUSG00000039462/ENSMUSG00000003814/ENSMUSG00000039952/ENSMUSG00000034205/ENSMUSG00000028047/ENSMUSG00000031548/ENSMUSG00000001119/ENSMUS

ENSMUSG00000021831/ENSMUSG00000048661/ENSMUSG00000056050/ENSMUSG00000069895/ENSMUSG00000039952/ENSMUSG00000027230/ENSMUSG00000026147/ENSMUSG00000034205/ENSMUS

ENSMUSG00000039462/ENSMUSG00000021831/ENSMUSG00000048661/ENSMUSG00000056050/ENSMUSG00000069895/ENSMUSG00000039952/ENSMUSG00000027230/ENSMUSG00000026147/ENSMUS

ENSMUSG00000031778/ENSMUSG00000004040/ENSMUSG00000019997/ENSMUSG00000031714/ENSMUSG00000020484/ENSMUSG00000037242/ENSMUSG00000023034/ENSMUSG00000027996/ENSMUS

ENSMUSG00000036356/ENSMUSG00000031849/ENSMUSG00000048616/ENSMUSG00000056501/ENSMUSG00000024098/ENSMUSG00000039662/ENSMUSG00000023951/ENSMUSG00000035673/ENSMUS

ENSMUSG00000037447/ENSMUSG00000024927/ENSMUSG00000034135/ENSMUSG00000030747/ENSMUSG00000050965/ENSMUSG00000021379/ENSMUSG00000019936/ENSMUSG00000021036/ENSMUS

ENSMUSG00000040945/ENSMUSG00000003814/ENSMUSG00000039952/ENSMUSG00000031548/ENSMUSG00000074934/ENSMUSG00000037902/ENSMUSG00000026812

ENSMUSG00000006445/ENSMUSG00000050965/ENSMUSG00000032702/ENSMUSG00000024486/ENSMUSG00000048661/ENSMUSG00000003814/ENSMUSG00000056050/ENSMUSG00000032487/ENSMUS

ENSMUSG00000006445/ENSMUSG00000050965/ENSMUSG00000032702/ENSMUSG00000024486/ENSMUSG00000048661/ENSMUSG00000003814/ENSMUSG00000056050/ENSMUSG00000032487/ENSMUS

ENSMUSG00000006445/ENSMUSG00000050965/ENSMUSG00000032702/ENSMUSG00000024486/ENSMUSG00000048661/ENSMUSG00000003814/ENSMUSG00000056050/ENSMUSG00000032487/ENSMUS

ENSMUSG00000029050/ENSMUSG00000034640/ENSMUSG00000035569/ENSMUSG00000027996/ENSMUSG00000036356/ENSMUSG00000031849/ENSMUSG00000048616/ENSMUSG00000007613/ENSMUS

ENSMUSG00000020745/ENSMUSG00000053797/ENSMUSG00000013076/ENSMUSG00000022443/ENSMUSG00000006445/ENSMUSG00000050965/ENSMUSG00000032702/ENSMUSG00000024486/ENSMUS

ENSMUSG00000034640/ENSMUSG00000048616/ENSMUSG00000007613/ENSMUSG00000033323/ENSMUSG00000022687/ENSMUSG00000023951/ENSMUSG00000022752/ENSMUSG00000018476/ENSMUS

ENSMUSG00000031393/ENSMUSG00000074934

ENSMUSG00000030341/ENSMUSG00000023951/ENSMUSG00000061436/ENSMUSG00000006445/ENSMUSG00000050965/ENSMUSG00000032487/ENSMUSG00000062232/ENSMUSG00000027230/ENSMUS

ENSMUSG00000039202/ENSMUSG00000032702/ENSMUSG00000021149/ENSMUSG00000036564/ENSMUSG00000003814/ENSMUSG00000056050/ENSMUSG00000039952/ENSMUSG00000029361/ENSMUS

ENSMUSG00000026147/ENSMUSG00000034205/ENSMUSG00000028047/ENSMUSG00000001119/ENSMUSG00000074934

ENSMUSG00000061436/ENSMUSG00000006445/ENSMUSG00000050965/ENSMUSG00000032487/ENSMUSG00000027230/ENSMUSG00000015839/ENSMUSG00000031393/ENSMUSG00000042903/ENSMUS

ENSMUSG00000048616/ENSMUSG00000045983/ENSMUSG00000005533/ENSMUSG00000025340/ENSMUSG00000048234/ENSMUSG00000039384/ENSMUSG00000040021/ENSMUSG00000020745/ENSMUS

ENSMUSG00000027996/ENSMUSG00000038400/ENSMUSG00000048616/ENSMUSG00000045983/ENSMUSG00000005533/ENSMUSG00000025340/ENSMUSG00000048234/ENSMUSG00000039384/ENSMUS

ENSMUSG00000039202/ENSMUSG00000022043/ENSMUSG00000032702/ENSMUSG00000021149/ENSMUSG00000036564/ENSMUSG00000003814/ENSMUSG00000056050/ENSMUSG00000039952/ENSMUS

ENSMUSG00000029249/ENSMUSG00000031548/ENSMUSG00000019943/ENSMUSG00000074934/ENSMUSG00000046791/ENSMUSG00000005871

ENSMUSG00000021149/ENSMUSG00000036564/ENSMUSG00000003814/ENSMUSG00000056050/ENSMUSG00000039952/ENSMUSG00000015839/ENSMUSG00000028211/ENSMUSG00000031548/ENSMUS

ENSMUSG00000024812/ENSMUSG00000042594/ENSMUSG00000034135/ENSMUSG00000019936/ENSMUSG00000026147/ENSMUSG00000028047/ENSMUSG00000001119/ENSMUSG00000074934

ENSMUSG00000007613/ENSMUSG00000033323/ENSMUSG00000022687/ENSMUSG00000023951/ENSMUSG00000022752/ENSMUSG00000018476/ENSMUSG00000028161/ENSMUSG00000034998/ENSMUS

ENSMUSG00000032702/ENSMUSG00000021149/ENSMUSG00000036564/ENSMUSG00000003814/ENSMUSG00000056050/ENSMUSG00000039952/ENSMUSG00000015839/ENSMUSG00000028211/ENSMUS

ENSMUSG00000031849/ENSMUSG00000048616/ENSMUSG00000005533/ENSMUSG00000005087/ENSMUSG00000039384/ENSMUSG00000085148/ENSMUSG00000039202/ENSMUSG00000042594/ENSMUS

ENSMUSG00000031548/ENSMUSG00000074934/ENSMUSG00000046791/ENSMUSG00000005871

ENSMUSG00000022687/ENSMUSG00000022752/ENSMUSG00000046058/ENSMUSG00000028161/ENSMUSG00000034998/ENSMUSG00000026313/ENSMUSG00000048661/ENSMUSG00000045098/ENSMUS

ENSMUSG00000085795/ENSMUSG00000063382/ENSMUSG00000074934/ENSMUSG00000024182

ENSMUSG00000023951/ENSMUSG00000073530/ENSMUSG00000034135/ENSMUSG00000032717/ENSMUSG00000040268/ENSMUSG00000026147/ENSMUSG00000028047/ENSMUSG00000031548/ENSMUS

ENSMUSG00000005087/ENSMUSG00000024098/ENSMUSG00000024927/ENSMUSG00000046223/ENSMUSG00000039662/ENSMUSG00000023951/ENSMUSG00000040021/ENSMUSG00000024085/ENSMUS

ENSMUSG00000032487/ENSMUSG00000015839/ENSMUSG00000044147/ENSMUSG00000031393

;G00000019936/ENSMUSG00000003814/ENSMUSG00000039952/ENSMUSG00000026147/ENSMUSG00000034205/ENSMUSG00000028047/ENSMUSG00000031548/ENSMUSG00000001119/ENSMUSG0000(
;G00000027712/ENSMUSG00000031386
;G00000025185/ENSMUSG00000074934/ENSMUSG00000032220
;G00000034205/ENSMUSG00000025185/ENSMUSG00000074934/ENSMUSG00000032220
;G00000022425/ENSMUSG00000030341/ENSMUSG00000007613/ENSMUSG00000045216/ENSMUSG00000023951/ENSMUSG00000044337/ENSMUSG00000023068/ENSMUSG00000023008/ENSMUSG0000(

;G00000006445/ENSMUSG00000034135/ENSMUSG00000021379/ENSMUSG00000026313/ENSMUSG00000056050/ENSMUSG00000032487/ENSMUSG00000027230/ENSMUSG00000026836/ENSMUSG0000(
;G00000056050/ENSMUSG00000027665/ENSMUSG00000026147/ENSMUSG00000034205/ENSMUSG00000028047/ENSMUSG00000001119/ENSMUSG00000074934

;G00000034205/ENSMUSG00000015839/ENSMUSG00000044147/ENSMUSG00000031393/ENSMUSG00000074934
;G00000034205/ENSMUSG00000015839/ENSMUSG00000044147/ENSMUSG00000031393/ENSMUSG00000074934

;G00000034205/ENSMUSG00000015839/ENSMUSG00000044147/ENSMUSG00000031393/ENSMUSG00000074934

;G00000048126/ENSMUSG00000028521/ENSMUSG00000037447/ENSMUSG00000024927/ENSMUSG00000023951/ENSMUSG00000020745/ENSMUSG00000073530/ENSMUSG00000024812/ENSMUSG0000(

;G00000048661/ENSMUSG00000040945/ENSMUSG00000003814/ENSMUSG00000056050/ENSMUSG00000032487/ENSMUSG00000026836/ENSMUSG00000034205/ENSMUSG00000015839/ENSMUSG0000(
;G00000028161/ENSMUSG00000034998/ENSMUSG00000026313/ENSMUSG00000048661/ENSMUSG00000000058/ENSMUSG00000026836/ENSMUSG00000024431/ENSMUSG00000044857/ENSMUSG0000(

;G00000028179/ENSMUSG00000015839/ENSMUSG00000031393/ENSMUSG00000042903/ENSMUSG00000074934

;G00000015839/ENSMUSG00000028211/ENSMUSG00000031548/ENSMUSG00000031393/ENSMUSG00000074934

;G00000074934
;G00000027177/ENSMUSG00000042594/ENSMUSG00000020044/ENSMUSG00000021070/ENSMUSG00000024423/ENSMUSG00000109511/ENSMUSG00000050965/ENSMUSG00000079037/ENSMUSG0000(
;G00000040021/ENSMUSG00000020745/ENSMUSG00000027177/ENSMUSG00000042594/ENSMUSG00000020044/ENSMUSG00000021070/ENSMUSG00000024423/ENSMUSG00000109511/ENSMUSG0000(
;G00000015839/ENSMUSG00000028211/ENSMUSG00000031548/ENSMUSG00000031393/ENSMUSG00000074934

;G00000031393/ENSMUSG00000074934

;G00000026313/ENSMUSG00000048661/ENSMUSG00000000058/ENSMUSG00000026836/ENSMUSG00000024431/ENSMUSG00000044857/ENSMUSG00000063382/ENSMUSG00000074934/ENSMUSG0000(
;G00000031548/ENSMUSG00000031393/ENSMUSG00000074934

;G00000022443/ENSMUSG00000050965/ENSMUSG00000030168/ENSMUSG00000032702/ENSMUSG00000024486/ENSMUSG00000008398/ENSMUSG00000056050/ENSMUSG00000026131/ENSMUSG0000(

;G00000000058/ENSMUSG00000078671/ENSMUSG00000024431/ENSMUSG00000042903/ENSMUSG00000044857/ENSMUSG00000063382/ENSMUSG00000074934/ENSMUSG00000050697/ENSMUSG0000(

;G00000001119/ENSMUSG00000074934/ENSMUSG00000024182
;G00000037820/ENSMUSG00000006445/ENSMUSG00000028042/ENSMUSG00000021379/ENSMUSG00000018634/ENSMUSG00000039952/ENSMUSG00000027665/ENSMUSG00000033454/ENSMUSG0000(

0068479/ENSMUSG00000027712/ENSMUSG00000031386

0013076/ENSMUSG00000022443/ENSMUSG00000061436/ENSMUSG00000006445/ENSMUSG00000050965/ENSMUSG00000030168/ENSMUSG00000024486/ENSMUSG00000048661/ENSMUSG0000000839

0029249/ENSMUSG00000028047/ENSMUSG00000031548/ENSMUSG00000019943/ENSMUSG00000074934/ENSMUSG00000046791/ENSMUSG00000005871

0042594/ENSMUSG00000006445/ENSMUSG00000034135/ENSMUSG00000032717/ENSMUSG00000050965/ENSMUSG00000019936/ENSMUSG00000026313/ENSMUSG00000040268/ENSMUSG0000005605

0044147/ENSMUSG00000031393/ENSMUSG00000074934
0063382/ENSMUSG00000074934/ENSMUSG00000050697/ENSMUSG00000026812/ENSMUSG00000026628/ENSMUSG00000005871

0015202/ENSMUSG00000053716/ENSMUSG00000039952/ENSMUSG00000044167/ENSMUSG00000029361/ENSMUSG00000031548/ENSMUSG00000039844/ENSMUSG00000044857/ENSMUSG0000007493
0050965/ENSMUSG00000026313/ENSMUSG00000079037/ENSMUSG00000015202/ENSMUSG00000053716/ENSMUSG00000039952/ENSMUSG00000044167/ENSMUSG00000029361/ENSMUSG0000003154

0050697/ENSMUSG00000026812/ENSMUSG00000026628/ENSMUSG00000005871

0033717/ENSMUSG00000015839/ENSMUSG00000026797/ENSMUSG00000056071/ENSMUSG00000027712

0026628/ENSMUSG00000005871

0031548/ENSMUSG00000044147/ENSMUSG00000024431/ENSMUSG00000085795/ENSMUSG00000025326/ENSMUSG00000041702/ENSMUSG00000004207/ENSMUSG00000005871

8/ENSMUSG00000056050/ENSMUSG00000032487/ENSMUSG00000027665/ENSMUSG00000027230/ENSMUSG00000026836/ENSMUSG0000003420

0/ENSMUSG00000026147/ENSMUSG00000034205/ENSMUSG00000028047/ENSMUSG00000031548/ENSMUSG00000001119/ENSMUSG0000007493

4/ENSMUSG00000037902/ENSMUSG00000026628/ENSMUSG00000005871
8/ENSMUSG00000039844/ENSMUSG00000044857/ENSMUSG00000074934/ENSMUSG00000037902/ENSMUSG00000035621/ENSMUSG0000002662

| Category | GOID | Description | GeneRatio | BgRatio | pvalue | padj | geneID | geneName |
|---|---|---|---|---|---|---|---|---|
| BP | GO:00420 | wound healing | 59\|1209 | 336\|1609 | 6.42E-10 | 8.27E-08 | ENSMUS | Col3a1/Col1a1/F3/Thbs1/Hmox1/Hmgcr/Serpine1/Plec/Plet1/I |
| BP | GO:00107 | negative regulation of cell development | 62\|1209 | 362\|1609 | 6.62E-10 | 8.33E-08 | ENSMUS | Col3a1/Ldlr/Rgs2/Sox9/Postn/Xylt1/Ifrd1/Efna1/Rtn4/Lgals1/Di |
| BP | GO:00900 | regulation of transmembrane receptor protein serine/threonin | 44\|1209 | 216\|1609 | 8.99E-10 | 1.09E-07 | ENSMUS | Thbs1/Vasn/Cilp/Inhba/Cd109/Cdh5/Aspn/Chrdl1/Cyr61/Bmp6 |
| BP | GO:00456 | regulation of osteoblast differentiation | 32\|1209 | 128\|1609 | 9.03E-10 | 1.09E-07 | ENSMUS | Igfbp5/Dlk1/Fgfr1/Smoc1/Noct/Cyr61/Lmna/Smad5/Bmp6/Fnd |
| BP | GO:00506 | epithelial cell proliferation | 66\|1209 | 401\|1609 | 1.03E-09 | 1.21E-07 | ENSMUS | Col8a1/Errfi1/Sulf2/F3/Thbs1/Hmox1/Myc/Igfbp5/Sox9/Dlk1/T |
| BP | GO:00071 | transmembrane receptor protein serine/threonine kinase signa | 56\|1209 | 315\|1609 | 1.10E-09 | 1.26E-07 | ENSMUS | Col3a1/Col1a2/Thbs1/Vasn/Cilp/Inhba/Egr1/Cd109/Jun/Cdh5/ |
| BP | GO:00016 | urogenital system development | 56\|1209 | 319\|1609 | 1.77E-09 | 2.00E-07 | ENSMUS | Col4a1/Sulf2/Myc/Sox9/Nid1/Egr1/Serpinf1/Fgfr1/Fem1b/Cdkn |
| BP | GO:00302 | bone mineralization | 29\|1209 | 113\|1609 | 2.87E-09 | 3.17E-07 | ENSMUS | Col1a2/Sox9/Dlk1/Omd/Aspn/Clec3b/Eif2ak3/Cyr61/Ank/Fgfr2/ |
| CC | GO:00324 | actin filament bundle | 26\|1232 | 93\|16313 | 2.96E-09 | 3.54E-07 | ENSMUS | Acta1/Marcks/Ablim3/Mylk/Rflna/Synpo/Myh11/Dlc1/Ldb3/Pls |
| BP | GO:00436 | regulation of DNA-templated transcription in response to stres | 19\|1209 | 53\|16097 | 3.82E-09 | 4.14E-07 | ENSMUS | Hmox1/Egr1/Atf4/Jun/Eif2ak3/Ddit3/Sesn2/Cd36/Cebpb/Rela/V |
| BP | GO:00436 | regulation of transcription from RNA polymerase II promoter ir | 18\|1209 | 48\|16097 | 4.26E-09 | 4.49E-07 | ENSMUS | Hmox1/Egr1/Atf4/Jun/Eif2ak3/Ddit3/Sesn2/Cd36/Cebpb/Vegfa/ |
| BP | GO:00160 | cell growth | 73\|1209 | 479\|1609 | 4.32E-09 | 4.49E-07 | ENSMUS | Col14a1/Igfbp5/Rgs2/Sox9/Postn/Emp1/Inhba/Ifrd1/Igfbp3/Tnf |
| BP | GO:00020 | chondrocyte development | 17\|1209 | 43\|16097 | 4.45E-09 | 4.54E-07 | ENSMUS | Sulf2/Col14a1/Sox9/Col12a1/Eif2ak3/Col6a2/Rflna/Chst11/Ma |
| BP | GO:00611 | morphogenesis of a branching epithelium | 42\|1209 | 212\|1609 | 5.18E-09 | 5.19E-07 | ENSMUS | Col4a1/Spry2/Myc/Sox9/Fgfr1/Fem1b/Met/Mmp14/Ncoa3/Fgfr |
| CC | GO:00055 | collagen trimer | 23\|1232 | 77\|16313 | 5.93E-09 | 5.91E-07 | ENSMUS | Col8a1/Col3a1/Col1a1/Col1a2/Col4a1/Col14a1/Col5a3/Col5a2 |
| BP | GO:00328 | cellular response to hormone stimulus | 72\|1209 | 474\|1609 | 6.37E-09 | 6.26E-07 | ENSMUS | Inhba/Lpl/Ywhag/Jun/Sorbs2/Eif4ebp1/Atp1a1/Ncoa3/Socs3/H |
| BP | GO:00017 | morphogenesis of a branching structure | 44\|1209 | 230\|1609 | 6.90E-09 | 6.66E-07 | ENSMUS | Col4a1/Spry2/Myc/Sox9/Fgfr1/Fem1b/Met/Rtn4/Mmp14/Ncoa |
| BP | GO:00105 | regulation of endothelial cell migration | 33\|1209 | 146\|1609 | 7.76E-09 | 7.36E-07 | ENSMUS | Thbs1/Hmox1/Map2k3/Klf4/Serpinf1/Fgfr1/Met/Efna1/Smoc2/ |
| BP | GO:00330 | muscle cell proliferation | 43\|1209 | 223\|1609 | 7.99E-09 | 7.45E-07 | ENSMUS | Thbs1/Hmox1/Hmgcr/Myc/Igfbp5/Trib1/Klf4/Rgs5/Egr1/Fgfr1/I |
| BP | GO:00714 | cellular response to external stimulus | 46\|1209 | 248\|1609 | 8.67E-09 | 7.95E-07 | ENSMUS | Col1a1/Fosl1/Sox9/Postn/Piezo1/Lpl/Slc2a1/Atf4/Jun/Atp1a1/ |
| BP | GO:00486 | smooth muscle cell proliferation | 35\|1209 | 162\|1609 | 9.62E-09 | 8.66E-07 | ENSMUS | Thbs1/Hmox1/Hmgcr/Myc/Igfbp5/Trib1/Klf4/Rgs5/Egr1/Igfbp3/ |
| MF | GO:00508 | extracellular matrix binding | 19\|1190 | 56\|15912 | 1.02E-08 | 1.32E-06 | ENSMUS | Thbs1/Eln/Nid1/Lgals3/Lgals1/Smoc1/Cyr61/Fbln2/Adgrg1/Olf |
| BP | GO:00457 | positive regulation of cell adhesion | 62\|1209 | 391\|1609 | 1.43E-08 | 1.27E-06 | ENSMUS | Col8a1/Thbs1/Itga5/Nid1/Piezo1/Has2/S100a10/Il6ra/Lgals1/S |
| BP | GO:00486 | regulation of smooth muscle cell proliferation | 34\|1209 | 157\|1609 | 1.47E-08 | 1.28E-06 | ENSMUS | Thbs1/Hmox1/Hmgcr/Myc/Igfbp5/Trib1/Klf4/Rgs5/Egr1/Igfbp3/ |
| BP | GO:00603 | cartilage development involved in endochondral bone morpho | 17\|1209 | 46\|16097 | 1.49E-08 | 1.28E-06 | ENSMUS | Col1a1/Thbs1/Col14a1/Sox9/Col12a1/Col6a2/Matn3/Hspg2/C |
| BP | GO:00434 | negative regulation of MAPK cascade | 35\|1209 | 165\|1609 | 1.58E-08 | 1.34E-06 | ENSMUS | Errfi1/Spry2/Hmgcr/Rgs2/Klf4/Dusp1/Efna1/Spred1/Xbp1/Hsph |
| BP | GO:00226 | gland morphogenesis | 32\|1209 | 143\|1609 | 1.69E-08 | 1.40E-06 | ENSMUS | Sulf2/Igfbp5/Sox9/Fgfr1/Fem1b/Met/Rtn4/Ncoa3/Fgfr2/Rxra/E |
| MF | GO:00051 | integrin binding | 29\|1190 | 122\|1591 | 1.71E-08 | 1.94E-06 | ENSMUS | Col3a1/Thbs1/Itga5/Lamb1/Mmp14/Cyr61/Cx3cl1/Thy1/Egfr/T |
| CC | GO:00426 | actomyosin | 25\|1232 | 95\|16313 | 2.28E-08 | 1.94E-06 | ENSMUS | Acta1/Ablim3/Mylk/Synpo/Myh11/Dlc1/Ldb3/Tlnrd1/Acta2/Ac |
| BP | GO:19040 | positive regulation of vasculature development | 38\|1209 | 191\|1609 | 2.45E-08 | 2.01E-06 | ENSMUS | F3/Thbs1/Hmox1/Serpine1/Klf4/Itga5/Egr1/Il6ra/Lgals3/Cdh5/ |
| BP | GO:00106 | positive regulation of epithelial cell migration | 31\|1209 | 138\|1609 | 2.56E-08 | 2.06E-06 | ENSMUS | Thbs1/Hmox1/Sox9/Map2k3/Serpine1/Fgfr1/Has2/Met/Jun/Rtn |
| BP | GO:00020 | cell migration involved in sprouting angiogenesis | 18\|1209 | 53\|16097 | 2.62E-08 | 2.09E-06 | ENSMUS | Thbs1/Hmox1/Klf4/Spred1/Meox2/Nr4a1/Nrp1/Robo1/Pik3r2/ |
| BP | GO:00901 | negative regulation of transmembrane receptor protein serine/ | 26\|1209 | 103\|1609 | 2.72E-08 | 2.13E-06 | ENSMUS | Vasn/Cilp/Cd109/Aspn/Chrdl1/Tgfbr3/Chst11/Xbp1/Ski/Sfrp2/ |
| BP | GO:00714 | cellular response to organonitrogen compound | 69\|1209 | 464\|1609 | 3.16E-08 | 2.44E-06 | ENSMUS | Col3a1/Col1a1/Col1a2/Col4a1/Tlr2/Igfbp5/Sox9/Col5a2/Egr1/ |
| BP | GO:00902 | negative regulation of cellular response to growth factor stimul | 30\|1209 | 132\|1609 | 3.23E-08 | 2.46E-06 | ENSMUS | Spry2/Sulf2/Thbs1/Vasn/Tnmd/Cd109/Aspn/Chrdl1/Tgfbr3/Chs |
| BP | GO:00486 | positive regulation of smooth muscle cell proliferation | 26\|1209 | 104\|1609 | 3.37E-08 | 2.53E-06 | ENSMUS | Thbs1/Hmox1/Hmgcr/Myc/Igfbp5/Egr1/Il6ra/Jun/Fgfr2/Cx3cl1/ |
| BP | GO:20001 | negative regulation of blood vessel morphogenesis | 27\|1209 | 111\|1609 | 3.44E-08 | 2.55E-06 | ENSMUS | Spry2/Thbs1/Serpine1/Tnmd/Klf4/Serpinf1/Thbs2/Col4a2/Rtn4 |
| BP | GO:00715 | cellular response to transforming growth factor beta stimulus | 38\|1209 | 194\|1609 | 3.79E-08 | 2.75E-06 | ENSMUS | Col3a1/Col1a2/Thbs1/Vasn/Sox9/Serpine1/Cilp/Cd109/Col4a2/ |
| BP | GO:00160 | Wnt signaling pathway | 61\|1209 | 392\|1609 | 3.81E-08 | 2.75E-06 | ENSMUS | Col1a1/Sulf2/Myc/Tlr2/Sox9/Klf4/Egr1/Wif1/Met/Ddit3/Abl2/F |
| MF | GO:00508 | cell adhesion molecule binding | 42\|1190 | 228\|1591 | 4.15E-08 | 4.18E-06 | ENSMUS | Col3a1/Thbs1/Postn/Itga5/Lamb1/Fgfr1/Cdh5/Rtn4/Mmp14/C |
| BP | GO:01987 | cell-cell signaling by wnt | 61\|1209 | 393\|1609 | 4.19E-08 | 2.98E-06 | ENSMUS | Col1a1/Sulf2/Myc/Tlr2/Sox9/Klf4/Egr1/Wif1/Met/Ddit3/Abl2/F |
| BP | GO:00308 | prostate gland development | 17\|1209 | 49\|16097 | 4.45E-08 | 3.12E-06 | ENSMUS | Sox9/Serpinf1/Fem1b/Fgfr2/Rxra/Tnc/Nog/Igf1r/Igf1/Cd44/Plau |
| BP | GO:00506 | chondroitin sulfate proteoglycan metabolic process | 13\|1209 | 29\|16097 | 4.94E-08 | 3.42E-06 | ENSMUS | Xylt1/Csgalnact2/Chpf/Chst11/Cytl1/Csgalnact1/Pxylp1/Slc35d |
| BP | GO:00082 | steroid metabolic process | 43\|1209 | 237\|1609 | 5.14E-08 | 3.51E-06 | ENSMUS | Hmgcr/Ldlr/Cyp51/Egr1/Fgfr1/Stard4/Scd1/Fdft1/Msmo1/Atp1 |
| BP | GO:00165 | negative regulation of angiogenesis | 26\|1209 | 107\|1609 | 6.30E-08 | 4.25E-06 | ENSMUS | Spry2/Thbs1/Serpine1/Tnmd/Klf4/Serpinf1/Thbs2/Col4a2/Spre |
| BP | GO:00486 | reproductive structure development | 62\|1209 | 407\|1609 | 6.55E-08 | 4.37E-06 | ENSMUS | Fosl1/Sox9/Serpine1/Inhba/Serpinf1/Xist/Fem1b/Met/Cdkn1c/ |
| BP | GO:00715 | response to transforming growth factor beta | 38\|1209 | 198\|1609 | 6.68E-08 | 4.40E-06 | ENSMUS | Col3a1/Col1a2/Thbs1/Vasn/Sox9/Serpine1/Cilp/Cd109/Col4a2/ |
| BP | GO:00426 | muscle cell differentiation | 59\|1209 | 380\|1609 | 6.91E-08 | 4.50E-06 | ENSMUS | Acta1/Col14a1/Igfbp5/Rgs2/Sox9/Cdon/Ifrd1/Met/Rcan1/Sorbs |
| BP | GO:19013 | negative regulation of vasculature development | 28\|1209 | 122\|1609 | 7.39E-08 | 4.75E-06 | ENSMUS | Spry2/Thbs1/Serpine1/Tnmd/Klf4/Serpinf1/Thbs2/Col4a2/Rtn4 |
| BP | GO:00614 | reproductive system development | 62\|1209 | 410\|1609 | 8.60E-08 | 5.40E-06 | ENSMUS | Fosl1/Sox9/Serpine1/Inhba/Serpinf1/Xist/Fem1b/Met/Cdkn1c/ |
| BP | GO:00456 | negative regulation of osteoblast differentiation | 17\|1209 | 51\|16097 | 8.76E-08 | 5.40E-06 | ENSMUS | Igfbp5/Dlk1/Fgfr1/Noct/Fndc3b/Ski/Nog/Twsg1/Nfatc1/Notch1 |

| Count | Up | Up_Gene_i | Down | Down_Gene_id |
|---|---|---|---|---|
| 59 | 25 | ENSMUSG0 | 34 | ENSMUSG00000028128/ENSMUSG00000040152/ENSMUSG00000005413/ENSMUSG00000021670/ENSMUSG00000037411/ENSMUSG00000022565/ENSMUSG0 |
| 62 | 22 | ENSMUSG0 | 40 | ENSMUSG00000032193/ENSMUSG00000026360/ENSMUSG00000000567/ENSMUSG00000030657/ENSMUSG00000001627/ENSMUSG00000027954/ENSMUSG0 |
| 44 | 18 | ENSMUSG0 | 26 | ENSMUSG00000040152/ENSMUSG00000039646/ENSMUSG00000041324/ENSMUSG00000046186/ENSMUSG00000039004/ENSMUSG00000029287/ENSMUSG0 |
| 32 | 10 | ENSMUSG0 | 22 | ENSMUSG00000040856/ENSMUSG00000031565/ENSMUSG00000021136/ENSMUSG00000023087/ENSMUSG00000028063/ENSMUSG00000021540/ENSMUSG0 |
| 66 | 27 | ENSMUSG0 | 39 | ENSMUSG00000068196/ENSMUSG00000028967/ENSMUSG00000006800/ENSMUSG00000028128/ENSMUSG00000040152/ENSMUSG00000005413/ENSMUSG0 |
| 56 | 23 | ENSMUSG0 | 33 | ENSMUSG00000040152/ENSMUSG00000039646/ENSMUSG00000041324/ENSMUSG00000038418/ENSMUSG00000046186/ENSMUSG00000021540/ENSMUSG0 |
| 56 | 25 | ENSMUSG0 | 31 | ENSMUSG00000006800/ENSMUSG00000022346/ENSMUSG00000000567/ENSMUSG00000038418/ENSMUSG00000000753/ENSMUSG00000031565/ENSMUSG0 |
| 29 | 14 | ENSMUSG0 | 15 | ENSMUSG00000000567/ENSMUSG00000040856/ENSMUSG00000031668/ENSMUSG00000022265/ENSMUSG00000030849/ENSMUSG00000039004/ENSMUSG0 |
| 26 | 19 | ENSMUSG0 | 7 | ENSMUSG00000031523/ENSMUSG00000070462/ENSMUSG00000054808/ENSMUSG00000022443/ENSMUSG00000032643/ENSMUSG00000026131/ENSMUSG0 |
| 19 | 3 | ENSMUSG0 | 16 | ENSMUSG00000005413/ENSMUSG00000038418/ENSMUSG00000042406/ENSMUSG00000031668/ENSMUSG00000025408/ENSMUSG00000028893/ENSMUSG0 |
| 18 | 3 | ENSMUSG0 | 15 | ENSMUSG00000005413/ENSMUSG00000038418/ENSMUSG00000042406/ENSMUSG00000031668/ENSMUSG00000025408/ENSMUSG00000028893/ENSMUSG0 |
| 73 | 28 | ENSMUSG0 | 45 | ENSMUSG00000026360/ENSMUSG00000000567/ENSMUSG00000030208/ENSMUSG00000041324/ENSMUSG00000001627/ENSMUSG00000023905/ENSMUSG0 |
| 17 | 8 | ENSMUSG0 | 9 | ENSMUSG00000006800/ENSMUSG00000000567/ENSMUSG00000031668/ENSMUSG00000020241/ENSMUSG00000027996/ENSMUSG00000031849/ENSMUSG0 |
| 42 | 20 | ENSMUSG0 | 22 | ENSMUSG00000022114/ENSMUSG00000022346/ENSMUSG00000000567/ENSMUSG00000031565/ENSMUSG00000032244/ENSMUSG00000009376/ENSMUSG0 |
| 23 | 17 | ENSMUSG0 | 6 | ENSMUSG00000068196/ENSMUSG00000020241/ENSMUSG00000048126/ENSMUSG00000039462/ENSMUSG00000026147/ENSMUSG00000001119 |
| 72 | 20 | ENSMUSG0 | 52 | ENSMUSG00000041324/ENSMUSG00000051391/ENSMUSG00000031490/ENSMUSG00000033161/ENSMUSG00000027678/ENSMUSG00000056758/ENSMUSG0 |
| 44 | 20 | ENSMUSG0 | 24 | ENSMUSG00000022114/ENSMUSG00000022346/ENSMUSG00000000567/ENSMUSG00000031565/ENSMUSG00000032244/ENSMUSG00000009376/ENSMUSG0 |
| 33 | 15 | ENSMUSG0 | 18 | ENSMUSG00000040152/ENSMUSG00000005413/ENSMUSG00000018932/ENSMUSG00000003032/ENSMUSG00000000753/ENSMUSG00000031565/ENSMUSG0 |
| 43 | 14 | ENSMUSG0 | 29 | ENSMUSG00000040152/ENSMUSG00000005413/ENSMUSG00000021670/ENSMUSG00000022346/ENSMUSG00000032501/ENSMUSG00000003032/ENSMUSG0 |
| 46 | 14 | ENSMUSG0 | 32 | ENSMUSG00000024912/ENSMUSG00000000567/ENSMUSG00000014444/ENSMUSG00000028645/ENSMUSG00000042406/ENSMUSG00000033161/ENSMUSG0 |
| 35 | 13 | ENSMUSG0 | 22 | ENSMUSG00000040152/ENSMUSG00000005413/ENSMUSG00000021670/ENSMUSG00000022346/ENSMUSG00000032501/ENSMUSG00000003032/ENSMUSG0 |
| 19 | 12 | ENSMUSG0 | 7 | ENSMUSG00000040152/ENSMUSG00000029675/ENSMUSG00000050335/ENSMUSG00000021136/ENSMUSG00000031785/ENSMUSG00000023951/ENSMUSG0 |
| 62 | 29 | ENSMUSG0 | 33 | ENSMUSG00000068196/ENSMUSG00000040152/ENSMUSG00000000555/ENSMUSG00000014444/ENSMUSG00000022367/ENSMUSG00000041959/ENSMUSG0 |
| 34 | 12 | ENSMUSG0 | 22 | ENSMUSG00000040152/ENSMUSG00000005413/ENSMUSG00000021670/ENSMUSG00000022346/ENSMUSG00000032501/ENSMUSG00000003032/ENSMUSG0 |
| 17 | 8 | ENSMUSG0 | 9 | ENSMUSG00000040152/ENSMUSG00000000567/ENSMUSG00000020241/ENSMUSG00000031849/ENSMUSG00000048126/ENSMUSG00000034135/ENSMUSG0 |
| 35 | 6 | ENSMUSG0 | 29 | ENSMUSG00000028967/ENSMUSG00000022114/ENSMUSG00000021670/ENSMUSG00000026360/ENSMUSG00000003032/ENSMUSG00000027954/ENSMUSG0 |
| 32 | 10 | ENSMUSG0 | 22 | ENSMUSG00000006800/ENSMUSG00000000567/ENSMUSG00000031565/ENSMUSG00000032244/ENSMUSG00000009376/ENSMUSG00000020458/ENSMUSG0 |
| 29 | 19 | ENSMUSG0 | 10 | ENSMUSG00000040152/ENSMUSG00000000555/ENSMUSG00000031778/ENSMUSG00000019997/ENSMUSG00000031849/ENSMUSG00000022817/ENSMUSG0 |
| 25 | 16 | ENSMUSG0 | 9 | ENSMUSG00000031523/ENSMUSG00000070462/ENSMUSG00000054808/ENSMUSG00000022443/ENSMUSG00000032643/ENSMUSG00000026313/ENSMUSG0 |
| 38 | 13 | ENSMUSG0 | 25 | ENSMUSG00000028128/ENSMUSG00000040152/ENSMUSG00000005413/ENSMUSG00000037411/ENSMUSG00000003032/ENSMUSG00000000555/ENSMUSG0 |
| 31 | 12 | ENSMUSG0 | 19 | ENSMUSG00000040152/ENSMUSG00000005413/ENSMUSG00000000567/ENSMUSG00000018932/ENSMUSG00000037411/ENSMUSG00000031565/ENSMUSG0 |
| 18 | 9 | ENSMUSG0 | 9 | ENSMUSG00000040152/ENSMUSG00000005413/ENSMUSG00000003032/ENSMUSG00000027351/ENSMUSG00000023034/ENSMUSG00000023951/ENSMUSG0 |
| 26 | 8 | ENSMUSG0 | 18 | ENSMUSG00000039646/ENSMUSG00000046186/ENSMUSG00000029287/ENSMUSG00000020484/ENSMUSG00000029050/ENSMUSG00000027996/ENSMUSG0 |
| 69 | 20 | ENSMUSG0 | 49 | ENSMUSG00000027995/ENSMUSG00000000567/ENSMUSG00000038418/ENSMUSG00000051391/ENSMUSG00000042406/ENSMUSG00000031490/ENSMUSG0 |
| 30 | 9 | ENSMUSG0 | 21 | ENSMUSG00000022114/ENSMUSG00000006800/ENSMUSG00000040152/ENSMUSG00000039646/ENSMUSG00000046186/ENSMUSG00000029287/ENSMUSG0 |
| 26 | 8 | ENSMUSG0 | 18 | ENSMUSG00000040152/ENSMUSG00000005413/ENSMUSG00000021670/ENSMUSG00000022346/ENSMUSG00000038418/ENSMUSG00000027947/ENSMUSG0 |
| 27 | 15 | ENSMUSG0 | 12 | ENSMUSG00000022114/ENSMUSG00000040152/ENSMUSG00000037411/ENSMUSG00000003032/ENSMUSG00000000753/ENSMUSG00000020458/ENSMUSG0 |
| 38 | 14 | ENSMUSG0 | 24 | ENSMUSG00000040152/ENSMUSG00000039646/ENSMUSG00000000567/ENSMUSG00000037411/ENSMUSG00000046186/ENSMUSG00000021540/ENSMUSG0 |
| 61 | 21 | ENSMUSG0 | 40 | ENSMUSG00000006800/ENSMUSG00000022346/ENSMUSG00000027995/ENSMUSG00000000567/ENSMUSG00000003032/ENSMUSG00000038418/ENSMUSG0 |
| 42 | 23 | ENSMUSG0 | 19 | ENSMUSG00000040152/ENSMUSG00000000555/ENSMUSG00000031565/ENSMUSG00000020458/ENSMUSG00000009687/ENSMUSG00000031778/ENSMUSG0 |
| 61 | 21 | ENSMUSG0 | 40 | ENSMUSG00000006800/ENSMUSG00000022346/ENSMUSG00000027995/ENSMUSG00000000567/ENSMUSG00000003032/ENSMUSG00000038418/ENSMUSG0 |
| 17 | 3 | ENSMUSG0 | 14 | ENSMUSG00000000567/ENSMUSG00000000753/ENSMUSG00000032244/ENSMUSG00000030849/ENSMUSG00000015846/ENSMUSG00000048616/ENSMUSG0 |
| 13 | 6 | ENSMUSG0 | 7 | ENSMUSG00000030657/ENSMUSG00000042042/ENSMUSG00000032997/ENSMUSG00000036356/ENSMUSG00000043587/ENSMUSG00000028521/ENSMUSG0 |
| 43 | 9 | ENSMUSG0 | 34 | ENSMUSG00000021670/ENSMUSG00000032193/ENSMUSG00000001467/ENSMUSG00000038418/ENSMUSG00000031565/ENSMUSG00000024378/ENSMUSG0 |
| 26 | 15 | ENSMUSG0 | 11 | ENSMUSG00000022114/ENSMUSG00000040152/ENSMUSG00000037411/ENSMUSG00000003032/ENSMUSG00000000753/ENSMUSG00000027351/ENSMUSG0 |
| 62 | 19 | ENSMUSG0 | 43 | ENSMUSG00000024912/ENSMUSG00000000567/ENSMUSG00000037411/ENSMUSG00000041324/ENSMUSG00000000753/ENSMUSG00000032244/ENSMUSG0 |
| 38 | 14 | ENSMUSG0 | 24 | ENSMUSG00000040152/ENSMUSG00000039646/ENSMUSG00000000567/ENSMUSG00000037411/ENSMUSG00000046186/ENSMUSG00000021540/ENSMUSG0 |
| 59 | 26 | ENSMUSG0 | 33 | ENSMUSG00000026360/ENSMUSG00000000567/ENSMUSG00000038119/ENSMUSG00000001627/ENSMUSG00000009376/ENSMUSG00000022951/ENSMUSG0 |
| 28 | 16 | ENSMUSG0 | 12 | ENSMUSG00000022114/ENSMUSG00000040152/ENSMUSG00000037411/ENSMUSG00000003032/ENSMUSG00000000753/ENSMUSG00000020458/ENSMUSG0 |
| 62 | 19 | ENSMUSG0 | 43 | ENSMUSG00000024912/ENSMUSG00000000567/ENSMUSG00000037411/ENSMUSG00000041324/ENSMUSG00000000753/ENSMUSG00000032244/ENSMUSG0 |
| 17 | 6 | ENSMUSG0 | 11 | ENSMUSG00000040856/ENSMUSG00000031565/ENSMUSG00000023087/ENSMUSG00000039286/ENSMUSG00000029050/ENSMUSG00000048616/ENSMUSG0 |

0000032068/ENSMUSG00000001627/ENSMUSG00000023905/ENSMUSG00000046186/ENSMUSG00000023067/ENSMUSG00000021950/ENSMUSG00000031778/ENSMUSG00000001131/ENSMUSG0000002
0000020458/ENSMUSG00000032064/ENSMUSG00000031785/ENSMUSG00000026259/ENSMUSG00000056758/ENSMUSG00000004040/ENSMUSG00000032340/ENSMUSG00000029050/ENSMUSG0000002
0000020484/ENSMUSG00000032340/ENSMUSG00000029050/ENSMUSG00000027996/ENSMUSG00000038400/ENSMUSG00000048616/ENSMUSG00000007613/ENSMUSG00000030530/ENSMUSG0000002
0000039004/ENSMUSG00000039286/ENSMUSG00000021756/ENSMUSG00000029050/ENSMUSG00000027996/ENSMUSG00000048616/ENSMUSG00000056501/ENSMUSG00000024098/ENSMUSG0000002
0000022346/ENSMUSG00000000567/ENSMUSG00000040856/ENSMUSG00000000753/ENSMUSG00000031565/ENSMUSG00000022367/ENSMUSG00000046186/ENSMUSG00000020458/ENSMUSG0000003
0000039004/ENSMUSG00000029287/ENSMUSG00000020484/ENSMUSG00000032340/ENSMUSG00000029050/ENSMUSG00000027996/ENSMUSG00000023845/ENSMUSG00000031849/ENSMUSG0000003
0000032244/ENSMUSG00000027947/ENSMUSG00000021540/ENSMUSG00000030849/ENSMUSG00000015846/ENSMUSG00000039004/ENSMUSG00000035342/ENSMUSG00000034640/ENSMUSG0000007
0000037465/ENSMUSG00000031849/ENSMUSG00000039662/ENSMUSG00000035673/ENSMUSG00000056050/ENSMUSG00000032487/ENSMUSG00000026836/ENSMUSG00000019943/ENSMUSG0000007
0000034201
0000056501/ENSMUSG00000024927/ENSMUSG00000023951/ENSMUSG00000034154/ENSMUSG00000024423/ENSMUSG00000036450/ENSMUSG00000027230/ENSMUSG00000015839/ENSMUSG0000002
0000056501/ENSMUSG00000023951/ENSMUSG00000034154/ENSMUSG00000024423/ENSMUSG00000036450/ENSMUSG00000027230/ENSMUSG00000015839/ENSMUSG00000025612/ENSMUSG0000002
0000020458/ENSMUSG00000030536/ENSMUSG00000023067/ENSMUSG00000027678/ENSMUSG00000038708/ENSMUSG00000049800/ENSMUSG00000028893/ENSMUSG00000019970/ENSMUSG0000002
0000048126/ENSMUSG00000056050/ENSMUSG00000001119
0000027678/ENSMUSG00000030849/ENSMUSG00000015846/ENSMUSG00000056758/ENSMUSG00000037242/ENSMUSG00000027996/ENSMUSG00000048616/ENSMUSG00000005087/ENSMUSG0000002

0000019970/ENSMUSG00000004040/ENSMUSG00000032715/ENSMUSG00000020484/ENSMUSG00000018736/ENSMUSG00000023034/ENSMUSG00000033863/ENSMUSG00000024789/ENSMUSG0000002
0000020458/ENSMUSG00000027678/ENSMUSG00000030849/ENSMUSG00000015846/ENSMUSG00000056758/ENSMUSG00000037242/ENSMUSG00000027996/ENSMUSG00000048616/ENSMUSG0000000
0000009376/ENSMUSG00000027954/ENSMUSG00000027351/ENSMUSG00000023951/ENSMUSG00000023068/ENSMUSG00000013076/ENSMUSG00000006445/ENSMUSG00000050965/ENSMUSG0000000
0000038418/ENSMUSG00000031565/ENSMUSG00000027947/ENSMUSG00000023067/ENSMUSG00000030849/ENSMUSG00000015846/ENSMUSG00000031778/ENSMUSG00000029287/ENSMUSG0000000
0000023067/ENSMUSG00000022463/ENSMUSG00000031668/ENSMUSG00000028893/ENSMUSG00000039004/ENSMUSG00000040463/ENSMUSG00000037465/ENSMUSG00000034354/ENSMUSG0000002
0000038418/ENSMUSG00000027947/ENSMUSG00000023067/ENSMUSG00000030849/ENSMUSG00000031778/ENSMUSG00000001419/ENSMUSG00000024789/ENSMUSG00000005533/ENSMUSG0000002
0000039952
0000027947/ENSMUSG00000030536/ENSMUSG00000021136/ENSMUSG00000031785/ENSMUSG00000031778/ENSMUSG00000022508/ENSMUSG00000020484/ENSMUSG00000021756/ENSMUSG0000000
0000038418/ENSMUSG00000027947/ENSMUSG00000023067/ENSMUSG00000030849/ENSMUSG00000031778/ENSMUSG00000001419/ENSMUSG00000024789/ENSMUSG00000005533/ENSMUSG0000002
0000026147/ENSMUSG00000028047/ENSMUSG00000001119
0000027351/ENSMUSG00000020484/ENSMUSG00000034765/ENSMUSG00000027996/ENSMUSG00000005533/ENSMUSG00000048234/ENSMUSG00000039384/ENSMUSG00000020745/ENSMUSG0000002
0000027678/ENSMUSG00000030849/ENSMUSG00000015846/ENSMUSG00000020484/ENSMUSG00000048616/ENSMUSG00000005533/ENSMUSG00000056501/ENSMUSG00000005087/ENSMUSG0000002
0000022443/ENSMUSG00000050965/ENSMUSG00000003814/ENSMUSG00000029816
0000026131/ENSMUSG00000034201/ENSMUSG00000034593
0000038418/ENSMUSG00000027947/ENSMUSG00000050335/ENSMUSG00000020458/ENSMUSG00000056758/ENSMUSG00000031778/ENSMUSG00000004040/ENSMUSG00000031714/ENSMUSG0000002
0000022367/ENSMUSG00000009376/ENSMUSG00000020458/ENSMUSG00000022425/ENSMUSG00000023951/ENSMUSG00000023068/ENSMUSG00000013076/ENSMUSG00000050965/ENSMUSG0000002
0000056050/ENSMUSG00000032487/ENSMUSG00000074934
0000038400/ENSMUSG00000048616/ENSMUSG00000024098/ENSMUSG00000046058/ENSMUSG00000061436/ENSMUSG00000048661/ENSMUSG00000000058/ENSMUSG00000026836/ENSMUSG0000003
0000037331/ENSMUSG00000042644/ENSMUSG00000028893/ENSMUSG00000019970/ENSMUSG00000028525/ENSMUSG00000004040/ENSMUSG00000032715/ENSMUSG00000020484/ENSMUSG0000001
0000020484/ENSMUSG00000029050/ENSMUSG00000027996/ENSMUSG00000038400/ENSMUSG00000048616/ENSMUSG00000024098/ENSMUSG00000046058/ENSMUSG00000061436/ENSMUSG0000004
0000030849/ENSMUSG00000031778/ENSMUSG00000001419/ENSMUSG00000024789/ENSMUSG00000005533/ENSMUSG00000023951/ENSMUSG00000037820/ENSMUSG00000050965/ENSMUSG0000002
0000027351/ENSMUSG00000068739/ENSMUSG00000006445/ENSMUSG00000027230/ENSMUSG00000031393/ENSMUSG00000042903
0000029287/ENSMUSG00000020484/ENSMUSG00000029050/ENSMUSG00000038400/ENSMUSG00000007613/ENSMUSG00000005533/ENSMUSG00000030530/ENSMUSG00000024098/ENSMUSG0000002
0000020218/ENSMUSG00000009376/ENSMUSG00000025408/ENSMUSG00000026596/ENSMUSG00000030849/ENSMUSG00000032064/ENSMUSG00000056758/ENSMUSG00000024811/ENSMUSG0000002
0000019997/ENSMUSG00000032340/ENSMUSG00000031849/ENSMUSG00000005125/ENSMUSG00000022817/ENSMUSG00000037815/ENSMUSG00000040511/ENSMUSG00000022443/ENSMUSG0000005
0000020218/ENSMUSG00000009376/ENSMUSG00000025408/ENSMUSG00000026596/ENSMUSG00000030849/ENSMUSG00000032064/ENSMUSG00000056758/ENSMUSG00000024811/ENSMUSG0000002
0000005533/ENSMUSG00000005087/ENSMUSG00000046223/ENSMUSG00000021379/ENSMUSG00000031548/ENSMUSG00000025326/ENSMUSG00000004207/ENSMUSG00000005871
0000029201
0000037071/ENSMUSG00000021273/ENSMUSG00000031604/ENSMUSG00000033161/ENSMUSG00000038615/ENSMUSG00000022463/ENSMUSG00000093930/ENSMUSG00000022351/ENSMUSG0000004
0000068739/ENSMUSG00000006445/ENSMUSG00000027230/ENSMUSG00000031393/ENSMUSG00000042903
0000009376/ENSMUSG00000027678/ENSMUSG00000030849/ENSMUSG00000031785/ENSMUSG00000015846/ENSMUSG00000056758/ENSMUSG00000039004/ENSMUSG00000034926/ENSMUSG0000003
0000029287/ENSMUSG00000020484/ENSMUSG00000029050/ENSMUSG00000038400/ENSMUSG00000007613/ENSMUSG00000005533/ENSMUSG00000030530/ENSMUSG00000024098/ENSMUSG0000002
0000028063/ENSMUSG00000030849/ENSMUSG00000029467/ENSMUSG00000015846/ENSMUSG00000050567/ENSMUSG00000030557/ENSMUSG00000020484/ENSMUSG00000032340/ENSMUSG0000002
0000027351/ENSMUSG00000068739/ENSMUSG00000006445/ENSMUSG00000027230/ENSMUSG00000031393/ENSMUSG00000042903
0000009376/ENSMUSG00000027678/ENSMUSG00000030849/ENSMUSG00000031785/ENSMUSG00000015846/ENSMUSG00000056758/ENSMUSG00000039004/ENSMUSG00000034926/ENSMUSG0000003
0000024098/ENSMUSG00000026313/ENSMUSG00000031548/ENSMUSG00000074934/ENSMUSG00000046791

20484/ENSMUSG00000031451/ENSMUSG00000031849/ENSMUSG00000048616/ENSMUSG00000005087/ENSMUSG00000039384/ENSMUSG00000042594/ENSMUSG00000022443/ENSMUSG00000050965/E
20889/ENSMUSG00000048616/ENSMUSG00000033323/ENSMUSG00000026728/ENSMUSG00000049792/ENSMUSG00000023951/ENSMUSG00000022752/ENSMUSG00000039384/ENSMUSG00000020745/E
24098/ENSMUSG00000046058/ENSMUSG00000061436/ENSMUSG00000048661/ENSMUSG00000000058/ENSMUSG00000026836/ENSMUSG00000031548/ENSMUSG00000044857/ENSMUSG00000085795/E
23951/ENSMUSG00000026313/ENSMUSG00000026836/ENSMUSG00000029249/ENSMUSG00000031548/ENSMUSG00000074934/ENSMUSG00000046791/ENSMUSG00000005871
30849/ENSMUSG00000056758/ENSMUSG00000039004/ENSMUSG00000029287/ENSMUSG00000004040/ENSMUSG00000020484/ENSMUSG00000023034/ENSMUSG00000033863/ENSMUSG00000027996/E
38400/ENSMUSG00000048616/ENSMUSG00000007613/ENSMUSG00000026728/ENSMUSG00000030530/ENSMUSG00000024098/ENSMUSG00000022817/ENSMUSG00000042377/ENSMUSG00000046058/E
70645/ENSMUSG00000048616/ENSMUSG00000007613/ENSMUSG00000005533/ENSMUSG00000005087/ENSMUSG00000046223/ENSMUSG00000023951/ENSMUSG00000021379/ENSMUSG00000026836/E
74934

25612/ENSMUSG00000026628
26628
28641/ENSMUSG00000002489/ENSMUSG00000022508/ENSMUSG00000026014/ENSMUSG00000020484/ENSMUSG00000018736/ENSMUSG00000027996/ENSMUSG00000026361/ENSMUSG00000007613/E

23951/ENSMUSG00000037820/ENSMUSG00000006445/ENSMUSG00000039952/ENSMUSG00000026836/ENSMUSG00000031548/ENSMUSG00000074934/ENSMUSG00000041702

24413/ENSMUSG00000070645/ENSMUSG00000020889/ENSMUSG00000005533/ENSMUSG00000024927/ENSMUSG00000048490/ENSMUSG00000040021/ENSMUSG00000039202/ENSMUSG00000028042/E
05087/ENSMUSG00000023951/ENSMUSG00000037820/ENSMUSG00000006445/ENSMUSG00000039952/ENSMUSG00000026836/ENSMUSG00000031548/ENSMUSG00000031393/ENSMUSG00000074934/E
03814/ENSMUSG00000032487/ENSMUSG00000015839/ENSMUSG00000031393
04040/ENSMUSG00000001419/ENSMUSG00000024789/ENSMUSG00000048616/ENSMUSG00000007613/ENSMUSG00000005533/ENSMUSG00000023951/ENSMUSG00000037820/ENSMUSG00000050965/E
20484/ENSMUSG00000029752/ENSMUSG00000031451/ENSMUSG00000027996/ENSMUSG00000045983/ENSMUSG00000024423/ENSMUSG00000039952/ENSMUSG00000034201/ENSMUSG00000035992/E
23951/ENSMUSG00000037820/ENSMUSG00000050965/ENSMUSG00000024486/ENSMUSG00000026313/ENSMUSG00000036564/ENSMUSG00000032487/ENSMUSG00000027665

07817/ENSMUSG00000024789/ENSMUSG00000022425/ENSMUSG00000005087/ENSMUSG00000024927/ENSMUSG00000023951/ENSMUSG00000000134/ENSMUSG00000039384/ENSMUSG00000037820/E
23951/ENSMUSG00000037820/ENSMUSG00000050965/ENSMUSG00000024486/ENSMUSG00000026313/ENSMUSG00000036564/ENSMUSG00000032487/ENSMUSG00000027665

27177/ENSMUSG00000042594/ENSMUSG00000020044/ENSMUSG00000109511/ENSMUSG00000050965/ENSMUSG00000015202/ENSMUSG00000053716/ENSMUSG00000039952/ENSMUSG00000044167/E
24098/ENSMUSG00000037820/ENSMUSG00000006445/ENSMUSG00000021379/ENSMUSG00000039952/ENSMUSG00000031548/ENSMUSG00000024431/ENSMUSG00000041702

20484/ENSMUSG00000027996/ENSMUSG00000030341/ENSMUSG00000023951/ENSMUSG00000061436/ENSMUSG00000050965/ENSMUSG00000032487/ENSMUSG00000062232/ENSMUSG00000028179/E
24486/ENSMUSG00000003814/ENSMUSG00000032487/ENSMUSG00000015839/ENSMUSG00000044147

31548/ENSMUSG00000044857/ENSMUSG00000063382/ENSMUSG00000074934
18736/ENSMUSG00000023034/ENSMUSG00000024789/ENSMUSG00000026728/ENSMUSG00000005533/ENSMUSG00000056501/ENSMUSG00000024927/ENSMUSG00000037815/ENSMUSG00000028042/E
48661/ENSMUSG00000000058/ENSMUSG00000027230/ENSMUSG00000031548/ENSMUSG00000044857/ENSMUSG00000063382/ENSMUSG00000074934
24486/ENSMUSG00000026313/ENSMUSG00000032487/ENSMUSG00000027665

22817/ENSMUSG00000046058/ENSMUSG00000061436/ENSMUSG00000048661/ENSMUSG00000000058/ENSMUSG00000026836/ENSMUSG00000024431/ENSMUSG00000085795/ENSMUSG00000063382/E
20271/ENSMUSG00000035342/ENSMUSG00000002489/ENSMUSG00000027996/ENSMUSG00000048616/ENSMUSG00000026361/ENSMUSG00000081683/ENSMUSG00000005087/ENSMUSG00000040021/E
50965/ENSMUSG00000003814/ENSMUSG00000029816/ENSMUSG00000026836/ENSMUSG00000045394
20271/ENSMUSG00000035342/ENSMUSG00000002489/ENSMUSG00000027996/ENSMUSG00000048616/ENSMUSG00000026361/ENSMUSG00000081683/ENSMUSG00000005087/ENSMUSG00000040021/E

45294/ENSMUSG00000039004/ENSMUSG00000026675/ENSMUSG00000034926/ENSMUSG00000024413/ENSMUSG00000034640/ENSMUSG00000020889/ENSMUSG00000005533/ENSMUSG00000032018/E

31714/ENSMUSG00000034640/ENSMUSG00000027996/ENSMUSG00000070645/ENSMUSG00000048616/ENSMUSG00000007613/ENSMUSG00000005533/ENSMUSG00000056501/ENSMUSG00000005087/E
22817/ENSMUSG00000046058/ENSMUSG00000061436/ENSMUSG00000048661/ENSMUSG00000000058/ENSMUSG00000026836/ENSMUSG00000024431/ENSMUSG00000085795/ENSMUSG00000063382/E
29050/ENSMUSG00000031849/ENSMUSG00000033323/ENSMUSG00000023951/ENSMUSG00000022752/ENSMUSG00000096054/ENSMUSG00000002052/ENSMUSG00000018476/ENSMUSG00000022443/E

31714/ENSMUSG00000034640/ENSMUSG00000027996/ENSMUSG00000070645/ENSMUSG00000048616/ENSMUSG00000007613/ENSMUSG00000005533/ENSMUSG00000056501/ENSMUSG00000005087/E

ENSMUSG00000030168/ENSMUSG00000032702/ENSMUSG00000024486/ENSMUSG00000008398/ENSMUSG00000056050/ENSMUSG00000026131/ENSMUSG00000033717/ENSMUSG00000015839/ENSMUS
ENSMUSG00000037815/ENSMUSG00000024812/ENSMUSG00000054808/ENSMUSG00000028161/ENSMUSG00000032702/ENSMUSG00000021379/ENSMUSG00000040945/ENSMUSG00000032593/ENSMUS
ENSMUSG00000063382/ENSMUSG00000074934/ENSMUSG00000024182

ENSMUSG00000020889/ENSMUSG00000048616/ENSMUSG00000026361/ENSMUSG00000007613/ENSMUSG00000056501/ENSMUSG00000023951/ENSMUSG00000039384/ENSMUSG00000047281/ENSMUS
ENSMUSG00000061436/ENSMUSG00000048661/ENSMUSG00000000058/ENSMUSG00000026836/ENSMUSG00000031548/ENSMUSG00000044857/ENSMUSG00000085795/ENSMUSG00000063382/ENSMUS
ENSMUSG00000031548/ENSMUSG00000025326/ENSMUSG00000074934/ENSMUSG00000026812/ENSMUSG00000004207/ENSMUSG00000045394/ENSMUSG00000005871/ENSMUSG00000032220

ENSMUSG00000033323/ENSMUSG00000045983/ENSMUSG00000005087/ENSMUSG00000023951/ENSMUSG00000022752/ENSMUSG00000020745/ENSMUSG00000021978/ENSMUSG00000047281/ENSMUS

ENSMUSG00000048249/ENSMUSG00000050965/ENSMUSG00000030168/ENSMUSG00000032702/ENSMUSG00000018634/ENSMUSG00000003814/ENSMUSG00000018965/ENSMUSG00000000058/ENSMUS
ENSMUSG00000041702

ENSMUSG00000024486/ENSMUSG00000026313/ENSMUSG00000036564/ENSMUSG00000032487/ENSMUSG00000027665/ENSMUSG00000005871
ENSMUSG00000044167/ENSMUSG00000029361/ENSMUSG00000015839/ENSMUSG00000063229/ENSMUSG00000028646/ENSMUSG00000026826/ENSMUSG00000050697/ENSMUSG00000026812/ENSMUS

ENSMUSG00000028042/ENSMUSG00000028161/ENSMUSG00000050965/ENSMUSG00000003814/ENSMUSG00000039952/ENSMUSG00000035356/ENSMUSG00000033454/ENSMUSG00000037902/ENSMUS

ENSMUSG00000031548/ENSMUSG00000039844/ENSMUSG00000044857/ENSMUSG00000037902/ENSMUSG00000026628/ENSMUSG00000005871

ENSMUSG00000015839/ENSMUSG00000074934

ENSMUSG00000024423/ENSMUSG00000050965/ENSMUSG00000032702/ENSMUSG00000018634/ENSMUSG00000079037/ENSMUSG00000062232/ENSMUSG00000000058/ENSMUSG00000038859/ENSMUS

ENSMUSG00000024182
ENSMUSG00000013076/ENSMUSG00000018583/ENSMUSG00000032717/ENSMUSG00000032702/ENSMUSG00000073563/ENSMUSG00000044167/ENSMUSG00000068740/ENSMUSG00000031548/ENSMUS

ENSMUSG00000013076/ENSMUSG00000018583/ENSMUSG00000032717/ENSMUSG00000032702/ENSMUSG00000073563/ENSMUSG00000044167/ENSMUSG00000068740/ENSMUSG00000031548/ENSMUS

ENSMUSG00000030747/ENSMUSG00000063142/ENSMUSG00000033105/ENSMUSG00000024924/ENSMUSG00000040268/ENSMUSG00000018965/ENSMUSG00000029249/ENSMUSG00000024431/ENSMUS

ENSMUSG00000035247/ENSMUSG00000045216/ENSMUSG00000046223/ENSMUSG00000023951/ENSMUSG00000048490/ENSMUSG00000030059/ENSMUSG00000074221/ENSMUSG00000079509/ENSMUS
ENSMUSG00000024182
ENSMUSG00000028161/ENSMUSG00000026313/ENSMUSG00000000058/ENSMUSG00000026836/ENSMUSG00000028179/ENSMUSG00000024431/ENSMUSG00000031393/ENSMUSG00000042903/ENSMUS

ENSMUSG00000035247/ENSMUSG00000045216/ENSMUSG00000046223/ENSMUSG00000023951/ENSMUSG00000048490/ENSMUSG00000030059/ENSMUSG00000074221/ENSMUSG00000079509/ENSMUS

G00000026797/ENSMUSG00000056071/ENSMUSG00000027712
G00000003814/ENSMUSG00000062232/ENSMUSG00000018965/ENSMUSG00000038539/ENSMUSG00000029249/ENSMUSG00000044147/ENSMUSG00000031393/ENSMUSG00000025326/ENSMUSG00000

G00000006445/ENSMUSG00000050965/ENSMUSG00000000058/ENSMUSG00000034205/ENSMUSG00000000184/ENSMUSG00000031548/ENSMUSG00000085795/ENSMUSG00000005871
G00000074934/ENSMUSG00000024182

G00000039953/ENSMUSG00000034154/ENSMUSG00000024423/ENSMUSG00000030168/ENSMUSG00000024486/ENSMUSG00000020132/ENSMUSG00000095098/ENSMUSG00000034201/ENSMUSG00000

G00000034201/ENSMUSG00000038859/ENSMUSG00000024052/ENSMUSG00000033717/ENSMUSG00000029249/ENSMUSG00000044167/ENSMUSG00000031548/ENSMUSG00000020091/ENSMUSG00000

G00000026628

G00000026812/ENSMUSG00000005871

G00000024052/ENSMUSG00000044167/ENSMUSG00000029361/ENSMUSG00000031548/ENSMUSG00000020091/ENSMUSG00000001119/ENSMUSG00000028646/ENSMUSG00000039844/ENSMUSG00000

G00000039844/ENSMUSG00000036432/ENSMUSG00000085795/ENSMUSG00000063382/ENSMUSG00000074934/ENSMUSG00000050697/ENSMUSG00000003345/ENSMUSG00000024182/ENSMUSG00000

G00000039844/ENSMUSG00000036432/ENSMUSG00000085795/ENSMUSG00000063382/ENSMUSG00000074934/ENSMUSG00000050697/ENSMUSG00000003345/ENSMUSG00000024182/ENSMUSG00000

G00000031835/ENSMUSG00000031393/ENSMUSG00000050697

G00000024073/ENSMUSG00000027832/ENSMUSG00000027465/ENSMUSG00000032717/ENSMUSG00000021379/ENSMUSG00000074656/ENSMUSG00000040268/ENSMUSG00000032487/ENSMUSG00000

G00000074934/ENSMUSG00000026812

G00000024073/ENSMUSG00000027832/ENSMUSG00000027465/ENSMUSG00000032717/ENSMUSG00000021379/ENSMUSG00000074656/ENSMUSG00000040268/ENSMUSG00000032487/ENSMUSG00000

)026812

)028179/ENSMUSG00000031548/ENSMUSG00000024431/ENSMUSG00000031393/ENSMUSG00000074934/ENSMUSG00000002083

)024431/ENSMUSG00000042903/ENSMUSG00000019943/ENSMUSG00000085795/ENSMUSG00000025326/ENSMUSG00000026826/ENSMUSG00000028661/ENSMUSG00000050697/ENSMUSG0000000588

)031393/ENSMUSG00000042903/ENSMUSG00000030287/ENSMUSG00000019943/ENSMUSG00000026826/ENSMUSG00000050697/ENSMUSG00000026812/ENSMUSG00000027712/ENSMUSG0000000587

)005871

)005871

)031548/ENSMUSG00000025326/ENSMUSG00000004207/ENSMUSG00000005871

)031548/ENSMUSG00000025326/ENSMUSG00000004207/ENSMUSG00000005871

6/ENSMUSG00000026812/ENSMUSG00000027712/ENSMUSG00000005871/ENSMUSG00000034593

1/ENSMUSG00000034593

| Category | GOID | Description | GeneRatio | BgRatio | pvalue | padj | geneID | geneName |
|---|---|---|---|---|---|---|---|---|
| BP | GO:004875 | branching morphogenesis of an epithelial tube | 35\|1209 | 176\|16097 | 8.78E-08 | 5.40E-06 | ENSMUS | Col4a1/Spry2/Myc/Sox9/Met/Mmp14/Ncoa3/Fgfr2/Hmga2/Tnc |
| BP | GO:003526 | organ growth | 38\|1209 | 200\|16097 | 8.80E-08 | 5.40E-06 | ENSMUS | Spry2/Thbs1/Col14a1/Rgs2/Sox9/Fgfr1/Col12a1/Sorbs2/Fgfr2/F |
| BP | GO:190511 | cell surface receptor signaling pathway involved in cell-cell sign | 69\|1209 | 480\|16097 | 1.21E-07 | 7.37E-06 | ENSMUS | Col1a1/Sulf2/Myc/Tlr2/Sox9/Klf4/Egr1/Wif1/Met/Ddit3/Abl2/F |
| BP | GO:005109 | positive regulation of binding | 37\|1209 | 195\|16097 | 1.34E-07 | 8.07E-06 | ENSMUS | Myc/Klf4/Hipk1/S100a10/Met/Cald1/Lgals3/Eif2ak3/Abl2/Tiam |
| MF | GO:003024 | carbohydrate binding | 41\|1190 | 229\|15912 | 1.35E-07 | 1.23E-05 | ENSMUS | Prg4/Crybg3/C4b/Lgals3/Lgals1/Vcan/Clec3b/Cd93/Cd34/P3h1 |
| BP | GO:001810 | peptidyl-tyrosine phosphorylation | 49\|1209 | 299\|16097 | 1.70E-07 | 1.01E-05 | ENSMUS | Errfi1/Itga5/Fgfr1/Il6ra/Efna1/Iqgap1/Pecam1/Abl2/Fgfr2/Socs3 |
| CC | GO:004512 | membrane raft | 54\|1232 | 342\|16313 | 1.70E-07 | 9.36E-06 | ENSMUS | Hspa1a/Hmox1/Ldlr/Tlr2/Rgs16/Cd14/Has2/S100a10/Pacsin2/S |
| CC | GO:000172 | stress fiber | 22\|1232 | 84\|16313 | 1.72E-07 | 9.36E-06 | ENSMUS | Acta1/Ablim3/Mylk/Synpo/Myh11/Dlc1/Ldb3/Tlnrd1/Acta2/Ac |
| CC | GO:009751 | contractile actin filament bundle | 22\|1232 | 84\|16313 | 1.72E-07 | 9.36E-06 | ENSMUS | Acta1/Ablim3/Mylk/Synpo/Myh11/Dlc1/Ldb3/Tlnrd1/Acta2/Ac |
| BP | GO:005076 | negative regulation of neurogenesis | 51\|1209 | 317\|16097 | 1.73E-07 | 1.02E-05 | ENSMUS | Col3a1/Ldlr/Sox9/Xylt1/Ifrd1/Efna1/Rtn4/Lgals1/Dixdc1/Adgrg |
| CC | GO:009858 | membrane region | 55\|1232 | 352\|16313 | 1.87E-07 | 9.36E-06 | ENSMUS | Hspa1a/Hmox1/Ldlr/Tlr2/Rgs16/Cd14/Has2/S100a10/Pacsin2/S |
| CC | GO:009885 | membrane microdomain | 54\|1232 | 343\|16313 | 1.88E-07 | 9.36E-06 | ENSMUS | Hspa1a/Hmox1/Ldlr/Tlr2/Rgs16/Cd14/Has2/S100a10/Pacsin2/S |
| BP | GO:005254 | regulation of peptidase activity | 54\|1209 | 345\|16097 | 1.90E-07 | 1.10E-05 | ENSMUS | Hspa1b/F3/Thbs1/Myc/Serpine1/Klf4/Herpud1/Serpinf1/A2m/C |
| BP | GO:009004 | regulation of cell migration involved in sprouting angiogenesis | 14\|1209 | 37\|16097 | 1.99E-07 | 1.15E-05 | ENSMUS | Thbs1/Hmox1/Klf4/Spred1/Meox2/Nrp1/Pik3r2/Vegfa/Notch1/ |
| BP | GO:000341 | growth plate cartilage development | 13\|1209 | 32\|16097 | 2.04E-07 | 1.16E-05 | ENSMUS | Thbs1/Col14a1/Col12a1/Col6a2/Matn3/Comp/Col6a3/Mmp13 |
| BP | GO:005067 | regulation of epithelial cell proliferation | 53\|1209 | 337\|16097 | 2.11E-07 | 1.19E-05 | ENSMUS | Errfi1/Sulf2/F3/Thbs1/Hmox1/Myc/Sox9/Dlk1/Tnmd/Serpinf1/F |
| BP | GO:004353 | positive regulation of blood vessel endothelial cell migration | 17\|1209 | 54\|16097 | 2.25E-07 | 1.26E-05 | ENSMUS | Thbs1/Hmox1/Map2k3/Fgfr1/Nrp1/Igf1/Vegfa/Nus1/Amotl1/Prl |
| BP | GO:001821 | peptidyl-tyrosine modification | 49\|1209 | 302\|16097 | 2.32E-07 | 1.27E-05 | ENSMUS | Errfi1/Itga5/Fgfr1/Il6ra/Efna1/Iqgap1/Pecam1/Abl2/Fgfr2/Socs3 |
| BP | GO:005196 | negative regulation of nervous system development | 53\|1209 | 338\|16097 | 2.32E-07 | 1.27E-05 | ENSMUS | Col3a1/Ldlr/Tlr2/Sox9/Xylt1/Ifrd1/Efna1/Rtn4/Lgals1/Eif2ak3/D |
| MF | GO:000202 | protease binding | 28\|1190 | 129\|15912 | 2.35E-07 | 1.94E-05 | ENSMUS | Col3a1/Col1a1/Col1a2/F3/Hspa1a/Ldlr/Serpine1/A2m/Cst3/Tim |
| BP | GO:005065 | chondroitin sulfate proteoglycan biosynthetic process | 10\|1209 | 19\|16097 | 2.72E-07 | 1.47E-05 | ENSMUS | Xylt1/Csgalnact2/Chpf/Chst11/Cytl1/Csgalnact1/Pxylp1/Slc35d |
| CC | GO:000590 | caveola | 23\|1232 | 93\|16313 | 2.82E-07 | 1.30E-05 | ENSMUS | Hmox1/Ldlr/Pacsin2/Slc2a1/Atp1a1/Cavin2/Lrp8/Jak2/Dlc1/Hk |
| BP | GO:003011 | regulation of Wnt signaling pathway | 46\|1209 | 278\|16097 | 3.03E-07 | 1.62E-05 | ENSMUS | Col1a1/Sulf2/Tlr2/Sox9/Egr1/Wif1/Ddit3/Abl2/Fgfr2/Dixdc1/Hr |
| BP | GO:004206 | gliogenesis | 49\|1209 | 305\|16097 | 3.15E-07 | 1.67E-05 | ENSMUS | Ldlr/Myc/Tlr2/Sox9/Lamb1/Rtn4/Vcan/Nfe2l1/Abl2/Adgrg1/Hm |
| BP | GO:003166 | cellular response to extracellular stimulus | 38\|1209 | 210\|16097 | 3.26E-07 | 1.71E-05 | ENSMUS | Fosl1/Postn/Lpl/Slc2a1/Atf4/Jun/Cdkn1a/Srebf2/Eif2ak3/Fos/Se |
| BP | GO:004544 | endothelial cell differentiation | 24\|1209 | 102\|16097 | 3.82E-07 | 1.98E-05 | ENSMUS | Tnmd/Met/Cdh5/Rtn4/Pecam1/Col18a1/Bmp6/Fasn/Clic4/Tnfr |
| MF | GO:000377 | actin binding | 60\|1190 | 411\|15912 | 4.12E-07 | 3.11E-05 | ENSMUS | Plec/Marcks/Cald1/Ablim3/Fxyd5/Tpm1/Tpm2/Epb41l1/Dixdc1 |
| BP | GO:004259 | response to starvation | 31\|1209 | 155\|16097 | 4.15E-07 | 2.14E-05 | ENSMUS | Slc2a1/Atf4/Jun/Eif4ebp1/Larp1/Cdkn1a/Srebf2/Eif2ak3/Ddit3/ |
| BP | GO:000341 | endochondral bone growth | 14\|1209 | 39\|16097 | 4.27E-07 | 2.18E-05 | ENSMUS | Thbs1/Col14a1/Col12a1/Col6a2/Matn3/Comp/Col6a3/Mmp13 |
| BP | GO:200123 | negative regulation of apoptotic signaling pathway | 39\|1209 | 221\|16097 | 4.48E-07 | 2.26E-05 | ENSMUS | Hmox1/Serpine1/Herpud1/Lgals3/Ier3/Lmna/Hyou1/Cx3cl1/Xb |
| BP | GO:000157 | vasculogenesis | 22\|1209 | 89\|16097 | 4.67E-07 | 2.34E-05 | ENSMUS | Fgfr1/Has2/Rtn4/Spred1/Fgfr2/Cd34/Tgfbr3/Zmiz1/Nrp1/Tiparp |
| BP | GO:007037 | ERK1 and ERK2 cascade | 47\|1209 | 292\|16097 | 5.16E-07 | 2.56E-05 | ENSMUS | Errfi1/Spry2/Hmgcr/Myc/Tlr2/Sox9/Klf4/Dusp1/Jun/Akap12/Sp |
| BP | GO:004544 | fat cell differentiation | 40\|1209 | 231\|16097 | 5.34E-07 | 2.59E-05 | ENSMUS | Rgs2/Dlk1/Klf4/Wif1/Lpl/Scd1/Noct/Eif2ak3/Ddit3/Insig1/Fndc |
| BP | GO:000698 | response to unfolded protein | 25\|1209 | 111\|16097 | 5.34E-07 | 2.59E-05 | ENSMUS | Hspa1b/Hspa1a/Thbs1/Chac1/Herpud1/Hspa9/Dnajb9/Atf4/Eif2 |
| BP | GO:000182 | kidney development | 43\|1209 | 257\|16097 | 5.37E-07 | 2.59E-05 | ENSMUS | Sulf2/Myc/Sox9/Nid1/Egr1/Fgfr1/Cdkn1c/Il6ra/Vcan/Smad5/Fg |
| CC | GO:009864 | complex of collagen trimers | 9\|1232 | 16\|16313 | 5.46E-07 | 2.33E-05 | ENSMUS | Col3a1/Col1a1/Col1a2/Col4a1/Col5a3/Col5a2/Col4a2/Lum/Co |
| BP | GO:004576 | positive regulation of angiogenesis | 33\|1209 | 173\|16097 | 5.57E-07 | 2.67E-05 | ENSMUS | F3/Thbs1/Hmox1/Serpine1/Klf4/Itga5/Lgals3/Cdh5/Rtn4/Smoc |
| BP | GO:000155 | regulation of cell growth | 59\|1209 | 404\|16097 | 5.88E-07 | 2.79E-05 | ENSMUS | Col14a1/Igfbp5/Rgs2/Inhba/Ifrd1/Igfbp3/Tnfrsf12a/Rtn4/Cdkn1 |
| BP | GO:007037 | regulation of ERK1 and ERK2 cascade | 45\|1209 | 276\|16097 | 6.22E-07 | 2.92E-05 | ENSMUS | Errfi1/Spry2/Hmgcr/Tlr2/Klf4/Dusp1/Jun/Akap12/Spred1/Cyr61 |
| BP | GO:000204 | sprouting angiogenesis | 25\|1209 | 112\|16097 | 6.40E-07 | 2.95E-05 | ENSMUS | Thbs1/Hmox1/Klf4/Itga5/Rtn4/Spred1/Meox2/Nr4a1/Nrp1/Flt4 |
| BP | GO:001000 | glial cell differentiation | 39\|1209 | 224\|16097 | 6.40E-07 | 2.95E-05 | ENSMUS | Ldlr/Tlr2/Sox9/Rtn4/Nfe2l1/Abl2/Hmga2/Nab2/Egfr/Stat3/B4ga |
| BP | GO:000717 | transforming growth factor beta receptor signaling pathway | 31\|1209 | 158\|16097 | 6.44E-07 | 2.95E-05 | ENSMUS | Col3a1/Col1a2/Thbs1/Vasn/Cd109/Jun/Cdh5/Aspn/Smad5/Fos/ |
| BP | GO:001481 | muscle cell migration | 24\|1209 | 105\|16097 | 6.74E-07 | 3.02E-05 | ENSMUS | F3/Myc/Igfbp5/Postn/Serpine1/Trib1/Egr1/Igfbp3/Has2/Met/Iq |
| BP | GO:004217 | regulation of protein catabolic process | 55\|1209 | 368\|16097 | 6.75E-07 | 3.02E-05 | ENSMUS | Hspa1b/Hspa1a/Hmgcr/Ldlr/Sox9/Trib1/Cyp51/Herpud1/Zfand |
| BP | GO:001076 | regulation of cell morphogenesis involved in differentiation | 51\|1209 | 331\|16097 | 6.76E-07 | 3.02E-05 | ENSMUS | Postn/Ifrd1/Has2/S100a10/Tnfrsf12a/Met/Efna1/Rtn4/Golga4/D |
| BP | GO:001491 | regulation of smooth muscle cell migration | 21\|1209 | 84\|16097 | 6.92E-07 | 3.07E-05 | ENSMUS | F3/Myc/Igfbp5/Postn/Serpine1/Trib1/Egr1/Igfbp3/Has2/Iqgap1/ |
| BP | GO:001490 | smooth muscle cell migration | 22\|1209 | 91\|16097 | 7.05E-07 | 3.10E-05 | ENSMUS | F3/Myc/Igfbp5/Postn/Serpine1/Trib1/Egr1/Igfbp3/Has2/Iqgap1/ |
| MF | GO:000196 | fibronectin binding | 12\|1190 | 30\|15912 | 7.06E-07 | 4.92E-05 | ENSMUS | Thbs1/Igfbp5/Igfbp3/Igfbp6/Ctsk/Tnc/Thbs4/Ctgf/Comp/Ssc5d/ |
| BP | GO:007200 | renal system development | 45\|1209 | 278\|16097 | 7.65E-07 | 3.34E-05 | ENSMUS | Col4a1/Sulf2/Myc/Sox9/Nid1/Egr1/Fgfr1/Cdkn1c/Il6ra/Vcan/Sm |
| BP | GO:200002 | regulation of animal organ morphogenesis | 35\|1209 | 192\|16097 | 7.87E-07 | 3.40E-05 | ENSMUS | Sulf2/Myc/Sox9/Chad/Fgfr1/Met/Aspn/Rtn4/Abl2/Fgfr2/Rxra/Hr |
| BP | GO:190303 | regulation of response to wounding | 32\|1209 | 168\|16097 | 8.51E-07 | 3.61E-05 | ENSMUS | Thbs1/Hmgcr/Serpine1/Klf4/Xylt1/Tnfrsf12a/Cd109/Rtn4/Serpi |

| Count | Up | Up_Gene_i | Down | Down_Gene_id |
|---|---|---|---|---|
| 35 | 19 | ENSMUSG0 | 16 | ENSMUSG00000022114/ENSMUSG00000022346/ENSMUSG00000000567/ENSMUSG00000009376/ENSMUSG00000027678/ENSMUSG00000030849/ENSMUSG0 |
| 38 | 15 | ENSMUSG0 | 23 | ENSMUSG00000022114/ENSMUSG00000040152/ENSMUSG00000026360/ENSMUSG00000000567/ENSMUSG00000031565/ENSMUSG00000030849/ENSMUSG0 |
| 69 | 24 | ENSMUSG0 | 45 | ENSMUSG00000006800/ENSMUSG00000022346/ENSMUSG00000027995/ENSMUSG00000000567/ENSMUSG00000003032/ENSMUSG00000038418/ENSMUSG0 |
| 37 | 13 | ENSMUSG0 | 24 | ENSMUSG00000022346/ENSMUSG00000003032/ENSMUSG00000008730/ENSMUSG00000041959/ENSMUSG00000009376/ENSMUSG00000050335/ENSMUSG0 |
| 41 | 18 | ENSMUSG0 | 23 | ENSMUSG00000006014/ENSMUSG00000022723/ENSMUSG00000073418/ENSMUSG00000050335/ENSMUSG00000028641/ENSMUSG00000027312/ENSMUSG0 |
| 49 | 18 | ENSMUSG0 | 31 | ENSMUSG00000028967/ENSMUSG00000000555/ENSMUSG00000031565/ENSMUSG00000027947/ENSMUSG00000027954/ENSMUSG00000030536/ENSMUSG0 |
| 54 | 21 | ENSMUSG0 | 33 | ENSMUSG00000005413/ENSMUSG00000032193/ENSMUSG00000027995/ENSMUSG00000051439/ENSMUSG00000022367/ENSMUSG00000041959/ENSMUSG0 |
| 22 | 15 | ENSMUSG0 | 7 | ENSMUSG00000031523/ENSMUSG00000070462/ENSMUSG00000054808/ENSMUSG00000022443/ENSMUSG00000032643/ENSMUSG00000026131/ENSMUSG0 |
| 22 | 15 | ENSMUSG0 | 7 | ENSMUSG00000031523/ENSMUSG00000070462/ENSMUSG00000054808/ENSMUSG00000022443/ENSMUSG00000032643/ENSMUSG00000026131/ENSMUSG0 |
| 51 | 18 | ENSMUSG0 | 33 | ENSMUSG00000032193/ENSMUSG00000000567/ENSMUSG00000030657/ENSMUSG00000001627/ENSMUSG00000027954/ENSMUSG00000020458/ENSMUSG0 |
| 55 | 21 | ENSMUSG0 | 34 | ENSMUSG00000005413/ENSMUSG00000032193/ENSMUSG00000027995/ENSMUSG00000051439/ENSMUSG00000022367/ENSMUSG00000041959/ENSMUSG0 |
| 54 | 21 | ENSMUSG0 | 33 | ENSMUSG00000005413/ENSMUSG00000032193/ENSMUSG00000027995/ENSMUSG00000051439/ENSMUSG00000022367/ENSMUSG00000041959/ENSMUSG0 |
| 54 | 14 | ENSMUSG0 | 40 | ENSMUSG00000028128/ENSMUSG00000040152/ENSMUSG00000022346/ENSMUSG00000037411/ENSMUSG00000003032/ENSMUSG00000031770/ENSMUSG0 |
| 14 | 8 | ENSMUSG0 | 6 | ENSMUSG00000040152/ENSMUSG00000005413/ENSMUSG00000003032/ENSMUSG00000027351/ENSMUSG00000023951/ENSMUSG00000032487 |
| 13 | 6 | ENSMUSG0 | 7 | ENSMUSG00000040152/ENSMUSG00000020241/ENSMUSG00000031849/ENSMUSG00000048126/ENSMUSG00000026147/ENSMUSG00000028047/ENSMUSG0 |
| 53 | 18 | ENSMUSG0 | 35 | ENSMUSG00000028967/ENSMUSG00000006800/ENSMUSG00000028128/ENSMUSG00000040152/ENSMUSG00000005413/ENSMUSG00000022346/ENSMUSG0 |
| 17 | 7 | ENSMUSG0 | 10 | ENSMUSG00000040152/ENSMUSG00000005413/ENSMUSG00000018932/ENSMUSG00000031565/ENSMUSG00000023951/ENSMUSG00000023068/ENSMUSG0 |
| 49 | 18 | ENSMUSG0 | 31 | ENSMUSG00000028967/ENSMUSG00000000555/ENSMUSG00000031565/ENSMUSG00000027947/ENSMUSG00000027954/ENSMUSG00000030536/ENSMUSG0 |
| 53 | 18 | ENSMUSG0 | 35 | ENSMUSG00000032193/ENSMUSG00000027995/ENSMUSG00000000567/ENSMUSG00000030657/ENSMUSG00000001627/ENSMUSG00000027954/ENSMUSG0 |
| 28 | 7 | ENSMUSG0 | 21 | ENSMUSG00000028128/ENSMUSG00000032193/ENSMUSG00000037411/ENSMUSG00000030111/ENSMUSG00000027447/ENSMUSG00000001131/ENSMUSG0 |
| 10 | 3 | ENSMUSG0 | 7 | ENSMUSG00000030657/ENSMUSG00000042042/ENSMUSG00000032997/ENSMUSG00000036356/ENSMUSG00000043587/ENSMUSG00000028521/ENSMUSG0 |
| 23 | 6 | ENSMUSG0 | 17 | ENSMUSG00000005413/ENSMUSG00000032193/ENSMUSG00000016664/ENSMUSG00000028645/ENSMUSG00000033161/ENSMUSG00000028613/ENSMUSG0 |
| 46 | 16 | ENSMUSG0 | 30 | ENSMUSG00000006800/ENSMUSG00000027995/ENSMUSG00000000567/ENSMUSG00000038418/ENSMUSG00000020218/ENSMUSG00000025408/ENSMUSG0 |
| 49 | 16 | ENSMUSG0 | 33 | ENSMUSG00000032193/ENSMUSG00000022346/ENSMUSG00000027995/ENSMUSG00000000567/ENSMUSG00000020458/ENSMUSG00000038615/ENSMUSG0 |
| 38 | 11 | ENSMUSG0 | 27 | ENSMUSG00000024912/ENSMUSG00000028645/ENSMUSG00000042406/ENSMUSG00000023067/ENSMUSG00000022463/ENSMUSG00000031668/ENSMUSG0 |
| 24 | 9 | ENSMUSG0 | 15 | ENSMUSG00000009376/ENSMUSG00000020458/ENSMUSG00000039004/ENSMUSG00000025153/ENSMUSG00000037242/ENSMUSG00000030341/ENSMUSG0 |
| 60 | 33 | ENSMUSG0 | 27 | ENSMUSG00000022565/ENSMUSG00000009687/ENSMUSG00000027624/ENSMUSG00000032064/ENSMUSG00000021950/ENSMUSG00000055322/ENSMUSG0 |
| 31 | 8 | ENSMUSG0 | 23 | ENSMUSG00000028645/ENSMUSG00000042406/ENSMUSG00000031490/ENSMUSG00000037331/ENSMUSG00000023067/ENSMUSG00000022463/ENSMUSG0 |
| 14 | 7 | ENSMUSG0 | 7 | ENSMUSG00000040152/ENSMUSG00000020241/ENSMUSG00000031849/ENSMUSG00000048126/ENSMUSG00000026147/ENSMUSG00000028047/ENSMUSG0 |
| 39 | 6 | ENSMUSG0 | 33 | ENSMUSG00000005413/ENSMUSG00000037411/ENSMUSG00000031770/ENSMUSG00000050335/ENSMUSG00000003541/ENSMUSG00000028063/ENSMUSG0 |
| 22 | 10 | ENSMUSG0 | 12 | ENSMUSG00000031565/ENSMUSG00000022367/ENSMUSG00000020458/ENSMUSG00000027351/ENSMUSG00000030849/ENSMUSG00000029287/ENSMUSG0 |
| 47 | 18 | ENSMUSG0 | 29 | ENSMUSG00000028967/ENSMUSG00000022114/ENSMUSG00000021670/ENSMUSG00000022346/ENSMUSG00000027995/ENSMUSG00000000567/ENSMUSG0 |
| 40 | 11 | ENSMUSG0 | 29 | ENSMUSG00000026360/ENSMUSG00000040856/ENSMUSG00000003032/ENSMUSG00000020218/ENSMUSG00000037071/ENSMUSG00000023087/ENSMUSG0 |
| 25 | 5 | ENSMUSG0 | 20 | ENSMUSG00000040152/ENSMUSG00000027313/ENSMUSG00000031770/ENSMUSG00000024359/ENSMUSG00000014905/ENSMUSG00000042406/ENSMUSG0 |
| 43 | 21 | ENSMUSG0 | 22 | ENSMUSG00000006800/ENSMUSG00000022346/ENSMUSG00000000567/ENSMUSG00000038418/ENSMUSG00000031565/ENSMUSG00000027947/ENSMUSG0 |
| 9 | 9 | ENSMUSG0 | 0 | |
| 33 | 12 | ENSMUSG0 | 21 | ENSMUSG00000028128/ENSMUSG00000040152/ENSMUSG00000005413/ENSMUSG00000037411/ENSMUSG00000003032/ENSMUSG00000000555/ENSMUSG0 |
| 59 | 20 | ENSMUSG0 | 39 | ENSMUSG00000026360/ENSMUSG00000041324/ENSMUSG00000001627/ENSMUSG00000023905/ENSMUSG00000020458/ENSMUSG00000023067/ENSMUSG0 |
| 45 | 18 | ENSMUSG0 | 27 | ENSMUSG00000028967/ENSMUSG00000022114/ENSMUSG00000021670/ENSMUSG00000027995/ENSMUSG00000003032/ENSMUSG00000027351/ENSMUSG0 |
| 25 | 11 | ENSMUSG0 | 14 | ENSMUSG00000040152/ENSMUSG00000005413/ENSMUSG00000003032/ENSMUSG00000000555/ENSMUSG00000020458/ENSMUSG00000027351/ENSMUSG0 |
| 39 | 12 | ENSMUSG0 | 27 | ENSMUSG00000032193/ENSMUSG00000027995/ENSMUSG00000000567/ENSMUSG00000020458/ENSMUSG00000038615/ENSMUSG00000026596/ENSMUSG0 |
| 31 | 11 | ENSMUSG0 | 20 | ENSMUSG00000040152/ENSMUSG00000039646/ENSMUSG00000046186/ENSMUSG00000021540/ENSMUSG00000029287/ENSMUSG00000020484/ENSMUSG0 |
| 24 | 10 | ENSMUSG0 | 14 | ENSMUSG00000028128/ENSMUSG00000022346/ENSMUSG00000037411/ENSMUSG00000032501/ENSMUSG00000038418/ENSMUSG00000022367/ENSMUSG0 |
| 55 | 12 | ENSMUSG0 | 43 | ENSMUSG00000021670/ENSMUSG00000032193/ENSMUSG00000000567/ENSMUSG00000032501/ENSMUSG00000001467/ENSMUSG00000031770/ENSMUSG0 |
| 51 | 19 | ENSMUSG0 | 32 | ENSMUSG00000001627/ENSMUSG00000022367/ENSMUSG00000041959/ENSMUSG00000023905/ENSMUSG00000009376/ENSMUSG00000027954/ENSMUSG0 |
| 21 | 8 | ENSMUSG0 | 13 | ENSMUSG00000028128/ENSMUSG00000022346/ENSMUSG00000037411/ENSMUSG00000032501/ENSMUSG00000038418/ENSMUSG00000022367/ENSMUSG0 |
| 22 | 9 | ENSMUSG0 | 13 | ENSMUSG00000028128/ENSMUSG00000022346/ENSMUSG00000037411/ENSMUSG00000032501/ENSMUSG00000038418/ENSMUSG00000022367/ENSMUSG0 |
| 12 | 8 | ENSMUSG0 | 4 | ENSMUSG00000040152/ENSMUSG00000019997/ENSMUSG00000031849/ENSMUSG00000023951 |
| 45 | 23 | ENSMUSG0 | 22 | ENSMUSG00000006800/ENSMUSG00000022346/ENSMUSG00000000567/ENSMUSG00000038418/ENSMUSG00000031565/ENSMUSG00000027947/ENSMUSG0 |
| 35 | 14 | ENSMUSG0 | 21 | ENSMUSG00000006800/ENSMUSG00000022346/ENSMUSG00000000567/ENSMUSG00000039084/ENSMUSG00000031565/ENSMUSG00000009376/ENSMUSG0 |
| 32 | 9 | ENSMUSG0 | 23 | ENSMUSG00000040152/ENSMUSG00000021670/ENSMUSG00000037411/ENSMUSG00000003032/ENSMUSG00000030657/ENSMUSG00000023905/ENSMUSG0 |

0000056758/ENSMUSG00000037242/ENSMUSG00000027996/ENSMUSG00000048616/ENSMUSG00000005087/ENSMUSG00000023951/ENSMUSG00000006445/ENSMUSG00000039952/ENSMUSG0000002
0000015846/ENSMUSG00000020241/ENSMUSG00000029287/ENSMUSG00000031849/ENSMUSG00000048616/ENSMUSG00000007613/ENSMUSG00000033323/ENSMUSG00000048126/ENSMUSG0000002
0000020218/ENSMUSG00000009376/ENSMUSG00000025408/ENSMUSG00000026596/ENSMUSG00000030849/ENSMUSG00000032064/ENSMUSG00000056758/ENSMUSG00000031778/ENSMUSG0000002
0000031668/ENSMUSG00000026596/ENSMUSG00000002489/ENSMUSG00000020349/ENSMUSG00000032715/ENSMUSG00000024789/ENSMUSG00000029050/ENSMUSG00000045983/ENSMUSG0000002
0000038168/ENSMUSG00000026670/ENSMUSG00000037012/ENSMUSG00000022425/ENSMUSG00000004846/ENSMUSG00000048578/ENSMUSG00000028673/ENSMUSG00000024085/ENSMUSG0000003
0000026596/ENSMUSG00000030849/ENSMUSG00000039004/ENSMUSG00000046733/ENSMUSG00000028613/ENSMUSG00000021756/ENSMUSG00000024789/ENSMUSG00000031451/ENSMUSG0000002
0000016664/ENSMUSG00000028645/ENSMUSG00000069833/ENSMUSG00000033161/ENSMUSG00000030536/ENSMUSG00000028613/ENSMUSG00000020349/ENSMUSG00000024789/ENSMUSG0000002
0000034201
0000034201
0000032064/ENSMUSG00000031785/ENSMUSG00000026259/ENSMUSG00000056758/ENSMUSG00000004040/ENSMUSG00000032340/ENSMUSG00000029050/ENSMUSG00000020889/ENSMUSG0000004
0000016664/ENSMUSG00000028645/ENSMUSG00000069833/ENSMUSG00000033161/ENSMUSG00000030536/ENSMUSG00000028613/ENSMUSG00000020349/ENSMUSG00000024789/ENSMUSG0000002
0000016664/ENSMUSG00000028645/ENSMUSG00000069833/ENSMUSG00000033161/ENSMUSG00000030536/ENSMUSG00000028613/ENSMUSG00000020349/ENSMUSG00000024789/ENSMUSG0000002
0000000753/ENSMUSG00000030111/ENSMUSG00000046186/ENSMUSG00000027954/ENSMUSG00000027447/ENSMUSG00000031668/ENSMUSG00000021950/ENSMUSG00000001131/ENSMUSG0000000

0000001119
0000000567/ENSMUSG00000040856/ENSMUSG00000000753/ENSMUSG00000031565/ENSMUSG00000022367/ENSMUSG00000046186/ENSMUSG00000020458/ENSMUSG00000030849/ENSMUSG0000005
0000013076/ENSMUSG00000050965/ENSMUSG00000032487/ENSMUSG00000015839
0000026596/ENSMUSG00000030849/ENSMUSG00000039004/ENSMUSG00000046733/ENSMUSG00000028613/ENSMUSG00000021756/ENSMUSG00000024789/ENSMUSG00000031451/ENSMUSG0000002
0000020458/ENSMUSG00000031668/ENSMUSG00000032064/ENSMUSG00000031785/ENSMUSG00000026259/ENSMUSG00000056758/ENSMUSG00000004040/ENSMUSG00000032340/ENSMUSG0000002
0000020484/ENSMUSG00000031849/ENSMUSG00000030341/ENSMUSG00000030530/ENSMUSG00000039100/ENSMUSG00000020044/ENSMUSG00000021070/ENSMUSG00000079037/ENSMUSG0000002
0000029201
0000024789/ENSMUSG00000031523/ENSMUSG00000037012/ENSMUSG00000007613/ENSMUSG00000005533/ENSMUSG00000004044/ENSMUSG00000037815/ENSMUSG00000063142/ENSMUSG0000003
0000026596/ENSMUSG00000030849/ENSMUSG00000032064/ENSMUSG00000056758/ENSMUSG00000024811/ENSMUSG00000035342/ENSMUSG00000002489/ENSMUSG00000027996/ENSMUSG0000004
0000026596/ENSMUSG00000031785/ENSMUSG00000056758/ENSMUSG00000025402/ENSMUSG00000031778/ENSMUSG00000004040/ENSMUSG00000017929/ENSMUSG00000002489/ENSMUSG0000002
0000028893/ENSMUSG00000039004/ENSMUSG00000040463/ENSMUSG00000037465/ENSMUSG00000034354/ENSMUSG00000020484/ENSMUSG00000029752/ENSMUSG00000031451/ENSMUSG0000002
0000007613/ENSMUSG00000004846/ENSMUSG00000023951/ENSMUSG00000024812/ENSMUSG00000018476/ENSMUSG00000022016/ENSMUSG00000062232/ENSMUSG00000026836/ENSMUSG0000003
0000070462/ENSMUSG00000037679/ENSMUSG00000037815/ENSMUSG00000096054/ENSMUSG00000054808/ENSMUSG00000023008/ENSMUSG00000022443/ENSMUSG00000024533/ENSMUSG0000003
0000031668/ENSMUSG00000025408/ENSMUSG00000028893/ENSMUSG00000040463/ENSMUSG00000037465/ENSMUSG00000034354/ENSMUSG00000020484/ENSMUSG00000029752/ENSMUSG0000003
0000001119
0000032115/ENSMUSG00000031778/ENSMUSG00000020484/ENSMUSG00000027996/ENSMUSG00000048616/ENSMUSG00000007613/ENSMUSG00000049792/ENSMUSG00000005087/ENSMUSG0000002
0000007817/ENSMUSG00000034640/ENSMUSG00000044337/ENSMUSG00000006445/ENSMUSG00000062232/ENSMUSG00000032220
0000003032/ENSMUSG00000027351/ENSMUSG00000026596/ENSMUSG00000030849/ENSMUSG00000031778/ENSMUSG00000002489/ENSMUSG00000019997/ENSMUSG00000020484/ENSMUSG0000003
0000031668/ENSMUSG00000025408/ENSMUSG00000045294/ENSMUSG00000039286/ENSMUSG00000032715/ENSMUSG00000020484/ENSMUSG00000023034/ENSMUSG00000020601/ENSMUSG0000002
0000031668/ENSMUSG00000025408/ENSMUSG00000020484/ENSMUSG00000031849/ENSMUSG00000021831/ENSMUSG00000048249/ENSMUSG00000043019/ENSMUSG00000003814/ENSMUSG0000002
0000021540/ENSMUSG00000030849/ENSMUSG00000039004/ENSMUSG00000035342/ENSMUSG00000034640/ENSMUSG00000070645/ENSMUSG00000048616/ENSMUSG00000007613/ENSMUSG0000000

0000050335/ENSMUSG00000020458/ENSMUSG00000056758/ENSMUSG00000031778/ENSMUSG00000004040/ENSMUSG00000031714/ENSMUSG00000020484/ENSMUSG00000027996/ENSMUSG0000003
0000027678/ENSMUSG00000038708/ENSMUSG00000049800/ENSMUSG00000028893/ENSMUSG00000019970/ENSMUSG00000028641/ENSMUSG00000022508/ENSMUSG00000020484/ENSMUSG0000001
0000026596/ENSMUSG00000030849/ENSMUSG00000031778/ENSMUSG00000002489/ENSMUSG00000019997/ENSMUSG00000020484/ENSMUSG00000031451/ENSMUSG00000005087/ENSMUSG0000002
0000023034/ENSMUSG00000023951/ENSMUSG00000006445/ENSMUSG00000056050/ENSMUSG00000032487/ENSMUSG00000027230/ENSMUSG00000034205/ENSMUSG00000074934
0000056758/ENSMUSG00000025402/ENSMUSG00000004040/ENSMUSG00000017929/ENSMUSG00000021756/ENSMUSG00000002881/ENSMUSG00000029050/ENSMUSG00000020889/ENSMUSG0000002
0000029050/ENSMUSG00000038400/ENSMUSG00000007613/ENSMUSG00000030530/ENSMUSG00000024098/ENSMUSG00000022817/ENSMUSG00000046058/ENSMUSG00000061436/ENSMUSG0000004
0000009376/ENSMUSG00000030536/ENSMUSG00000021756/ENSMUSG00000026313/ENSMUSG00000036564/ENSMUSG00000015839/ENSMUSG00000042903/ENSMUSG00000034593
0000053581/ENSMUSG00000027954/ENSMUSG00000038615/ENSMUSG00000003541/ENSMUSG00000027447/ENSMUSG00000025373/ENSMUSG00000001131/ENSMUSG00000020349/ENSMUSG0000003
0000020458/ENSMUSG00000038708/ENSMUSG00000032064/ENSMUSG00000026259/ENSMUSG00000035671/ENSMUSG00000019970/ENSMUSG00000002489/ENSMUSG00000028613/ENSMUSG0000001
0000030536/ENSMUSG00000021756/ENSMUSG00000026313/ENSMUSG00000036564/ENSMUSG00000015839/ENSMUSG00000042903/ENSMUSG00000034593
0000030536/ENSMUSG00000021756/ENSMUSG00000026313/ENSMUSG00000036564/ENSMUSG00000015839/ENSMUSG00000042903/ENSMUSG00000034593

0000021540/ENSMUSG00000030849/ENSMUSG00000039004/ENSMUSG00000035342/ENSMUSG00000034640/ENSMUSG00000070645/ENSMUSG00000048616/ENSMUSG00000007613/ENSMUSG0000000
0000020458/ENSMUSG00000026596/ENSMUSG00000030849/ENSMUSG00000015846/ENSMUSG00000056758/ENSMUSG00000020484/ENSMUSG00000027996/ENSMUSG00000048616/ENSMUSG0000000
0000046186/ENSMUSG00000020458/ENSMUSG00000023067/ENSMUSG00000020484/ENSMUSG00000018736/ENSMUSG00000032340/ENSMUSG00000005533/ENSMUSG00000039384/ENSMUSG0000004

26836/ENSMUSG00000074934
22752/ENSMUSG00000040021/ENSMUSG00000026147/ENSMUSG00000028047/ENSMUSG00000001119/ENSMUSG00000024431/ENSMUSG00000025326/ENSMUSG00000004207/ENSMUSG00000005871
24811/ENSMUSG00000020271/ENSMUSG00000035342/ENSMUSG00000002489/ENSMUSG00000027996/ENSMUSG00000048616/ENSMUSG00000026361/ENSMUSG00000081683/ENSMUSG00000005087/E
26796/ENSMUSG00000006585/ENSMUSG00000046223/ENSMUSG00000030738/ENSMUSG00000027177/ENSMUSG00000061436/ENSMUSG00000022365/ENSMUSG00000062232/ENSMUSG00000044147/E
38296/ENSMUSG00000027832/ENSMUSG00000059991/ENSMUSG00000018906/ENSMUSG00000023175/ENSMUSG00000003814/ENSMUSG00000030835/ENSMUSG00000034205/ENSMUSG00000004609
27996/ENSMUSG00000037012/ENSMUSG00000027293/ENSMUSG00000022425/ENSMUSG00000030341/ENSMUSG00000005533/ENSMUSG00000005087/ENSMUSG00000023951/ENSMUSG00000042594/E
24413/ENSMUSG00000031523/ENSMUSG00000037012/ENSMUSG00000030341/ENSMUSG00000007613/ENSMUSG00000005533/ENSMUSG00000030530/ENSMUSG00000004044/ENSMUSG00000037815/E

48616/ENSMUSG00000026728/ENSMUSG00000049792/ENSMUSG00000039384/ENSMUSG00000020745/ENSMUSG00000037815/ENSMUSG00000028161/ENSMUSG00000032702/ENSMUSG00000021379/E
24413/ENSMUSG00000031523/ENSMUSG00000037012/ENSMUSG00000030341/ENSMUSG00000007613/ENSMUSG00000005533/ENSMUSG00000030530/ENSMUSG00000004044/ENSMUSG00000037815/E
24413/ENSMUSG00000031523/ENSMUSG00000037012/ENSMUSG00000030341/ENSMUSG00000007613/ENSMUSG00000005533/ENSMUSG00000030530/ENSMUSG00000004044/ENSMUSG00000037815/E
04040/ENSMUSG00000019997/ENSMUSG00000034926/ENSMUSG00000020349/ENSMUSG00000023034/ENSMUSG00000024789/ENSMUSG00000067780/ENSMUSG00000031451/ENSMUSG00000027996/E

56758/ENSMUSG00000039004/ENSMUSG00000029287/ENSMUSG00000004040/ENSMUSG00000020484/ENSMUSG00000023034/ENSMUSG00000033863/ENSMUSG00000027996/ENSMUSG00000020889/E

27996/ENSMUSG00000037012/ENSMUSG00000027293/ENSMUSG00000022425/ENSMUSG00000030341/ENSMUSG00000005533/ENSMUSG00000005087/ENSMUSG00000023951/ENSMUSG00000042594/E
29050/ENSMUSG00000020889/ENSMUSG00000048616/ENSMUSG00000026728/ENSMUSG00000049792/ENSMUSG00000039384/ENSMUSG00000020745/ENSMUSG00000037815/ENSMUSG00000028161/E
22365/ENSMUSG00000047126/ENSMUSG00000025209/ENSMUSG00000022074/ENSMUSG00000024807/ENSMUSG00000004207/ENSMUSG00000005871

32487/ENSMUSG00000000058/ENSMUSG00000029361
48616/ENSMUSG00000026361/ENSMUSG00000040021/ENSMUSG00000018583/ENSMUSG00000032717/ENSMUSG00000032702/ENSMUSG00000073563/ENSMUSG00000044167/ENSMUSG00000031548/E
28613/ENSMUSG00000021756/ENSMUSG00000002881/ENSMUSG00000029050/ENSMUSG00000020889/ENSMUSG00000022425/ENSMUSG00000048616/ENSMUSG00000026728/ENSMUSG00000005125/E
27996/ENSMUSG00000045983/ENSMUSG00000024423/ENSMUSG00000034201/ENSMUSG00000035992/ENSMUSG00000044167/ENSMUSG00000015839/ENSMUSG00000063229/ENSMUSG00000028646/E
39844
32643/ENSMUSG00000034154/ENSMUSG00000024423/ENSMUSG00000040852/ENSMUSG00000063142/ENSMUSG00000001630/ENSMUSG00000026131/ENSMUSG00000018965/ENSMUSG00000039952/E
31451/ENSMUSG00000024423/ENSMUSG00000034201/ENSMUSG00000035992/ENSMUSG00000044167/ENSMUSG00000015839/ENSMUSG00000028646/ENSMUSG00000050697/ENSMUSG00000026628

24927/ENSMUSG00000046223/ENSMUSG00000023951/ENSMUSG00000044337/ENSMUSG00000037815/ENSMUSG00000024073/ENSMUSG00000021070/ENSMUSG00000022037/ENSMUSG00000032487/E

31451/ENSMUSG00000005087/ENSMUSG00000023951/ENSMUSG00000044337/ENSMUSG00000039384/ENSMUSG00000006445/ENSMUSG00000020044/ENSMUSG00000050965/ENSMUSG00000036564/E
27996/ENSMUSG00000020889/ENSMUSG00000056501/ENSMUSG00000023951/ENSMUSG00000039384/ENSMUSG00000028042/ENSMUSG00000021831/ENSMUSG00000021379/ENSMUSG00000032487/E
22365/ENSMUSG00000027230/ENSMUSG00000020571/ENSMUSG00000015839/ENSMUSG00000024807/ENSMUSG00000026628
05087/ENSMUSG00000023951/ENSMUSG00000031548/ENSMUSG00000074934/ENSMUSG00000026812/ENSMUSG00000045394/ENSMUSG00000005871/ENSMUSG00000032220

30341/ENSMUSG00000023951/ENSMUSG00000061436/ENSMUSG00000050965/ENSMUSG00000032487/ENSMUSG00000015839/ENSMUSG00000074934
18736/ENSMUSG00000027996/ENSMUSG00000026361/ENSMUSG00000007613/ENSMUSG00000033323/ENSMUSG00000045983/ENSMUSG00000005087/ENSMUSG00000023951/ENSMUSG00000022752/E
23951/ENSMUSG00000044337/ENSMUSG00000039384/ENSMUSG00000006445/ENSMUSG00000020044/ENSMUSG00000050965/ENSMUSG00000036564/ENSMUSG00000015202/ENSMUSG00000062232/E

22425/ENSMUSG00000048616/ENSMUSG00000026728/ENSMUSG00000005125/ENSMUSG00000024927/ENSMUSG00000039384/ENSMUSG00000021379/ENSMUSG00000053398/ENSMUSG00000022037/E
48661/ENSMUSG00000000058/ENSMUSG00000026836/ENSMUSG00000085795/ENSMUSG00000063382/ENSMUSG00000024182

32715/ENSMUSG00000020484/ENSMUSG00000020601/ENSMUSG00000034640/ENSMUSG00000037458/ENSMUSG00000049792/ENSMUSG00000030530/ENSMUSG00000038764/ENSMUSG00000024927/E
18736/ENSMUSG00000022425/ENSMUSG00000026728/ENSMUSG00000023951/ENSMUSG00000020745/ENSMUSG00000036606/ENSMUSG00000096054/ENSMUSG00000054808/ENSMUSG00000028161/E

05087/ENSMUSG00000023951/ENSMUSG00000031548/ENSMUSG00000074934/ENSMUSG00000026812/ENSMUSG00000045394/ENSMUSG00000005871/ENSMUSG00000032220
07613/ENSMUSG00000023951/ENSMUSG00000026836/ENSMUSG00000031548/ENSMUSG00000074934/ENSMUSG00000041702/ENSMUSG00000005871
42594/ENSMUSG00000022443/ENSMUSG00000050965/ENSMUSG00000032702/ENSMUSG00000024486/ENSMUSG00000033717/ENSMUSG00000015839/ENSMUSG00000056071/ENSMUSG00000027712

ENSMUSG00000040021/ENSMUSG00000096054/ENSMUSG00000013076/ENSMUSG00000018583/ENSMUSG00000028161/ENSMUSG00000032717/ENSMUSG00000032702/ENSMUSG00000073563/ENSMUS
ENSMUSG00000009630

ENSMUSG00000050965/ENSMUSG00000024486/ENSMUSG00000079037/ENSMUSG00000000058/ENSMUSG00000033717/ENSMUSG00000026836/ENSMUSG00000031548/ENSMUSG00000074934
ENSMUSG00000004267/ENSMUSG00000023175/ENSMUSG00000050965/ENSMUSG00000063142/ENSMUSG00000079037/ENSMUSG00000032487/ENSMUSG00000039952/ENSMUSG00000000058/ENSMUS

ENSMUSG00000032593/ENSMUSG00000003814/ENSMUSG00000062232/ENSMUSG00000018965/ENSMUSG00000038539/ENSMUSG00000029249/ENSMUSG00000044147/ENSMUSG00000031393/ENSMUS
ENSMUSG00000004267/ENSMUSG00000023175/ENSMUSG00000109511/ENSMUSG00000050965/ENSMUSG00000063142/ENSMUSG00000079037/ENSMUSG00000032487/ENSMUSG00000039952/ENSMUS
ENSMUSG00000004267/ENSMUSG00000023175/ENSMUSG00000050965/ENSMUSG00000063142/ENSMUSG00000079037/ENSMUSG00000032487/ENSMUSG00000039952/ENSMUSG00000000058/ENSMUS
ENSMUSG00000031523/ENSMUSG00000035199/ENSMUSG00000030530/ENSMUSG00000005087/ENSMUSG00000046223/ENSMUSG00000023951/ENSMUSG00000027834/ENSMUSG00000024073/ENSMUS

ENSMUSG00000048616/ENSMUSG00000026361/ENSMUSG00000007613/ENSMUSG00000023951/ENSMUSG00000039384/ENSMUSG00000047281/ENSMUSG00000050965/ENSMUSG00000000058/ENSMUS

ENSMUSG00000050965/ENSMUSG00000024486/ENSMUSG00000079037/ENSMUSG00000000058/ENSMUSG00000033717/ENSMUSG00000026836/ENSMUSG00000031548/ENSMUSG00000074934
ENSMUSG00000032702/ENSMUSG00000021379/ENSMUSG00000032593/ENSMUSG00000003814/ENSMUSG00000062232/ENSMUSG00000018965/ENSMUSG00000038539/ENSMUSG00000029249/ENSMUS

ENSMUSG00000039844/ENSMUSG00000036432/ENSMUSG00000085795/ENSMUSG00000074934/ENSMUSG00000003345/ENSMUSG00000024182/ENSMUSG00000005871
ENSMUSG00000024927/ENSMUSG00000039384/ENSMUSG00000020745/ENSMUSG00000021379/ENSMUSG00000053398/ENSMUSG00000022037/ENSMUSG00000039952/ENSMUSG00000038539/ENSMUS
ENSMUSG00000026826/ENSMUSG00000050697/ENSMUSG00000026812/ENSMUSG00000026628

ENSMUSG00000038859/ENSMUSG00000034216/ENSMUSG00000032220/ENSMUSG00000034593

ENSMUSG00000000282/ENSMUSG00000027230/ENSMUSG00000026836/ENSMUSG00000028179/ENSMUSG00000032350/ENSMUSG00000015839/ENSMUSG00000009291/ENSMUSG00000036432/ENSMUS

ENSMUSG00000015202/ENSMUSG00000062232/ENSMUSG00000029816/ENSMUSG00000039844/ENSMUSG00000037902/ENSMUSG00000026628
ENSMUSG00000095098/ENSMUSG00000038539/ENSMUSG00000044167/ENSMUSG00000031548/ENSMUSG00000026826/ENSMUSG00000024182

ENSMUSG00000020745/ENSMUSG00000021978/ENSMUSG00000047281/ENSMUSG00000039953/ENSMUSG00000034154/ENSMUSG00000030168/ENSMUSG00000024486/ENSMUSG00000020132/ENSMUS
ENSMUSG00000029816/ENSMUSG00000039844/ENSMUSG00000037902/ENSMUSG00000026628

ENSMUSG00000039952/ENSMUSG00000038539/ENSMUSG00000031393/ENSMUSG00000056071

ENSMUSG00000030059/ENSMUSG00000040021/ENSMUSG00000061436/ENSMUSG00000020044/ENSMUSG00000048249/ENSMUSG00000022037/ENSMUSG00000025234/ENSMUSG00000033717/ENSMUS
ENSMUSG00000032702/ENSMUSG00000060012/ENSMUSG00000040945/ENSMUSG00000003814/ENSMUSG00000062232/ENSMUSG00000020132/ENSMUSG00000018965/ENSMUSG00000025326/ENSMUS

;G00000044167/ENSMUSG00000068740/ENSMUSG00000031548/ENSMUSG00000039844/ENSMUSG00000036432/ENSMUSG00000031393/ENSMUSG00000085795/ENSMUSG00000063382/ENSMUSG0000(

;G00000029361/ENSMUSG00000019943

;G00000025326/ENSMUSG00000026812
;G00000000058/ENSMUSG00000029361/ENSMUSG00000019943
;G00000029361/ENSMUSG00000019943
;G00000047281/ENSMUSG00000020044/ENSMUSG00000032487/ENSMUSG00000035992/ENSMUSG00000029249/ENSMUSG00000035900/ENSMUSG00000036432/ENSMUSG00000056071/ENSMUSG0000(

;G00000000184/ENSMUSG00000031548/ENSMUSG00000085795/ENSMUSG00000005871

;G00000044147/ENSMUSG00000031393/ENSMUSG00000025326/ENSMUSG00000026812

;G00000031393/ENSMUSG00000056071

;G00000026826/ENSMUSG00000024807

;G00000095098/ENSMUSG00000034201/ENSMUSG00000028179/ENSMUSG00000031548/ENSMUSG00000024431/ENSMUSG00000031393/ENSMUSG00000074934/ENSMUSG00000002083

;G00000028793/ENSMUSG00000044167/ENSMUSG00000032350/ENSMUSG00000042303/ENSMUSG00000038068/ENSMUSG00000009291/ENSMUSG00000024425/ENSMUSG00000025326/ENSMUSG0000(
;G00000025372

0074934/ENSMUSG00000050697/ENSMUSG00000025372/ENSMUSG00000003345/ENSMUSG00000024182/ENSMUSG00000005871

0002083

0028277/ENSMUSG00000024182/ENSMUSG00000002365/ENSMUSG00000005871

| Category | GOID | Description | GeneRatio | BgRatio | pvalue | padj | geneID | geneName |
|---|---|---|---|---|---|---|---|---|
| BP | GO:003432 | cell junction assembly | 36\|1209 | 201\|16097 | 8.54E-07 | 3.61E-05 | ENSMUS | Plec/Itga5/Hipk1/S100a10/Lamc1/Cdh5/Iqgap1/Pecam1/Mmp1 |
| BP | GO:200123 | regulation of apoptotic signaling pathway | 58\|1209 | 399\|16097 | 8.55E-07 | 3.61E-05 | ENSMUS | Thbs1/Hmox1/Myc/Serpine1/Inhba/Herpud1/Fgfr1/Fem1b/Tnfr |
| BP | GO:000801 | blood circulation | 61\|1209 | 429\|16097 | 9.54E-07 | 3.97E-05 | ENSMUS | Col1a2/Hmox1/Hmgcr/Rgs2/Postn/Map2k3/Myl1/Met/Cdh5/At |
| BP | GO:000751 | skeletal muscle tissue development | 33\|1209 | 177\|16097 | 9.56E-07 | 3.97E-05 | ENSMUS | Acta1/Hmgcr/Myc/Eln/Cdon/Hivep3/Egr1/Mylpf/Met/Rcan1/Pla |
| BP | GO:000301 | circulatory system process | 62\|1209 | 439\|16097 | 9.83E-07 | 4.05E-05 | ENSMUS | Col1a2/Hmox1/Hmgcr/Rgs2/Postn/Map2k3/Myl1/Met/Cdh5/At |
| BP | GO:004358 | skin development | 39\|1209 | 228\|16097 | 1.02E-06 | 4.15E-05 | ENSMUS | Col3a1/Errfi1/Col1a1/Col1a2/Igfbp5/Sox9/Inhba/Col5a2/Met/C |
| BP | GO:003296 | collagen metabolic process | 23\|1209 | 100\|16097 | 1.02E-06 | 4.15E-05 | ENSMUS | Errfi1/Col1a1/Col1a2/Tnxb/Mfap4/Serpine1/Il6ra/Mmp14/Adar |
| BP | GO:001059 | positive regulation of endothelial cell migration | 22\|1209 | 93\|16097 | 1.05E-06 | 4.18E-05 | ENSMUS | Thbs1/Hmox1/Map2k3/Fgfr1/Met/Smoc2/Nrp1/Igf1/Flt4/Vegfa/ |
| BP | GO:007016 | regulation of biomineral tissue development | 22\|1209 | 93\|16097 | 1.05E-06 | 4.18E-05 | ENSMUS | Sox9/Omd/Atf4/Aspn/Cyr61/Ank/Bmp6/Rflna/Gas6/Comp/Cebp |
| BP | GO:000704 | cell-substrate junction assembly | 21\|1209 | 86\|16097 | 1.05E-06 | 4.18E-05 | ENSMUS | Plec/Itga5/S100a10/Lamc1/Iqgap1/Mmp14/Thy1/Nrp1/Dlc1/Tr |
| BP | GO:006074 | prostate gland epithelium morphogenesis | 12\|1209 | 31\|16097 | 1.12E-06 | 4.44E-05 | ENSMUS | Sox9/Fem1b/Fgfr2/Rxra/Tnc/Nog/Igf1r/Igf1/Cd44/Notch1/Id4/S |
| BP | GO:009886 | bone growth | 14\|1209 | 42\|16097 | 1.21E-06 | 4.73E-05 | ENSMUS | Thbs1/Col14a1/Col12a1/Col6a2/Matn3/Comp/Col6a3/Mmp13 |
| BP | GO:003444 | substrate adhesion-dependent cell spreading | 23\|1209 | 101\|16097 | 1.23E-06 | 4.77E-05 | ENSMUS | Postn/Lamb1/Has2/S100a10/Lamc1/Efna1/Fndc3b/Nrp1/Hspg2 |
| BP | GO:000301 | muscle system process | 55\|1209 | 375\|16097 | 1.23E-06 | 4.77E-05 | ENSMUS | Errfi1/Sulf2/Col14a1/Igfbp5/Rgs2/Map2k3/Myl1/Klf4/Met/Cald |
| BP | GO:004870 | astrocyte differentiation | 20\|1209 | 80\|16097 | 1.27E-06 | 4.86E-05 | ENSMUS | Ldlr/Sox9/Abl2/Hmga2/Egfr/Stat3/Il6st/Nr1d1/Nog/Vim/Notch |
| BP | GO:007190 | regulation of protein serine/threonine kinase activity | 58\|1209 | 404\|16097 | 1.29E-06 | 4.90E-05 | ENSMUS | Spry2/Tnxb/Thbs1/Hmgcr/Rgs2/Map2k3/Trib1/Gadd45a/Fgfr1/I |
| BP | GO:003433 | cell junction organization | 41\|1209 | 248\|16097 | 1.35E-06 | 5.10E-05 | ENSMUS | Plec/Itga5/Hipk1/S100a10/Lamc1/Cdh5/Iqgap1/Pecam1/Mmp1 |
| BP | GO:005114 | striated muscle cell differentiation | 47\|1209 | 302\|16097 | 1.38E-06 | 5.18E-05 | ENSMUS | Acta1/Col14a1/Igfbp5/Rgs2/Cdon/Met/Rcan1/Sorbs2/Mmp14/I |
| BP | GO:006156 | axon development | 64\|1209 | 463\|16097 | 1.40E-06 | 5.21E-05 | ENSMUS | Klf4/Xylt1/Ifrd1/Tnfrsf12a/Jun/Efna1/Rtn4/Lama2/Golga4/Cdh1 |
| BP | GO:000716 | cell-matrix adhesion | 35\|1209 | 197\|16097 | 1.47E-06 | 5.43E-05 | ENSMUS | Col3a1/Tnxb/Thbs1/Postn/Serpine1/Plet1/Col5a3/Nid1/S100a1 |
| BP | GO:001097 | positive regulation of neuron projection development | 51\|1209 | 340\|16097 | 1.53E-06 | 5.64E-05 | ENSMUS | Rgs2/Serpinf1/Fgfr1/Mapk6/Tnfrsf12a/Met/Rtn4/Iqgap1/Golga4 |
| BP | GO:006056 | epithelial tube morphogenesis | 53\|1209 | 359\|16097 | 1.57E-06 | 5.74E-05 | ENSMUS | Col4a1/Spry2/Myc/Sox9/Met/Mmp14/Ncoa3/Abl2/Fgfr2/Hmga |
| BP | GO:005109 | regulation of binding | 56\|1209 | 388\|16097 | 1.65E-06 | 6.00E-05 | ENSMUS | Hmox1/Myc/Cdon/Klf4/Hipk1/S100a10/Met/Cald1/Lgals3/Jun/ |
| BP | GO:006051 | prostate gland morphogenesis | 12\|1209 | 32\|16097 | 1.68E-06 | 6.04E-05 | ENSMUS | Sox9/Fem1b/Fgfr2/Rxra/Tnc/Nog/Igf1r/Igf1/Cd44/Notch1/Id4/S |
| BP | GO:004350 | regulation of muscle adaptation | 22\|1209 | 96\|16097 | 1.86E-06 | 6.66E-05 | ENSMUS | Errfi1/Col14a1/Igfbp5/Rgs2/Klf4/Lmna/Atp2a2/Mef2a/Lmcd1/T |
| BP | GO:004328 | regulation of cysteine-type endopeptidase activity involved in a | 34\|1209 | 191\|16097 | 1.97E-06 | 6.88E-05 | ENSMUS | Hspa1b/F3/Thbs1/Myc/Klf4/Herpud1/Eif2ak3/Cyr61/Ctgf/Dhcr2 |
| BP | GO:000341 | chondrocyte morphogenesis involved in endochondral bone m | 8\|1209 | 14\|16097 | 1.97E-06 | 6.88E-05 | ENSMUS | Col14a1/Col12a1/Col6a2/Matn3/Col6a3/Matn1/Col6a1/Vit |
| BP | GO:000342 | growth plate cartilage chondrocyte morphogenesis | 8\|1209 | 14\|16097 | 1.97E-06 | 6.88E-05 | ENSMUS | Col14a1/Col12a1/Col6a2/Matn3/Col6a3/Matn1/Col6a1/Vit |
| BP | GO:009017 | chondrocyte morphogenesis | 8\|1209 | 14\|16097 | 1.97E-06 | 6.88E-05 | ENSMUS | Col14a1/Col12a1/Col6a2/Matn3/Col6a3/Matn1/Col6a1/Vit |
| BP | GO:001046 | negative regulation of peptidase activity | 33\|1209 | 183\|16097 | 2.07E-06 | 7.17E-05 | ENSMUS | Hspa1b/Thbs1/Serpine1/Klf4/Herpud1/Serpinf1/A2m/Cd109/Se |
| BP | GO:005073 | regulation of peptidyl-tyrosine phosphorylation | 40\|1209 | 244\|16097 | 2.25E-06 | 7.74E-05 | ENSMUS | Errfi1/Itga5/Il6ra/Efna1/Iqgap1/Pecam1/Socs3/Bmp6/Gprc5a/T |
| BP | GO:003051 | regulation of BMP signaling pathway | 21\|1209 | 90\|16097 | 2.33E-06 | 7.96E-05 | ENSMUS | Cdh5/Chrdl1/Cyr61/Tgfbr3/Neo1/Ski/Sfrp2/Nog/Twsg1/Notch1 |
| BP | GO:003027 | negative regulation of ossification | 20\|1209 | 83\|16097 | 2.36E-06 | 8.03E-05 | ENSMUS | Igfbp5/Sox9/Dlk1/Fgfr1/Noct/Rflna/Fndc3b/Ski/Nog/Twsg1/Nfa |
| BP | GO:006053 | skeletal muscle organ development | 33\|1209 | 185\|16097 | 2.65E-06 | 8.95E-05 | ENSMUS | Acta1/Hmgcr/Myc/Eln/Cdon/Hivep3/Egr1/Mylpf/Met/Rcan1/Pla |
| BP | GO:004341 | positive regulation of MAPK cascade | 64\|1209 | 472\|16097 | 2.70E-06 | 9.07E-05 | ENSMUS | Spry2/Thbs1/Hmgcr/Tlr2/Map2k3/Cdon/Gadd45a/Fgfr1/Igfbp3/ |
| BP | GO:005254 | regulation of endopeptidase activity | 45\|1209 | 291\|16097 | 2.74E-06 | 9.16E-05 | ENSMUS | Hspa1b/F3/Thbs1/Myc/Serpine1/Klf4/Herpud1/Serpinf1/Efna1/ |
| BP | GO:190351 | mucopolysaccharide metabolic process | 16\|1209 | 57\|16097 | 2.82E-06 | 9.37E-05 | ENSMUS | Xylt1/Has2/Tmem2/Chpf/Itih5/Chst11/Csgalnact1/Slc35d1/Cd4 |
| BP | GO:001071 | regulation of epithelial to mesenchymal transition | 20\|1209 | 84\|16097 | 2.89E-06 | 9.51E-05 | ENSMUS | Col1a1/Vasn/Efna1/Phldb1/Tgfbr3/Tiam1/Ppp2ca/Sfrp2/Nog/Tg |
| BP | GO:006138 | trabecula morphogenesis | 15\|1209 | 51\|16097 | 3.03E-06 | 9.92E-05 | ENSMUS | Col1a1/Chad/Tgfbr3/Nog/Tgfbr1/Vegfa/Nfatc1/Sbno2/Notch1/A |
| BP | GO:009719 | intrinsic apoptotic signaling pathway | 44\|1209 | 283\|16097 | 3.05E-06 | 9.95E-05 | ENSMUS | Hmox1/Myc/Chac1/Fnip2/Hipk1/Herpud1/Atf4/Cdkn1a/Ier3/Ei |
| BP | GO:006048 | mesenchyme development | 41\|1209 | 256\|16097 | 3.09E-06 | 0.0001 | ENSMUS | Col1a1/Thbs1/Myc/Vasn/Sox9/Fgfr1/Has2/Efna1/Rtn4/Phldb1/I |
| CC | GO:004485 | plasma membrane raft | 25\|1232 | 121\|16313 | 3.20E-06 | 0.00012 | ENSMUS | Hmox1/Ldlr/Has2/Pacsin2/Slc2a1/Atp1a1/Cavin2/Lrp8/Jak2/Dl |
| BP | GO:011011 | positive regulation of animal organ morphogenesis | 21\|1209 | 92\|16097 | 3.39E-06 | 0.00011 | ENSMUS | Myc/Sox9/Fgfr1/Rtn4/Fgfr2/Hmga2/Cd34/Xbp1/Agtr2/Maged1/ |
| CC | GO:000558 | fibrillar collagen trimer | 7\|1232 | 11\|16313 | 3.47E-06 | 0.00012 | ENSMUS | Col3a1/Col1a1/Col1a2/Col5a3/Col5a2/Lum/Col11a1 |
| CC | GO:009864 | banded collagen fibril | 7\|1232 | 11\|16313 | 3.47E-06 | 0.00012 | ENSMUS | Col3a1/Col1a1/Col1a2/Col5a3/Col5a2/Lum/Col11a1 |
| BP | GO:000183 | epithelial to mesenchymal transition | 26\|1209 | 130\|16097 | 3.49E-06 | 0.00011 | ENSMUS | Col1a1/Vasn/Sox9/Has2/Efna1/Rtn4/Phldb1/Fgfr2/Tgfbr3/Tiam |
| BP | GO:004559 | regulation of fat cell differentiation | 26\|1209 | 130\|16097 | 3.49E-06 | 0.00011 | ENSMUS | Dlk1/Wif1/Lpl/Noct/Ddit3/Insig1/Fndc3b/Trib3/Xbp1/Trib2/Sfr |
| BP | GO:000315 | endothelium development | 24\|1209 | 115\|16097 | 3.78E-06 | 0.00012 | ENSMUS | Tnmd/Met/Cdh5/Rtn4/Pecam1/Col18a1/Bmp6/Fasn/Clic4/Tnfr |
| BP | GO:006084 | artery development | 22\|1209 | 100\|16097 | 3.82E-06 | 0.00012 | ENSMUS | Col3a1/Ldlr/Rtn4/Mylk/Zmiz1/Nrp1/Notch3/Adgrf5/Comp/Ltb |
| BP | GO:000342 | growth plate cartilage morphogenesis | 8\|1209 | 15\|16097 | 3.95E-06 | 0.00012 | ENSMUS | Col14a1/Col12a1/Col6a2/Matn3/Col6a3/Matn1/Col6a1/Vit |
| BP | GO:003592 | cellular response to vascular endothelial growth factor stimulu | 15\|1209 | 52\|16097 | 3.96E-06 | 0.00012 | ENSMUS | Spry2/Map2k3/Smoc2/Gab1/Xbp1/Nr4a1/Nrp1/Flt4/Robo1/Re |

| Count | Up | Up_Gene_i | Down | Down_Gene_id |
|---|---|---|---|---|
| 36 | 16 | ENSMUSG0 | 20 | ENSMUSG00000022565/ENSMUSG00000000555/ENSMUSG00000008730/ENSMUSG00000041959/ENSMUSG00000030536/ENSMUSG00000031523/ENSMUSG0 |
| 58 | 9 | ENSMUSG0 | 49 | ENSMUSG00000040152/ENSMUSG00000005413/ENSMUSG00000022346/ENSMUSG00000037411/ENSMUSG00000041324/ENSMUSG00000031770/ENSMUSG0 |
| 61 | 24 | ENSMUSG0 | 37 | ENSMUSG00000005413/ENSMUSG00000021670/ENSMUSG00000026360/ENSMUSG00000018932/ENSMUSG00000009376/ENSMUSG00000033161/ENSMUSG0 |
| 33 | 11 | ENSMUSG0 | 22 | ENSMUSG00000021670/ENSMUSG00000022346/ENSMUSG00000029675/ENSMUSG00000038119/ENSMUSG00000028634/ENSMUSG00000038418/ENSMUSG0 |
| 62 | 25 | ENSMUSG0 | 37 | ENSMUSG00000005413/ENSMUSG00000021670/ENSMUSG00000026360/ENSMUSG00000018932/ENSMUSG00000009376/ENSMUSG00000033161/ENSMUSG0 |
| 39 | 15 | ENSMUSG0 | 24 | ENSMUSG00000028967/ENSMUSG00000000567/ENSMUSG00000041324/ENSMUSG00000009376/ENSMUSG00000046186/ENSMUSG00000037071/ENSMUSG0 |
| 23 | 10 | ENSMUSG0 | 13 | ENSMUSG00000028967/ENSMUSG00000037411/ENSMUSG00000027947/ENSMUSG00000029287/ENSMUSG00000028641/ENSMUSG00000006931/ENSMUSG0 |
| 22 | 10 | ENSMUSG0 | 12 | ENSMUSG00000040152/ENSMUSG00000005413/ENSMUSG00000018932/ENSMUSG00000031565/ENSMUSG00000009376/ENSMUSG00000023951/ENSMUSG0 |
| 22 | 11 | ENSMUSG0 | 11 | ENSMUSG00000000567/ENSMUSG00000042406/ENSMUSG00000022265/ENSMUSG00000039004/ENSMUSG00000031451/ENSMUSG00000031849/ENSMUSG0 |
| 21 | 9 | ENSMUSG0 | 12 | ENSMUSG00000022565/ENSMUSG00000000555/ENSMUSG00000041959/ENSMUSG00000030536/ENSMUSG00000031523/ENSMUSG00000055322/ENSMUSG0 |
| 12 | 3 | ENSMUSG0 | 9 | ENSMUSG00000000567/ENSMUSG00000032244/ENSMUSG00000030849/ENSMUSG00000015846/ENSMUSG00000048616/ENSMUSG00000005533/ENSMUSG0 |
| 14 | 7 | ENSMUSG0 | 7 | ENSMUSG00000040152/ENSMUSG00000020241/ENSMUSG00000031849/ENSMUSG00000048126/ENSMUSG00000026147/ENSMUSG00000028047/ENSMUSG0 |
| 23 | 13 | ENSMUSG0 | 10 | ENSMUSG00000022367/ENSMUSG00000041959/ENSMUSG00000027954/ENSMUSG00000039286/ENSMUSG00000022425/ENSMUSG00000054808/ENSMUSG0 |
| 55 | 26 | ENSMUSG0 | 29 | ENSMUSG00000028967/ENSMUSG00000006800/ENSMUSG00000026360/ENSMUSG00000018932/ENSMUSG00000003032/ENSMUSG00000009376/ENSMUSG0 |
| 20 | 7 | ENSMUSG0 | 13 | ENSMUSG00000032193/ENSMUSG00000000567/ENSMUSG00000026596/ENSMUSG00000056758/ENSMUSG00000004040/ENSMUSG00000021756/ENSMUSG0 |
| 58 | 16 | ENSMUSG0 | 42 | ENSMUSG00000022114/ENSMUSG00000040152/ENSMUSG00000021670/ENSMUSG00000026360/ENSMUSG00000018932/ENSMUSG00000032501/ENSMUSG0 |
| 41 | 17 | ENSMUSG0 | 24 | ENSMUSG00000022565/ENSMUSG00000000555/ENSMUSG00000008730/ENSMUSG00000041959/ENSMUSG00000030536/ENSMUSG00000039004/ENSMUSG0 |
| 47 | 23 | ENSMUSG0 | 24 | ENSMUSG00000026360/ENSMUSG00000038119/ENSMUSG00000009376/ENSMUSG00000022951/ENSMUSG00000028063/ENSMUSG00000015846/ENSMUSG0 |
| 64 | 26 | ENSMUSG0 | 38 | ENSMUSG00000003032/ENSMUSG00000030657/ENSMUSG00000001627/ENSMUSG00000023905/ENSMUSG00000027954/ENSMUSG00000020458/ENSMUSG0 |
| 35 | 18 | ENSMUSG0 | 17 | ENSMUSG00000040152/ENSMUSG00000037411/ENSMUSG00000032068/ENSMUSG00000041959/ENSMUSG00000030536/ENSMUSG00000002489/ENSMUSG0 |
| 51 | 16 | ENSMUSG0 | 35 | ENSMUSG00000026360/ENSMUSG00000000753/ENSMUSG00000031565/ENSMUSG00000042688/ENSMUSG00000023905/ENSMUSG00000009376/ENSMUSG0 |
| 53 | 27 | ENSMUSG0 | 26 | ENSMUSG00000022114/ENSMUSG00000022346/ENSMUSG00000000567/ENSMUSG00000009376/ENSMUSG00000027678/ENSMUSG00000026596/ENSMUSG0 |
| 56 | 18 | ENSMUSG0 | 38 | ENSMUSG00000005413/ENSMUSG00000022346/ENSMUSG00000038119/ENSMUSG00000003032/ENSMUSG00000008730/ENSMUSG00000041959/ENSMUSG0 |
| 12 | 3 | ENSMUSG0 | 9 | ENSMUSG00000000567/ENSMUSG00000032244/ENSMUSG00000030849/ENSMUSG00000015846/ENSMUSG00000048616/ENSMUSG00000005533/ENSMUSG0 |
| 22 | 8 | ENSMUSG0 | 14 | ENSMUSG00000028967/ENSMUSG00000026360/ENSMUSG00000003032/ENSMUSG00000028063/ENSMUSG00000029467/ENSMUSG00000030557/ENSMUSG0 |
| 34 | 8 | ENSMUSG0 | 26 | ENSMUSG00000028128/ENSMUSG00000040152/ENSMUSG00000022346/ENSMUSG00000003032/ENSMUSG00000031770/ENSMUSG00000031668/ENSMUSG0 |
| 8 | 5 | ENSMUSG0 | 3 | ENSMUSG00000020241/ENSMUSG00000048126/ENSMUSG00000001119 |
| 8 | 5 | ENSMUSG0 | 3 | ENSMUSG00000020241/ENSMUSG00000048126/ENSMUSG00000001119 |
| 8 | 5 | ENSMUSG0 | 3 | ENSMUSG00000020241/ENSMUSG00000048126/ENSMUSG00000001119 |
| 33 | 8 | ENSMUSG0 | 25 | ENSMUSG00000040152/ENSMUSG00000037411/ENSMUSG00000003032/ENSMUSG00000031770/ENSMUSG00000000753/ENSMUSG00000030111/ENSMUSG0 |
| 40 | 14 | ENSMUSG0 | 26 | ENSMUSG00000028967/ENSMUSG00000000555/ENSMUSG00000027947/ENSMUSG00000027954/ENSMUSG00000030536/ENSMUSG00000039004/ENSMUSG0 |
| 21 | 10 | ENSMUSG0 | 11 | ENSMUSG00000029287/ENSMUSG00000032340/ENSMUSG00000029050/ENSMUSG00000027996/ENSMUSG00000048616/ENSMUSG00000024098/ENSMUSG0 |
| 20 | 8 | ENSMUSG0 | 12 | ENSMUSG00000000567/ENSMUSG00000040856/ENSMUSG00000031565/ENSMUSG00000023087/ENSMUSG00000039286/ENSMUSG00000029050/ENSMUSG0 |
| 33 | 11 | ENSMUSG0 | 22 | ENSMUSG00000021670/ENSMUSG00000022346/ENSMUSG00000029675/ENSMUSG00000038119/ENSMUSG00000028634/ENSMUSG00000038418/ENSMUSG0 |
| 64 | 24 | ENSMUSG0 | 40 | ENSMUSG00000022114/ENSMUSG00000040152/ENSMUSG00000021670/ENSMUSG00000027995/ENSMUSG00000018932/ENSMUSG00000038119/ENSMUSG0 |
| 45 | 10 | ENSMUSG0 | 35 | ENSMUSG00000028128/ENSMUSG00000040152/ENSMUSG00000022346/ENSMUSG00000037411/ENSMUSG00000003032/ENSMUSG00000031770/ENSMUSG0 |
| 16 | 5 | ENSMUSG0 | 11 | ENSMUSG00000030657/ENSMUSG00000022367/ENSMUSG00000024754/ENSMUSG00000032997/ENSMUSG00000036356/ENSMUSG00000028521/ENSMUSG0 |
| 20 | 3 | ENSMUSG0 | 17 | ENSMUSG00000039646/ENSMUSG00000027954/ENSMUSG00000048537/ENSMUSG00000029287/ENSMUSG00000002489/ENSMUSG00000020349/ENSMUSG0 |
| 15 | 6 | ENSMUSG0 | 9 | ENSMUSG00000039084/ENSMUSG00000029287/ENSMUSG00000048616/ENSMUSG00000007613/ENSMUSG00000023951/ENSMUSG00000035673/ENSMUSG0 |
| 44 | 4 | ENSMUSG0 | 40 | ENSMUSG00000005413/ENSMUSG00000022346/ENSMUSG00000027313/ENSMUSG00000061175/ENSMUSG00000008730/ENSMUSG00000031770/ENSMUSG0 |
| 41 | 15 | ENSMUSG0 | 26 | ENSMUSG00000040152/ENSMUSG00000022346/ENSMUSG00000039646/ENSMUSG00000000567/ENSMUSG00000031565/ENSMUSG00000022367/ENSMUSG0 |
| 25 | 6 | ENSMUSG0 | 19 | ENSMUSG00000005413/ENSMUSG00000032193/ENSMUSG00000022367/ENSMUSG00000016664/ENSMUSG00000028645/ENSMUSG00000033161/ENSMUSG0 |
| 21 | 9 | ENSMUSG0 | 12 | ENSMUSG00000022346/ENSMUSG00000000567/ENSMUSG00000031565/ENSMUSG00000020458/ENSMUSG00000030849/ENSMUSG00000056758/ENSMUSG0 |
| 7 | 7 | ENSMUSG0 | 0 | |
| 7 | 7 | ENSMUSG0 | 0 | |
| 26 | 5 | ENSMUSG0 | 21 | ENSMUSG00000039646/ENSMUSG00000000567/ENSMUSG00000022367/ENSMUSG00000027954/ENSMUSG00000020458/ENSMUSG00000048537/ENSMUSG0 |
| 26 | 5 | ENSMUSG0 | 21 | ENSMUSG00000040856/ENSMUSG00000020218/ENSMUSG00000023087/ENSMUSG00000025408/ENSMUSG00000045294/ENSMUSG00000039286/ENSMUSG0 |
| 24 | 9 | ENSMUSG0 | 15 | ENSMUSG00000009376/ENSMUSG00000020458/ENSMUSG00000039004/ENSMUSG00000025153/ENSMUSG00000037242/ENSMUSG00000030341/ENSMUSG0 |
| 22 | 13 | ENSMUSG0 | 9 | ENSMUSG00000032193/ENSMUSG00000020458/ENSMUSG00000007817/ENSMUSG00000031849/ENSMUSG00000048616/ENSMUSG00000007613/ENSMUSG0 |
| 8 | 5 | ENSMUSG0 | 3 | ENSMUSG00000020241/ENSMUSG00000048126/ENSMUSG00000001119 |
| 15 | 7 | ENSMUSG0 | 8 | ENSMUSG00000022114/ENSMUSG00000018932/ENSMUSG00000031714/ENSMUSG00000020484/ENSMUSG00000023034/ENSMUSG00000024927/ENSMUSG0 |

0000055322/ENSMUSG00000022425/ENSMUSG00000023951/ENSMUSG00000037815/ENSMUSG00000054808/ENSMUSG00000006445/ENSMUSG00000050965/ENSMUSG00000040945/ENSMUSG0000006
0000031565/ENSMUSG00000032244/ENSMUSG00000023905/ENSMUSG00000050335/ENSMUSG00000003541/ENSMUSG00000031668/ENSMUSG00000028063/ENSMUSG00000025408/ENSMUSG0000003
0000003541/ENSMUSG00000021540/ENSMUSG00000029467/ENSMUSG00000039004/ENSMUSG00000028525/ENSMUSG00000031778/ENSMUSG00000029287/ENSMUSG00000030557/ENSMUSG0000001
0000009376/ENSMUSG00000022951/ENSMUSG00000019817/ENSMUSG00000001419/ENSMUSG00000020423/ENSMUSG00000023034/ENSMUSG00000029050/ENSMUSG00000028161/ENSMUSG0000003
0000003541/ENSMUSG00000021540/ENSMUSG00000029467/ENSMUSG00000039004/ENSMUSG00000028525/ENSMUSG00000031778/ENSMUSG00000029287/ENSMUSG00000030557/ENSMUSG0000001
0000027678/ENSMUSG00000030849/ENSMUSG00000034926/ENSMUSG00000037242/ENSMUSG00000074794/ENSMUSG00000029135/ENSMUSG00000031849/ENSMUSG00000024927/ENSMUSG0000004
0000019997/ENSMUSG00000038168/ENSMUSG00000041779/ENSMUSG00000026728/ENSMUSG00000004846/ENSMUSG00000020682/ENSMUSG00000027230
0000023068/ENSMUSG00000013076/ENSMUSG00000050965/ENSMUSG00000003814/ENSMUSG00000032487/ENSMUSG00000015839
0000056501/ENSMUSG00000056050/ENSMUSG00000026836/ENSMUSG00000019943/ENSMUSG00000074934
0000022425/ENSMUSG00000023951/ENSMUSG00000040945/ENSMUSG00000026131/ENSMUSG00000074934/ENSMUSG00000026812
0000005087/ENSMUSG00000021379/ENSMUSG00000031548
0000001119
0000032702/ENSMUSG00000040945/ENSMUSG00000003814/ENSMUSG00000031548
0000033161/ENSMUSG00000028063/ENSMUSG00000021540/ENSMUSG00000029467/ENSMUSG00000028525/ENSMUSG00000030557/ENSMUSG00000002489/ENSMUSG00000019997/ENSMUSG0000003
0000020889/ENSMUSG00000048616/ENSMUSG00000026728/ENSMUSG00000021379/ENSMUSG00000038539/ENSMUSG00000031393/ENSMUSG00000056071
0000036390/ENSMUSG00000031565/ENSMUSG00000009376/ENSMUSG00000030536/ENSMUSG00000023067/ENSMUSG00000027351/ENSMUSG00000028893/ENSMUSG00000002489/ENSMUSG0000003
0000031523/ENSMUSG00000055322/ENSMUSG00000022425/ENSMUSG00000007613/ENSMUSG00000031788/ENSMUSG00000023951/ENSMUSG00000037815/ENSMUSG00000054808/ENSMUSG0000000
0000050567/ENSMUSG00000030557/ENSMUSG00000020484/ENSMUSG00000032340/ENSMUSG00000029050/ENSMUSG00000031849/ENSMUSG00000033323/ENSMUSG00000023951/ENSMUSG0000002
0000038708/ENSMUSG00000030849/ENSMUSG00000032064/ENSMUSG00000035671/ENSMUSG00000017929/ENSMUSG00000002489/ENSMUSG00000026014/ENSMUSG00000018736/ENSMUSG0000002
0000022508/ENSMUSG00000019997/ENSMUSG00000031523/ENSMUSG00000022425/ENSMUSG00000023951/ENSMUSG00000022817/ENSMUSG00000040945/ENSMUSG00000039952/ENSMUSG0000007
0000020458/ENSMUSG00000030536/ENSMUSG00000038708/ENSMUSG00000026596/ENSMUSG00000032064/ENSMUSG00000019970/ENSMUSG00000031778/ENSMUSG00000029426/ENSMUSG0000002
0000030849/ENSMUSG00000056758/ENSMUSG00000035342/ENSMUSG00000037242/ENSMUSG00000029050/ENSMUSG00000027996/ENSMUSG00000031523/ENSMUSG00000048616/ENSMUSG0000000
0000009376/ENSMUSG00000050335/ENSMUSG00000031668/ENSMUSG00000025408/ENSMUSG00000026596/ENSMUSG00000029287/ENSMUSG00000002489/ENSMUSG00000020349/ENSMUSG0000003
0000005087/ENSMUSG00000021379/ENSMUSG00000031548
0000030341/ENSMUSG00000033323/ENSMUSG00000022752/ENSMUSG00000028161/ENSMUSG00000050965/ENSMUSG00000026313/ENSMUSG00000044167/ENSMUSG00000024431
0000019997/ENSMUSG00000034926/ENSMUSG00000020349/ENSMUSG00000023034/ENSMUSG00000024789/ENSMUSG00000031451/ENSMUSG00000027996/ENSMUSG00000031523/ENSMUSG0000003

0000046186/ENSMUSG00000027447/ENSMUSG00000021950/ENSMUSG00000001131/ENSMUSG00000034926/ENSMUSG00000023034/ENSMUSG00000067780/ENSMUSG00000031451/ENSMUSG0000002
0000046733/ENSMUSG00000028613/ENSMUSG00000021756/ENSMUSG00000024789/ENSMUSG00000031451/ENSMUSG00000027996/ENSMUSG00000027293/ENSMUSG00000022425/ENSMUSG0000003
0000061436/ENSMUSG00000048661/ENSMUSG00000031548/ENSMUSG00000044857/ENSMUSG00000074934
0000048616/ENSMUSG00000024098/ENSMUSG00000026313/ENSMUSG00000031548/ENSMUSG00000074934/ENSMUSG00000046791
0000009376/ENSMUSG00000022951/ENSMUSG00000019817/ENSMUSG00000001419/ENSMUSG00000020423/ENSMUSG00000023034/ENSMUSG00000029050/ENSMUSG00000028161/ENSMUSG0000003
0000036390/ENSMUSG00000031565/ENSMUSG00000009376/ENSMUSG00000027947/ENSMUSG00000027954/ENSMUSG00000030536/ENSMUSG00000026596/ENSMUSG00000030849/ENSMUSG0000003
0000000753/ENSMUSG00000027954/ENSMUSG00000031668/ENSMUSG00000021950/ENSMUSG00000001131/ENSMUSG00000004040/ENSMUSG00000019997/ENSMUSG00000034926/ENSMUSG0000002
0000005087/ENSMUSG00000003665/ENSMUSG00000029201/ENSMUSG00000047126/ENSMUSG00000039497
0000027996/ENSMUSG00000048616/ENSMUSG00000007613/ENSMUSG00000023951/ENSMUSG00000039952/ENSMUSG00000026836/ENSMUSG00000034205/ENSMUSG00000031548/ENSMUSG0000008
0000028047/ENSMUSG00000031548/ENSMUSG00000074934
0000042406/ENSMUSG00000023067/ENSMUSG00000003541/ENSMUSG00000031668/ENSMUSG00000025408/ENSMUSG00000032115/ENSMUSG00000026510/ENSMUSG00000040463/ENSMUSG0000003
0000027954/ENSMUSG00000020458/ENSMUSG00000048537/ENSMUSG00000030849/ENSMUSG00000015846/ENSMUSG00000029287/ENSMUSG00000002489/ENSMUSG00000020349/ENSMUSG0000002
0000028613/ENSMUSG00000024789/ENSMUSG00000031523/ENSMUSG00000037012/ENSMUSG00000007613/ENSMUSG00000005533/ENSMUSG00000004044/ENSMUSG00000037815/ENSMUSG0000006
0000020484/ENSMUSG00000048616/ENSMUSG00000007613/ENSMUSG00000023951/ENSMUSG00000026836/ENSMUSG00000074934

0000030849/ENSMUSG00000029287/ENSMUSG00000002489/ENSMUSG00000020349/ENSMUSG00000027996/ENSMUSG00000048616/ENSMUSG00000007613/ENSMUSG00000023951/ENSMUSG0000003
0000032715/ENSMUSG00000020484/ENSMUSG00000020601/ENSMUSG00000027996/ENSMUSG00000020889/ENSMUSG00000056501/ENSMUSG00000023951/ENSMUSG00000039384/ENSMUSG0000002
0000007613/ENSMUSG00000004846/ENSMUSG00000023951/ENSMUSG00000024812/ENSMUSG00000018476/ENSMUSG00000022016/ENSMUSG00000062232/ENSMUSG00000026836/ENSMUSG0000003
0000035247/ENSMUSG00000023951/ENSMUSG00000031540

0000023951/ENSMUSG00000023068

52232/ENSMUSG00000026131/ENSMUSG00000039844/ENSMUSG00000085795/ENSMUSG00000074934/ENSMUSG00000026812
32115/ENSMUSG00000031778/ENSMUSG00000020484/ENSMUSG00000024789/ENSMUSG00000027996/ENSMUSG00000048616/ENSMUSG00000007613/ENSMUSG00000049792/ENSMUSG00000026389/E
19997/ENSMUSG00000024789/ENSMUSG00000034765/ENSMUSG00000046410/ENSMUSG00000031451/ENSMUSG00000070645/ENSMUSG00000031849/ENSMUSG00000004846/ENSMUSG00000023951/E
34998/ENSMUSG00000026313/ENSMUSG00000048661/ENSMUSG00000000058/ENSMUSG00000044857/ENSMUSG00000063382/ENSMUSG00000050697/ENSMUSG00000026628
19997/ENSMUSG00000024789/ENSMUSG00000034765/ENSMUSG00000046410/ENSMUSG00000031451/ENSMUSG00000070645/ENSMUSG00000031849/ENSMUSG00000004846/ENSMUSG00000023951/E
40021/ENSMUSG00000053797/ENSMUSG00000033508/ENSMUSG00000047281/ENSMUSG00000006445/ENSMUSG00000039473/ENSMUSG00000032487/ENSMUSG00000004207/ENSMUSG00000005871/E

31849/ENSMUSG00000030341/ENSMUSG00000033323/ENSMUSG00000022752/ENSMUSG00000021070/ENSMUSG00000028161/ENSMUSG00000050965/ENSMUSG00000063142/ENSMUSG00000026313/E

31714/ENSMUSG00000020349/ENSMUSG00000032715/ENSMUSG00000020601/ENSMUSG00000024789/ENSMUSG00000034765/ENSMUSG00000027996/ENSMUSG00000007613/ENSMUSG00000005533/E
06445/ENSMUSG00000050965/ENSMUSG00000040945/ENSMUSG00000062232/ENSMUSG00000026131/ENSMUSG00000044147/ENSMUSG00000039844/ENSMUSG00000085795/ENSMUSG00000074934/E
22752/ENSMUSG00000018476/ENSMUSG00000022443/ENSMUSG00000028161/ENSMUSG00000026313/ENSMUSG00000000058/ENSMUSG00000026836/ENSMUSG00000024431/ENSMUSG00000074934/E
24789/ENSMUSG00000032340/ENSMUSG00000035764/ENSMUSG00000048616/ENSMUSG00000026728/ENSMUSG00000005533/ENSMUSG00000022687/ENSMUSG00000023951/ENSMUSG00000020745/E
74934/ENSMUSG00000037902/ENSMUSG00000026812
28641/ENSMUSG00000002489/ENSMUSG00000028613/ENSMUSG00000018736/ENSMUSG00000005533/ENSMUSG00000023951/ENSMUSG00000027834/ENSMUSG00000020745/ENSMUSG00000036606/E
05087/ENSMUSG00000035247/ENSMUSG00000023951/ENSMUSG00000036606/ENSMUSG00000006445/ENSMUSG00000029009/ENSMUSG00000039952/ENSMUSG00000026836/ENSMUSG00000031548/E
32715/ENSMUSG00000024789/ENSMUSG00000029050/ENSMUSG00000048616/ENSMUSG00000007613/ENSMUSG00000045983/ENSMUSG00000026796/ENSMUSG00000006585/ENSMUSG00000046223/E

35199/ENSMUSG00000005087/ENSMUSG00000046223/ENSMUSG00000023951/ENSMUSG00000047281/ENSMUSG00000032487/ENSMUSG00000035992/ENSMUSG00000029249/ENSMUSG00000035900/E

27996/ENSMUSG00000005087/ENSMUSG00000046223/ENSMUSG00000023951/ENSMUSG00000027834/ENSMUSG00000024073/ENSMUSG00000047281/ENSMUSG00000020044/ENSMUSG00000032487/E
30341/ENSMUSG00000005087/ENSMUSG00000023951/ENSMUSG00000042594/ENSMUSG00000050965/ENSMUSG00000024486/ENSMUSG00000079037/ENSMUSG00000000058/ENSMUSG00000033717/E

34998/ENSMUSG00000026313/ENSMUSG00000048661/ENSMUSG00000000058/ENSMUSG00000044857/ENSMUSG00000063382/ENSMUSG00000050697/ENSMUSG00000026628
32064/ENSMUSG00000031778/ENSMUSG00000002489/ENSMUSG00000019997/ENSMUSG00000031714/ENSMUSG00000024789/ENSMUSG00000031451/ENSMUSG00000007613/ENSMUSG00000035199/E
20349/ENSMUSG00000023034/ENSMUSG00000024789/ENSMUSG00000031451/ENSMUSG00000027996/ENSMUSG00000031523/ENSMUSG00000035199/ENSMUSG00000030530/ENSMUSG00000005087/E

35795/ENSMUSG00000063382/ENSMUSG00000074934

32715/ENSMUSG00000020484/ENSMUSG00000024789/ENSMUSG00000030341/ENSMUSG00000049792/ENSMUSG00000035199/ENSMUSG00000056501/ENSMUSG00000026389/ENSMUSG00000005087/E
27996/ENSMUSG00000048616/ENSMUSG00000007613/ENSMUSG00000046223/ENSMUSG00000023951/ENSMUSG00000039952/ENSMUSG00000026836/ENSMUSG00000034205/ENSMUSG00000031548/E
53142/ENSMUSG00000032487/ENSMUSG00000039952/ENSMUSG00000000058/ENSMUSG00000029361

39952/ENSMUSG00000026836/ENSMUSG00000034205/ENSMUSG00000031548/ENSMUSG00000085795/ENSMUSG00000063382/ENSMUSG00000074934
28042/ENSMUSG00000021379/ENSMUSG00000032487/ENSMUSG00000095098/ENSMUSG00000044167/ENSMUSG00000031548/ENSMUSG00000024182
39844

ENSMUSG00000005087/ENSMUSG00000024927/ENSMUSG00000046223/ENSMUSG00000023951/ENSMUSG00000044337/ENSMUSG00000037815/ENSMUSG00000024073/ENSMUSG00000020044/ENSMUS
ENSMUSG00000021070/ENSMUSG00000050965/ENSMUSG00000063142/ENSMUSG00000024486/ENSMUSG00000026313/ENSMUSG00000032487/ENSMUSG00000033717/ENSMUSG00000032350/ENSMUS

ENSMUSG00000021070/ENSMUSG00000050965/ENSMUSG00000063142/ENSMUSG00000024486/ENSMUSG00000026313/ENSMUSG00000032487/ENSMUSG00000033717/ENSMUSG00000032350/ENSMUS
ENSMUSG00000034593

ENSMUSG00000032487/ENSMUSG00000033717/ENSMUSG00000044167/ENSMUSG00000029361/ENSMUSG00000024431/ENSMUSG00000019943

ENSMUSG00000081683/ENSMUSG00000023951/ENSMUSG00000039384/ENSMUSG00000040021/ENSMUSG00000027177/ENSMUSG00000042594/ENSMUSG00000047281/ENSMUSG00000036707/ENSMUS
ENSMUSG00000026812
ENSMUSG00000026812
ENSMUSG00000036606/ENSMUSG00000006445/ENSMUSG00000023175/ENSMUSG00000050965/ENSMUSG00000060012/ENSMUSG00000025582/ENSMUSG00000020132/ENSMUSG00000026131/ENSMUS

ENSMUSG00000024924/ENSMUSG00000036564/ENSMUSG00000062232/ENSMUSG00000020132/ENSMUSG00000015839/ENSMUSG00000042211/ENSMUSG00000024431/ENSMUSG00000039844/ENSMUS
ENSMUSG00000024431/ENSMUSG00000074934/ENSMUSG00000026812
ENSMUSG00000030738/ENSMUSG00000027177/ENSMUSG00000061436/ENSMUSG00000028161/ENSMUSG00000032717/ENSMUSG00000021379/ENSMUSG00000026313/ENSMUSG00000021149/ENSMUS

ENSMUSG00000036432/ENSMUSG00000056071/ENSMUSG00000002083

ENSMUSG00000035992/ENSMUSG00000036432
ENSMUSG00000026836/ENSMUSG00000031548/ENSMUSG00000074934

ENSMUSG00000005533/ENSMUSG00000081683/ENSMUSG00000005087/ENSMUSG00000023951/ENSMUSG00000044337/ENSMUSG00000061436/ENSMUSG00000032717/ENSMUSG00000050965/ENSMUS
ENSMUSG00000046223/ENSMUSG00000023951/ENSMUSG00000027834/ENSMUSG00000047281/ENSMUSG00000020044/ENSMUSG00000032487/ENSMUSG00000035992/ENSMUSG00000029249/ENSMUS

ENSMUSG00000046223/ENSMUSG00000044337/ENSMUSG00000061436/ENSMUSG00000047281/ENSMUSG00000006445/ENSMUSG00000021070/ENSMUSG00000021831/ENSMUSG00000050965/ENSMUS
ENSMUSG00000085795/ENSMUSG00000063382/ENSMUSG00000074934

;G00000021070/ENSMUSG00000022037/ENSMUSG00000032487/ENSMUSG00000000282/ENSMUSG00000027230/ENSMUSG00000026836/ENSMUSG00000028179/ENSMUSG00000032350/ENSMUSG0000(
;G00000029361/ENSMUSG00000024935/ENSMUSG00000024431/ENSMUSG00000020684/ENSMUSG00000031393/ENSMUSG00000019943

;G00000029361/ENSMUSG00000024935/ENSMUSG00000024431/ENSMUSG00000020684/ENSMUSG00000031393/ENSMUSG00000019943

;G00000032717/ENSMUSG00000109511/ENSMUSG00000050965/ENSMUSG00000021149/ENSMUSG00000062232/ENSMUSG00000053716/ENSMUSG00000033717/ENSMUSG00000000184/ENSMUSG0000(

;G00000039952/ENSMUSG00000041570/ENSMUSG00000026797/ENSMUSG00000026826/ENSMUSG00000028661/ENSMUSG00000021728/ENSMUSG00000005871

;G00000031393/ENSMUSG00000023027/ENSMUSG00000056071/ENSMUSG00000025372

;G00000022365/ENSMUSG00000062232/ENSMUSG00000029249/ENSMUSG00000044147/ENSMUSG00000002658/ENSMUSG00000031393/ENSMUSG00000009630

;G00000036564/ENSMUSG00000062232/ENSMUSG00000029816/ENSMUSG00000000058/ENSMUSG00000033717/ENSMUSG00000039844/ENSMUSG00000028661/ENSMUSG00000004207/ENSMUSG0000(
;G00000035900/ENSMUSG00000036432/ENSMUSG00000056071/ENSMUSG00000002083

;G00000022037/ENSMUSG00000032487/ENSMUSG00000027230/ENSMUSG00000015839/ENSMUSG00000030609/ENSMUSG00000009291/ENSMUSG00000056071/ENSMUSG00000002083/ENSMUSG0000(

)015839/ENSMUSG00000028211/ENSMUSG00000031548/ENSMUSG00000009291/ENSMUSG00000036432/ENSMUSG00000056071/ENSMUSG00000026826/ENSMUSG00000002083/ENSMUSG0000002480

)031548/ENSMUSG00000024182/ENSMUSG00000005871

)024182

)024807

7/ENSMUSG00000026628

| Category | GOID | Description | GeneRatio | BgRatio | pvalue | padj | geneID | geneName |
|---|---|---|---|---|---|---|---|---|
| BP | GO:00020( | epithelial cell development | 37\|1209 | 223\|16097 | 4.00E-06 | 0.00012 | ENSMUS | Sox9/Tnmd/Fgfr1/Fem1b/Met/Cdh5/Rtn4/Pecam1/Col18a1/Fgf |
| BP | GO:00018 | endothelial cell development | 17\|1209 | 65\|16097 | 4.05E-06 | 0.00012 | ENSMUS | Tnmd/Met/Cdh5/Rtn4/Pecam1/Col18a1/Fasn/Clic4/Tnfrsf1a/Pl |
| BP | GO:00360( | positive regulation of transcription from RNA polymerase II pro | 10\|1209 | 24\|16097 | 4.07E-06 | 0.00012 | ENSMUS | Atf4/Eif2ak3/Ddit3/Sesn2/Cebpb/Vegfa/Notch1/Creb3l1/Nfe2l2 |
| BP | GO:00713 | cellular response to lipid | 60\|1209 | 439\|16097 | 4.28E-06 | 0.00013 | ENSMUS | Ldlr/Sox9/Serpine1/Trib1/Inhba/Cd14/Lpl/Eif4ebp1/Atp1a1/Nf |
| BP | GO:00349 | response to endoplasmic reticulum stress | 38\|1209 | 233\|16097 | 4.54E-06 | 0.00014 | ENSMUS | Thbs1/Chac1/Herpud1/Dnajb9/Ppp1r15b/Atf4/Jun/Nfe2l1/Eif2 |
| BP | GO:00310 | regeneration | 27\|1209 | 141\|16097 | 5.40E-06 | 0.00016 | ENSMUS | Sulf2/Hmox1/Serpine1/Klf4/Xylt1/Ifrd1/Jun/Rtn4/Cdkn1a/Thy1 |
| BP | GO:00019 | endochondral ossification | 11\|1209 | 30\|16097 | 5.90E-06 | 0.00018 | ENSMUS | Col1a1/Mmp14/Nab2/Bmp6/Mef2d/Nab1/Hspg2/Csgalnact1/M |
| BP | GO:00360 | replacement ossification | 11\|1209 | 30\|16097 | 5.90E-06 | 0.00018 | ENSMUS | Col1a1/Mmp14/Nab2/Bmp6/Mef2d/Nab1/Hspg2/Csgalnact1/M |
| BP | GO:00350 | cardiocyte differentiation | 31\|1209 | 175\|16097 | 6.11E-06 | 0.00018 | ENSMUS | Col14a1/Rgs2/Met/Sorbs2/Lmna/Rxra/Maml1/Myh11/Tgfbr3/E |
| BP | GO:00600 | canonical Wnt signaling pathway | 41\|1209 | 263\|16097 | 6.14E-06 | 0.00018 | ENSMUS | Col1a1/Sulf2/Myc/Sox9/Klf4/Egr1/Ddit3/Fgfr2/Dixdc1/Egfr/Tnk |
| BP | GO:00092 | cellular response to starvation | 26\|1209 | 134\|16097 | 6.24E-06 | 0.00018 | ENSMUS | Slc2a1/Atf4/Jun/Cdkn1a/Srebf2/Eif2ak3/Sesn2/Mybbp1a/Klf10/ |
| BP | GO:00359 | response to topologically incorrect protein | 26\|1209 | 134\|16097 | 6.24E-06 | 0.00018 | ENSMUS | Hspa1b/Hspa1a/Thbs1/Chac1/Herpud1/Hspa9/Dnajb9/Atf4/Eif2 |
| BP | GO:00850 | extracellular matrix assembly | 10\|1209 | 25\|16097 | 6.32E-06 | 0.00018 | ENSMUS | Col1a2/Tnxb/Sox9/Mfap4/Has2/Myh11/Lox/Fbln5/Has1/Notch |
| BP | GO:00148 | striated muscle hypertrophy | 22\|1209 | 103\|16097 | 6.38E-06 | 0.00018 | ENSMUS | Errfi1/Col14a1/Igfbp5/Rgs2/Sorbs2/Lmna/Atp2a2/Mef2a/Tiam1 |
| BP | GO:00060 | proteoglycan metabolic process | 18\|1209 | 74\|16097 | 6.49E-06 | 0.00019 | ENSMUS | Sulf2/Xylt1/Csgalnact2/Chpf/Chst11/Cytl1/Csgalnact1/Pxylp1/S |
| CC | GO:00156 | actin cytoskeleton | 63\|1232 | 472\|16313 | 6.51E-06 | 0.00022 | ENSMUS | Acta1/Myl1/Mylpf/Marcks/Cald1/Slc2a1/Ahnak/Iqgap1/Ablim3, |
| BP | GO:00456 | regulation of glial cell differentiation | 19\|1209 | 82\|16097 | 7.78E-06 | 0.00022 | ENSMUS | Ldlr/Tlr2/Rtn4/Hmga2/Il6st/Nr1d1/Enpp2/Nog/Rela/Dusp10/N |
| CC | GO:00059 | cell-cell junction | 60\|1232 | 445\|16313 | 7.86E-06 | 0.00025 | ENSMUS | Itga5/Pacsin2/Slc2a1/Cdh5/Ahnak/Atp1a1/Iqgap1/Pecam1/Cdh |
| BP | GO:00359 | skeletal muscle cell differentiation | 18\|1209 | 75\|16097 | 7.95E-06 | 0.00022 | ENSMUS | Myc/Cdon/Hivep3/Egr1/Plagl1/Fos/Mef2d/Btg2/Nr4a1/S100b/N |
| BP | GO:00457 | positive regulation of protein catabolic process | 34\|1209 | 204\|16097 | 8.81E-06 | 0.00025 | ENSMUS | Hspa1b/Hspa1a/Ldlr/Sox9/Trib1/Herpud1/Zfand2a/Ier3/Wwp2, |
| BP | GO:00148 | muscle hypertrophy | 22\|1209 | 105\|16097 | 8.85E-06 | 0.00025 | ENSMUS | Errfi1/Col14a1/Igfbp5/Rgs2/Sorbs2/Lmna/Atp2a2/Mef2a/Tiam1 |
| MF | GO:00504 | transforming growth factor beta binding | 9\|1190 | 21\|15912 | 9.15E-06 | 0.00059 | ENSMUS | Thbs1/Vasn/Cd109/Tgfbr3/Ltbp1/Cd36/Tgfbr1/Twsg1/Acvr1 |
| BP | GO:00305( | BMP signaling pathway | 27\|1209 | 145\|16097 | 9.29E-06 | 0.00026 | ENSMUS | Egr1/Cdh5/Chrdl1/Cyr61/Smad5/Bmp6/Tgfbr3/Neo1/Ski/Sfrp2/ |
| BP | GO:00302( | chondroitin sulfate metabolic process | 9\|1209 | 21\|16097 | 9.48E-06 | 0.00026 | ENSMUS | Xylt1/Chpf/Chst11/Csgalnact1/Slc35d1/Ugdh/Eglam/Spock2/N |
| BP | GO:19015 | positive regulation of transcription from RNA polymerase II pro | 9\|1209 | 21\|16097 | 9.48E-06 | 0.00026 | ENSMUS | Atf4/Smad5/Rxra/Sesn2/Xbp1/Rela/Vegfa/Notch1/Nfe2l2 |
| BP | GO:00140 | regulation of gliogenesis | 26\|1209 | 137\|16097 | 9.49E-06 | 0.00026 | ENSMUS | Ldlr/Myc/Tlr2/Rtn4/Hmga2/Tiam1/Il6st/Ski/Nr1d1/Enpp2/Nog/ |
| BP | GO:00603 | trabecula formation | 10\|1209 | 26\|16097 | 9.59E-06 | 0.00026 | ENSMUS | Col1a1/Chad/Tgfbr3/Vegfa/Adgrg6/Adamts1/Tek/Thbs3/Sfrp1/G |
| BP | GO:00434( | regulation of MAP kinase activity | 42\|1209 | 277\|16097 | 9.63E-06 | 0.00026 | ENSMUS | Spry2/Tnxb/Thbs1/Hmgcr/Rgs2/Map2k3/Trib1/Gadd45a/Fgfr1/ |
| BP | GO:00018 | mesonephros development | 21\|1209 | 98\|16097 | 9.76E-06 | 0.00026 | ENSMUS | Myc/Sox9/Fgfr1/Vcan/Smad5/Fgfr2/Lzts2/Agtr2/Maged1/Ren1/ |
| BP | GO:00321( | negative regulation of response to external stimulus | 45\|1209 | 306\|16097 | 1.05E-05 | 0.00028 | ENSMUS | Thbs1/Hmgcr/Ldlr/Serpine1/Trib1/Xylt1/Serpinf1/Sec14l1/Dus |
| BP | GO:00060 | aminoglycan metabolic process | 20\|1209 | 91\|16097 | 1.06E-05 | 0.00028 | ENSMUS | Xylt1/Has2/Tmem2/Chpf/Itih5/Chst11/B4galt5/Csgalnact1/Pxyl |
| BP | GO:00071 | negative regulation of cell adhesion | 41\|1209 | 269\|16097 | 1.08E-05 | 0.00029 | ENSMUS | Col1a1/Thbs1/Postn/Serpine1/Klf4/Plet1/Lamb1/Dusp1/Lgals3, |
| BP | GO:00085 | epidermis development | 40\|1209 | 260\|16097 | 1.10E-05 | 0.00029 | ENSMUS | Errfi1/Igfbp5/Sox9/Klf4/Inhba/Fgfr1/Cd109/Mafg/Ncoa3/Fgfr2/ |
| BP | GO:00608 | regulation of canonical Wnt signaling pathway | 35\|1209 | 215\|16097 | 1.11E-05 | 0.00029 | ENSMUS | Col1a1/Sulf2/Sox9/Egr1/Ddit3/Fgfr2/Egfr/Tnks2/Lzts2/Sfrp2/No |
| MF | GO:00191 | transmembrane receptor protein kinase activity | 17\|1190 | 70\|15912 | 1.13E-05 | 0.00068 | ENSMUS | Fgfr1/Met/Fgfr2/Tgfbr3/Egfr/Nrp1/Tgfbr1/Igf1r/Flt4/Axl/Epha2/ |
| BP | GO:00033( | cardiac muscle hypertrophy | 21\|1209 | 99\|16097 | 1.15E-05 | 0.0003 | ENSMUS | Errfi1/Col14a1/Rgs2/Sorbs2/Lmna/Atp2a2/Mef2a/Tiam1/Agtr2/ |
| BP | GO:00487 | mesenchymal cell differentiation | 34\|1209 | 207\|16097 | 1.21E-05 | 0.00032 | ENSMUS | Col1a1/Vasn/Sox9/Fgfr1/Has2/Efna1/Rtn4/Phldb1/Fgfr2/Tgfbr3 |
| BP | GO:00107 | positive regulation of cell morphogenesis involved in differenti | 31\|1209 | 181\|16097 | 1.24E-05 | 0.00032 | ENSMUS | Has2/S100a10/Tnfrsf12a/Met/Rtn4/Golga4/Dixdc1/Sgk1/Tiam1 |
| BP | GO:00550( | cardiac muscle cell differentiation | 26\|1209 | 139\|16097 | 1.24E-05 | 0.00032 | ENSMUS | Col14a1/Rgs2/Met/Sorbs2/Lmna/Rxra/Maml1/Myh11/Mef2a/A |
| BP | GO:00720 | kidney epithelium development | 25\|1209 | 131\|16097 | 1.27E-05 | 0.00033 | ENSMUS | Myc/Sox9/Fgfr1/Vcan/Smad5/Fgfr2/Lzts2/Agtr2/Maged1/Nog/C |
| BP | GO:00456 | positive regulation of neuron differentiation | 57\|1209 | 425\|16097 | 1.31E-05 | 0.00034 | ENSMUS | Rgs2/Cdon/Serpinf1/Fgfr1/Mapk6/Tnfrsf12a/Met/Rtn4/Iqgap1/ |
| BP | GO:00421 | cellular response to glucose starvation | 12\|1209 | 38\|16097 | 1.32E-05 | 0.00034 | ENSMUS | Slc2a1/Atf4/Eif2ak3/Sesn2/Mybbp1a/Mtmr3/Xbp1/Sik1/Asns/Ir |
| BP | GO:00550( | muscle cell development | 33\|1209 | 199\|16097 | 1.32E-05 | 0.00034 | ENSMUS | Acta1/Col14a1/Rgs2/Met/Rcan1/Sorbs2/Lmna/Atp2a2/Maml1/ |
| BP | GO:00514( | stress-activated MAPK cascade | 38\|1209 | 244\|16097 | 1.36E-05 | 0.00034 | ENSMUS | Tnxb/Hmgcr/Map2k3/Trib1/Gadd45a/Dusp1/Met/Dixdc1/Nrk/ |
| BP | GO:00510 | negative regulation of transport | 64\|1209 | 496\|16097 | 1.38E-05 | 0.00035 | ENSMUS | Gem/Thbs1/Hmox1/Hmgcr/Myc/Tlr2/Rgs2/Cyp51/Igfbp3/Pacsi |
| BP | GO:00316 | cellular response to nutrient levels | 31\|1209 | 182\|16097 | 1.39E-05 | 0.00035 | ENSMUS | Postn/Lpl/Slc2a1/Atf4/Jun/Cdkn1a/Srebf2/Eif2ak3/Sesn2/Mybb |
| BP | GO:00302( | glycosaminoglycan metabolic process | 18\|1209 | 78\|16097 | 1.42E-05 | 0.00035 | ENSMUS | Xylt1/Has2/Tmem2/Chpf/Itih5/Chst11/Csgalnact1/Pxylp1/Slc3 |
| BP | GO:19033 | regulation of adherens junction organization | 16\|1209 | 64\|16097 | 1.44E-05 | 0.00036 | ENSMUS | S100a10/Iqgap1/Mmp14/Bmp6/Thy1/Nrp1/Dlc1/Enpp2/Vegfa/ |
| BP | GO:00074( | axonogenesis | 57\|1209 | 427\|16097 | 1.51E-05 | 0.00037 | ENSMUS | Ifrd1/Tnfrsf12a/Efna1/Rtn4/Lama2/Golga4/Cdh11/Fgfr2/Dixdc1 |
| BP | GO:19016 | cellular response to peptide | 41\|1209 | 273\|16097 | 1.55E-05 | 0.00038 | ENSMUS | Lpl/Ywhag/Eif4ebp1/Socs3/Sgk1/Stat3/Trib3/Xbp1/Ndel1/Nr4a |
| BP | GO:00352 | exocrine system development | 14\|1209 | 51\|16097 | 1.56E-05 | 0.00038 | ENSMUS | Sox9/Fgfr1/Met/Fgfr2/Egfr/Xbp1/Nrp1/Igf1r/Igf1/Twsg1/Plxnd1 |

| Count | Up | Up_Gene_id | Down | Down_Gene_id |
|---|---|---|---|---|
| 37 | 10 | ENSMUSG0 | 27 | ENSMUSG00000000567/ENSMUSG00000031565/ENSMUSG00000032244/ENSMUSG00000009376/ENSMUSG00000020458/ENSMUSG00000030849/ENSMUSG0 |
| 17 | 6 | ENSMUSG0 | 11 | ENSMUSG00000009376/ENSMUSG00000020458/ENSMUSG00000025153/ENSMUSG00000037242/ENSMUSG00000030341/ENSMUSG00000004846/ENSMUSG0 |
| 10 | 1 | ENSMUSG0 | 9 | ENSMUSG00000042406/ENSMUSG00000031668/ENSMUSG00000025408/ENSMUSG00000028893/ENSMUSG00000056501/ENSMUSG00000023951/ENSMUSG0 |
| 60 | 14 | ENSMUSG0 | 46 | ENSMUSG00000032193/ENSMUSG00000000567/ENSMUSG00000037411/ENSMUSG00000032501/ENSMUSG00000041324/ENSMUSG00000051439/ENSMUSG0 |
| 38 | 5 | ENSMUSG0 | 33 | ENSMUSG00000040152/ENSMUSG00000027313/ENSMUSG00000031770/ENSMUSG00000014905/ENSMUSG00000046062/ENSMUSG00000042406/ENSMUSG0 |
| 27 | 13 | ENSMUSG0 | 14 | ENSMUSG00000006800/ENSMUSG00000005413/ENSMUSG00000037411/ENSMUSG00000003032/ENSMUSG00000030657/ENSMUSG00000001627/ENSMUSG0 |
| 11 | 5 | ENSMUSG0 | 6 | ENSMUSG00000025402/ENSMUSG00000039004/ENSMUSG00000001419/ENSMUSG00000002881/ENSMUSG00000036356/ENSMUSG00000034135 |
| 11 | 5 | ENSMUSG0 | 6 | ENSMUSG00000025402/ENSMUSG00000039004/ENSMUSG00000001419/ENSMUSG00000002881/ENSMUSG00000036356/ENSMUSG00000034135 |
| 31 | 15 | ENSMUSG0 | 16 | ENSMUSG00000026360/ENSMUSG00000009376/ENSMUSG00000028063/ENSMUSG00000015846/ENSMUSG00000050567/ENSMUSG00000029287/ENSMUSG0 |
| 41 | 14 | ENSMUSG0 | 27 | ENSMUSG00000006800/ENSMUSG00000022346/ENSMUSG00000000567/ENSMUSG00000003032/ENSMUSG00000038418/ENSMUSG00000025408/ENSMUSG0 |
| 26 | 6 | ENSMUSG0 | 20 | ENSMUSG00000028645/ENSMUSG00000042406/ENSMUSG00000023067/ENSMUSG00000022463/ENSMUSG00000031668/ENSMUSG00000028893/ENSMUSG0 |
| 26 | 5 | ENSMUSG0 | 21 | ENSMUSG00000040152/ENSMUSG00000027313/ENSMUSG00000031770/ENSMUSG00000024359/ENSMUSG00000014905/ENSMUSG00000042406/ENSMUSG0 |
| 10 | 7 | ENSMUSG0 | 3 | ENSMUSG00000000567/ENSMUSG00000022367/ENSMUSG00000003665 |
| 22 | 9 | ENSMUSG0 | 13 | ENSMUSG00000028967/ENSMUSG00000026360/ENSMUSG00000028063/ENSMUSG00000029467/ENSMUSG00000030557/ENSMUSG00000002489/ENSMUSG0 |
| 18 | 7 | ENSMUSG0 | 11 | ENSMUSG00000006800/ENSMUSG00000030657/ENSMUSG00000042042/ENSMUSG00000032997/ENSMUSG00000036356/ENSMUSG00000043587/ENSMUSG0 |
| 63 | 38 | ENSMUSG0 | 25 | ENSMUSG00000028645/ENSMUSG00000069833/ENSMUSG00000030536/ENSMUSG00000026596/ENSMUSG00000022241/ENSMUSG00000031523/ENSMUSG0 |
| 19 | 5 | ENSMUSG0 | 14 | ENSMUSG00000032193/ENSMUSG00000027995/ENSMUSG00000020458/ENSMUSG00000056758/ENSMUSG00000021756/ENSMUSG00000020889/ENSMUSG0 |
| 60 | 22 | ENSMUSG0 | 38 | ENSMUSG00000000555/ENSMUSG00000016664/ENSMUSG00000028645/ENSMUSG00000069833/ENSMUSG00000033161/ENSMUSG00000030536/ENSMUSG0 |
| 18 | 5 | ENSMUSG0 | 13 | ENSMUSG00000022346/ENSMUSG00000038119/ENSMUSG00000028634/ENSMUSG00000038418/ENSMUSG00000019817/ENSMUSG00000001419/ENSMUSG0 |
| 34 | 9 | ENSMUSG0 | 25 | ENSMUSG00000032193/ENSMUSG00000000567/ENSMUSG00000032501/ENSMUSG00000031770/ENSMUSG00000053581/ENSMUSG00000003541/ENSMUSG0 |
| 22 | 9 | ENSMUSG0 | 13 | ENSMUSG00000028967/ENSMUSG00000026360/ENSMUSG00000028063/ENSMUSG00000029467/ENSMUSG00000030557/ENSMUSG00000002489/ENSMUSG0 |
| 9 | 2 | ENSMUSG0 | 7 | ENSMUSG00000040152/ENSMUSG00000039646/ENSMUSG00000046186/ENSMUSG00000029287/ENSMUSG00000007613/ENSMUSG00000024098/ENSMUSG0 |
| 27 | 10 | ENSMUSG0 | 17 | ENSMUSG00000038418/ENSMUSG00000021540/ENSMUSG00000039004/ENSMUSG00000029287/ENSMUSG00000032340/ENSMUSG00000029050/ENSMUSG0 |
| 9 | 4 | ENSMUSG0 | 5 | ENSMUSG00000030657/ENSMUSG00000032997/ENSMUSG00000036356/ENSMUSG00000028521/ENSMUSG00000029201 |
| 9 | 1 | ENSMUSG0 | 8 | ENSMUSG00000042406/ENSMUSG00000021540/ENSMUSG00000015846/ENSMUSG00000028893/ENSMUSG00000020484/ENSMUSG00000024927/ENSMUSG0 |
| 26 | 8 | ENSMUSG0 | 18 | ENSMUSG00000032193/ENSMUSG00000022346/ENSMUSG00000027995/ENSMUSG00000020458/ENSMUSG00000056758/ENSMUSG00000002489/ENSMUSG0 |
| 10 | 4 | ENSMUSG0 | 6 | ENSMUSG00000039084/ENSMUSG00000029287/ENSMUSG00000023951/ENSMUSG00000028047/ENSMUSG00000031548/ENSMUSG00000074934 |
| 42 | 10 | ENSMUSG0 | 32 | ENSMUSG00000022114/ENSMUSG00000040152/ENSMUSG00000021670/ENSMUSG00000026360/ENSMUSG00000018932/ENSMUSG00000032501/ENSMUSG0 |
| 21 | 8 | ENSMUSG0 | 13 | ENSMUSG00000022346/ENSMUSG00000000567/ENSMUSG00000031565/ENSMUSG00000021540/ENSMUSG00000030849/ENSMUSG00000035342/ENSMUSG0 |
| 45 | 19 | ENSMUSG0 | 26 | ENSMUSG00000040152/ENSMUSG00000021670/ENSMUSG00000032193/ENSMUSG00000037411/ENSMUSG00000032501/ENSMUSG00000030657/ENSMUSG0 |
| 20 | 6 | ENSMUSG0 | 14 | ENSMUSG00000030657/ENSMUSG00000022367/ENSMUSG00000024754/ENSMUSG00000032997/ENSMUSG00000017929/ENSMUSG00000036356/ENSMUSG0 |
| 41 | 12 | ENSMUSG0 | 29 | ENSMUSG00000040152/ENSMUSG00000037411/ENSMUSG00000003032/ENSMUSG00000032068/ENSMUSG00000050335/ENSMUSG00000026596/ENSMUSG0 |
| 40 | 10 | ENSMUSG0 | 30 | ENSMUSG00000028967/ENSMUSG00000000567/ENSMUSG00000003032/ENSMUSG00000041324/ENSMUSG00000031565/ENSMUSG00000046186/ENSMUSG0 |
| 35 | 13 | ENSMUSG0 | 22 | ENSMUSG00000006800/ENSMUSG00000000567/ENSMUSG00000038418/ENSMUSG00000025408/ENSMUSG00000030849/ENSMUSG00000024811/ENSMUSG0 |
| 17 | 8 | ENSMUSG0 | 9 | ENSMUSG00000031565/ENSMUSG00000009376/ENSMUSG00000030849/ENSMUSG00000029287/ENSMUSG00000007613/ENSMUSG00000005533/ENSMUSG0 |
| 21 | 8 | ENSMUSG0 | 13 | ENSMUSG00000028967/ENSMUSG00000026360/ENSMUSG00000028063/ENSMUSG00000029467/ENSMUSG00000030557/ENSMUSG00000002489/ENSMUSG0 |
| 34 | 11 | ENSMUSG0 | 23 | ENSMUSG00000039646/ENSMUSG00000000567/ENSMUSG00000031565/ENSMUSG00000022367/ENSMUSG00000027954/ENSMUSG00000020458/ENSMUSG0 |
| 31 | 13 | ENSMUSG0 | 18 | ENSMUSG00000022367/ENSMUSG00000041959/ENSMUSG00000023905/ENSMUSG00000009376/ENSMUSG00000020458/ENSMUSG00000038708/ENSMUSG0 |
| 26 | 12 | ENSMUSG0 | 14 | ENSMUSG00000026360/ENSMUSG00000009376/ENSMUSG00000028063/ENSMUSG00000015846/ENSMUSG00000050567/ENSMUSG00000030557/ENSMUSG0 |
| 25 | 12 | ENSMUSG0 | 13 | ENSMUSG00000022346/ENSMUSG00000000567/ENSMUSG00000031565/ENSMUSG00000021540/ENSMUSG00000030849/ENSMUSG00000035342/ENSMUSG0 |
| 57 | 16 | ENSMUSG0 | 41 | ENSMUSG00000026360/ENSMUSG00000038119/ENSMUSG00000000753/ENSMUSG00000031565/ENSMUSG00000042688/ENSMUSG00000023905/ENSMUSG0 |
| 12 | 1 | ENSMUSG0 | 11 | ENSMUSG00000028645/ENSMUSG00000042406/ENSMUSG00000031668/ENSMUSG00000028893/ENSMUSG00000040463/ENSMUSG00000034354/ENSMUSG0 |
| 33 | 17 | ENSMUSG0 | 16 | ENSMUSG00000026360/ENSMUSG00000009376/ENSMUSG00000022951/ENSMUSG00000028063/ENSMUSG00000029467/ENSMUSG00000050567/ENSMUSG0 |
| 38 | 14 | ENSMUSG0 | 24 | ENSMUSG00000021670/ENSMUSG00000018932/ENSMUSG00000032501/ENSMUSG00000036390/ENSMUSG00000009376/ENSMUSG00000032064/ENSMUSG0 |
| 64 | 25 | ENSMUSG0 | 39 | ENSMUSG00000028214/ENSMUSG00000040152/ENSMUSG00000005413/ENSMUSG00000021670/ENSMUSG00000022346/ENSMUSG00000027995/ENSMUSG0 |
| 31 | 9 | ENSMUSG0 | 22 | ENSMUSG00000028645/ENSMUSG00000042406/ENSMUSG00000023067/ENSMUSG00000022463/ENSMUSG00000031668/ENSMUSG00000028893/ENSMUSG0 |
| 18 | 5 | ENSMUSG0 | 13 | ENSMUSG00000030657/ENSMUSG00000022367/ENSMUSG00000024754/ENSMUSG00000032997/ENSMUSG00000036356/ENSMUSG00000043587/ENSMUSG0 |
| 16 | 6 | ENSMUSG0 | 10 | ENSMUSG00000041959/ENSMUSG00000030536/ENSMUSG00000039004/ENSMUSG00000031523/ENSMUSG00000022425/ENSMUSG00000023951/ENSMUSG0 |
| 57 | 23 | ENSMUSG0 | 34 | ENSMUSG00000001627/ENSMUSG00000023905/ENSMUSG00000027954/ENSMUSG00000020458/ENSMUSG00000038708/ENSMUSG00000030849/ENSMUSG0 |
| 41 | 11 | ENSMUSG0 | 30 | ENSMUSG00000051391/ENSMUSG00000031490/ENSMUSG00000019970/ENSMUSG00000004040/ENSMUSG00000032715/ENSMUSG00000020484/ENSMUSG0 |
| 14 | 4 | ENSMUSG0 | 10 | ENSMUSG00000000567/ENSMUSG00000031565/ENSMUSG00000009376/ENSMUSG00000030849/ENSMUSG00000020484/ENSMUSG00000005533/ENSMUSG0 |

0000039004/ENSMUSG00000025153/ENSMUSG00000020484/ENSMUSG00000037242/ENSMUSG00000029135/ENSMUSG00000030341/ENSMUSG00000026728/ENSMUSG00000004846/ENSMUSG0000002
0000023951/ENSMUSG00000024812/ENSMUSG00000022016/ENSMUSG00000062232/ENSMUSG00000039844
0000027230/ENSMUSG00000015839/ENSMUSG00000026628
0000031490/ENSMUSG00000033161/ENSMUSG00000038615/ENSMUSG00000027678/ENSMUSG00000026596/ENSMUSG00000056758/ENSMUSG00000045294/ENSMUSG00000019970/ENSMUSG0000003
0000038615/ENSMUSG00000031668/ENSMUSG00000025408/ENSMUSG00000032115/ENSMUSG00000028893/ENSMUSG00000032715/ENSMUSG00000020484/ENSMUSG00000014959/ENSMUSG0000005
0000020458/ENSMUSG00000023067/ENSMUSG00000018736/ENSMUSG00000024789/ENSMUSG00000032340/ENSMUSG00000005533/ENSMUSG00000056501/ENSMUSG00000039384

0000030557/ENSMUSG00000033323/ENSMUSG00000023951/ENSMUSG00000022752/ENSMUSG00000018476/ENSMUSG00000026836/ENSMUSG00000029249/ENSMUSG00000024431/ENSMUSG0000007
0000030849/ENSMUSG00000032064/ENSMUSG00000024811/ENSMUSG00000035342/ENSMUSG00000027996/ENSMUSG00000048616/ENSMUSG00000081683/ENSMUSG00000040021/ENSMUSG0000001
0000040463/ENSMUSG00000037465/ENSMUSG00000034354/ENSMUSG00000020484/ENSMUSG00000029752/ENSMUSG00000031451/ENSMUSG00000024423/ENSMUSG00000034201/ENSMUSG0000003
0000031668/ENSMUSG00000025408/ENSMUSG00000020484/ENSMUSG00000031849/ENSMUSG00000021831/ENSMUSG00000048249/ENSMUSG00000043019/ENSMUSG00000003814/ENSMUSG0000002

0000030341/ENSMUSG00000033323/ENSMUSG00000022752/ENSMUSG00000028161/ENSMUSG00000050965/ENSMUSG00000044167/ENSMUSG00000024431
0000028521/ENSMUSG00000045216/ENSMUSG00000021978/ENSMUSG00000029201/ENSMUSG00000039497
0000032232/ENSMUSG00000070462/ENSMUSG00000037815/ENSMUSG00000054808/ENSMUSG00000022443/ENSMUSG00000032643/ENSMUSG00000040852/ENSMUSG00000026313/ENSMUSG0000003
0000022425/ENSMUSG00000048616/ENSMUSG00000024927/ENSMUSG00000039384/ENSMUSG00000021379/ENSMUSG00000039952/ENSMUSG00000038539/ENSMUSG00000031393
0000029467/ENSMUSG00000046798/ENSMUSG00000002489/ENSMUSG00000039701/ENSMUSG00000031714/ENSMUSG00000037242/ENSMUSG00000032232/ENSMUSG00000007613/ENSMUSG0000000
0000020423/ENSMUSG00000023034/ENSMUSG00000034998/ENSMUSG00000048661/ENSMUSG00000044857/ENSMUSG00000063382/ENSMUSG00000026628
0000025373/ENSMUSG00000032715/ENSMUSG00000020484/ENSMUSG00000020601/ENSMUSG00000034640/ENSMUSG00000048249/ENSMUSG00000022037/ENSMUSG00000025234/ENSMUSG0000003
0000030341/ENSMUSG00000033323/ENSMUSG00000022752/ENSMUSG00000028161/ENSMUSG00000050965/ENSMUSG00000044167/ENSMUSG00000024431
0000026836
0000027996/ENSMUSG00000031849/ENSMUSG00000048616/ENSMUSG00000024098/ENSMUSG00000042377/ENSMUSG00000061436/ENSMUSG00000048661/ENSMUSG00000026836/ENSMUSG0000003

0000023951/ENSMUSG00000015839
0000021756/ENSMUSG00000029050/ENSMUSG00000020889/ENSMUSG00000022425/ENSMUSG00000048616/ENSMUSG00000026728/ENSMUSG00000024927/ENSMUSG00000039384/ENSMUSG0000002

0000036390/ENSMUSG00000031565/ENSMUSG00000009376/ENSMUSG00000030536/ENSMUSG00000027351/ENSMUSG00000002489/ENSMUSG00000031714/ENSMUSG00000032715/ENSMUSG0000002
0000070645/ENSMUSG00000048616/ENSMUSG00000005087/ENSMUSG00000023951/ENSMUSG00000031548/ENSMUSG00000074934/ENSMUSG00000045394
0000000753/ENSMUSG00000020823/ENSMUSG00000046186/ENSMUSG00000020458/ENSMUSG00000023067/ENSMUSG00000003541/ENSMUSG00000039899/ENSMUSG00000031778/ENSMUSG0000003
0000043587/ENSMUSG00000028521/ENSMUSG00000005087/ENSMUSG00000003665/ENSMUSG00000021978/ENSMUSG00000029201/ENSMUSG00000047126/ENSMUSG00000039497
0000039899/ENSMUSG00000039004/ENSMUSG00000022508/ENSMUSG00000024789/ENSMUSG00000031523/ENSMUSG00000022425/ENSMUSG00000056501/ENSMUSG00000005087/ENSMUSG0000002
0000051510/ENSMUSG00000027678/ENSMUSG00000030849/ENSMUSG00000025402/ENSMUSG00000031714/ENSMUSG00000037242/ENSMUSG00000002881/ENSMUSG00000029135/ENSMUSG0000000
0000035342/ENSMUSG00000027996/ENSMUSG00000048616/ENSMUSG00000040021/ENSMUSG00000018583/ENSMUSG00000032702/ENSMUSG00000073563/ENSMUSG00000044167/ENSMUSG0000003
0000006445/ENSMUSG00000026836/ENSMUSG00000028661
0000030341/ENSMUSG00000033323/ENSMUSG00000022752/ENSMUSG00000028161/ENSMUSG00000050965/ENSMUSG00000044167/ENSMUSG00000024431
0000048537/ENSMUSG00000030849/ENSMUSG00000029287/ENSMUSG00000002489/ENSMUSG00000020349/ENSMUSG00000027996/ENSMUSG00000048616/ENSMUSG00000007613/ENSMUSG0000004
0000032064/ENSMUSG00000019970/ENSMUSG00000002489/ENSMUSG00000028613/ENSMUSG00000018736/ENSMUSG00000022425/ENSMUSG00000023951/ENSMUSG00000020745/ENSMUSG0000003
0000033323/ENSMUSG00000023951/ENSMUSG00000022752/ENSMUSG00000018476/ENSMUSG00000026836/ENSMUSG00000024431/ENSMUSG00000074934/ENSMUSG00000026812
0000048616/ENSMUSG00000005087/ENSMUSG00000023951/ENSMUSG00000031548/ENSMUSG00000074934/ENSMUSG00000045394/ENSMUSG00000032220
0000009376/ENSMUSG00000020458/ENSMUSG00000030536/ENSMUSG00000038708/ENSMUSG00000026596/ENSMUSG00000032064/ENSMUSG00000019970/ENSMUSG00000039004/ENSMUSG0000003
0000020484/ENSMUSG00000029752/ENSMUSG00000024423/ENSMUSG00000015839/ENSMUSG00000050697
0000030557/ENSMUSG00000029050/ENSMUSG00000031849/ENSMUSG00000033323/ENSMUSG00000023951/ENSMUSG00000022752/ENSMUSG00000028161/ENSMUSG00000026313/ENSMUSG0000000
0000002489/ENSMUSG00000019997/ENSMUSG00000031714/ENSMUSG00000027996/ENSMUSG00000035199/ENSMUSG00000005533/ENSMUSG00000081683/ENSMUSG00000023951/ENSMUSG0000003
0000026360/ENSMUSG00000001467/ENSMUSG00000016664/ENSMUSG00000050335/ENSMUSG00000042406/ENSMUSG00000022463/ENSMUSG00000003541/ENSMUSG00000045294/ENSMUSG0000003
0000040463/ENSMUSG00000037465/ENSMUSG00000034354/ENSMUSG00000020484/ENSMUSG00000029752/ENSMUSG00000031451/ENSMUSG00000045983/ENSMUSG00000024423/ENSMUSG0000003
0000028521/ENSMUSG00000005087/ENSMUSG00000003665/ENSMUSG00000021978/ENSMUSG00000029201/ENSMUSG00000047126/ENSMUSG00000039497
0000040945/ENSMUSG00000044147/ENSMUSG00000074934/ENSMUSG00000026812
0000032064/ENSMUSG00000035671/ENSMUSG00000017929/ENSMUSG00000002489/ENSMUSG00000026014/ENSMUSG00000018736/ENSMUSG00000032340/ENSMUSG00000035764/ENSMUSG0000004
0000018736/ENSMUSG00000023034/ENSMUSG00000024789/ENSMUSG00000026728/ENSMUSG00000005533/ENSMUSG00000024927/ENSMUSG00000028042/ENSMUSG00000050965/ENSMUSG0000003
0000024098/ENSMUSG00000037820/ENSMUSG00000039952/ENSMUSG00000041702

23951/ENSMUSG00000020745/ENSMUSG00000024812/ENSMUSG00000027465/ENSMUSG00000047281/ENSMUSG00000006445/ENSMUSG00000074818/ENSMUSG00000022016/ENSMUSG00000062232/E

39004/ENSMUSG00000028525/ENSMUSG00000031778/ENSMUSG00000020484/ENSMUSG00000033863/ENSMUSG00000024789/ENSMUSG00000024413/ENSMUSG00000020889/ENSMUSG00000026361/E
56501/ENSMUSG00000052656/ENSMUSG00000039100/ENSMUSG00000025823/ENSMUSG00000021831/ENSMUSG00000048249/ENSMUSG00000043019/ENSMUSG00000026924/ENSMUSG00000003814/E

74934/ENSMUSG00000026812
18583/ENSMUSG00000032702/ENSMUSG00000073563/ENSMUSG00000044167/ENSMUSG00000031548/ENSMUSG00000039844/ENSMUSG00000036432/ENSMUSG00000085795/ENSMUSG00000063382/E
35992/ENSMUSG00000044167/ENSMUSG00000015839/ENSMUSG00000028646/ENSMUSG00000050697/ENSMUSG00000026628
22365/ENSMUSG00000022037/ENSMUSG00000027230/ENSMUSG00000020571/ENSMUSG00000015839/ENSMUSG00000024807/ENSMUSG00000026628

37351/ENSMUSG00000001630/ENSMUSG00000026131/ENSMUSG00000034201/ENSMUSG00000038859/ENSMUSG00000025372/ENSMUSG00000026812/ENSMUSG00000034216/ENSMUSG00000002365/E

05125/ENSMUSG00000031788/ENSMUSG00000033955/ENSMUSG00000023951/ENSMUSG00000037815/ENSMUSG00000040511/ENSMUSG00000024812/ENSMUSG00000054808/ENSMUSG00000013076/E

33717/ENSMUSG00000028793/ENSMUSG00000044167/ENSMUSG00000032350/ENSMUSG00000042303/ENSMUSG00000038068/ENSMUSG00000009291/ENSMUSG00000024425/ENSMUSG00000024182/E

31548/ENSMUSG00000044857/ENSMUSG00000074934

21379/ENSMUSG00000039952/ENSMUSG00000038539/ENSMUSG00000031393

20601/ENSMUSG00000024789/ENSMUSG00000034765/ENSMUSG00000027996/ENSMUSG00000007613/ENSMUSG00000005533/ENSMUSG00000081683/ENSMUSG00000023951/ENSMUSG00000039384/E

32340/ENSMUSG00000020889/ENSMUSG00000030341/ENSMUSG00000005087/ENSMUSG00000025340/ENSMUSG00000039384/ENSMUSG00000020682/ENSMUSG00000042594/ENSMUSG00000024425/E

24098/ENSMUSG00000023951/ENSMUSG00000036606/ENSMUSG00000054808/ENSMUSG00000042594/ENSMUSG00000028042/ENSMUSG00000032702/ENSMUSG00000021149/ENSMUSG00000079037/E
05533/ENSMUSG00000004846/ENSMUSG00000024927/ENSMUSG00000040021/ENSMUSG00000020745/ENSMUSG00000053797/ENSMUSG00000047281/ENSMUSG00000006445/ENSMUSG00000063142/E
31548/ENSMUSG00000039844/ENSMUSG00000036432/ENSMUSG00000085795/ENSMUSG00000074934/ENSMUSG00000003345/ENSMUSG00000024182/ENSMUSG00000005871

46223/ENSMUSG00000023951/ENSMUSG00000039952/ENSMUSG00000026836/ENSMUSG00000034205/ENSMUSG00000031548/ENSMUSG00000085795/ENSMUSG00000063382/ENSMUSG00000074934
36606/ENSMUSG00000003814/ENSMUSG00000020132/ENSMUSG00000025372

31778/ENSMUSG00000029426/ENSMUSG00000028641/ENSMUSG00000002489/ENSMUSG00000022508/ENSMUSG00000028613/ENSMUSG00000018736/ENSMUSG00000045983/ENSMUSG00000005533/E

00058/ENSMUSG00000024431
39384/ENSMUSG00000020745/ENSMUSG00000027177/ENSMUSG00000061436/ENSMUSG00000032717/ENSMUSG00000044167/ENSMUSG00000031548/ENSMUSG00000039844/ENSMUSG00000037902/E
31778/ENSMUSG00000020349/ENSMUSG00000032340/ENSMUSG00000031451/ENSMUSG00000030341/ENSMUSG00000037434/ENSMUSG00000035199/ENSMUSG00000025340/ENSMUSG00000028161/E
34201/ENSMUSG00000035992/ENSMUSG00000044167/ENSMUSG00000015839/ENSMUSG00000028646/ENSMUSG00000050697/ENSMUSG00000026812/ENSMUSG00000026628

48616/ENSMUSG00000026728/ENSMUSG00000005533/ENSMUSG00000022687/ENSMUSG00000023951/ENSMUSG00000020745/ENSMUSG00000036606/ENSMUSG00000006445/ENSMUSG00000023175/E
32702/ENSMUSG00000018634/ENSMUSG00000079037/ENSMUSG00000000058/ENSMUSG00000038859/ENSMUSG00000024052/ENSMUSG00000044167/ENSMUSG00000020091/ENSMUSG00000042903/E

ENSMUSG00000029249/ENSMUSG00000039844/ENSMUSG00000004207/ENSMUSG00000032220

ENSMUSG00000026728/ENSMUSG00000056501/ENSMUSG00000037447/ENSMUSG00000024927/ENSMUSG00000048490/ENSMUSG00000040021/ENSMUSG00000035673/ENSMUSG00000039202/ENSMUS
ENSMUSG00000022365/ENSMUSG00000022037/ENSMUSG00000027230/ENSMUSG00000020571/ENSMUSG00000015839/ENSMUSG00000009630/ENSMUSG00000028277/ENSMUSG00000002083/ENSMUS

ENSMUSG00000074934/ENSMUSG00000003345/ENSMUSG00000024182/ENSMUSG00000005871

ENSMUSG00000032220/ENSMUSG00000034593

ENSMUSG00000022443/ENSMUSG00000006445/ENSMUSG00000028161/ENSMUSG00000050965/ENSMUSG00000039473/ENSMUSG00000062232/ENSMUSG00000026131/ENSMUSG00000018965/ENSMUS

ENSMUSG00000002365/ENSMUSG00000005871

ENSMUSG00000027177/ENSMUSG00000042594/ENSMUSG00000032717/ENSMUSG00000109511/ENSMUSG00000050965/ENSMUSG00000053716/ENSMUSG00000033717/ENSMUSG00000031548/ENSMUS

ENSMUSG00000074934/ENSMUSG00000037902/ENSMUSG00000027712

ENSMUSG00000040945/ENSMUSG00000056050/ENSMUSG00000029816/ENSMUSG00000024425/ENSMUSG00000085795/ENSMUSG00000045394
ENSMUSG00000039473/ENSMUSG00000074818/ENSMUSG00000032487/ENSMUSG00000029249/ENSMUSG00000004207/ENSMUSG00000005871/ENSMUSG00000034593

ENSMUSG00000023951/ENSMUSG00000027834/ENSMUSG00000020745/ENSMUSG00000036606/ENSMUSG00000024423/ENSMUSG00000024924/ENSMUSG00000036564/ENSMUSG00000062232/ENSMUS

ENSMUSG00000024182
ENSMUSG00000050965/ENSMUSG00000018634/ENSMUSG00000009907/ENSMUSG00000032487/ENSMUSG00000033717/ENSMUSG00000029249/ENSMUSG00000032350/ENSMUSG00000029361/ENSMUS

ENSMUSG00000050965/ENSMUSG00000060012/ENSMUSG00000025582/ENSMUSG00000020132/ENSMUSG00000026131/ENSMUSG00000039952/ENSMUSG00000026797/ENSMUSG00000026826/ENSMUS
ENSMUSG00000019943/ENSMUSG00000026826/ENSMUSG00000050697/ENSMUSG00000026812/ENSMUSG00000027712/ENSMUSG00000005871/ENSMUSG00000034593

G00000048249/ENSMUSG00000030747/ENSMUSG00000050965/ENSMUSG00000003814/ENSMUSG00000022037/ENSMUSG00000016024/ENSMUSG00000018965/ENSMUSG00000029249/ENSMUSG0000
G00000024807/ENSMUSG00000026628

G00000039952/ENSMUSG00000042303/ENSMUSG00000062901/ENSMUSG00000027712/ENSMUSG00000045394/ENSMUSG00000005871/ENSMUSG00000032220

G00000024182

G00000020132/ENSMUSG00000029249/ENSMUSG00000015839/ENSMUSG00000042211/ENSMUSG00000024431/ENSMUSG00000039844/ENSMUSG00000031393/ENSMUSG00000023027/ENSMUSG0000

G00000031548/ENSMUSG00000035828/ENSMUSG00000024425/ENSMUSG00000028277/ENSMUSG00000037902/ENSMUSG00000035621/ENSMUSG00000027712/ENSMUSG00000035640

G00000028661/ENSMUSG00000021728/ENSMUSG00000005871

)031548/ENSMUSG00000024431/ENSMUSG00000085795/ENSMUSG00000025326/ENSMUSG00000050697/ENSMUSG00000037902/ENSMUSG00000005886

)056071/ENSMUSG00000025372

| Category | GOID | Description | GeneRatio | BgRatio | pvalue | padj | geneID | geneName |
|---|---|---|---|---|---|---|---|---|
| BP | GO:004349 | protein kinase B signaling | 30\|1209 | 175\|16097 | 1.67E-05 | 0.00041 | ENSMUS | Spry2/F3/Thbs1/Igfbp5/Sox9/Klf4/Rtn4/Ddit3/Sesn2/Rnf41/Cx3 |
| BP | GO:004871 | regulation of astrocyte differentiation | 11\|1209 | 33\|16097 | 1.69E-05 | 0.00041 | ENSMUS | Ldlr/Hmga2/Il6st/Nr1d1/Nog/Notch1/Id4/Epha4/Atf5/Hes1/Me |
| BP | GO:000961 | response to mechanical stimulus | 26\|1209 | 142\|16097 | 1.84E-05 | 0.00044 | ENSMUS | Col1a1/Thbs1/Myc/Sox9/Piezo1/Jun/Atp1a1/Abl2/Fos/Usp53/C |
| BP | GO:003017 | negative regulation of Wnt signaling pathway | 26\|1209 | 142\|16097 | 1.84E-05 | 0.00044 | ENSMUS | Sox9/Egr1/Wif1/Ddit3/Hmga2/Lzts2/Sfrp2/Nog/Lats1/Notch1/S |
| BP | GO:190265 | secondary alcohol metabolic process | 23\|1209 | 118\|16097 | 1.95E-05 | 0.00047 | ENSMUS | Hmgcr/Ldlr/Cyp51/Fgfr1/Fdft1/Nfe2l1/Srebf2/Hmgcs1/Sqle/Insi |
| BP | GO:002179 | cerebral cortex cell migration | 14\|1209 | 52\|16097 | 1.99E-05 | 0.00047 | ENSMUS | Lamb1/Fgfr1/Pex5/Rtn4/Dixdc1/Adgrg1/Egfr/Lrp8/Ndel1/Nrp1/ |
| BP | GO:200123 | regulation of extrinsic apoptotic signaling pathway | 27\|1209 | 151\|16097 | 2.00E-05 | 0.00047 | ENSMUS | Thbs1/Hmox1/Serpine1/Inhba/Fgfr1/Fem1b/Tnfrsf12a/Lgals3/L |
| BP | GO:003109 | stress-activated protein kinase signaling cascade | 41\|1209 | 276\|16097 | 2.01E-05 | 0.00048 | ENSMUS | Errfi1/Tnxb/Hmgcr/Map2k3/Trib1/Gadd45a/Dusp1/Met/Dixdc1 |
| BP | GO:003287 | regulation of stress-activated MAPK cascade | 35\|1209 | 221\|16097 | 2.04E-05 | 0.00048 | ENSMUS | Tnxb/Hmgcr/Gadd45a/Dusp1/Met/Dixdc1/Nrk/Egfr/Tiam1/Ctgf/ |
| BP | GO:003576 | endothelial cell chemotaxis | 10\|1209 | 28\|16097 | 2.06E-05 | 0.00048 | ENSMUS | Thbs1/Fgfr1/Met/Smoc2/Nr4a1/Nrp1/Vegfa/Notch1/Nov/Kdr |
| MF | GO:000551 | collagen binding | 16\|1190 | 66\|15912 | 2.08E-05 | 0.00118 | ENSMUS | Tnxb/Thbs1/Nid1/Pcolce/Aspn/Ctsk/Adgrg1/Abi3bp/Thbs4/Com |
| BP | GO:000165 | ureteric bud development | 20\|1209 | 95\|16097 | 2.08E-05 | 0.00048 | ENSMUS | Myc/Sox9/Fgfr1/Vcan/Smad5/Fgfr2/Lzts2/Agtr2/Maged1/Nog/C |
| BP | GO:007216 | mesonephric epithelium development | 20\|1209 | 95\|16097 | 2.08E-05 | 0.00048 | ENSMUS | Myc/Sox9/Fgfr1/Vcan/Smad5/Fgfr2/Lzts2/Agtr2/Maged1/Nog/C |
| BP | GO:007216 | mesonephric tubule development | 20\|1209 | 95\|16097 | 2.08E-05 | 0.00048 | ENSMUS | Myc/Sox9/Fgfr1/Vcan/Smad5/Fgfr2/Lzts2/Agtr2/Maged1/Nog/C |
| BP | GO:001474 | regulation of muscle hypertrophy | 17\|1209 | 73\|16097 | 2.16E-05 | 0.0005 | ENSMUS | Errfi1/Col14a1/Igfbp5/Rgs2/Lmna/Mef2a/Lmcd1/Tnfrsf1a/Ctdp |
| BP | GO:000170 | in utero embryonic development | 63\|1209 | 493\|16097 | 2.17E-05 | 0.0005 | ENSMUS | Fosl1/Fgfr1/Xist/Cdkn1c/Rtn4/Mafg/Ncoa3/Cyr61/Tpm1/Fgfr2/ |
| BP | GO:005134 | negative regulation of hydrolase activity | 47\|1209 | 334\|16097 | 2.18E-05 | 0.0005 | ENSMUS | Hspa1b/Spry2/Thbs1/Rgs2/Serpine1/Klf4/Herpud1/Serpinf1/A2 |
| BP | GO:004203 | regulation of cytokine biosynthetic process | 21\|1209 | 103\|16097 | 2.18E-05 | 0.0005 | ENSMUS | Errfi1/Prg4/Thbs1/Tlr2/Map2k3/Klf4/Egr1/Akap12/Stat3/Cebpg |
| BP | GO:190367 | regulation of sprouting angiogenesis | 16\|1209 | 66\|16097 | 2.19E-05 | 0.0005 | ENSMUS | Thbs1/Hmox1/Klf4/Itga5/Spred1/Meox2/Nrp1/Pik3r2/Vegfa/Nc |
| BP | GO:190121 | negative regulation of neuron death | 36\|1209 | 231\|16097 | 2.24E-05 | 0.0005 | ENSMUS | Mt1/Hmox1/Serpinf1/Atf4/Jun/Aars/Hyou1/Cd34/Cx3cl1/Stat3 |
| MF | GO:004339 | proteoglycan binding | 11\|1190 | 34\|15912 | 2.24E-05 | 0.0012 | ENSMUS | Col5a3/Nid1/Lpl/Ctsk/Tnc/Comp/Gpnmb/Fst/Ptn/Cfh/Grem1 |
| BP | GO:005067 | positive regulation of epithelial cell proliferation | 31\|1209 | 187\|16097 | 2.41E-05 | 0.00054 | ENSMUS | F3/Hmox1/Myc/Sox9/Fgfr1/Has2/Jun/Rtn4/Fgfr2/Hmga2/Bmp6 |
| BP | GO:007137 | cellular response to peptide hormone stimulus | 36\|1209 | 232\|16097 | 2.46E-05 | 0.00055 | ENSMUS | Lpl/Ywhag/Eif4ebp1/Socs3/Sgk1/Stat3/Trib3/Xbp1/Ndel1/Nr4a |
| BP | GO:003085 | regulation of epithelial cell differentiation | 25\|1209 | 136\|16097 | 2.47E-05 | 0.00055 | ENSMUS | Errfi1/Sox9/Serpine1/Cd109/Cdkn1c/Cdh5/Mafg/Ncoa3/Bmp6/ |
| BP | GO:007030 | regulation of stress-activated protein kinase signaling cascade | 35\|1209 | 223\|16097 | 2.48E-05 | 0.00055 | ENSMUS | Tnxb/Hmgcr/Gadd45a/Dusp1/Met/Dixdc1/Nrk/Egfr/Tiam1/Ctgf/ |
| BP | GO:007140 | cellular response to organic cyclic compound | 53\|1209 | 395\|16097 | 2.58E-05 | 0.00057 | ENSMUS | Igfbp5/Inhba/Egr1/Atf4/Eif4ebp1/Atp1a1/Larp1/Nfe2l1/Ncoa3/ |
| BP | GO:000989 | positive regulation of catabolic process | 54\|1209 | 405\|16097 | 2.59E-05 | 0.00057 | ENSMUS | Hspa1b/Hspa1a/Hmox1/Ldlr/Myc/Tlr2/Sox9/Trib1/Herpud1/Zfa |
| BP | GO:005073 | positive regulation of peptidyl-tyrosine phosphorylation | 30\|1209 | 179\|16097 | 2.61E-05 | 0.00057 | ENSMUS | Itga5/Il6ra/Efna1/Iqgap1/Pecam1/Bmp6/Thbs4/Lrp8/Il6st/Hsph |
| BP | GO:002188 | forebrain cell migration | 16\|1209 | 67\|16097 | 2.68E-05 | 0.00058 | ENSMUS | Lamb1/Fgfr1/Pex5/Rtn4/Dixdc1/Adgrg1/Egfr/Lrp8/Ndel1/Nrp1/ |
| BP | GO:200011 | regulation of cysteine-type endopeptidase activity | 34\|1209 | 215\|16097 | 2.75E-05 | 0.0006 | ENSMUS | Hspa1b/F3/Thbs1/Myc/Klf4/Herpud1/Eif2ak3/Cyr61/Ctgf/Dhcr2 |
| BP | GO:000999 | response to extracellular stimulus | 48\|1209 | 347\|16097 | 2.80E-05 | 0.00061 | ENSMUS | Fosl1/Ldlr/Postn/Plec/Lpl/Slc2a1/Atf4/Jun/Aacs/Eif4ebp1/Larp |
| BP | GO:001701 | regulation of transforming growth factor beta receptor signalin | 20\|1209 | 97\|16097 | 2.86E-05 | 0.00062 | ENSMUS | Thbs1/Vasn/Cd109/Aspn/Tgfbr3/Chst11/Xbp1/Ski/Pmepa1/Ltbp |
| BP | GO:007177 | response to BMP | 27\|1209 | 154\|16097 | 2.88E-05 | 0.00062 | ENSMUS | Egr1/Cdh5/Chrdl1/Cyr61/Smad5/Bmp6/Tgfbr3/Neo1/Ski/Sfrp2/ |
| BP | GO:007177 | cellular response to BMP stimulus | 27\|1209 | 154\|16097 | 2.88E-05 | 0.00062 | ENSMUS | Egr1/Cdh5/Chrdl1/Cyr61/Smad5/Bmp6/Tgfbr3/Neo1/Ski/Sfrp2/ |
| BP | GO:006097 | coronary vasculature morphogenesis | 10\|1209 | 29\|16097 | 2.94E-05 | 0.00062 | ENSMUS | Fgfr1/Spred1/Fgfr2/Rxra/Tgfbr3/Nrp1/Tgfbr1/Vegfa/Notch1/Ace |
| BP | GO:000669 | steroid biosynthetic process | 23\|1209 | 121\|16097 | 2.97E-05 | 0.00063 | ENSMUS | Hmgcr/Cyp51/Egr1/Stard4/Fdft1/Msmo1/Atp1a1/Hmgcs1/Sqle/ |
| BP | GO:001612 | sterol metabolic process | 23\|1209 | 121\|16097 | 2.97E-05 | 0.00063 | ENSMUS | Hmgcr/Ldlr/Cyp51/Fdft1/Msmo1/Nfe2l1/Srebf2/Hmgcs1/Sqle/I |
| BP | GO:000820 | cholesterol metabolic process | 22\|1209 | 113\|16097 | 2.99E-05 | 0.00063 | ENSMUS | Hmgcr/Ldlr/Cyp51/Fdft1/Nfe2l1/Srebf2/Hmgcs1/Sqle/Insig1/Hs |
| BP | GO:009009 | negative regulation of canonical Wnt signaling pathway | 22\|1209 | 113\|16097 | 2.99E-05 | 0.00063 | ENSMUS | Sox9/Egr1/Ddit3/Lzts2/Sfrp2/Nog/Lats1/Notch1/Sfrp4/G3bp1/ |
| MF | GO:000471 | transmembrane receptor protein tyrosine kinase activity | 14\|1190 | 54\|15912 | 3.01E-05 | 0.00152 | ENSMUS | Fgfr1/Met/Fgfr2/Egfr/Nrp1/Igf1r/Flt4/Axl/Epha2/Epha4/Tek/Tie1 |
| BP | GO:000193 | endothelial cell proliferation | 25\|1209 | 138\|16097 | 3.19E-05 | 0.00067 | ENSMUS | F3/Thbs1/Hmox1/Dlk1/Tnmd/Fgfr1/Jun/Cd34/Bmp6/Stat3/Thb |
| BP | GO:001491 | positive regulation of smooth muscle cell migration | 15\|1209 | 61\|16097 | 3.29E-05 | 0.00068 | ENSMUS | F3/Myc/Igfbp5/Postn/Egr1/Has2/Iqgap1/Il6st/Igf1/Adamts1/Hd |
| BP | GO:005105 | negative regulation of small GTPase mediated signal transducti | 15\|1209 | 61\|16097 | 3.29E-05 | 0.00068 | ENSMUS | Spry2/Met/Abl2/Bcl6/Dlc1/Cgnl1/Rabgef1/Rasa2/Nup62/Kank1 |
| BP | GO:006104 | regulation of wound healing | 24\|1209 | 130\|16097 | 3.31E-05 | 0.00068 | ENSMUS | Thbs1/Hmgcr/Serpine1/Tnfrsf12a/Cd109/Serping1/Smoc2/Cdkn |
| BP | GO:003433 | adherens junction organization | 23\|1209 | 122\|16097 | 3.40E-05 | 0.00069 | ENSMUS | Hipk1/S100a10/Cdh5/Iqgap1/Mmp14/Cdh11/Bmp6/Thy1/Nrp |
| BP | GO:007208 | stem cell proliferation | 23\|1209 | 122\|16097 | 3.40E-05 | 0.00069 | ENSMUS | Prg4/Fgfr1/Ncoa3/Fgfr2/Hmga2/Cd34/Cx3cl1/Sfrp2/Vegfa/Nfat |
| BP | GO:001003 | response to inorganic substance | 55\|1209 | 419\|16097 | 3.41E-05 | 0.00069 | ENSMUS | Mt1/Thbs1/Hmox1/Met/Ppp1r15b/Ddi2/Jun/Iqgap1/Nfe2l1/Fo |
| BP | GO:000188 | liver development | 22\|1209 | 114\|16097 | 3.44E-05 | 0.0007 | ENSMUS | Sulf2/Hmox1/Sox9/Met/Jun/Aacs/Rtn4/Tgfbr3/Xbp1/Cebpb/Rel |
| BP | GO:190188 | regulation of cell junction assembly | 18\|1209 | 83\|16097 | 3.50E-05 | 0.00071 | ENSMUS | S100a10/Iqgap1/Mmp14/Thy1/Nrp1/Dlc1/Enpp2/Vegfa/Epha2/ |
| BP | GO:004315 | negative regulation of cysteine-type endopeptidase activity inv | 17\|1209 | 76\|16097 | 3.76E-05 | 0.00076 | ENSMUS | Hspa1b/Thbs1/Klf4/Herpud1/Dhcr24/Nr4a1/Gas6/Sfrp2/Igf1/Co |
| BP | GO:004566 | negative regulation of neuron differentiation | 37\|1209 | 246\|16097 | 3.83E-05 | 0.00077 | ENSMUS | Sox9/Xylt1/Ifrd1/Efna1/Rtn4/Lgals1/Dixdc1/Ngef/Thy1/Nrp1/N |

| Count | Up | Up_Gene_i | Down | Down_Gene_id |
|---|---|---|---|---|
| 30 | 8 | ENSMUSG0 | 22 | ENSMUSG00000022114/ENSMUSG00000028128/ENSMUSG00000040152/ENSMUSG00000000567/ENSMUSG00000003032/ENSMUSG00000020458/ENSMUSG0 |
| 11 | 3 | ENSMUSG0 | 8 | ENSMUSG00000032193/ENSMUSG00000056758/ENSMUSG00000021756/ENSMUSG00000020889/ENSMUSG00000048616/ENSMUSG00000021379/ENSMUSG0 |
| 26 | 14 | ENSMUSG0 | 12 | ENSMUSG00000040152/ENSMUSG00000022346/ENSMUSG00000000567/ENSMUSG00000014444/ENSMUSG00000033161/ENSMUSG00000026596/ENSMUSG0 |
| 26 | 8 | ENSMUSG0 | 18 | ENSMUSG00000000567/ENSMUSG00000038418/ENSMUSG00000020218/ENSMUSG00000025408/ENSMUSG00000056758/ENSMUSG00000035342/ENSMUSG0 |
| 23 | 4 | ENSMUSG0 | 19 | ENSMUSG00000021670/ENSMUSG00000032193/ENSMUSG00000001467/ENSMUSG00000031565/ENSMUSG00000021273/ENSMUSG00000038615/ENSMUSG0 |
| 14 | 5 | ENSMUSG0 | 9 | ENSMUSG00000031565/ENSMUSG00000005069/ENSMUSG00000020458/ENSMUSG00000032064/ENSMUSG00000031785/ENSMUSG00000028613/ENSMUSG0 |
| 27 | 5 | ENSMUSG0 | 22 | ENSMUSG00000040152/ENSMUSG00000005413/ENSMUSG00000037411/ENSMUSG00000041324/ENSMUSG00000031565/ENSMUSG00000032244/ENSMUSG0 |
| 41 | 14 | ENSMUSG0 | 27 | ENSMUSG00000028967/ENSMUSG00000021670/ENSMUSG00000018932/ENSMUSG00000032501/ENSMUSG00000036390/ENSMUSG00000009376/ENSMUSG0 |
| 35 | 13 | ENSMUSG0 | 22 | ENSMUSG00000021670/ENSMUSG00000036390/ENSMUSG00000009376/ENSMUSG00000032064/ENSMUSG00000002489/ENSMUSG00000019997/ENSMUSG0 |
| 10 | 5 | ENSMUSG0 | 5 | ENSMUSG00000040152/ENSMUSG00000031565/ENSMUSG00000009376/ENSMUSG00000023034/ENSMUSG00000023951 |
| 16 | 13 | ENSMUSG0 | 3 | ENSMUSG00000040152/ENSMUSG00000031785/ENSMUSG00000031849 |
| 20 | 8 | ENSMUSG0 | 12 | ENSMUSG00000022346/ENSMUSG00000000567/ENSMUSG00000031565/ENSMUSG00000021540/ENSMUSG00000030849/ENSMUSG00000035342/ENSMUSG0 |
| 20 | 8 | ENSMUSG0 | 12 | ENSMUSG00000022346/ENSMUSG00000000567/ENSMUSG00000031565/ENSMUSG00000021540/ENSMUSG00000030849/ENSMUSG00000035342/ENSMUSG0 |
| 20 | 8 | ENSMUSG0 | 12 | ENSMUSG00000022346/ENSMUSG00000000567/ENSMUSG00000031565/ENSMUSG00000021540/ENSMUSG00000030849/ENSMUSG00000035342/ENSMUSG0 |
| 17 | 6 | ENSMUSG0 | 11 | ENSMUSG00000028967/ENSMUSG00000026360/ENSMUSG00000028063/ENSMUSG00000030557/ENSMUSG00000030341/ENSMUSG00000033323/ENSMUSG0 |
| 63 | 22 | ENSMUSG0 | 41 | ENSMUSG00000024912/ENSMUSG00000031565/ENSMUSG00000020458/ENSMUSG00000051510/ENSMUSG00000027678/ENSMUSG00000030849/ENSMUSG0 |
| 47 | 14 | ENSMUSG0 | 33 | ENSMUSG00000022114/ENSMUSG00000040152/ENSMUSG00000026360/ENSMUSG00000037411/ENSMUSG00000003032/ENSMUSG00000031770/ENSMUSG0 |
| 21 | 4 | ENSMUSG0 | 17 | ENSMUSG00000028967/ENSMUSG00000006014/ENSMUSG00000040152/ENSMUSG00000027995/ENSMUSG00000018932/ENSMUSG00000003032/ENSMUSG0 |
| 16 | 8 | ENSMUSG0 | 8 | ENSMUSG00000040152/ENSMUSG00000005413/ENSMUSG00000003032/ENSMUSG00000000555/ENSMUSG00000027351/ENSMUSG00000023951/ENSMUSG0 |
| 36 | 11 | ENSMUSG0 | 25 | ENSMUSG00000031765/ENSMUSG00000005413/ENSMUSG00000000753/ENSMUSG00000042406/ENSMUSG00000031960/ENSMUSG00000032115/ENSMUSG0 |
| 11 | 8 | ENSMUSG0 | 3 | ENSMUSG00000031849/ENSMUSG00000029816/ENSMUSG00000074934 |
| 31 | 11 | ENSMUSG0 | 20 | ENSMUSG00000028128/ENSMUSG00000005413/ENSMUSG00000022346/ENSMUSG00000000567/ENSMUSG00000031565/ENSMUSG00000022367/ENSMUSG0 |
| 36 | 8 | ENSMUSG0 | 28 | ENSMUSG00000051391/ENSMUSG00000031490/ENSMUSG00000019970/ENSMUSG00000004040/ENSMUSG00000032715/ENSMUSG00000020484/ENSMUSG0 |
| 25 | 11 | ENSMUSG0 | 14 | ENSMUSG00000028967/ENSMUSG00000000567/ENSMUSG00000037411/ENSMUSG00000046186/ENSMUSG00000051510/ENSMUSG00000027678/ENSMUSG0 |
| 35 | 13 | ENSMUSG0 | 22 | ENSMUSG00000021670/ENSMUSG00000036390/ENSMUSG00000009376/ENSMUSG00000032064/ENSMUSG00000002489/ENSMUSG00000019997/ENSMUSG0 |
| 53 | 13 | ENSMUSG0 | 40 | ENSMUSG00000041324/ENSMUSG00000038418/ENSMUSG00000042406/ENSMUSG00000031490/ENSMUSG00000033161/ENSMUSG00000037331/ENSMUSG0 |
| 54 | 14 | ENSMUSG0 | 40 | ENSMUSG00000005413/ENSMUSG00000032193/ENSMUSG00000022346/ENSMUSG00000027995/ENSMUSG00000000567/ENSMUSG00000032501/ENSMUSG0 |
| 30 | 10 | ENSMUSG0 | 20 | ENSMUSG00000000555/ENSMUSG00000027947/ENSMUSG00000027954/ENSMUSG00000030536/ENSMUSG00000039004/ENSMUSG00000028613/ENSMUSG0 |
| 16 | 7 | ENSMUSG0 | 9 | ENSMUSG00000031565/ENSMUSG00000005069/ENSMUSG00000020458/ENSMUSG00000032064/ENSMUSG00000031785/ENSMUSG00000028613/ENSMUSG0 |
| 34 | 8 | ENSMUSG0 | 26 | ENSMUSG00000028128/ENSMUSG00000040152/ENSMUSG00000022346/ENSMUSG00000003032/ENSMUSG00000031770/ENSMUSG00000031668/ENSMUSG0 |
| 48 | 13 | ENSMUSG0 | 35 | ENSMUSG00000024912/ENSMUSG00000032193/ENSMUSG00000022565/ENSMUSG00000028645/ENSMUSG00000042406/ENSMUSG00000029482/ENSMUSG0 |
| 20 | 5 | ENSMUSG0 | 15 | ENSMUSG00000040152/ENSMUSG00000039646/ENSMUSG00000029287/ENSMUSG00000020484/ENSMUSG00000029050/ENSMUSG0 |
| 27 | 10 | ENSMUSG0 | 17 | ENSMUSG00000038418/ENSMUSG00000021540/ENSMUSG00000039004/ENSMUSG00000029287/ENSMUSG00000032340/ENSMUSG00000029050/ENSMUSG0 |
| 27 | 10 | ENSMUSG0 | 17 | ENSMUSG00000038418/ENSMUSG00000021540/ENSMUSG00000039004/ENSMUSG00000029287/ENSMUSG00000032340/ENSMUSG00000029050/ENSMUSG0 |
| 10 | 3 | ENSMUSG0 | 7 | ENSMUSG00000031565/ENSMUSG00000027351/ENSMUSG00000030849/ENSMUSG00000015846/ENSMUSG00000029287/ENSMUSG00000007613/ENSMUSG0 |
| 23 | 3 | ENSMUSG0 | 20 | ENSMUSG00000021670/ENSMUSG00000001467/ENSMUSG00000038418/ENSMUSG00000024378/ENSMUSG00000021273/ENSMUSG00000031604/ENSMUSG0 |
| 23 | 4 | ENSMUSG0 | 19 | ENSMUSG00000021670/ENSMUSG00000032193/ENSMUSG00000001467/ENSMUSG00000021273/ENSMUSG00000031604/ENSMUSG00000038615/ENSMUSG0 |
| 22 | 4 | ENSMUSG0 | 18 | ENSMUSG00000021670/ENSMUSG00000032193/ENSMUSG00000001467/ENSMUSG00000021273/ENSMUSG00000038615/ENSMUSG00000022463/ENSMUSG0 |
| 22 | 7 | ENSMUSG0 | 15 | ENSMUSG00000000567/ENSMUSG00000038418/ENSMUSG00000025408/ENSMUSG00000035342/ENSMUSG00000027996/ENSMUSG00000048616/ENSMUSG0 |
| 14 | 8 | ENSMUSG0 | 6 | ENSMUSG00000031565/ENSMUSG00000009376/ENSMUSG00000030849/ENSMUSG00000005533/ENSMUSG00000006445/ENSMUSG00000028661 |
| 25 | 10 | ENSMUSG0 | 15 | ENSMUSG00000028128/ENSMUSG00000040152/ENSMUSG00000005413/ENSMUSG00000040856/ENSMUSG00000031565/ENSMUSG00000039004/ENSMUSG0 |
| 15 | 6 | ENSMUSG0 | 9 | ENSMUSG00000028128/ENSMUSG00000022346/ENSMUSG00000038418/ENSMUSG00000022367/ENSMUSG00000030536/ENSMUSG00000021756/ENSMUSG0 |
| 15 | 2 | ENSMUSG0 | 13 | ENSMUSG00000022114/ENSMUSG00000009376/ENSMUSG00000026596/ENSMUSG00000022508/ENSMUSG00000031523/ENSMUSG00000032232/ENSMUSG0 |
| 24 | 7 | ENSMUSG0 | 17 | ENSMUSG00000040152/ENSMUSG00000021670/ENSMUSG00000037411/ENSMUSG00000023905/ENSMUSG00000046186/ENSMUSG00000023067/ENSMUSG0 |
| 23 | 10 | ENSMUSG0 | 13 | ENSMUSG00000008730/ENSMUSG00000041959/ENSMUSG00000030536/ENSMUSG00000039004/ENSMUSG00000031523/ENSMUSG00000022425/ENSMUSG0 |
| 23 | 7 | ENSMUSG0 | 16 | ENSMUSG00000006014/ENSMUSG00000031565/ENSMUSG00000027678/ENSMUSG00000030849/ENSMUSG00000056758/ENSMUSG00000031778/ENSMUSG0 |
| 55 | 19 | ENSMUSG0 | 36 | ENSMUSG00000031765/ENSMUSG00000040152/ENSMUSG00000005413/ENSMUSG00000009376/ENSMUSG00000046062/ENSMUSG00000078515/ENSMUSG0 |
| 22 | 8 | ENSMUSG0 | 14 | ENSMUSG00000006800/ENSMUSG00000005413/ENSMUSG00000000567/ENSMUSG00000009376/ENSMUSG00000029482/ENSMUSG00000020458/ENSMUSG0 |
| 18 | 7 | ENSMUSG0 | 11 | ENSMUSG00000041959/ENSMUSG00000030536/ENSMUSG00000031523/ENSMUSG00000022425/ENSMUSG00000023951/ENSMUSG00000006445/ENSMUSG0 |
| 17 | 3 | ENSMUSG0 | 14 | ENSMUSG00000040152/ENSMUSG00000003032/ENSMUSG00000031770/ENSMUSG00000034926/ENSMUSG00000023034/ENSMUSG00000031451/ENSMUSG0 |
| 37 | 14 | ENSMUSG0 | 23 | ENSMUSG00000000567/ENSMUSG00000030657/ENSMUSG00000001627/ENSMUSG00000027954/ENSMUSG00000020458/ENSMUSG00000032064/ENSMUSG0 |

0000025408/ENSMUSG00000028893/ENSMUSG00000025373/ENSMUSG00000031778/ENSMUSG00000031451/ENSMUSG00000007613/ENSMUSG00000005533/ENSMUSG00000023951/ENSMUSG0000004
0000038539/ENSMUSG00000031393
0000039701/ENSMUSG00000024927/ENSMUSG00000074818/ENSMUSG00000039952/ENSMUSG00000029249/ENSMUSG00000029361
0000027996/ENSMUSG00000048616/ENSMUSG00000040021/ENSMUSG00000018583/ENSMUSG00000032717/ENSMUSG00000044167/ENSMUSG00000031548/ENSMUSG00000039844/ENSMUSG0000003
0000022463/ENSMUSG00000093930/ENSMUSG00000022351/ENSMUSG00000045294/ENSMUSG00000026675/ENSMUSG00000034926/ENSMUSG00000024413/ENSMUSG00000032018/ENSMUSG0000003
0000018736/ENSMUSG00000035764/ENSMUSG00000020745
0000023905/ENSMUSG00000050335/ENSMUSG00000028063/ENSMUSG00000031778/ENSMUSG00000027996/ENSMUSG00000007613/ENSMUSG00000024927/ENSMUSG00000023951/ENSMUSG0000003
0000032064/ENSMUSG00000002489/ENSMUSG00000019997/ENSMUSG00000031714/ENSMUSG00000027996/ENSMUSG00000035199/ENSMUSG00000005533/ENSMUSG00000081683/ENSMUSG0000002
0000031714/ENSMUSG00000027996/ENSMUSG00000035199/ENSMUSG00000005533/ENSMUSG00000081683/ENSMUSG00000023951/ENSMUSG00000039384/ENSMUSG00000020745/ENSMUSG0000002

0000048616/ENSMUSG00000005087/ENSMUSG00000023951/ENSMUSG00000031548/ENSMUSG00000074934/ENSMUSG00000045394
0000048616/ENSMUSG00000005087/ENSMUSG00000023951/ENSMUSG00000031548/ENSMUSG00000074934/ENSMUSG00000045394
0000048616/ENSMUSG00000005087/ENSMUSG00000023951/ENSMUSG00000031548/ENSMUSG00000074934/ENSMUSG00000045394
0000022752/ENSMUSG00000028161/ENSMUSG00000050965/ENSMUSG00000044167/ENSMUSG00000024431
0000015846/ENSMUSG00000029287/ENSMUSG00000031714/ENSMUSG00000039234/ENSMUSG00000018736/ENSMUSG00000007817/ENSMUSG00000039262/ENSMUSG00000035569/ENSMUSG0000004
0000000753/ENSMUSG00000030111/ENSMUSG00000046186/ENSMUSG00000027351/ENSMUSG00000027447/ENSMUSG00000021950/ENSMUSG00000001131/ENSMUSG00000034926/ENSMUSG0000002
0000038418/ENSMUSG00000004040/ENSMUSG00000056216/ENSMUSG00000020601/ENSMUSG00000024789/ENSMUSG00000056501/ENSMUSG00000024927/ENSMUSG00000003810/ENSMUSG0000001
0000032487/ENSMUSG00000027230
0000031778/ENSMUSG00000004040/ENSMUSG00000020423/ENSMUSG00000024789/ENSMUSG00000032340/ENSMUSG00000056501/ENSMUSG00000023951/ENSMUSG00000061436/ENSMUSG0000002

0000020458/ENSMUSG00000030849/ENSMUSG00000056758/ENSMUSG00000039004/ENSMUSG00000004040/ENSMUSG00000020484/ENSMUSG00000023034/ENSMUSG00000048616/ENSMUSG0000002
0000018736/ENSMUSG00000023034/ENSMUSG00000024789/ENSMUSG00000005533/ENSMUSG00000024927/ENSMUSG00000028042/ENSMUSG00000050965/ENSMUSG00000032702/ENSMUSG0000001
0000039004/ENSMUSG00000030341/ENSMUSG00000007613/ENSMUSG00000056501/ENSMUSG00000023951/ENSMUSG00000047281/ENSMUSG00000022016/ENSMUSG00000005871
0000031714/ENSMUSG00000027996/ENSMUSG00000035199/ENSMUSG00000005533/ENSMUSG00000081683/ENSMUSG00000023951/ENSMUSG00000039384/ENSMUSG00000020745/ENSMUSG0000002
0000038615/ENSMUSG00000027678/ENSMUSG00000021540/ENSMUSG00000042644/ENSMUSG00000056758/ENSMUSG00000045294/ENSMUSG00000019970/ENSMUSG00000028525/ENSMUSG0000000
0000031770/ENSMUSG00000053581/ENSMUSG00000037331/ENSMUSG00000003541/ENSMUSG00000025408/ENSMUSG00000028893/ENSMUSG00000025373/ENSMUSG00000032715/ENSMUSG0000002
0000021756/ENSMUSG00000024789/ENSMUSG00000031451/ENSMUSG00000027293/ENSMUSG00000022425/ENSMUSG00000030341/ENSMUSG00000005087/ENSMUSG00000023951/ENSMUSG0000002
0000018736/ENSMUSG00000035764/ENSMUSG00000020745
0000019997/ENSMUSG00000034926/ENSMUSG00000020349/ENSMUSG00000023034/ENSMUSG00000024789/ENSMUSG00000031451/ENSMUSG00000027996/ENSMUSG00000031523/ENSMUSG0000003
0000031490/ENSMUSG00000037331/ENSMUSG00000023087/ENSMUSG00000023067/ENSMUSG00000022463/ENSMUSG00000031668/ENSMUSG00000025408/ENSMUSG00000028893/ENSMUSG0000003
0000038400/ENSMUSG00000030530/ENSMUSG00000046058/ENSMUSG00000061436/ENSMUSG00000048661/ENSMUSG00000000058/ENSMUSG00000085795/ENSMUSG00000063382/ENSMUSG0000002
0000027996/ENSMUSG00000031849/ENSMUSG00000048616/ENSMUSG00000024098/ENSMUSG00000042377/ENSMUSG00000061436/ENSMUSG00000048661/ENSMUSG00000026836/ENSMUSG0000003
0000027996/ENSMUSG00000031849/ENSMUSG00000048616/ENSMUSG00000024098/ENSMUSG00000042377/ENSMUSG00000061436/ENSMUSG00000048661/ENSMUSG00000026836/ENSMUSG0000003
0000023951
0000033161/ENSMUSG00000093930/ENSMUSG00000022351/ENSMUSG00000045294/ENSMUSG00000039004/ENSMUSG00000026675/ENSMUSG00000034926/ENSMUSG00000020889/ENSMUSG0000000
0000022463/ENSMUSG00000093930/ENSMUSG00000022351/ENSMUSG00000045294/ENSMUSG00000026675/ENSMUSG00000034926/ENSMUSG00000024413/ENSMUSG00000032018/ENSMUSG0000003
0000093930/ENSMUSG00000022351/ENSMUSG00000045294/ENSMUSG00000026675/ENSMUSG00000034926/ENSMUSG00000024413/ENSMUSG00000032018/ENSMUSG00000030747/ENSMUSG0000003
0000040021/ENSMUSG00000018583/ENSMUSG00000044167/ENSMUSG00000031548/ENSMUSG00000039844/ENSMUSG00000036432/ENSMUSG00000074934/ENSMUSG00000024182/ENSMUSG0000000

0000004040/ENSMUSG00000020484/ENSMUSG00000023034/ENSMUSG00000007613/ENSMUSG00000023951/ENSMUSG00000006445/ENSMUSG00000050965/ENSMUSG00000000058/ENSMUSG0000003
0000026313/ENSMUSG00000042903/ENSMUSG00000034593
0000025340/ENSMUSG00000032413/ENSMUSG00000109511/ENSMUSG00000032702/ENSMUSG00000039844/ENSMUSG00000009630/ENSMUSG00000031453
0000020484/ENSMUSG00000039384/ENSMUSG00000042594/ENSMUSG00000022443/ENSMUSG00000050965/ENSMUSG00000032702/ENSMUSG00000024486/ENSMUSG00000033717/ENSMUSG0000001
0000031788/ENSMUSG00000023951/ENSMUSG00000040945/ENSMUSG00000044147/ENSMUSG00000085795/ENSMUSG00000074934/ENSMUSG00000026812
0000027996/ENSMUSG00000023951/ENSMUSG00000020745/ENSMUSG00000037815/ENSMUSG00000036606/ENSMUSG00000021379/ENSMUSG00000069895/ENSMUSG00000058070/ENSMUSG0000004
0000030536/ENSMUSG00000038615/ENSMUSG00000042644/ENSMUSG00000039004/ENSMUSG00000030557/ENSMUSG00000020349/ENSMUSG00000037242/ENSMUSG00000023034/ENSMUSG0000002
0000029287/ENSMUSG00000020484/ENSMUSG00000056501/ENSMUSG00000024927/ENSMUSG00000039662/ENSMUSG00000024085/ENSMUSG00000027665/ENSMUSG00000044147
0000040945/ENSMUSG00000062232/ENSMUSG00000039844/ENSMUSG00000074934/ENSMUSG00000026812
0000027996/ENSMUSG00000005087/ENSMUSG00000046223/ENSMUSG00000023951/ENSMUSG00000047281/ENSMUSG00000032487/ENSMUSG00000035992/ENSMUSG00000036432
0000026259/ENSMUSG00000032340/ENSMUSG00000026728/ENSMUSG00000049792/ENSMUSG00000020745/ENSMUSG00000028161/ENSMUSG00000032702/ENSMUSG00000021379/ENSMUSG0000003

42594/ENSMUSG00000006445/ENSMUSG00000024486/ENSMUSG00000040268/ENSMUSG00000039952/ENSMUSG00000027665/ENSMUSG00000039844/ENSMUSG00000044857

36432/ENSMUSG00000074934/ENSMUSG00000024182/ENSMUSG00000005871
30747/ENSMUSG00000033105/ENSMUSG00000024924/ENSMUSG00000031835/ENSMUSG00000050697

37815/ENSMUSG00000024073/ENSMUSG00000020044/ENSMUSG00000026836/ENSMUSG00000032350/ENSMUSG00000031548/ENSMUSG00000036432/ENSMUSG00000026628
23951/ENSMUSG00000039384/ENSMUSG00000020745/ENSMUSG00000027177/ENSMUSG00000061436/ENSMUSG00000032717/ENSMUSG00000024383/ENSMUSG00000044167/ENSMUSG00000031548/E
27177/ENSMUSG00000061436/ENSMUSG00000032717/ENSMUSG00000044167/ENSMUSG00000031548/ENSMUSG00000039844/ENSMUSG00000037902/ENSMUSG00000024182

48616/ENSMUSG00000007613/ENSMUSG00000037434/ENSMUSG00000056501/ENSMUSG00000004846/ENSMUSG00000035247/ENSMUSG00000045216/ENSMUSG00000039662/ENSMUSG00000023951/E
23034/ENSMUSG00000067780/ENSMUSG00000031451/ENSMUSG00000027996/ENSMUSG00000081683/ENSMUSG00000005087/ENSMUSG00000046223/ENSMUSG00000023951/ENSMUSG00000027834/E
16024/ENSMUSG00000028277/ENSMUSG00000031438

22270/ENSMUSG00000018634/ENSMUSG00000029816/ENSMUSG00000027665/ENSMUSG00000029249/ENSMUSG00000032350/ENSMUSG00000029361/ENSMUSG00000026797/ENSMUSG00000031393/E

23951/ENSMUSG00000050965/ENSMUSG00000000058/ENSMUSG00000000184/ENSMUSG00000031548/ENSMUSG00000085795
18634/ENSMUSG00000000058/ENSMUSG00000038859/ENSMUSG00000024052/ENSMUSG00000044167/ENSMUSG00000020091/ENSMUSG00000042903/ENSMUSG00000019943/ENSMUSG00000026826/E

27177/ENSMUSG00000061436/ENSMUSG00000032717/ENSMUSG00000044167/ENSMUSG00000031548/ENSMUSG00000039844/ENSMUSG00000037902/ENSMUSG00000024182
04040/ENSMUSG00000033863/ENSMUSG00000024789/ENSMUSG00000024413/ENSMUSG00000034640/ENSMUSG00000048490/ENSMUSG00000030059/ENSMUSG00000040021/ENSMUSG00000037815/E
20484/ENSMUSG00000049323/ENSMUSG00000020423/ENSMUSG00000020601/ENSMUSG00000034640/ENSMUSG00000022708/ENSMUSG00000048249/ENSMUSG00000021036/ENSMUSG00000022037/E
24486/ENSMUSG00000079037/ENSMUSG00000000058/ENSMUSG00000033717/ENSMUSG00000026836/ENSMUSG00000074934

35199/ENSMUSG00000005087/ENSMUSG00000046223/ENSMUSG00000023951/ENSMUSG00000047281/ENSMUSG00000032487/ENSMUSG00000035992/ENSMUSG00000029249/ENSMUSG00000035900/E
39004/ENSMUSG00000040463/ENSMUSG00000037465/ENSMUSG00000034354/ENSMUSG00000020484/ENSMUSG00000029752/ENSMUSG00000031451/ENSMUSG00000027996/ENSMUSG00000045983/E
24182
31548/ENSMUSG00000044857/ENSMUSG00000074934
31548/ENSMUSG00000044857/ENSMUSG00000074934

05533/ENSMUSG00000032018/ENSMUSG00000033105/ENSMUSG00000029249/ENSMUSG00000024431/ENSMUSG00000050697
30747/ENSMUSG00000033105/ENSMUSG00000024924/ENSMUSG00000031835/ENSMUSG00000050697
33105/ENSMUSG00000024924/ENSMUSG00000031835/ENSMUSG00000050697
05871

34205

15839/ENSMUSG00000056071/ENSMUSG00000027712

40025/ENSMUSG00000045394
24413/ENSMUSG00000037434/ENSMUSG00000024927/ENSMUSG00000018476/ENSMUSG00000028161/ENSMUSG00000024423/ENSMUSG00000063142/ENSMUSG00000079037/ENSMUSG00000025582/E

32593/ENSMUSG00000003814/ENSMUSG00000062232/ENSMUSG00000018965/ENSMUSG00000029249/ENSMUSG00000044147/ENSMUSG00000031393/ENSMUSG00000025326/ENSMUSG00000026812

ENSMUSG00000039844/ENSMUSG00000037902/ENSMUSG00000024182/ENSMUSG00000074923

ENSMUSG00000040021/ENSMUSG00000074221/ENSMUSG00000024085/ENSMUSG00000038279/ENSMUSG00000024073/ENSMUSG00000002052/ENSMUSG00000022443/ENSMUSG00000020346/ENSMUS
ENSMUSG00000024073/ENSMUSG00000027832/ENSMUSG00000047281/ENSMUSG00000020044/ENSMUSG00000040945/ENSMUSG00000032487/ENSMUSG00000035992/ENSMUSG00000029361/ENSMUS

ENSMUSG00000026826/ENSMUSG00000026812

ENSMUSG00000050697/ENSMUSG00000026812/ENSMUSG00000027712/ENSMUSG00000005871/ENSMUSG00000034593

ENSMUSG00000039202/ENSMUSG00000048249/ENSMUSG00000024423/ENSMUSG00000003814/ENSMUSG00000022037/ENSMUSG00000062232/ENSMUSG00000018965/ENSMUSG00000029249/ENSMUS
ENSMUSG00000025234/ENSMUSG00000033717/ENSMUSG00000028793/ENSMUSG00000044167/ENSMUSG00000032350/ENSMUSG00000028211/ENSMUSG00000042303/ENSMUSG00000040025/ENSMUS

ENSMUSG00000036432/ENSMUSG00000056071/ENSMUSG00000002083
ENSMUSG00000005533/ENSMUSG00000024423/ENSMUSG00000034201/ENSMUSG00000035992/ENSMUSG00000044167/ENSMUSG00000015839/ENSMUSG00000063229/ENSMUSG00000028646/ENSMUS

ENSMUSG00000040268/ENSMUSG00000044167/ENSMUSG00000034205/ENSMUSG00000015839/ENSMUSG00000013275/ENSMUSG00000031393/ENSMUSG00000021794/ENSMUSG00000009630/ENSMUS

G00000032717/ENSMUSG00000024486/ENSMUSG00000074656/ENSMUSG00000026254/ENSMUSG00000026836/ENSMUSG00000024807/ENSMUSG00000024182/ENSMUSG00000035007/ENSMUSG0000
G00000036432/ENSMUSG00000026812

G00000029361/ENSMUSG00000031548/ENSMUSG00000024431/ENSMUSG00000039844/ENSMUSG00000031393/ENSMUSG00000030287/ENSMUSG00000085795/ENSMUSG00000025326/ENSMUSG0000
G00000038068/ENSMUSG00000009291/ENSMUSG00000024425/ENSMUSG00000050697/ENSMUSG00000026812/ENSMUSG00000024182/ENSMUSG00000039318/ENSMUSG00000002365/ENSMUSG0000

G00000026826/ENSMUSG00000050697/ENSMUSG00000026812/ENSMUSG00000026628

G00000026826/ENSMUSG00000050697/ENSMUSG00000037902/ENSMUSG00000004207/ENSMUSG00000027712

)031386/ENSMUSG00000032220

)005886
)005871

| Category | GOID | Description | GeneRatio | BgRatio | pvalue | padj | geneID | geneName |
|---|---|---|---|---|---|---|---|---|
| BP | GO:190384 | regulation of cellular response to transforming growth factor b | 20\|1209 | 99\|16097 | 3.90E-05 | 0.00078 | ENSMUS | Thbs1/Vasn/Cd109/Aspn/Tgfbr3/Chst11/Xbp1/Ski/Pmepa1/Ltbp |
| BP | GO:004658 | negative regulation of Ras protein signal transduction | 14\|1209 | 55\|16097 | 3.95E-05 | 0.00078 | ENSMUS | Spry2/Met/Abl2/Bcl6/Dlc1/Rabgef1/Rasa2/Nup62/Kank1/Stmn |
| BP | GO:004208 | cytokine biosynthetic process | 22\|1209 | 115\|16097 | 3.96E-05 | 0.00078 | ENSMUS | Errfi1/Prg4/Thbs1/Tlr2/Map2k3/Klf4/Egr1/Akap12/Stat3/Cebpg |
| BP | GO:004350 | muscle adaptation | 22\|1209 | 115\|16097 | 3.96E-05 | 0.00078 | ENSMUS | Errfi1/Col14a1/Igfbp5/Rgs2/Klf4/Lmna/Atp2a2/Mef2a/Lmcd1/T |
| BP | GO:004873 | cardiac muscle tissue development | 37\|1209 | 247\|16097 | 4.18E-05 | 0.00083 | ENSMUS | Col14a1/Rgs2/Fgfr1/Met/Sorbs2/Lmna/Tpm1/Fgfr2/Rxra/Maml |
| BP | GO:006135 | neural precursor cell proliferation | 28\|1209 | 166\|16097 | 4.26E-05 | 0.00084 | ENSMUS | Cdon/Fgfr1/Fgfr2/Dixdc1/Adgrg1/Hmga2/Cx3cl1/Btg2/Nog/Tac |
| BP | GO:004586 | positive regulation of proteolysis | 44\|1209 | 315\|16097 | 4.72E-05 | 0.00092 | ENSMUS | Hspa1b/F3/Hspa1a/Myc/Trib1/Herpud1/Zfand2a/Efna1/Mmp14 |
| BP | GO:001061 | regulation of cardiac muscle hypertrophy | 16\|1209 | 70\|16097 | 4.78E-05 | 0.00093 | ENSMUS | Errfi1/Col14a1/Rgs2/Lmna/Mef2a/Lmcd1/Tnfrsf1a/Ctdp1/Igf1/T |
| BP | GO:000697 | response to oxidative stress | 51\|1209 | 384\|16097 | 4.78E-05 | 0.00093 | ENSMUS | Srxn1/Thbs1/Hmox1/Met/Rcan1/Ppp1r15b/Atf4/Jun/Nfe2l1/Cs |
| BP | GO:009025 | regulation of muscle system process | 35\|1209 | 230\|16097 | 4.80E-05 | 0.00093 | ENSMUS | Errfi1/Col14a1/Igfbp5/Rgs2/Klf4/Cald1/Atp1a1/Lmna/Atp2a2/P |
| BP | GO:003433 | adherens junction assembly | 18\|1209 | 85\|16097 | 4.89E-05 | 0.00094 | ENSMUS | Hipk1/S100a10/Iqgap1/Mmp14/Thy1/Nrp1/Dlc1/Enpp2/Actn2 |
| BP | GO:005500 | striated muscle cell development | 30\|1209 | 185\|16097 | 4.96E-05 | 0.00094 | ENSMUS | Acta1/Col14a1/Rgs2/Met/Rcan1/Sorbs2/Lmna/Maml1/Myh11/ |
| BP | GO:003569 | cellular response to drug | 43\|1209 | 306\|16097 | 4.97E-05 | 0.00094 | ENSMUS | Thbs1/Hmox1/Myc/Sox9/Serpine1/Egr1/Met/Atf4/Ddi2/Sorbs2, |
| BP | GO:006005 | angiogenesis involved in wound healing | 7\|1209 | 15\|16097 | 4.98E-05 | 0.00094 | ENSMUS | Serpine1/Smoc2/Cx3cl1/Xbp1/Adipor2/Prcp/Ndnf |
| BP | GO:006143 | bone trabecula morphogenesis | 7\|1209 | 15\|16097 | 4.98E-05 | 0.00094 | ENSMUS | Col1a1/Chad/Vegfa/Sbno2/Thbs3/Sfrp1/Grem1 |
| BP | GO:003032 | lung development | 31\|1209 | 194\|16097 | 4.99E-05 | 0.00094 | ENSMUS | Errfi1/Spry2/Igfbp5/Sox9/Stk40/Fgfr1/Mmp14/Adamts2/Fgfr2/ |
| BP | GO:004434 | type B pancreatic cell proliferation | 9\|1209 | 25\|16097 | 5.00E-05 | 0.00094 | ENSMUS | Errfi1/Igfbp5/Igfbp3/Igfbp4/Nr4a1/Nr1d1/Nov/Sfrp1/Ptprv |
| BP | GO:005189 | positive regulation of focal adhesion assembly | 9\|1209 | 25\|16097 | 5.00E-05 | 0.00094 | ENSMUS | S100a10/Iqgap1/Thy1/Nrp1/Enpp2/Vegfa/Tek/Kdr/Tsc1 |
| BP | GO:190153 | regulation of hematopoietic progenitor cell differentiation | 9\|1209 | 25\|16097 | 5.00E-05 | 0.00094 | ENSMUS | Hspa9/Vegfa/Notch1/Zbtb1/Fnip1/Nfe2l2/Ythdf2/Hes1/Kdr |
| BP | GO:190165 | response to peptide | 48\|1209 | 355\|16097 | 5.04E-05 | 0.00094 | ENSMUS | Egr1/Lpl/Ywhag/Eif4ebp1/Atp2a2/Socs3/Sesn2/Sgk1/Tgfbr3/Sta |
| CC | GO:000561 | interstitial matrix | 7\|1232 | 15\|16313 | 5.16E-05 | 0.00154 | ENSMUS | Col14a1/Smoc2/Abi3bp/Tnc/Igf1/Egflam/Vit |
| BP | GO:006100 | hepaticobiliary system development | 22\|1209 | 117\|16097 | 5.21E-05 | 0.00097 | ENSMUS | Sulf2/Hmox1/Sox9/Met/Jun/Aacs/Rtn4/Tgfbr3/Xbp1/Cebpb/Rel |
| BP | GO:004343 | response to peptide hormone | 42\|1209 | 297\|16097 | 5.22E-05 | 0.00097 | ENSMUS | Egr1/Lpl/Ywhag/Eif4ebp1/Atp2a2/Socs3/Sesn2/Sgk1/Tgfbr3/Sta |
| BP | GO:000602 | glycosaminoglycan biosynthetic process | 12\|1209 | 43\|16097 | 5.25E-05 | 0.00097 | ENSMUS | Xylt1/Has2/Chpf/Chst11/Csgalnact1/Pxylp1/Slc35d1/Has1/Extl |
| BP | GO:000165 | temperature homeostasis | 28\|1209 | 168\|16097 | 5.31E-05 | 0.00098 | ENSMUS | Egr1/Scd1/Atf4/Ddit3/Stat3/Jak2/Arrdc3/Fabp4/Lnpep/Nr1d1/ |
| BP | GO:000341 | growth plate cartilage chondrocyte differentiation | 8\|1209 | 20\|16097 | 5.50E-05 | 0.001 | ENSMUS | Col14a1/Col12a1/Col6a2/Matn3/Col6a3/Matn1/Col6a1/Vit |
| BP | GO:000343 | chondrocyte development involved in endochondral bone mor | 8\|1209 | 20\|16097 | 5.50E-05 | 0.001 | ENSMUS | Col14a1/Col12a1/Col6a2/Matn3/Col6a3/Matn1/Col6a1/Vit |
| BP | GO:009005 | positive regulation of cell migration involved in sprouting angi | 8\|1209 | 20\|16097 | 5.50E-05 | 0.001 | ENSMUS | Hmox1/Nrp1/Vegfa/Ptgs2/Kdr/Hdac7/Pik3c2a/Rhoj |
| BP | GO:190339 | positive regulation of adherens junction organization | 10\|1209 | 31\|16097 | 5.66E-05 | 0.00103 | ENSMUS | S100a10/Iqgap1/Thy1/Nrp1/Enpp2/Vegfa/Tek/Arf6/Kdr/Tsc1 |
| BP | GO:003134 | positive regulation of cell projection organization | 56\|1209 | 437\|16097 | 5.69E-05 | 0.00103 | ENSMUS | Rgs2/Serpinf1/Fgfr1/Mapk6/Tnfrsf12a/Met/Rtn4/Iqgap1/Golga4 |
| BP | GO:004884 | artery morphogenesis | 16\|1209 | 71\|16097 | 5.74E-05 | 0.00104 | ENSMUS | Col3a1/Ldlr/Rtn4/Mylk/Zmiz1/Nrp1/Notch3/Adgrf5/Comp/Nog |
| MF | GO:003002 | extracellular matrix structural constituent conferring compres | 6\|1190 | 11\|15912 | 5.75E-05 | 0.00274 | ENSMUS | Aspn/Vcan/Prelp/Hspg2/Lum/Spock2 |
| BP | GO:003020 | chondroitin sulfate biosynthetic process | 6\|1209 | 11\|16097 | 5.89E-05 | 0.00105 | ENSMUS | Xylt1/Chpf/Chst11/Csgalnact1/Slc35d1/Ugdh |
| BP | GO:006034 | bone trabecula formation | 6\|1209 | 11\|16097 | 5.89E-05 | 0.00105 | ENSMUS | Col1a1/Chad/Vegfa/Thbs3/Sfrp1/Grem1 |
| BP | GO:199044 | positive regulation of transcription from RNA polymerase II pro | 6\|1209 | 11\|16097 | 5.89E-05 | 0.00105 | ENSMUS | Atf4/Eif2ak3/Ddit3/Cebpb/Creb3l1/Atf3 |
| BP | GO:000761 | learning or memory | 39\|1209 | 270\|16097 | 5.98E-05 | 0.00106 | ENSMUS | Hmgcr/Ldlr/Tlr2/Itga5/Egr1/Serpinf1/Lamb1/Rcan1/Jun/Abl2/It |
| BP | GO:002202 | telencephalon cell migration | 15\|1209 | 64\|16097 | 6.00E-05 | 0.00106 | ENSMUS | Lamb1/Fgfr1/Pex5/Rtn4/Dixdc1/Adgrg1/Egfr/Lrp8/Ndel1/Nrp1/ |
| BP | GO:006068 | regulation of morphogenesis of a branching structure | 15\|1209 | 64\|16097 | 6.00E-05 | 0.00106 | ENSMUS | Sox9/Fgfr1/Met/Rtn4/Fgfr2/Rxra/Agtr2/Maged1/Nog/Vegfa/Sfrp |
| BP | GO:004828 | lung alveolus development | 14\|1209 | 57\|16097 | 6.06E-05 | 0.00106 | ENSMUS | Errfi1/Igfbp5/Stk40/Fgfr2/Igf1/Flt4/Hs6st1/Foxp2/Vegfa/Cic/Ma |
| BP | GO:005189 | regulation of focal adhesion assembly | 14\|1209 | 57\|16097 | 6.06E-05 | 0.00106 | ENSMUS | S100a10/Iqgap1/Mmp14/Thy1/Nrp1/Dlc1/Enpp2/Vegfa/Tek/Rc |
| BP | GO:009010 | regulation of cell-substrate junction assembly | 14\|1209 | 57\|16097 | 6.06E-05 | 0.00106 | ENSMUS | S100a10/Iqgap1/Mmp14/Thy1/Nrp1/Dlc1/Enpp2/Vegfa/Tek/Rc |
| BP | GO:000195 | regulation of cell-matrix adhesion | 21\|1209 | 110\|16097 | 6.08E-05 | 0.00106 | ENSMUS | Thbs1/Postn/Serpine1/Plet1/S100a10/Iqgap1/Mmp14/Thy1/Bc |
| BP | GO:005189 | regulation of protein kinase B signaling | 25\|1209 | 144\|16097 | 6.63E-05 | 0.00115 | ENSMUS | Spry2/F3/Thbs1/Igfbp5/Klf4/Rtn4/Ddit3/Rnf41/Cx3cl1/Egfr/C1 |
| BP | GO:190121 | regulation of neuron death | 47\|1209 | 349\|16097 | 6.64E-05 | 0.00115 | ENSMUS | Mt1/Hmox1/Egr1/Serpinf1/Atf4/Jun/Aars/Ddit3/Fos/Hyou1/Cd |
| BP | GO:003032 | respiratory tube development | 31\|1209 | 197\|16097 | 6.72E-05 | 0.00116 | ENSMUS | Errfi1/Spry2/Igfbp5/Sox9/Stk40/Fgfr1/Mmp14/Adamts2/Fgfr2/ |
| BP | GO:007054 | response to fatty acid | 12\|1209 | 44\|16097 | 6.73E-05 | 0.00116 | ENSMUS | Ldlr/Tlr2/Lpl/Scd1/Aacs/Scd2/Tgfbr3/Cd36/Dgat2/Id3/Gnas/Prk |
| BP | GO:004210 | cytokine metabolic process | 22\|1209 | 119\|16097 | 6.80E-05 | 0.00117 | ENSMUS | Errfi1/Prg4/Thbs1/Tlr2/Map2k3/Klf4/Egr1/Akap12/Stat3/Cebpg |
| MF | GO:000122 | DNA-binding transcription activator activity, RNA polymerase I | 50\|1190 | 381\|15912 | 6.88E-05 | 0.00312 | ENSMUS | Fosl1/Myc/Sox9/Klf4/Egr1/Atf4/Jun/Plagl1/Mafg/Nfe2l1/Ncoa3, |
| BP | GO:005500 | cardiac cell development | 20\|1209 | 103\|16097 | 7.04E-05 | 0.00121 | ENSMUS | Col14a1/Rgs2/Met/Sorbs2/Lmna/Maml1/Myh11/Tgfbr3/Mef2a/ |
| BP | GO:000602 | aminoglycan biosynthetic process | 13\|1209 | 51\|16097 | 7.30E-05 | 0.00124 | ENSMUS | Xylt1/Has2/Chpf/Chst11/B4galt5/Csgalnact1/Pxylp1/Slc35d1/H |
| BP | GO:003016 | proteoglycan biosynthetic process | 13\|1209 | 51\|16097 | 7.30E-05 | 0.00124 | ENSMUS | Xylt1/Csgalnact2/Chpf/Chst11/Cytl1/Csgalnact1/Pxylp1/Slc35d |

| Count | Up | Up_Gene_i | Down | Down_Gene_id |
|---|---|---|---|---|
| 20 | 5 | ENSMUSG0 | 15 | ENSMUSG00000040152/ENSMUSG00000039646/ENSMUSG00000046186/ENSMUSG00000029287/ENSMUSG00000020484/ENSMUSG00000029050/ENSMUSG0 |
| 14 | 2 | ENSMUSG0 | 12 | ENSMUSG00000022114/ENSMUSG00000009376/ENSMUSG00000026596/ENSMUSG00000022508/ENSMUSG00000031523/ENSMUSG00000025340/ENSMUSG0 |
| 22 | 5 | ENSMUSG0 | 17 | ENSMUSG00000028967/ENSMUSG00000006014/ENSMUSG00000040152/ENSMUSG00000027995/ENSMUSG00000018932/ENSMUSG00000003032/ENSMUSG0 |
| 22 | 9 | ENSMUSG0 | 13 | ENSMUSG00000028967/ENSMUSG00000026360/ENSMUSG00000003032/ENSMUSG00000028063/ENSMUSG00000029467/ENSMUSG00000030557/ENSMUSG0 |
| 37 | 17 | ENSMUSG0 | 20 | ENSMUSG00000026360/ENSMUSG00000031565/ENSMUSG00000009376/ENSMUSG00000028063/ENSMUSG00000030849/ENSMUSG00000015846/ENSMUSG0 |
| 28 | 8 | ENSMUSG0 | 20 | ENSMUSG00000038119/ENSMUSG00000031565/ENSMUSG00000030849/ENSMUSG00000032064/ENSMUSG00000031785/ENSMUSG00000056758/ENSMUSG0 |
| 44 | 13 | ENSMUSG0 | 31 | ENSMUSG00000028128/ENSMUSG00000022346/ENSMUSG00000032501/ENSMUSG00000031770/ENSMUSG00000053581/ENSMUSG00000027954/ENSMUSG0 |
| 16 | 5 | ENSMUSG0 | 11 | ENSMUSG00000028967/ENSMUSG00000026360/ENSMUSG00000028063/ENSMUSG00000030557/ENSMUSG00000030341/ENSMUSG00000033323/ENSMUSG0 |
| 51 | 15 | ENSMUSG0 | 36 | ENSMUSG00000032802/ENSMUSG00000040152/ENSMUSG00000005413/ENSMUSG00000009376/ENSMUSG00000022951/ENSMUSG00000046062/ENSMUSG0 |
| 35 | 14 | ENSMUSG0 | 21 | ENSMUSG00000028967/ENSMUSG00000026360/ENSMUSG00000003032/ENSMUSG00000033161/ENSMUSG00000028063/ENSMUSG00000029467/ENSMUSG0 |
| 18 | 8 | ENSMUSG0 | 10 | ENSMUSG00000008730/ENSMUSG00000041959/ENSMUSG00000030536/ENSMUSG00000031523/ENSMUSG00000022425/ENSMUSG00000023951/ENSMUSG0 |
| 30 | 15 | ENSMUSG0 | 15 | ENSMUSG00000026360/ENSMUSG00000009376/ENSMUSG00000022951/ENSMUSG00000028063/ENSMUSG00000050567/ENSMUSG00000030557/ENSMUSG0 |
| 43 | 10 | ENSMUSG0 | 33 | ENSMUSG00000040152/ENSMUSG00000005413/ENSMUSG00000022346/ENSMUSG00000000567/ENSMUSG00000037411/ENSMUSG00000038418/ENSMUSG0 |
| 7 | 3 | ENSMUSG0 | 4 | ENSMUSG00000037411/ENSMUSG00000031778/ENSMUSG00000020484/ENSMUSG00000030168 |
| 7 | 1 | ENSMUSG0 | 6 | ENSMUSG00000039084/ENSMUSG00000023951/ENSMUSG00000035673/ENSMUSG00000028047/ENSMUSG00000031548/ENSMUSG00000074934 |
| 31 | 14 | ENSMUSG0 | 17 | ENSMUSG00000028967/ENSMUSG00000022114/ENSMUSG00000000567/ENSMUSG00000042608/ENSMUSG00000031565/ENSMUSG00000030849/ENSMUSG0 |
| 9 | 5 | ENSMUSG0 | 4 | ENSMUSG00000028967/ENSMUSG00000023034/ENSMUSG00000020889/ENSMUSG00000031548 |
| 9 | 4 | ENSMUSG0 | 5 | ENSMUSG00000041959/ENSMUSG00000030536/ENSMUSG00000022425/ENSMUSG00000023951/ENSMUSG00000026812 |
| 9 | 3 | ENSMUSG0 | 6 | ENSMUSG00000024359/ENSMUSG00000023951/ENSMUSG00000033454/ENSMUSG00000035992/ENSMUSG00000015839/ENSMUSG00000040025 |
| 48 | 13 | ENSMUSG0 | 35 | ENSMUSG00000038418/ENSMUSG00000051391/ENSMUSG00000031490/ENSMUSG00000029467/ENSMUSG00000028893/ENSMUSG00000019970/ENSMUSG0 |
| 7 | 7 | ENSMUSG0 | 0 | |
| 22 | 8 | ENSMUSG0 | 14 | ENSMUSG00000006800/ENSMUSG00000005413/ENSMUSG00000000567/ENSMUSG00000009376/ENSMUSG00000029482/ENSMUSG00000020458/ENSMUSG0 |
| 42 | 9 | ENSMUSG0 | 33 | ENSMUSG00000038418/ENSMUSG00000051391/ENSMUSG00000031490/ENSMUSG00000029467/ENSMUSG00000028893/ENSMUSG00000019970/ENSMUSG0 |
| 12 | 1 | ENSMUSG0 | 11 | ENSMUSG00000030657/ENSMUSG00000022367/ENSMUSG00000032997/ENSMUSG00000036356/ENSMUSG00000043587/ENSMUSG00000028521/ENSMUSG0 |
| 28 | 9 | ENSMUSG0 | 19 | ENSMUSG00000038418/ENSMUSG00000037071/ENSMUSG00000042406/ENSMUSG00000025408/ENSMUSG00000004040/ENSMUSG00000024789/ENSMUSG0 |
| 8 | 5 | ENSMUSG0 | 3 | ENSMUSG00000020241/ENSMUSG00000048126/ENSMUSG00000001119 |
| 8 | 5 | ENSMUSG0 | 3 | ENSMUSG00000020241/ENSMUSG00000048126/ENSMUSG00000001119 |
| 8 | 5 | ENSMUSG0 | 3 | ENSMUSG00000005413/ENSMUSG00000023951/ENSMUSG00000032487 |
| 10 | 4 | ENSMUSG0 | 6 | ENSMUSG00000041959/ENSMUSG00000030536/ENSMUSG00000022425/ENSMUSG00000023951/ENSMUSG00000044147/ENSMUSG00000026812 |
| 56 | 17 | ENSMUSG0 | 39 | ENSMUSG00000026360/ENSMUSG00000000753/ENSMUSG00000031565/ENSMUSG00000042688/ENSMUSG00000023905/ENSMUSG00000009376/ENSMUSG0 |
| 16 | 8 | ENSMUSG0 | 8 | ENSMUSG00000032193/ENSMUSG00000020458/ENSMUSG00000007817/ENSMUSG00000031849/ENSMUSG00000048616/ENSMUSG00000007613/ENSMUSG0 |
| 6 | 6 | ENSMUSG0 | 0 | |
| 6 | 1 | ENSMUSG0 | 5 | ENSMUSG00000030657/ENSMUSG00000032997/ENSMUSG00000036356/ENSMUSG00000028521/ENSMUSG00000029201 |
| 6 | 1 | ENSMUSG0 | 5 | ENSMUSG00000039084/ENSMUSG00000023951/ENSMUSG00000028047/ENSMUSG00000031548/ENSMUSG00000074934 |
| 6 | 0 | | 6 | ENSMUSG00000042406/ENSMUSG00000031668/ENSMUSG00000025408/ENSMUSG00000056501/ENSMUSG00000027230/ENSMUSG00000026628 |
| 39 | 14 | ENSMUSG0 | 25 | ENSMUSG00000021670/ENSMUSG00000032193/ENSMUSG00000027995/ENSMUSG00000000555/ENSMUSG00000038418/ENSMUSG00000000753/ENSMUSG0 |
| 15 | 6 | ENSMUSG0 | 9 | ENSMUSG00000031565/ENSMUSG00000005069/ENSMUSG00000020458/ENSMUSG00000032064/ENSMUSG00000031785/ENSMUSG00000028613/ENSMUSG0 |
| 15 | 3 | ENSMUSG0 | 12 | ENSMUSG00000000567/ENSMUSG00000031565/ENSMUSG00000009376/ENSMUSG00000020458/ENSMUSG00000030849/ENSMUSG00000015846/ENSMUSG0 |
| 14 | 6 | ENSMUSG0 | 8 | ENSMUSG00000028967/ENSMUSG00000042608/ENSMUSG00000030849/ENSMUSG00000045216/ENSMUSG00000023951/ENSMUSG00000005442/ENSMUSG0 |
| 14 | 6 | ENSMUSG0 | 8 | ENSMUSG00000041959/ENSMUSG00000030536/ENSMUSG00000031523/ENSMUSG00000022425/ENSMUSG00000023951/ENSMUSG00000040945/ENSMUSG0 |
| 14 | 6 | ENSMUSG0 | 8 | ENSMUSG00000041959/ENSMUSG00000030536/ENSMUSG00000031523/ENSMUSG00000022425/ENSMUSG00000023951/ENSMUSG00000040945/ENSMUSG0 |
| 21 | 8 | ENSMUSG0 | 13 | ENSMUSG00000040152/ENSMUSG00000037411/ENSMUSG00000032068/ENSMUSG00000041959/ENSMUSG00000030536/ENSMUSG00000022508/ENSMUSG0 |
| 25 | 6 | ENSMUSG0 | 19 | ENSMUSG00000022114/ENSMUSG00000028128/ENSMUSG00000040152/ENSMUSG00000003032/ENSMUSG00000020458/ENSMUSG00000025408/ENSMUSG0 |
| 47 | 12 | ENSMUSG0 | 35 | ENSMUSG00000031765/ENSMUSG00000005413/ENSMUSG00000038418/ENSMUSG00000000753/ENSMUSG00000042406/ENSMUSG00000031960/ENSMUSG0 |
| 31 | 14 | ENSMUSG0 | 17 | ENSMUSG00000028967/ENSMUSG00000022114/ENSMUSG00000000567/ENSMUSG00000042608/ENSMUSG00000031565/ENSMUSG00000030849/ENSMUSG0 |
| 12 | 4 | ENSMUSG0 | 8 | ENSMUSG00000032193/ENSMUSG00000027995/ENSMUSG00000037071/ENSMUSG00000029482/ENSMUSG00000025203/ENSMUSG00000029287/ENSMUSG0 |
| 22 | 5 | ENSMUSG0 | 17 | ENSMUSG00000028967/ENSMUSG00000006014/ENSMUSG00000040152/ENSMUSG00000027995/ENSMUSG00000018932/ENSMUSG00000003032/ENSMUSG0 |
| 50 | 12 | ENSMUSG0 | 38 | ENSMUSG00000024912/ENSMUSG00000022346/ENSMUSG00000000567/ENSMUSG00000003032/ENSMUSG00000038418/ENSMUSG00000042406/ENSMUSG0 |
| 20 | 10 | ENSMUSG0 | 10 | ENSMUSG00000026360/ENSMUSG00000009376/ENSMUSG00000028063/ENSMUSG00000050567/ENSMUSG00000029287/ENSMUSG00000030557/ENSMUSG0 |
| 13 | 1 | ENSMUSG0 | 12 | ENSMUSG00000030657/ENSMUSG00000022367/ENSMUSG00000032997/ENSMUSG00000017929/ENSMUSG00000036356/ENSMUSG00000043587/ENSMUSG0 |
| 13 | 3 | ENSMUSG0 | 10 | ENSMUSG00000030657/ENSMUSG00000042042/ENSMUSG00000032997/ENSMUSG00000036356/ENSMUSG00000043587/ENSMUSG00000028521/ENSMUSG0 |

0000038400/ENSMUSG00000030530/ENSMUSG00000046058/ENSMUSG00000061436/ENSMUSG00000048661/ENSMUSG00000000058/ENSMUSG00000085795/ENSMUSG00000063382/ENSMUSG0000002
0000032413/ENSMUSG00000109511/ENSMUSG00000032702/ENSMUSG00000039844/ENSMUSG00000009630/ENSMUSG00000031453
0000038418/ENSMUSG00000004040/ENSMUSG00000056216/ENSMUSG00000020601/ENSMUSG00000024789/ENSMUSG00000056501/ENSMUSG00000024927/ENSMUSG00000003810/ENSMUSG0000001
0000030341/ENSMUSG00000033323/ENSMUSG00000022752/ENSMUSG00000028161/ENSMUSG00000050965/ENSMUSG00000044167/ENSMUSG00000024431
0000050567/ENSMUSG00000029287/ENSMUSG00000030557/ENSMUSG00000048616/ENSMUSG00000007613/ENSMUSG00000033323/ENSMUSG00000023951/ENSMUSG00000022752/ENSMUSG0000001
0000031778/ENSMUSG00000020423/ENSMUSG00000048616/ENSMUSG00000030852/ENSMUSG00000023951/ENSMUSG00000020745/ENSMUSG00000037815/ENSMUSG00000036606/ENSMUSG0000002
0000031668/ENSMUSG00000004040/ENSMUSG00000020271/ENSMUSG00000019997/ENSMUSG00000020349/ENSMUSG00000032715/ENSMUSG00000020484/ENSMUSG00000020601/ENSMUSG0000002
0000022752/ENSMUSG00000028161/ENSMUSG00000050965/ENSMUSG00000044167/ENSMUSG00000024431
0000042406/ENSMUSG00000038615/ENSMUSG00000027447/ENSMUSG00000029467/ENSMUSG00000028893/ENSMUSG00000027312/ENSMUSG00000031714/ENSMUSG00000020484/ENSMUSG0000002
0000028525/ENSMUSG00000030557/ENSMUSG00000019997/ENSMUSG00000030341/ENSMUSG00000033323/ENSMUSG00000022752/ENSMUSG00000028161/ENSMUSG00000050965/ENSMUSG0000006
0000040945/ENSMUSG00000085795/ENSMUSG00000074934/ENSMUSG00000026812
0000029050/ENSMUSG00000031849/ENSMUSG00000033323/ENSMUSG00000023951/ENSMUSG00000022752/ENSMUSG00000028161/ENSMUSG00000026313/ENSMUSG00000000058/ENSMUSG0000002
0000009376/ENSMUSG00000042406/ENSMUSG00000078515/ENSMUSG00000031490/ENSMUSG00000037331/ENSMUSG00000028525/ENSMUSG00000033863/ENSMUSG00000024789/ENSMUSG0000003

0000056758/ENSMUSG00000039286/ENSMUSG00000019997/ENSMUSG00000048616/ENSMUSG00000004846/ENSMUSG00000045216/ENSMUSG00000023951/ENSMUSG00000005442/ENSMUSG0000002

0000029287/ENSMUSG00000004040/ENSMUSG00000032715/ENSMUSG00000020484/ENSMUSG00000018736/ENSMUSG00000023034/ENSMUSG00000024789/ENSMUSG00000026728/ENSMUSG0000000

0000029287/ENSMUSG00000020484/ENSMUSG00000056501/ENSMUSG00000024927/ENSMUSG00000039662/ENSMUSG00000024085/ENSMUSG00000027665/ENSMUSG00000044147
0000029287/ENSMUSG00000004040/ENSMUSG00000032715/ENSMUSG00000020484/ENSMUSG00000018736/ENSMUSG00000023034/ENSMUSG00000024789/ENSMUSG00000005533/ENSMUSG0000002
0000003665/ENSMUSG00000021978/ENSMUSG00000029201/ENSMUSG00000047126/ENSMUSG00000039497
0000074794/ENSMUSG00000023845/ENSMUSG00000020889/ENSMUSG00000005533/ENSMUSG00000056501/ENSMUSG00000023951/ENSMUSG00000000134/ENSMUSG00000018476/ENSMUSG0000002

0000020458/ENSMUSG00000030536/ENSMUSG00000038708/ENSMUSG00000026596/ENSMUSG00000032064/ENSMUSG00000019970/ENSMUSG00000031778/ENSMUSG00000029426/ENSMUSG0000002
0000023951/ENSMUSG00000031540

0000022951/ENSMUSG00000026596/ENSMUSG00000042644/ENSMUSG00000019970/ENSMUSG00000020423/ENSMUSG00000048616/ENSMUSG00000020745/ENSMUSG00000005442/ENSMUSG0000005
0000018736/ENSMUSG00000035764/ENSMUSG00000020745
0000048616/ENSMUSG00000023951/ENSMUSG00000031548/ENSMUSG00000031393/ENSMUSG00000074934/ENSMUSG00000041702
0000024085/ENSMUSG00000069895
0000074934/ENSMUSG00000026812
0000074934/ENSMUSG00000026812
0000031523/ENSMUSG00000022425/ENSMUSG00000023951/ENSMUSG00000040945/ENSMUSG00000039952/ENSMUSG00000074934/ENSMUSG00000026812
0000025373/ENSMUSG00000031778/ENSMUSG00000031451/ENSMUSG00000007613/ENSMUSG00000005533/ENSMUSG00000023951/ENSMUSG00000042594/ENSMUSG00000006445/ENSMUSG0000002
0000025408/ENSMUSG00000032115/ENSMUSG00000031778/ENSMUSG00000026510/ENSMUSG00000004040/ENSMUSG00000020423/ENSMUSG00000024789/ENSMUSG00000032340/ENSMUSG0000003
0000056758/ENSMUSG00000039286/ENSMUSG00000019997/ENSMUSG00000048616/ENSMUSG00000004846/ENSMUSG00000045216/ENSMUSG00000023951/ENSMUSG00000005442/ENSMUSG0000002
0000030747/ENSMUSG00000050697
0000038418/ENSMUSG00000004040/ENSMUSG00000056216/ENSMUSG00000020601/ENSMUSG00000024789/ENSMUSG00000056501/ENSMUSG00000024927/ENSMUSG00000003810/ENSMUSG0000001
0000019817/ENSMUSG00000051510/ENSMUSG00000038615/ENSMUSG00000027678/ENSMUSG00000025408/ENSMUSG00000046711/ENSMUSG00000056758/ENSMUSG00000000078/ENSMUSG0000000
0000033323/ENSMUSG00000023951/ENSMUSG00000022752/ENSMUSG00000024431
0000028521/ENSMUSG00000003665/ENSMUSG00000021978/ENSMUSG00000029201/ENSMUSG00000047126/ENSMUSG00000039497
0000045216/ENSMUSG00000021978/ENSMUSG00000029201/ENSMUSG00000039497

24182

16024/ENSMUSG00000028277/ENSMUSG00000031438

18476/ENSMUSG00000026836/ENSMUSG00000024431/ENSMUSG00000074934/ENSMUSG00000026812/ENSMUSG00000005871

21379/ENSMUSG00000058070/ENSMUSG00000038539/ENSMUSG00000065954/ENSMUSG00000044167/ENSMUSG00000039844

24789/ENSMUSG00000027996/ENSMUSG00000031523/ENSMUSG00000035199/ENSMUSG00000022443/ENSMUSG00000022037/ENSMUSG00000025234/ENSMUSG00000033717/ENSMUSG00000029249/E

24789/ENSMUSG00000049792/ENSMUSG00000035199/ENSMUSG00000024927/ENSMUSG00000018339/ENSMUSG00000018476/ENSMUSG00000024423/ENSMUSG00000050965/ENSMUSG00000079037/E

63142/ENSMUSG00000026313/ENSMUSG00000032487/ENSMUSG00000033717/ENSMUSG00000044167/ENSMUSG00000024431/ENSMUSG00000019943

24431

31451/ENSMUSG00000026728/ENSMUSG00000024927/ENSMUSG00000048234/ENSMUSG00000037815/ENSMUSG00000018476/ENSMUSG00000028161/ENSMUSG00000024423/ENSMUSG00000079037/E

24085/ENSMUSG00000069895/ENSMUSG00000039952

05533/ENSMUSG00000024927/ENSMUSG00000028042/ENSMUSG00000050965/ENSMUSG00000032702/ENSMUSG00000018634/ENSMUSG00000079037/ENSMUSG00000000058/ENSMUSG00000038859/E

24927/ENSMUSG00000028042/ENSMUSG00000050965/ENSMUSG00000032702/ENSMUSG00000018634/ENSMUSG00000000058/ENSMUSG00000038859/ENSMUSG00000024052/ENSMUSG00000044167/E

28042/ENSMUSG00000030168/ENSMUSG00000032487/ENSMUSG00000018707/ENSMUSG00000005871

28641/ENSMUSG00000002489/ENSMUSG00000028613/ENSMUSG00000018736/ENSMUSG00000022425/ENSMUSG00000007613/ENSMUSG00000005533/ENSMUSG00000023951/ENSMUSG00000027834/E

69991/ENSMUSG00000050965/ENSMUSG00000018634/ENSMUSG00000036564/ENSMUSG00000079037/ENSMUSG00000069895/ENSMUSG00000032487/ENSMUSG00000000184/ENSMUSG00000020091/E

24486/ENSMUSG00000040268/ENSMUSG00000039952/ENSMUSG00000039844/ENSMUSG00000044857

30341/ENSMUSG00000056501/ENSMUSG00000023951/ENSMUSG00000061436/ENSMUSG00000022270/ENSMUSG00000063142/ENSMUSG00000018634/ENSMUSG00000026313/ENSMUSG00000079037/E

24085/ENSMUSG00000069895/ENSMUSG00000039952

16024/ENSMUSG00000028277/ENSMUSG00000031438

04040/ENSMUSG00000056216/ENSMUSG00000052942/ENSMUSG00000001419/ENSMUSG00000030557/ENSMUSG00000037465/ENSMUSG00000023034/ENSMUSG00000033863/ENSMUSG00000029135/E

ENSMUSG00000028793/ENSMUSG00000032350/ENSMUSG00000035900/ENSMUSG00000038068/ENSMUSG00000056071/ENSMUSG00000002083/ENSMUSG00000024182/ENSMUSG00000002365

ENSMUSG00000040268/ENSMUSG00000032487/ENSMUSG00000029249/ENSMUSG00000044167/ENSMUSG00000032350/ENSMUSG00000015839/ENSMUSG00000028211/ENSMUSG00000009630/ENSMUS

ENSMUSG00000040268/ENSMUSG00000029249/ENSMUSG00000044167/ENSMUSG00000015839/ENSMUSG00000028211/ENSMUSG00000031548/ENSMUSG00000024431/ENSMUSG00000030287/ENSMUS

ENSMUSG00000024052/ENSMUSG00000044167/ENSMUSG00000029361/ENSMUSG00000020091/ENSMUSG00000042903/ENSMUSG00000019943/ENSMUSG00000026826/ENSMUSG00000050697/ENSMUS

ENSMUSG00000029361/ENSMUSG00000020091/ENSMUSG00000042903/ENSMUSG00000019943/ENSMUSG00000026826/ENSMUSG00000050697/ENSMUSG00000026812/ENSMUSG00000027712/ENSMUS

ENSMUSG00000020745/ENSMUSG00000036606/ENSMUSG00000024924/ENSMUSG00000026313/ENSMUSG00000036564/ENSMUSG00000062232/ENSMUSG00000020132/ENSMUSG00000015839/ENSMUS

ENSMUSG00000031393/ENSMUSG00000074923

ENSMUSG00000022037/ENSMUSG00000029816/ENSMUSG00000027665/ENSMUSG00000029249/ENSMUSG00000032350/ENSMUSG00000029361/ENSMUSG00000024431/ENSMUSG00000026797/ENSMUS

ENSMUSG00000056501/ENSMUSG00000052040/ENSMUSG00000051495/ENSMUSG00000024927/ENSMUSG00000000134/ENSMUSG00000028042/ENSMUSG00000048249/ENSMUSG00000008398/ENSMUS

;G00000026826/ENSMUSG00000050697/ENSMUSG00000037902/ENSMUSG00000026812/ENSMUSG00000004207

;G00000037902/ENSMUSG00000004207

;G00000026812/ENSMUSG00000027712/ENSMUSG00000005871/ENSMUSG00000034593

;G00000005871/ENSMUSG00000034593

;G00000042211/ENSMUSG00000024431/ENSMUSG00000039844/ENSMUSG00000031393/ENSMUSG00000023027/ENSMUSG00000056071/ENSMUSG00000025372/ENSMUSG00000005871

;G00000031393/ENSMUSG00000026826/ENSMUSG00000002083/ENSMUSG00000026812

;G00000027230/ENSMUSG00000029249/ENSMUSG00000015839/ENSMUSG00000025612/ENSMUSG00000024431/ENSMUSG00000023027/ENSMUSG00000026826

| Category | GOID | Description | GeneRatio | BgRatio | pvalue | padj | geneID | geneName |
|---|---|---|---|---|---|---|---|---|
| BP | GO:00311( | axon regeneration | 13\|1209 | 51\|16097 | 7.30E-05 | 0.00124 | ENSMUS | Klf4/Xylt1/Jun/Rtn4/Tnc/Ndel1/Jak2/Neo1/Igf1r/Lamb2/Epha4/ |
| BP | GO:004677 | protein autophosphorylation | 35\|1209 | 235\|16097 | 7.52E-05 | 0.00127 | ENSMUS | Errfi1/Fgfr1/Met/Hspa9/Jun/Iqgap1/Eif2ak3/Abl2/Fgfr2/Thy1/Eg |
| BP | GO:001474 | negative regulation of muscle hypertrophy | 10\|1209 | 32\|16097 | 7.68E-05 | 0.00129 | ENSMUS | Errfi1/Igfbp5/Rgs2/Lmna/Tnfrsf1a/Ctdp1/Tomm70a/Pi16/Notch |
| BP | GO:004568 | negative regulation of glial cell differentiation | 10\|1209 | 32\|16097 | 7.68E-05 | 0.00129 | ENSMUS | Ldlr/Hmga2/Nr1d1/Nog/Dusp10/Notch1/Id4/Atf5/Hes1/Mecp2 |
| BP | GO:01061( | cold-induced thermogenesis | 24\|1209 | 137\|16097 | 7.95E-05 | 0.00133 | ENSMUS | Scd1/Atf4/Ddit3/Jak2/Arrdc3/Fabp4/Lnpep/Nr1d1/Cd36/Igf1r/ |
| BP | GO:012016 | regulation of cold-induced thermogenesis | 24\|1209 | 137\|16097 | 7.95E-05 | 0.00133 | ENSMUS | Scd1/Atf4/Ddit3/Jak2/Arrdc3/Fabp4/Lnpep/Nr1d1/Cd36/Igf1r/ |
| CC | GO:004329 | contractile fiber | 33\|1232 | 216\|16313 | 8.03E-05 | 0.00229 | ENSMUS | Acta1/Myl1/Plec/Slc2a1/Ahnak/Pecam1/Cst3/Tpm1/Tpm2/Pde |
| MF | GO:003525 | glucocorticoid receptor binding | 8\|1190 | 21\|15912 | 8.05E-05 | 0.00347 | ENSMUS | Stat3/Nr4a1/Cebpb/Nrip1/Ywhah/Tacc1/Nr4a2/Ncoa2 |
| BP | GO:000188 | endothelial cell morphogenesis | 7\|1209 | 16\|16097 | 8.28E-05 | 0.00138 | ENSMUS | Tnmd/Met/Pecam1/Col18a1/Clic4/Plod3/Notch4 |
| BP | GO:00030( | heart morphogenesis | 37\|1209 | 255\|16097 | 8.31E-05 | 0.00138 | ENSMUS | Thbs1/Sox9/Has2/Jun/Efna1/Rtn4/Cyr61/Tpm1/Fgfr2/Rxra/Tgfb |
| MF | GO:00052( | structural constituent of cytoskeleton | 14\|1190 | 59\|15912 | 8.68E-05 | 0.00357 | ENSMUS | Tubb2a/Plec/Tuba1a/Tubb4b/Tpm1/Tpm2/Tubb2b/Vim/Tuba1 |
| BP | GO:007099 | neuron death | 50\|1209 | 383\|16097 | 8.78E-05 | 0.00146 | ENSMUS | Mt1/Hmox1/Egr1/Serpinf1/Atf4/Jun/Aars/Ddit3/Fos/Hyou1/Cd |
| MF | GO:000471 | protein tyrosine kinase activity | 22\|1190 | 122\|15912 | 9.38E-05 | 0.0037 | ENSMUS | Map2k3/Hipk1/Fgfr1/Met/Abl2/Fgfr2/Egfr/Jak2/Nrp1/Igf1r/Flt4 |
| BP | GO:003009 | myeloid cell differentiation | 49\|1209 | 374\|16097 | 9.41E-05 | 0.00156 | ENSMUS | Myc/Tlr2/Trib1/Inhba/Hspa9/Cd109/Cdkn1c/Jun/Nfe2l1/Smad |
| BP | GO:003268 | regulation of tumor necrosis factor production | 26\|1209 | 156\|16097 | 9.69E-05 | 0.0016 | ENSMUS | Errfi1/Thbs1/Tlr2/Cd14/Lpl/Akap12/Cd34/Arfgef2/Cx3cl1/Jak2/ |
| MF | GO:004633 | SMAD binding | 17\|1190 | 82\|15912 | 9.85E-05 | 0.00372 | ENSMUS | Col3a1/Col1a2/Col5a2/Jun/Fos/Tgfbr3/Mef2a/Ski/Pmepa1/Tgfb |
| BP | GO:000166 | response to hypoxia | 30\|1209 | 192\|16097 | 9.98E-05 | 0.00164 | ENSMUS | Hmox1/Myc/Vasn/Egr1/Eif4ebp1/Rtn4/Hyou1/Cd34/Fosl2/Fnd |
| BP | GO:000341 | chondrocyte differentiation involved in endochondral bone m | 9\|1209 | 27\|16097 | 0.0001 | 0.00164 | ENSMUS | Col14a1/Sox9/Col12a1/Col6a2/Matn3/Col6a3/Matn1/Col6a1/ |
| BP | GO:005109 | regulation of DNA-binding transcription factor activity | 49\|1209 | 375\|16097 | 0.0001 | 0.00164 | ENSMUS | Fosl1/Hmox1/Tlr2/Trib1/Klf4/Ifrd1/Ncoa3/Lrrfip1/Ddit3/Hmga |
| BP | GO:001095 | negative regulation of endopeptidase activity | 24\|1209 | 139\|16097 | 0.0001 | 0.00164 | ENSMUS | Hspa1b/Thbs1/Serpine1/Klf4/Herpud1/Serpinf1/Serping1/Anxa |
| BP | GO:190189 | positive regulation of cell junction assembly | 10\|1209 | 33\|16097 | 0.0001 | 0.00167 | ENSMUS | S100a10/Iqgap1/Thy1/Nrp1/Enpp2/Vegfa/Epha2/Tek/Kdr/Tsc1 |
| BP | GO:001401 | positive regulation of gliogenesis | 17\|1209 | 82\|16097 | 0.0001 | 0.00168 | ENSMUS | Myc/Tlr2/Rtn4/Tiam1/Il6st/Enpp2/Nog/Vim/Igf1/Rela/Notch1/ |
| BP | GO:004586 | negative regulation of proteolysis | 40\|1209 | 287\|16097 | 0.00011 | 0.00174 | ENSMUS | Hspa1b/Thbs1/Serpine1/Chac1/Klf4/Herpud1/Serpinf1/A2m/Cd |
| BP | GO:001612 | sterol biosynthetic process | 12\|1209 | 46\|16097 | 0.00011 | 0.00174 | ENSMUS | Hmgcr/Cyp51/Fdft1/Msmo1/Hmgcs1/Sqle/Insig1/Hsd17b7/Dhc |
| BP | GO:003629 | response to decreased oxygen levels | 31\|1209 | 202\|16097 | 0.00011 | 0.00174 | ENSMUS | Hmox1/Myc/Vasn/Egr1/Eif4ebp1/Rtn4/Hyou1/Cd34/Fosl2/Fnd |
| BP | GO:007037 | positive regulation of ERK1 and ERK2 cascade | 30\|1209 | 193\|16097 | 0.00011 | 0.00176 | ENSMUS | Spry2/Hmgcr/Tlr2/Jun/Akap12/Abl2/Fgfr2/Cx3cl1/Egfr/Ctgf/C1 |
| BP | GO:001081 | negative regulation of cell-substrate adhesion | 14\|1209 | 60\|16097 | 0.00011 | 0.00176 | ENSMUS | Col1a1/Thbs1/Postn/Serpine1/Plet1/Lgals1/Mmp14/Bcl6/Dlc1/ |
| BP | GO:00311( | neuron projection regeneration | 14\|1209 | 60\|16097 | 0.00011 | 0.00176 | ENSMUS | Klf4/Xylt1/Jun/Rtn4/Thy1/Tnc/Ndel1/Jak2/Neo1/Igf1r/Lamb2/ |
| BP | GO:004592 | negative regulation of growth | 36\|1209 | 249\|16097 | 0.00011 | 0.00176 | ENSMUS | Igfbp5/Rgs2/Inhba/Ifrd1/Rtn4/Cdkn1a/Sertad2/Sesn2/P3h1/Bcl |
| BP | GO:00321( | positive regulation of response to external stimulus | 40\|1209 | 288\|16097 | 0.00012 | 0.00184 | ENSMUS | Thbs1/Ldlr/Tlr2/Serpine1/Fgfr1/Lpl/Met/Rtn4/Smoc2/Bmp6/Cx |
| BP | GO:000181 | positive regulation of cytokine production | 52\|1209 | 408\|16097 | 0.00012 | 0.00188 | ENSMUS | Sulf2/Thbs1/Tlr2/Postn/Dlk1/Serpine1/Egr1/Sec14l1/Cd14/Lpl/ |
| BP | GO:190355 | regulation of tumor necrosis factor superfamily cytokine produ | 26\|1209 | 158\|16097 | 0.00012 | 0.00188 | ENSMUS | Errfi1/Thbs1/Tlr2/Cd14/Lpl/Akap12/Cd34/Arfgef2/Cx3cl1/Jak2/ |
| BP | GO:003462 | cellular response to unfolded protein | 17\|1209 | 83\|16097 | 0.00012 | 0.00188 | ENSMUS | Hspa1b/Hspa1a/Herpud1/Hspa9/Atf4/Eif2ak3/Ddit3/Xbp1/Ccn |
| BP | GO:190403 | regulation of epithelial cell apoptotic process | 17\|1209 | 83\|16097 | 0.00012 | 0.00188 | ENSMUS | Thbs1/Hmox1/Serpine1/Col18a1/Abl2/Xbp1/Jak2/Gas6/Angptl |
| BP | GO:200011 | negative regulation of cysteine-type endopeptidase activity | 17\|1209 | 83\|16097 | 0.00012 | 0.00188 | ENSMUS | Hspa1b/Thbs1/Klf4/Herpud1/Dhcr24/Nr4a1/Gas6/Sfrp2/Igf1/C |
| BP | GO:000726 | nitric oxide mediated signal transduction | 8\|1209 | 22\|16097 | 0.00012 | 0.00188 | ENSMUS | Mt1/Thbs1/Egfr/Cd36/Vegfa/Rasd1/Nos1/Ndnf |
| BP | GO:007187 | adrenergic receptor signaling pathway | 8\|1209 | 22\|16097 | 0.00012 | 0.00188 | ENSMUS | Rgs2/Pde4b/Arrdc3/Gnai2/Rapgef2/Adra2a/Nos1/Gnas |
| BP | GO:19033( | negative regulation of regulated secretory pathway | 8\|1209 | 22\|16097 | 0.00012 | 0.00188 | ENSMUS | Hmox1/Rabgef1/Notch1/Gnai2/Lgals9/Adra2a/Rest/Cbarp |
| BP | GO:00719( | negative regulation of protein serine/threonine kinase activity | 20\|1209 | 107\|16097 | 0.00012 | 0.00189 | ENSMUS | Spry2/Hmgcr/Rgs2/Dusp1/Cdkn1a/Spred1/Dusp5/Sfrp2/Dusp1 |
| BP | GO:000743 | salivary gland development | 11\|1209 | 40\|16097 | 0.00012 | 0.00191 | ENSMUS | Fgfr1/Met/Fgfr2/Egfr/Xbp1/Nrp1/Twsg1/Plxnd1/Tgm2/Dag1/Bt |
| BP | GO:003313 | positive regulation of peptidyl-serine phosphorylation | 18\|1209 | 91\|16097 | 0.00012 | 0.00191 | ENSMUS | Spry2/Fnip2/Met/Egfr/Gas6/Sfrp2/Eif4g1/Tnks1bp1/Cd44/Vegfa |
| BP | GO:190303 | positive regulation of response to wounding | 15\|1209 | 68\|16097 | 0.00013 | 0.00192 | ENSMUS | Thbs1/Serpine1/Rtn4/Smoc2/Mylk/Xbp1/Ndel1/Cd36/Igf1r/Kar |
| BP | GO:003313 | regulation of peptidyl-serine phosphorylation | 22\|1209 | 124\|16097 | 0.00013 | 0.00195 | ENSMUS | Spry2/Fnip2/Met/Egfr/Gas6/Sfrp2/Eif4g1/Tnks1bp1/Cd44/Vegfa |
| BP | GO:000701 | actin filament organization | 50\|1209 | 389\|16097 | 0.00013 | 0.00197 | ENSMUS | Acta1/Tlr2/Eln/Marcks/S100a10/Met/Cald1/Sorbs2/Pecam1/Tp |
| BP | GO:004689 | regulation of lipid biosynthetic process | 25\|1209 | 150\|16097 | 0.00013 | 0.00198 | ENSMUS | Ldlr/Egr1/Stard4/Atp1a1/Acsl3/Cyr61/Insig1/Bmp6/Trib3/Sik1/ |
| BP | GO:004871 | negative regulation of astrocyte differentiation | 7\|1209 | 17\|16097 | 0.00013 | 0.00199 | ENSMUS | Ldlr/Hmga2/Nr1d1/Nog/Id4/Atf5/Mecp2 |
| MF | GO:004323 | laminin binding | 9\|1190 | 28\|15912 | 0.00013 | 0.00483 | ENSMUS | Thbs1/Nid1/Lgals3/Lgals1/Thbs4/Ssc5d/Adgrg6/Dag1/Itga2 |
| BP | GO:003264 | tumor necrosis factor production | 26\|1209 | 159\|16097 | 0.00013 | 0.00201 | ENSMUS | Errfi1/Thbs1/Tlr2/Cd14/Lpl/Akap12/Cd34/Arfgef2/Cx3cl1/Jak2/ |
| BP | GO:00456( | regulation of endothelial cell differentiation | 10\|1209 | 34\|16097 | 0.00014 | 0.00203 | ENSMUS | Cdh5/Bmp6/Tnfrsf1a/Tgfbr1/Vegfa/Notch1/Cldn5/Apold1/Akap |
| BP | GO:000704 | cell-substrate adherens junction assembly | 16\|1209 | 76\|16097 | 0.00014 | 0.00203 | ENSMUS | S100a10/Iqgap1/Mmp14/Thy1/Nrp1/Dlc1/Enpp2/Actn2/Vegfa/ |
| BP | GO:004804 | focal adhesion assembly | 16\|1209 | 76\|16097 | 0.00014 | 0.00203 | ENSMUS | S100a10/Iqgap1/Mmp14/Thy1/Nrp1/Dlc1/Enpp2/Actn2/Vegfa/ |

| Count | Up | Up_Gene_id | Down | Down_Gene_id |
|---|---|---|---|---|
| 13 | 6 | ENSMUSG0 | 7 | ENSMUSG00000003032/ENSMUSG00000030657/ENSMUSG00000020458/ENSMUSG00000018736/ENSMUSG00000024789/ENSMUSG00000032340/ENSMUSG0 |
| 35 | 11 | ENSMUSG0 | 24 | ENSMUSG00000028967/ENSMUSG00000031565/ENSMUSG00000009376/ENSMUSG00000024359/ENSMUSG00000030536/ENSMUSG00000031668/ENSMUSG0 |
| 10 | 3 | ENSMUSG0 | 7 | ENSMUSG00000028967/ENSMUSG00000026360/ENSMUSG00000028063/ENSMUSG00000030341/ENSMUSG00000033323/ENSMUSG00000022752/ENSMUSG0 |
| 10 | 2 | ENSMUSG0 | 8 | ENSMUSG00000032193/ENSMUSG00000056758/ENSMUSG00000020889/ENSMUSG00000048616/ENSMUSG00000039384/ENSMUSG00000021379/ENSMUSG0 |
| 24 | 8 | ENSMUSG0 | 16 | ENSMUSG00000037071/ENSMUSG00000042406/ENSMUSG00000025408/ENSMUSG00000024789/ENSMUSG00000074794/ENSMUSG00000023845/ENSMUSG0 |
| 24 | 8 | ENSMUSG0 | 16 | ENSMUSG00000037071/ENSMUSG00000042406/ENSMUSG00000025408/ENSMUSG00000024789/ENSMUSG00000074794/ENSMUSG00000023845/ENSMUSG0 |
| 33 | 18 | ENSMUSG0 | 15 | ENSMUSG00000022565/ENSMUSG00000028645/ENSMUSG00000069833/ENSMUSG00000027447/ENSMUSG00000028525/ENSMUSG00000096054/ENSMUSG0 |
| 8 | 0 | | 8 | ENSMUSG00000004040/ENSMUSG00000023034/ENSMUSG00000056501/ENSMUSG00000048490/ENSMUSG00000018965/ENSMUSG00000065954/ENSMUSG0 |
| 7 | 4 | ENSMUSG0 | 3 | ENSMUSG00000009376/ENSMUSG00000037242/ENSMUSG00000004846 |
| 37 | 20 | ENSMUSG0 | 17 | ENSMUSG00000040152/ENSMUSG00000000567/ENSMUSG00000022367/ENSMUSG00000027954/ENSMUSG00000020458/ENSMUSG00000030849/ENSMUSG0 |
| 14 | 11 | ENSMUSG0 | 3 | ENSMUSG00000022565/ENSMUSG00000026728/ENSMUSG00000053797 |
| 50 | 13 | ENSMUSG0 | 37 | ENSMUSG00000031765/ENSMUSG00000005413/ENSMUSG00000038418/ENSMUSG00000000753/ENSMUSG00000042406/ENSMUSG00000031960/ENSMUSG0 |
| 22 | 10 | ENSMUSG0 | 12 | ENSMUSG00000018932/ENSMUSG00000008730/ENSMUSG00000031565/ENSMUSG00000009376/ENSMUSG00000026596/ENSMUSG00000030849/ENSMUSG0 |
| 49 | 15 | ENSMUSG0 | 34 | ENSMUSG00000022346/ENSMUSG00000027995/ENSMUSG00000032501/ENSMUSG00000041324/ENSMUSG00000024359/ENSMUSG00000046186/ENSMUSG0 |
| 26 | 9 | ENSMUSG0 | 17 | ENSMUSG00000028967/ENSMUSG00000040152/ENSMUSG00000027995/ENSMUSG00000051439/ENSMUSG00000074582/ENSMUSG00000031778/ENSMUSG0 |
| 17 | 7 | ENSMUSG0 | 10 | ENSMUSG00000029287/ENSMUSG00000030557/ENSMUSG00000029050/ENSMUSG00000038400/ENSMUSG00000007613/ENSMUSG00000046058/ENSMUSG0 |
| 30 | 10 | ENSMUSG0 | 20 | ENSMUSG00000005413/ENSMUSG00000022346/ENSMUSG00000039646/ENSMUSG00000038418/ENSMUSG00000031490/ENSMUSG00000020458/ENSMUSG0 |
| 9 | 5 | ENSMUSG0 | 4 | ENSMUSG00000000567/ENSMUSG00000020241/ENSMUSG00000048126/ENSMUSG00000001119 |
| 49 | 14 | ENSMUSG0 | 35 | ENSMUSG00000024912/ENSMUSG00000005413/ENSMUSG00000027995/ENSMUSG00000032501/ENSMUSG00000003032/ENSMUSG00000001627/ENSMUSG0 |
| 24 | 4 | ENSMUSG0 | 20 | ENSMUSG00000040152/ENSMUSG00000037411/ENSMUSG00000003032/ENSMUSG00000031770/ENSMUSG00000000753/ENSMUSG00000021950/ENSMUSG0 |
| 10 | 4 | ENSMUSG0 | 6 | ENSMUSG00000041959/ENSMUSG00000030536/ENSMUSG00000022425/ENSMUSG00000023951/ENSMUSG00000006445/ENSMUSG00000026812 |
| 17 | 7 | ENSMUSG0 | 10 | ENSMUSG00000022346/ENSMUSG00000027995/ENSMUSG00000020458/ENSMUSG00000002489/ENSMUSG00000021756/ENSMUSG00000022425/ENSMUSG0 |
| 40 | 9 | ENSMUSG0 | 31 | ENSMUSG00000040152/ENSMUSG00000037411/ENSMUSG00000027313/ENSMUSG00000003032/ENSMUSG00000031770/ENSMUSG00000000753/ENSMUSG0 |
| 12 | 0 | | 12 | ENSMUSG00000021670/ENSMUSG00000001467/ENSMUSG00000021273/ENSMUSG00000031604/ENSMUSG00000093930/ENSMUSG00000022351/ENSMUSG0 |
| 31 | 10 | ENSMUSG0 | 21 | ENSMUSG00000005413/ENSMUSG00000022346/ENSMUSG00000039646/ENSMUSG00000038418/ENSMUSG00000031490/ENSMUSG00000020458/ENSMUSG0 |
| 30 | 14 | ENSMUSG0 | 16 | ENSMUSG00000022114/ENSMUSG00000021670/ENSMUSG00000027995/ENSMUSG00000026596/ENSMUSG00000030849/ENSMUSG00000031778/ENSMUSG0 |
| 14 | 5 | ENSMUSG0 | 9 | ENSMUSG00000040152/ENSMUSG00000037411/ENSMUSG00000032068/ENSMUSG00000022508/ENSMUSG00000031523/ENSMUSG00000022425/ENSMUSG0 |
| 14 | 7 | ENSMUSG0 | 7 | ENSMUSG00000003032/ENSMUSG00000030657/ENSMUSG00000020458/ENSMUSG00000018736/ENSMUSG00000024789/ENSMUSG00000032340/ENSMUSG0 |
| 36 | 13 | ENSMUSG0 | 23 | ENSMUSG00000026360/ENSMUSG00000041324/ENSMUSG00000001627/ENSMUSG00000020458/ENSMUSG00000023067/ENSMUSG00000049800/ENSMUSG0 |
| 40 | 13 | ENSMUSG0 | 27 | ENSMUSG00000040152/ENSMUSG00000032193/ENSMUSG00000027995/ENSMUSG00000037411/ENSMUSG00000031565/ENSMUSG00000009376/ENSMUSG0 |
| 52 | 14 | ENSMUSG0 | 38 | ENSMUSG00000006800/ENSMUSG00000040152/ENSMUSG00000027995/ENSMUSG00000040856/ENSMUSG00000037411/ENSMUSG00000038418/ENSMUSG0 |
| 26 | 9 | ENSMUSG0 | 17 | ENSMUSG00000028967/ENSMUSG00000040152/ENSMUSG00000027995/ENSMUSG00000051439/ENSMUSG00000074582/ENSMUSG00000031778/ENSMUSG0 |
| 17 | 3 | ENSMUSG0 | 14 | ENSMUSG00000031770/ENSMUSG00000024359/ENSMUSG00000042406/ENSMUSG00000031668/ENSMUSG00000025408/ENSMUSG00000020484/ENSMUSG0 |
| 17 | 8 | ENSMUSG0 | 9 | ENSMUSG00000040152/ENSMUSG00000005413/ENSMUSG00000037411/ENSMUSG00000026596/ENSMUSG00000020484/ENSMUSG00000024789/ENSMUSG0 |
| 17 | 3 | ENSMUSG0 | 14 | ENSMUSG00000040152/ENSMUSG00000003032/ENSMUSG00000031770/ENSMUSG00000034926/ENSMUSG00000023034/ENSMUSG00000031451/ENSMUSG0 |
| 8 | 4 | ENSMUSG0 | 4 | ENSMUSG00000031765/ENSMUSG00000040152/ENSMUSG00000023951/ENSMUSG00000029361 |
| 8 | 2 | ENSMUSG0 | 6 | ENSMUSG00000026360/ENSMUSG00000028525/ENSMUSG00000074794/ENSMUSG00000062232/ENSMUSG00000033717/ENSMUSG00000029361 |
| 8 | 3 | ENSMUSG0 | 5 | ENSMUSG00000005413/ENSMUSG00000025340/ENSMUSG00000033717/ENSMUSG00000029249/ENSMUSG00000035640 |
| 20 | 4 | ENSMUSG0 | 16 | ENSMUSG00000022114/ENSMUSG00000021670/ENSMUSG00000026360/ENSMUSG00000023067/ENSMUSG00000027351/ENSMUSG00000034765/ENSMUSG0 |
| 11 | 3 | ENSMUSG0 | 8 | ENSMUSG00000031565/ENSMUSG00000009376/ENSMUSG00000030849/ENSMUSG00000020484/ENSMUSG00000024098/ENSMUSG00000037820/ENSMUSG0 |
| 18 | 3 | ENSMUSG0 | 15 | ENSMUSG00000022114/ENSMUSG00000061175/ENSMUSG00000009376/ENSMUSG00000031451/ENSMUSG00000027996/ENSMUSG00000045983/ENSMUSG0 |
| 15 | 4 | ENSMUSG0 | 11 | ENSMUSG00000040152/ENSMUSG00000037411/ENSMUSG00000020458/ENSMUSG00000020484/ENSMUSG00000018736/ENSMUSG00000005533/ENSMUSG0 |
| 22 | 4 | ENSMUSG0 | 18 | ENSMUSG00000022114/ENSMUSG00000061175/ENSMUSG00000009376/ENSMUSG00000031451/ENSMUSG00000027996/ENSMUSG00000045983/ENSMUSG0 |
| 50 | 28 | ENSMUSG0 | 22 | ENSMUSG00000027995/ENSMUSG00000029675/ENSMUSG00000041959/ENSMUSG00000009376/ENSMUSG00000026596/ENSMUSG00000031778/ENSMUSG0 |
| 25 | 7 | ENSMUSG0 | 18 | ENSMUSG00000032193/ENSMUSG00000038418/ENSMUSG00000024378/ENSMUSG00000033161/ENSMUSG00000032883/ENSMUSG00000045294/ENSMUSG0 |
| 7 | 0 | | 7 | ENSMUSG00000032193/ENSMUSG00000056758/ENSMUSG00000020889/ENSMUSG00000048616/ENSMUSG00000021379/ENSMUSG00000038539/ENSMUSG0 |
| 9 | 6 | ENSMUSG0 | 3 | ENSMUSG00000040152/ENSMUSG00000050335/ENSMUSG00000039952 |
| 26 | 9 | ENSMUSG0 | 17 | ENSMUSG00000028967/ENSMUSG00000040152/ENSMUSG00000027995/ENSMUSG00000051439/ENSMUSG00000074582/ENSMUSG00000031778/ENSMUSG0 |
| 10 | 5 | ENSMUSG0 | 5 | ENSMUSG00000039004/ENSMUSG00000030341/ENSMUSG00000007613/ENSMUSG00000023951/ENSMUSG00000022016 |
| 16 | 8 | ENSMUSG0 | 8 | ENSMUSG00000041959/ENSMUSG00000030536/ENSMUSG00000031523/ENSMUSG00000022425/ENSMUSG00000023951/ENSMUSG00000040945/ENSMUSG0 |
| 16 | 8 | ENSMUSG0 | 8 | ENSMUSG00000041959/ENSMUSG00000030536/ENSMUSG00000031523/ENSMUSG00000022425/ENSMUSG00000023951/ENSMUSG00000040945/ENSMUSG0 |

0000005533
0000026596/ENSMUSG00000030849/ENSMUSG00000020349/ENSMUSG00000024789/ENSMUSG00000037012/ENSMUSG00000007613/ENSMUSG00000005533/ENSMUSG00000033955/ENSMUSG0000002
0000044167
0000038539/ENSMUSG00000031393
0000020889/ENSMUSG00000005533/ENSMUSG00000056501/ENSMUSG00000023951/ENSMUSG00000000134/ENSMUSG00000018476/ENSMUSG00000028042/ENSMUSG00000030168/ENSMUSG0000001
0000020889/ENSMUSG00000005533/ENSMUSG00000056501/ENSMUSG00000023951/ENSMUSG00000000134/ENSMUSG00000018476/ENSMUSG00000028042/ENSMUSG00000030168/ENSMUSG0000001
0000054808/ENSMUSG00000032643/ENSMUSG00000028161/ENSMUSG00000026313/ENSMUSG00000026131/ENSMUSG00000039952/ENSMUSG00000029361/ENSMUSG00000027712/ENSMUSG0000003
0000026826/ENSMUSG00000005886

0000015846/ENSMUSG00000029287/ENSMUSG00000007817/ENSMUSG00000027996/ENSMUSG00000031523/ENSMUSG00000048616/ENSMUSG00000007613/ENSMUSG00000023951/ENSMUSG0000002

0000025408/ENSMUSG00000032115/ENSMUSG00000031778/ENSMUSG00000026510/ENSMUSG00000004040/ENSMUSG00000039701/ENSMUSG00000020423/ENSMUSG00000024789/ENSMUSG0000003
0000024789/ENSMUSG00000005533/ENSMUSG00000027177/ENSMUSG00000061436/ENSMUSG00000006445/ENSMUSG00000028661
0000038615/ENSMUSG00000021540/ENSMUSG00000025373/ENSMUSG00000029287/ENSMUSG00000004040/ENSMUSG00000056216/ENSMUSG00000025153/ENSMUSG00000022508/ENSMUSG0000003
0000024789/ENSMUSG00000031451/ENSMUSG00000030341/ENSMUSG00000037447/ENSMUSG00000022708/ENSMUSG00000022037/ENSMUSG00000016024/ENSMUSG00000029816/ENSMUSG0000002
0000061436/ENSMUSG00000027230/ENSMUSG00000026836/ENSMUSG00000024182
0000032115/ENSMUSG00000029135/ENSMUSG00000023951/ENSMUSG00000063142/ENSMUSG00000024924/ENSMUSG00000018634/ENSMUSG00000036450/ENSMUSG00000029249/ENSMUSG0000003

0000027678/ENSMUSG00000026305/ENSMUSG00000025408/ENSMUSG00000056758/ENSMUSG00000025373/ENSMUSG00000031778/ENSMUSG00000029287/ENSMUSG00000004040/ENSMUSG0000005
0000001131/ENSMUSG00000034926/ENSMUSG00000023034/ENSMUSG00000031451/ENSMUSG00000027996/ENSMUSG00000005087/ENSMUSG00000046223/ENSMUSG00000023951/ENSMUSG0000002

0000048616/ENSMUSG00000026728/ENSMUSG00000024927/ENSMUSG00000039952
0000030111/ENSMUSG00000046186/ENSMUSG00000027954/ENSMUSG00000027447/ENSMUSG00000021950/ENSMUSG00000001131/ENSMUSG00000034926/ENSMUSG00000023034/ENSMUSG0000006
0000045294/ENSMUSG00000026675/ENSMUSG00000034926/ENSMUSG00000032018/ENSMUSG00000033105/ENSMUSG00000050697
0000032115/ENSMUSG00000029135/ENSMUSG00000023951/ENSMUSG00000063142/ENSMUSG00000024924/ENSMUSG00000018634/ENSMUSG00000036450/ENSMUSG00000029249/ENSMUSG0000003
0000019997/ENSMUSG00000031451/ENSMUSG00000005087/ENSMUSG00000023951/ENSMUSG00000044337/ENSMUSG00000050965/ENSMUSG00000036564/ENSMUSG00000062232/ENSMUSG0000002
0000054808/ENSMUSG00000032702/ENSMUSG00000040945
0000005533
0000028893/ENSMUSG00000028641/ENSMUSG00000022508/ENSMUSG00000027996/ENSMUSG00000048616/ENSMUSG00000033323/ENSMUSG00000022752/ENSMUSG00000039384/ENSMUSG0000003
0000020458/ENSMUSG00000039004/ENSMUSG00000031778/ENSMUSG00000002489/ENSMUSG00000018736/ENSMUSG00000024789/ENSMUSG00000031451/ENSMUSG00000020594/ENSMUSG0000003
0000020823/ENSMUSG00000051439/ENSMUSG00000027947/ENSMUSG00000042406/ENSMUSG00000020458/ENSMUSG00000031668/ENSMUSG00000025408/ENSMUSG00000026596/ENSMUSG0000002
0000024789/ENSMUSG00000031451/ENSMUSG00000030341/ENSMUSG00000037447/ENSMUSG00000022708/ENSMUSG00000022037/ENSMUSG00000016024/ENSMUSG00000029816/ENSMUSG0000002
0000021831/ENSMUSG00000048249/ENSMUSG00000003814/ENSMUSG00000022365/ENSMUSG00000027230/ENSMUSG00000020571/ENSMUSG00000015839/ENSMUSG00000026628
0000031451/ENSMUSG00000005533/ENSMUSG00000015839
0000027996/ENSMUSG00000005087/ENSMUSG00000046223/ENSMUSG00000023951/ENSMUSG00000047281/ENSMUSG00000032487/ENSMUSG00000035992/ENSMUSG00000036432

0000027996/ENSMUSG00000039384/ENSMUSG00000040021/ENSMUSG00000027177/ENSMUSG00000042594/ENSMUSG00000109511/ENSMUSG00000050965/ENSMUSG00000053716/ENSMUSG0000003
0000039952/ENSMUSG00000041702
0000033955/ENSMUSG00000005087/ENSMUSG00000023951/ENSMUSG00000040021/ENSMUSG00000032487/ENSMUSG00000027665/ENSMUSG00000035992/ENSMUSG00000029361/ENSMUSG0000002
0000032702/ENSMUSG00000024486/ENSMUSG00000033717/ENSMUSG00000015839/ENSMUSG00000056071
0000033955/ENSMUSG00000005087/ENSMUSG00000023951/ENSMUSG00000040021/ENSMUSG00000021070/ENSMUSG00000015202/ENSMUSG00000032487/ENSMUSG00000027665/ENSMUSG0000003
0000019997/ENSMUSG00000031523/ENSMUSG00000032232/ENSMUSG00000007613/ENSMUSG00000022817/ENSMUSG00000040021/ENSMUSG00000054808/ENSMUSG00000024533/ENSMUSG0000004
0000039004/ENSMUSG00000032715/ENSMUSG00000020889/ENSMUSG00000005533/ENSMUSG00000022708/ENSMUSG00000021770/ENSMUSG00000030747/ENSMUSG00000032349/ENSMUSG0000003
0000031393

0000024789/ENSMUSG00000031451/ENSMUSG00000030341/ENSMUSG00000037447/ENSMUSG00000022708/ENSMUSG00000022037/ENSMUSG00000016024/ENSMUSG00000029816/ENSMUSG0000002

0000074934/ENSMUSG00000026812
0000074934/ENSMUSG00000026812

23951/ENSMUSG00000030655/ENSMUSG00000024423/ENSMUSG00000050965/ENSMUSG00000073563/ENSMUSG00000029816/ENSMUSG00000035828/ENSMUSG00000074934/ENSMUSG00000028661/E

18707/ENSMUSG00000005871
18707/ENSMUSG00000005871
31543

26836/ENSMUSG00000031540/ENSMUSG00000044857

32340/ENSMUSG00000030341/ENSMUSG00000049792/ENSMUSG00000056501/ENSMUSG00000023951/ENSMUSG00000061436/ENSMUSG00000022270/ENSMUSG00000063142/ENSMUSG00000018634/E

37465/ENSMUSG00000024789/ENSMUSG00000026361/ENSMUSG00000056501/ENSMUSG00000052040/ENSMUSG00000023951/ENSMUSG00000000134/ENSMUSG00000020745/ENSMUSG00000035673/E
27551/ENSMUSG00000028277/ENSMUSG00000037902

34205/ENSMUSG00000015839/ENSMUSG00000025612/ENSMUSG00000031393/ENSMUSG00000030287/ENSMUSG00000026826

56216/ENSMUSG00000028613/ENSMUSG00000020601/ENSMUSG00000024789/ENSMUSG00000031451/ENSMUSG00000030103/ENSMUSG00000005533/ENSMUSG00000022096/ENSMUSG00000024927/E
27834/ENSMUSG00000047281/ENSMUSG00000020044/ENSMUSG00000032487/ENSMUSG00000035992/ENSMUSG00000036432

57780/ENSMUSG00000031451/ENSMUSG00000027996/ENSMUSG00000049792/ENSMUSG00000005087/ENSMUSG00000038764/ENSMUSG00000046223/ENSMUSG00000023951/ENSMUSG00000027834/E

34205/ENSMUSG00000015839/ENSMUSG00000025612/ENSMUSG00000031393/ENSMUSG00000030287/ENSMUSG00000026826/ENSMUSG00000026812
29816/ENSMUSG00000039844

30168/ENSMUSG00000095098/ENSMUSG00000034201/ENSMUSG00000028179/ENSMUSG00000031548/ENSMUSG00000031393/ENSMUSG00000074934/ENSMUSG00000002083/ENSMUSG00000005871
30341/ENSMUSG00000005533/ENSMUSG00000056501/ENSMUSG00000021508/ENSMUSG00000023951/ENSMUSG00000037820/ENSMUSG00000050965/ENSMUSG00000003814/ENSMUSG00000032487/E
28525/ENSMUSG00000074582/ENSMUSG00000031778/ENSMUSG00000004040/ENSMUSG00000056216/ENSMUSG00000020484/ENSMUSG00000024789/ENSMUSG00000037012/ENSMUSG00000020594/E
27551/ENSMUSG00000028277/ENSMUSG00000037902

31548/ENSMUSG00000005871

24182

35992/ENSMUSG00000029361/ENSMUSG00000050697/ENSMUSG00000024182
40852/ENSMUSG00000032702/ENSMUSG00000038859/ENSMUSG00000044147/ENSMUSG00000025372/ENSMUSG00000037902/ENSMUSG00000026812/ENSMUSG00000002365
32487/ENSMUSG00000029249/ENSMUSG00000026623/ENSMUSG00000024431

27551/ENSMUSG00000028277/ENSMUSG00000037902

ENSMUSG00000003345

ENSMUSG00000026313/ENSMUSG00000079037/ENSMUSG00000022037/ENSMUSG00000029816/ENSMUSG00000027665/ENSMUSG00000029249/ENSMUSG00000032350/ENSMUSG00000029361/ENSMUS

ENSMUSG00000024812/ENSMUSG00000042594/ENSMUSG00000022443/ENSMUSG00000006445/ENSMUSG00000050965/ENSMUSG00000034201/ENSMUSG00000031548/ENSMUSG00000031540/ENSMUS

ENSMUSG00000023951/ENSMUSG00000061436/ENSMUSG00000028161/ENSMUSG00000055491/ENSMUSG00000030527/ENSMUSG00000026313/ENSMUSG00000079037/ENSMUSG00000022037/ENSMUS

ENSMUSG00000024073/ENSMUSG00000061436/ENSMUSG00000047281/ENSMUSG00000020044/ENSMUSG00000032487/ENSMUSG00000035992/ENSMUSG00000036432/ENSMUSG00000028277

ENSMUSG00000016024/ENSMUSG00000035356/ENSMUSG00000056071/ENSMUSG00000022808
ENSMUSG00000000776/ENSMUSG00000030341/ENSMUSG00000030530/ENSMUSG00000056501/ENSMUSG00000037447/ENSMUSG00000022708/ENSMUSG00000020075/ENSMUSG00000024927/ENSMUS

;G00000024431/ENSMUSG00000026797/ENSMUSG00000031393/ENSMUSG00000026826/ENSMUSG00000002083/ENSMUSG00000026812

;G00000024425/ENSMUSG00000031543/ENSMUSG00000005871

;G00000034297/ENSMUSG00000028179/ENSMUSG00000031540/ENSMUSG00000074934

;G00000030059/ENSMUSG00000022037/ENSMUSG00000032487/ENSMUSG00000016024/ENSMUSG00000027551/ENSMUSG00000033717/ENSMUSG00000043421

| Category | GOID | Description | GeneRatio | BgRatio | pvalue | padj | geneID | geneName |
|---|---|---|---|---|---|---|---|---|
| BP | GO:007257 | endothelial cell apoptotic process | 13\|1209 | 54\|16097 | 0.00014 | 0.00205 | ENSMUS | Thbs1/Serpine1/Hipk1/Col18a1/Abl2/Xbp1/Gas6/Angptl4/Cd24 |
| BP | GO:004881 | regulation of dendrite morphogenesis | 20\|1209 | 108\|16097 | 0.00014 | 0.00208 | ENSMUS | Met/Efna1/Ngef/Sgk1/Tiam1/Lrp8/Robo1/Pafah1b1/Nedd4/Syn |
| BP | GO:004639 | carboxylic acid biosynthetic process | 43\|1209 | 320\|16097 | 0.00014 | 0.00208 | ENSMUS | Myc/Acly/Stard4/Has2/Lpl/Scd1/Ier3/Acsl3/Scd2/Insig1/Psat1/S |
| BP | GO:000110 | response to acid chemical | 36\|1209 | 252\|16097 | 0.00014 | 0.00209 | ENSMUS | Col3a1/Col1a1/Col1a2/Col4a1/Ldlr/Tlr2/Sox9/Klf4/Mest/Col5a |
| BP | GO:002195 | central nervous system neuron development | 17\|1209 | 84\|16097 | 0.00014 | 0.00209 | ENSMUS | Fgfr1/Cdh11/Fgfr2/B4galt5/Btg2/Ndel1/Nrp1/Fbxo45/Robo1/P |
| BP | GO:000646 | negative regulation of protein kinase activity | 29\|1209 | 187\|16097 | 0.00015 | 0.00217 | ENSMUS | Errfi1/Spry2/Hmgcr/Rgs2/Trib1/Gadd45a/Dusp1/Cdkn1a/Spred |
| BP | GO:001605 | organic acid biosynthetic process | 43\|1209 | 321\|16097 | 0.00015 | 0.00221 | ENSMUS | Myc/Acly/Stard4/Has2/Lpl/Scd1/Ier3/Acsl3/Scd2/Insig1/Psat1/S |
| BP | GO:005089 | cognition | 41\|1209 | 302\|16097 | 0.00016 | 0.00232 | ENSMUS | Hmgcr/Ldlr/Tlr2/Itga5/Egr1/Serpinf1/Lamb1/Rcan1/Jun/Abl2/It |
| CC | GO:004444 | contractile fiber part | 30\|1232 | 196\|16313 | 0.00016 | 0.00437 | ENSMUS | Acta1/Slc2a1/Ahnak/Pecam1/Tpm1/Tpm2/Pde4b/Synpo/Myh1 |
| BP | GO:003283 | glomerulus development | 13\|1209 | 55\|16097 | 0.00017 | 0.00245 | ENSMUS | Sulf2/Nid1/Egr1/Il6ra/Agtr2/Notch3/Nog/Notch1/Lamb2/Tek/H |
| BP | GO:001820 | peptidyl-serine modification | 41\|1209 | 303\|16097 | 0.00017 | 0.00248 | ENSMUS | Spry2/Fnip2/Hipk1/Met/Mast4/Eif2ak3/Sgk1/Egfr/Gas6/Sfrp2/H |
| BP | GO:000268 | positive regulation of leukocyte migration | 23\|1209 | 135\|16097 | 0.00017 | 0.00248 | ENSMUS | Thbs1/Tlr2/Serpine1/Lgals3/Rtn4/Pecam1/Mmp14/Cx3cl1/Thy |
| BP | GO:003808 | vascular endothelial growth factor signaling pathway | 10\|1209 | 35\|16097 | 0.00018 | 0.00257 | ENSMUS | Spry2/Smoc2/Gab1/Nrp1/Flt4/Robo1/Vegfa/Nus1/Ptp4a3/Kdr |
| BP | GO:003030 | negative regulation of cell growth | 29\|1209 | 189\|16097 | 0.00018 | 0.00258 | ENSMUS | Rgs2/Inhba/Ifrd1/Rtn4/Cdkn1a/Sertad2/Sesn2/P3h1/Bcl6/Nrp1 |
| BP | GO:007170 | tumor necrosis factor superfamily cytokine production | 26\|1209 | 162\|16097 | 0.00018 | 0.00262 | ENSMUS | Errfi1/Thbs1/Tlr2/Cd14/Lpl/Akap12/Cd34/Arfgef2/Cx3cl1/Jak2/ |
| BP | GO:004887 | multicellular organismal homeostasis | 54\|1209 | 436\|16097 | 0.00019 | 0.00268 | ENSMUS | Col14a1/Sox9/Dlk1/Egr1/Met/Scd1/Atf4/Ddit3/Ctsk/Cd34/Bmp |
| BP | GO:003050 | regulation of bone mineralization | 16\|1209 | 78\|16097 | 0.00019 | 0.00268 | ENSMUS | Sox9/Omd/Cyr61/Ank/Bmp6/Rflna/Comp/Mgp/Mia3/Matn1/Fz |
| BP | GO:006044 | branching involved in prostate gland morphogenesis | 6\|1209 | 13\|16097 | 0.00019 | 0.00273 | ENSMUS | Fem1b/Fgfr2/Rxra/Igf1/Cd44/Sfrp1 |
| BP | GO:006141 | regulation of transcription from RNA polymerase II promoter ir | 6\|1209 | 13\|16097 | 0.00019 | 0.00273 | ENSMUS | Egr1/Vegfa/Notch1/Hif1an/Nfe2l2/Bach1 |
| BP | GO:005077 | regulation of dendrite development | 28\|1209 | 181\|16097 | 0.0002 | 0.00283 | ENSMUS | Mapk6/Met/Efna1/Iqgap1/Ngef/Sgk1/Tiam1/Lrp8/Robo1/Pafah |
| BP | GO:009003 | regulation of steroid hormone biosynthetic process | 7\|1209 | 18\|16097 | 0.0002 | 0.00285 | ENSMUS | Atp1a1/Bmp6/Igf1r/Igf1/Dab2/Rest/Nr3c1 |
| BP | GO:006157 | actin filament bundle organization | 26\|1209 | 163\|16097 | 0.0002 | 0.00285 | ENSMUS | Eln/Marcks/S100a10/Met/Cald1/Tpm1/Abl2/Rflna/Synpo/Cx3cl |
| BP | GO:001401 | negative regulation of gliogenesis | 12\|1209 | 49\|16097 | 0.00021 | 0.00291 | ENSMUS | Ldlr/Hmga2/Ski/Nr1d1/Nog/Dusp10/Notch1/Id4/Atf5/Ptn/Hes1 |
| BP | GO:003501 | somatic stem cell population maintenance | 12\|1209 | 49\|16097 | 0.00021 | 0.00291 | ENSMUS | Sox9/Hmga2/Klf10/Ski/Nog/Ldb2/Rest/Sfrp1/Hes1/Kat6a/Bcl9l/ |
| BP | GO:005092 | regulation of chemotaxis | 30\|1209 | 200\|16097 | 0.00021 | 0.00294 | ENSMUS | Thbs1/Serpine1/Fgfr1/Dusp1/Met/Smoc2/Zswim4/Cx3cl1/Thbs |
| BP | GO:000761 | memory | 23\|1209 | 137\|16097 | 0.00021 | 0.00299 | ENSMUS | Ldlr/Itga5/Egr1/Serpinf1/Rcan1/Itpr3/Sgk1/Ehmt2/S100b/Nog/ |
| BP | GO:005104 | negative regulation of secretion | 34\|1209 | 238\|16097 | 0.00021 | 0.00299 | ENSMUS | Hmox1/Hmgcr/Cyp51/Igfbp3/Cd34/Cx3cl1/Agtr2/C1qtnf3/Neo |
| CC | GO:004323 | receptor complex | 46\|1232 | 355\|16313 | 0.00022 | 0.00581 | ENSMUS | Ldlr/Tlr2/Itga5/Fgfr1/Cd14/Met/Il6ra/Fgfr2/Itpr3/Rxra/Gprc5a/ |
| BP | GO:000861 | lipid biosynthetic process | 58\|1209 | 481\|16097 | 0.00022 | 0.00312 | ENSMUS | Hmgcr/Ldlr/Sgms2/Acly/Cyp51/Egr1/Stard4/Lpl/Scd1/Fdft1/Ms |
| BP | GO:000743 | salivary gland morphogenesis | 10\|1209 | 36\|16097 | 0.00023 | 0.00318 | ENSMUS | Fgfr1/Met/Fgfr2/Egfr/Nrp1/Twsg1/Plxnd1/Tgm2/Dag1/Btbd7 |
| BP | GO:007048 | response to oxygen levels | 32\|1209 | 220\|16097 | 0.00023 | 0.00323 | ENSMUS | Hmox1/Myc/Vasn/Egr1/Eif4ebp1/Rtn4/Hyou1/Cd34/Fosl2/Fnd |
| MF | GO:000097 | RNA polymerase II proximal promoter sequence-specific DNA b | 59\|1190 | 495\|15912 | 0.00024 | 0.0081 | ENSMUS | Fosl1/Myc/Sox9/Klf4/Atf4/Jun/Mafg/Srebf2/Lrrfip1/Ddit3/Mafk |
| MF | GO:000504 | scavenger receptor activity | 11\|1190 | 43\|15912 | 0.00024 | 0.0081 | ENSMUS | Prg4/Scarb2/Enpp2/Cd36/Ssc5d/Ackr3/Scarf2/Ackr4/Loxl2/Enp |
| BP | GO:000910 | glycoprotein metabolic process | 41\|1209 | 308\|16097 | 0.00024 | 0.00335 | ENSMUS | Sulf2/Xylt1/Csgalnact2/Tet3/Cst3/Rpn1/Chpf/Pcsk6/Chst11/B4 |
| BP | GO:009010 | positive regulation of transmembrane receptor protein serine/ | 19\|1209 | 104\|16097 | 0.00024 | 0.00336 | ENSMUS | Thbs1/Inhba/Cdh5/Cyr61/Bmp6/Tgfbr3/Neo1/Tgfbr1/Furin/Tws |
| BP | GO:006053 | cartilage morphogenesis | 8\|1209 | 24\|16097 | 0.00024 | 0.00336 | ENSMUS | Col14a1/Col12a1/Col6a2/Matn3/Col6a3/Matn1/Col6a1/Vit |
| BP | GO:004338 | positive regulation of DNA binding | 13\|1209 | 57\|16097 | 0.00025 | 0.00337 | ENSMUS | Myc/Klf4/Hipk1/Jak2/Ski/Fam129b/Igf1/Plaur/Hipk3/Hipk2/Lga |
| BP | GO:005110 | regulation of DNA binding | 21\|1209 | 121\|16097 | 0.00025 | 0.00338 | ENSMUS | Hmox1/Myc/Klf4/Hipk1/Jun/Ddit3/Jak2/Ski/Fam129b/Cdt1/Igf |
| BP | GO:001061 | negative regulation of cardiac muscle hypertrophy | 9\|1209 | 30\|16097 | 0.00025 | 0.00338 | ENSMUS | Errfi1/Rgs2/Lmna/Tnfrsf1a/Ctdp1/Tomm70a/Pi16/Notch1/Foxc |
| BP | GO:004592 | negative regulation of exocytosis | 9\|1209 | 30\|16097 | 0.00025 | 0.00338 | ENSMUS | Hmox1/Rabgef1/Notch1/Gnai2/Vps4b/Lgals9/Adra2a/Rest/Cbar |
| BP | GO:190403 | negative regulation of epithelial cell apoptotic process | 11\|1209 | 43\|16097 | 0.00025 | 0.0034 | ENSMUS | Hmox1/Serpine1/Abl2/Gas6/Angptl4/Igf1r/Igf1/Tek/Nfe2l2/Kdr/ |
| BP | GO:190290 | regulation of supramolecular fiber organization | 44\|1209 | 339\|16097 | 0.00026 | 0.0035 | ENSMUS | Ldlr/Tlr2/Eln/S100a10/Met/Cdh5/Pecam1/Tpm1/Sgk1/Synpo/C |
| BP | GO:190353 | negative regulation of secretion by cell | 31\|1209 | 212\|16097 | 0.00026 | 0.00355 | ENSMUS | Hmox1/Hmgcr/Cyp51/Igfbp3/Cd34/Cx3cl1/Agtr2/C1qtnf3/Neo |
| BP | GO:003286 | cellular response to insulin stimulus | 28\|1209 | 184\|16097 | 0.00026 | 0.00355 | ENSMUS | Lpl/Ywhag/Eif4ebp1/Socs3/Sgk1/Trib3/Xbp1/Ndel1/Igf1r/Pik3r |
| BP | GO:000181 | negative regulation of cytokine production | 34\|1209 | 241\|16097 | 0.00027 | 0.00365 | ENSMUS | Errfi1/Prg4/Thbs1/Hmox1/Tlr2/Klf4/Fgfr1/Met/Sars/Cd34/Cx3cl |
| BP | GO:003096 | endoplasmic reticulum unfolded protein response | 14\|1209 | 65\|16097 | 0.00027 | 0.00365 | ENSMUS | Herpud1/Atf4/Eif2ak3/Ddit3/Xbp1/Ccnd1/Ero1l/Crebrf/Calr/De |
| BP | GO:001810 | peptidyl-serine phosphorylation | 38\|1209 | 280\|16097 | 0.00027 | 0.00365 | ENSMUS | Spry2/Fnip2/Hipk1/Met/Mast4/Eif2ak3/Sgk1/Egfr/Gas6/Sfrp2/H |
| BP | GO:002198 | cerebral cortex development | 19\|1209 | 105\|16097 | 0.00028 | 0.0037 | ENSMUS | Col3a1/Cdon/Lamb1/Fgfr1/Pex5/Rtn4/Dixdc1/Adgrg1/Egfr/Lrp |
| BP | GO:005501 | cardiac muscle cell development | 18\|1209 | 97\|16097 | 0.00029 | 0.00383 | ENSMUS | Col14a1/Rgs2/Met/Sorbs2/Lmna/Maml1/Myh11/Mef2a/Agtr2/ |
| BP | GO:003085 | positive regulation of epithelial cell differentiation | 13\|1209 | 58\|16097 | 0.0003 | 0.00392 | ENSMUS | Sox9/Serpine1/Cdh5/Ncoa3/Bmp6/Notch1/Cldn5/Sfrp4/Sfn/Ak |
| BP | GO:007020 | protein heterotrimerization | 7\|1209 | 19\|16097 | 0.0003 | 0.00395 | ENSMUS | Col1a1/Col1a2/Col6a2/Ppp2ca/C1qtnf6/Nup62/Col6a1 |

| Count | Up | Up_Gene_id | Down | Down_Gene_id |
|---|---|---|---|---|
| 13 | 6 | ENSMUSG0 | 7 | ENSMUSG00000040152/ENSMUSG00000037411/ENSMUSG00000008730/ENSMUSG00000026596/ENSMUSG00000020484/ENSMUSG00000031451/ENSMUSG0 |
| 20 | 6 | ENSMUSG0 | 14 | ENSMUSG00000009376/ENSMUSG00000027954/ENSMUSG00000026259/ENSMUSG00000019970/ENSMUSG00000002489/ENSMUSG00000028613/ENSMUSG0 |
| 43 | 6 | ENSMUSG0 | 37 | ENSMUSG00000022346/ENSMUSG00000020917/ENSMUSG00000024378/ENSMUSG00000022367/ENSMUSG00000037071/ENSMUSG00000003541/ENSMUSG0 |
| 36 | 15 | ENSMUSG0 | 21 | ENSMUSG00000032193/ENSMUSG00000027995/ENSMUSG00000000567/ENSMUSG00000003032/ENSMUSG00000038418/ENSMUSG00000041959/ENSMUSG0 |
| 17 | 6 | ENSMUSG0 | 11 | ENSMUSG00000031565/ENSMUSG00000030849/ENSMUSG00000017929/ENSMUSG00000020423/ENSMUSG00000018736/ENSMUSG00000035764/ENSMUSG0 |
| 29 | 6 | ENSMUSG0 | 23 | ENSMUSG00000028967/ENSMUSG00000022114/ENSMUSG00000021670/ENSMUSG00000026360/ENSMUSG00000032501/ENSMUSG00000036390/ENSMUSG0 |
| 43 | 6 | ENSMUSG0 | 37 | ENSMUSG00000022346/ENSMUSG00000020917/ENSMUSG00000024378/ENSMUSG00000022367/ENSMUSG00000037071/ENSMUSG00000003541/ENSMUSG0 |
| 41 | 15 | ENSMUSG0 | 26 | ENSMUSG00000021670/ENSMUSG00000032193/ENSMUSG00000027995/ENSMUSG00000000555/ENSMUSG00000038418/ENSMUSG00000000753/ENSMUSG0 |
| 30 | 17 | ENSMUSG0 | 13 | ENSMUSG00000028645/ENSMUSG00000069833/ENSMUSG00000028525/ENSMUSG00000096054/ENSMUSG00000054808/ENSMUSG00000032643/ENSMUSG0 |
| 13 | 8 | ENSMUSG0 | 5 | ENSMUSG00000006800/ENSMUSG00000038418/ENSMUSG00000027947/ENSMUSG00000048616/ENSMUSG00000032220 |
| 41 | 8 | ENSMUSG0 | 33 | ENSMUSG00000022114/ENSMUSG00000061175/ENSMUSG00000008730/ENSMUSG00000009376/ENSMUSG00000034751/ENSMUSG00000031668/ENSMUSG0 |
| 23 | 11 | ENSMUSG0 | 12 | ENSMUSG00000040152/ENSMUSG00000027995/ENSMUSG00000037411/ENSMUSG00000050335/ENSMUSG00000020458/ENSMUSG00000031778/ENSMUSG0 |
| 10 | 6 | ENSMUSG0 | 4 | ENSMUSG00000022114/ENSMUSG00000031714/ENSMUSG00000023951/ENSMUSG00000023068 |
| 29 | 9 | ENSMUSG0 | 20 | ENSMUSG00000026360/ENSMUSG00000041324/ENSMUSG00000001627/ENSMUSG00000020458/ENSMUSG00000023067/ENSMUSG00000049800/ENSMUSG0 |
| 26 | 9 | ENSMUSG0 | 17 | ENSMUSG00000028967/ENSMUSG00000040152/ENSMUSG00000027995/ENSMUSG00000051439/ENSMUSG00000074582/ENSMUSG00000031778/ENSMUSG0 |
| 54 | 21 | ENSMUSG0 | 33 | ENSMUSG00000000567/ENSMUSG00000040856/ENSMUSG00000038418/ENSMUSG00000009376/ENSMUSG00000037071/ENSMUSG00000042406/ENSMUSG0 |
| 16 | 8 | ENSMUSG0 | 8 | ENSMUSG00000000567/ENSMUSG00000022265/ENSMUSG00000039004/ENSMUSG00000031849/ENSMUSG00000056050/ENSMUSG00000026836/ENSMUSG0 |
| 6 | 1 | ENSMUSG0 | 5 | ENSMUSG00000032244/ENSMUSG00000030849/ENSMUSG00000015846/ENSMUSG00000005087/ENSMUSG00000031548 |
| 6 | 1 | ENSMUSG0 | 5 | ENSMUSG00000038418/ENSMUSG00000023951/ENSMUSG00000036450/ENSMUSG00000015839/ENSMUSG00000025612 |
| 28 | 7 | ENSMUSG0 | 21 | ENSMUSG00000042688/ENSMUSG00000009376/ENSMUSG00000027954/ENSMUSG00000030536/ENSMUSG00000026259/ENSMUSG00000019970/ENSMUSG0 |
| 7 | 2 | ENSMUSG0 | 5 | ENSMUSG00000033161/ENSMUSG00000039004/ENSMUSG00000005533/ENSMUSG00000029249/ENSMUSG00000024431 |
| 26 | 12 | ENSMUSG0 | 14 | ENSMUSG00000029675/ENSMUSG00000041959/ENSMUSG00000009376/ENSMUSG00000026596/ENSMUSG00000031778/ENSMUSG00000019997/ENSMUSG0 |
| 12 | 3 | ENSMUSG0 | 9 | ENSMUSG00000032193/ENSMUSG00000056758/ENSMUSG00000029050/ENSMUSG00000020889/ENSMUSG00000048616/ENSMUSG00000039384/ENSMUSG0 |
| 12 | 2 | ENSMUSG0 | 10 | ENSMUSG00000000567/ENSMUSG00000056758/ENSMUSG00000037465/ENSMUSG00000029050/ENSMUSG00000048616/ENSMUSG00000029249/ENSMUSG0 |
| 30 | 15 | ENSMUSG0 | 15 | ENSMUSG00000040152/ENSMUSG00000037411/ENSMUSG00000031565/ENSMUSG00000009376/ENSMUSG00000035671/ENSMUSG00000031778/ENSMUSG0 |
| 23 | 7 | ENSMUSG0 | 16 | ENSMUSG00000032193/ENSMUSG00000000555/ENSMUSG00000038418/ENSMUSG00000000753/ENSMUSG00000022951/ENSMUSG00000042644/ENSMUSG0 |
| 34 | 14 | ENSMUSG0 | 20 | ENSMUSG00000005413/ENSMUSG00000021670/ENSMUSG00000001467/ENSMUSG00000031778/ENSMUSG00000032340/ENSMUSG00000031451/ENSMUSG0 |
| 46 | 17 | ENSMUSG0 | 29 | ENSMUSG00000032193/ENSMUSG00000027995/ENSMUSG00000000555/ENSMUSG00000031565/ENSMUSG00000051439/ENSMUSG00000009376/ENSMUSG0 |
| 58 | 12 | ENSMUSG0 | 46 | ENSMUSG00000021670/ENSMUSG00000032193/ENSMUSG00000050931/ENSMUSG00000020917/ENSMUSG00000001467/ENSMUSG00000038418/ENSMUSG0 |
| 10 | 3 | ENSMUSG0 | 7 | ENSMUSG00000031565/ENSMUSG00000009376/ENSMUSG00000030849/ENSMUSG00000024098/ENSMUSG00000037820/ENSMUSG00000039952/ENSMUSG0 |
| 32 | 10 | ENSMUSG0 | 22 | ENSMUSG00000005413/ENSMUSG00000022346/ENSMUSG00000039646/ENSMUSG00000038418/ENSMUSG00000031490/ENSMUSG00000020458/ENSMUSG0 |
| 59 | 15 | ENSMUSG0 | 44 | ENSMUSG00000024912/ENSMUSG00000022346/ENSMUSG00000000567/ENSMUSG00000003032/ENSMUSG00000042406/ENSMUSG00000051510/ENSMUSG0 |
| 11 | 5 | ENSMUSG0 | 6 | ENSMUSG00000006014/ENSMUSG00000029426/ENSMUSG00000022425/ENSMUSG00000044337/ENSMUSG00000034205/ENSMUSG00000025185 |
| 41 | 9 | ENSMUSG0 | 32 | ENSMUSG00000006800/ENSMUSG00000030657/ENSMUSG00000042042/ENSMUSG00000034832/ENSMUSG00000027447/ENSMUSG00000030062/ENSMUSG0 |
| 19 | 8 | ENSMUSG0 | 11 | ENSMUSG00000040152/ENSMUSG00000041324/ENSMUSG00000039004/ENSMUSG00000029287/ENSMUSG00000032340/ENSMUSG00000007613/ENSMUSG0 |
| 8 | 5 | ENSMUSG0 | 3 | ENSMUSG00000020241/ENSMUSG00000048126/ENSMUSG00000001119 |
| 13 | 4 | ENSMUSG0 | 9 | ENSMUSG00000022346/ENSMUSG00000003032/ENSMUSG00000008730/ENSMUSG00000024789/ENSMUSG00000029050/ENSMUSG00000026796/ENSMUSG0 |
| 21 | 6 | ENSMUSG0 | 15 | ENSMUSG00000005413/ENSMUSG00000022346/ENSMUSG00000003032/ENSMUSG00000008730/ENSMUSG00000025408/ENSMUSG00000024789/ENSMUSG0 |
| 9 | 2 | ENSMUSG0 | 7 | ENSMUSG00000028967/ENSMUSG00000026360/ENSMUSG00000028063/ENSMUSG00000030341/ENSMUSG00000033323/ENSMUSG00000022752/ENSMUSG0 |
| 9 | 3 | ENSMUSG0 | 6 | ENSMUSG00000005413/ENSMUSG00000025340/ENSMUSG00000009907/ENSMUSG00000033717/ENSMUSG00000029249/ENSMUSG00000035640 |
| 11 | 5 | ENSMUSG0 | 6 | ENSMUSG00000005413/ENSMUSG00000037411/ENSMUSG00000026596/ENSMUSG00000031451/ENSMUSG00000005533/ENSMUSG00000015839 |
| 44 | 20 | ENSMUSG0 | 24 | ENSMUSG00000032193/ENSMUSG00000027995/ENSMUSG00000029675/ENSMUSG00000041959/ENSMUSG00000009376/ENSMUSG00000019970/ENSMUSG0 |
| 31 | 11 | ENSMUSG0 | 20 | ENSMUSG00000005413/ENSMUSG00000021670/ENSMUSG00000001467/ENSMUSG00000031778/ENSMUSG00000032340/ENSMUSG00000031451/ENSMUSG0 |
| 28 | 6 | ENSMUSG0 | 22 | ENSMUSG00000051391/ENSMUSG00000031490/ENSMUSG00000019970/ENSMUSG00000032715/ENSMUSG00000020484/ENSMUSG00000018736/ENSMUSG0 |
| 34 | 9 | ENSMUSG0 | 25 | ENSMUSG00000028967/ENSMUSG00000006014/ENSMUSG00000040152/ENSMUSG00000005413/ENSMUSG00000027995/ENSMUSG00000003032/ENSMUSG0 |
| 14 | 1 | ENSMUSG0 | 13 | ENSMUSG00000031770/ENSMUSG00000042406/ENSMUSG00000031668/ENSMUSG00000025408/ENSMUSG00000020484/ENSMUSG00000021831/ENSMUSG0 |
| 38 | 5 | ENSMUSG0 | 33 | ENSMUSG00000022114/ENSMUSG00000061175/ENSMUSG00000008730/ENSMUSG00000009376/ENSMUSG00000034751/ENSMUSG00000031668/ENSMUSG0 |
| 19 | 6 | ENSMUSG0 | 13 | ENSMUSG00000038119/ENSMUSG00000031565/ENSMUSG00000005069/ENSMUSG00000020458/ENSMUSG00000032064/ENSMUSG00000031785/ENSMUSG0 |
| 18 | 9 | ENSMUSG0 | 9 | ENSMUSG00000026360/ENSMUSG00000009376/ENSMUSG00000028063/ENSMUSG00000050567/ENSMUSG00000030557/ENSMUSG00000033323/ENSMUSG0 |
| 13 | 6 | ENSMUSG0 | 7 | ENSMUSG00000000567/ENSMUSG00000037411/ENSMUSG00000027678/ENSMUSG00000039004/ENSMUSG00000047281/ENSMUSG00000022016/ENSMUSG0 |
| 7 | 3 | ENSMUSG0 | 4 | ENSMUSG00000020241/ENSMUSG00000020349/ENSMUSG00000109511/ENSMUSG00000001119 |

0000015839
0000020745/ENSMUSG00000096054/ENSMUSG00000028161/ENSMUSG00000062232/ENSMUSG00000020132/ENSMUSG00000018965/ENSMUSG00000025326/ENSMUSG00000025372
0000032883/ENSMUSG00000025203/ENSMUSG00000045294/ENSMUSG00000024640/ENSMUSG00000004040/ENSMUSG00000025153/ENSMUSG00000032715/ENSMUSG00000029752/ENSMUSG0000003
0000037071/ENSMUSG00000029482/ENSMUSG00000025203/ENSMUSG00000026596/ENSMUSG00000015846/ENSMUSG00000028893/ENSMUSG00000029287/ENSMUSG00000020484/ENSMUSG0000000
0000020745/ENSMUSG00000050965/ENSMUSG00000062232/ENSMUSG00000038539/ENSMUSG00000026826
0000023067/ENSMUSG00000027351/ENSMUSG00000046733/ENSMUSG00000032715/ENSMUSG00000049323/ENSMUSG00000034765/ENSMUSG00000027996/ENSMUSG00000005533/ENSMUSG0000003
0000032883/ENSMUSG00000025203/ENSMUSG00000045294/ENSMUSG00000024640/ENSMUSG00000004040/ENSMUSG00000025153/ENSMUSG00000032715/ENSMUSG00000029752/ENSMUSG0000003
0000022951/ENSMUSG00000026596/ENSMUSG00000042644/ENSMUSG00000019970/ENSMUSG00000020423/ENSMUSG00000048616/ENSMUSG00000020142/ENSMUSG00000020745/ENSMUSG0000000
0000028161/ENSMUSG00000026313/ENSMUSG00000026131/ENSMUSG00000039952/ENSMUSG00000029361/ENSMUSG00000027712/ENSMUSG00000031543

0000019970/ENSMUSG00000031451/ENSMUSG00000027996/ENSMUSG00000037012/ENSMUSG00000007613/ENSMUSG00000045983/ENSMUSG00000033955/ENSMUSG00000005087/ENSMUSG0000002
0000031451/ENSMUSG00000021508/ENSMUSG00000023951/ENSMUSG00000003814/ENSMUSG00000056050/ENSMUSG00000016024

0000028893/ENSMUSG00000028641/ENSMUSG00000022508/ENSMUSG00000027996/ENSMUSG00000033323/ENSMUSG00000022752/ENSMUSG00000030168/ENSMUSG00000095098/ENSMUSG0000003
0000024789/ENSMUSG00000031451/ENSMUSG00000030341/ENSMUSG00000037447/ENSMUSG00000022708/ENSMUSG00000022037/ENSMUSG00000016024/ENSMUSG00000029816/ENSMUSG0000002
0000025408/ENSMUSG00000039004/ENSMUSG00000004040/ENSMUSG00000019997/ENSMUSG00000024789/ENSMUSG00000074794/ENSMUSG00000035569/ENSMUSG00000023845/ENSMUSG0000002
0000019943/ENSMUSG00000074934

0000002489/ENSMUSG00000028613/ENSMUSG00000020745/ENSMUSG00000096054/ENSMUSG00000028161/ENSMUSG00000024924/ENSMUSG00000062232/ENSMUSG00000020132/ENSMUSG0000001

0000031523/ENSMUSG00000007613/ENSMUSG00000022817/ENSMUSG00000054808/ENSMUSG00000024533/ENSMUSG00000038859/ENSMUSG00000025372/ENSMUSG00000026812
0000021379/ENSMUSG00000038539/ENSMUSG00000031393
0000031548/ENSMUSG00000031540/ENSMUSG00000063382/ENSMUSG00000005871
0000002489/ENSMUSG00000031451/ENSMUSG00000021508/ENSMUSG00000023951/ENSMUSG00000020682/ENSMUSG00000050965/ENSMUSG00000003814/ENSMUSG00000016024/ENSMUSG0000007
0000019970/ENSMUSG00000048616/ENSMUSG00000005442/ENSMUSG00000018634/ENSMUSG00000069895/ENSMUSG00000032487/ENSMUSG00000000184/ENSMUSG00000020091/ENSMUSG0000003
0000030341/ENSMUSG00000037434/ENSMUSG00000025340/ENSMUSG00000028161/ENSMUSG00000018634/ENSMUSG00000009907/ENSMUSG00000033717/ENSMUSG00000029249/ENSMUSG0000002
0000027947/ENSMUSG00000030849/ENSMUSG00000042644/ENSMUSG00000015846/ENSMUSG00000046733/ENSMUSG00000029287/ENSMUSG00000028613/ENSMUSG00000021756/ENSMUSG0000003
0000024378/ENSMUSG00000037071/ENSMUSG00000021273/ENSMUSG00000031604/ENSMUSG00000033161/ENSMUSG00000032883/ENSMUSG00000093930/ENSMUSG00000022351/ENSMUSG0000002
0000041702
0000032115/ENSMUSG00000029135/ENSMUSG00000023951/ENSMUSG00000063142/ENSMUSG00000024924/ENSMUSG00000018634/ENSMUSG00000036450/ENSMUSG00000029249/ENSMUSG0000004
0000022463/ENSMUSG00000026305/ENSMUSG00000025408/ENSMUSG00000018143/ENSMUSG00000021540/ENSMUSG00000068739/ENSMUSG00000015846/ENSMUSG00000000078/ENSMUSG0000000

0000032997/ENSMUSG00000030513/ENSMUSG00000017929/ENSMUSG00000024413/ENSMUSG00000036356/ENSMUSG00000043587/ENSMUSG00000028521/ENSMUSG00000004846/ENSMUSG0000004
0000030530/ENSMUSG00000024098/ENSMUSG00000061436/ENSMUSG00000026836/ENSMUSG00000024182

0000046223/ENSMUSG00000027177/ENSMUSG00000061436
0000029050/ENSMUSG00000026796/ENSMUSG00000006585/ENSMUSG00000046223/ENSMUSG00000027177/ENSMUSG00000061436/ENSMUSG00000032717/ENSMUSG00000021379/ENSMUSG0000002
0000044167

0000031778/ENSMUSG00000019997/ENSMUSG00000031523/ENSMUSG00000007613/ENSMUSG00000040021/ENSMUSG00000024533/ENSMUSG00000040852/ENSMUSG00000032702/ENSMUSG0000002
0000030341/ENSMUSG00000037434/ENSMUSG00000025340/ENSMUSG00000028161/ENSMUSG00000018634/ENSMUSG00000009907/ENSMUSG00000033717/ENSMUSG00000029249/ENSMUSG0000002
0000005533/ENSMUSG00000024927/ENSMUSG00000028042/ENSMUSG00000050965/ENSMUSG00000032702/ENSMUSG00000000058/ENSMUSG00000038859/ENSMUSG00000024052/ENSMUSG0000004
0000031565/ENSMUSG00000009376/ENSMUSG00000068739/ENSMUSG00000031778/ENSMUSG00000022508/ENSMUSG00000020601/ENSMUSG00000031451/ENSMUSG00000030341/ENSMUSG0000003
0000048249/ENSMUSG00000003814/ENSMUSG00000022365/ENSMUSG00000027230/ENSMUSG00000020571/ENSMUSG00000015839/ENSMUSG00000026628
0000019970/ENSMUSG00000031451/ENSMUSG00000027996/ENSMUSG00000037012/ENSMUSG00000007613/ENSMUSG00000045983/ENSMUSG00000033955/ENSMUSG00000005087/ENSMUSG0000002
0000028613/ENSMUSG00000018736/ENSMUSG00000035764/ENSMUSG00000030852/ENSMUSG00000020745/ENSMUSG00000065954/ENSMUSG00000026812
0000023951/ENSMUSG00000022752/ENSMUSG00000024431
0000005871

37012/ENSMUSG00000020889/ENSMUSG00000030268/ENSMUSG00000004846/ENSMUSG00000022708/ENSMUSG00000003665/ENSMUSG00000039202/ENSMUSG00000029201/ENSMUSG00000004267/E
05533/ENSMUSG00000056501/ENSMUSG00000023951/ENSMUSG00000030747/ENSMUSG00000001119/ENSMUSG00000028646/ENSMUSG00000050697

39384/ENSMUSG00000040021/ENSMUSG00000027177/ENSMUSG00000042594/ENSMUSG00000109511/ENSMUSG00000050965/ENSMUSG00000053716/ENSMUSG00000031548/ENSMUSG00000005871
37012/ENSMUSG00000020889/ENSMUSG00000030268/ENSMUSG00000004846/ENSMUSG00000022708/ENSMUSG00000003665/ENSMUSG00000039202/ENSMUSG00000029201/ENSMUSG00000004267/E
05442/ENSMUSG00000059991/ENSMUSG00000050965/ENSMUSG00000018634/ENSMUSG00000036564/ENSMUSG00000079037/ENSMUSG00000069895/ENSMUSG00000032487/ENSMUSG00000000184/E

23951/ENSMUSG00000040021/ENSMUSG00000030655/ENSMUSG00000027177/ENSMUSG00000061436/ENSMUSG00000036707/ENSMUSG00000021070/ENSMUSG00000050965/ENSMUSG00000003810/E

34201/ENSMUSG00000028179/ENSMUSG00000031548/ENSMUSG00000031393/ENSMUSG00000074934/ENSMUSG00000002083
27551/ENSMUSG00000028277/ENSMUSG00000037902
20889/ENSMUSG00000045983/ENSMUSG00000005533/ENSMUSG00000056501/ENSMUSG00000023951/ENSMUSG00000000134/ENSMUSG00000053797/ENSMUSG00000018476/ENSMUSG00000047281/E

18965/ENSMUSG00000041570/ENSMUSG00000044147/ENSMUSG00000024431/ENSMUSG00000031393/ENSMUSG00000025326/ENSMUSG00000025372

74934
31393/ENSMUSG00000074923
29361/ENSMUSG00000031548/ENSMUSG00000035828/ENSMUSG00000035621/ENSMUSG00000027712/ENSMUSG00000035640
30341/ENSMUSG00000007613/ENSMUSG00000005533/ENSMUSG00000054027/ENSMUSG00000005087/ENSMUSG00000022817/ENSMUSG00000036606/ENSMUSG00000006445/ENSMUSG00000039462/E
25203/ENSMUSG00000045294/ENSMUSG00000039004/ENSMUSG00000026675/ENSMUSG00000025153/ENSMUSG00000017929/ENSMUSG00000034926/ENSMUSG00000032715/ENSMUSG00000020889/E

44167/ENSMUSG00000034205/ENSMUSG00000015839/ENSMUSG00000025612/ENSMUSG00000031393/ENSMUSG00000030287/ENSMUSG00000026826/ENSMUSG00000026812
04040/ENSMUSG00000040463/ENSMUSG00000056216/ENSMUSG00000052942/ENSMUSG00000001419/ENSMUSG00000030557/ENSMUSG00000037465/ENSMUSG00000020484/ENSMUSG00000023034/E

45216/ENSMUSG00000020205/ENSMUSG00000023068/ENSMUSG00000024085/ENSMUSG00000021978/ENSMUSG00000038296/ENSMUSG00000027832/ENSMUSG00000029201/ENSMUSG00000020346/E

29249

22037/ENSMUSG00000041570/ENSMUSG00000034201/ENSMUSG00000038859/ENSMUSG00000044147/ENSMUSG00000031393/ENSMUSG00000025372/ENSMUSG00000026812/ENSMUSG00000002365/E
29361/ENSMUSG00000031548/ENSMUSG00000035828/ENSMUSG00000035621/ENSMUSG00000027712/ENSMUSG00000035640
44167/ENSMUSG00000020091/ENSMUSG00000042903/ENSMUSG00000019943/ENSMUSG00000050697/ENSMUSG00000026812/ENSMUSG00000005871/ENSMUSG00000034593
30530/ENSMUSG00000025340/ENSMUSG00000024098/ENSMUSG00000006445/ENSMUSG00000079037/ENSMUSG00000016024/ENSMUSG00000029816/ENSMUSG00000019943/ENSMUSG00000024425/E

23951/ENSMUSG00000040021/ENSMUSG00000030655/ENSMUSG00000027177/ENSMUSG00000061436/ENSMUSG00000036707/ENSMUSG00000021070/ENSMUSG00000050965/ENSMUSG00000003810/E

ENSMUSG00000032349/ENSMUSG00000053398/ENSMUSG00000026313/ENSMUSG00000029009/ENSMUSG00000038633/ENSMUSG00000032487/ENSMUSG00000047126/ENSMUSG00000028179/ENSMUS

ENSMUSG00000032349/ENSMUSG00000053398/ENSMUSG00000026313/ENSMUSG00000029009/ENSMUSG00000038633/ENSMUSG00000032487/ENSMUSG00000047126/ENSMUSG00000028179/ENSMUS
ENSMUSG00000020091/ENSMUSG00000031393/ENSMUSG00000074923

ENSMUSG00000073563/ENSMUSG00000001630/ENSMUSG00000015202/ENSMUSG00000032487/ENSMUSG00000027665/ENSMUSG00000035992/ENSMUSG00000029361/ENSMUSG00000050697/ENSMUS

ENSMUSG00000028042/ENSMUSG00000050965/ENSMUSG00000030168/ENSMUSG00000030527/ENSMUSG00000032487/ENSMUSG00000027665/ENSMUSG00000018707/ENSMUSG00000026147/ENSMUS

ENSMUSG00000024924/ENSMUSG00000033717/ENSMUSG00000026836/ENSMUSG00000030287/ENSMUSG00000025185/ENSMUSG00000028661
ENSMUSG00000005533/ENSMUSG00000030275/ENSMUSG00000022708/ENSMUSG00000023068/ENSMUSG00000039202/ENSMUSG00000021770/ENSMUSG00000032018/ENSMUSG00000058258/ENSMUS

ENSMUSG00000029135/ENSMUSG00000030103/ENSMUSG00000020889/ENSMUSG00000056501/ENSMUSG00000052040/ENSMUSG00000024927/ENSMUSG00000048490/ENSMUSG00000000134/ENSMUS

ENSMUSG00000043019/ENSMUSG00000024486/ENSMUSG00000003746/ENSMUSG00000039497/ENSMUSG00000048920/ENSMUSG00000028277/ENSMUSG00000039357/ENSMUSG00000024807/ENSMUS

ENSMUSG00000005871

ENSMUSG00000037902/ENSMUSG00000031438

ENSMUSG00000073563/ENSMUSG00000001630/ENSMUSG00000015202/ENSMUSG00000032487/ENSMUSG00000027665/ENSMUSG00000035992/ENSMUSG00000029361/ENSMUSG00000050697/ENSMUS

G00000039497/ENSMUSG00000063229/ENSMUSG00000026623/ENSMUSG00000021794/ENSMUSG00000050697/ENSMUSG00000034593

G00000039497/ENSMUSG00000063229/ENSMUSG00000026623/ENSMUSG00000021794/ENSMUSG00000050697/ENSMUSG00000034593

G00000003345/ENSMUSG00000024182

G00000050697/ENSMUSG00000005871

G00000030747/ENSMUSG00000032349/ENSMUSG00000033105/ENSMUSG00000021036/ENSMUSG00000038633/ENSMUSG00000032487/ENSMUSG00000027665/ENSMUSG00000024052/ENSMUSG0000

G00000052056/ENSMUSG00000018476/ENSMUSG00000028042/ENSMUSG00000026313/ENSMUSG00000008398/ENSMUSG00000027230/ENSMUSG00000029249/ENSMUSG00000078671/ENSMUSG0000

G00000034951

G00000003345/ENSMUSG00000024182

0075703/ENSMUSG00000029249/ENSMUSG00000005615/ENSMUSG00000026623/ENSMUSG00000024431/ENSMUSG00000050697/ENSMUSG00000034593

0035397/ENSMUSG00000024431/ENSMUSG00000026826/ENSMUSG00000005886/ENSMUSG00000026628

| Category | GOID | Description | GeneRatio | BgRatio | pvalue | padj | geneID | geneName |
|---|---|---|---|---|---|---|---|---|
| BP | GO:006054 | respiratory system development | 32\|1209 | 223\|16097 | 0.0003 | 0.00395 | ENSMUS | Errfi1/Spry2/Igfbp5/Sox9/Stk40/Fgfr1/Mmp14/Adamts2/Fgfr2/ |
| BP | GO:006161 | pri-miRNA transcription by RNA polymerase II | 12\|1209 | 51\|16097 | 0.00031 | 0.00407 | ENSMUS | Fosl1/Sox9/Klf4/Jun/Srebf2/Fos/Stat3/Tgfbr1/Rela/Hdac4/Ddx1 |
| BP | GO:200035 | regulation of endothelial cell apoptotic process | 12\|1209 | 51\|16097 | 0.00031 | 0.00407 | ENSMUS | Thbs1/Serpine1/Col18a1/Abl2/Xbp1/Gas6/Angptl4/Cd248/Tek/ |
| BP | GO:006041 | ventricular septum morphogenesis | 11\|1209 | 44\|16097 | 0.00031 | 0.00407 | ENSMUS | Fgfr2/Tgfbr3/Nog/Tgfbr1/Robo1/Nfatc1/Notch1/Robo2/Acvr1/ |
| BP | GO:003286 | response to insulin | 31\|1209 | 214\|16097 | 0.00031 | 0.00407 | ENSMUS | Egr1/Lpl/Ywhag/Eif4ebp1/Socs3/Sesn2/Sgk1/Trib3/Xbp1/Ndel1, |
| BP | GO:006074 | epithelial cell differentiation involved in prostate gland develo | 6\|1209 | 14\|16097 | 0.00031 | 0.00407 | ENSMUS | Fem1b/Fgfr2/Rxra/Plaur/Notch1/Psap |
| BP | GO:006097 | vasculogenesis involved in coronary vascular morphogenesis | 6\|1209 | 14\|16097 | 0.00031 | 0.00407 | ENSMUS | Fgfr1/Spred1/Fgfr2/Tgfbr3/Nrp1/Notch1 |
| BP | GO:009011 | cargo loading into COPII-coated vesicle | 6\|1209 | 14\|16097 | 0.00031 | 0.00407 | ENSMUS | Insig1/Sec24d/Sec23a/Tbc1d20/Mia3/Sec24a |
| BP | GO:190401 | epithelial cell apoptotic process | 19\|1209 | 106\|16097 | 0.00031 | 0.00407 | ENSMUS | Thbs1/Hmox1/Serpine1/Hipk1/Col18a1/Abl2/Xbp1/Jak2/Gas6/ |
| BP | GO:000301 | vascular process in circulatory system | 28\|1209 | 186\|16097 | 0.00032 | 0.00408 | ENSMUS | Hmgcr/Rgs2/Cdh5/Akap12/Bmp6/Cx3cl1/Tgfbr3/Egfr/Agtr2/Du |
| BP | GO:004340 | negative regulation of MAP kinase activity | 14\|1209 | 66\|16097 | 0.00032 | 0.00415 | ENSMUS | Spry2/Hmgcr/Rgs2/Dusp1/Spred1/Dusp5/Sfrp2/Dusp10/Hipk3/ |
| BP | GO:005068 | negative regulation of epithelial cell proliferation | 22\|1209 | 132\|16097 | 0.00032 | 0.00415 | ENSMUS | Thbs1/Sox9/Dlk1/Tnmd/Serpinf1/Cd109/Fgfr2/Tgfbr3/Klf9/Mag |
| BP | GO:004632 | regulation of JNK cascade | 27\|1209 | 177\|16097 | 0.00032 | 0.00415 | ENSMUS | Tnxb/Gadd45a/Dixdc1/Nrk/Egfr/Tiam1/Ctgf/Gab1/Sfrp2/Igf1r/F |
| MF | GO:003802 | cargo receptor activity | 16\|1190 | 82\|15912 | 0.00033 | 0.01055 | ENSMUS | Prg4/Ldlr/Scarb2/Lrp8/Enpp2/Cd36/Ssc5d/Ackr3/Scarf2/Vldlr/ |
| BP | GO:001580 | neutral amino acid transport | 9\|1209 | 31\|16097 | 0.00033 | 0.00418 | ENSMUS | Rgs2/Slc3a2/Nfe2l1/Slc38a2/Slc6a17/Slc6a6/Slc38a3/Slc1a4/Sl |
| BP | GO:005077 | positive regulation of axonogenesis | 18\|1209 | 98\|16097 | 0.00033 | 0.00419 | ENSMUS | Tnfrsf12a/Rtn4/Golga4/Dixdc1/Tiam1/Tubb2b/Ndel1/Nrp1/Cxc |
| BP | GO:199084 | adaptive thermogenesis | 24\|1209 | 150\|16097 | 0.00033 | 0.00423 | ENSMUS | Scd1/Atf4/Ddit3/Jak2/Arrdc3/Fabp4/Lnpep/Nr1d1/Cd36/Igf1r/ |
| BP | GO:009719 | extrinsic apoptotic signaling pathway | 31\|1209 | 215\|16097 | 0.00034 | 0.0043 | ENSMUS | Thbs1/Hmox1/Serpine1/Hipk1/Inhba/Fgfr1/Fem1b/Tnfrsf12a/L |
| BP | GO:003243 | positive regulation of proteasomal ubiquitin-dependent protei | 16\|1209 | 82\|16097 | 0.00034 | 0.00433 | ENSMUS | Hspa1b/Hspa1a/Trib1/Herpud1/Zfand2a/Trib3/Trib2/Nkd2/Rnf |
| BP | GO:000193 | regulation of endothelial cell proliferation | 21\|1209 | 124\|16097 | 0.00035 | 0.00443 | ENSMUS | F3/Thbs1/Hmox1/Dlk1/Tnmd/Fgfr1/Jun/Bmp6/Stat3/Thbs4/Nr |
| BP | GO:003051 | negative regulation of transforming growth factor beta recepto | 13\|1209 | 59\|16097 | 0.00035 | 0.00445 | ENSMUS | Vasn/Cd109/Aspn/Tgfbr3/Chst11/Xbp1/Ski/Pmepa1/Ltbp1/Eid2 |
| BP | GO:190161 | organic hydroxy compound metabolic process | 51\|1209 | 416\|16097 | 0.00036 | 0.00447 | ENSMUS | Aoc2/Hmgcr/Ldlr/Moxd1/Cyp51/Fgfr1/Stard4/Fdft1/Msmo1/N |
| BP | GO:007200 | nephron development | 22\|1209 | 133\|16097 | 0.00036 | 0.00453 | ENSMUS | Sulf2/Myc/Sox9/Nid1/Egr1/Il6ra/Lzts2/Agtr2/Notch3/Maged1/ |
| BP | GO:002154 | pallium development | 23\|1209 | 142\|16097 | 0.00037 | 0.00456 | ENSMUS | Col3a1/Cdon/Lamb1/Fgfr1/Pex5/Rtn4/Dixdc1/Adgrg1/Egfr/Lrp |
| BP | GO:004255 | myelination | 23\|1209 | 142\|16097 | 0.00037 | 0.00456 | ENSMUS | Tlr2/Eif2ak3/Mpz/Rxra/Nab2/Atrn/B4galt5/Nab1/Ski/S100b/Nd |
| BP | GO:005101 | actin filament bundle assembly | 25\|1209 | 160\|16097 | 0.00037 | 0.00456 | ENSMUS | Eln/Marcks/S100a10/Met/Cald1/Tpm1/Abl2/Synpo/Cx3cl1/Ctg |
| BP | GO:003367 | negative regulation of kinase activity | 30\|1209 | 207\|16097 | 0.00038 | 0.00477 | ENSMUS | Errfi1/Spry2/Hmgcr/Rgs2/Trib1/Gadd45a/Dusp1/Cdkn1a/Spred |
| BP | GO:003195 | regulation of protein autophosphorylation | 11\|1209 | 45\|16097 | 0.00039 | 0.00478 | ENSMUS | Errfi1/Jun/Iqgap1/Ppp2ca/Tnks1bp1/Vegfa/Impact/Gpnmb/Ace |
| BP | GO:190370 | regulation of hemopoiesis | 47\|1209 | 376\|16097 | 0.00039 | 0.00479 | ENSMUS | Myc/Trib1/Inhba/Hspa9/Jun/Mmp14/Fos/Fgl2/Rnf41/Stat3/Bcl |
| BP | GO:200123 | negative regulation of extrinsic apoptotic signaling pathway | 17\|1209 | 91\|16097 | 0.00039 | 0.00479 | ENSMUS | Hmox1/Serpine1/Lgals3/Lmna/Cx3cl1/Nrp1/Sfrp2/Tgfbr1/Igf1/ |
| BP | GO:001097 | negative regulation of neuron projection development | 26\|1209 | 170\|16097 | 0.0004 | 0.00488 | ENSMUS | Xylt1/Ifrd1/Efna1/Rtn4/Lgals1/Ngef/Thy1/Nrp1/Neo1/Sema3b/ |
| BP | GO:004881 | dendrite morphogenesis | 26\|1209 | 170\|16097 | 0.0004 | 0.00488 | ENSMUS | Met/Efna1/Abl2/Ngef/Sgk1/Mef2a/Tiam1/Lrp8/Nrp1/Robo1/Pa |
| BP | GO:190186 | regulation of muscle tissue development | 25\|1209 | 161\|16097 | 0.0004 | 0.00495 | ENSMUS | Hmgcr/Col14a1/Igfbp5/Rgs2/Cdon/Fgfr1/Fgfr2/Tgfbr3/S100b/N |
| BP | GO:007138 | cellular response to steroid hormone stimulus | 23\|1209 | 143\|16097 | 0.00041 | 0.00497 | ENSMUS | Eif4ebp1/Atp1a1/Ncoa3/Hmga2/Sgk1/Agtr2/Klf9/Jak2/Npc1/Igf |
| BP | GO:000725 | JNK cascade | 28\|1209 | 189\|16097 | 0.00041 | 0.00504 | ENSMUS | Tnxb/Trib1/Gadd45a/Dixdc1/Nrk/Egfr/Tiam1/Ctgf/Gab1/Sfrp2/ |
| BP | GO:004800 | platelet-derived growth factor receptor signaling pathway | 13\|1209 | 60\|16097 | 0.00042 | 0.00511 | ENSMUS | F3/Iqgap1/Abl2/Gab1/Jak2/Nrp1/Tiparp/Lox/Vegfa/Ndrg4/Plek |
| BP | GO:007005 | intrinsic apoptotic signaling pathway in response to endoplasn | 13\|1209 | 60\|16097 | 0.00042 | 0.00511 | ENSMUS | Chac1/Herpud1/Atf4/Eif2ak3/Ddit3/Hyou1/Trib3/Xbp1/Cebpb/ |
| BP | GO:000979 | post-embryonic development | 20\|1209 | 117\|16097 | 0.00042 | 0.00513 | ENSMUS | Dlk1/Klf4/Abl2/Fgfr2/Selenop/Chst11/Jak2/Tiparp/Tgfbr1/Foxp |
| BP | GO:007188 | adenylate cyclase-activating adrenergic receptor signaling path | 7\|1209 | 20\|16097 | 0.00043 | 0.00518 | ENSMUS | Pde4b/Arrdc3/Gnai2/Rapgef2/Adra2a/Nos1/Gnas |
| BP | GO:200102 | regulation of endothelial cell chemotaxis | 7\|1209 | 20\|16097 | 0.00043 | 0.00518 | ENSMUS | Thbs1/Fgfr1/Met/Smoc2/Vegfa/Notch1/Kdr |
| BP | GO:004858 | developmental cell growth | 34\|1209 | 247\|16097 | 0.00043 | 0.0052 | ENSMUS | Col14a1/Rgs2/Sox9/Postn/Ifrd1/Tnfrsf12a/Sorbs2/Rtn4/Iqgap1, |
| BP | GO:007039 | response to lipoteichoic acid | 5\|1209 | 10\|16097 | 0.00043 | 0.0052 | ENSMUS | Tlr2/Cd14/Cd36/Rela/Lbp |
| BP | GO:007122 | cellular response to lipoteichoic acid | 5\|1209 | 10\|16097 | 0.00043 | 0.0052 | ENSMUS | Tlr2/Cd14/Cd36/Rela/Lbp |
| BP | GO:190161 | organic hydroxy compound biosynthetic process | 29\|1209 | 199\|16097 | 0.00043 | 0.0052 | ENSMUS | Hmgcr/Moxd1/Cyp51/Stard4/Fdft1/Msmo1/Hmgcs1/Sqle/Insig |
| MF | GO:000552 | insulin-like growth factor binding | 8\|1190 | 26\|15912 | 0.00044 | 0.01379 | ENSMUS | Igfbp5/Igfbp3/Igfbp6/Igfbp4/Cyr61/Ctgf/Igf1r/Nov |
| BP | GO:003276 | positive regulation of tumor necrosis factor production | 17\|1209 | 92\|16097 | 0.00044 | 0.00529 | ENSMUS | Thbs1/Tlr2/Cd14/Lpl/Akap12/Arfgef2/Jak2/Cd36/Tnfrsf1a/Arid5 |
| BP | GO:000727 | ensheathment of neurons | 23\|1209 | 144\|16097 | 0.00045 | 0.00534 | ENSMUS | Tlr2/Eif2ak3/Mpz/Rxra/Nab2/Atrn/B4galt5/Nab1/Ski/S100b/Nd |
| BP | GO:000836 | axon ensheathment | 23\|1209 | 144\|16097 | 0.00045 | 0.00534 | ENSMUS | Tlr2/Eif2ak3/Mpz/Rxra/Nab2/Atrn/B4galt5/Nab1/Ski/S100b/Nd |
| BP | GO:000268 | regulation of leukocyte migration | 28\|1209 | 190\|16097 | 0.00045 | 0.00534 | ENSMUS | Thbs1/Tlr2/Serpine1/Dusp1/Lgals3/Rtn4/Pecam1/Mmp14/Cx3c |
| BP | GO:190002 | regulation of substrate adhesion-dependent cell spreading | 12\|1209 | 53\|16097 | 0.00045 | 0.00535 | ENSMUS | Postn/Has2/S100a10/Nrp1/Enpp2/Actn4/Nedd9/Kank1/Rcc2/C |
| BP | GO:000863 | intrinsic apoptotic signaling pathway in response to DNA dama | 19\|1209 | 109\|16097 | 0.00045 | 0.00535 | ENSMUS | Hmox1/Myc/Fnip2/Hipk1/Cdkn1a/Ier3/Cxcl12/Tnfrsf1a/Steap3, |

| Count | Up | Up_Gene_i | Down | Down_Gene_id |
|---|---|---|---|---|
| 32 | 14 | ENSMUSG0 | 18 | ENSMUSG00000028967/ENSMUSG00000022114/ENSMUSG00000000567/ENSMUSG00000042608/ENSMUSG00000031565/ENSMUSG00000030849/ENSMUSG0 |
| 12 | 3 | ENSMUSG0 | 9 | ENSMUSG00000024912/ENSMUSG00000000567/ENSMUSG00000003032/ENSMUSG00000022463/ENSMUSG00000004040/ENSMUSG00000007613/ENSMUSG0 |
| 12 | 6 | ENSMUSG0 | 6 | ENSMUSG00000040152/ENSMUSG00000037411/ENSMUSG00000026596/ENSMUSG00000020484/ENSMUSG00000031451/ENSMUSG00000015839 |
| 11 | 6 | ENSMUSG0 | 5 | ENSMUSG00000030849/ENSMUSG00000029287/ENSMUSG00000048616/ENSMUSG00000007613/ENSMUSG00000026836 |
| 31 | 7 | ENSMUSG0 | 24 | ENSMUSG00000038418/ENSMUSG00000051391/ENSMUSG00000031490/ENSMUSG00000028893/ENSMUSG00000019970/ENSMUSG00000032715/ENSMUSG0 |
| 6 | 1 | ENSMUSG0 | 5 | ENSMUSG00000032244/ENSMUSG00000030849/ENSMUSG00000015846/ENSMUSG00000046223/ENSMUSG00000004207 |
| 6 | 2 | ENSMUSG0 | 4 | ENSMUSG00000031565/ENSMUSG00000027351/ENSMUSG00000030849/ENSMUSG00000029287 |
| 6 | 0 | | 6 | ENSMUSG00000045294/ENSMUSG00000039234/ENSMUSG00000020986/ENSMUSG00000027465/ENSMUSG00000056050/ENSMUSG00000036391 |
| 19 | 8 | ENSMUSG0 | 11 | ENSMUSG00000040152/ENSMUSG00000005413/ENSMUSG00000037411/ENSMUSG00000008730/ENSMUSG00000026596/ENSMUSG00000020484/ENSMUSG0 |
| 28 | 11 | ENSMUSG0 | 17 | ENSMUSG00000021670/ENSMUSG00000026360/ENSMUSG00000039004/ENSMUSG00000031778/ENSMUSG00000029287/ENSMUSG00000034765/ENSMUSG0 |
| 14 | 1 | ENSMUSG0 | 13 | ENSMUSG00000022114/ENSMUSG00000021670/ENSMUSG00000026360/ENSMUSG00000027351/ENSMUSG00000034765/ENSMUSG00000027996/ENSMUSG0 |
| 22 | 6 | ENSMUSG0 | 16 | ENSMUSG00000040152/ENSMUSG00000000567/ENSMUSG00000040856/ENSMUSG00000000753/ENSMUSG00000046186/ENSMUSG00000030849/ENSMUSG0 |
| 27 | 10 | ENSMUSG0 | 17 | ENSMUSG00000036390/ENSMUSG00000032064/ENSMUSG00000002489/ENSMUSG00000019997/ENSMUSG00000031714/ENSMUSG00000027996/ENSMUSG0 |
| 16 | 6 | ENSMUSG0 | 10 | ENSMUSG00000006014/ENSMUSG00000032193/ENSMUSG00000029426/ENSMUSG00000028613/ENSMUSG00000022425/ENSMUSG00000044337/ENSMUSG0 |
| 9 | 1 | ENSMUSG0 | 8 | ENSMUSG00000026360/ENSMUSG00000010095/ENSMUSG00000038615/ENSMUSG00000022462/ENSMUSG00000027894/ENSMUSG00000030096/ENSMUSG0 |
| 18 | 9 | ENSMUSG0 | 9 | ENSMUSG00000023905/ENSMUSG00000020458/ENSMUSG00000038708/ENSMUSG00000032064/ENSMUSG00000002489/ENSMUSG00000018736/ENSMUSG0 |
| 24 | 8 | ENSMUSG0 | 16 | ENSMUSG00000037071/ENSMUSG00000042406/ENSMUSG00000025408/ENSMUSG00000024789/ENSMUSG00000074794/ENSMUSG00000023845/ENSMUSG0 |
| 31 | 5 | ENSMUSG0 | 26 | ENSMUSG00000040152/ENSMUSG00000005413/ENSMUSG00000037411/ENSMUSG00000008730/ENSMUSG00000041324/ENSMUSG00000031565/ENSMUSG0 |
| 16 | 5 | ENSMUSG0 | 11 | ENSMUSG00000032501/ENSMUSG00000031770/ENSMUSG00000053581/ENSMUSG00000032715/ENSMUSG00000020601/ENSMUSG00000022037/ENSMUSG0 |
| 21 | 9 | ENSMUSG0 | 12 | ENSMUSG00000028128/ENSMUSG00000040152/ENSMUSG00000005413/ENSMUSG00000040856/ENSMUSG00000031565/ENSMUSG00000039004/ENSMUSG0 |
| 13 | 3 | ENSMUSG0 | 10 | ENSMUSG00000039646/ENSMUSG00000046186/ENSMUSG00000029287/ENSMUSG00000020484/ENSMUSG00000029050/ENSMUSG00000038400/ENSMUSG0 |
| 51 | 14 | ENSMUSG0 | 37 | ENSMUSG00000078651/ENSMUSG00000021670/ENSMUSG00000032193/ENSMUSG00000020000/ENSMUSG00000001467/ENSMUSG00000031565/ENSMUSG0 |
| 22 | 11 | ENSMUSG0 | 11 | ENSMUSG00000006800/ENSMUSG00000022346/ENSMUSG00000000567/ENSMUSG00000038418/ENSMUSG00000027947/ENSMUSG00000035342/ENSMUSG0 |
| 23 | 7 | ENSMUSG0 | 16 | ENSMUSG00000038119/ENSMUSG00000031565/ENSMUSG00000005069/ENSMUSG00000020458/ENSMUSG00000032064/ENSMUSG00000031785/ENSMUSG0 |
| 23 | 7 | ENSMUSG0 | 16 | ENSMUSG00000027995/ENSMUSG00000031668/ENSMUSG00000015846/ENSMUSG00000025402/ENSMUSG00000027312/ENSMUSG00000017929/ENSMUSG0 |
| 25 | 11 | ENSMUSG0 | 14 | ENSMUSG00000029675/ENSMUSG00000041959/ENSMUSG00000009376/ENSMUSG00000026596/ENSMUSG00000031778/ENSMUSG00000019997/ENSMUSG0 |
| 30 | 6 | ENSMUSG0 | 24 | ENSMUSG00000028967/ENSMUSG00000022114/ENSMUSG00000021670/ENSMUSG00000026360/ENSMUSG00000032501/ENSMUSG00000036390/ENSMUSG0 |
| 11 | 3 | ENSMUSG0 | 8 | ENSMUSG00000028967/ENSMUSG00000030536/ENSMUSG00000020349/ENSMUSG00000033955/ENSMUSG00000023951/ENSMUSG00000024423/ENSMUSG0 |
| 47 | 15 | ENSMUSG0 | 32 | ENSMUSG00000022346/ENSMUSG00000032501/ENSMUSG00000041324/ENSMUSG00000024359/ENSMUSG00000039899/ENSMUSG00000025373/ENSMUSG0 |
| 17 | 3 | ENSMUSG0 | 14 | ENSMUSG00000005413/ENSMUSG00000037411/ENSMUSG00000050335/ENSMUSG00000028063/ENSMUSG00000031778/ENSMUSG00000027996/ENSMUSG0 |
| 26 | 8 | ENSMUSG0 | 18 | ENSMUSG00000030657/ENSMUSG00000001627/ENSMUSG00000027954/ENSMUSG00000020458/ENSMUSG00000026259/ENSMUSG00000032340/ENSMUSG0 |
| 26 | 8 | ENSMUSG0 | 18 | ENSMUSG00000009376/ENSMUSG00000027954/ENSMUSG00000026259/ENSMUSG00000019970/ENSMUSG00000030557/ENSMUSG0 |
| 25 | 9 | ENSMUSG0 | 16 | ENSMUSG00000021670/ENSMUSG00000026360/ENSMUSG00000038119/ENSMUSG00000031565/ENSMUSG00000030849/ENSMUSG00000029287/ENSMUSG0 |
| 23 | 5 | ENSMUSG0 | 18 | ENSMUSG00000031490/ENSMUSG00000033161/ENSMUSG00000027678/ENSMUSG00000056758/ENSMUSG00000019970/ENSMUSG00000033863/ENSMUSG0 |
| 28 | 10 | ENSMUSG0 | 18 | ENSMUSG00000032501/ENSMUSG00000036390/ENSMUSG00000032064/ENSMUSG00000002489/ENSMUSG00000019997/ENSMUSG00000031714/ENSMUSG0 |
| 13 | 2 | ENSMUSG0 | 11 | ENSMUSG00000028128/ENSMUSG00000030536/ENSMUSG00000026596/ENSMUSG00000031714/ENSMUSG00000024789/ENSMUSG00000034640/ENSMUSG0 |
| 13 | 0 | | 13 | ENSMUSG00000027313/ENSMUSG00000031770/ENSMUSG00000042406/ENSMUSG00000031668/ENSMUSG00000025408/ENSMUSG00000032115/ENSMUSG0 |
| 20 | 6 | ENSMUSG0 | 14 | ENSMUSG00000040856/ENSMUSG00000003032/ENSMUSG00000026596/ENSMUSG00000030849/ENSMUSG00000024789/ENSMUSG00000034640/ENSMUSG0 |
| 7 | 2 | ENSMUSG0 | 5 | ENSMUSG00000028525/ENSMUSG00000074794/ENSMUSG00000062232/ENSMUSG00000033717/ENSMUSG00000029361 |
| 7 | 3 | ENSMUSG0 | 4 | ENSMUSG00000040152/ENSMUSG00000031565/ENSMUSG00000009376/ENSMUSG00000023951 |
| 34 | 16 | ENSMUSG0 | 18 | ENSMUSG00000026360/ENSMUSG00000000567/ENSMUSG00000001627/ENSMUSG00000023905/ENSMUSG00000020458/ENSMUSG00000030536/ENSMUSG0 |
| 5 | 1 | ENSMUSG0 | 4 | ENSMUSG00000027995/ENSMUSG00000051439/ENSMUSG00000024927/ENSMUSG00000016024 |
| 5 | 1 | ENSMUSG0 | 4 | ENSMUSG00000027995/ENSMUSG00000051439/ENSMUSG00000024927/ENSMUSG00000016024 |
| 29 | 4 | ENSMUSG0 | 25 | ENSMUSG00000021670/ENSMUSG00000020000/ENSMUSG00000001467/ENSMUSG00000024378/ENSMUSG00000021273/ENSMUSG00000031604/ENSMUSG0 |
| 8 | 6 | ENSMUSG0 | 2 | ENSMUSG00000019997/ENSMUSG00000005533 |
| 17 | 6 | ENSMUSG0 | 11 | ENSMUSG00000040152/ENSMUSG00000027995/ENSMUSG00000051439/ENSMUSG00000074582/ENSMUSG00000024789/ENSMUSG00000030341/ENSMUSG0 |
| 23 | 7 | ENSMUSG0 | 16 | ENSMUSG00000027995/ENSMUSG00000031668/ENSMUSG00000015846/ENSMUSG00000025402/ENSMUSG00000027312/ENSMUSG00000017929/ENSMUSG0 |
| 23 | 7 | ENSMUSG0 | 16 | ENSMUSG00000027995/ENSMUSG00000031668/ENSMUSG00000015846/ENSMUSG00000025402/ENSMUSG00000027312/ENSMUSG00000017929/ENSMUSG0 |
| 28 | 13 | ENSMUSG0 | 15 | ENSMUSG00000040152/ENSMUSG00000027995/ENSMUSG00000037411/ENSMUSG00000050335/ENSMUSG00000020458/ENSMUSG00000031778/ENSMUSG0 |
| 12 | 5 | ENSMUSG0 | 7 | ENSMUSG00000022367/ENSMUSG00000041959/ENSMUSG00000022425/ENSMUSG00000054808/ENSMUSG00000032702/ENSMUSG00000040945/ENSMUSG0 |
| 19 | 3 | ENSMUSG0 | 16 | ENSMUSG00000005413/ENSMUSG00000022346/ENSMUSG00000061175/ENSMUSG00000008730/ENSMUSG00000023067/ENSMUSG00000003541/ENSMUSG0 |

0000056758/ENSMUSG00000039286/ENSMUSG00000019997/ENSMUSG00000029050/ENSMUSG00000048616/ENSMUSG00000004846/ENSMUSG00000045216/ENSMUSG00000023951/ENSMUSG0000000
0000024927/ENSMUSG00000026313/ENSMUSG00000024431

0000020484/ENSMUSG00000018736/ENSMUSG00000005533/ENSMUSG00000024927/ENSMUSG00000028042/ENSMUSG00000050965/ENSMUSG00000032702/ENSMUSG00000000058/ENSMUSG0000000

0000024789/ENSMUSG00000031451/ENSMUSG00000005533/ENSMUSG00000015839/ENSMUSG00000044147
0000031849/ENSMUSG00000004846/ENSMUSG00000023951/ENSMUSG00000021070/ENSMUSG00000063142/ENSMUSG00000032487/ENSMUSG00000033717/ENSMUSG00000032350/ENSMUSG0000000
0000039384/ENSMUSG00000027177/ENSMUSG00000042594/ENSMUSG00000109511/ENSMUSG00000050965/ENSMUSG00000053716/ENSMUSG00000031548
0000029287/ENSMUSG00000033863/ENSMUSG00000027996/ENSMUSG00000026361/ENSMUSG00000007613/ENSMUSG00000039384/ENSMUSG00000047281/ENSMUSG00000000058/ENSMUSG0000000
0000005533/ENSMUSG00000081683/ENSMUSG00000039384/ENSMUSG00000020745/ENSMUSG00000027177/ENSMUSG00000061436/ENSMUSG00000032717/ENSMUSG00000031548/ENSMUSG0000000
0000024924/ENSMUSG00000056050/ENSMUSG00000034205/ENSMUSG00000025185
0000020142/ENSMUSG00000024935
0000023951/ENSMUSG00000020745/ENSMUSG00000036606
0000020889/ENSMUSG00000005533/ENSMUSG00000056501/ENSMUSG00000023951/ENSMUSG00000000134/ENSMUSG00000018476/ENSMUSG00000028042/ENSMUSG00000030168/ENSMUSG0000000
0000032244/ENSMUSG00000023905/ENSMUSG00000050335/ENSMUSG00000028063/ENSMUSG00000025373/ENSMUSG00000031778/ENSMUSG00000024789/ENSMUSG00000027996/ENSMUSG0000000
0000025234/ENSMUSG00000028793/ENSMUSG00000032350/ENSMUSG00000038068/ENSMUSG00000024182
0000004040/ENSMUSG00000023034/ENSMUSG00000007613/ENSMUSG00000023951/ENSMUSG00000050965/ENSMUSG00000000058
0000046058/ENSMUSG00000048661/ENSMUSG00000000058/ENSMUSG00000063382
0000024378/ENSMUSG00000021273/ENSMUSG00000031604/ENSMUSG00000038615/ENSMUSG00000022463/ENSMUSG00000093930/ENSMUSG00000022351/ENSMUSG00000045294/ENSMUSG0000000
0000048616/ENSMUSG00000005087/ENSMUSG00000023951/ENSMUSG00000074934/ENSMUSG00000032220
0000028613/ENSMUSG00000020423/ENSMUSG00000018736/ENSMUSG00000035764/ENSMUSG00000030852/ENSMUSG00000020745/ENSMUSG00000018476/ENSMUSG00000021379/ENSMUSG0000000
0000002881/ENSMUSG00000029050/ENSMUSG00000005125/ENSMUSG00000028995/ENSMUSG00000021379/ENSMUSG00000022037/ENSMUSG00000039952/ENSMUSG00000026812/ENSMUSG0000000
0000031523/ENSMUSG00000007613/ENSMUSG00000022817/ENSMUSG00000054808/ENSMUSG00000024533/ENSMUSG00000038859/ENSMUSG00000025372/ENSMUSG00000026812
0000023067/ENSMUSG00000027351/ENSMUSG00000046733/ENSMUSG00000032715/ENSMUSG00000049323/ENSMUSG00000034765/ENSMUSG00000027996/ENSMUSG00000005533/ENSMUSG0000000
0000029816/ENSMUSG00000074934
0000004040/ENSMUSG00000022508/ENSMUSG00000037465/ENSMUSG00000020484/ENSMUSG00000007817/ENSMUSG00000031451/ENSMUSG00000026361/ENSMUSG00000056501/ENSMUSG0000000
0000007613/ENSMUSG00000024927/ENSMUSG00000023951/ENSMUSG00000037815/ENSMUSG00000024073/ENSMUSG00000026836/ENSMUSG00000032350/ENSMUSG00000036432
0000026728/ENSMUSG00000049792/ENSMUSG00000020745/ENSMUSG00000028161/ENSMUSG00000032702/ENSMUSG00000032593/ENSMUSG00000062232/ENSMUSG00000018965/ENSMUSG0000000
0000002489/ENSMUSG00000028613/ENSMUSG00000020745/ENSMUSG00000096054/ENSMUSG00000028161/ENSMUSG00000024924/ENSMUSG00000062232/ENSMUSG00000020132/ENSMUSG0000000
0000048616/ENSMUSG00000007613/ENSMUSG00000033323/ENSMUSG00000022687/ENSMUSG00000022752/ENSMUSG00000026313/ENSMUSG00000024431/ENSMUSG00000074934/ENSMUSG0000000
0000024789/ENSMUSG00000024413/ENSMUSG00000040021/ENSMUSG00000039202/ENSMUSG00000048249/ENSMUSG00000003814/ENSMUSG00000018965/ENSMUSG00000029249/ENSMUSG0000000
0000027996/ENSMUSG00000005533/ENSMUSG00000081683/ENSMUSG00000039384/ENSMUSG00000020745/ENSMUSG00000027177/ENSMUSG00000061436/ENSMUSG00000032717/ENSMUSG0000000
0000023951/ENSMUSG00000036564/ENSMUSG00000040268/ENSMUSG00000039844/ENSMUSG00000032220
0000032715/ENSMUSG00000020484/ENSMUSG00000056501/ENSMUSG00000021831/ENSMUSG00000027230/ENSMUSG00000002083/ENSMUSG00000024807
0000007613/ENSMUSG00000023951/ENSMUSG00000079509/ENSMUSG00000040268/ENSMUSG00000038539/ENSMUSG00000031393/ENSMUSG00000026826/ENSMUSG00000032220

0000038708/ENSMUSG00000002489/ENSMUSG00000026014/ENSMUSG00000018736/ENSMUSG00000033323/ENSMUSG00000023951/ENSMUSG00000022752/ENSMUSG00000020745/ENSMUSG0000000

0000093930/ENSMUSG00000022351/ENSMUSG00000045294/ENSMUSG00000039004/ENSMUSG00000026675/ENSMUSG00000034926/ENSMUSG00000020889/ENSMUSG00000023068/ENSMUSG0000000

0000037447/ENSMUSG00000022708/ENSMUSG00000022037/ENSMUSG00000016024/ENSMUSG00000027551
0000002881/ENSMUSG00000029050/ENSMUSG00000005125/ENSMUSG00000028995/ENSMUSG00000021379/ENSMUSG00000022037/ENSMUSG00000039952/ENSMUSG00000026812/ENSMUSG0000000
0000002881/ENSMUSG00000029050/ENSMUSG00000005125/ENSMUSG00000028995/ENSMUSG00000021379/ENSMUSG00000022037/ENSMUSG00000039952/ENSMUSG00000026812/ENSMUSG0000000
0000031451/ENSMUSG00000021508/ENSMUSG00000025340/ENSMUSG00000023951/ENSMUSG00000020682/ENSMUSG00000003814/ENSMUSG00000056050/ENSMUSG00000016024/ENSMUSG0000000
0000003814
0000030341/ENSMUSG00000026389/ENSMUSG00000005087/ENSMUSG00000044337/ENSMUSG00000061436/ENSMUSG00000047281/ENSMUSG00000006445/ENSMUSG00000022037/ENSMUSG0000000

5442/ENSMUSG00000024085/ENSMUSG00000069895/ENSMUSG00000039952

8859/ENSMUSG00000024052/ENSMUSG00000044167/ENSMUSG00000020091/ENSMUSG00000042903/ENSMUSG00000019943/ENSMUSG00000050697/ENSMUSG00000026812/ENSMUSG00000005871/E

9361/ENSMUSG00000024431/ENSMUSG00000019943

1548/ENSMUSG00000005871
9844/ENSMUSG00000037902/ENSMUSG00000024182

8707/ENSMUSG00000005871
7613/ENSMUSG00000024927/ENSMUSG00000023951/ENSMUSG00000037815/ENSMUSG00000024073/ENSMUSG00000020044/ENSMUSG00000026836/ENSMUSG00000032350/ENSMUSG00000031548/E

9004/ENSMUSG00000026675/ENSMUSG00000089774/ENSMUSG00000034926/ENSMUSG00000024413/ENSMUSG00000020889/ENSMUSG00000023068/ENSMUSG00000032018/ENSMUSG00000030747/E

5954/ENSMUSG00000026812
4207/ENSMUSG00000034593

9384/ENSMUSG00000040021/ENSMUSG00000027177/ENSMUSG00000042594/ENSMUSG00000109511/ENSMUSG00000050965/ENSMUSG00000053716/ENSMUSG00000031548/ENSMUSG00000035621/E

2040/ENSMUSG00000005087/ENSMUSG00000023951/ENSMUSG00000000134/ENSMUSG00000039384/ENSMUSG00000024812/ENSMUSG00000028042/ENSMUSG00000050965/ENSMUSG00000069895/E

4147/ENSMUSG00000031393/ENSMUSG00000025326/ENSMUSG00000026812
8965/ENSMUSG00000068740/ENSMUSG00000025326/ENSMUSG00000025372
0697/ENSMUSG00000005871
1548/ENSMUSG00000024431/ENSMUSG00000025326/ENSMUSG00000005886
1548/ENSMUSG00000039844/ENSMUSG00000037902/ENSMUSG00000024182

4423/ENSMUSG00000020132/ENSMUSG00000024431/ENSMUSG00000031393

2018/ENSMUSG00000033105/ENSMUSG00000003752/ENSMUSG00000021036/ENSMUSG00000062232/ENSMUSG00000029249/ENSMUSG00000063229/ENSMUSG00000026826/ENSMUSG00000050697/E

4207/ENSMUSG00000034593
4207/ENSMUSG00000034593
4934

0609/ENSMUSG00000002083

ENSMUSG00000034593

ENSMUSG00000036432/ENSMUSG00000022074/ENSMUSG00000026628

ENSMUSG00000063142/ENSMUSG00000033105/ENSMUSG00000024924/ENSMUSG00000003752/ENSMUSG00000021036/ENSMUSG00000062232/ENSMUSG00000029249/ENSMUSG00000063229/ENSMUS

ENSMUSG00000005871

ENSMUSG00000034201/ENSMUSG00000035356/ENSMUSG00000033454/ENSMUSG00000035992/ENSMUSG00000015839/ENSMUSG00000040025/ENSMUSG00000031548/ENSMUSG00000024425/ENSMUS

ENSMUSG00000024210/ENSMUSG00000034593

G00000031835/ENSMUSG00000031393/ENSMUSG00000026826/ENSMUSG00000050697/ENSMUSG00000024210/ENSMUSG00000034593

G00000005871

| Category | GOID | Description | GeneRatio | BgRatio | pvalue | padj | geneID | geneName |
|---|---|---|---|---|---|---|---|---|
| BP | GO:001071 | negative regulation of epithelial to mesenchymal transition | 8\|1209 | 26\|16097 | 0.00045 | 0.00535 | ENSMUS | Vasn/Efna1/Tgfbr3/Ppp2ca/Sfrp2/Nog/Vegfa/Sfrp1 |
| BP | GO:190367 | positive regulation of sprouting angiogenesis | 10\|1209 | 39\|16097 | 0.00047 | 0.00549 | ENSMUS | Hmox1/Klf4/Itga5/Nrp1/Vegfa/Ptgs2/Kdr/Hdac7/Pik3c2a/Rhoj |
| BP | GO:00003( | response to reactive oxygen species | 28\|1209 | 191\|1609 | 0.00049 | 0.00576 | ENSMUS | Thbs1/Hmox1/Met/Ppp1r15b/Jun/Fos/Sesn2/Egfr/Hsph1/Cd36, |
| BP | GO:190384 | negative regulation of cellular response to transforming growth | 13\|1209 | 61\|16097 | 0.0005 | 0.00583 | ENSMUS | Vasn/Cd109/Aspn/Tgfbr3/Chst11/Xbp1/Ski/Pmepa1/Ltbp1/Eid2 |
| MF | GO:005142 | hormone receptor binding | 25\|1190 | 164\|1591 | 0.0005 | 0.01517 | ENSMUS | Ncoa3/Hmga1/Rxra/Stat3/Nr4a1/Jak2/Fabp4/Gpha2/Tacc2/Ceb |
| BP | GO:190355 | positive regulation of tumor necrosis factor superfamily cytoki | 17\|1209 | 93\|16097 | 0.0005 | 0.0059 | ENSMUS | Thbs1/Tlr2/Cd14/Lpl/Akap12/Arfgef2/Jak2/Cd36/Tnfrsf1a/Arid5 |
| BP | GO:00309( | forebrain development | 43\|1209 | 340\|1609 | 0.00053 | 0.00613 | ENSMUS | Col3a1/Cdon/Inhba/Lamb1/Fgfr1/Pex5/Rtn4/Fgfr2/Dixdc1/Adg |
| BP | GO:00032{ | ventricular septum development | 15\|1209 | 77\|16097 | 0.00053 | 0.00614 | ENSMUS | Cyr61/Fgfr2/Tgfbr3/Ltbp1/Nog/Tgfbr1/Hectd1/Robo1/Nfatc1/N |
| CC | GO:000591 | adherens junction | 38\|1232 | 288\|1631 | 0.00053 | 0.01331 | ENSMUS | Plec/Itga5/Cdh5/Iqgap1/Cdh11/Atp2a2/Jak2/Nrp1/Dlc1/Tns1/N |
| BP | GO:003017 | positive regulation of Wnt signaling pathway | 21\|1209 | 128\|1609 | 0.00054 | 0.00631 | ENSMUS | Col1a1/Sulf2/Tlr2/Abl2/Fgfr2/Dixdc1/Egfr/Tnks2/Sfrp2/Cdc73/S |
| MF | GO:000371 | transcription coregulator activity | 51\|1190 | 426\|1591 | 0.00056 | 0.01641 | ENSMUS | Hspa1a/Hipk1/Med13l/Sqstm1/Jun/Nfe2l1/Ncoa3/Hmga1/Serta |
| BP | GO:00066 | cholesterol biosynthetic process | 10\|1209 | 40\|16097 | 0.00058 | 0.00672 | ENSMUS | Hmgcr/Cyp51/Fdft1/Hmgcs1/Insig1/Hsd17b7/Dhcr24/Sc5d/Lss |
| BP | GO:190495 | negative regulation of establishment of protein localization | 28\|1209 | 193\|1609 | 0.00058 | 0.00672 | ENSMUS | Hmgcr/Cyp51/Insig1/Wwp2/Cd34/Cx3cl1/Ppp2ca/Hsph1/C1qt |
| BP | GO:005126 | protein homooligomerization | 44\|1209 | 352\|1609 | 0.00059 | 0.00674 | ENSMUS | Hmox1/Scube3/Marcks/Pex5/Eif2ak3/Itpr3/Rxra/Thbs4/Steap4/ |
| BP | GO:004308 | regulation of GTPase activity | 37\|1209 | 281\|1609 | 0.00059 | 0.00677 | ENSMUS | Spry2/Rgs16/S100a10/Iqgap1/Pecam1/Ngef/Cx3cl1/Thy1/Tiam |
| BP | GO:001025 | endomembrane system organization | 46\|1209 | 373\|1609 | 0.0006 | 0.00685 | ENSMUS | Atp8b2/Pacsin2/Sqstm1/Pex5/Rtn4/Dnm1/Eif2ak3/Lmna/Atp2a |
| BP | GO:200073 | negative regulation of stem cell differentiation | 7\|1209 | 21\|16097 | 0.0006 | 0.0069 | ENSMUS | Hspa9/Ncoa3/Stat3/Notch1/Rest/Nfe2l2/Hes1 |
| BP | GO:006044 | branching involved in salivary gland morphogenesis | 8\|1209 | 27\|16097 | 0.0006 | 0.0069 | ENSMUS | Fgfr1/Met/Fgfr2/Nrp1/Plxnd1/Tgm2/Dag1/Btbd7 |
| CC | GO:000013 | Golgi membrane | 32\|1232 | 231\|1631 | 0.00062 | 0.01479 | ENSMUS | Sgms2/Xylt1/Csgalnact2/Chpf/Arfgef2/Gorasp2/Sec23a/Rab2a/( |
| BP | GO:005072 | regulation of inflammatory response | 41\|1209 | 324\|1609 | 0.00069 | 0.00791 | ENSMUS | Ldlr/Tlr2/Serpine1/Serpinf1/A2m/Lpl/Cdh5/Serping1/Nfe2l1/Ie |
| BP | GO:003051 | negative regulation of BMP signaling pathway | 11\|1209 | 48\|16097 | 0.0007 | 0.00791 | ENSMUS | Chrdl1/Ski/Sfrp2/Nog/Twsg1/Notch1/Hipk2/Lemd3/Sfrp1/Lem |
| BP | GO:004667 | negative regulation of insulin secretion | 11\|1209 | 48\|16097 | 0.0007 | 0.00791 | ENSMUS | Hmgcr/Ppp3ca/Nov/Adra2a/Rest/Nos1/Sfrp1/Ucp2/Pim3/Midn |
| BP | GO:000315 | outflow tract morphogenesis | 15\|1209 | 79\|16097 | 0.0007 | 0.00795 | ENSMUS | Thbs1/Jun/Fgfr2/Tgfbr3/Nrp1/Sfrp2/Nog/Robo1/Vegfa/Plxnd1/ |
| BP | GO:00610 | regulation of protein tyrosine kinase activity | 14\|1209 | 71\|16097 | 0.00071 | 0.00798 | ENSMUS | Errfi1/Efna1/Gprc5a/Thy1/Lrp8/Hsph1/Gas6/Nedd9/Epha4/Prn |
| BP | GO:00610 | positive regulation of protein tyrosine kinase activity | 10\|1209 | 41\|16097 | 0.00072 | 0.00807 | ENSMUS | Efna1/Lrp8/Hsph1/Gas6/Nedd9/Epha4/Prnp/Adra2a/Ace/Grem |
| BP | GO:190265 | secondary alcohol biosynthetic process | 10\|1209 | 41\|16097 | 0.00072 | 0.00807 | ENSMUS | Hmgcr/Cyp51/Fdft1/Hmgcs1/Insig1/Hsd17b7/Dhcr24/Sc5d/Lss |
| BP | GO:004352 | regulation of neuron apoptotic process | 34\|1209 | 254\|1609 | 0.00072 | 0.00807 | ENSMUS | Mt1/Hmox1/Egr1/Atf4/Jun/Aars/Ddit3/Hyou1/Cx3cl1/Hsph1/B |
| BP | GO:00018 | placenta development | 24\|1209 | 158\|1609 | 0.00072 | 0.00807 | ENSMUS | Fosl1/Serpine1/Xist/Met/Cdkn1c/Ncoa3/Cyr61/Fgfr2/Nrk/Rxra/ |
| BP | GO:00162( | regulation of striated muscle tissue development | 24\|1209 | 158\|1609 | 0.00072 | 0.00807 | ENSMUS | Hmgcr/Col14a1/Rgs2/Cdon/Fgfr1/Fgfr2/Tgfbr3/S100b/Nog/Tgfb |
| CC | GO:003125 | cell leading edge | 45\|1232 | 364\|1631 | 0.00073 | 0.01609 | ENSMUS | Itga5/Tnfrsf12a/Atf4/Iqgap1/Pecam1/Ablim3/Abl2/S100a6/Myl |
| MF | GO:003199 | insulin-like growth factor I binding | 5\|1190 | 11\|15912 | 0.00073 | 0.02071 | ENSMUS | Igfbp5/Igfbp3/Igfbp6/Igfbp4/Igf1r |
| BP | GO:007163 | positive regulation of transforming growth factor beta product | 6\|1209 | 16\|16097 | 0.00074 | 0.0082 | ENSMUS | Thbs1/Cd34/Cx3cl1/Furin/Lum/Ptgs2 |
| BP | GO:009005 | negative regulation of cell migration involved in sprouting angi | 6\|1209 | 16\|16097 | 0.00074 | 0.0082 | ENSMUS | Thbs1/Klf4/Spred1/Meox2/Pik3r2/Notch1 |
| BP | GO:004361 | regulation of transcription from RNA polymerase II promoter ir | 5\|1209 | 11\|16097 | 0.00075 | 0.00825 | ENSMUS | Hmox1/Atf4/Sesn2/Cd36/Nfe2l2 |
| BP | GO:006052 | prostate glandular acinus development | 5\|1209 | 11\|16097 | 0.00075 | 0.00825 | ENSMUS | Fgfr2/Rxra/Igf1/Notch1/Sfrp1 |
| BP | GO:006073 | prostate gland growth | 5\|1209 | 11\|16097 | 0.00075 | 0.00825 | ENSMUS | Sox9/Fgfr2/Igf1/Ube3a/Psap |
| BP | GO:007083 | basement membrane assembly | 5\|1209 | 11\|16097 | 0.00075 | 0.00825 | ENSMUS | Lamb1/Phldb1/Plod3/Lamb2/Dag1 |
| BP | GO:001921 | regulation of lipid metabolic process | 39\|1209 | 305\|1609 | 0.00076 | 0.00835 | ENSMUS | Ldlr/Egr1/Stard4/Scd1/Atp1a1/Nfe2l1/Acsl3/Eif2ak3/Cyr61/Insi |
| CC | GO:00164 | myosin II complex | 6\|1232 | 16\|16313 | 0.00076 | 0.01609 | ENSMUS | Myh11/Myh9/Myh14/Ttn/Limch1/Myl9 |
| CC | GO:000152 | microfibril | 5\|1232 | 11\|16313 | 0.00077 | 0.01609 | ENSMUS | Mfap4/Mfap5/Ltbp1/Adamts10/Mfap1a |
| BP | GO:009027 | negative regulation of peptide hormone secretion | 12\|1209 | 56\|16097 | 0.00077 | 0.00848 | ENSMUS | Hmgcr/Ppp3ca/Nov/Adra2a/Rest/Nos1/Sfrp1/Ucp2/Pim3/Midn |
| BP | GO:006032 | cell chemotaxis | 35\|1209 | 265\|1609 | 0.00077 | 0.00849 | ENSMUS | Thbs1/Serpine1/Fgfr1/Dusp1/Met/Lgals3/Smoc2/Pde4b/Cx3cl1 |
| CC | GO:00423{ | sarcolemma | 24\|1232 | 158\|1631 | 0.00078 | 0.01609 | ENSMUS | Plec/Slc2a1/Ahnak/Atp1a1/Lama2/Slc38a2/Col6a2/Cd36/Slc30 |
| BP | GO:002153 | telencephalon development | 30\|1209 | 216\|1609 | 0.00079 | 0.00859 | ENSMUS | Col3a1/Cdon/Inhba/Lamb1/Fgfr1/Pex5/Rtn4/Dixdc1/Adgrg1/E |
| BP | GO:000692 | substrate-dependent cell migration | 8\|1209 | 28\|16097 | 0.00079 | 0.00859 | ENSMUS | Fgfr1/Tnfrsf12a/Abl2/Nrp1/Robo1/Itga11/Epha8/Itga2 |
| BP | GO:004253 | tumor necrosis factor biosynthetic process | 8\|1209 | 28\|16097 | 0.00079 | 0.00859 | ENSMUS | Errfi1/Thbs1/Tlr2/Akap12/Jak2/Lbp/App/Ube2j1 |
| BP | GO:004253 | regulation of tumor necrosis factor biosynthetic process | 8\|1209 | 28\|16097 | 0.00079 | 0.00859 | ENSMUS | Errfi1/Thbs1/Tlr2/Akap12/Jak2/Lbp/App/Ube2j1 |
| BP | GO:200035 | negative regulation of endothelial cell apoptotic process | 8\|1209 | 28\|16097 | 0.00079 | 0.00859 | ENSMUS | Serpine1/Abl2/Gas6/Angptl4/Tek/Nfe2l2/Kdr/Ndnf |
| BP | GO:00468 | positive regulation of lipid biosynthetic process | 15\|1209 | 80\|16097 | 0.0008 | 0.0087 | ENSMUS | Ldlr/Stard4/Acsl3/Cyr61/Bmp6/Adgrf5/Nr1d1/Igf1r/Igf1/Zbtb2( |
| BP | GO:190303 | negative regulation of response to wounding | 15\|1209 | 80\|16097 | 0.0008 | 0.0087 | ENSMUS | Thbs1/Hmgcr/Serpine1/Xylt1/Cd109/Rtn4/Serping1/Cdkn1a/Cc |
| BP | GO:00082{ | insulin receptor signaling pathway | 19\|1209 | 114\|1609 | 0.0008 | 0.0087 | ENSMUS | Eif4ebp1/Socs3/Ndel1/Igf1r/Pik3r2/Rela/Gnai2/Zbtb7b/Prkca/K |

| Count | Up | Up_Gene_i | Down | Down_Gene_id |
| --- | --- | --- | --- | --- |
| 8 | 0 | | 8 | ENSMUSG00000039646/ENSMUSG00000027954/ENSMUSG00000029287/ENSMUSG00000020349/ENSMUSG00000027996/ENSMUSG00000048616/ENSMUSG0 |
| 10 | 5 | ENSMUSG0 | 5 | ENSMUSG00000005413/ENSMUSG00000003032/ENSMUSG00000000555/ENSMUSG00000023951/ENSMUSG00000032487 |
| 28 | 12 | ENSMUSG0 | 16 | ENSMUSG00000040152/ENSMUSG00000005413/ENSMUSG00000009376/ENSMUSG00000046062/ENSMUSG00000028893/ENSMUSG00000024927/ENSMUSG0 |
| 13 | 3 | ENSMUSG0 | 10 | ENSMUSG00000039646/ENSMUSG00000046186/ENSMUSG00000029287/ENSMUSG00000020484/ENSMUSG00000029050/ENSMUSG00000038400/ENSMUSG0 |
| 25 | 3 | ENSMUSG0 | 22 | ENSMUSG00000027678/ENSMUSG00000046711/ENSMUSG00000015846/ENSMUSG00000004040/ENSMUSG00000023034/ENSMUSG00000024789/ENSMUSG0 |
| 17 | 6 | ENSMUSG0 | 11 | ENSMUSG00000040152/ENSMUSG00000027995/ENSMUSG00000051439/ENSMUSG00000074582/ENSMUSG00000024789/ENSMUSG00000030341/ENSMUSG0 |
| 43 | 17 | ENSMUSG0 | 26 | ENSMUSG00000038119/ENSMUSG00000041324/ENSMUSG00000031565/ENSMUSG00000005069/ENSMUSG00000020458/ENSMUSG00000030849/ENSMUSG0 |
| 15 | 9 | ENSMUSG0 | 6 | ENSMUSG00000030849/ENSMUSG00000029287/ENSMUSG00000048616/ENSMUSG00000007613/ENSMUSG00000035247/ENSMUSG00000026836 |
| 38 | 15 | ENSMUSG0 | 23 | ENSMUSG00000022565/ENSMUSG00000000555/ENSMUSG00000030536/ENSMUSG00000029467/ENSMUSG00000024789/ENSMUSG00000031523/ENSMUSG0 |
| 21 | 9 | ENSMUSG0 | 12 | ENSMUSG00000006800/ENSMUSG00000027995/ENSMUSG00000026596/ENSMUSG00000030849/ENSMUSG00000032064/ENSMUSG00000024811/ENSMUSG0 |
| 51 | 13 | ENSMUSG0 | 38 | ENSMUSG00000008730/ENSMUSG00000018076/ENSMUSG00000015837/ENSMUSG00000038615/ENSMUSG00000027678/ENSMUSG00000046711/ENSMUSG0 |
| 10 | 0 | | 10 | ENSMUSG00000021670/ENSMUSG00000001467/ENSMUSG00000021273/ENSMUSG00000093930/ENSMUSG00000045294/ENSMUSG00000026675/ENSMUSG0 |
| 28 | 10 | ENSMUSG0 | 18 | ENSMUSG00000021670/ENSMUSG00000001467/ENSMUSG00000045294/ENSMUSG00000031778/ENSMUSG00000020349/ENSMUSG00000032340/ENSMUSG0 |
| 44 | 16 | ENSMUSG0 | 28 | ENSMUSG00000005413/ENSMUSG00000038677/ENSMUSG00000005069/ENSMUSG00000031668/ENSMUSG00000042644/ENSMUSG00000015846/ENSMUSG0 |
| 37 | 12 | ENSMUSG0 | 25 | ENSMUSG00000022114/ENSMUSG00000041959/ENSMUSG00000030536/ENSMUSG00000026259/ENSMUSG00000031778/ENSMUSG00000002489/ENSMUSG0 |
| 46 | 4 | ENSMUSG0 | 42 | ENSMUSG00000060671/ENSMUSG00000016664/ENSMUSG00000015837/ENSMUSG00000005069/ENSMUSG00000020458/ENSMUSG00000031668/ENSMUSG0 |
| 7 | 2 | ENSMUSG0 | 5 | ENSMUSG00000024359/ENSMUSG00000027678/ENSMUSG00000004040/ENSMUSG00000029249/ENSMUSG00000015839 |
| 8 | 2 | ENSMUSG0 | 6 | ENSMUSG00000031565/ENSMUSG00000009376/ENSMUSG00000030849/ENSMUSG00000037820/ENSMUSG00000039952/ENSMUSG00000041702 |
| 32 | 5 | ENSMUSG0 | 27 | ENSMUSG00000050931/ENSMUSG00000030657/ENSMUSG00000042042/ENSMUSG00000032997/ENSMUSG00000074582/ENSMUSG00000014959/ENSMUSG0 |
| 41 | 14 | ENSMUSG0 | 27 | ENSMUSG00000032193/ENSMUSG00000027995/ENSMUSG00000037411/ENSMUSG00000000753/ENSMUSG00000030111/ENSMUSG00000038615/ENSMUSG0 |
| 11 | 2 | ENSMUSG0 | 9 | ENSMUSG00000029050/ENSMUSG00000027996/ENSMUSG00000048616/ENSMUSG00000024098/ENSMUSG00000061436/ENSMUSG00000048661/ENSMUSG0 |
| 11 | 3 | ENSMUSG0 | 8 | ENSMUSG00000021670/ENSMUSG00000028161/ENSMUSG00000033717/ENSMUSG00000029249/ENSMUSG00000029361/ENSMUSG00000031548/ENSMUSG0 |
| 15 | 9 | ENSMUSG0 | 6 | ENSMUSG00000040152/ENSMUSG00000030849/ENSMUSG00000029287/ENSMUSG00000027996/ENSMUSG00000048616/ENSMUSG00000023951 |
| 14 | 6 | ENSMUSG0 | 8 | ENSMUSG00000028967/ENSMUSG00000027954/ENSMUSG00000046733/ENSMUSG00000028613/ENSMUSG00000031451/ENSMUSG00000079037/ENSMUSG0 |
| 10 | 4 | ENSMUSG0 | 6 | ENSMUSG00000027954/ENSMUSG00000028613/ENSMUSG00000031451/ENSMUSG00000079037/ENSMUSG00000033717/ENSMUSG00000074934 |
| 10 | 0 | | 10 | ENSMUSG00000021670/ENSMUSG00000001467/ENSMUSG00000021273/ENSMUSG00000093930/ENSMUSG00000045294/ENSMUSG00000026675/ENSMUSG0 |
| 34 | 8 | ENSMUSG0 | 26 | ENSMUSG00000031765/ENSMUSG00000005413/ENSMUSG00000038418/ENSMUSG00000042406/ENSMUSG00000031960/ENSMUSG00000025408/ENSMUSG0 |
| 24 | 10 | ENSMUSG0 | 14 | ENSMUSG00000024912/ENSMUSG00000037411/ENSMUSG00000009376/ENSMUSG00000027678/ENSMUSG00000030849/ENSMUSG00000015846/ENSMUSG0 |
| 24 | 8 | ENSMUSG0 | 16 | ENSMUSG00000021670/ENSMUSG00000026360/ENSMUSG00000038119/ENSMUSG00000031565/ENSMUSG00000030849/ENSMUSG00000029287/ENSMUSG0 |
| 45 | 15 | ENSMUSG0 | 30 | ENSMUSG00000000555/ENSMUSG00000023905/ENSMUSG00000042406/ENSMUSG00000030536/ENSMUSG00000026596/ENSMUSG00000001025/ENSMUSG0 |
| 5 | 4 | ENSMUSG0 | 1 | ENSMUSG00000005533 |
| 6 | 2 | ENSMUSG0 | 4 | ENSMUSG00000040152/ENSMUSG00000031778/ENSMUSG00000030530/ENSMUSG00000032487 |
| 6 | 3 | ENSMUSG0 | 3 | ENSMUSG00000040152/ENSMUSG00000003032/ENSMUSG00000027351 |
| 5 | 1 | ENSMUSG0 | 4 | ENSMUSG00000005413/ENSMUSG00000042406/ENSMUSG00000028893/ENSMUSG00000015839 |
| 5 | 2 | ENSMUSG0 | 3 | ENSMUSG00000030849/ENSMUSG00000015846/ENSMUSG00000031548 |
| 5 | 1 | ENSMUSG0 | 4 | ENSMUSG00000000567/ENSMUSG00000030849/ENSMUSG00000025326/ENSMUSG00000004207 |
| 5 | 2 | ENSMUSG0 | 3 | ENSMUSG00000048537/ENSMUSG00000004846/ENSMUSG00000039952 |
| 39 | 11 | ENSMUSG0 | 28 | ENSMUSG00000032193/ENSMUSG00000038418/ENSMUSG00000024378/ENSMUSG00000037071/ENSMUSG00000033161/ENSMUSG00000038615/ENSMUSG0 |
| 6 | 5 | ENSMUSG0 | 1 | ENSMUSG00000022443 |
| 5 | 4 | ENSMUSG0 | 1 | ENSMUSG00000068479 |
| 12 | 3 | ENSMUSG0 | 9 | ENSMUSG00000021670/ENSMUSG00000028161/ENSMUSG00000033717/ENSMUSG00000029249/ENSMUSG00000029361/ENSMUSG00000031548/ENSMUSG0 |
| 35 | 11 | ENSMUSG0 | 24 | ENSMUSG00000040152/ENSMUSG00000037411/ENSMUSG00000031565/ENSMUSG00000009376/ENSMUSG00000050335/ENSMUSG00000028525/ENSMUSG0 |
| 24 | 5 | ENSMUSG0 | 19 | ENSMUSG00000022565/ENSMUSG00000028645/ENSMUSG00000069833/ENSMUSG00000033161/ENSMUSG00000022462/ENSMUSG00000020241/ENSMUSG0 |
| 30 | 11 | ENSMUSG0 | 19 | ENSMUSG00000038119/ENSMUSG00000041324/ENSMUSG00000031565/ENSMUSG00000005069/ENSMUSG00000020458/ENSMUSG00000032064/ENSMUSG0 |
| 8 | 4 | ENSMUSG0 | 4 | ENSMUSG00000031565/ENSMUSG00000023905/ENSMUSG00000026596/ENSMUSG00000028661 |
| 8 | 2 | ENSMUSG0 | 6 | ENSMUSG00000028967/ENSMUSG00000040152/ENSMUSG00000027995/ENSMUSG00000024789/ENSMUSG00000016024/ENSMUSG00000028277 |
| 8 | 2 | ENSMUSG0 | 6 | ENSMUSG00000028967/ENSMUSG00000040152/ENSMUSG00000027995/ENSMUSG00000024789/ENSMUSG00000016024/ENSMUSG00000028277 |
| 8 | 4 | ENSMUSG0 | 4 | ENSMUSG00000037411/ENSMUSG00000026596/ENSMUSG00000031451/ENSMUSG00000015839 |
| 15 | 4 | ENSMUSG0 | 11 | ENSMUSG00000032193/ENSMUSG00000024378/ENSMUSG00000032883/ENSMUSG00000039004/ENSMUSG00000020889/ENSMUSG00000005533/ENSMUSG0 |
| 15 | 4 | ENSMUSG0 | 11 | ENSMUSG00000040152/ENSMUSG00000021670/ENSMUSG00000037411/ENSMUSG00000030657/ENSMUSG00000046186/ENSMUSG00000020458/ENSMUSG0 |
| 19 | 4 | ENSMUSG0 | 15 | ENSMUSG00000031490/ENSMUSG00000018736/ENSMUSG00000005533/ENSMUSG00000024927/ENSMUSG00000028042/ENSMUSG00000050965/ENSMUSG0 |

0000023951/ENSMUSG00000031548

0000018476/ENSMUSG00000024423/ENSMUSG00000050965/ENSMUSG00000040268/ENSMUSG00000044167/ENSMUSG00000015839/ENSMUSG00000009630/ENSMUSG00000050697/ENSMUSG0000003
0000046058/ENSMUSG00000048661/ENSMUSG00000000058/ENSMUSG00000063382
0000024784/ENSMUSG00000030852/ENSMUSG00000056501/ENSMUSG00000037447/ENSMUSG00000022096/ENSMUSG00000048490/ENSMUSG00000040021/ENSMUSG00000054808/ENSMUSG0000010
0000037447/ENSMUSG00000022708/ENSMUSG00000022037/ENSMUSG00000016024/ENSMUSG00000027551
0000032064/ENSMUSG00000031785/ENSMUSG00000056758/ENSMUSG00000028613/ENSMUSG00000020423/ENSMUSG00000018736/ENSMUSG00000029050/ENSMUSG00000031523/ENSMUSG0000003

0000055322/ENSMUSG00000005125/ENSMUSG00000026796/ENSMUSG00000031788/ENSMUSG00000033955/ENSMUSG00000023951/ENSMUSG00000022817/ENSMUSG00000037815/ENSMUSG0000004
0000027996/ENSMUSG00000026361/ENSMUSG00000032702/ENSMUSG00000073563/ENSMUSG00000031548/ENSMUSG00000003345
0000049800/ENSMUSG00000025066/ENSMUSG00000050567/ENSMUSG00000040463/ENSMUSG00000030557/ENSMUSG00000032715/ENSMUSG00000007817/ENSMUSG00000029050/ENSMUSG0000003
0000034926/ENSMUSG00000032018/ENSMUSG00000033105/ENSMUSG00000050697
0000031451/ENSMUSG00000025340/ENSMUSG00000028161/ENSMUSG00000033717/ENSMUSG00000029249/ENSMUSG00000029361/ENSMUSG00000031548/ENSMUSG00000035828/ENSMUSG0000002
0000029050/ENSMUSG00000027293/ENSMUSG00000070738/ENSMUSG00000030499/ENSMUSG00000018339/ENSMUSG00000037820/ENSMUSG00000063142/ENSMUSG00000030527/ENSMUSG0000007
0000022508/ENSMUSG00000018736/ENSMUSG00000081683/ENSMUSG00000020745/ENSMUSG00000036606/ENSMUSG00000027465/ENSMUSG00000006445/ENSMUSG00000032413/ENSMUSG0000004
0000028063/ENSMUSG00000029467/ENSMUSG00000021950/ENSMUSG00000074582/ENSMUSG00000014959/ENSMUSG00000018736/ENSMUSG00000041779/ENSMUSG00000047187/ENSMUSG0000004

0000020986/ENSMUSG00000047187/ENSMUSG00000036356/ENSMUSG00000038400/ENSMUSG00000030341/ENSMUSG00000030530/ENSMUSG00000079111/ENSMUSG00000010392/ENSMUSG0000003
0000003541/ENSMUSG00000031778/ENSMUSG00000022508/ENSMUSG00000024789/ENSMUSG00000020889/ENSMUSG00000030341/ENSMUSG00000056501/ENSMUSG00000005087/ENSMUSG0000002
0000031548/ENSMUSG00000044857/ENSMUSG00000074934
0000035828/ENSMUSG00000035621

0000033717/ENSMUSG00000074934

0000034926/ENSMUSG00000032018/ENSMUSG00000033105/ENSMUSG00000050697
0000032115/ENSMUSG00000031778/ENSMUSG00000020423/ENSMUSG00000024789/ENSMUSG00000030341/ENSMUSG00000056501/ENSMUSG00000023951/ENSMUSG00000061436/ENSMUSG0000002
0000031714/ENSMUSG00000056501/ENSMUSG00000035247/ENSMUSG00000045216/ENSMUSG00000074221/ENSMUSG00000024073/ENSMUSG00000032717/ENSMUSG00000032487
0000048616/ENSMUSG00000007613/ENSMUSG00000033323/ENSMUSG00000022687/ENSMUSG00000022752/ENSMUSG00000026313/ENSMUSG00000024431/ENSMUSG00000074934/ENSMUSG0000005
0000002489/ENSMUSG00000026014/ENSMUSG00000018736/ENSMUSG00000024270/ENSMUSG00000031523/ENSMUSG00000026728/ENSMUSG00000005087/ENSMUSG00000022817/ENSMUSG0000002

0000032883/ENSMUSG00000031668/ENSMUSG00000045294/ENSMUSG00000039004/ENSMUSG00000034354/ENSMUSG00000032715/ENSMUSG00000020889/ENSMUSG00000005533/ENSMUSG0000002

0000035828/ENSMUSG00000035621/ENSMUSG00000027712
0000031778/ENSMUSG00000002489/ENSMUSG00000031714/ENSMUSG00000023034/ENSMUSG00000031451/ENSMUSG00000022425/ENSMUSG00000021508/ENSMUSG00000022651/ENSMUSG0000002
0000037434/ENSMUSG00000048126/ENSMUSG00000005533/ENSMUSG00000023175/ENSMUSG00000026131/ENSMUSG00000039952/ENSMUSG00000000058/ENSMUSG00000047126/ENSMUSG0000002
0000031785/ENSMUSG00000028613/ENSMUSG00000020423/ENSMUSG00000018736/ENSMUSG00000029050/ENSMUSG00000035764/ENSMUSG00000030852/ENSMUSG00000020745/ENSMUSG0000001

0000022708/ENSMUSG00000030747/ENSMUSG00000032349/ENSMUSG00000032487/ENSMUSG00000026623
0000023067/ENSMUSG00000032340/ENSMUSG00000039384/ENSMUSG00000042594/ENSMUSG00000027712
0000032702/ENSMUSG00000000058/ENSMUSG00000038859/ENSMUSG00000044167/ENSMUSG00000020091/ENSMUSG00000042903/ENSMUSG00000050697/ENSMUSG00000026812/ENSMUSG0000000

37902/ENSMUSG00000004207

09511/ENSMUSG00000003814/ENSMUSG00000034297/ENSMUSG00000018965/ENSMUSG00000034201/ENSMUSG00000065954/ENSMUSG00000026826/ENSMUSG00000005886

35764/ENSMUSG00000048616/ENSMUSG00000030852/ENSMUSG00000024098/ENSMUSG00000020745/ENSMUSG00000018476/ENSMUSG00000021379/ENSMUSG00000062232/ENSMUSG00000038539/E

40511/ENSMUSG00000022443/ENSMUSG00000006445/ENSMUSG00000026131/ENSMUSG00000039952/ENSMUSG00000000058/ENSMUSG00000062901/ENSMUSG00000005871/ENSMUSG00000032220

30103/ENSMUSG00000020889/ENSMUSG00000026796/ENSMUSG00000037447/ENSMUSG00000051495/ENSMUSG00000022096/ENSMUSG00000048490/ENSMUSG00000038025/ENSMUSG00000027177/E

24425/ENSMUSG00000028277/ENSMUSG00000035621/ENSMUSG00000027712
79037/ENSMUSG00000001910/ENSMUSG00000022365/ENSMUSG00000022037/ENSMUSG00000033454/ENSMUSG00000029249/ENSMUSG00000028179/ENSMUSG00000024935/ENSMUSG00000025209/E
40945/ENSMUSG00000062232/ENSMUSG00000000058/ENSMUSG00000020130/ENSMUSG00000042303/ENSMUSG00000039844/ENSMUSG00000031453/ENSMUSG00000026812/ENSMUSG00000035007/E
49792/ENSMUSG00000005125/ENSMUSG00000031788/ENSMUSG00000014867/ENSMUSG00000030059/ENSMUSG00000020745/ENSMUSG00000096054/ENSMUSG00000027465/ENSMUSG00000022443/E

32458/ENSMUSG00000058152/ENSMUSG00000029502/ENSMUSG00000024085/ENSMUSG00000027465/ENSMUSG00000021770/ENSMUSG00000020346/ENSMUSG00000003746/ENSMUSG00000020132/E
25340/ENSMUSG00000024927/ENSMUSG00000039384/ENSMUSG00000035673/ENSMUSG00000037820/ENSMUSG00000050965/ENSMUSG00000032487/ENSMUSG00000016024/ENSMUSG00000035356/E

22270/ENSMUSG00000063142/ENSMUSG00000026313/ENSMUSG00000079037/ENSMUSG00000027665/ENSMUSG00000032350/ENSMUSG00000029361/ENSMUSG00000024431/ENSMUSG00000026797/E

50697/ENSMUSG00000005871
20745/ENSMUSG00000037815/ENSMUSG00000013076/ENSMUSG00000022443/ENSMUSG00000006445/ENSMUSG00000040852/ENSMUSG00000032702/ENSMUSG00000040268/ENSMUSG00000039952/E

22708/ENSMUSG00000021770/ENSMUSG00000030747/ENSMUSG00000032349/ENSMUSG00000063142/ENSMUSG00000030527/ENSMUSG00000032487/ENSMUSG00000033717/ENSMUSG00000029249/E

23951/ENSMUSG00000044337/ENSMUSG00000020682/ENSMUSG00000006445/ENSMUSG00000050965/ENSMUSG00000024486/ENSMUSG00000003814/ENSMUSG00000016024/ENSMUSG00000056071/E
29361/ENSMUSG00000001119/ENSMUSG00000048920/ENSMUSG00000027712/ENSMUSG00000031543
18476/ENSMUSG00000021379/ENSMUSG00000038539/ENSMUSG00000065954/ENSMUSG00000026812

05871

ENSMUSG00000065954/ENSMUSG00000026826/ENSMUSG00000026812

ENSMUSG00000054808/ENSMUSG00000061436/ENSMUSG00000045482/ENSMUSG00000055491/ENSMUSG00000026313/ENSMUSG00000034297/ENSMUSG00000059288/ENSMUSG00000024052/ENSMUS

ENSMUSG00000002083/ENSMUSG00000024182/ENSMUSG00000027712/ENSMUSG00000026608/ENSMUSG00000005871
ENSMUSG00000039318/ENSMUSG00000002365
ENSMUSG00000030095/ENSMUSG00000048661/ENSMUSG00000009907/ENSMUSG00000026924/ENSMUSG00000056050/ENSMUSG00000022037/ENSMUSG00000000058/ENSMUSG00000041570/ENSMUS

ENSMUSG00000024687/ENSMUSG00000025484/ENSMUSG00000024425/ENSMUSG00000039357
ENSMUSG00000056071/ENSMUSG00000024425/ENSMUSG00000037902/ENSMUSG00000022808

ENSMUSG00000031393/ENSMUSG00000026826/ENSMUSG00000002083

ENSMUSG00000027665/ENSMUSG00000044147/ENSMUSG00000019943/ENSMUSG00000026812/ENSMUSG00000031543/ENSMUSG00000005871/ENSMUSG00000034593

ENSMUSG00000026623/ENSMUSG00000024431/ENSMUSG00000028661/ENSMUSG00000050697/ENSMUSG00000004207

ENSMUSG00000074934

G00000026604/ENSMUSG00000031540/ENSMUSG00000024431/ENSMUSG00000063382/ENSMUSG00000025326/ENSMUSG00000005886/ENSMUSG00000031386

G00000038859/ENSMUSG00000047126/ENSMUSG00000073639/ENSMUSG00000056071/ENSMUSG00000044857/ENSMUSG00000025372/ENSMUSG00000004207/ENSMUSG00000039318/ENSMUSG0000[illegible]

)034951/ENSMUSG00000034216/ENSMUSG00000002365

| Category | GOID | Description | GeneRatio | BgRatio | pvalue | padj | geneID | geneName |
|---|---|---|---|---|---|---|---|---|
| BP | GO:00426[illegible] | cholesterol homeostasis | 14\|1209 | 72\|16097 | 0.00082 | 0.00877 | ENSMUS | Ldlr/Lpl/Nfe2l1/Srebf2/Insig1/Xbp1/Npc1/Fabp4/Nr1d1/Nus1/[illegible] |
| BP | GO:00703[illegible] | negative regulation of ERK1 and ERK2 cascade | 14\|1209 | 72\|16097 | 0.00082 | 0.00877 | ENSMUS | Errfi1/Spry2/Klf4/Dusp1/Spred1/Xbp1/Igf1/Dusp10/Timp3/Cnk[illegible] |
| BP | GO:00016[illegible] | eye development | 42\|1209 | 337\|1609[illegible] | 0.00082 | 0.00877 | ENSMUS | Col8a1/Col4a1/Sox9/Cdon/Klf4/Hipk1/Inhba/Col5a2/Crybg3/C[illegible] |
| BP | GO:00550[illegible] | lipid homeostasis | 20\|1209 | 123\|1609[illegible] | 0.00082 | 0.00877 | ENSMUS | Ldlr/Lpl/Rtn4/Nfe2l1/Srebf2/Sesn2/Insig1/Xbp1/C1qtnf3/Npc1,[illegible] |
| BP | GO:00073[illegible] | regulation of mitotic cell cycle | 55\|1209 | 474\|1609[illegible] | 0.00082 | 0.00877 | ENSMUS | Hspa1b/Hspa1a/Myc/Klf4/Gadd45a/Fgfr1/Met/Cdkn1c/Eif4ebp[illegible] |
| BP | GO:00308[illegible] | mammary gland development | 21\|1209 | 132\|1609[illegible] | 0.00082 | 0.00877 | ENSMUS | Igfbp5/Sox9/Rtn4/Ncoa3/Eif2ak3/Fgfr2/Fasn/Xbp1/Jak2/Igf1r/C[illegible] |
| BP | GO:00605[illegible] | developmental growth involved in morphogenesis | 34\|1209 | 256\|1609[illegible] | 0.00083 | 0.00877 | ENSMUS | Spry2/Sox9/Postn/Ifrd1/Tnfrsf12a/Rtn4/Iqgap1/Golga4/Fgfr2/T[illegible] |
| BP | GO:00031[illegible] | aortic valve development | 7\|1209 | 22\|16097 | 0.00083 | 0.00877 | ENSMUS | Sox9/Efna1/Tnfrsf1a/Robo1/Nfatc1/Notch1/Robo2 |
| BP | GO:00303[illegible] | adrenal gland development | 7\|1209 | 22\|16097 | 0.00083 | 0.00877 | ENSMUS | Cdkn1c/Hmga2/Igf1r/Pbx1/Crhr1/Ly6e/Nr3c1 |
| BP | GO:00901[illegible] | positive regulation of branching involved in ureteric bud morp | 7\|1209 | 22\|16097 | 0.00083 | 0.00877 | ENSMUS | Sox9/Agtr2/Maged1/Nog/Vegfa/Grem1/Sox8 |
| MF | GO:00192[illegible] | kinase regulator activity | 26\|1190 | 179\|1591[illegible] | 0.00083 | 0.02272 | ENSMUS | Spry2/Trib1/Klf4/Cdkn1c/Iqgap1/Cdkn1a/Spred1/Socs3/Ibtk/G[illegible] |
| BP | GO:00457[illegible] | positive regulation of ossification | 17\|1209 | 97\|16097 | 0.00083 | 0.00881 | ENSMUS | Cyr61/Lmna/Smad5/Bmp6/Il6st/Sfrp2/Cebpb/Igf1/Vegfa/Mia3/[illegible] |
| BP | GO:00512[illegible] | negative regulation of protein transport | 27\|1209 | 188\|1609[illegible] | 0.00085 | 0.00893 | ENSMUS | Hmgcr/Cyp51/Insig1/Wwp2/Cd34/Cx3cl1/Ppp2ca/C1qtnf3/Ne[illegible] |
| MF | GO:00167[illegible] | oxidoreductase activity, acting on paired donors, with incorpo | 20\|1190 | 124\|1591[illegible] | 0.00086 | 0.02297 | ENSMUS | Hmox1/Moxd1/Cyp51/Scd1/Msmo1/Tet3/Sqle/Scd2/P3h1/P3h[illegible] |
| BP | GO:00018[illegible] | embryonic placenta development | 18\|1209 | 106\|1609[illegible] | 0.00087 | 0.00918 | ENSMUS | Xist/Cdkn1c/Ncoa3/Cyr61/Fgfr2/Nrk/Socs3/Egfr/Gab1/Cebpb/H[illegible] |
| BP | GO:00217[illegible] | cerebral cortex radially oriented cell migration | 9\|1209 | 35\|16097 | 0.00087 | 0.00918 | ENSMUS | Lamb1/Rtn4/Dixdc1/Adgrg1/Lrp8/Ndel1/Fbxo45/Pafah1b1/Srg[illegible] |
| BP | GO:00507[illegible] | regulation of axonogenesis | 28\|1209 | 198\|1609[illegible] | 0.00088 | 0.00919 | ENSMUS | Ifrd1/Tnfrsf12a/Efna1/Rtn4/Golga4/Dixdc1/Zswim4/Thy1/Tiam[illegible] |
| BP | GO:00901[illegible] | positive regulation of kidney development | 10\|1209 | 42\|16097 | 0.00088 | 0.00919 | ENSMUS | Myc/Sox9/Egr1/Il6ra/Agtr2/Maged1/Nog/Vegfa/Grem1/Sox8 |
| BP | GO:19035[illegible] | negative regulation of response to endoplasmic reticulum stres | 10\|1209 | 42\|16097 | 0.00088 | 0.00919 | ENSMUS | Herpud1/Ppp1r15b/Hyou1/Xbp1/Crebrf/Clu/Creb3l1/Pdia6/Ub[illegible] |
| BP | GO:00454[illegible] | myoblast differentiation | 16\|1209 | 89\|16097 | 0.00088 | 0.00919 | ENSMUS | Sox9/Cdon/Igfbp3/Lgals1/Ddit3/Maml1/Cxcl14/Igf1/Boc/Notch[illegible] |
| BP | GO:00100[illegible] | response to metal ion | 33\|1209 | 247\|1609[illegible] | 0.00088 | 0.00919 | ENSMUS | Mt1/Thbs1/Hmox1/Jun/Iqgap1/Nfe2l1/Fos/Itpr3/Bmp6/Mylk/E[illegible] |
| BP | GO:00485[illegible] | rhythmic process | 34\|1209 | 257\|1609[illegible] | 0.00089 | 0.0092 | ENSMUS | Inhba/Egr1/Atf4/Noct/Egfr/Mybbp1a/Fbxw11/Klf10/Klf9/Sik1/[illegible] |
| BP | GO:00434[illegible] | negative regulation of DNA-binding transcription factor activit | 23\|1209 | 151\|1609[illegible] | 0.00089 | 0.0092 | ENSMUS | Hmox1/Trib1/Klf4/Ddit3/Wwp2/Cebpg/Trib2/Sik1/Gas6/Bhlhe[illegible] |
| BP | GO:00316[illegible] | response to nutrient levels | 40\|1209 | 318\|1609[illegible] | 0.0009 | 0.00932 | ENSMUS | Ldlr/Postn/Plec/Lpl/Slc2a1/Atf4/Jun/Aacs/Eif4ebp1/Larp1/Cdk[illegible] |
| BP | GO:00435[illegible] | negative regulation of neuron apoptotic process | 25\|1209 | 170\|1609[illegible] | 0.00092 | 0.00947 | ENSMUS | Mt1/Hmox1/Jun/Aars/Hyou1/Cx3cl1/Hsph1/Btg2/Jak2/Nrp1/C[illegible] |
| BP | GO:00723[illegible] | monocarboxylic acid biosynthetic process | 31\|1209 | 228\|1609[illegible] | 0.00093 | 0.00961 | ENSMUS | Myc/Acly/Stard4/Lpl/Scd1/Ier3/Acsl3/Scd2/Insig1/Stat3/Fasn/T[illegible] |
| BP | GO:00075[illegible] | sex differentiation | 32\|1209 | 238\|1609[illegible] | 0.00094 | 0.00961 | ENSMUS | Sox9/Serpine1/Inhba/Ncoa3/Smad5/Adgrg1/Hmga2/Bmp6/Dhc[illegible] |
| BP | GO:00513[illegible] | negative regulation of transferase activity | 32\|1209 | 238\|1609[illegible] | 0.00094 | 0.00961 | ENSMUS | Errfi1/Spry2/Hmgcr/Rgs2/Trib1/Gadd45a/Dusp1/Cdkn1a/Spred[illegible] |
| BP | GO:00550[illegible] | regulation of cardiac muscle tissue growth | 14\|1209 | 73\|16097 | 0.00094 | 0.00961 | ENSMUS | Col14a1/Rgs2/Fgfr1/Fgfr2/Tgfbr3/Nog/Tgfbr1/Ctdp1/Igf1/Tomm[illegible] |
| BP | GO:00550[illegible] | sterol homeostasis | 14\|1209 | 73\|16097 | 0.00094 | 0.00961 | ENSMUS | Ldlr/Lpl/Nfe2l1/Srebf2/Insig1/Xbp1/Npc1/Fabp4/Nr1d1/Nus1/[illegible] |
| BP | GO:00060[illegible] | alcohol metabolic process | 36\|1209 | 278\|1609[illegible] | 0.00094 | 0.00961 | ENSMUS | Hmgcr/Ldlr/Moxd1/Cyp51/Fgfr1/Fdft1/Nfe2l1/Srebf2/Hmgcs1/[illegible] |
| BP | GO:00486[illegible] | regulation of muscle organ development | 24\|1209 | 161\|1609[illegible] | 0.00095 | 0.00961 | ENSMUS | Hmgcr/Col14a1/Rgs2/Cdon/Fgfr1/Fgfr2/Tgfbr3/S100b/Nog/Tgf[illegible] |
| BP | GO:00316[illegible] | regulation of protein stability | 35\|1209 | 268\|1609[illegible] | 0.00095 | 0.00961 | ENSMUS | Ddi2/Efna1/Rtn4/Lmna/Usp36/P3h1/B4galt5/Fbxw11/Xbp1/Nr[illegible] |
| BP | GO:00310[illegible] | actomyosin structure organization | 28\|1209 | 199\|1609[illegible] | 0.00095 | 0.00961 | ENSMUS | Acta1/Eln/S100a10/Met/Tpm1/Epb41l1/Synpo/Mef2a/Ctgf/Nrp[illegible] |
| BP | GO:00313[illegible] | negative regulation of cell projection organization | 28\|1209 | 199\|1609[illegible] | 0.00095 | 0.00961 | ENSMUS | Spry2/Xylt1/Ifrd1/Efna1/Rtn4/Lgals1/Ngef/Thy1/Nrp1/Neo1/Se[illegible] |
| BP | GO:00032[illegible] | cardiac chamber development | 26\|1209 | 180\|1609[illegible] | 0.00096 | 0.00969 | ENSMUS | Cyr61/Tpm1/Fgfr2/Rxra/Maml1/Tgfbr3/Nrp1/Sfrp2/Ltbp1/Nog/[illegible] |
| BP | GO:00219[illegible] | central nervous system neuron differentiation | 26\|1209 | 180\|1609[illegible] | 0.00096 | 0.00969 | ENSMUS | Inhba/Fgfr1/Pex5/Nfe2l1/Cdh11/Fgfr2/B4galt5/Btg2/Ndel1/Nrp[illegible] |
| BP | GO:01500[illegible] | visual system development | 42\|1209 | 340\|1609[illegible] | 0.00098 | 0.00986 | ENSMUS | Col8a1/Col4a1/Sox9/Cdon/Klf4/Hipk1/Inhba/Col5a2/Crybg3/C[illegible] |
| BP | GO:19029[illegible] | negative regulation of supramolecular fiber organization | 22\|1209 | 143\|1609[illegible] | 0.00099 | 0.00999 | ENSMUS | Ldlr/Tlr2/Met/Cdh5/Pecam1/Tpm1/Sgk1/Dlc1/Chadl/Tmsb10/[illegible] |
| CC | GO:00988[illegible] | endoplasmic reticulum subcompartment | 56\|1232 | 486\|1631[illegible] | 0.001 | 0.01987 | ENSMUS | Hmgcr/Sgms2/Moxd1/Herpud1/Scd1/Fdft1/Msmo1/Rtn4/Nfe2l[illegible] |
| BP | GO:00061[illegible] | regulation of carbohydrate metabolic process | 25\|1209 | 171\|1609[illegible] | 0.001 | 0.01002 | ENSMUS | Myc/Igfbp3/Has2/Nfe2l1/Ier3/Igfbp4/Sesn2/Scarb2/Stat3/C1qt[illegible] |
| BP | GO:00192[illegible] | regulation of steroid metabolic process | 16\|1209 | 90\|16097 | 0.001 | 0.01002 | ENSMUS | Ldlr/Egr1/Stard4/Scd1/Atp1a1/Insig1/Bmp6/Nr1d1/Igf1r/Igf1/[illegible] |
| BP | GO:00329[illegible] | collagen biosynthetic process | 11\|1209 | 50\|16097 | 0.001 | 0.01002 | ENSMUS | Errfi1/Col1a1/Serpine1/Il6ra/P3h4/Ctgf/Tram2/Ltbp1/Vim/Cre[illegible] |
| BP | GO:00509[illegible] | negative regulation of chemotaxis | 11\|1209 | 50\|16097 | 0.001 | 0.01002 | ENSMUS | Thbs1/Dusp1/Nrp1/Sema3b/Robo1/Mmp28/Notch1/Robo2/N[illegible] |
| BP | GO:00032[illegible] | cardiac chamber morphogenesis | 21\|1209 | 134\|1609[illegible] | 0.001 | 0.01002 | ENSMUS | Cyr61/Tpm1/Fgfr2/Rxra/Tgfbr3/Nrp1/Sfrp2/Nog/Tgfbr1/Robo1/[illegible] |
| BP | GO:00435[illegible] | negative regulation of blood vessel endothelial cell migration | 8\|1209 | 29\|16097 | 0.00102 | 0.01015 | ENSMUS | Thbs1/Klf4/Spred1/Meox2/Pik3r2/Notch1/Vash1/Mecp2 |
| BP | GO:00313[illegible] | positive regulation of cellular catabolic process | 42\|1209 | 341\|1609[illegible] | 0.00104 | 0.0103 | ENSMUS | Hspa1b/Hspa1a/Hmox1/Ldlr/Myc/Tlr2/Trib1/Herpud1/Zfand2a,[illegible] |
| MF | GO:00171[illegible] | Wnt-protein binding | 9\|1190 | 36\|15912 | 0.00105 | 0.02729 | ENSMUS | Wif1/Met/Sfrp2/Fzd10/Axl/Sfrp4/Fzd9/Sfrp1/Fzd2 |
| BP | GO:00354[illegible] | cargo loading into vesicle | 6\|1209 | 17\|16097 | 0.00107 | 0.01053 | ENSMUS | Insig1/Sec24d/Sec23a/Tbc1d20/Mia3/Sec24a |
| BP | GO:00507[illegible] | negative regulation of dendrite morphogenesis | 6\|1209 | 17\|16097 | 0.00107 | 0.01053 | ENSMUS | Efna1/Ngef/Ppp3ca/Rapgef2/Ywhah/Ube3a |
| BP | GO:00328[illegible] | negative regulation of stress-activated MAPK cascade | 10\|1209 | 43\|16097 | 0.00107 | 0.01053 | ENSMUS | Dusp1/Hsph1/Sfrp2/Dusp10/Pafah1b1/Sfrp4/Hipk3/Foxo1/Sfrp[illegible] |

| Count | Up | Up_Gene_id | Down | Down_Gene_id |
|---|---|---|---|---|
| 14 | 3 | ENSMUSG0 | 11 | ENSMUSG00000032193/ENSMUSG00000038615/ENSMUSG00000022463/ENSMUSG00000045294/ENSMUSG00000020484/ENSMUSG00000024413/ENSMUSG0 |
| 14 | 3 | ENSMUSG0 | 11 | ENSMUSG00000028967/ENSMUSG00000022114/ENSMUSG00000003032/ENSMUSG00000027351/ENSMUSG00000020484/ENSMUSG00000039384/ENSMUSG0 |
| 42 | 18 | ENSMUSG0 | 24 | ENSMUSG00000068196/ENSMUSG00000000567/ENSMUSG00000038119/ENSMUSG00000003032/ENSMUSG00000008730/ENSMUSG00000041324/ENSMUSG0 |
| 20 | 5 | ENSMUSG0 | 15 | ENSMUSG00000032193/ENSMUSG00000020458/ENSMUSG00000038615/ENSMUSG00000022463/ENSMUSG00000028893/ENSMUSG00000045294/ENSMUSG0 |
| 55 | 19 | ENSMUSG0 | 36 | ENSMUSG00000022346/ENSMUSG00000003032/ENSMUSG00000036390/ENSMUSG00000031565/ENSMUSG00000009376/ENSMUSG00000031490/ENSMUSG0 |
| 21 | 5 | ENSMUSG0 | 16 | ENSMUSG00000000567/ENSMUSG00000020458/ENSMUSG00000027678/ENSMUSG00000031668/ENSMUSG00000030849/ENSMUSG00000025153/ENSMUSG0 |
| 34 | 14 | ENSMUSG0 | 20 | ENSMUSG00000022114/ENSMUSG00000000567/ENSMUSG00000001627/ENSMUSG00000023905/ENSMUSG00000020458/ENSMUSG00000030536/ENSMUSG0 |
| 7 | 4 | ENSMUSG0 | 3 | ENSMUSG00000000567/ENSMUSG00000027954/ENSMUSG00000030341 |
| 7 | 3 | ENSMUSG0 | 4 | ENSMUSG00000056758/ENSMUSG00000005533/ENSMUSG00000018634/ENSMUSG00000024431 |
| 7 | 3 | ENSMUSG0 | 4 | ENSMUSG00000000567/ENSMUSG00000048616/ENSMUSG00000023951/ENSMUSG00000074934 |
| 26 | 7 | ENSMUSG0 | 19 | ENSMUSG00000022114/ENSMUSG00000032501/ENSMUSG00000003032/ENSMUSG00000030536/ENSMUSG00000023067/ENSMUSG00000027351/ENSMUSG0 |
| 17 | 7 | ENSMUSG0 | 10 | ENSMUSG00000028063/ENSMUSG00000021540/ENSMUSG00000039004/ENSMUSG00000021756/ENSMUSG00000027996/ENSMUSG00000056501/ENSMUSG0 |
| 27 | 9 | ENSMUSG0 | 18 | ENSMUSG00000021670/ENSMUSG00000001467/ENSMUSG00000045294/ENSMUSG00000031778/ENSMUSG00000020349/ENSMUSG00000032340/ENSMUSG0 |
| 20 | 2 | ENSMUSG0 | 18 | ENSMUSG00000005413/ENSMUSG00000020000/ENSMUSG00000001467/ENSMUSG00000037071/ENSMUSG00000031604/ENSMUSG00000034832/ENSMUSG0 |
| 18 | 9 | ENSMUSG0 | 9 | ENSMUSG00000027678/ENSMUSG00000030849/ENSMUSG00000031714/ENSMUSG00000056501/ENSMUSG00000035247/ENSMUSG00000045216/ENSMUSG0 |
| 9 | 2 | ENSMUSG0 | 7 | ENSMUSG00000020458/ENSMUSG00000032064/ENSMUSG00000031785/ENSMUSG00000028613/ENSMUSG00000018736/ENSMUSG00000035764/ENSMUSG0 |
| 28 | 13 | ENSMUSG0 | 15 | ENSMUSG00000001627/ENSMUSG00000023905/ENSMUSG00000027954/ENSMUSG00000020458/ENSMUSG00000038708/ENSMUSG00000032064/ENSMUSG0 |
| 10 | 3 | ENSMUSG0 | 7 | ENSMUSG00000022346/ENSMUSG00000000567/ENSMUSG00000038418/ENSMUSG00000027947/ENSMUSG00000048616/ENSMUSG00000023951/ENSMUSG0 |
| 10 | 0 | | 10 | ENSMUSG00000031770/ENSMUSG00000046062/ENSMUSG00000032115/ENSMUSG00000020484/ENSMUSG00000048249/ENSMUSG00000022037/ENSMUSG0 |
| 16 | 7 | ENSMUSG0 | 9 | ENSMUSG00000000567/ENSMUSG00000038119/ENSMUSG00000025408/ENSMUSG00000050567/ENSMUSG00000021508/ENSMUSG00000022687/ENSMUSG0 |
| 33 | 9 | ENSMUSG0 | 24 | ENSMUSG00000031765/ENSMUSG00000040152/ENSMUSG00000005413/ENSMUSG00000030536/ENSMUSG00000038615/ENSMUSG00000042644/ENSMUSG0 |
| 34 | 10 | ENSMUSG0 | 24 | ENSMUSG00000041324/ENSMUSG00000038418/ENSMUSG00000042406/ENSMUSG00000023087/ENSMUSG00000040463/ENSMUSG00000020271/ENSMUSG0 |
| 23 | 9 | ENSMUSG0 | 14 | ENSMUSG00000005413/ENSMUSG00000032501/ENSMUSG00000003032/ENSMUSG00000025408/ENSMUSG00000056216/ENSMUSG00000020601/ENSMUSG0 |
| 40 | 11 | ENSMUSG0 | 29 | ENSMUSG00000032193/ENSMUSG00000022565/ENSMUSG00000028645/ENSMUSG00000042406/ENSMUSG00000029482/ENSMUSG00000031490/ENSMUSG0 |
| 25 | 8 | ENSMUSG0 | 17 | ENSMUSG00000031765/ENSMUSG00000005413/ENSMUSG00000031960/ENSMUSG00000032115/ENSMUSG00000031778/ENSMUSG00000020423/ENSMUSG0 |
| 31 | 5 | ENSMUSG0 | 26 | ENSMUSG00000022346/ENSMUSG00000020917/ENSMUSG00000024378/ENSMUSG00000037071/ENSMUSG00000003541/ENSMUSG00000032883/ENSMUSG0 |
| 32 | 8 | ENSMUSG0 | 24 | ENSMUSG00000000567/ENSMUSG00000037411/ENSMUSG00000041324/ENSMUSG00000027678/ENSMUSG00000021540/ENSMUSG00000031785/ENSMUSG0 |
| 32 | 7 | ENSMUSG0 | 25 | ENSMUSG00000028967/ENSMUSG00000022114/ENSMUSG00000021670/ENSMUSG00000026360/ENSMUSG00000032501/ENSMUSG00000036390/ENSMUSG0 |
| 14 | 4 | ENSMUSG0 | 10 | ENSMUSG00000026360/ENSMUSG00000031565/ENSMUSG00000030849/ENSMUSG00000029287/ENSMUSG00000048616/ENSMUSG00000007613/ENSMUSG0 |
| 14 | 3 | ENSMUSG0 | 11 | ENSMUSG00000032193/ENSMUSG00000038615/ENSMUSG00000022463/ENSMUSG00000045294/ENSMUSG00000020484/ENSMUSG00000024413/ENSMUSG0 |
| 36 | 7 | ENSMUSG0 | 29 | ENSMUSG00000021670/ENSMUSG00000032193/ENSMUSG00000020000/ENSMUSG00000001467/ENSMUSG00000031565/ENSMUSG00000021273/ENSMUSG0 |
| 24 | 8 | ENSMUSG0 | 16 | ENSMUSG00000021670/ENSMUSG00000026360/ENSMUSG00000038119/ENSMUSG00000031565/ENSMUSG00000030849/ENSMUSG00000029287/ENSMUSG0 |
| 35 | 5 | ENSMUSG0 | 30 | ENSMUSG00000078515/ENSMUSG00000027954/ENSMUSG00000020458/ENSMUSG00000028063/ENSMUSG00000033909/ENSMUSG00000028641/ENSMUSG0 |
| 28 | 17 | ENSMUSG0 | 11 | ENSMUSG00000029675/ENSMUSG00000041959/ENSMUSG00000009376/ENSMUSG00000027624/ENSMUSG00000030557/ENSMUSG00000019997/ENSMUSG0 |
| 28 | 8 | ENSMUSG0 | 20 | ENSMUSG00000022114/ENSMUSG00000030657/ENSMUSG00000001627/ENSMUSG00000027954/ENSMUSG00000020458/ENSMUSG00000026259/ENSMUSG0 |
| 26 | 17 | ENSMUSG0 | 9 | ENSMUSG00000030849/ENSMUSG00000015846/ENSMUSG00000050567/ENSMUSG00000029287/ENSMUSG00000027996/ENSMUSG00000048616/ENSMUSG0 |
| 26 | 9 | ENSMUSG0 | 17 | ENSMUSG00000041324/ENSMUSG00000031565/ENSMUSG00000005069/ENSMUSG00000038615/ENSMUSG00000030849/ENSMUSG00000017929/ENSMUSG0 |
| 42 | 18 | ENSMUSG0 | 24 | ENSMUSG00000068196/ENSMUSG00000000567/ENSMUSG00000038119/ENSMUSG00000003032/ENSMUSG00000008730/ENSMUSG00000041324/ENSMUSG0 |
| 22 | 11 | ENSMUSG0 | 11 | ENSMUSG00000032193/ENSMUSG00000027995/ENSMUSG00000009376/ENSMUSG00000019970/ENSMUSG00000031523/ENSMUSG00000040852/ENSMUSG0 |
| 56 | 6 | ENSMUSG0 | 50 | ENSMUSG00000021670/ENSMUSG00000050931/ENSMUSG00000020000/ENSMUSG00000031770/ENSMUSG00000037071/ENSMUSG00000021273/ENSMUSG0 |
| 25 | 8 | ENSMUSG0 | 17 | ENSMUSG00000022346/ENSMUSG00000022367/ENSMUSG00000038615/ENSMUSG00000003541/ENSMUSG00000028893/ENSMUSG00000029426/ENSMUSG0 |
| 16 | 3 | ENSMUSG0 | 13 | ENSMUSG00000032193/ENSMUSG00000038418/ENSMUSG00000024378/ENSMUSG00000037071/ENSMUSG00000033161/ENSMUSG00000045294/ENSMUSG0 |
| 11 | 3 | ENSMUSG0 | 8 | ENSMUSG00000028967/ENSMUSG00000037411/ENSMUSG00000027947/ENSMUSG00000006931/ENSMUSG00000019997/ENSMUSG00000041779/ENSMUSG0 |
| 11 | 8 | ENSMUSG0 | 3 | ENSMUSG00000040152/ENSMUSG00000020682/ENSMUSG00000074934 |
| 21 | 14 | ENSMUSG0 | 7 | ENSMUSG00000030849/ENSMUSG00000015846/ENSMUSG00000029287/ENSMUSG00000027996/ENSMUSG00000048616/ENSMUSG00000007613/ENSMUSG0 |
| 8 | 4 | ENSMUSG0 | 4 | ENSMUSG00000040152/ENSMUSG00000003032/ENSMUSG00000027351/ENSMUSG00000031393 |
| 42 | 10 | ENSMUSG0 | 32 | ENSMUSG00000005413/ENSMUSG00000032193/ENSMUSG00000022346/ENSMUSG00000027995/ENSMUSG00000032501/ENSMUSG00000031770/ENSMUSG0 |
| 9 | 4 | ENSMUSG0 | 5 | ENSMUSG00000020218/ENSMUSG00000009376/ENSMUSG00000027996/ENSMUSG00000081683/ENSMUSG00000031548 |
| 6 | 0 | | 6 | ENSMUSG00000045294/ENSMUSG00000039234/ENSMUSG00000020986/ENSMUSG00000027465/ENSMUSG00000056050/ENSMUSG00000036391 |
| 6 | 0 | | 6 | ENSMUSG00000027954/ENSMUSG00000026259/ENSMUSG00000028161/ENSMUSG00000062232/ENSMUSG00000018965/ENSMUSG00000025326 |
| 10 | 3 | ENSMUSG0 | 7 | ENSMUSG00000027996/ENSMUSG00000039384/ENSMUSG00000020745/ENSMUSG00000027177/ENSMUSG00000044167/ENSMUSG00000031548/ENSMUSG0 |

0000020889/ENSMUSG00000023068/ENSMUSG00000030747/ENSMUSG00000034297/ENSMUSG00000036391
0000020044/ENSMUSG00000015202/ENSMUSG00000039844/ENSMUSG00000037902/ENSMUSG00000026628
0000022723/ENSMUSG00000021136/ENSMUSG00000030849/ENSMUSG00000041592/ENSMUSG00000015846/ENSMUSG00000039004/ENSMUSG00000004040/ENSMUSG00000037242/ENSMUSG0000002
0000020484/ENSMUSG00000024413/ENSMUSG00000020889/ENSMUSG00000022708/ENSMUSG00000023068/ENSMUSG00000030747/ENSMUSG00000034297/ENSMUSG00000036391/ENSMUSG0000005
0000030536/ENSMUSG00000023067/ENSMUSG00000038615/ENSMUSG00000003541/ENSMUSG00000030849/ENSMUSG00000078429/ENSMUSG00000020423/ENSMUSG00000029752/ENSMUSG0000002
0000020484/ENSMUSG00000024789/ENSMUSG00000005533/ENSMUSG00000056501/ENSMUSG00000023951/ENSMUSG00000040021/ENSMUSG00000006445/ENSMUSG00000028042/ENSMUSG0000002
0000038708/ENSMUSG00000030849/ENSMUSG00000035342/ENSMUSG00000002489/ENSMUSG00000026014/ENSMUSG00000018736/ENSMUSG00000027996/ENSMUSG00000023951/ENSMUSG0000002

0000035941/ENSMUSG00000046733/ENSMUSG00000032715/ENSMUSG00000049323/ENSMUSG00000020601/ENSMUSG00000031451/ENSMUSG00000011960/ENSMUSG00000036707/ENSMUSG0000002
0000023951/ENSMUSG00000056050/ENSMUSG00000026836/ENSMUSG00000019943
0000031451/ENSMUSG00000025340/ENSMUSG00000028161/ENSMUSG00000033717/ENSMUSG00000029249/ENSMUSG00000029361/ENSMUSG00000031548/ENSMUSG00000035828/ENSMUSG0000002
0000022351/ENSMUSG00000025203/ENSMUSG00000028641/ENSMUSG00000038168/ENSMUSG00000004846/ENSMUSG00000032018/ENSMUSG00000018906/ENSMUSG00000036450/ENSMUSG0000003
0000074221/ENSMUSG00000024073/ENSMUSG00000032717
0000020745
0000035671/ENSMUSG00000002489/ENSMUSG00000018736/ENSMUSG00000026728/ENSMUSG00000023951/ENSMUSG00000020745/ENSMUSG00000036606/ENSMUSG00000060012/ENSMUSG0000002
0000074934
0000027230/ENSMUSG00000020571/ENSMUSG00000028277/ENSMUSG00000024807
0000036450/ENSMUSG00000029249/ENSMUSG00000074934
0000039004/ENSMUSG00000030557/ENSMUSG00000020349/ENSMUSG00000037242/ENSMUSG00000024413/ENSMUSG00000037434/ENSMUSG00000028161/ENSMUSG00000063142/ENSMUSG0000007
0000037465/ENSMUSG00000033863/ENSMUSG00000030103/ENSMUSG00000020889/ENSMUSG00000022425/ENSMUSG00000005533/ENSMUSG00000048490/ENSMUSG00000022124/ENSMUSG0000002
0000031451/ENSMUSG00000030103/ENSMUSG00000005533/ENSMUSG00000022096/ENSMUSG00000026313/ENSMUSG00000079037/ENSMUSG00000034297/ENSMUSG00000031540
0000037331/ENSMUSG00000023067/ENSMUSG00000022463/ENSMUSG00000031668/ENSMUSG00000025408/ENSMUSG00000028893/ENSMUSG00000040463/ENSMUSG00000037465/ENSMUSG0000003
0000024789/ENSMUSG00000056501/ENSMUSG00000023951/ENSMUSG00000061436/ENSMUSG00000022270/ENSMUSG00000027665/ENSMUSG00000032350/ENSMUSG00000029361/ENSMUSG0000002
0000025203/ENSMUSG00000045294/ENSMUSG00000004040/ENSMUSG00000025153/ENSMUSG00000032715/ENSMUSG00000029752/ENSMUSG00000037012/ENSMUSG00000020889/ENSMUSG0000002
0000056758/ENSMUSG00000039004/ENSMUSG00000034926/ENSMUSG00000034640/ENSMUSG00000027996/ENSMUSG00000070645/ENSMUSG00000007613/ENSMUSG00000056501/ENSMUSG0000002
0000023067/ENSMUSG00000027351/ENSMUSG00000046733/ENSMUSG00000032715/ENSMUSG00000049323/ENSMUSG00000034765/ENSMUSG00000027996/ENSMUSG00000049792/ENSMUSG0000000
0000033323/ENSMUSG00000022752/ENSMUSG00000024431/ENSMUSG00000005871
0000020889/ENSMUSG00000023068/ENSMUSG00000030747/ENSMUSG00000034297/ENSMUSG00000036391
0000038615/ENSMUSG00000022463/ENSMUSG00000093930/ENSMUSG00000022351/ENSMUSG00000045294/ENSMUSG00000039004/ENSMUSG00000026675/ENSMUSG00000089774/ENSMUSG0000003
0000048616/ENSMUSG00000007613/ENSMUSG00000033323/ENSMUSG00000022687/ENSMUSG00000022752/ENSMUSG00000026313/ENSMUSG00000024431/ENSMUSG00000074934/ENSMUSG0000005
0000017929/ENSMUSG00000020271/ENSMUSG00000020484/ENSMUSG00000020889/ENSMUSG00000026361/ENSMUSG00000049792/ENSMUSG00000048234/ENSMUSG00000022124/ENSMUSG0000003
0000031523/ENSMUSG00000007613/ENSMUSG00000022817/ENSMUSG00000022443/ENSMUSG00000026812
0000032340/ENSMUSG00000026728/ENSMUSG00000049792/ENSMUSG00000020745/ENSMUSG00000028161/ENSMUSG00000032702/ENSMUSG00000079037/ENSMUSG00000032593/ENSMUSG0000006
0000007613/ENSMUSG00000035247/ENSMUSG00000026836
0000020423/ENSMUSG00000018736/ENSMUSG00000027996/ENSMUSG00000035764/ENSMUSG00000020745/ENSMUSG00000050965/ENSMUSG00000021379/ENSMUSG00000062232/ENSMUSG0000003
0000022723/ENSMUSG00000021136/ENSMUSG00000030849/ENSMUSG00000041592/ENSMUSG00000015846/ENSMUSG00000039004/ENSMUSG00000004040/ENSMUSG00000037242/ENSMUSG0000002
0000032702/ENSMUSG00000022037/ENSMUSG00000041570/ENSMUSG00000034201/ENSMUSG00000005871
0000031604/ENSMUSG00000020458/ENSMUSG00000038615/ENSMUSG00000022463/ENSMUSG00000031668/ENSMUSG00000025203/ENSMUSG00000030062/ENSMUSG00000029467/ENSMUSG0000004
0000004040/ENSMUSG00000020889/ENSMUSG00000022708/ENSMUSG00000030747/ENSMUSG00000026313/ENSMUSG00000047126/ENSMUSG00000044167/ENSMUSG00000024431/ENSMUSG0000005
0000039004/ENSMUSG00000020889/ENSMUSG00000005533/ENSMUSG00000030747/ENSMUSG00000063142/ENSMUSG00000029249/ENSMUSG00000024431
0000026728/ENSMUSG00000027230

0000026836

0000053581/ENSMUSG00000037331/ENSMUSG00000025408/ENSMUSG00000028893/ENSMUSG00000032715/ENSMUSG00000020484/ENSMUSG00000049323/ENSMUSG00000020423/ENSMUSG0000002

0000037902

29050/ENSMUSG00000007613/ENSMUSG00000026728/ENSMUSG00000024098/ENSMUSG00000023951/ENSMUSG00000024085/ENSMUSG00000027465/ENSMUSG00000061436/ENSMUSG00000006445/E
50697
26361/ENSMUSG00000045983/ENSMUSG00000005533/ENSMUSG00000006585/ENSMUSG00000038764/ENSMUSG00000020745/ENSMUSG00000022043/ENSMUSG00000034154/ENSMUSG00000109511/E
24431/ENSMUSG00000085795
20745/ENSMUSG00000074221/ENSMUSG00000024423/ENSMUSG00000020132/ENSMUSG00000031548/ENSMUSG00000031393

22016/ENSMUSG00000027665/ENSMUSG00000000184/ENSMUSG00000074934/ENSMUSG00000005871

24425/ENSMUSG00000028277/ENSMUSG00000035621/ENSMUSG00000027712
38633/ENSMUSG00000032487/ENSMUSG00000029361/ENSMUSG00000046791

20132

79037/ENSMUSG00000025582/ENSMUSG00000034205/ENSMUSG00000015839/ENSMUSG00000013275/ENSMUSG00000031393/ENSMUSG00000021794/ENSMUSG00000009630/ENSMUSG00000050697/E
27832/ENSMUSG00000063142/ENSMUSG00000021379/ENSMUSG00000040268/ENSMUSG00000025261/ENSMUSG00000038539/ENSMUSG00000036432/ENSMUSG00000025326/ENSMUSG00000050697/E

34354/ENSMUSG00000020484/ENSMUSG00000029752/ENSMUSG00000031451/ENSMUSG00000045983/ENSMUSG00000005533/ENSMUSG00000024423/ENSMUSG00000034201/ENSMUSG00000035992/E
26797/ENSMUSG00000031393/ENSMUSG00000026826
22708/ENSMUSG00000039202/ENSMUSG00000004267/ENSMUSG00000032349/ENSMUSG00000026313/ENSMUSG00000038633/ENSMUSG00000032487/ENSMUSG00000039497/ENSMUSG00000063229/E
23951/ENSMUSG00000048490/ENSMUSG00000030059/ENSMUSG00000079509/ENSMUSG00000027832/ENSMUSG00000027465/ENSMUSG00000074656/ENSMUSG00000040268/ENSMUSG00000031548/E
05533/ENSMUSG00000039384/ENSMUSG00000040021/ENSMUSG00000027177/ENSMUSG00000042594/ENSMUSG00000109511/ENSMUSG00000050965/ENSMUSG00000053716/ENSMUSG00000031548/E

34926/ENSMUSG00000024413/ENSMUSG00000023068/ENSMUSG00000032018/ENSMUSG00000030747/ENSMUSG00000063142/ENSMUSG00000033105/ENSMUSG00000024924/ENSMUSG00000003752/E
50697/ENSMUSG00000005871
32431/ENSMUSG00000033105/ENSMUSG00000021149/ENSMUSG00000079037/ENSMUSG00000026924/ENSMUSG00000003814/ENSMUSG00000029447/ENSMUSG00000022365/ENSMUSG00000022037/E

52232/ENSMUSG00000018965/ENSMUSG00000044147/ENSMUSG00000031393/ENSMUSG00000025326/ENSMUSG00000026812

38539/ENSMUSG00000031548/ENSMUSG00000026826
29050/ENSMUSG00000007613/ENSMUSG00000026728/ENSMUSG00000024098/ENSMUSG00000023951/ENSMUSG00000024085/ENSMUSG00000027465/ENSMUSG00000061436/ENSMUSG00000006445/E

42644/ENSMUSG00000045294/ENSMUSG00000025373/ENSMUSG00000020484/ENSMUSG00000014959/ENSMUSG00000047187/ENSMUSG00000030530/ENSMUSG00000079111/ENSMUSG00000039100/E
50697/ENSMUSG00000005886/ENSMUSG00000035621

20601/ENSMUSG00000022708/ENSMUSG00000021036/ENSMUSG00000022037/ENSMUSG00000025234/ENSMUSG00000033717/ENSMUSG00000028793/ENSMUSG00000044167/ENSMUSG00000032350/E

:NSMUSG00000005871

:NSMUSG00000045482/ENSMUSG00000050965/ENSMUSG00000040945/ENSMUSG00000009907/ENSMUSG00000018965/ENSMUSG00000029816/ENSMUSG00000018707/ENSMUSG00000047126/ENSMUS

:NSMUSG00000027712
:NSMUSG00000005886

:NSMUSG00000044167/ENSMUSG00000015839/ENSMUSG00000028646/ENSMUSG00000050697/ENSMUSG00000026812/ENSMUSG00000026628

:NSMUSG00000026623/ENSMUSG00000050697/ENSMUSG00000034593
:NSMUSG00000025326
:NSMUSG00000035621/ENSMUSG00000005871

:NSMUSG00000021036/ENSMUSG00000029249/ENSMUSG00000031835/ENSMUSG00000031393/ENSMUSG00000050697/ENSMUSG00000024210

:NSMUSG00000026797/ENSMUSG00000032612/ENSMUSG00000026812/ENSMUSG00000024807/ENSMUSG00000034951/ENSMUSG00000031386/ENSMUSG00000031438

:NSMUSG00000005871

:NSMUSG00000023068/ENSMUSG00000027465/ENSMUSG00000021770/ENSMUSG00000032018/ENSMUSG00000022270/ENSMUSG00000021831/ENSMUSG00000021427/ENSMUSG00000030747/ENSMUS

:NSMUSG00000028211/ENSMUSG00000040025/ENSMUSG00000038068/ENSMUSG00000009291/ENSMUSG00000050697/ENSMUSG00000026812/ENSMUSG00000024182/ENSMUSG00000039318/ENSMUS

G00000000184/ENSMUSG00000035828/ENSMUSG00000031393/ENSMUSG00000042903/ENSMUSG00000005871

G00000032349/ENSMUSG00000036564/ENSMUSG00000021036/ENSMUSG00000026924/ENSMUSG00000003814/ENSMUSG00000056050/ENSMUSG00000022365/ENSMUSG00000026131/ENSMUSG0000[illegible]

G00000002365

)030835/ENSMUSG00000027230/ENSMUSG00000024052/ENSMUSG00000075703/ENSMUSG00000073639/ENSMUSG00000029361/ENSMUSG00000005615/ENSMUSG00000030287/ENSMUSG0000002480

7/ENSMUSG00000047554/ENSMUSG00000027366

| Category | GOID | Description | GeneRatio | BgRatio | pvalue | padj | geneID | geneName |
|---|---|---|---|---|---|---|---|---|
| BP | GO:00610 | establishment of endothelial barrier | 10\|1209 | 43\|16097 | 0.00107 | 0.01053 | ENSMUS | Cdh5/Rtn4/Fasn/Tnfrsf1a/Vegfa/Tjp2/Cldn5/Akap11/Rapgef2/Ra |
| BP | GO:00703 | negative regulation of stress-activated protein kinase signaling | 10\|1209 | 43\|16097 | 0.00107 | 0.01053 | ENSMUS | Dusp1/Hsph1/Sfrp2/Dusp10/Pafah1b1/Sfrp4/Hipk3/Foxo1/Sfrp |
| BP | GO:00514 | neuron apoptotic process | 36\|1209 | 280\|1609 | 0.00107 | 0.01055 | ENSMUS | Mt1/Hmox1/Egr1/Atf4/Jun/Aars/Ddit3/Hyou1/Cx3cl1/Usp53/H |
| BP | GO:00345 | cellular response to oxidative stress | 34\|1209 | 260\|1609 | 0.00109 | 0.01065 | ENSMUS | Srxn1/Thbs1/Hmox1/Met/Atf4/Jun/Nfe2l1/Fos/Atp2a2/Sesn2/E |
| CC | GO:00701 | anchoring junction | 38\|1232 | 299\|1631 | 0.00109 | 0.02106 | ENSMUS | Plec/Itga5/Cdh5/Iqgap1/Cdh11/Atp2a2/Jak2/Nrp1/Dlc1/Tns1/N |
| BP | GO:00020 | lens development in camera-type eye | 13\|1209 | 66\|16097 | 0.00109 | 0.01069 | ENSMUS | Cdon/Hipk1/Crybg3/Cdkn1c/Fgfr2/Ski/Tgfbr1/Vim/Tbc1d20/Hi |
| BP | GO:00486 | response to axon injury | 13\|1209 | 66\|16097 | 0.00109 | 0.01069 | ENSMUS | Klf4/Xylt1/Jun/Rtn4/Tnc/Ndel1/Jak2/Neo1/Igf1r/Lamb2/Epha4, |
| BP | GO:00717 | basement membrane organization | 7\|1209 | 23\|16097 | 0.00111 | 0.01086 | ENSMUS | Col4a1/Lamb1/Phldb1/Plod3/Lamb2/Dag1/Cma1 |
| BP | GO:01201 | positive regulation of cold-induced thermogenesis | 16\|1209 | 91\|16097 | 0.00113 | 0.011 | ENSMUS | Scd1/Jak2/Fabp4/Cd36/Igf1r/Cebpb/Vegfa/Kdm6b/Zbtb7b/Aca |
| BP | GO:00488 | sensory system development | 42\|1209 | 343\|1609 | 0.00116 | 0.01131 | ENSMUS | Col8a1/Col4a1/Sox9/Cdon/Klf4/Hipk1/Inhba/Col5a2/Crybg3/C |
| MF | GO:00484 | platelet-derived growth factor binding | 5\|1190 | 12\|15912 | 0.00118 | 0.02962 | ENSMUS | Col3a1/Col1a1/Col1a2/Col4a1/Col6a1 |
| BP | GO:00019 | positive regulation of cell-matrix adhesion | 11\|1209 | 51\|16097 | 0.00119 | 0.01152 | ENSMUS | S100a10/Iqgap1/Thy1/Nrp1/Enpp2/Cd36/Vegfa/Tek/Dag1/Kdr/ |
| BP | GO:00426 | ovulation cycle | 11\|1209 | 51\|16097 | 0.00119 | 0.01152 | ENSMUS | Inhba/Egr1/Egfr/Enpp2/Igf1r/Nrip1/Axl/Ptx3/Adamts1/Plekha1/ |
| BP | GO:00903 | positive regulation of wound healing | 11\|1209 | 51\|16097 | 0.00119 | 0.01152 | ENSMUS | Thbs1/Serpine1/Smoc2/Mylk/Xbp1/Cd36/Kank1/Hbegf/Adra2a/ |
| BP | GO:00097 | hormone-mediated signaling pathway | 22\|1209 | 145\|1609 | 0.0012 | 0.01152 | ENSMUS | Ncoa3/Hmga2/Sgk1/Klf9/Jak2/Ren1/Nr1d1/Igf1/Lats1/Abhd2/C |
| BP | GO:00147 | negative regulation of muscle adaptation | 5\|1209 | 12\|16097 | 0.0012 | 0.01152 | ENSMUS | Errfi1/Igfbp5/Klf4/Lmna/Foxo1 |
| BP | GO:00459 | negative regulation of calcium ion-dependent exocytosis | 5\|1209 | 12\|16097 | 0.0012 | 0.01152 | ENSMUS | Notch1/Gnai2/Adra2a/Rest/Cbarp |
| BP | GO:20012 | negative regulation of neuron migration | 5\|1209 | 12\|16097 | 0.0012 | 0.01152 | ENSMUS | Col3a1/Adgrg1/Stat3/Tnn/Srgap2 |
| BP | GO:00604 | heart growth | 18\|1209 | 109\|1609 | 0.00122 | 0.01167 | ENSMUS | Col14a1/Rgs2/Fgfr1/Sorbs2/Fgfr2/Rxra/Tgfbr3/Agtr2/Nog/Tgfbr |
| BP | GO:00359 | aorta development | 12\|1209 | 59\|16097 | 0.00125 | 0.012 | ENSMUS | Col3a1/Mylk/Ltbp1/Lox/Hectd1/Robo1/Plxnd1/Notch1/Robo2 |
| BP | GO:00485 | response to steroid hormone | 27\|1209 | 193\|1609 | 0.00126 | 0.01205 | ENSMUS | Eif4ebp1/Atp1a1/Ncoa3/Rxra/Hmga2/Sgk1/Agtr2/Klf9/Jak2/Np |
| BP | GO:20000 | positive regulation of ubiquitin-dependent protein catabolic p | 16\|1209 | 92\|16097 | 0.00127 | 0.01214 | ENSMUS | Hspa1b/Hspa1a/Trib1/Herpud1/Zfand2a/Trib3/Trib2/Nkd2/Rnf |
| BP | GO:00303 | positive regulation of cell growth | 25\|1209 | 174\|1609 | 0.00128 | 0.01222 | ENSMUS | Tnfrsf12a/Rtn4/Mmp14/Ncoa3/Golga4/Sgk1/Egfr/Ndel1/Nrp1/S |
| BP | GO:00104 | regulation of mesenchymal cell proliferation | 10\|1209 | 44\|16097 | 0.00129 | 0.01228 | ENSMUS | Myc/Sox9/Fgfr1/Lmna/Fgfr2/Foxp2/Vegfa/Syne1/Ptn/Kdr |
| MF | GO:00199 | cytokine binding | 19\|1190 | 119\|1591 | 0.0013 | 0.03067 | ENSMUS | Thbs1/Vasn/A2m/Cd109/Il6ra/Tgfbr3/Il6st/Comp/Ltbp1/Cd36/ |
| BP | GO:00603 | cardiac epithelial to mesenchymal transition | 8\|1209 | 30\|16097 | 0.0013 | 0.01233 | ENSMUS | Has2/Efna1/Rtn4/Tgfbr3/Nog/Tgfbr1/Notch1/Acvr1 |
| MF | GO:00009 | RNA polymerase II core promoter sequence-specific DNA bindir | 9\|1190 | 37\|15912 | 0.0013 | 0.03067 | ENSMUS | Egr1/Gadd45a/Fos/Cebpb/Zfp521/Zfp142/Zfp64/Rest/Sox8 |
| CC | GO:00319 | Golgi cisterna | 12\|1232 | 59\|16313 | 0.00132 | 0.02441 | ENSMUS | Xylt1/Csgalnact2/Chpf/Gorasp2/Csgalnact1/Furin/Gosr1/Chsy3, |
| MF | GO:00012 | DNA-binding transcription repressor activity, RNA polymerase | 29\|1190 | 214\|1591 | 0.00132 | 0.03067 | ENSMUS | Myc/Srebf2/Lrrfip1/Glis3/Bcl6/Btg2/Bhlhe40/Nr1d1/Cebpb/Zbt |
| BP | GO:00550 | cardiac muscle tissue growth | 17\|1209 | 101\|1609 | 0.00133 | 0.01256 | ENSMUS | Col14a1/Rgs2/Fgfr1/Sorbs2/Fgfr2/Rxra/Tgfbr3/Agtr2/Nog/Tgfbr |
| BP | GO:00425 | glucose homeostasis | 31\|1209 | 233\|1609 | 0.00133 | 0.01258 | ENSMUS | Hmgcr/Igfbp5/Met/Aacs/Sesn2/Stat3/Tiam1/Ppp2ca/Xbp1/C1q |
| BP | GO:00486 | regulation of developmental growth | 45\|1209 | 377\|1609 | 0.00134 | 0.01258 | ENSMUS | Col14a1/Rgs2/Serpine1/Ifrd1/Fgfr1/Tnfrsf12a/Pex5/Rtn4/Cdkn1 |
| BP | GO:00082 | regulation of blood pressure | 24\|1209 | 165\|1609 | 0.00134 | 0.01258 | ENSMUS | Col1a2/Hmox1/Postn/Atp1a1/Ier3/Tpm1/Cd34/Agtr2/Kcnk6/G |
| BP | GO:00019 | negative regulation of cell-matrix adhesion | 9\|1209 | 37\|16097 | 0.00134 | 0.01258 | ENSMUS | Thbs1/Postn/Serpine1/Plet1/Mmp14/Bcl6/Dlc1/Enpp2/Rcc2 |
| BP | GO:00305 | positive regulation of BMP signaling pathway | 9\|1209 | 37\|16097 | 0.00134 | 0.01258 | ENSMUS | Cdh5/Cyr61/Tgfbr3/Neo1/Twsg1/Notch1/Kcp/Hes1/Kdr |
| BP | GO:00342 | regulation of cellular amide metabolic process | 43\|1209 | 356\|1609 | 0.00134 | 0.01258 | ENSMUS | Thbs1/Igfbp5/Rgs2/Ppp1r15b/Aars/Eif4ebp1/Efna1/Larp1/Eif2a |
| BP | GO:00085 | regulation of Notch signaling pathway | 15\|1209 | 84\|16097 | 0.00135 | 0.01266 | ENSMUS | Postn/Dlk1/Chac1/Mmp14/Stat3/Bcl6/Il6st/Zmiz1/Robo1/Notc |
| CC | GO:00098 | cytoplasmic side of plasma membrane | 23\|1232 | 155\|1631 | 0.00137 | 0.02441 | ENSMUS | Errfi1/Gem/Rgs2/Ptp4a1/Iqgap1/Atp2a2/S100a6/Tiam1/Ptpn3/ |
| CC | GO:00300 | myofibril | 28\|1232 | 203\|1631 | 0.0014 | 0.02441 | ENSMUS | Acta1/Myl1/Slc2a1/Ahnak/Tpm1/Tpm2/Pde4b/Synpo/Tnni2/Ld |
| MF | GO:00010 | core promoter binding | 12\|1190 | 60\|15912 | 0.00141 | 0.03191 | ENSMUS | Myc/Egr1/Gadd45a/Fos/Klf10/Cebpb/Zfp521/Zfp142/Zfp64/Res |
| BP | GO:00455 | negative regulation of fat cell differentiation | 11\|1209 | 52\|16097 | 0.00141 | 0.01319 | ENSMUS | Dlk1/Ddit3/Insig1/Trib3/Trib2/Vegfa/Id4/Ccdc85b/Foxo1/Enpp |
| BP | GO:00335 | carbohydrate homeostasis | 31\|1209 | 234\|1609 | 0.00143 | 0.01329 | ENSMUS | Hmgcr/Igfbp5/Met/Aacs/Sesn2/Stat3/Tiam1/Ppp2ca/Xbp1/C1q |
| BP | GO:00433 | regulation of protein binding | 31\|1209 | 234\|1609 | 0.00143 | 0.01329 | ENSMUS | Myc/Cdon/Met/Cald1/Lgals3/Eif2ak3/Abl2/Tgfbr3/Tiam1/Ppp2 |
| CC | GO:00057 | endoplasmic reticulum membrane | 54\|1232 | 472\|1631 | 0.00143 | 0.02441 | ENSMUS | Hmgcr/Sgms2/Moxd1/Herpud1/Scd1/Fdft1/Msmo1/Rtn4/Nfe2l |
| BP | GO:00074 | neuroblast proliferation | 12\|1209 | 60\|16097 | 0.00146 | 0.01351 | ENSMUS | Fgfr1/Fgfr2/Cx3cl1/Vegfa/Pafah1b1/Notch1/Ctnna1/Plxnb2/Id4 |
| BP | GO:00181 | protein hydroxylation | 7\|1209 | 24\|16097 | 0.00147 | 0.01351 | ENSMUS | P3h1/P3h4/P3h2/Plod3/Crtap/P4ha2/Hif1an |
| BP | GO:00612 | positive regulation of mesonephros development | 7\|1209 | 24\|16097 | 0.00147 | 0.01351 | ENSMUS | Sox9/Agtr2/Maged1/Nog/Vegfa/Grem1/Sox8 |
| BP | GO:00614 | renal system vasculature development | 7\|1209 | 24\|16097 | 0.00147 | 0.01351 | ENSMUS | Egr1/Il6ra/Nrp1/Notch3/Notch1/Tek/Hes1 |
| BP | GO:00614 | kidney vasculature development | 7\|1209 | 24\|16097 | 0.00147 | 0.01351 | ENSMUS | Egr1/Il6ra/Nrp1/Notch3/Notch1/Tek/Hes1 |
| BP | GO:00701 | negative regulation of biomineral tissue development | 7\|1209 | 24\|16097 | 0.00147 | 0.01351 | ENSMUS | Sox9/Aspn/Rflna/Gas6/Notch1/Enpp1/Grem1 |
| BP | GO:19058 | negative regulation of synapse organization | 7\|1209 | 24\|16097 | 0.00147 | 0.01351 | ENSMUS | Tlr2/Efna1/Ngef/Prnp/Fzd9/Ptn/Ube3a |

| Count | Up | Up_Gene_i | Down | Down_Gene_id |
|---|---|---|---|---|
| 10 | 2 | ENSMUSG0 | 8 | ENSMUSG00000020458/ENSMUSG00000025153/ENSMUSG00000030341/ENSMUSG00000023951/ENSMUSG00000024812/ENSMUSG00000022016/ENSMUSG0 |
| 10 | 3 | ENSMUSG0 | 7 | ENSMUSG00000027996/ENSMUSG00000039384/ENSMUSG00000020745/ENSMUSG00000027177/ENSMUSG00000044167/ENSMUSG00000031548/ENSMUSG0 |
| 36 | 9 | ENSMUSG0 | 27 | ENSMUSG00000031765/ENSMUSG00000005413/ENSMUSG00000038418/ENSMUSG00000042406/ENSMUSG00000031960/ENSMUSG00000025408/ENSMUSG0 |
| 34 | 9 | ENSMUSG0 | 25 | ENSMUSG00000032802/ENSMUSG00000040152/ENSMUSG00000005413/ENSMUSG00000009376/ENSMUSG00000042406/ENSMUSG00000038615/ENSMUSG0 |
| 38 | 15 | ENSMUSG0 | 23 | ENSMUSG00000022565/ENSMUSG00000000555/ENSMUSG00000030536/ENSMUSG00000029467/ENSMUSG00000024789/ENSMUSG00000031523/ENSMUSG0 |
| 13 | 3 | ENSMUSG0 | 10 | ENSMUSG00000038119/ENSMUSG00000008730/ENSMUSG00000022723/ENSMUSG00000030849/ENSMUSG00000029050/ENSMUSG00000007613/ENSMUSG0 |
| 13 | 6 | ENSMUSG0 | 7 | ENSMUSG00000003032/ENSMUSG00000030657/ENSMUSG00000020458/ENSMUSG00000018736/ENSMUSG00000024789/ENSMUSG00000032340/ENSMUSG0 |
| 7 | 4 | ENSMUSG0 | 3 | ENSMUSG00000048537/ENSMUSG00000004846/ENSMUSG00000039952 |
| 16 | 6 | ENSMUSG0 | 10 | ENSMUSG00000037071/ENSMUSG00000024789/ENSMUSG00000005533/ENSMUSG00000056501/ENSMUSG00000023951/ENSMUSG00000018476/ENSMUSG0 |
| 42 | 18 | ENSMUSG0 | 24 | ENSMUSG00000068196/ENSMUSG00000000567/ENSMUSG00000038119/ENSMUSG00000003032/ENSMUSG00000008730/ENSMUSG00000041324/ENSMUSG0 |
| 5 | 4 | ENSMUSG0 | 1 | ENSMUSG00000001119 |
| 11 | 5 | ENSMUSG0 | 6 | ENSMUSG00000041959/ENSMUSG00000030536/ENSMUSG00000022425/ENSMUSG00000023951/ENSMUSG00000039952/ENSMUSG00000026812 |
| 11 | 4 | ENSMUSG0 | 7 | ENSMUSG00000041324/ENSMUSG00000038418/ENSMUSG00000022425/ENSMUSG00000005533/ENSMUSG00000048490/ENSMUSG00000027832/ENSMUSG0 |
| 11 | 3 | ENSMUSG0 | 8 | ENSMUSG00000040152/ENSMUSG00000037411/ENSMUSG00000020484/ENSMUSG00000032702/ENSMUSG00000024486/ENSMUSG00000033717/ENSMUSG0 |
| 22 | 5 | ENSMUSG0 | 17 | ENSMUSG00000027678/ENSMUSG00000056758/ENSMUSG00000019970/ENSMUSG00000033863/ENSMUSG00000024789/ENSMUSG00000070645/ENSMUSG0 |
| 5 | 1 | ENSMUSG0 | 4 | ENSMUSG00000028967/ENSMUSG00000003032/ENSMUSG00000028063/ENSMUSG00000044167 |
| 5 | 2 | ENSMUSG0 | 3 | ENSMUSG00000033717/ENSMUSG00000029249/ENSMUSG00000035640 |
| 5 | 3 | ENSMUSG0 | 2 | ENSMUSG00000031785/ENSMUSG00000004040 |
| 18 | 7 | ENSMUSG0 | 11 | ENSMUSG00000026360/ENSMUSG00000031565/ENSMUSG00000030849/ENSMUSG00000015846/ENSMUSG00000029287/ENSMUSG00000048616/ENSMUSG0 |
| 12 | 10 | ENSMUSG0 | 2 | ENSMUSG00000035247/ENSMUSG00000031540 |
| 27 | 6 | ENSMUSG0 | 21 | ENSMUSG00000031490/ENSMUSG00000033161/ENSMUSG00000027678/ENSMUSG00000015846/ENSMUSG00000056758/ENSMUSG00000019970/ENSMUSG0 |
| 16 | 5 | ENSMUSG0 | 11 | ENSMUSG00000032501/ENSMUSG00000031770/ENSMUSG00000053581/ENSMUSG00000032715/ENSMUSG00000020601/ENSMUSG00000022037/ENSMUSG0 |
| 25 | 7 | ENSMUSG0 | 18 | ENSMUSG00000023905/ENSMUSG00000020458/ENSMUSG00000027678/ENSMUSG00000038708/ENSMUSG00000019970/ENSMUSG00000018736/ENSMUSG0 |
| 10 | 3 | ENSMUSG0 | 7 | ENSMUSG00000022346/ENSMUSG00000000567/ENSMUSG00000031565/ENSMUSG00000028063/ENSMUSG00000030849/ENSMUSG00000023951/ENSMUSG0 |
| 19 | 3 | ENSMUSG0 | 16 | ENSMUSG00000040152/ENSMUSG00000039646/ENSMUSG00000030111/ENSMUSG00000046186/ENSMUSG00000027947/ENSMUSG00000029287/ENSMUSG0 |
| 8 | 1 | ENSMUSG0 | 7 | ENSMUSG00000022367/ENSMUSG00000027954/ENSMUSG00000020458/ENSMUSG00000029287/ENSMUSG00000048616/ENSMUSG00000007613/ENSMUSG0 |
| 9 | 3 | ENSMUSG0 | 6 | ENSMUSG00000038418/ENSMUSG00000036390/ENSMUSG00000056501/ENSMUSG00000026135/ENSMUSG00000027551/ENSMUSG00000029249 |
| 12 | 1 | ENSMUSG0 | 11 | ENSMUSG00000030657/ENSMUSG00000042042/ENSMUSG00000032997/ENSMUSG00000014959/ENSMUSG00000036356/ENSMUSG00000030530/ENSMUSG0 |
| 29 | 8 | ENSMUSG0 | 21 | ENSMUSG00000022346/ENSMUSG00000022463/ENSMUSG00000026305/ENSMUSG00000052942/ENSMUSG00000022508/ENSMUSG00000020423/ENSMUSG0 |
| 17 | 6 | ENSMUSG0 | 11 | ENSMUSG00000026360/ENSMUSG00000031565/ENSMUSG00000030849/ENSMUSG00000015846/ENSMUSG00000029287/ENSMUSG00000048616/ENSMUSG0 |
| 31 | 10 | ENSMUSG0 | 21 | ENSMUSG00000021670/ENSMUSG00000009376/ENSMUSG00000029482/ENSMUSG00000028893/ENSMUSG00000004040/ENSMUSG00000002489/ENSMUSG0 |
| 45 | 14 | ENSMUSG0 | 31 | ENSMUSG00000026360/ENSMUSG00000037411/ENSMUSG00000001627/ENSMUSG00000031565/ENSMUSG00000023905/ENSMUSG00000005069/ENSMUSG0 |
| 24 | 12 | ENSMUSG0 | 12 | ENSMUSG00000005413/ENSMUSG00000033161/ENSMUSG00000003541/ENSMUSG00000046410/ENSMUSG00000031451/ENSMUSG00000070645/ENSMUSG0 |
| 9 | 2 | ENSMUSG0 | 7 | ENSMUSG00000040152/ENSMUSG00000037411/ENSMUSG00000032068/ENSMUSG00000022508/ENSMUSG00000031523/ENSMUSG00000022425/ENSMUSG0 |
| 9 | 6 | ENSMUSG0 | 3 | ENSMUSG00000029287/ENSMUSG00000032340/ENSMUSG00000024098 |
| 43 | 9 | ENSMUSG0 | 34 | ENSMUSG00000040152/ENSMUSG00000026360/ENSMUSG00000046062/ENSMUSG00000031960/ENSMUSG00000031490/ENSMUSG00000027954/ENSMUSG0 |
| 15 | 8 | ENSMUSG0 | 7 | ENSMUSG00000040856/ENSMUSG00000027313/ENSMUSG00000004040/ENSMUSG00000022508/ENSMUSG00000021756/ENSMUSG00000007817/ENSMUSG0 |
| 23 | 6 | ENSMUSG0 | 17 | ENSMUSG00000028967/ENSMUSG00000028214/ENSMUSG00000026360/ENSMUSG00000030536/ENSMUSG00000029467/ENSMUSG00000001025/ENSMUSG0 |
| 28 | 15 | ENSMUSG0 | 13 | ENSMUSG00000028645/ENSMUSG00000069833/ENSMUSG00000028525/ENSMUSG00000096054/ENSMUSG00000054808/ENSMUSG00000032643/ENSMUSG0 |
| 12 | 3 | ENSMUSG0 | 9 | ENSMUSG00000022346/ENSMUSG00000038418/ENSMUSG00000036390/ENSMUSG00000037465/ENSMUSG00000056501/ENSMUSG00000026135/ENSMUSG0 |
| 11 | 1 | ENSMUSG0 | 10 | ENSMUSG00000040856/ENSMUSG00000025408/ENSMUSG00000045294/ENSMUSG00000032715/ENSMUSG00000020601/ENSMUSG00000023951/ENSMUSG0 |
| 31 | 10 | ENSMUSG0 | 21 | ENSMUSG00000021670/ENSMUSG00000009376/ENSMUSG00000029482/ENSMUSG00000028893/ENSMUSG00000004040/ENSMUSG00000002489/ENSMUSG0 |
| 31 | 11 | ENSMUSG0 | 20 | ENSMUSG00000022346/ENSMUSG00000038119/ENSMUSG00000009376/ENSMUSG00000050335/ENSMUSG00000031668/ENSMUSG00000026596/ENSMUSG0 |
| 54 | 6 | ENSMUSG0 | 48 | ENSMUSG00000021670/ENSMUSG00000050931/ENSMUSG00000020000/ENSMUSG00000031770/ENSMUSG00000037071/ENSMUSG00000021273/ENSMUSG0 |
| 12 | 3 | ENSMUSG0 | 9 | ENSMUSG00000031565/ENSMUSG00000030849/ENSMUSG00000031778/ENSMUSG00000023951/ENSMUSG00000020745/ENSMUSG00000037815/ENSMUSG0 |
| 7 | 0 | | 7 | ENSMUSG00000028641/ENSMUSG00000006931/ENSMUSG00000038168/ENSMUSG00000004846/ENSMUSG00000032431/ENSMUSG00000018906/ENSMUSG0 |
| 7 | 3 | ENSMUSG0 | 4 | ENSMUSG00000000567/ENSMUSG00000048616/ENSMUSG00000023951/ENSMUSG00000074934 |
| 7 | 5 | ENSMUSG0 | 2 | ENSMUSG00000038418/ENSMUSG00000027947 |
| 7 | 5 | ENSMUSG0 | 2 | ENSMUSG00000038418/ENSMUSG00000027947 |
| 7 | 4 | ENSMUSG0 | 3 | ENSMUSG00000000567/ENSMUSG00000031451/ENSMUSG00000074934 |
| 7 | 2 | ENSMUSG0 | 5 | ENSMUSG00000027995/ENSMUSG00000027954/ENSMUSG00000026259/ENSMUSG00000079037/ENSMUSG00000025326 |

0000062232/ENSMUSG00000039844
0000037902
0000032115/ENSMUSG00000031778/ENSMUSG00000039701/ENSMUSG00000020423/ENSMUSG00000024789/ENSMUSG00000030341/ENSMUSG00000056501/ENSMUSG00000023951/ENSMUSG0000006
0000029467/ENSMUSG00000028893/ENSMUSG00000020484/ENSMUSG00000024789/ENSMUSG00000049792/ENSMUSG00000035199/ENSMUSG00000024927/ENSMUSG00000018476/ENSMUSG0000002
0000055322/ENSMUSG00000005125/ENSMUSG00000026796/ENSMUSG00000031788/ENSMUSG00000033955/ENSMUSG00000023951/ENSMUSG00000022817/ENSMUSG00000037815/ENSMUSG0000004
0000026728/ENSMUSG00000027465/ENSMUSG00000061436/ENSMUSG00000006445
0000005533

0000028042/ENSMUSG00000030168/ENSMUSG00000018707/ENSMUSG00000005871
0000022723/ENSMUSG00000021136/ENSMUSG00000030849/ENSMUSG00000041592/ENSMUSG00000015846/ENSMUSG00000039004/ENSMUSG00000004040/ENSMUSG00000037242/ENSMUSG0000002

0000040268
0000015839/ENSMUSG00000056071
0000020889/ENSMUSG00000040021/ENSMUSG00000039202/ENSMUSG00000048249/ENSMUSG00000030168/ENSMUSG00000003814/ENSMUSG00000018965/ENSMUSG00000031548/ENSMUSG0000002

0000007613/ENSMUSG00000033323/ENSMUSG00000022752/ENSMUSG00000024431/ENSMUSG00000005871

0000033863/ENSMUSG00000024789/ENSMUSG00000024413/ENSMUSG00000040021/ENSMUSG00000039202/ENSMUSG00000048249/ENSMUSG00000063142/ENSMUSG00000003814/ENSMUSG0000002
0000025234/ENSMUSG00000028793/ENSMUSG00000032350/ENSMUSG00000038068/ENSMUSG00000024182
0000027996/ENSMUSG00000007613/ENSMUSG00000045983/ENSMUSG00000023951/ENSMUSG00000020745/ENSMUSG00000021978/ENSMUSG00000047281/ENSMUSG00000034154/ENSMUSG0000002
0000096054
0000021756/ENSMUSG00000031849/ENSMUSG00000048616/ENSMUSG00000030341/ENSMUSG00000007613/ENSMUSG00000005087/ENSMUSG00000024098/ENSMUSG00000044337/ENSMUSG0000002
0000026836

0000010392/ENSMUSG00000058152/ENSMUSG00000024085/ENSMUSG00000020132/ENSMUSG00000025484
0000030103/ENSMUSG00000020889/ENSMUSG00000056501/ENSMUSG00000022708/ENSMUSG00000024927/ENSMUSG00000052056/ENSMUSG00000008398/ENSMUSG00000000282/ENSMUSG0000002
0000007613/ENSMUSG00000033323/ENSMUSG00000022752/ENSMUSG00000024431/ENSMUSG00000005871
0000020349/ENSMUSG00000020484/ENSMUSG00000031451/ENSMUSG00000037012/ENSMUSG00000020889/ENSMUSG00000005533/ENSMUSG00000022708/ENSMUSG00000028161/ENSMUSG0000003
0000020458/ENSMUSG00000023067/ENSMUSG00000038708/ENSMUSG00000030849/ENSMUSG00000056758/ENSMUSG00000029287/ENSMUSG00000004040/ENSMUSG00000024811/ENSMUSG0000002
0000021070/ENSMUSG00000032487/ENSMUSG00000029361/ENSMUSG00000020684/ENSMUSG00000031393/ENSMUSG00000019943
0000040945

0000037331/ENSMUSG00000031668/ENSMUSG00000015846/ENSMUSG00000028893/ENSMUSG00000004040/ENSMUSG00000020423/ENSMUSG00000035093/ENSMUSG00000020594/ENSMUSG0000002
0000036450
0000002489/ENSMUSG00000038764/ENSMUSG00000059991/ENSMUSG00000028161/ENSMUSG00000020132/ENSMUSG00000026131/ENSMUSG00000000058/ENSMUSG00000019943/ENSMUSG0000003
0000028161/ENSMUSG00000026313/ENSMUSG00000026131/ENSMUSG00000039952/ENSMUSG00000029361/ENSMUSG00000027712/ENSMUSG00000031543
0000027551/ENSMUSG00000029249/ENSMUSG00000024431
0000021379/ENSMUSG00000095098/ENSMUSG00000044167/ENSMUSG00000024182
0000020349/ENSMUSG00000020484/ENSMUSG00000031451/ENSMUSG00000037012/ENSMUSG00000020889/ENSMUSG00000005533/ENSMUSG00000022708/ENSMUSG00000028161/ENSMUSG0000003
0000029287/ENSMUSG00000002489/ENSMUSG00000020349/ENSMUSG00000032715/ENSMUSG00000048616/ENSMUSG00000007613/ENSMUSG00000061436/ENSMUSG00000026313/ENSMUSG0000002
0000031604/ENSMUSG00000020458/ENSMUSG00000038615/ENSMUSG00000022463/ENSMUSG00000031668/ENSMUSG00000025203/ENSMUSG00000030062/ENSMUSG00000029467/ENSMUSG0000004
0000036606/ENSMUSG00000021379/ENSMUSG00000058070
0000036450

61436/ENSMUSG00000022270/ENSMUSG00000063142/ENSMUSG00000026313/ENSMUSG00000079037/ENSMUSG00000027665/ENSMUSG00000032350/ENSMUSG00000029361/ENSMUSG00000024431/E
24423/ENSMUSG00000040268/ENSMUSG00000029249/ENSMUSG00000044167/ENSMUSG00000015839/ENSMUSG00000028211/ENSMUSG00000026826/ENSMUSG00000050697/ENSMUSG00000037902/E
40511/ENSMUSG00000022443/ENSMUSG00000006445/ENSMUSG00000026131/ENSMUSG00000039952/ENSMUSG00000000058/ENSMUSG00000062901/ENSMUSG00000005871/ENSMUSG00000032220

29050/ENSMUSG00000007613/ENSMUSG00000026728/ENSMUSG00000024098/ENSMUSG00000023951/ENSMUSG00000024085/ENSMUSG00000027465/ENSMUSG00000061436/ENSMUSG00000006445/E

24431/ENSMUSG00000025326/ENSMUSG00000005886

28370/ENSMUSG00000018965/ENSMUSG00000029249/ENSMUSG00000031548/ENSMUSG00000024431/ENSMUSG00000025326/ENSMUSG00000005886

24486/ENSMUSG00000031548/ENSMUSG00000024431/ENSMUSG00000031393

26836/ENSMUSG00000074934

27230/ENSMUSG00000029249/ENSMUSG00000044167/ENSMUSG00000025612/ENSMUSG00000035397/ENSMUSG00000031393/ENSMUSG00000026628

30168/ENSMUSG00000027665/ENSMUSG00000033717/ENSMUSG00000044167/ENSMUSG00000035828/ENSMUSG00000042903/ENSMUSG00000050697
27312/ENSMUSG00000018736/ENSMUSG00000027996/ENSMUSG00000048616/ENSMUSG00000007613/ENSMUSG00000033323/ENSMUSG00000023951/ENSMUSG00000022752/ENSMUSG00000039384/E

26728/ENSMUSG00000045983/ENSMUSG00000003135/ENSMUSG00000033499/ENSMUSG00000030738/ENSMUSG00000021770/ENSMUSG00000024423/ENSMUSG00000050965/ENSMUSG00000079037/E

31453/ENSMUSG00000002365/ENSMUSG00000031543

30168/ENSMUSG00000027665/ENSMUSG00000033717/ENSMUSG00000044167/ENSMUSG00000035828/ENSMUSG00000042903/ENSMUSG00000050697
22365/ENSMUSG00000062232/ENSMUSG00000044147/ENSMUSG00000002658/ENSMUSG00000031393/ENSMUSG00000009630
42644/ENSMUSG00000045294/ENSMUSG00000020484/ENSMUSG00000014959/ENSMUSG00000047187/ENSMUSG00000030530/ENSMUSG00000079111/ENSMUSG00000039100/ENSMUSG00000023068/E

ENSMUSG00000026797/ENSMUSG00000031393/ENSMUSG00000026826/ENSMUSG00000002083
ENSMUSG00000026812/ENSMUSG00000004207

ENSMUSG00000005871

ENSMUSG00000040021/ENSMUSG00000020745/ENSMUSG00000020132/ENSMUSG00000027665/ENSMUSG00000031548/ENSMUSG00000024431/ENSMUSG00000031393/ENSMUSG00000005871

ENSMUSG00000026254/ENSMUSG00000003814/ENSMUSG00000022037/ENSMUSG00000045896/ENSMUSG00000015839/ENSMUSG00000040025/ENSMUSG00000020091/ENSMUSG00000022412/ENSMUS

ENSMUSG00000027465/ENSMUSG00000021770/ENSMUSG00000032018/ENSMUSG00000022270/ENSMUSG00000021831/ENSMUSG00000021427/ENSMUSG00000030747/ENSMUSG00000032349/ENSMUS

;G00000021838/ENSMUSG00000056071/ENSMUSG00000026812

;G00000036564/ENSMUSG00000021036/ENSMUSG00000026924/ENSMUSG00000003814/ENSMUSG00000056050/ENSMUSG00000022365/ENSMUSG00000026131/ENSMUSG00000030835/ENSMUSG0000(

)027230/ENSMUSG00000024052/ENSMUSG00000075703/ENSMUSG00000029361/ENSMUSG00000005615/ENSMUSG00000030287/ENSMUSG00000024807/ENSMUSG00000047554/ENSMUSG0000002736

| Category | GOID | Description | GeneRatio | BgRatio | pvalue | padj | geneID | geneName |
|---|---|---|---|---|---|---|---|---|
| BP | GO:00507 | negative regulation of protein secretion | 21\|1209 | 138\|1609 | 0.00147 | 0.01351 | ENSMUS | Hmgcr/Cyp51/Cd34/Cx3cl1/C1qtnf3/Neo1/Gas6/Ssc5d/Rabgef1 |
| BP | GO:00031 | aortic valve morphogenesis | 6\|1209 | 18\|16097 | 0.0015 | 0.01372 | ENSMUS | Sox9/Efna1/Robo1/Nfatc1/Notch1/Robo2 |
| BP | GO:00511 | regulation of muscle cell differentiation | 23\|1209 | 157\|1609 | 0.00152 | 0.01386 | ENSMUS | Col14a1/Rgs2/Mmp14/Fgfr2/Maml1/Xbp1/Sik1/Cxcl12/Ctdp1/ |
| BP | GO:004301 | camera-type eye development | 37\|1209 | 296\|1609 | 0.00154 | 0.01406 | ENSMUS | Col8a1/Col4a1/Sox9/Cdon/Klf4/Hipk1/Inhba/Crybg3/Cdkn1c/J |
| BP | GO:006061 | adipose tissue development | 10\|1209 | 45\|16097 | 0.00155 | 0.01415 | ENSMUS | Lmna/Hmga2/Xbp1/Arrdc3/Dgat2/Sptlc2/Ebf2/Fto/Pik3ca/Sox |
| BP | GO:00098 | negative regulation of catabolic process | 36\|1209 | 286\|1609 | 0.00157 | 0.01425 | ENSMUS | Hmox1/Hmgcr/Cyp51/Efna1/Larp1/Noct/Ier3/Cst3/Rnf41/Usp3 |
| BP | GO:00329 | regulation of actin filament-based process | 45\|1209 | 380\|1609 | 0.00157 | 0.01427 | ENSMUS | Tlr2/Eln/S100a10/Met/Atp1a1/Pecam1/Tpm1/Dixdc1/Atp2a2/ |
| BP | GO:00458 | positive regulation of protein kinase activity | 48\|1209 | 412\|1609 | 0.00157 | 0.01428 | ENSMUS | Thbs1/Map2k3/Egr1/Gadd45a/Fgfr1/Met/Il6ra/Efna1/Iqgap1/Co |
| MF | GO:00313 | translation initiation factor binding | 9\|1190 | 38\|15912 | 0.00159 | 0.03523 | ENSMUS | Myc/Eif4ebp1/Larp1/Eif2ak3/Eif4g1/Eif3c/Eif2s2/Eif4ebp2/Hhex |
| BP | GO:20012 | negative regulation of intrinsic apoptotic signaling pathway | 16\|1209 | 94\|16097 | 0.00161 | 0.01457 | ENSMUS | Herpud1/Hyou1/Xbp1/Hsph1/Cxcl12/Bag5/Cd44/Plaur/Ackr3/ |
| BP | GO:00712 | cellular response to metal ion | 21\|1209 | 139\|1609 | 0.00161 | 0.01459 | ENSMUS | Mt1/Hmox1/Jun/Iqgap1/Nfe2l1/Fos/Bmp6/Mylk/Egfr/Mef2a/Cli |
| BP | GO:00466 | response to cadmium ion | 8\|1209 | 31\|16097 | 0.00164 | 0.01478 | ENSMUS | Mt1/Hmox1/Jun/Fos/Egfr/Npc1/Slc30a1/Prnp |
| BP | GO:00518 | negative regulation of protein kinase B signaling | 9\|1209 | 38\|16097 | 0.00164 | 0.01478 | ENSMUS | Klf4/Ddit3/Igf1r/Sh2b3/Epha2/Plekha1/Dag1/Rapgef1/Lemd2 |
| BP | GO:00705 | regulation of neuron projection regeneration | 9\|1209 | 38\|16097 | 0.00164 | 0.01478 | ENSMUS | Klf4/Xylt1/Rtn4/Thy1/Ndel1/Neo1/Igf1r/Epha4/Ptn |
| BP | GO:000991 | epidermal cell differentiation | 23\|1209 | 158\|1609 | 0.00165 | 0.01481 | ENSMUS | Errfi1/Klf4/Fgfr1/Cd109/Mafg/Ncoa3/Clic4/Fosl2/Lats1/Pafah1 |
| BP | GO:00434 | steroid hormone mediated signaling pathway | 17\|1209 | 103\|1609 | 0.00165 | 0.01481 | ENSMUS | Ncoa3/Hmga2/Sgk1/Klf9/Jak2/Igf1/Lats1/Abhd2/Crebrf/Calr/Da |
| BP | GO:19018 | positive regulation of proteasomal protein catabolic process | 17\|1209 | 103\|1609 | 0.00165 | 0.01481 | ENSMUS | Hspa1b/Hspa1a/Trib1/Herpud1/Zfand2a/Trib3/Xbp1/Trib2/Nkd |
| BP | GO:00609 | endocrine hormone secretion | 11\|1209 | 53\|16097 | 0.00166 | 0.01486 | ENSMUS | Inhba/Fgfr1/Bmp6/Agtr2/C1qtnf3/Ren1/Fam129b/Tmf1/Crhr1/ |
| BP | GO:00101 | body morphogenesis | 12\|1209 | 61\|16097 | 0.0017 | 0.0151 | ENSMUS | Col1a1/Cdon/Crispld2/Phldb1/Ski/Tiparp/Ankrd11/Nog/Plekha |
| BP | GO:00603 | pathway-restricted SMAD protein phosphorylation | 12\|1209 | 61\|16097 | 0.0017 | 0.0151 | ENSMUS | Inhba/Bmp6/Tgfbr3/Xbp1/Pmepa1/Nog/Tgfbr1/Twsg1/Dab2/Lg |
| BP | GO:00604 | lung morphogenesis | 12\|1209 | 61\|16097 | 0.0017 | 0.0151 | ENSMUS | Spry2/Sox9/Stk40/Fgfr2/Hmga2/Tnc/Nog/Igf1/Plod3/Foxp2/Da |
| CC | GO:00995 | region of cytosol | 9\|1232 | 38\|16313 | 0.00171 | 0.02837 | ENSMUS | Eif4ebp1/Hnrnpa2b1/Fbxo45/Marcksl1/Nedd4/Prkca/Dag1/Dbr |
| BP | GO:00725 | reactive oxygen species metabolic process | 34\|1209 | 267\|1609 | 0.00171 | 0.01521 | ENSMUS | Thbs1/Tlr2/Klf4/Gadd45a/Met/Cdkn1a/Ier3/Cyr61/Sesn2/Cd34 |
| BP | GO:00065 | cellular amino acid metabolic process | 31\|1209 | 237\|1609 | 0.00175 | 0.01554 | ENSMUS | Aars/Gars/Sars/Nars/Tars/Psat1/Yars/Asns/Azin1/Iars/Bcat1/Ploc |
| MF | GO:00198 | protein kinase regulator activity | 22\|1190 | 150\|1591 | 0.00177 | 0.03783 | ENSMUS | Spry2/Trib1/Cdkn1c/Iqgap1/Cdkn1a/Spred1/Ibtk/Gprc5a/Trib3 |
| CC | GO:00306 | Golgi-associated vesicle membrane | 12\|1232 | 61\|16313 | 0.00178 | 0.02877 | ENSMUS | Sec24d/Sec23a/Furin/Kdelr2/Copb2/Sec16a/Copg2/Cltc/Cope/ |
| MF | GO:00445 | S100 protein binding | 5\|1190 | 13\|15912 | 0.0018 | 0.03783 | ENSMUS | Ahnak/Iqgap1/Atp2a2/S100a6/S100b |
| BP | GO:00459 | positive regulation of mitotic cell cycle | 23\|1209 | 159\|1609 | 0.0018 | 0.01589 | ENSMUS | Fgfr1/Met/Eif4ebp1/Smoc2/Egfr/Asns/Eif4g1/Igf1r/Cdt1/Igf1/Pa |
| BP | GO:00107 | negative regulation of cell morphogenesis involved in differenti | 16\|1209 | 95\|16097 | 0.0018 | 0.01593 | ENSMUS | Postn/Ifrd1/Efna1/Rtn4/Ngef/Thy1/Nrp1/Sema3b/Actn4/Ppp3c |
| BP | GO:006201 | regulation of small molecule metabolic process | 40\|1209 | 330\|1609 | 0.00182 | 0.01602 | ENSMUS | Ldlr/Myc/Egr1/Igfbp3/Stard4/Has2/Nfe2l1/Ier3/Igfbp4/Eif2ak3/ |
| BP | GO:00380 | collagen-activated signaling pathway | 5\|1209 | 13\|16097 | 0.00183 | 0.01611 | ENSMUS | Col1a1/Col4a1/Col4a2/Itga11/Itga2 |
| BP | GO:00613 | renal tubule morphogenesis | 14\|1209 | 78\|16097 | 0.00184 | 0.01614 | ENSMUS | Col4a1/Myc/Sox9/Lzts2/Agtr2/Maged1/Nog/Cd44/Vegfa/Pbx1/ |
| BP | GO:00359 | cellular response to topologically incorrect protein | 17\|1209 | 104\|1609 | 0.00184 | 0.01614 | ENSMUS | Hspa1b/Hspa1a/Herpud1/Hspa9/Atf4/Eif2ak3/Ddit3/Xbp1/Ccn |
| BP | GO:19000 | positive regulation of dendrite development | 17\|1209 | 104\|1609 | 0.00184 | 0.01614 | ENSMUS | Mapk6/Met/Iqgap1/Sgk1/Tiam1/Lrp8/Pafah1b1/Ptprd/Vldlr/Ep |
| BP | GO:00031 | endocardial cushion development | 10\|1209 | 46\|16097 | 0.00185 | 0.01617 | ENSMUS | Thbs1/Sox9/Nog/Tgfbr1/Robo1/Notch1/Nedd4/Robo2/Acvr1/M |
| BP | GO:00701 | positive regulation of biomineral tissue development | 10\|1209 | 46\|16097 | 0.00185 | 0.01617 | ENSMUS | Atf4/Cyr61/Bmp6/Cebpb/Mia3/Fzd9/Acvr1/Ptn/Atp2b1/Fbxo5 |
| BP | GO:01100 | regulation of actin filament organization | 32\|1209 | 248\|1609 | 0.00186 | 0.0162 | ENSMUS | Tlr2/Eln/S100a10/Met/Pecam1/Tpm1/Synpo/Cx3cl1/Ctgf/Nrp1 |
| BP | GO:00017 | neuron migration | 24\|1209 | 169\|1609 | 0.00186 | 0.0162 | ENSMUS | Col3a1/Met/Pex5/Adgrg1/Stat3/Tubb2b/Ndel1/Nrp1/Neo1/Gas |
| BP | GO:00712 | cellular response to inorganic substance | 24\|1209 | 169\|1609 | 0.00186 | 0.0162 | ENSMUS | Mt1/Thbs1/Hmox1/Ddi2/Jun/Iqgap1/Nfe2l1/Fos/Bmp6/Mylk/E |
| BP | GO:00336 | positive regulation of kinase activity | 51\|1209 | 448\|1609 | 0.00189 | 0.01637 | ENSMUS | Thbs1/Map2k3/Egr1/Gadd45a/Fgfr1/Met/Il6ra/Efna1/Iqgap1/Co |
| BP | GO:00181 | peptide cross-linking | 7\|1209 | 25\|16097 | 0.00191 | 0.01638 | ENSMUS | Col3a1/Thbs1/F13a1/Tgm2/Egflam/Spock2/Ndnf |
| BP | GO:00350 | negative regulation of Rho protein signal transduction | 7\|1209 | 25\|16097 | 0.00191 | 0.01638 | ENSMUS | Met/Abl2/Bcl6/Dlc1/Kank1/Stmn1/Rasip1 |
| BP | GO:00714 | hematopoietic stem cell proliferation | 7\|1209 | 25\|16097 | 0.00191 | 0.01638 | ENSMUS | Prg4/Cd34/Sfrp2/Fubp1/Atxn1l/Ace/Ythdf2 |
| BP | GO:009011 | COPII-coated vesicle budding | 7\|1209 | 25\|16097 | 0.00191 | 0.01638 | ENSMUS | Insig1/Sec24d/Sec23a/Tbc1d20/Sec16a/Mia3/Sec24a |
| BP | GO:00901 | regulation of branching involved in ureteric bud morphogenesi | 7\|1209 | 25\|16097 | 0.00191 | 0.01638 | ENSMUS | Sox9/Agtr2/Maged1/Nog/Vegfa/Grem1/Sox8 |
| BP | GO:190367 | negative regulation of sprouting angiogenesis | 7\|1209 | 25\|16097 | 0.00191 | 0.01638 | ENSMUS | Thbs1/Klf4/Spred1/Meox2/Pik3r2/Notch1/Creb3l1 |
| BP | GO:200005 | regulation of non-canonical Wnt signaling pathway | 7\|1209 | 25\|16097 | 0.00191 | 0.01638 | ENSMUS | Abl2/Tiam1/Sfrp2/Sfrp4/Nkd1/Dab2/Sfrp1 |
| BP | GO:00163 | dendrite development | 35\|1209 | 279\|1609 | 0.00191 | 0.01638 | ENSMUS | Mapk6/Met/Efna1/Iqgap1/Abl2/Ngef/Sgk1/Mef2a/Tiam1/Lrp8/ |
| BP | GO:00326 | regulation of interleukin-6 production | 21\|1209 | 141\|1609 | 0.00193 | 0.01652 | ENSMUS | Prg4/Tlr2/Lpl/Met/Il6ra/Stat3/Xbp1/C1qtnf3/Gas6/Cd36/Tnfrsf |
| BP | GO:00342 | negative regulation of cellular amide metabolic process | 21\|1209 | 141\|1609 | 0.00193 | 0.01652 | ENSMUS | Igfbp5/Rgs2/Eif4ebp1/Larp1/Eif2ak3/Sesn2/Stat3/Btg2/Eif4g1/I |
| CC | GO:00995 | postsynaptic cytosol | 7\|1232 | 25\|16313 | 0.00197 | 0.03106 | ENSMUS | Eif4ebp1/Hnrnpa2b1/Fbxo45/Nedd4/Dag1/Dbn1/Baiap2 |

| Count | Up | Up_Gene_i | Down | Down_Gene_id |
|---|---|---|---|---|
| 21 | 7 | ENSMUSG0 | 14 | ENSMUSG00000021670/ENSMUSG00000001467/ENSMUSG00000031778/ENSMUSG00000032340/ENSMUSG00000031451/ENSMUSG00000025340/ENSMUSG0 |
| 6 | 4 | ENSMUSG0 | 2 | ENSMUSG00000000567/ENSMUSG00000027954 |
| 23 | 10 | ENSMUSG0 | 13 | ENSMUSG00000026360/ENSMUSG00000030849/ENSMUSG00000050567/ENSMUSG00000020484/ENSMUSG00000033323/ENSMUSG00000022752/ENSMUSG0 |
| 37 | 16 | ENSMUSG0 | 21 | ENSMUSG00000068196/ENSMUSG00000000567/ENSMUSG00000038119/ENSMUSG00000003032/ENSMUSG00000008730/ENSMUSG00000041324/ENSMUSG0 |
| 10 | 3 | ENSMUSG0 | 7 | ENSMUSG00000028063/ENSMUSG00000056758/ENSMUSG00000020484/ENSMUSG00000074794/ENSMUSG00000030747/ENSMUSG00000021036/ENSMUSG0 |
| 36 | 6 | ENSMUSG0 | 30 | ENSMUSG00000005413/ENSMUSG00000021670/ENSMUSG00000001467/ENSMUSG00000027954/ENSMUSG00000037331/ENSMUSG00000023087/ENSMUSG0 |
| 45 | 21 | ENSMUSG0 | 24 | ENSMUSG00000027995/ENSMUSG00000029675/ENSMUSG00000041959/ENSMUSG00000009376/ENSMUSG00000033161/ENSMUSG00000032064/ENSMUSG0 |
| 48 | 14 | ENSMUSG0 | 34 | ENSMUSG00000040152/ENSMUSG00000018932/ENSMUSG00000038418/ENSMUSG00000036390/ENSMUSG00000031565/ENSMUSG00000009376/ENSMUSG0 |
| 9 | 1 | ENSMUSG0 | 8 | ENSMUSG00000022346/ENSMUSG00000031490/ENSMUSG00000037331/ENSMUSG00000031668/ENSMUSG00000045983/ENSMUSG00000030738/ENSMUSG0 |
| 16 | 2 | ENSMUSG0 | 14 | ENSMUSG00000031770/ENSMUSG00000032115/ENSMUSG00000020484/ENSMUSG00000049792/ENSMUSG00000005087/ENSMUSG00000046223/ENSMUSG0 |
| 21 | 7 | ENSMUSG0 | 14 | ENSMUSG00000031765/ENSMUSG00000005413/ENSMUSG00000030536/ENSMUSG00000038615/ENSMUSG00000039004/ENSMUSG00000030557/ENSMUSG0 |
| 8 | 3 | ENSMUSG0 | 5 | ENSMUSG00000031765/ENSMUSG00000005413/ENSMUSG00000024413/ENSMUSG00000037434/ENSMUSG00000079037 |
| 9 | 0 | | 9 | ENSMUSG00000003032/ENSMUSG00000025408/ENSMUSG00000005533/ENSMUSG00000042594/ENSMUSG00000006445/ENSMUSG00000040268/ENSMUSG0 |
| 9 | 3 | ENSMUSG0 | 6 | ENSMUSG00000003032/ENSMUSG00000030657/ENSMUSG00000020458/ENSMUSG00000018736/ENSMUSG00000032340/ENSMUSG00000005533 |
| 23 | 4 | ENSMUSG0 | 19 | ENSMUSG00000028967/ENSMUSG00000003032/ENSMUSG00000031565/ENSMUSG00000046186/ENSMUSG00000051510/ENSMUSG00000027678/ENSMUSG0 |
| 17 | 3 | ENSMUSG0 | 14 | ENSMUSG00000027678/ENSMUSG00000056758/ENSMUSG00000019970/ENSMUSG00000033863/ENSMUSG00000024789/ENSMUSG00000040021/ENSMUSG0 |
| 17 | 5 | ENSMUSG0 | 12 | ENSMUSG00000032501/ENSMUSG00000031770/ENSMUSG00000053581/ENSMUSG00000032715/ENSMUSG00000020484/ENSMUSG00000020601/ENSMUSG0 |
| 11 | 4 | ENSMUSG0 | 7 | ENSMUSG00000041324/ENSMUSG00000031565/ENSMUSG00000039004/ENSMUSG00000070645/ENSMUSG00000026796/ENSMUSG00000030059/ENSMUSG0 |
| 12 | 2 | ENSMUSG0 | 10 | ENSMUSG00000038119/ENSMUSG00000031825/ENSMUSG00000048537/ENSMUSG00000029050/ENSMUSG00000034640/ENSMUSG00000035569/ENSMUSG0 |
| 12 | 2 | ENSMUSG0 | 10 | ENSMUSG00000041324/ENSMUSG00000039004/ENSMUSG00000029287/ENSMUSG00000020484/ENSMUSG00000038400/ENSMUSG00000048616/ENSMUSG0 |
| 12 | 4 | ENSMUSG0 | 8 | ENSMUSG00000022114/ENSMUSG00000000567/ENSMUSG00000042608/ENSMUSG00000030849/ENSMUSG00000056758/ENSMUSG00000048616/ENSMUSG0 |
| 9 | 4 | ENSMUSG0 | 5 | ENSMUSG00000031490/ENSMUSG00000035764/ENSMUSG00000050965/ENSMUSG00000039952/ENSMUSG00000025372 |
| 34 | 13 | ENSMUSG0 | 21 | ENSMUSG00000040152/ENSMUSG00000027995/ENSMUSG00000003032/ENSMUSG00000036390/ENSMUSG00000009376/ENSMUSG00000023067/ENSMUSG0 |
| 31 | 4 | ENSMUSG0 | 27 | ENSMUSG00000031960/ENSMUSG00000029777/ENSMUSG00000068739/ENSMUSG00000024587/ENSMUSG00000022241/ENSMUSG00000024640/ENSMUSG0 |
| 22 | 5 | ENSMUSG0 | 17 | ENSMUSG00000022114/ENSMUSG00000032501/ENSMUSG00000030536/ENSMUSG00000023067/ENSMUSG00000027351/ENSMUSG00000035941/ENSMUSG0 |
| 12 | 2 | ENSMUSG0 | 10 | ENSMUSG00000039234/ENSMUSG00000020986/ENSMUSG00000030530/ENSMUSG00000079111/ENSMUSG00000032458/ENSMUSG00000026924/ENSMUSG0 |
| 5 | 1 | ENSMUSG0 | 4 | ENSMUSG00000069833/ENSMUSG00000030536/ENSMUSG00000029467/ENSMUSG00000001025 |
| 23 | 9 | ENSMUSG0 | 14 | ENSMUSG00000031565/ENSMUSG00000009376/ENSMUSG00000031490/ENSMUSG00000029752/ENSMUSG00000045983/ENSMUSG00000005533/ENSMUSG0 |
| 16 | 5 | ENSMUSG0 | 11 | ENSMUSG00000001627/ENSMUSG00000027954/ENSMUSG00000020458/ENSMUSG00000026259/ENSMUSG00000054808/ENSMUSG00000028161/ENSMUSG0 |
| 40 | 10 | ENSMUSG0 | 30 | ENSMUSG00000032193/ENSMUSG00000022346/ENSMUSG00000038418/ENSMUSG00000024378/ENSMUSG00000022367/ENSMUSG00000038615/ENSMUSG0 |
| 5 | 5 | ENSMUSG0 | 0 | |
| 14 | 7 | ENSMUSG0 | 7 | ENSMUSG00000022346/ENSMUSG00000000567/ENSMUSG00000035342/ENSMUSG00000048616/ENSMUSG00000005087/ENSMUSG00000023951/ENSMUSG0 |
| 17 | 3 | ENSMUSG0 | 14 | ENSMUSG00000031770/ENSMUSG00000024359/ENSMUSG00000042406/ENSMUSG00000031668/ENSMUSG00000025408/ENSMUSG00000020484/ENSMUSG0 |
| 17 | 5 | ENSMUSG0 | 12 | ENSMUSG00000042688/ENSMUSG00000009376/ENSMUSG00000030536/ENSMUSG00000019970/ENSMUSG00000002489/ENSMUSG00000028613/ENSMUSG0 |
| 10 | 5 | ENSMUSG0 | 5 | ENSMUSG00000040152/ENSMUSG00000000567/ENSMUSG00000048616/ENSMUSG00000007613/ENSMUSG00000026836 |
| 10 | 4 | ENSMUSG0 | 6 | ENSMUSG00000042406/ENSMUSG00000039004/ENSMUSG00000056501/ENSMUSG00000056050/ENSMUSG00000026836/ENSMUSG00000019943 |
| 32 | 15 | ENSMUSG0 | 17 | ENSMUSG00000027995/ENSMUSG00000029675/ENSMUSG00000041959/ENSMUSG00000009376/ENSMUSG00000031778/ENSMUSG00000019997/ENSMUSG0 |
| 24 | 9 | ENSMUSG0 | 15 | ENSMUSG00000009376/ENSMUSG00000005069/ENSMUSG00000031785/ENSMUSG00000004040/ENSMUSG00000018736/ENSMUSG00000032340/ENSMUSG0 |
| 24 | 7 | ENSMUSG0 | 17 | ENSMUSG00000031765/ENSMUSG00000040152/ENSMUSG00000005413/ENSMUSG00000078515/ENSMUSG00000030536/ENSMUSG00000038615/ENSMUSG0 |
| 51 | 15 | ENSMUSG0 | 36 | ENSMUSG00000040152/ENSMUSG00000018932/ENSMUSG00000038418/ENSMUSG00000036390/ENSMUSG00000031565/ENSMUSG00000009376/ENSMUSG0 |
| 7 | 5 | ENSMUSG0 | 2 | ENSMUSG00000040152/ENSMUSG00000037820 |
| 7 | 2 | ENSMUSG0 | 5 | ENSMUSG00000009376/ENSMUSG00000026596/ENSMUSG00000022508/ENSMUSG00000031523/ENSMUSG00000032702 |
| 7 | 3 | ENSMUSG0 | 4 | ENSMUSG00000006014/ENSMUSG00000027996/ENSMUSG00000069895/ENSMUSG00000040025 |
| 7 | 0 | | 7 | ENSMUSG00000045294/ENSMUSG00000039234/ENSMUSG00000020986/ENSMUSG00000027465/ENSMUSG00000026924/ENSMUSG00000056050/ENSMUSG0 |
| 7 | 3 | ENSMUSG0 | 4 | ENSMUSG00000000567/ENSMUSG00000048616/ENSMUSG00000023951/ENSMUSG00000074934 |
| 7 | 3 | ENSMUSG0 | 4 | ENSMUSG00000040152/ENSMUSG00000003032/ENSMUSG00000027351/ENSMUSG00000027230 |
| 7 | 3 | ENSMUSG0 | 4 | ENSMUSG00000026596/ENSMUSG00000002489/ENSMUSG00000027996/ENSMUSG00000031548 |
| 35 | 10 | ENSMUSG0 | 25 | ENSMUSG00000042688/ENSMUSG00000009376/ENSMUSG00000027954/ENSMUSG00000030536/ENSMUSG00000026596/ENSMUSG00000026259/ENSMUSG0 |
| 21 | 6 | ENSMUSG0 | 15 | ENSMUSG00000006014/ENSMUSG00000027995/ENSMUSG00000009376/ENSMUSG00000027947/ENSMUSG00000004040/ENSMUSG00000020484/ENSMUSG0 |
| 21 | 3 | ENSMUSG0 | 18 | ENSMUSG00000026360/ENSMUSG00000031490/ENSMUSG00000037331/ENSMUSG00000031668/ENSMUSG00000028893/ENSMUSG00000004040/ENSMUSG0 |
| 7 | 3 | ENSMUSG0 | 4 | ENSMUSG00000031490/ENSMUSG00000035764/ENSMUSG00000039952/ENSMUSG00000025372 |

0000028161/ENSMUSG00000033717/ENSMUSG00000029249/ENSMUSG00000029361/ENSMUSG00000031548/ENSMUSG00000035828/ENSMUSG00000035621/ENSMUSG00000027712

0000002052/ENSMUSG00000026313/ENSMUSG00000028179/ENSMUSG00000024431/ENSMUSG00000031393/ENSMUSG00000042903/ENSMUSG00000074934
0000022723/ENSMUSG00000030849/ENSMUSG00000041592/ENSMUSG00000015846/ENSMUSG00000037242/ENSMUSG00000029050/ENSMUSG00000007613/ENSMUSG00000026728/ENSMUSG0000002
0000027665
0000003541/ENSMUSG00000027447/ENSMUSG00000025373/ENSMUSG00000033909/ENSMUSG00000001131/ENSMUSG00000004040/ENSMUSG00000049323/ENSMUSG00000024413/ENSMUSG0000003
0000029467/ENSMUSG00000028525/ENSMUSG00000031778/ENSMUSG00000019997/ENSMUSG00000031523/ENSMUSG00000007613/ENSMUSG00000081683/ENSMUSG00000040021/ENSMUSG0000002
0000027947/ENSMUSG00000027954/ENSMUSG00000030536/ENSMUSG00000023067/ENSMUSG00000025402/ENSMUSG00000002489/ENSMUSG00000028613/ENSMUSG00000031714/ENSMUSG0000002
0000074656/ENSMUSG00000020091
0000044337/ENSMUSG00000021070/ENSMUSG00000022037/ENSMUSG00000032487/ENSMUSG00000027230/ENSMUSG00000015839/ENSMUSG00000009291/ENSMUSG00000024807
0000037242/ENSMUSG00000079037/ENSMUSG00000025582/ENSMUSG00000015839/ENSMUSG00000013275/ENSMUSG00000031393/ENSMUSG00000050697/ENSMUSG00000027712

0000039952/ENSMUSG00000039844/ENSMUSG00000044857

0000037242/ENSMUSG00000029135/ENSMUSG00000040021/ENSMUSG00000020745/ENSMUSG00000053797/ENSMUSG00000047281/ENSMUSG00000006445/ENSMUSG00000063142/ENSMUSG0000003
0000039202/ENSMUSG00000048249/ENSMUSG00000003814/ENSMUSG00000018965/ENSMUSG00000031548/ENSMUSG00000024431/ENSMUSG00000025326/ENSMUSG00000005886
0000022037/ENSMUSG00000025234/ENSMUSG00000028793/ENSMUSG00000032350/ENSMUSG00000038068/ENSMUSG00000024182
0000018634
0000048616/ENSMUSG00000040268/ENSMUSG00000039952/ENSMUSG00000031540
0000007613/ENSMUSG00000024098/ENSMUSG00000026836/ENSMUSG00000074934
0000004846/ENSMUSG00000039952

0000003541/ENSMUSG00000028893/ENSMUSG00000025373/ENSMUSG00000004040/ENSMUSG00000019997/ENSMUSG00000024789/ENSMUSG00000018339/ENSMUSG00000027832/ENSMUSG0000002
0000028811/ENSMUSG00000029752/ENSMUSG00000037458/ENSMUSG00000037851/ENSMUSG00000030268/ENSMUSG00000004846/ENSMUSG00000039662/ENSMUSG00000022299/ENSMUSG0000003
0000046733/ENSMUSG00000032715/ENSMUSG00000049323/ENSMUSG00000020601/ENSMUSG00000031451/ENSMUSG00000011960/ENSMUSG00000036707/ENSMUSG00000027665/ENSMUSG0000000
0000047126/ENSMUSG00000036391/ENSMUSG00000032096/ENSMUSG00000027366

0000006585/ENSMUSG00000020745/ENSMUSG00000109511/ENSMUSG00000050965/ENSMUSG00000040945/ENSMUSG00000009907/ENSMUSG00000000184/ENSMUSG00000031393
0000032702/ENSMUSG00000040945/ENSMUSG00000062232/ENSMUSG00000018965/ENSMUSG00000025326
0000003541/ENSMUSG00000031668/ENSMUSG00000028893/ENSMUSG00000045294/ENSMUSG00000039004/ENSMUSG00000004040/ENSMUSG00000032715/ENSMUSG00000020889/ENSMUSG0000002

0000074934
0000021831/ENSMUSG00000048249/ENSMUSG00000003814/ENSMUSG00000022365/ENSMUSG00000027230/ENSMUSG00000020571/ENSMUSG00000015839/ENSMUSG00000026628
0000020745/ENSMUSG00000024924/ENSMUSG00000020132/ENSMUSG00000024431/ENSMUSG00000031393/ENSMUSG00000025372

0000031523/ENSMUSG00000007613/ENSMUSG00000040021/ENSMUSG00000024533/ENSMUSG00000040852/ENSMUSG00000032702/ENSMUSG00000038859/ENSMUSG00000044147/ENSMUSG0000002
0000031451/ENSMUSG00000035764/ENSMUSG00000022305/ENSMUSG00000023951/ENSMUSG00000020745/ENSMUSG00000036606/ENSMUSG00000062232/ENSMUSG00000068740/ENSMUSG0000002
0000039004/ENSMUSG00000030557/ENSMUSG00000037242/ENSMUSG00000079037/ENSMUSG00000025582/ENSMUSG00000044167/ENSMUSG00000015839/ENSMUSG00000013275/ENSMUSG0000003
0000027947/ENSMUSG00000027954/ENSMUSG00000030536/ENSMUSG00000023067/ENSMUSG00000025402/ENSMUSG00000002489/ENSMUSG00000028613/ENSMUSG00000031714/ENSMUSG0000002

0000036391

0000019970/ENSMUSG00000030557/ENSMUSG00000002489/ENSMUSG00000028613/ENSMUSG00000020745/ENSMUSG00000096054/ENSMUSG00000024533/ENSMUSG00000028161/ENSMUSG0000002
0000031451/ENSMUSG00000030341/ENSMUSG00000056501/ENSMUSG00000037447/ENSMUSG00000022708/ENSMUSG00000025340/ENSMUSG00000016024/ENSMUSG00000027551/ENSMUSG0000003
0000020423/ENSMUSG00000045983/ENSMUSG00000050965/ENSMUSG00000079037/ENSMUSG00000026254/ENSMUSG00000003814/ENSMUSG00000022037/ENSMUSG00000045896/ENSMUSG0000004

24098/ENSMUSG00000023951/ENSMUSG00000024085/ENSMUSG00000027465/ENSMUSG00000061436/ENSMUSG00000006445/ENSMUSG00000005871

37458/ENSMUSG00000049792/ENSMUSG00000045983/ENSMUSG00000030530/ENSMUSG00000038764/ENSMUSG00000024927/ENSMUSG00000023951/ENSMUSG00000034575/ENSMUSG00000061436/E
22443/ENSMUSG00000024533/ENSMUSG00000040852/ENSMUSG00000032702/ENSMUSG00000038859/ENSMUSG00000029361/ENSMUSG00000044147/ENSMUSG00000025372/ENSMUSG00000026812/E
20349/ENSMUSG00000024789/ENSMUSG00000031451/ENSMUSG00000007613/ENSMUSG00000081683/ENSMUSG00000023951/ENSMUSG00000036707/ENSMUSG00000032717/ENSMUSG00000024924/E

39473/ENSMUSG00000074818/ENSMUSG00000032487/ENSMUSG00000029249/ENSMUSG00000004207

26313/ENSMUSG00000022037/ENSMUSG00000032487/ENSMUSG00000044167/ENSMUSG00000029361/ENSMUSG00000015839/ENSMUSG00000037902
32431/ENSMUSG00000021831/ENSMUSG00000018906/ENSMUSG00000053398/ENSMUSG00000029009/ENSMUSG00000028179/ENSMUSG00000032350/ENSMUSG00000029361/ENSMUSG00000031700/E
00184/ENSMUSG00000074934/ENSMUSG00000005871

22708/ENSMUSG00000030747/ENSMUSG00000032349/ENSMUSG00000063142/ENSMUSG00000026313/ENSMUSG00000032487/ENSMUSG00000047126/ENSMUSG00000029249/ENSMUSG00000044167/E

25372/ENSMUSG00000026812/ENSMUSG00000002365
26826
31393/ENSMUSG00000050697/ENSMUSG00000027712
20349/ENSMUSG00000024789/ENSMUSG00000031451/ENSMUSG00000007613/ENSMUSG00000081683/ENSMUSG00000005087/ENSMUSG00000023951/ENSMUSG00000036707/ENSMUSG00000032717/E

24924/ENSMUSG00000062232/ENSMUSG00000020132/ENSMUSG00000018965/ENSMUSG00000041570/ENSMUSG00000068740/ENSMUSG00000044147/ENSMUSG00000024431/ENSMUSG00000031393/E
37902
40025/ENSMUSG00000020091/ENSMUSG00000021838/ENSMUSG00000026812

ENSMUSG00000020044/ENSMUSG00000030527/ENSMUSG00000026313/ENSMUSG00000033717/ENSMUSG00000050697/ENSMUSG00000028277/ENSMUSG00000026812
ENSMUSG00000002365
ENSMUSG00000024383/ENSMUSG00000079037/ENSMUSG00000022037/ENSMUSG00000062232/ENSMUSG00000039952/ENSMUSG00000033717/ENSMUSG00000000184/ENSMUSG00000074934/ENSMUS

ENSMUSG00000031393/ENSMUSG00000021794/ENSMUSG00000010755/ENSMUSG00000005871

ENSMUSG00000029361/ENSMUSG00000026623/ENSMUSG00000024431/ENSMUSG00000050697/ENSMUSG00000005886/ENSMUSG00000035621/ENSMUSG00000005871

ENSMUSG00000024924/ENSMUSG00000024383/ENSMUSG00000079037/ENSMUSG00000022037/ENSMUSG00000062232/ENSMUSG00000039952/ENSMUSG00000033717/ENSMUSG00000000184/ENSMUS

ENSMUSG00000025326/ENSMUSG00000025372

;G00000024182/ENSMUSG00000074923/ENSMUSG00000002365

;G00000074934/ENSMUSG00000028661/ENSMUSG00000024182/ENSMUSG00000074923/ENSMUSG00000002365

| Category | GOID | Description | GeneRatio | BgRatio | pvalue | padj | geneID | geneName |
|---|---|---|---|---|---|---|---|---|
| BP | GO:005092 | positive regulation of chemotaxis | 20\|1209 | 132\|16097 | 0.00199 | 0.01696 | ENSMUS | Thbs1/Serpine1/Fgfr1/Met/Smoc2/Cx3cl1/Thbs4/Tiam1/Tubb2 |
| BP | GO:002195 | central nervous system neuron axonogenesis | 9\|1209 | 39\|16097 | 0.002 | 0.01699 | ENSMUS | Cdh11/B4galt5/Ndel1/Fbxo45/Pafah1b1/Prkca/Epha4/Zeb2/Nr |
| BP | GO:000834 | adult locomotory behavior | 16\|1209 | 96\|16097 | 0.00202 | 0.01711 | ENSMUS | Dnm1/Abl2/Npc1/Cxcl12/Pafah1b1/Hipk2/Kcnma1/Epha4/Atp |
| BP | GO:005189 | positive regulation of protein kinase B signaling | 16\|1209 | 96\|16097 | 0.00202 | 0.01711 | ENSMUS | Spry2/F3/Thbs1/Igfbp5/Rtn4/Cx3cl1/Egfr/C1qtnf3/Gas6/Tgfbr1/ |
| BP | GO:002184 | cell proliferation in forebrain | 8\|1209 | 32\|16097 | 0.00204 | 0.01726 | ENSMUS | Fgfr1/Fgfr2/Dixdc1/Hmga2/Tacc2/Tacc1/Zeb2/Hhex |
| BP | GO:007163 | regulation of transforming growth factor beta production | 8\|1209 | 32\|16097 | 0.00204 | 0.01726 | ENSMUS | Thbs1/Met/Cd34/Cx3cl1/Ltbp1/Furin/Lum/Ptgs2 |
| BP | GO:200017 | negative regulation of dendrite development | 8\|1209 | 32\|16097 | 0.00204 | 0.01726 | ENSMUS | Efna1/Ngef/Ppp3ca/Rapgef2/Ywhah/Arf6/Mecp2/Ube3a |
| BP | GO:004253 | positive regulation of tumor necrosis factor biosynthetic proce | 6\|1209 | 19\|16097 | 0.00205 | 0.0173 | ENSMUS | Thbs1/Tlr2/Akap12/Jak2/Lbp/App |
| CC | GO:004319 | dendritic spine | 25\|1232 | 179\|16313 | 0.00207 | 0.03171 | ENSMUS | Cald1/Lama2/Dnm1/Abl2/Pde4b/Arfgef2/Synpo/Tiam1/Nr1d1/ |
| BP | GO:006042 | regulation of heart growth | 14\|1209 | 79\|16097 | 0.00208 | 0.01753 | ENSMUS | Col14a1/Rgs2/Fgfr1/Fgfr2/Tgfbr3/Nog/Tgfbr1/Ctdp1/Igf1/Tomm |
| BP | GO:004516 | cell fate commitment | 31\|1209 | 240\|16097 | 0.00214 | 0.01799 | ENSMUS | Spry2/Sox9/Cdon/Fgfr1/Chrdl1/Nfe2l1/Smad5/Fgfr2/Stat3/AW |
| BP | GO:000268 | negative regulation of immune system process | 50\|1209 | 440\|16097 | 0.00216 | 0.01811 | ENSMUS | Col3a1/Thbs1/Hmox1/Ldlr/Myc/Sox9/Trib1/A2m/Sec14l1/Dusp |
| BP | GO:000193 | positive regulation of endothelial cell proliferation | 15\|1209 | 88\|16097 | 0.00219 | 0.01835 | ENSMUS | F3/Hmox1/Fgfr1/Jun/Bmp6/Stat3/Thbs4/Nr4a1/Cxcl12/Igf1/Flt |
| MF | GO:003525 | nuclear hormone receptor binding | 21\|1190 | 143\|15912 | 0.00219 | 0.04508 | ENSMUS | Ncoa3/Hmga1/Rxra/Stat3/Nr4a1/Tacc2/Cebpb/Arid5a/Hr/Nrip1 |
| BP | GO:000803 | neuron recognition | 10\|1209 | 47\|16097 | 0.0022 | 0.01838 | ENSMUS | Rtn4/Nrp1/Robo1/Robo2/Bsg/Epha4/Tnn/App/Gap43/Emb |
| BP | GO:003209 | positive regulation of protein binding | 17\|1209 | 106\|16097 | 0.00227 | 0.01899 | ENSMUS | Cald1/Lgals3/Eif2ak3/Abl2/Tiam1/Ppp2ca/Trib3/Plxnd1/Hipk2/ |
| CC | GO:004217 | nuclear outer membrane-endoplasmic reticulum membrane ne | 55\|1232 | 493\|16313 | 0.00228 | 0.03402 | ENSMUS | Hmgcr/Sgms2/Moxd1/Herpud1/Scd1/Fdft1/Msmo1/Rtn4/Nfe2l |
| CC | GO:009856 | cytoplasmic side of membrane | 24\|1232 | 171\|16313 | 0.00235 | 0.03431 | ENSMUS | Errfi1/Gem/Rgs2/Ptp4a1/Iqgap1/Atp2a2/S100a6/Tiam1/Ptpn3/ |
| BP | GO:004688 | regulation of hormone secretion | 33\|1209 | 262\|16097 | 0.00237 | 0.01981 | ENSMUS | Hmgcr/Inhba/Fgfr1/Igfbp3/Aacs/Hmga2/Bmp6/Egfr/Tiam1/Agtr |
| BP | GO:000323 | cardiac ventricle development | 20\|1209 | 134\|16097 | 0.00239 | 0.01984 | ENSMUS | Cyr61/Tpm1/Fgfr2/Rxra/Tgfbr3/Sfrp2/Ltbp1/Nog/Tgfbr1/Hectd |
| BP | GO:000705 | cell cycle arrest | 20\|1209 | 134\|16097 | 0.00239 | 0.01984 | ENSMUS | Thbs1/Myc/Inhba/Gadd45a/Cdkn1c/Cdkn1a/Ddit3/Mybbp1a/N |
| MF | GO:004696 | thyroid hormone receptor binding | 7\|1190 | 26\|15912 | 0.00239 | 0.04811 | ENSMUS | Arid5a/Hr/Nup62/Med13/Gas2l1/Tacc1/Ncoa2 |
| BP | GO:001403 | Schwann cell differentiation | 9\|1209 | 40\|16097 | 0.00241 | 0.01992 | ENSMUS | Nab2/Nab1/Ski/Ndrg1/Rela/Adgrg6/Bag1/Lamb2/Dag1 |
| BP | GO:190305 | regulation of extracellular matrix organization | 9\|1209 | 40\|16097 | 0.00241 | 0.01992 | ENSMUS | Sox9/Has2/Phldb1/Abl2/Tnfrsf1a/Chadl/Notch1/Lemd3/Dag1 |
| BP | GO:190352 | negative regulation of blood circulation | 9\|1209 | 40\|16097 | 0.00241 | 0.01992 | ENSMUS | Atp1a1/Atp2a2/Cx3cl1/Agtr2/Jak2/Dusp5/Prkca/Atp1a2/Nos1 |
| BP | GO:004582 | negative regulation of heart contraction | 7\|1209 | 26\|16097 | 0.00245 | 0.02019 | ENSMUS | Atp1a1/Atp2a2/Agtr2/Jak2/Prkca/Atp1a2/Nos1 |
| BP | GO:005119 | positive regulation of cofactor metabolic process | 7\|1209 | 26\|16097 | 0.00245 | 0.02019 | ENSMUS | Myc/Eif2ak3/Igf1/Zbtb20/Nfe2l2/App/Prkaa1 |
| BP | GO:004616 | alcohol biosynthetic process | 19\|1209 | 125\|16097 | 0.00245 | 0.02019 | ENSMUS | Hmgcr/Moxd1/Cyp51/Fdft1/Hmgcs1/Insig1/Bmp6/Hsd17b7/Dh |
| BP | GO:001490 | myotube differentiation | 18\|1209 | 116\|16097 | 0.0025 | 0.02047 | ENSMUS | Acta1/Cdon/Met/Rcan1/Mmp14/Maml1/Xbp1/Sik1/Neo1/Ski/ |
| BP | GO:004230 | molting cycle | 18\|1209 | 116\|16097 | 0.0025 | 0.02047 | ENSMUS | Igfbp5/Sox9/Inhba/Cd109/Fgfr2/Egfr/Rela/Nfatc1/Notch1/Krt1 |
| BP | GO:004263 | hair cycle | 18\|1209 | 116\|16097 | 0.0025 | 0.02047 | ENSMUS | Igfbp5/Sox9/Inhba/Cd109/Fgfr2/Egfr/Rela/Nfatc1/Notch1/Krt1 |
| BP | GO:003261 | interleukin-1 production | 16\|1209 | 98\|16097 | 0.00251 | 0.02047 | ENSMUS | Errfi1/Tlr2/Egr1/Lpl/Cx3cl1/Jak2/Gas6/Hk1/Cd36/Igf1/Lgals9/P |
| BP | GO:007200 | nephron epithelium development | 16\|1209 | 98\|16097 | 0.00251 | 0.02047 | ENSMUS | Myc/Sox9/Lzts2/Agtr2/Maged1/Nog/Cd44/Vegfa/Notch1/Pbx1/ |
| MF | GO:000104 | core promoter sequence-specific DNA binding | 10\|1190 | 48\|15912 | 0.00251 | 0.04946 | ENSMUS | Egr1/Gadd45a/Fos/Klf10/Cebpb/Zfp521/Zfp142/Zfp64/Rest/Sox |
| BP | GO:190533 | regulation of morphogenesis of an epithelium | 17\|1209 | 107\|16097 | 0.00252 | 0.02047 | ENSMUS | Sox9/Fgfr1/Met/Abl2/Rxra/Agtr2/Maged1/Sfrp2/Nog/Vegfa/Nkd |
| BP | GO:000320 | endocardial cushion morphogenesis | 8\|1209 | 33\|16097 | 0.00252 | 0.02047 | ENSMUS | Sox9/Nog/Tgfbr1/Robo1/Notch1/Robo2/Acvr1/Mdm4 |
| BP | GO:000340 | axis elongation | 8\|1209 | 33\|16097 | 0.00252 | 0.02047 | ENSMUS | Spry2/Sox9/Fgfr2/Tnc/Sfrp2/Zfp568/Nkd1/Sfrp1 |
| BP | GO:000698 | ER-nucleus signaling pathway | 8\|1209 | 33\|16097 | 0.00252 | 0.02047 | ENSMUS | Ppp1r15b/Atf4/Eif2ak3/Ddit3/Atp2a2/Insig1/Zbtb7b/Nfe2l2 |
| BP | GO:004800 | insulin-like growth factor receptor signaling pathway | 8\|1209 | 33\|16097 | 0.00252 | 0.02047 | ENSMUS | Igfbp5/Cilp/Igfbp3/Igfbp6/Igfbp4/Eif2ak3/Igf1r/Igf1 |
| BP | GO:004568 | positive regulation of glial cell differentiation | 10\|1209 | 48\|16097 | 0.00259 | 0.02097 | ENSMUS | Tlr2/Rtn4/Il6st/Enpp2/Rela/Notch1/Bag1/Dag1/Ptn/Hes1 |
| BP | GO:005087 | brown fat cell differentiation | 10\|1209 | 48\|16097 | 0.00259 | 0.02097 | ENSMUS | Rgs2/Scd1/Fabp4/Cebpb/Dusp10/Zbtb7b/Ero1l/Ebf2/Ptgs2/Fto |
| BP | GO:006067 | ureteric bud morphogenesis | 12\|1209 | 64\|16097 | 0.0026 | 0.02098 | ENSMUS | Myc/Sox9/Lzts2/Agtr2/Maged1/Nog/Cd44/Vegfa/Pbx1/Hes1/Gr |
| BP | GO:006103 | regulation of cartilage development | 12\|1209 | 64\|16097 | 0.0026 | 0.02098 | ENSMUS | Sox9/Cyr61/Ctsk/Bmp6/Rflna/Ctgf/Nog/Tgfbr1/Chadl/Rela/Loxl |
| BP | GO:000736 | gastrulation | 22\|1209 | 154\|16097 | 0.00262 | 0.02109 | ENSMUS | Klf4/Itga5/Inhba/Col5a2/Fgfr1/Col12a1/Phldb1/Sfrp2/Nog/Cdc |
| BP | GO:000317 | heart valve development | 11\|1209 | 56\|16097 | 0.00264 | 0.02123 | ENSMUS | Sox9/Efna1/Cyr61/Tnfrsf1a/Hectd1/Robo1/Nfatc1/Notch1/Rob |
| BP | GO:007020 | protein trimerization | 11\|1209 | 56\|16097 | 0.00264 | 0.02123 | ENSMUS | Col1a1/Col1a2/Col6a2/Steap4/Ppp2ca/C1qtnf3/Ski/C1qtnf6/N |
| BP | GO:009013 | epithelial cell-cell adhesion | 5\|1209 | 14\|16097 | 0.00267 | 0.02137 | ENSMUS | Thbs4/Kifc3/Itgb5/Ctnna1/Nov |
| BP | GO:190155 | regulation of endothelial cell development | 5\|1209 | 14\|16097 | 0.00267 | 0.02137 | ENSMUS | Cdh5/Tnfrsf1a/Vegfa/Cldn5/Akap11 |
| BP | GO:190314 | regulation of establishment of endothelial barrier | 5\|1209 | 14\|16097 | 0.00267 | 0.02137 | ENSMUS | Cdh5/Tnfrsf1a/Vegfa/Cldn5/Akap11 |
| BP | GO:004325 | regulation of protein complex assembly | 49\|1209 | 434\|16097 | 0.00271 | 0.02163 | ENSMUS | Fosl1/Tlr2/Eln/Fnip2/Piezo1/Met/Pex5/Jun/Cdh5/Eif4ebp1/Pec |
| BP | GO:000279 | negative regulation of peptide secretion | 21\|1209 | 145\|16097 | 0.00274 | 0.02179 | ENSMUS | Hmgcr/Cyp51/Cd34/Cx3cl1/C1qtnf3/Neo1/Gas6/Ssc5d/Rabgef1 |

| Count | Up | Up_Gene_id | Down | Down_Gene_id |
|---|---|---|---|---|
| 20 | 8 | ENSMUSG0 | 12 | ENSMUSG00000040152/ENSMUSG00000037411/ENSMUSG00000031565/ENSMUSG00000009376/ENSMUSG00000031778/ENSMUSG00000002489/ENSMUSG0 |
| 9 | 3 | ENSMUSG0 | 6 | ENSMUSG00000017929/ENSMUSG00000018736/ENSMUSG00000035764/ENSMUSG00000020745/ENSMUSG00000050965/ENSMUSG00000026826 |
| 16 | 5 | ENSMUSG0 | 11 | ENSMUSG00000026596/ENSMUSG00000024413/ENSMUSG00000020745/ENSMUSG00000061436/ENSMUSG00000063142/ENSMUSG00000000184/ENSMUSG0 |
| 16 | 6 | ENSMUSG0 | 10 | ENSMUSG00000022114/ENSMUSG00000028128/ENSMUSG00000040152/ENSMUSG00000020458/ENSMUSG00000031778/ENSMUSG00000031451/ENSMUSG0 |
| 8 | 2 | ENSMUSG0 | 6 | ENSMUSG00000031565/ENSMUSG00000030849/ENSMUSG00000032064/ENSMUSG00000056758/ENSMUSG00000030852/ENSMUSG00000065954 |
| 8 | 3 | ENSMUSG0 | 5 | ENSMUSG00000040152/ENSMUSG00000009376/ENSMUSG00000031778/ENSMUSG00000030530/ENSMUSG00000032487 |
| 8 | 0 | | 8 | ENSMUSG00000027954/ENSMUSG00000026259/ENSMUSG00000028161/ENSMUSG00000062232/ENSMUSG00000018965/ENSMUSG00000044147/ENSMUSG0 |
| 6 | 2 | ENSMUSG0 | 4 | ENSMUSG00000040152/ENSMUSG00000027995/ENSMUSG00000024789/ENSMUSG00000016024 |
| 25 | 12 | ENSMUSG0 | 13 | ENSMUSG00000026596/ENSMUSG00000028525/ENSMUSG00000074582/ENSMUSG00000002489/ENSMUSG00000020889/ENSMUSG00000096054/ENSMUSG0 |
| 14 | 4 | ENSMUSG0 | 10 | ENSMUSG00000026360/ENSMUSG00000031565/ENSMUSG00000030849/ENSMUSG00000029287/ENSMUSG00000048616/ENSMUSG00000007613/ENSMUSG0 |
| 31 | 13 | ENSMUSG0 | 18 | ENSMUSG00000022114/ENSMUSG00000000567/ENSMUSG00000038119/ENSMUSG00000031565/ENSMUSG00000038615/ENSMUSG00000021540/ENSMUSG0 |
| 50 | 16 | ENSMUSG0 | 34 | ENSMUSG00000040152/ENSMUSG00000005413/ENSMUSG00000032193/ENSMUSG00000022346/ENSMUSG00000000567/ENSMUSG00000032501/ENSMUSG0 |
| 15 | 6 | ENSMUSG0 | 9 | ENSMUSG00000028128/ENSMUSG00000005413/ENSMUSG00000031565/ENSMUSG00000039004/ENSMUSG00000004040/ENSMUSG00000023034/ENSMUSG0 |
| 21 | 1 | ENSMUSG0 | 20 | ENSMUSG00000027678/ENSMUSG00000046711/ENSMUSG00000015846/ENSMUSG00000004040/ENSMUSG00000023034/ENSMUSG00000030852/ENSMUSG0 |
| 10 | 7 | ENSMUSG0 | 3 | ENSMUSG00000020458/ENSMUSG00000023175/ENSMUSG00000021728 |
| 17 | 6 | ENSMUSG0 | 11 | ENSMUSG00000050335/ENSMUSG00000031668/ENSMUSG00000026596/ENSMUSG00000002489/ENSMUSG00000020349/ENSMUSG00000032715/ENSMUSG0 |
| 55 | 6 | ENSMUSG0 | 49 | ENSMUSG00000021670/ENSMUSG00000050931/ENSMUSG00000020000/ENSMUSG00000031770/ENSMUSG00000037071/ENSMUSG00000021273/ENSMUSG0 |
| 24 | 6 | ENSMUSG0 | 18 | ENSMUSG00000028967/ENSMUSG00000028214/ENSMUSG00000026360/ENSMUSG00000030536/ENSMUSG00000029467/ENSMUSG00000001025/ENSMUSG0 |
| 33 | 10 | ENSMUSG0 | 23 | ENSMUSG00000021670/ENSMUSG00000041324/ENSMUSG00000031565/ENSMUSG00000029482/ENSMUSG00000056758/ENSMUSG00000039004/ENSMUSG0 |
| 20 | 12 | ENSMUSG0 | 8 | ENSMUSG00000030849/ENSMUSG00000015846/ENSMUSG00000029287/ENSMUSG00000027996/ENSMUSG00000048616/ENSMUSG00000007613/ENSMUSG0 |
| 20 | 7 | ENSMUSG0 | 13 | ENSMUSG00000040152/ENSMUSG00000022346/ENSMUSG00000041324/ENSMUSG00000036390/ENSMUSG00000023067/ENSMUSG00000025408/ENSMUSG0 |
| 7 | 0 | | 7 | ENSMUSG00000037447/ENSMUSG00000022096/ENSMUSG00000109511/ENSMUSG00000034297/ENSMUSG00000034201/ENSMUSG00000065954/ENSMUSG0 |
| 9 | 3 | ENSMUSG0 | 6 | ENSMUSG00000025402/ENSMUSG00000002881/ENSMUSG00000029050/ENSMUSG00000005125/ENSMUSG00000024927/ENSMUSG00000039952 |
| 9 | 2 | ENSMUSG0 | 7 | ENSMUSG00000000567/ENSMUSG00000022367/ENSMUSG00000048537/ENSMUSG00000026596/ENSMUSG00000030341/ENSMUSG00000048661/ENSMUSG0 |
| 9 | 2 | ENSMUSG0 | 7 | ENSMUSG00000033161/ENSMUSG00000029467/ENSMUSG00000031778/ENSMUSG00000024789/ENSMUSG00000034765/ENSMUSG00000050965/ENSMUSG0 |
| 7 | 2 | ENSMUSG0 | 5 | ENSMUSG00000033161/ENSMUSG00000029467/ENSMUSG00000024789/ENSMUSG00000050965/ENSMUSG00000029361 |
| 7 | 2 | ENSMUSG0 | 5 | ENSMUSG00000022346/ENSMUSG00000031668/ENSMUSG00000022708/ENSMUSG00000015839/ENSMUSG00000050697 |
| 19 | 2 | ENSMUSG0 | 17 | ENSMUSG00000021670/ENSMUSG00000020000/ENSMUSG00000001467/ENSMUSG00000021273/ENSMUSG00000093930/ENSMUSG00000045294/ENSMUSG0 |
| 18 | 7 | ENSMUSG0 | 11 | ENSMUSG00000038119/ENSMUSG00000009376/ENSMUSG00000022951/ENSMUSG00000050567/ENSMUSG00000020484/ENSMUSG00000032340/ENSMUSG0 |
| 18 | 9 | ENSMUSG0 | 9 | ENSMUSG00000000567/ENSMUSG00000041324/ENSMUSG00000046186/ENSMUSG00000030849/ENSMUSG00000024927/ENSMUSG00000053797/ENSMUSG0 |
| 18 | 9 | ENSMUSG0 | 9 | ENSMUSG00000000567/ENSMUSG00000041324/ENSMUSG00000046186/ENSMUSG00000030849/ENSMUSG00000024927/ENSMUSG00000053797/ENSMUSG0 |
| 16 | 8 | ENSMUSG0 | 8 | ENSMUSG00000028967/ENSMUSG00000027995/ENSMUSG00000038418/ENSMUSG00000031778/ENSMUSG00000024789/ENSMUSG00000031451/ENSMUSG0 |
| 16 | 8 | ENSMUSG0 | 8 | ENSMUSG00000022346/ENSMUSG00000000567/ENSMUSG00000035342/ENSMUSG00000048616/ENSMUSG00000005087/ENSMUSG00000023951/ENSMUSG0 |
| 10 | 3 | ENSMUSG0 | 7 | ENSMUSG00000038418/ENSMUSG00000036390/ENSMUSG00000037465/ENSMUSG00000056501/ENSMUSG00000026135/ENSMUSG00000027551/ENSMUSG0 |
| 17 | 6 | ENSMUSG0 | 11 | ENSMUSG00000000567/ENSMUSG00000031565/ENSMUSG00000009376/ENSMUSG00000026596/ENSMUSG00000015846/ENSMUSG00000027996/ENSMUSG0 |
| 8 | 4 | ENSMUSG0 | 4 | ENSMUSG00000000567/ENSMUSG00000048616/ENSMUSG00000007613/ENSMUSG00000026836 |
| 8 | 2 | ENSMUSG0 | 6 | ENSMUSG00000022114/ENSMUSG00000000567/ENSMUSG00000030849/ENSMUSG00000027996/ENSMUSG00000074221/ENSMUSG00000031548 |
| 8 | 0 | | 8 | ENSMUSG00000046062/ENSMUSG00000042406/ENSMUSG00000031668/ENSMUSG00000025408/ENSMUSG00000029467/ENSMUSG00000045294/ENSMUSG0 |
| 8 | 6 | ENSMUSG0 | 2 | ENSMUSG00000031668/ENSMUSG00000005533 |
| 10 | 4 | ENSMUSG0 | 6 | ENSMUSG00000027995/ENSMUSG00000020458/ENSMUSG00000021756/ENSMUSG00000022425/ENSMUSG00000024927/ENSMUSG00000039952 |
| 10 | 3 | ENSMUSG0 | 7 | ENSMUSG00000026360/ENSMUSG00000037071/ENSMUSG00000056501/ENSMUSG00000039384/ENSMUSG00000028042/ENSMUSG00000021831/ENSMUSG0 |
| 12 | 5 | ENSMUSG0 | 7 | ENSMUSG00000022346/ENSMUSG00000000567/ENSMUSG00000035342/ENSMUSG00000048616/ENSMUSG00000005087/ENSMUSG00000023951/ENSMUSG0 |
| 12 | 4 | ENSMUSG0 | 8 | ENSMUSG00000000567/ENSMUSG00000039004/ENSMUSG00000019997/ENSMUSG00000048616/ENSMUSG00000007613/ENSMUSG00000024927/ENSMUSG0 |
| 22 | 5 | ENSMUSG0 | 17 | ENSMUSG00000003032/ENSMUSG00000000555/ENSMUSG00000041324/ENSMUSG00000031565/ENSMUSG00000048537/ENSMUSG00000027996/ENSMUSG0 |
| 11 | 6 | ENSMUSG0 | 5 | ENSMUSG00000000567/ENSMUSG00000027954/ENSMUSG00000030341/ENSMUSG00000035247/ENSMUSG00000026836 |
| 11 | 6 | ENSMUSG0 | 5 | ENSMUSG00000020241/ENSMUSG00000020349/ENSMUSG00000029050/ENSMUSG00000109511/ENSMUSG00000001119 |
| 5 | 2 | ENSMUSG0 | 3 | ENSMUSG00000031788/ENSMUSG00000022817/ENSMUSG00000037815 |
| 5 | 2 | ENSMUSG0 | 3 | ENSMUSG00000030341/ENSMUSG00000023951/ENSMUSG00000022016 |
| 5 | 2 | ENSMUSG0 | 3 | ENSMUSG00000030341/ENSMUSG00000023951/ENSMUSG00000022016 |
| 49 | 16 | ENSMUSG0 | 33 | ENSMUSG00000024912/ENSMUSG00000027995/ENSMUSG00000029675/ENSMUSG00000061175/ENSMUSG00000014444/ENSMUSG00000009376/ENSMUSG0 |
| 21 | 7 | ENSMUSG0 | 14 | ENSMUSG00000021670/ENSMUSG00000001467/ENSMUSG00000031778/ENSMUSG00000032340/ENSMUSG00000031451/ENSMUSG00000025340/ENSMUSG0 |

0000031451/ENSMUSG00000021508/ENSMUSG00000023951/ENSMUSG00000050965/ENSMUSG00000003814/ENSMUSG00000016024

0000031393/ENSMUSG00000026826/ENSMUSG00000026812/ENSMUSG00000024182/ENSMUSG00000032096
0000007613/ENSMUSG00000005533/ENSMUSG00000023951/ENSMUSG00000024486

0000031393/ENSMUSG00000025326

0000039953/ENSMUSG00000028161/ENSMUSG00000024431/ENSMUSG00000019943/ENSMUSG00000025372/ENSMUSG00000005886/ENSMUSG00000025979
0000033323/ENSMUSG00000022752/ENSMUSG00000024431/ENSMUSG00000005871
0000030849/ENSMUSG00000004040/ENSMUSG00000027996/ENSMUSG00000026361/ENSMUSG00000007613/ENSMUSG00000056501/ENSMUSG00000040021/ENSMUSG00000018476/ENSMUSG0000002
0000030111/ENSMUSG00000020823/ENSMUSG00000024359/ENSMUSG00000050335/ENSMUSG00000039899/ENSMUSG00000031778/ENSMUSG00000022508/ENSMUSG00000020889/ENSMUSG0000002
0000023951/ENSMUSG00000050965/ENSMUSG00000000058
0000056501/ENSMUSG00000037447/ENSMUSG00000022096/ENSMUSG00000048490/ENSMUSG00000040021/ENSMUSG00000054808/ENSMUSG00000109511/ENSMUSG00000003814/ENSMUSG0000003

0000061436/ENSMUSG00000022365/ENSMUSG00000062232/ENSMUSG00000044147/ENSMUSG00000009630
0000031604/ENSMUSG00000020458/ENSMUSG00000038615/ENSMUSG00000022463/ENSMUSG00000031668/ENSMUSG00000025203/ENSMUSG00000030062/ENSMUSG00000029467/ENSMUSG0000004
0000002489/ENSMUSG00000038764/ENSMUSG00000059991/ENSMUSG00000028161/ENSMUSG00000020132/ENSMUSG00000026131/ENSMUSG00000000058/ENSMUSG00000019943/ENSMUSG0000003
0000002489/ENSMUSG00000024789/ENSMUSG00000070645/ENSMUSG00000020889/ENSMUSG00000030059/ENSMUSG00000028161/ENSMUSG00000018634/ENSMUSG00000033717/ENSMUSG0000002
0000035247/ENSMUSG00000026836
0000040463/ENSMUSG00000003814/ENSMUSG00000034201/ENSMUSG00000028211/ENSMUSG00000042303/ENSMUSG00000042903/ENSMUSG00000005871
0000005886

0000039952
0000029361

0000039004/ENSMUSG00000026675/ENSMUSG00000034926/ENSMUSG00000023068/ENSMUSG00000032018/ENSMUSG00000033105/ENSMUSG00000003752/ENSMUSG00000021036/ENSMUSG0000002
0000029050/ENSMUSG00000022443/ENSMUSG00000028161/ENSMUSG00000026313/ENSMUSG00000000058
0000032487/ENSMUSG00000005871/ENSMUSG00000034593
0000032487/ENSMUSG00000005871/ENSMUSG00000034593
0000037012/ENSMUSG00000037902
0000074934/ENSMUSG00000032220
0000029249
0000048616/ENSMUSG00000023951/ENSMUSG00000031548/ENSMUSG00000074934/ENSMUSG00000041702

0000028042/ENSMUSG00000015839

0000032487
0000074934
0000034205/ENSMUSG00000074934
0000048616/ENSMUSG00000026361/ENSMUSG00000024098/ENSMUSG00000074221/ENSMUSG00000018476/ENSMUSG00000029201/ENSMUSG00000006445/ENSMUSG00000039952/ENSMUSG0000002

0000005069/ENSMUSG00000031490/ENSMUSG00000045294/ENSMUSG00000019970/ENSMUSG00000038400/ENSMUSG00000045983/ENSMUSG00000006585/ENSMUSG00000023951/ENSMUSG0000004
0000028161/ENSMUSG00000033717/ENSMUSG00000029249/ENSMUSG00000029361/ENSMUSG00000031548/ENSMUSG00000035828/ENSMUSG00000035621/ENSMUSG00000027712

28042/ENSMUSG00000026836/ENSMUSG00000031548/ENSMUSG00000005871
26361/ENSMUSG00000056501/ENSMUSG00000052040/ENSMUSG00000005087/ENSMUSG00000025340/ENSMUSG00000024098/ENSMUSG00000039384/ENSMUSG00000020682/ENSMUSG00000024812/E

34297/ENSMUSG00000018965/ENSMUSG00000034201/ENSMUSG00000065954/ENSMUSG00000026826/ENSMUSG00000005886

42644/ENSMUSG00000045294/ENSMUSG00000020484/ENSMUSG00000014959/ENSMUSG00000047187/ENSMUSG00000030530/ENSMUSG00000079111/ENSMUSG00000039100/ENSMUSG00000023068/E
31453/ENSMUSG00000027366/ENSMUSG00000002365/ENSMUSG00000031543
29249/ENSMUSG00000029361/ENSMUSG00000031548/ENSMUSG00000035828/ENSMUSG00000020684/ENSMUSG00000021794/ENSMUSG00000035621/ENSMUSG00000027712/ENSMUSG00000022808

29249/ENSMUSG00000050697/ENSMUSG00000024210

26836/ENSMUSG00000031548/ENSMUSG00000024182

40021/ENSMUSG00000018583/ENSMUSG00000024533/ENSMUSG00000024423/ENSMUSG00000032702/ENSMUSG00000022037/ENSMUSG00000041570/ENSMUSG00000038859/ENSMUSG00000035992/E

ENSMUSG00000028042/ENSMUSG00000079037/ENSMUSG00000056050/ENSMUSG00000029816/ENSMUSG00000034201/ENSMUSG00000035900/ENSMUSG00000015839/ENSMUSG00000040025/ENSMUS

ENSMUSG00000027465/ENSMUSG00000021770/ENSMUSG00000032018/ENSMUSG00000022270/ENSMUSG00000021831/ENSMUSG00000021427/ENSMUSG00000030747/ENSMUSG00000032349/ENSMUS

ENSMUSG00000044147/ENSMUSG00000026797/ENSMUSG00000031393/ENSMUSG00000050697/ENSMUSG00000025372/ENSMUSG00000002083/ENSMUSG00000034216/ENSMUSG00000002365/ENSMUS

;G00000031548/ENSMUSG00000024425/ENSMUSG00000074934

;G00000109511/ENSMUSG00000036564/ENSMUSG00000021036/ENSMUSG00000026924/ENSMUSG00000003814/ENSMUSG00000056050/ENSMUSG00000022365/ENSMUSG00000026131/ENSMUSG0000(

;G00000031386/ENSMUSG00000005871

0030835/ENSMUSG00000027230/ENSMUSG00000024052/ENSMUSG00000075703/ENSMUSG00000029361/ENSMUSG00000005615/ENSMUSG00000030287/ENSMUSG00000024807/ENSMUSG0000004755

4/ENSMUSG00000027366

| Category | GOID | Description | GeneRatio | BgRatio | pvalue | padj | geneID | geneName |
|---|---|---|---|---|---|---|---|---|
| BP | GO:001095 | positive regulation of endopeptidase activity | 21\|1209 | 145\|16097 | 0.00274 | 0.02179 | ENSMUS | F3/Myc/Efna1/Eif2ak3/Cyr61/Stat3/Ctgf/Ppp2ca/Jak2/Sfrp2/Dl |
| BP | GO:000194 | lymphangiogenesis | 6\|1209 | 20\|16097 | 0.00275 | 0.02182 | ENSMUS | Flt4/Vegfa/Nfatc1/Vash1/Epha2/Ptpn14 |
| BP | GO:000327 | cardiac septum development | 18\|1209 | 117\|16097 | 0.00275 | 0.02182 | ENSMUS | Cyr61/Fgfr2/Maml1/Tgfbr3/Nrp1/Ltbp1/Nog/Tgfbr1/Hectd1/Ro |
| CC | GO:003001 | sarcomere | 25\|1232 | 183\|16313 | 0.0028 | 0.03965 | ENSMUS | Acta1/Slc2a1/Tpm1/Tpm2/Pde4b/Synpo/Tnni2/Ldb3/Actn2/Sy |
| BP | GO:002240 | regulation of cell-cell adhesion | 43\|1209 | 370\|16097 | 0.00281 | 0.02229 | ENSMUS | Klf4/Piezo1/Has2/Il6ra/Lgals3/Lgals1/Abl2/Fgl2/Bmp6/Cx3cl1/T |
| BP | GO:004568 | regulation of epidermis development | 13\|1209 | 73\|16097 | 0.00283 | 0.02238 | ENSMUS | Errfi1/Inhba/Cd109/Mafg/Ncoa3/Nab2/Nab1/Notch1/Sfrp4/Sfn |
| BP | GO:006097 | coronary vasculature development | 13\|1209 | 73\|16097 | 0.00283 | 0.02238 | ENSMUS | Fgfr1/Spred1/Fgfr2/Rxra/Tgfbr3/Nrp1/Ltbp1/Tgfbr1/Robo1/Veg |
| CC | GO:003001 | Z disc | 19\|1232 | 126\|16313 | 0.00286 | 0.03965 | ENSMUS | Slc2a1/Pde4b/Synpo/Ldb3/Actn2/Actn4/Fhl3/Ppp3ca/Hdac4/D |
| BP | GO:000329 | physiological muscle hypertrophy | 9\|1209 | 41\|16097 | 0.00288 | 0.02264 | ENSMUS | Col14a1/Rgs2/Sorbs2/Agtr2/Ctdp1/Igf1/Tomm70a/Pi16/Nr3c1 |
| BP | GO:000330 | physiological cardiac muscle hypertrophy | 9\|1209 | 41\|16097 | 0.00288 | 0.02264 | ENSMUS | Col14a1/Rgs2/Sorbs2/Agtr2/Ctdp1/Igf1/Tomm70a/Pi16/Nr3c1 |
| BP | GO:000641 | tRNA aminoacylation for protein translation | 9\|1209 | 41\|16097 | 0.00288 | 0.02264 | ENSMUS | Aars/Gars/Sars/Nars/Tars/Yars/Iars/Dars/Cars |
| BP | GO:006104 | cell growth involved in cardiac muscle cell development | 9\|1209 | 41\|16097 | 0.00288 | 0.02264 | ENSMUS | Col14a1/Rgs2/Sorbs2/Agtr2/Ctdp1/Igf1/Tomm70a/Pi16/Nr3c1 |
| CC | GO:007016 | tight junction | 18\|1232 | 117\|16313 | 0.00292 | 0.03965 | ENSMUS | Cdh5/Cldn12/Synpo/Amotl2/Usp53/Cgnl1/Tgfbr1/Esam/Ccnd1/ |
| BP | GO:007217 | mesonephric tubule morphogenesis | 12\|1209 | 65\|16097 | 0.00298 | 0.02335 | ENSMUS | Myc/Sox9/Lzts2/Agtr2/Maged1/Nog/Cd44/Vegfa/Pbx1/Hes1/Gr |
| BP | GO:006041 | muscle tissue morphogenesis | 14\|1209 | 82\|16097 | 0.00298 | 0.02337 | ENSMUS | Col3a1/Tpm1/Fgfr2/Rxra/Mylk/Tgfbr3/Nog/Tgfbr1/Notch1/Col1 |
| BP | GO:009874 | cell-cell adhesion via plasma-membrane adhesion molecules | 25\|1209 | 185\|16097 | 0.00301 | 0.02359 | ENSMUS | Klf4/Itga5/Cdh5/Lgals1/Pecam1/Cdh11/Mpz/Sdk2/Cx3cl1/Robo |
| CC | GO:004430 | neuron spine | 25\|1232 | 184\|16313 | 0.00302 | 0.04007 | ENSMUS | Cald1/Lama2/Dnm1/Abl2/Pde4b/Arfgef2/Synpo/Tiam1/Nr1d1/ |
| BP | GO:004513 | development of primary sexual characteristics | 26\|1209 | 195\|16097 | 0.00304 | 0.02372 | ENSMUS | Sox9/Serpine1/Inhba/Adgrg1/Hmga2/Tiparp/Sfrp2/Ren1/Tgfbr1 |
| BP | GO:007213 | mesenchyme morphogenesis | 10\|1209 | 49\|16097 | 0.00304 | 0.02373 | ENSMUS | Myc/Sox9/Fgfr1/Nog/Tgfbr1/Robo1/Notch1/Robo2/Acvr1/Mdm |
| BP | GO:000834 | glial cell migration | 11\|1209 | 57\|16097 | 0.00306 | 0.02373 | ENSMUS | Lamb1/Rtn4/Vcan/Adgrg1/Tiam1/Lrp8/Vim/Pafah1b1/Epha4/Fu |
| BP | GO:005114 | negative regulation of muscle cell differentiation | 11\|1209 | 57\|16097 | 0.00306 | 0.02373 | ENSMUS | Rgs2/Xbp1/Ctdp1/Tomm70a/Pi16/Nfatc1/Notch1/Hdac4/Nov/ |
| BP | GO:007145 | cellular response to hypoxia | 15\|1209 | 91\|16097 | 0.00306 | 0.02373 | ENSMUS | Myc/Vasn/Egr1/Eif4ebp1/Rtn4/Hyou1/Cd34/Fndc1/Vegfa/Notc |
| BP | GO:004867 | regulation of axon regeneration | 8\|1209 | 34\|16097 | 0.00308 | 0.02373 | ENSMUS | Klf4/Xylt1/Rtn4/Ndel1/Neo1/Igf1r/Epha4/Ptn |
| BP | GO:006138 | heart trabecula morphogenesis | 8\|1209 | 34\|16097 | 0.00308 | 0.02373 | ENSMUS | Tgfbr3/Nog/Tgfbr1/Nfatc1/Notch1/Adgrg6/Adamts1/Tek |
| BP | GO:007160 | transforming growth factor beta production | 8\|1209 | 34\|16097 | 0.00308 | 0.02373 | ENSMUS | Thbs1/Met/Cd34/Cx3cl1/Ltbp1/Furin/Lum/Ptgs2 |
| BP | GO:005105 | regulation of small GTPase mediated signal transduction | 32\|1209 | 256\|16097 | 0.00308 | 0.02373 | ENSMUS | Col3a1/Spry2/Cdon/Tiam2/Met/Sipa1l2/Rtn4/Abl2/Adgrg1/Ng |
| BP | GO:003629 | cellular response to decreased oxygen levels | 16\|1209 | 100\|16097 | 0.00309 | 0.02373 | ENSMUS | Myc/Vasn/Egr1/Eif4ebp1/Rtn4/Hyou1/Cd34/Fndc1/Vegfa/Notc |
| BP | GO:001097 | positive regulation of G2/M transition of mitotic cell cycle | 7\|1209 | 27\|16097 | 0.00309 | 0.02373 | ENSMUS | Ccnd1/Pbx1/Rcc2/Vps4b/App/Mecp2/Fbxo5 |
| BP | GO:005509 | acylglycerol homeostasis | 7\|1209 | 27\|16097 | 0.00309 | 0.02373 | ENSMUS | Lpl/Sesn2/Xbp1/C1qtnf3/Angptl4/Dgat2/Med13 |
| BP | GO:006121 | regulation of mesonephros development | 7\|1209 | 27\|16097 | 0.00309 | 0.02373 | ENSMUS | Sox9/Agtr2/Maged1/Nog/Vegfa/Grem1/Sox8 |
| BP | GO:007032 | triglyceride homeostasis | 7\|1209 | 27\|16097 | 0.00309 | 0.02373 | ENSMUS | Lpl/Sesn2/Xbp1/C1qtnf3/Angptl4/Dgat2/Med13 |
| BP | GO:001081 | regulation of hormone levels | 52\|1209 | 470\|16097 | 0.00313 | 0.02394 | ENSMUS | Hmgcr/Inhba/Egr1/Fgfr1/Igfbp3/Aacs/Atp1a1/Pcsk6/Hmga2/Bm |
| BP | GO:004583 | positive regulation of lipid metabolic process | 20\|1209 | 138\|16097 | 0.00338 | 0.02585 | ENSMUS | Ldlr/Stard4/Scd1/Acsl3/Cyr61/Bmp6/Adgrf5/Nr1d1/Cd36/Angp |
| BP | GO:004566 | positive regulation of osteoblast differentiation | 12\|1209 | 66\|16097 | 0.00339 | 0.02591 | ENSMUS | Cyr61/Lmna/Smad5/Bmp6/Il6st/Sfrp2/Cebpb/Igf1/Vegfa/Acvr1/ |
| BP | GO:200037 | positive regulation of reactive oxygen species metabolic proces | 16\|1209 | 101\|16097 | 0.00342 | 0.02606 | ENSMUS | Thbs1/Tlr2/Klf4/Gadd45a/Cdkn1a/Rnf41/Egfr/Jak2/Cd36/Ddah |
| BP | GO:000156 | branching involved in blood vessel morphogenesis | 9\|1209 | 42\|16097 | 0.00343 | 0.02606 | ENSMUS | Col4a1/Nrp1/Sfrp2/Cxcl12/Vegfa/Plxnd1/Tek/Acvr1/Notch4 |
| BP | GO:190204 | regulation of extrinsic apoptotic signaling pathway via death d | 9\|1209 | 42\|16097 | 0.00343 | 0.02606 | ENSMUS | Thbs1/Hmox1/Serpine1/Fem1b/Lgals3/Sfrp2/Timp3/Sfrp1/Atf3 |
| BP | GO:006039 | regulation of pathway-restricted SMAD protein phosphorylatic | 11\|1209 | 58\|16097 | 0.00352 | 0.02674 | ENSMUS | Inhba/Bmp6/Xbp1/Pmepa1/Nog/Tgfbr1/Twsg1/Dab2/Lgals9/Ac |
| BP | GO:004866 | negative regulation of smooth muscle cell proliferation | 10\|1209 | 50\|16097 | 0.00355 | 0.02692 | ENSMUS | Hmox1/Igfbp5/Trib1/Klf4/Rgs5/Igfbp3/Cdkn1a/Vipr2/Npr1/Ndr |
| CC | GO:000592 | bicellular tight junction | 17\|1232 | 110\|16313 | 0.00358 | 0.04578 | ENSMUS | Cdh5/Cldn12/Synpo/Amotl2/Usp53/Cgnl1/Tgfbr1/Esam/Ccnd1/ |
| CC | GO:003167 | I band | 20\|1232 | 138\|16313 | 0.0036 | 0.04578 | ENSMUS | Slc2a1/Pde4b/Synpo/Ldb3/Actn2/Actn4/Fhl3/Ppp3ca/Hdac4/D |
| BP | GO:003630 | lymph vessel morphogenesis | 6\|1209 | 21\|16097 | 0.0036 | 0.02728 | ENSMUS | Flt4/Vegfa/Nfatc1/Vash1/Epha2/Ptpn14 |
| BP | GO:006039 | SMAD protein signal transduction | 13\|1209 | 75\|16097 | 0.00362 | 0.02734 | ENSMUS | Cilp/Inhba/Jun/Smad5/Fos/Bmp6/Ski/Lnpep/Tgfbr1/Vim/Hipk2 |
| CC | GO:001645 | myosin complex | 11\|1232 | 58\|16313 | 0.00367 | 0.04578 | ENSMUS | Myl1/Mylpf/Myh11/Cgnl1/Myh9/Myh14/Ttn/Limch1/Myl9/My |
| BP | GO:000257 | myeloid leukocyte differentiation | 26\|1209 | 198\|16097 | 0.00374 | 0.02819 | ENSMUS | Myc/Tlr2/Trib1/Cd109/Jun/Fos/Fasn/Klf10/Camk4/Cebpb/Vegfa |
| BP | GO:000170 | formation of primary germ layer | 14\|1209 | 84\|16097 | 0.00374 | 0.02819 | ENSMUS | Itga5/Inhba/Col5a2/Fgfr1/Col12a1/Sfrp2/Nog/Cdc73/Twsg1/Kd |
| BP | GO:004688 | negative regulation of hormone secretion | 14\|1209 | 84\|16097 | 0.00374 | 0.02819 | ENSMUS | Hmgcr/Igfbp3/Ppp3ca/Crhr1/Nov/Adra2a/Rest/Nos1/Sfrp1/Ucp |
| BP | GO:004560 | positive regulation of endothelial cell differentiation | 5\|1209 | 15\|16097 | 0.00376 | 0.02821 | ENSMUS | Cdh5/Bmp6/Notch1/Cldn5/Akap11 |
| BP | GO:007140 | cellular response to low-density lipoprotein particle stimulus | 5\|1209 | 15\|16097 | 0.00376 | 0.02821 | ENSMUS | Ldlr/Lpl/Hmgcs1/Npc1/Cd36 |
| BP | GO:200009 | regulation of Wnt signaling pathway, planar cell polarity pathw | 5\|1209 | 15\|16097 | 0.00376 | 0.02821 | ENSMUS | Abl2/Sfrp2/Nkd1/Dab2/Sfrp1 |
| BP | GO:006132 | renal tubule development | 15\|1209 | 93\|16097 | 0.00379 | 0.02841 | ENSMUS | Col4a1/Myc/Sox9/Lzts2/Agtr2/Maged1/Nog/Cd44/Vegfa/Notch |

| Count | Up | Up_Gene_i | Down | Down_Gene_id |
|---|---|---|---|---|
| 21 | 6 | ENSMUSG0 | 15 | ENSMUSG00000028128/ENSMUSG00000022346/ENSMUSG00000027954/ENSMUSG00000031668/ENSMUSG00000004040/ENSMUSG00000019997/ENSMUSG0 |
| 6 | 3 | ENSMUSG0 | 3 | ENSMUSG00000023951/ENSMUSG00000006445/ENSMUSG00000026604 |
| 18 | 11 | ENSMUSG0 | 7 | ENSMUSG00000030849/ENSMUSG00000050567/ENSMUSG00000029287/ENSMUSG00000048616/ENSMUSG00000007613/ENSMUSG00000035247/ENSMUSG0 |
| 25 | 14 | ENSMUSG0 | 11 | ENSMUSG00000028645/ENSMUSG00000028525/ENSMUSG00000096054/ENSMUSG00000054808/ENSMUSG00000032643/ENSMUSG00000028161/ENSMUSG0 |
| 43 | 9 | ENSMUSG0 | 34 | ENSMUSG00000003032/ENSMUSG00000014444/ENSMUSG00000022367/ENSMUSG00000027947/ENSMUSG00000050335/ENSMUSG00000026596/ENSMUSG0 |
| 13 | 5 | ENSMUSG0 | 8 | ENSMUSG00000028967/ENSMUSG00000041324/ENSMUSG00000046186/ENSMUSG00000051510/ENSMUSG00000027678/ENSMUSG00000025402/ENSMUSG0 |
| 13 | 6 | ENSMUSG0 | 7 | ENSMUSG00000031565/ENSMUSG00000027351/ENSMUSG00000030849/ENSMUSG00000015846/ENSMUSG00000029287/ENSMUSG00000007613/ENSMUSG0 |
| 19 | 9 | ENSMUSG0 | 10 | ENSMUSG00000028645/ENSMUSG00000028525/ENSMUSG00000054808/ENSMUSG00000032643/ENSMUSG00000028161/ENSMUSG00000026313/ENSMUSG0 |
| 9 | 5 | ENSMUSG0 | 4 | ENSMUSG00000026360/ENSMUSG00000033323/ENSMUSG00000022752/ENSMUSG00000024431 |
| 9 | 5 | ENSMUSG0 | 4 | ENSMUSG00000026360/ENSMUSG00000033323/ENSMUSG00000022752/ENSMUSG00000024431 |
| 9 | 1 | ENSMUSG0 | 8 | ENSMUSG00000031960/ENSMUSG00000029777/ENSMUSG00000068739/ENSMUSG00000024587/ENSMUSG00000022241/ENSMUSG00000028811/ENSMUSG0 |
| 9 | 5 | ENSMUSG0 | 4 | ENSMUSG00000026360/ENSMUSG00000033323/ENSMUSG00000022752/ENSMUSG00000024431 |
| 18 | 7 | ENSMUSG0 | 11 | ENSMUSG00000046798/ENSMUSG00000039701/ENSMUSG00000032232/ENSMUSG00000007613/ENSMUSG00000024812/ENSMUSG00000013076/ENSMUSG0 |
| 12 | 5 | ENSMUSG0 | 7 | ENSMUSG00000022346/ENSMUSG00000000567/ENSMUSG00000035342/ENSMUSG00000048616/ENSMUSG00000005087/ENSMUSG00000023951/ENSMUSG0 |
| 14 | 9 | ENSMUSG0 | 5 | ENSMUSG00000030849/ENSMUSG00000015846/ENSMUSG00000029287/ENSMUSG00000048616/ENSMUSG00000007613 |
| 25 | 12 | ENSMUSG0 | 13 | ENSMUSG00000003032/ENSMUSG00000000555/ENSMUSG00000041592/ENSMUSG00000031778/ENSMUSG00000036606/ENSMUSG00000040511/ENSMUSG0 |
| 25 | 12 | ENSMUSG0 | 13 | ENSMUSG00000026596/ENSMUSG00000028525/ENSMUSG00000074582/ENSMUSG00000002489/ENSMUSG00000020889/ENSMUSG00000096054/ENSMUSG0 |
| 26 | 6 | ENSMUSG0 | 20 | ENSMUSG00000000567/ENSMUSG00000037411/ENSMUSG00000041324/ENSMUSG00000031785/ENSMUSG00000056758/ENSMUSG00000034640/ENSMUSG0 |
| 10 | 4 | ENSMUSG0 | 6 | ENSMUSG00000022346/ENSMUSG00000000567/ENSMUSG00000031565/ENSMUSG00000048616/ENSMUSG00000007613/ENSMUSG00000026836 |
| 11 | 5 | ENSMUSG0 | 6 | ENSMUSG00000020458/ENSMUSG00000031785/ENSMUSG00000002489/ENSMUSG00000028613/ENSMUSG00000026728/ENSMUSG00000020745 |
| 11 | 4 | ENSMUSG0 | 7 | ENSMUSG00000026360/ENSMUSG00000020484/ENSMUSG00000033323/ENSMUSG00000022752/ENSMUSG00000026313/ENSMUSG00000031393/ENSMUSG0 |
| 15 | 4 | ENSMUSG0 | 11 | ENSMUSG00000022346/ENSMUSG00000039646/ENSMUSG00000038418/ENSMUSG00000031490/ENSMUSG00000020458/ENSMUSG00000032115/ENSMUSG0 |
| 8 | 2 | ENSMUSG0 | 6 | ENSMUSG00000003032/ENSMUSG00000030657/ENSMUSG00000020458/ENSMUSG00000018736/ENSMUSG00000032340/ENSMUSG00000005533 |
| 8 | 5 | ENSMUSG0 | 3 | ENSMUSG00000029287/ENSMUSG00000048616/ENSMUSG00000007613 |
| 8 | 3 | ENSMUSG0 | 5 | ENSMUSG00000040152/ENSMUSG00000009376/ENSMUSG00000031778/ENSMUSG00000030530/ENSMUSG00000032487 |
| 32 | 9 | ENSMUSG0 | 23 | ENSMUSG00000022114/ENSMUSG00000038119/ENSMUSG00000023800/ENSMUSG00000009376/ENSMUSG00000001995/ENSMUSG00000020458/ENSMUSG0 |
| 16 | 4 | ENSMUSG0 | 12 | ENSMUSG00000022346/ENSMUSG00000039646/ENSMUSG00000038418/ENSMUSG00000031490/ENSMUSG00000020458/ENSMUSG00000032115/ENSMUSG0 |
| 7 | 4 | ENSMUSG0 | 3 | ENSMUSG00000040945/ENSMUSG00000009907/ENSMUSG00000031393 |
| 7 | 3 | ENSMUSG0 | 4 | ENSMUSG00000028893/ENSMUSG00000020484/ENSMUSG00000030747/ENSMUSG00000034297 |
| 7 | 3 | ENSMUSG0 | 4 | ENSMUSG00000000567/ENSMUSG00000048616/ENSMUSG00000023951/ENSMUSG00000074934 |
| 7 | 3 | ENSMUSG0 | 4 | ENSMUSG00000028893/ENSMUSG00000020484/ENSMUSG00000030747/ENSMUSG00000034297 |
| 52 | 16 | ENSMUSG0 | 36 | ENSMUSG00000021670/ENSMUSG00000041324/ENSMUSG00000038418/ENSMUSG00000031565/ENSMUSG00000029482/ENSMUSG00000033161/ENSMUSG0 |
| 20 | 7 | ENSMUSG0 | 13 | ENSMUSG00000032193/ENSMUSG00000024378/ENSMUSG00000037071/ENSMUSG00000032883/ENSMUSG00000039004/ENSMUSG00000020889/ENSMUSG0 |
| 12 | 4 | ENSMUSG0 | 8 | ENSMUSG00000028063/ENSMUSG00000021540/ENSMUSG00000039004/ENSMUSG00000021756/ENSMUSG00000027996/ENSMUSG00000056501/ENSMUSG0 |
| 16 | 4 | ENSMUSG0 | 12 | ENSMUSG00000040152/ENSMUSG00000027995/ENSMUSG00000003032/ENSMUSG00000036390/ENSMUSG00000023067/ENSMUSG00000025373/ENSMUSG0 |
| 9 | 6 | ENSMUSG0 | 3 | ENSMUSG00000027996/ENSMUSG00000023951/ENSMUSG00000026836 |
| 9 | 0 | | 9 | ENSMUSG00000040152/ENSMUSG00000005413/ENSMUSG00000037411/ENSMUSG00000032244/ENSMUSG00000050335/ENSMUSG00000027996/ENSMUSG0 |
| 11 | 2 | ENSMUSG0 | 9 | ENSMUSG00000041324/ENSMUSG00000039004/ENSMUSG00000020484/ENSMUSG00000038400/ENSMUSG00000048616/ENSMUSG00000007613/ENSMUSG0 |
| 10 | 5 | ENSMUSG0 | 5 | ENSMUSG00000005413/ENSMUSG00000032501/ENSMUSG00000003032/ENSMUSG00000023067/ENSMUSG00000036564 |
| 17 | 7 | ENSMUSG0 | 10 | ENSMUSG00000046798/ENSMUSG00000039701/ENSMUSG00000032232/ENSMUSG00000007613/ENSMUSG00000024812/ENSMUSG00000013076/ENSMUSG0 |
| 20 | 10 | ENSMUSG0 | 10 | ENSMUSG00000028645/ENSMUSG00000028525/ENSMUSG00000054808/ENSMUSG00000032643/ENSMUSG00000028161/ENSMUSG00000026313/ENSMUSG0 |
| 6 | 3 | ENSMUSG0 | 3 | ENSMUSG00000023951/ENSMUSG00000006445/ENSMUSG00000026604 |
| 13 | 5 | ENSMUSG0 | 8 | ENSMUSG00000041324/ENSMUSG00000021540/ENSMUSG00000039004/ENSMUSG00000029050/ENSMUSG00000023845/ENSMUSG00000007613/ENSMUSG0 |
| 11 | 7 | ENSMUSG0 | 4 | ENSMUSG00000032232/ENSMUSG00000022443/ENSMUSG00000032220/ENSMUSG00000034593 |
| 26 | 9 | ENSMUSG0 | 17 | ENSMUSG00000022346/ENSMUSG00000027995/ENSMUSG00000032501/ENSMUSG00000046186/ENSMUSG00000025153/ENSMUSG00000037465/ENSMUSG0 |
| 14 | 3 | ENSMUSG0 | 11 | ENSMUSG00000000555/ENSMUSG00000041324/ENSMUSG00000031565/ENSMUSG00000027996/ENSMUSG00000048616/ENSMUSG00000026361/ENSMUSG0 |
| 14 | 4 | ENSMUSG0 | 10 | ENSMUSG00000021670/ENSMUSG00000028161/ENSMUSG00000018634/ENSMUSG00000033717/ENSMUSG00000029249/ENSMUSG00000029361/ENSMUSG0 |
| 5 | 3 | ENSMUSG0 | 2 | ENSMUSG00000039004/ENSMUSG00000022016 |
| 5 | 2 | ENSMUSG0 | 3 | ENSMUSG00000032193/ENSMUSG00000093930/ENSMUSG00000024413 |
| 5 | 2 | ENSMUSG0 | 3 | ENSMUSG00000026596/ENSMUSG00000027996/ENSMUSG00000031548 |
| 15 | 8 | ENSMUSG0 | 7 | ENSMUSG00000022346/ENSMUSG00000000567/ENSMUSG00000035342/ENSMUSG00000048616/ENSMUSG00000005087/ENSMUSG00000023951/ENSMUSG0 |

0000020349/ENSMUSG00000024789/ENSMUSG00000027996/ENSMUSG00000031523/ENSMUSG00000035199/ENSMUSG00000029249/ENSMUSG00000035900/ENSMUSG00000056071/ENSMUSG0000000

0000026836
0000026313/ENSMUSG00000026131/ENSMUSG00000029361/ENSMUSG00000027712/ENSMUSG00000031543
0000039899/ENSMUSG00000039004/ENSMUSG00000031778/ENSMUSG00000022508/ENSMUSG00000020484/ENSMUSG00000021756/ENSMUSG00000007817/ENSMUSG00000024789/ENSMUSG0000000
0000002881/ENSMUSG00000047281
0000023951
0000026131/ENSMUSG00000029361/ENSMUSG00000027712/ENSMUSG00000031543

0000037851/ENSMUSG00000010755

0000006445/ENSMUSG00000039473/ENSMUSG00000062232/ENSMUSG00000045394/ENSMUSG00000005871
0000074934

0000039953/ENSMUSG00000023175/ENSMUSG00000032593/ENSMUSG00000026836/ENSMUSG00000068740/ENSMUSG00000045394/ENSMUSG00000021728
0000039953/ENSMUSG00000028161/ENSMUSG00000024431/ENSMUSG00000019943/ENSMUSG00000025372/ENSMUSG00000005886/ENSMUSG00000025979
0000027996/ENSMUSG00000070645/ENSMUSG00000007613/ENSMUSG00000056501/ENSMUSG00000023951/ENSMUSG00000048490/ENSMUSG00000030059/ENSMUSG00000079509/ENSMUSG0000000

0000042903
0000023951/ENSMUSG00000024924/ENSMUSG00000036450/ENSMUSG00000015839/ENSMUSG00000025612

0000026596/ENSMUSG00000031785/ENSMUSG00000026259/ENSMUSG00000074582/ENSMUSG00000002489/ENSMUSG00000022508/ENSMUSG00000031523/ENSMUSG00000032232/ENSMUSG0000000
0000023951/ENSMUSG00000024924/ENSMUSG00000036450/ENSMUSG00000015839/ENSMUSG00000025612/ENSMUSG00000026812

0000030513/ENSMUSG00000056758/ENSMUSG00000039004/ENSMUSG00000002489/ENSMUSG00000024789/ENSMUSG00000034640/ENSMUSG00000070645/ENSMUSG00000020889/ENSMUSG0000000
0000005533/ENSMUSG00000022708/ENSMUSG00000030747/ENSMUSG00000032349/ENSMUSG00000032487/ENSMUSG00000026623/ENSMUSG00000028661
0000023951/ENSMUSG00000026836
0000024789/ENSMUSG00000027832/ENSMUSG00000026313/ENSMUSG00000022037/ENSMUSG00000032487/ENSMUSG00000015839

0000020044/ENSMUSG00000031548/ENSMUSG00000026628
0000024098/ENSMUSG00000026836/ENSMUSG00000074934

0000039473/ENSMUSG00000062232/ENSMUSG00000045394/ENSMUSG00000005871
0000026131/ENSMUSG00000029361/ENSMUSG00000027712/ENSMUSG00000031543

0000026728/ENSMUSG00000061436

0000056501/ENSMUSG00000023951/ENSMUSG00000000134/ENSMUSG00000020745/ENSMUSG00000035673/ENSMUSG00000024812/ENSMUSG00000006445/ENSMUSG00000050965/ENSMUSG0000000
0000024098/ENSMUSG00000018476/ENSMUSG00000006445/ENSMUSG00000026836/ENSMUSG00000024182
0000031548/ENSMUSG00000035828/ENSMUSG00000035621/ENSMUSG00000027712

0000074934

)2083

;6501/ENSMUSG00000005087/ENSMUSG00000024098/ENSMUSG00000024927/ENSMUSG00000023951/ENSMUSG00000039384/ENSMUSG00000042594/ENSMUSG00000028042/ENSMUSG00000050965/E

?7832/ENSMUSG00000027465/ENSMUSG00000074656/ENSMUSG00000040268/ENSMUSG00000031548/ENSMUSG00000025326

?5340/ENSMUSG00000039662/ENSMUSG00000032413/ENSMUSG00000109511/ENSMUSG00000032702/ENSMUSG00000044147/ENSMUSG00000039844/ENSMUSG00000009630/ENSMUSG00000031453

?6796/ENSMUSG00000005533/ENSMUSG00000030530/ENSMUSG00000030059/ENSMUSG00000028161/ENSMUSG00000063142/ENSMUSG00000018634/ENSMUSG00000040268/ENSMUSG00000018965/E

31548/ENSMUSG00000024425/ENSMUSG00000005871

ENSMUSG00000021149/ENSMUSG00000079037/ENSMUSG00000056050/ENSMUSG00000029816/ENSMUSG00000035356/ENSMUSG00000033454/ENSMUSG00000068740/ENSMUSG00000024425/ENSMUS

ENSMUSG00000033717/ENSMUSG00000029249/ENSMUSG00000029361/ENSMUSG00000031548/ENSMUSG00000035828/ENSMUSG00000024431/ENSMUSG00000020684/ENSMUSG00000031393/ENSMUS

;G00000085795/ENSMUSG00000037902/ENSMUSG00000045394

;G00000021794/ENSMUSG00000035621/ENSMUSG00000027712/ENSMUSG00000022808/ENSMUSG00000034593

| Category | GOID | Description | GeneRatio | BgRatio | pvalue | padj | geneID | geneName |
|---|---|---|---|---|---|---|---|---|
| CC | GO:003258 | Golgi cisterna membrane | 5\|1232 | 15\|16313 | 0.00385 | 0.04704 | ENSMUS | Csgalnact2/Chpf/Csgalnact1/Chsy3/Rab21 |
| BP | GO:000194 | lymph vessel development | 7\|1209 | 28\|16097 | 0.00386 | 0.02876 | ENSMUS | Flt4/Vegfa/Nfatc1/Vash1/Epha2/Ptpn14/Kdr |
| BP | GO:004594 | positive regulation of steroid metabolic process | 7\|1209 | 28\|16097 | 0.00386 | 0.02876 | ENSMUS | Stard4/Scd1/Bmp6/Nr1d1/Igf1r/Igf1/Dab2 |
| BP | GO:006103 | negative regulation of cartilage development | 7\|1209 | 28\|16097 | 0.00386 | 0.02876 | ENSMUS | Sox9/Ctsk/Rflna/Nog/Tgfbr1/Chadl/Grem1 |
| BP | GO:006105 | regulation of cell growth involved in cardiac muscle cell develo | 7\|1209 | 28\|16097 | 0.00386 | 0.02876 | ENSMUS | Col14a1/Rgs2/Ctdp1/Igf1/Tomm70a/Pi16/Nr3c1 |
| BP | GO:003053 | adult behavior | 22\|1209 | 159\|1609 | 0.0039 | 0.02892 | ENSMUS | Met/Mafg/Dnm1/Abl2/Npc1/Cxcl12/Ehmt2/Pafah1b1/Hipk2/K |
| BP | GO:004580 | positive regulation of endocytosis | 22\|1209 | 159\|1609 | 0.0039 | 0.02892 | ENSMUS | Serpine1/Cd14/Dnm1/Gas6/Cd36/Vegfa/Axl/Syne1/Ptx3/Sfrp4/ |
| BP | GO:004560 | positive regulation of fat cell differentiation | 11\|1209 | 59\|16097 | 0.00404 | 0.02997 | ENSMUS | Wif1/Lpl/Noct/Fndc3b/Xbp1/Sfrp2/Cebpb/Igf1/Zbtb7b/Ptgs2/S |
| BP | GO:004856 | embryonic organ development | 49\|1209 | 443\|1609 | 0.00407 | 0.03006 | ENSMUS | Spry2/Myc/Sox9/Hipk1/Col5a2/Fgfr1/Xist/Cdkn1c/Efna1/Mmp1 |
| BP | GO:004427 | sulfur compound biosynthetic process | 13\|1209 | 76\|16097 | 0.00407 | 0.03006 | ENSMUS | Acly/Xylt1/Eif2ak3/Chpf/Chst11/Csgalnact1/Slc35d1/Ugdh/Mth |
| BP | GO:006041 | cardiac septum morphogenesis | 13\|1209 | 76\|16097 | 0.00407 | 0.03006 | ENSMUS | Cyr61/Fgfr2/Tgfbr3/Nrp1/Nog/Tgfbr1/Robo1/Nfatc1/Notch1/R |
| CC | GO:004329 | apical junction complex | 19\|1232 | 130\|1631 | 0.00408 | 0.04874 | ENSMUS | Cdh5/Cldn12/Synpo/Amotl2/Usp53/Cgnl1/Tgfbr1/Kifc3/Ctnna |
| BP | GO:001056 | regulation of cellular ketone metabolic process | 17\|1209 | 112\|1609 | 0.00409 | 0.0302 | ENSMUS | Egr1/Stard4/Nfe2l1/Eif2ak3/Insig1/Bmp6/Trib3/Nr1d1/Acadl/D |
| BP | GO:003263 | interleukin-6 production | 21\|1209 | 150\|1609 | 0.00412 | 0.03033 | ENSMUS | Prg4/Tlr2/Lpl/Met/Il6ra/Stat3/Xbp1/C1qtnf3/Gas6/Cd36/Tnfrsf |
| BP | GO:006100 | regulation of dendritic spine morphogenesis | 10\|1209 | 51\|16097 | 0.00413 | 0.03035 | ENSMUS | Efna1/Ngef/Tiam1/Lrp8/Pafah1b1/Epha4/Dbn1/Shank3/Ube3a/ |
| BP | GO:000963 | response to toxic substance | 36\|1209 | 303\|1609 | 0.00416 | 0.03054 | ENSMUS | Mt1/Hmox1/Egr1/Met/Ppp1r15b/Aacs/Hsph1/Cd36/Prdx6/Slc |
| BP | GO:003265 | regulation of interleukin-1 production | 14\|1209 | 85\|16097 | 0.00418 | 0.03066 | ENSMUS | Errfi1/Tlr2/Egr1/Lpl/Cx3cl1/Jak2/Gas6/Hk1/Igf1/Lgals9/Pycard/ |
| BP | GO:001585 | organic hydroxy compound transport | 26\|1209 | 200\|1609 | 0.00428 | 0.03138 | ENSMUS | Ldlr/Myc/Igfbp3/Stard4/Lgals3/Rtn4/Srebf2/Bmp6/Slc5a3/Xbp |
| BP | GO:007122 | cellular response to acid chemical | 24\|1209 | 180\|1609 | 0.00429 | 0.03139 | ENSMUS | Col3a1/Col1a1/Col1a2/Col4a1/Ldlr/Sox9/Col5a2/Egr1/S100a1 |
| BP | GO:001714 | negative regulation of translation | 18\|1209 | 122\|1609 | 0.00435 | 0.03172 | ENSMUS | Igfbp5/Rgs2/Eif4ebp1/Larp1/Eif2ak3/Sesn2/Stat3/Btg2/Eif4g1/ |
| BP | GO:004864 | negative regulation of developmental growth | 18\|1209 | 122\|1609 | 0.00435 | 0.03172 | ENSMUS | Rgs2/Ifrd1/Rtn4/Cdkn1a/Nrp1/Sfrp2/Sema3b/Nog/Ctdp1/Tomn |
| BP | GO:004328 | positive regulation of cysteine-type endopeptidase activity inv | 17\|1209 | 113\|1609 | 0.00449 | 0.03263 | ENSMUS | F3/Myc/Eif2ak3/Cyr61/Ctgf/Ppp2ca/Jak2/Dlc1/Arl6ip5/Robo1/ |
| BP | GO:007145 | cellular response to oxygen levels | 17\|1209 | 113\|1609 | 0.00449 | 0.03263 | ENSMUS | Myc/Vasn/Egr1/Eif4ebp1/Rtn4/Hyou1/Cd34/Fndc1/Vegfa/Notc |
| BP | GO:000205 | positive regulation of mesenchymal cell proliferation | 8\|1209 | 36\|16097 | 0.0045 | 0.03263 | ENSMUS | Myc/Sox9/Fgfr1/Fgfr2/Foxp2/Vegfa/Syne1/Kdr |
| BP | GO:004688 | regulation of hormone biosynthetic process | 8\|1209 | 36\|16097 | 0.0045 | 0.03263 | ENSMUS | Egr1/Atp1a1/Bmp6/Igf1r/Igf1/Dab2/Rest/Nr3c1 |
| BP | GO:004666 | male sex differentiation | 19\|1209 | 132\|1609 | 0.00455 | 0.03293 | ENSMUS | Sox9/Inhba/Smad5/Adgrg1/Hmga2/Bmp6/Dhcr24/Sfrp2/Ren1/ |
| BP | GO:001087 | lipid localization | 38\|1209 | 326\|1609 | 0.00455 | 0.03293 | ENSMUS | Thbs1/Ldlr/Atp8b2/Abca8a/Inhba/Mest/Igfbp3/Stard4/Lpl/Sreb |
| BP | GO:000840 | gonad development | 25\|1209 | 191\|1609 | 0.0046 | 0.03296 | ENSMUS | Sox9/Serpine1/Inhba/Adgrg1/Hmga2/Tiparp/Sfrp2/Ren1/Tgfbr1 |
| BP | GO:200037 | regulation of reactive oxygen species metabolic process | 25\|1209 | 191\|1609 | 0.0046 | 0.03296 | ENSMUS | Thbs1/Tlr2/Klf4/Gadd45a/Cdkn1a/Ier3/Cd34/Rnf41/Egfr/Stat3/ |
| BP | GO:000821 | mineralocorticoid metabolic process | 4\|1209 | 10\|16097 | 0.0046 | 0.03296 | ENSMUS | Bmp6/Kcnma1/Dab2/Rest |
| BP | GO:001061 | regulation of cardiac muscle adaptation | 4\|1209 | 10\|16097 | 0.0046 | 0.03296 | ENSMUS | Errfi1/Lmna/Ppp3ca/Foxo1 |
| BP | GO:002180 | cerebral cortex tangential migration | 4\|1209 | 10\|16097 | 0.0046 | 0.03296 | ENSMUS | Fgfr1/Nrp1/Fbxo45/Robo1 |
| BP | GO:003234 | aldosterone metabolic process | 4\|1209 | 10\|16097 | 0.0046 | 0.03296 | ENSMUS | Bmp6/Kcnma1/Dab2/Rest |
| BP | GO:004358 | skin morphogenesis | 4\|1209 | 10\|16097 | 0.0046 | 0.03296 | ENSMUS | Errfi1/Col1a1/Col1a2/Itga2 |
| BP | GO:190545 | regulation of lymphoid progenitor cell differentiation | 4\|1209 | 10\|16097 | 0.0046 | 0.03296 | ENSMUS | Notch1/Zbtb1/Fnip1/Hes1 |
| BP | GO:001089 | positive regulation of steroid biosynthetic process | 6\|1209 | 22\|16097 | 0.00464 | 0.03308 | ENSMUS | Stard4/Bmp6/Nr1d1/Igf1r/Igf1/Dab2 |
| BP | GO:001491 | negative regulation of smooth muscle cell migration | 6\|1209 | 22\|16097 | 0.00464 | 0.03308 | ENSMUS | Igfbp5/Serpine1/Trib1/Igfbp3/Ndrg4/Nfe2l2 |
| BP | GO:003090 | notochord development | 6\|1209 | 22\|16097 | 0.00464 | 0.03308 | ENSMUS | Sox9/Col5a2/Efna1/Nog/Epha2/Id3 |
| BP | GO:004356 | regulation of insulin-like growth factor receptor signaling path | 6\|1209 | 22\|16097 | 0.00464 | 0.03308 | ENSMUS | Igfbp5/Cilp/Igfbp3/Igfbp6/Igfbp4/Igf1 |
| BP | GO:004211 | macrophage activation | 14\|1209 | 86\|16097 | 0.00466 | 0.03312 | ENSMUS | Thbs1/Ldlr/Tlr2/Jun/Cx3cl1/Jak2/Adgrf5/Nr1d1/Sbno2/Crtc3/C |
| BP | GO:004667 | response to antibiotic | 29\|1209 | 232\|1609 | 0.00469 | 0.03329 | ENSMUS | Hmox1/Egr1/Met/Ppp1r15b/Aacs/Larp1/Hsph1/Jak2/Igf1r/Igf1/ |
| BP | GO:005109 | positive regulation of DNA-binding transcription factor activity | 29\|1209 | 232\|1609 | 0.00469 | 0.03329 | ENSMUS | Fosl1/Tlr2/Ifrd1/Ncoa3/Lrrfip1/Ddit3/Hmga2/Rnf41/Cx3cl1/Tg |
| BP | GO:004303 | tRNA aminoacylation | 9\|1209 | 44\|16097 | 0.00476 | 0.03369 | ENSMUS | Aars/Gars/Sars/Nars/Tars/Yars/Iars/Dars/Cars |
| BP | GO:190289 | regulation of pri-miRNA transcription by RNA polymerase II | 9\|1209 | 44\|16097 | 0.00476 | 0.03369 | ENSMUS | Sox9/Jun/Srebf2/Fos/Stat3/Tgfbr1/Rela/Hdac4/Nr3c1 |
| BP | GO:001060 | posttranscriptional regulation of gene expression | 47\|1209 | 425\|1609 | 0.00485 | 0.03425 | ENSMUS | Thbs1/Igfbp5/Rgs2/Ppp1r15b/Aars/Eif4ebp1/Larp1/Noct/Eif2ak |
| BP | GO:190305 | positive regulation of proteolysis involved in cellular protein c | 17\|1209 | 114\|1609 | 0.00492 | 0.03471 | ENSMUS | Hspa1b/Hspa1a/Trib1/Herpud1/Zfand2a/Trib3/Xbp1/Trib2/Nkd |
| BP | GO:000680 | nitric oxide biosynthetic process | 12\|1209 | 69\|16097 | 0.00494 | 0.0348 | ENSMUS | Tlr2/Klf4/Cd34/Jak2/Cd36/Igf1/Ddah2/Ptx3/Clu/Ptgs2/Nos1/Si |
| BP | GO:190336 | positive regulation of cellular protein catabolic process | 19\|1209 | 133\|1609 | 0.00494 | 0.0348 | ENSMUS | Hspa1b/Hspa1a/Ldlr/Trib1/Herpud1/Zfand2a/Trib3/Xbp1/Trib2 |
| BP | GO:000693 | muscle contraction | 33\|1209 | 275\|1609 | 0.00507 | 0.03566 | ENSMUS | Sulf2/Rgs2/Map2k3/Myl1/Met/Cald1/Atp1a1/Tpm1/Smad5/Tpr |
| BP | GO:000759 | blood coagulation | 20\|1209 | 143\|1609 | 0.00509 | 0.03575 | ENSMUS | F3/Thbs1/Serpine1/Serping1/F13a1/Anxa8/Cd34/Cx3cl1/Gas6/ |
| BP | GO:001473 | regulation of skeletal muscle adaptation | 5\|1209 | 16\|16097 | 0.00514 | 0.03585 | ENSMUS | Igfbp5/Atp2a2/Nfatc1/Gtf2i/Ppp3ca |

| Count | Up | Up_Gene_i | Down | Down_Gene_id |
|---|---|---|---|---|
| 5 | 0 | | 5 | ENSMUSG00000042042/ENSMUSG00000032997/ENSMUSG00000036356/ENSMUSG00000058152/ENSMUSG00000020132 |
| 7 | 4 | ENSMUSG0 | 3 | ENSMUSG00000023951/ENSMUSG00000006445/ENSMUSG00000026604 |
| 7 | 2 | ENSMUSG0 | 5 | ENSMUSG00000024378/ENSMUSG00000037071/ENSMUSG00000039004/ENSMUSG00000020889/ENSMUSG00000005533 |
| 7 | 3 | ENSMUSG0 | 4 | ENSMUSG00000000567/ENSMUSG00000048616/ENSMUSG00000007613/ENSMUSG00000074934 |
| 7 | 3 | ENSMUSG0 | 4 | ENSMUSG00000026360/ENSMUSG00000033323/ENSMUSG00000022752/ENSMUSG00000024431 |
| 22 | 7 | ENSMUSG0 | 15 | ENSMUSG00000009376/ENSMUSG00000051510/ENSMUSG00000026596/ENSMUSG00000024413/ENSMUSG00000020745/ENSMUSG00000061436/ENSMUSG0 |
| 22 | 7 | ENSMUSG0 | 15 | ENSMUSG00000037411/ENSMUSG00000051439/ENSMUSG00000031451/ENSMUSG00000023951/ENSMUSG00000096054/ENSMUSG00000027832/ENSMUSG0 |
| 11 | 2 | ENSMUSG0 | 9 | ENSMUSG00000020218/ENSMUSG00000023087/ENSMUSG00000039286/ENSMUSG00000020484/ENSMUSG00000027996/ENSMUSG00000056501/ENSMUSG0 |
| 49 | 22 | ENSMUSG0 | 27 | ENSMUSG00000022114/ENSMUSG00000022346/ENSMUSG00000000567/ENSMUSG00000008730/ENSMUSG00000031565/ENSMUSG00000027954/ENSMUSG0 |
| 13 | 1 | ENSMUSG0 | 12 | ENSMUSG00000020917/ENSMUSG00000030657/ENSMUSG00000031668/ENSMUSG00000032997/ENSMUSG00000036356/ENSMUSG00000028521/ENSMUSG0 |
| 13 | 8 | ENSMUSG0 | 5 | ENSMUSG00000030849/ENSMUSG00000029287/ENSMUSG00000048616/ENSMUSG00000007613/ENSMUSG00000026836 |
| 19 | 7 | ENSMUSG0 | 12 | ENSMUSG00000046798/ENSMUSG00000039701/ENSMUSG00000032232/ENSMUSG00000007613/ENSMUSG00000031788/ENSMUSG00000037815/ENSMUSG0 |
| 17 | 2 | ENSMUSG0 | 15 | ENSMUSG00000038418/ENSMUSG00000024378/ENSMUSG00000038615/ENSMUSG00000031668/ENSMUSG00000045294/ENSMUSG00000039004/ENSMUSG0 |
| 21 | 6 | ENSMUSG0 | 15 | ENSMUSG00000006014/ENSMUSG00000027995/ENSMUSG00000009376/ENSMUSG00000027947/ENSMUSG00000004040/ENSMUSG00000020484/ENSMUSG0 |
| 10 | 3 | ENSMUSG0 | 7 | ENSMUSG00000027954/ENSMUSG00000026259/ENSMUSG00000002489/ENSMUSG00000028613/ENSMUSG00000020745/ENSMUSG00000025326/ENSMUSG0 |
| 36 | 12 | ENSMUSG0 | 24 | ENSMUSG00000031765/ENSMUSG00000005413/ENSMUSG00000038418/ENSMUSG00000009376/ENSMUSG00000046062/ENSMUSG00000029482/ENSMUSG0 |
| 14 | 6 | ENSMUSG0 | 8 | ENSMUSG00000028967/ENSMUSG00000027995/ENSMUSG00000038418/ENSMUSG00000031778/ENSMUSG00000024789/ENSMUSG00000031451/ENSMUSG0 |
| 26 | 7 | ENSMUSG0 | 19 | ENSMUSG00000032193/ENSMUSG00000022346/ENSMUSG00000024378/ENSMUSG00000050335/ENSMUSG00000020458/ENSMUSG00000022463/ENSMUSG0 |
| 24 | 11 | ENSMUSG0 | 13 | ENSMUSG00000032193/ENSMUSG00000000567/ENSMUSG00000038418/ENSMUSG00000041959/ENSMUSG00000026596/ENSMUSG00000028893/ENSMUSG0 |
| 18 | 2 | ENSMUSG0 | 16 | ENSMUSG00000026360/ENSMUSG00000031490/ENSMUSG00000037331/ENSMUSG00000031668/ENSMUSG00000028893/ENSMUSG00000004040/ENSMUSG0 |
| 18 | 6 | ENSMUSG0 | 12 | ENSMUSG00000026360/ENSMUSG00000001627/ENSMUSG00000020458/ENSMUSG00000023067/ENSMUSG00000027996/ENSMUSG00000048616/ENSMUSG0 |
| 17 | 5 | ENSMUSG0 | 12 | ENSMUSG00000028128/ENSMUSG00000022346/ENSMUSG00000031668/ENSMUSG00000019997/ENSMUSG00000020349/ENSMUSG00000024789/ENSMUSG0 |
| 17 | 4 | ENSMUSG0 | 13 | ENSMUSG00000022346/ENSMUSG00000039646/ENSMUSG00000038418/ENSMUSG00000031490/ENSMUSG00000020458/ENSMUSG00000032115/ENSMUSG0 |
| 8 | 2 | ENSMUSG0 | 6 | ENSMUSG00000022346/ENSMUSG00000000567/ENSMUSG00000031565/ENSMUSG00000030849/ENSMUSG00000023951/ENSMUSG00000096054 |
| 8 | 2 | ENSMUSG0 | 6 | ENSMUSG00000038418/ENSMUSG00000033161/ENSMUSG00000039004/ENSMUSG00000005533/ENSMUSG00000029249/ENSMUSG00000024431 |
| 19 | 4 | ENSMUSG0 | 15 | ENSMUSG00000000567/ENSMUSG00000041324/ENSMUSG00000021540/ENSMUSG00000031785/ENSMUSG00000056758/ENSMUSG00000039004/ENSMUSG0 |
| 38 | 13 | ENSMUSG0 | 25 | ENSMUSG00000040152/ENSMUSG00000032193/ENSMUSG00000060671/ENSMUSG00000041324/ENSMUSG00000024378/ENSMUSG00000022463/ENSMUSG0 |
| 25 | 5 | ENSMUSG0 | 20 | ENSMUSG00000000567/ENSMUSG00000037411/ENSMUSG00000041324/ENSMUSG00000031785/ENSMUSG00000056758/ENSMUSG00000034640/ENSMUSG0 |
| 25 | 9 | ENSMUSG0 | 16 | ENSMUSG00000040152/ENSMUSG00000027995/ENSMUSG00000003032/ENSMUSG00000036390/ENSMUSG00000023067/ENSMUSG00000003541/ENSMUSG0 |
| 4 | 1 | ENSMUSG0 | 3 | ENSMUSG00000039004/ENSMUSG00000063142/ENSMUSG00000029249 |
| 4 | 0 | | 4 | ENSMUSG00000028967/ENSMUSG00000028063/ENSMUSG00000028161/ENSMUSG00000044167 |
| 4 | 2 | ENSMUSG0 | 2 | ENSMUSG00000031565/ENSMUSG00000035764 |
| 4 | 1 | ENSMUSG0 | 3 | ENSMUSG00000039004/ENSMUSG00000063142/ENSMUSG00000029249 |
| 4 | 3 | ENSMUSG0 | 1 | ENSMUSG00000028967 |
| 4 | 2 | ENSMUSG0 | 2 | ENSMUSG00000033454/ENSMUSG00000035992 |
| 6 | 2 | ENSMUSG0 | 4 | ENSMUSG00000024378/ENSMUSG00000039004/ENSMUSG00000020889/ENSMUSG00000005533 |
| 6 | 2 | ENSMUSG0 | 4 | ENSMUSG00000037411/ENSMUSG00000032501/ENSMUSG00000036564/ENSMUSG00000015839 |
| 6 | 2 | ENSMUSG0 | 4 | ENSMUSG00000000567/ENSMUSG00000027954/ENSMUSG00000048616/ENSMUSG00000006445 |
| 6 | 6 | ENSMUSG0 | 0 | |
| 14 | 4 | ENSMUSG0 | 10 | ENSMUSG00000040152/ENSMUSG00000032193/ENSMUSG00000027995/ENSMUSG00000031778/ENSMUSG00000024789/ENSMUSG00000020889/ENSMUSG0 |
| 29 | 7 | ENSMUSG0 | 22 | ENSMUSG00000005413/ENSMUSG00000038418/ENSMUSG00000009376/ENSMUSG00000046062/ENSMUSG00000029482/ENSMUSG00000037331/ENSMUSG0 |
| 29 | 6 | ENSMUSG0 | 23 | ENSMUSG00000024912/ENSMUSG00000027995/ENSMUSG00000001627/ENSMUSG00000027678/ENSMUSG00000026305/ENSMUSG00000025408/ENSMUSG0 |
| 9 | 1 | ENSMUSG0 | 8 | ENSMUSG00000031960/ENSMUSG00000029777/ENSMUSG00000068739/ENSMUSG00000024587/ENSMUSG00000022241/ENSMUSG00000028811/ENSMUSG0 |
| 9 | 2 | ENSMUSG0 | 7 | ENSMUSG00000000567/ENSMUSG00000022463/ENSMUSG00000004040/ENSMUSG00000007613/ENSMUSG00000024927/ENSMUSG00000026313/ENSMUSG0 |
| 47 | 11 | ENSMUSG0 | 36 | ENSMUSG00000040152/ENSMUSG00000026360/ENSMUSG00000046062/ENSMUSG00000031960/ENSMUSG00000031490/ENSMUSG00000037331/ENSMUSG0 |
| 17 | 5 | ENSMUSG0 | 12 | ENSMUSG00000032501/ENSMUSG00000031770/ENSMUSG00000053581/ENSMUSG00000032715/ENSMUSG00000020484/ENSMUSG00000020601/ENSMUSG0 |
| 12 | 4 | ENSMUSG0 | 8 | ENSMUSG00000027995/ENSMUSG00000003032/ENSMUSG00000024789/ENSMUSG00000027832/ENSMUSG00000022037/ENSMUSG00000032487/ENSMUSG0 |
| 19 | 5 | ENSMUSG0 | 14 | ENSMUSG00000032193/ENSMUSG00000032501/ENSMUSG00000031770/ENSMUSG00000053581/ENSMUSG00000032715/ENSMUSG00000020484/ENSMUSG0 |
| 33 | 15 | ENSMUSG0 | 18 | ENSMUSG00000006800/ENSMUSG00000026360/ENSMUSG00000018932/ENSMUSG00000009376/ENSMUSG00000033161/ENSMUSG00000021540/ENSMUSG0 |
| 20 | 7 | ENSMUSG0 | 13 | ENSMUSG00000028128/ENSMUSG00000040152/ENSMUSG00000037411/ENSMUSG00000021950/ENSMUSG00000031778/ENSMUSG00000031451/ENSMUSG0 |
| 5 | 3 | ENSMUSG0 | 2 | ENSMUSG00000029467/ENSMUSG00000028161 |

0000063142/ENSMUSG00000018634/ENSMUSG00000029361/ENSMUSG00000000184/ENSMUSG00000031393/ENSMUSG00000026826/ENSMUSG00000026812/ENSMUSG00000024182/ENSMUSG0000003
0000054808/ENSMUSG00000028161/ENSMUSG00000003814/ENSMUSG00000022037/ENSMUSG00000016024/ENSMUSG00000020132/ENSMUSG00000039844/ENSMUSG00000074934/ENSMUSG0000003
0000028042/ENSMUSG00000032487/ENSMUSG00000031548
0000027678/ENSMUSG00000030849/ENSMUSG00000045294/ENSMUSG00000031714/ENSMUSG00000048616/ENSMUSG00000007613/ENSMUSG00000056501/ENSMUSG00000035247/ENSMUSG0000004
0000029201/ENSMUSG00000029009/ENSMUSG00000028179/ENSMUSG00000039497/ENSMUSG00000032350/ENSMUSG00000015839

0000024812/ENSMUSG00000013076/ENSMUSG00000039473/ENSMUSG00000062232/ENSMUSG00000045394/ENSMUSG00000005871
0000032715/ENSMUSG00000020889/ENSMUSG00000030747/ENSMUSG00000032349/ENSMUSG00000063142/ENSMUSG00000032487/ENSMUSG00000029249/ENSMUSG00000026623/ENSMUSG0000000
0000031451/ENSMUSG00000030341/ENSMUSG00000056501/ENSMUSG00000037447/ENSMUSG00000022708/ENSMUSG00000025340/ENSMUSG00000016024/ENSMUSG00000027551/ENSMUSG0000003
0000025372
0000037434/ENSMUSG00000005533/ENSMUSG00000032724/ENSMUSG00000024927/ENSMUSG00000018476/ENSMUSG00000024423/ENSMUSG00000050965/ENSMUSG00000063142/ENSMUSG0000001
0000037012/ENSMUSG00000037902
0000039004/ENSMUSG00000089774/ENSMUSG00000020484/ENSMUSG00000024413/ENSMUSG00000070645/ENSMUSG00000030059/ENSMUSG00000023068/ENSMUSG00000018634/ENSMUSG0000000
0000020484/ENSMUSG00000056501/ENSMUSG00000023951/ENSMUSG00000030747/ENSMUSG00000001119/ENSMUSG00000028646/ENSMUSG00000050697
0000020423/ENSMUSG00000045983/ENSMUSG00000050965/ENSMUSG00000026254/ENSMUSG00000003814/ENSMUSG00000045896/ENSMUSG00000040025/ENSMUSG00000020091/ENSMUSG0000002
0000033323/ENSMUSG00000022752/ENSMUSG00000039384/ENSMUSG00000031548/ENSMUSG00000031393/ENSMUSG00000005871
0000031523/ENSMUSG00000035199/ENSMUSG00000029249/ENSMUSG00000035900/ENSMUSG00000056071/ENSMUSG00000002083
0000023951/ENSMUSG00000024924/ENSMUSG00000036450/ENSMUSG00000044167/ENSMUSG00000015839/ENSMUSG00000025612/ENSMUSG00000026812

0000034926/ENSMUSG00000027996/ENSMUSG00000070645/ENSMUSG00000007613/ENSMUSG00000030059/ENSMUSG00000027465/ENSMUSG00000074656/ENSMUSG00000040268/ENSMUSG0000003
0000032883/ENSMUSG00000022351/ENSMUSG00000039004/ENSMUSG00000029426/ENSMUSG00000024413/ENSMUSG00000070645/ENSMUSG00000048490/ENSMUSG00000030059/ENSMUSG0000002
0000027996/ENSMUSG00000070645/ENSMUSG00000007613/ENSMUSG00000056501/ENSMUSG00000023951/ENSMUSG00000048490/ENSMUSG00000030059/ENSMUSG00000079509/ENSMUSG0000002
0000025373/ENSMUSG00000004040/ENSMUSG00000024789/ENSMUSG00000027832/ENSMUSG00000026313/ENSMUSG00000022037/ENSMUSG00000032487/ENSMUSG00000044167/ENSMUSG0000001

0000035673/ENSMUSG00000030527/ENSMUSG00000022037/ENSMUSG00000016024
0000024789/ENSMUSG00000005533/ENSMUSG00000024927/ENSMUSG00000018476/ENSMUSG00000024423/ENSMUSG00000050965/ENSMUSG00000018634/ENSMUSG00000040268/ENSMUSG0000002
0000056758/ENSMUSG00000025373/ENSMUSG00000031778/ENSMUSG00000029287/ENSMUSG00000004040/ENSMUSG00000028613/ENSMUSG00000024789/ENSMUSG00000024927/ENSMUSG0000002
0000037851/ENSMUSG00000010755
0000024431
0000023087/ENSMUSG00000031668/ENSMUSG00000015846/ENSMUSG00000028893/ENSMUSG00000004040/ENSMUSG00000020423/ENSMUSG00000035093/ENSMUSG00000020594/ENSMUSG0000002
0000022037/ENSMUSG00000025234/ENSMUSG00000028793/ENSMUSG00000032350/ENSMUSG00000038068/ENSMUSG00000024182
0000029361/ENSMUSG00000037902
0000020601/ENSMUSG00000022037/ENSMUSG00000025234/ENSMUSG00000028793/ENSMUSG00000032350/ENSMUSG00000038068/ENSMUSG00000009291/ENSMUSG00000024182
0000029467/ENSMUSG00000028525/ENSMUSG00000019997/ENSMUSG00000031849/ENSMUSG00000021070/ENSMUSG00000050965/ENSMUSG00000063142/ENSMUSG00000026313/ENSMUSG0000003
0000031849/ENSMUSG00000042594/ENSMUSG00000050965/ENSMUSG00000015839/ENSMUSG00000026797/ENSMUSG00000056071/ENSMUSG00000027712

32096
37902

45216/ENSMUSG00000023951/ENSMUSG00000074221/ENSMUSG00000024073/ENSMUSG00000042594/ENSMUSG00000061436/ENSMUSG00000006445/ENSMUSG00000032717/ENSMUSG00000074818/E

05871
37902

18634/ENSMUSG00000040268/ENSMUSG00000015839/ENSMUSG00000028211/ENSMUSG00000030287/ENSMUSG00000009630/ENSMUSG00000026826/ENSMUSG00000050697/ENSMUSG00000037902/E

09907/ENSMUSG00000024687/ENSMUSG00000029361/ENSMUSG00000031393/ENSMUSG00000021728

21838/ENSMUSG00000026812

31548
23068/ENSMUSG00000021070/ENSMUSG00000030747/ENSMUSG00000024924/ENSMUSG00000018634/ENSMUSG00000009907/ENSMUSG00000016024/ENSMUSG00000024687/ENSMUSG00000037606/E
27832/ENSMUSG00000027465/ENSMUSG00000074656/ENSMUSG00000040268/ENSMUSG00000031548/ENSMUSG00000025326
15839/ENSMUSG00000037902

22037/ENSMUSG00000015839/ENSMUSG00000028211/ENSMUSG00000030287/ENSMUSG00000009630/ENSMUSG00000050697/ENSMUSG00000037902/ENSMUSG00000004207
23951/ENSMUSG00000061436/ENSMUSG00000028161/ENSMUSG00000055491/ENSMUSG00000030527/ENSMUSG00000026313/ENSMUSG00000022037/ENSMUSG00000028179/ENSMUSG00000074934

26728/ENSMUSG00000045983/ENSMUSG00000003135/ENSMUSG00000033499/ENSMUSG00000023951/ENSMUSG00000030738/ENSMUSG00000034575/ENSMUSG00000028161/ENSMUSG00000024423/E

32487/ENSMUSG00000033717/ENSMUSG00000029361/ENSMUSG00000019943

ENSMUSG00000026836/ENSMUSG00000029249/ENSMUSG00000031540/ENSMUSG00000024182

ENSMUSG00000004207

ENSMUSG00000043421/ENSMUSG00000004207

ENSMUSG00000050965/ENSMUSG00000042109/ENSMUSG00000026254/ENSMUSG00000003814/ENSMUSG00000045896/ENSMUSG00000040025/ENSMUSG00000020091/ENSMUSG00000022412/ENSMUS

;G00000021838/ENSMUSG00000031393/ENSMUSG00000056071/ENSMUSG00000026812/ENSMUSG00000000384

| Category | GOID | Description | GeneRatio | BgRatio | pvalue | padj | geneID | geneName |
|---|---|---|---|---|---|---|---|---|
| BP | GO:00359 | chondrocyte proliferation | 5\|1209 | 16\|16097 | 0.00514 | 0.03585 | ENSMUS | Mmp14/Ctgf/Comp/Nov/Col9a1 |
| BP | GO:19900 | amyloid fibril formation | 5\|1209 | 16\|16097 | 0.00514 | 0.03585 | ENSMUS | Ldlr/Cd36/Clu/Cryab/App |
| BP | GO:20008 | regulation of corticosteroid hormone secretion | 5\|1209 | 16\|16097 | 0.00514 | 0.03585 | ENSMUS | Bmp6/Agtr2/Ren1/Crhr1/Dab2 |
| BP | GO:20010 | positive regulation of endothelial cell chemotaxis | 5\|1209 | 16\|16097 | 0.00514 | 0.03585 | ENSMUS | Fgfr1/Met/Smoc2/Vegfa/Kdr |
| BP | GO:00550 | regulation of cardiac muscle tissue development | 15\|1209 | 96\|16097 | 0.00515 | 0.03587 | ENSMUS | Col14a1/Rgs2/Fgfr1/Fgfr2/Tgfbr3/Nog/Tgfbr1/Ctdp1/Igf1/Tomm |
| BP | GO:00459 | regulation of embryonic development | 18\|1209 | 124\|1609 | 0.00517 | 0.03599 | ENSMUS | Klf4/Col5a2/Fgfr1/Lama2/Noct/Phldb1/Nrk/B4galt5/Sfrp2/Igf1/ |
| BP | GO:00218 | forebrain neuron differentiation | 11\|1209 | 61\|16097 | 0.00526 | 0.03653 | ENSMUS | Inhba/Fgfr1/Fgfr2/Nrp1/Robo1/Smarcc2/Robo2/Rapgef2/Atf5/ |
| BP | GO:00454 | regulation of nitric oxide biosynthetic process | 11\|1209 | 61\|16097 | 0.00526 | 0.03653 | ENSMUS | Tlr2/Klf4/Cd34/Jak2/Cd36/Igf1/Ddah2/Ptx3/Clu/Ptgs2/Sirpa |
| BP | GO:00341 | erythrocyte homeostasis | 19\|1209 | 134\|1609 | 0.00537 | 0.03719 | ENSMUS | Hmox1/Inhba/Hspa9/Nfe2l1/Smad5/Tgfbr3/Stat3/Cebpg/Bcl6/J |
| BP | GO:00027 | regulation of peptide secretion | 51\|1209 | 472\|1609 | 0.00544 | 0.03768 | ENSMUS | Hmgcr/Tlr2/Postn/Cyp51/Cd14/Lpl/Aacs/Akap12/Abl2/Hmga2/ |
| BP | GO:00508 | regulation of body fluid levels | 34\|1209 | 287\|1609 | 0.00548 | 0.03775 | ENSMUS | F3/Thbs1/Serpine1/Met/Scd1/Serping1/Eif2ak3/F13a1/Anxa8/C |
| BP | GO:00468 | hormone secretion | 37\|1209 | 319\|1609 | 0.00549 | 0.03775 | ENSMUS | Hmgcr/Inhba/Fgfr1/Igfbp3/Aacs/Hmga2/Bmp6/Egfr/Tiam1/Agtr |
| BP | GO:00075 | neuromuscular junction development | 10\|1209 | 53\|16097 | 0.0055 | 0.03775 | ENSMUS | Col4a1/Sorbs2/Fgfr2/Tnc/Nedd4/Lamb2/Fzd9/Ptn/Shank3/App |
| BP | GO:00355 | non-canonical Wnt signaling pathway | 10\|1209 | 53\|16097 | 0.0055 | 0.03775 | ENSMUS | Abl2/Tiam1/Sfrp2/Fzd10/Sfrp4/Nkd1/Dab2/Fzd9/Sfrp1/Fzd2 |
| BP | GO:19047 | regulation of vascular smooth muscle cell proliferation | 10\|1209 | 53\|16097 | 0.0055 | 0.03775 | ENSMUS | Hmox1/Igfbp5/Rgs5/Jun/Cdkn1a/Mef2d/Jak2/Igf1/Gnai2/Adam |
| BP | GO:19908 | vascular smooth muscle cell proliferation | 10\|1209 | 53\|16097 | 0.0055 | 0.03775 | ENSMUS | Hmox1/Igfbp5/Rgs5/Jun/Cdkn1a/Mef2d/Jak2/Igf1/Gnai2/Adam |
| BP | GO:00430 | amino acid activation | 9\|1209 | 45\|16097 | 0.00556 | 0.0381 | ENSMUS | Aars/Gars/Sars/Nars/Tars/Yars/Iars/Dars/Cars |
| BP | GO:19909 | response to amino acid starvation | 9\|1209 | 45\|16097 | 0.00556 | 0.0381 | ENSMUS | Atf4/Eif4ebp1/Larp1/Cdkn1a/Eif2ak3/Sesn2/Impact/Rragc/Atf3 |
| BP | GO:19030 | regulation of proteolysis involved in cellular protein catabolic | 26\|1209 | 204\|1609 | 0.00557 | 0.0381 | ENSMUS | Hspa1b/Hspa1a/Trib1/Herpud1/Zfand2a/Efna1/Nfe2l1/Trib3/Xb |
| BP | GO:00109 | positive regulation of peptidase activity | 21\|1209 | 154\|1609 | 0.0056 | 0.03829 | ENSMUS | F3/Myc/Efna1/Eif2ak3/Cyr61/Stat3/Ctgf/Ppp2ca/Jak2/Sfrp2/Dl |
| BP | GO:19033 | regulation of cellular protein catabolic process | 29\|1209 | 235\|1609 | 0.00562 | 0.03837 | ENSMUS | Hspa1b/Hspa1a/Ldlr/Trib1/Herpud1/Zfand2a/Efna1/Nfe2l1/Trib |
| BP | GO:00064 | regulation of translation | 36\|1209 | 309\|1609 | 0.00569 | 0.0388 | ENSMUS | Thbs1/Igfbp5/Rgs2/Ppp1r15b/Aars/Eif4ebp1/Larp1/Eif2ak3/Rxr |
| BP | GO:00072 | integrin-mediated signaling pathway | 14\|1209 | 88\|16097 | 0.00574 | 0.03908 | ENSMUS | Col3a1/Itga5/Thy1/Timp1/Ctgf/Nrp1/Itgb5/Ctnna1/Itgbl1/Rcc2 |
| BP | GO:00512 | maintenance of location | 34\|1209 | 288\|1609 | 0.00578 | 0.03933 | ENSMUS | Mest/Stard4/Lpl/Srebf2/Ddit3/Sqle/Itpr3/Insig1/Ibtk/Cx3cl1/Th |
| BP | GO:00162 | negative regulation of macroautophagy | 7\|1209 | 30\|16097 | 0.00582 | 0.0394 | ENSMUS | Hmox1/Rnf41/Usp36/Smcr8/Npc1/Ehmt2/Tsc1 |
| BP | GO:00610 | myeloid leukocyte cytokine production | 7\|1209 | 30\|16097 | 0.00582 | 0.0394 | ENSMUS | Hmox1/Tlr2/Rtn4/Bcl6/Gas6/Cd36/Ddx21 |
| BP | GO:19027 | positive regulation of cell cycle G2/M phase transition | 7\|1209 | 30\|16097 | 0.00582 | 0.0394 | ENSMUS | Ccnd1/Pbx1/Rcc2/Vps4b/App/Mecp2/Fbxo5 |
| BP | GO:190531 | semi-lunar valve development | 7\|1209 | 30\|16097 | 0.00582 | 0.0394 | ENSMUS | Sox9/Efna1/Tnfrsf1a/Robo1/Nfatc1/Notch1/Robo2 |
| BP | GO:000681 | phosphate ion transport | 6\|1209 | 23\|16097 | 0.00589 | 0.03951 | ENSMUS | Slc20a1/Fgfr1/Atf4/Ank/Cebpb/Sfrp4 |
| BP | GO:00218 | cerebral cortex radial glia guided migration | 6\|1209 | 23\|16097 | 0.00589 | 0.03951 | ENSMUS | Lamb1/Rtn4/Adgrg1/Lrp8/Pafah1b1/Srgap2 |
| BP | GO:00220 | telencephalon glial cell migration | 6\|1209 | 23\|16097 | 0.00589 | 0.03951 | ENSMUS | Lamb1/Rtn4/Adgrg1/Lrp8/Pafah1b1/Srgap2 |
| BP | GO:00319 | positive regulation of protein autophosphorylation | 6\|1209 | 23\|16097 | 0.00589 | 0.03951 | ENSMUS | Iqgap1/Tnks1bp1/Vegfa/Gpnmb/Ace/Grem1 |
| BP | GO:00359 | steroid hormone secretion | 6\|1209 | 23\|16097 | 0.00589 | 0.03951 | ENSMUS | Inhba/Bmp6/Agtr2/Ren1/Crhr1/Dab2 |
| BP | GO:00435 | skeletal muscle adaptation | 6\|1209 | 23\|16097 | 0.00589 | 0.03951 | ENSMUS | Igfbp5/Atp2a2/Nfatc1/Gtf2i/Ppp3ca/Myoz2 |
| BP | GO:007201 | glomerulus vasculature development | 6\|1209 | 23\|16097 | 0.00589 | 0.03951 | ENSMUS | Egr1/Il6ra/Notch3/Notch1/Tek/Hes1 |
| BP | GO:001931 | hexose metabolic process | 26\|1209 | 205\|1609 | 0.00594 | 0.03976 | ENSMUS | Myc/Igfbp3/Atf4/Nfe2l1/Igfbp4/Sesn2/C1qtnf3/Sik1/Hk1/Nr1d1 |
| BP | GO:00075 | hemostasis | 20\|1209 | 145\|1609 | 0.00595 | 0.03981 | ENSMUS | F3/Thbs1/Serpine1/Serping1/F13a1/Anxa8/Cd34/Cx3cl1/Gas6/ |
| BP | GO:01500 | neuroinflammatory response | 11\|1209 | 62\|16097 | 0.00598 | 0.03993 | ENSMUS | Ldlr/Tlr2/Jun/Cx3cl1/Egfr/Jak2/Nr1d1/Igf1/Clu/Ptgs2/App |
| BP | GO:00507 | regulation of protein secretion | 48\|1209 | 441\|1609 | 0.006 | 0.04003 | ENSMUS | Hmgcr/Tlr2/Postn/Cyp51/Cd14/Lpl/Aacs/Akap12/Abl2/Cd34/C |
| BP | GO:20012 | regulation of intrinsic apoptotic signaling pathway | 22\|1209 | 165\|1609 | 0.00608 | 0.04048 | ENSMUS | Herpud1/Eif2ak3/Ddit3/Hyou1/Xbp1/Hsph1/Cxcl12/Bag5/Stea |
| BP | GO:00224 | negative regulation of cell-cell adhesion | 23\|1209 | 175\|1609 | 0.00608 | 0.04048 | ENSMUS | Klf4/Lgals3/Abl2/Fgl2/Bmp6/Bcl6/Jak2/Cxcl12/Cebpb/Cd44/Tw |
| BP | GO:00217 | glial cell development | 16\|1209 | 107\|1609 | 0.00609 | 0.04048 | ENSMUS | Ldlr/Tlr2/Egfr/B4galt5/Ski/Nr1d1/Vim/Ndrg1/Adgrg6/Lamb2/Id |
| BP | GO:00457 | positive regulation of cell cycle | 38\|1209 | 332\|1609 | 0.00614 | 0.04075 | ENSMUS | Fosl1/Myc/Fgfr1/Met/Eif4ebp1/Smoc2/Cdkn1a/Fgfr2/Egfr/Mybl |
| BP | GO:00327 | monocarboxylic acid metabolic process | 50\|1209 | 464\|1609 | 0.00619 | 0.04105 | ENSMUS | Tnxb/Myc/Acly/Stard4/Lpl/Scd1/Pex5/Aacs/Nfe2l1/Ier3/Acsl3/E |
| BP | GO:00511 | smooth muscle cell differentiation | 12\|1209 | 71\|16097 | 0.00625 | 0.04135 | ENSMUS | Sox9/Fgfr2/Comp/Vegfa/Nfatc1/Notch1/Acvr1/Cth/Tmsb4x/Hes |
| BP | GO:19000 | regulation of hemostasis | 12\|1209 | 71\|16097 | 0.00625 | 0.04135 | ENSMUS | Thbs1/Serpine1/Serping1/Cd34/Comp/Cd36/Sh2b3/Tfpi/Prkca/ |
| BP | GO:00322 | regulation of protein polymerization | 26\|1209 | 206\|1609 | 0.00632 | 0.04178 | ENSMUS | Tlr2/Eln/Met/Cdh5/Pecam1/Tpm1/Sgk1/Pik3r2/Lats1/Tmsb10/ |
| BP | GO:00435 | regulation of JUN kinase activity | 14\|1209 | 89\|16097 | 0.00635 | 0.04183 | ENSMUS | Tnxb/Nrk/Tiam1/Gab1/Sfrp2/Fzd10/Dusp10/Hipk3/Mdfi/Epha4 |
| BP | GO:00712 | cellular response to mechanical stimulus | 8\|1209 | 38\|16097 | 0.00636 | 0.04183 | ENSMUS | Col1a1/Sox9/Piezo1/Atp1a1/Dag1/Atp1a2/Cnn2/Nos1 |
| BP | GO:00970 | energy homeostasis | 8\|1209 | 38\|16097 | 0.00636 | 0.04183 | ENSMUS | Stat3/Cd36/Eif4g1/Crtc3/Pik3ca/Ampd2/Prcp/Prkaa1 |
| BP | GO:19000 | positive regulation of substrate adhesion-dependent cell sprea | 8\|1209 | 38\|16097 | 0.00636 | 0.04183 | ENSMUS | Has2/S100a10/Nrp1/Enpp2/Nedd9/Calr/Dab2/Dbn1 |

| Count | Up | Up_Gene_i | Down | Down_Gene_id |
|---|---|---|---|---|
| 5 | 2 | ENSMUSG0 | 3 | ENSMUSG00000019997/ENSMUSG00000031849/ENSMUSG00000026147 |
| 5 | 3 | ENSMUSG0 | 2 | ENSMUSG00000032193/ENSMUSG00000022037 |
| 5 | 2 | ENSMUSG0 | 3 | ENSMUSG00000039004/ENSMUSG00000070645/ENSMUSG00000018634 |
| 5 | 2 | ENSMUSG0 | 3 | ENSMUSG00000031565/ENSMUSG00000009376/ENSMUSG00000023951 |
| 15 | 4 | ENSMUSG0 | 11 | ENSMUSG00000026360/ENSMUSG00000031565/ENSMUSG00000030849/ENSMUSG00000029287/ENSMUSG00000048616/ENSMUSG00000007613/ENSMUSG0 |
| 18 | 8 | ENSMUSG0 | 10 | ENSMUSG00000003032/ENSMUSG00000031565/ENSMUSG00000023087/ENSMUSG00000048537/ENSMUSG00000017929/ENSMUSG00000027996/ENSMUSG0 |
| 11 | 6 | ENSMUSG0 | 5 | ENSMUSG00000041324/ENSMUSG00000031565/ENSMUSG00000030849/ENSMUSG00000062232/ENSMUSG00000038539 |
| 11 | 4 | ENSMUSG0 | 7 | ENSMUSG00000027995/ENSMUSG00000003032/ENSMUSG00000024789/ENSMUSG00000027832/ENSMUSG00000022037/ENSMUSG00000032487/ENSMUSG0 |
| 19 | 3 | ENSMUSG0 | 16 | ENSMUSG00000005413/ENSMUSG00000041324/ENSMUSG00000024359/ENSMUSG00000038615/ENSMUSG00000021540/ENSMUSG00000029287/ENSMUSG0 |
| 51 | 20 | ENSMUSG0 | 31 | ENSMUSG00000021670/ENSMUSG00000027995/ENSMUSG00000001467/ENSMUSG00000051439/ENSMUSG00000029482/ENSMUSG00000026596/ENSMUSG0 |
| 34 | 11 | ENSMUSG0 | 23 | ENSMUSG00000028128/ENSMUSG00000040152/ENSMUSG00000037411/ENSMUSG00000009376/ENSMUSG00000037071/ENSMUSG00000031668/ENSMUSG0 |
| 37 | 12 | ENSMUSG0 | 25 | ENSMUSG00000021670/ENSMUSG00000041324/ENSMUSG00000031565/ENSMUSG00000029482/ENSMUSG00000056758/ENSMUSG00000039004/ENSMUSG0 |
| 10 | 9 | ENSMUSG0 | 1 | ENSMUSG00000030849 |
| 10 | 5 | ENSMUSG0 | 5 | ENSMUSG00000026596/ENSMUSG00000002489/ENSMUSG00000027996/ENSMUSG00000081683/ENSMUSG00000031548 |
| 10 | 6 | ENSMUSG0 | 4 | ENSMUSG00000005413/ENSMUSG00000023067/ENSMUSG00000001419/ENSMUSG00000024789 |
| 10 | 6 | ENSMUSG0 | 4 | ENSMUSG00000005413/ENSMUSG00000023067/ENSMUSG00000001419/ENSMUSG00000024789 |
| 9 | 1 | ENSMUSG0 | 8 | ENSMUSG00000031960/ENSMUSG00000029777/ENSMUSG00000068739/ENSMUSG00000024587/ENSMUSG00000022241/ENSMUSG00000028811/ENSMUSG0 |
| 9 | 0 | | 9 | ENSMUSG00000042406/ENSMUSG00000031490/ENSMUSG00000037331/ENSMUSG00000023067/ENSMUSG00000031668/ENSMUSG00000028893/ENSMUSG0 |
| 26 | 6 | ENSMUSG0 | 20 | ENSMUSG00000032501/ENSMUSG00000031770/ENSMUSG00000053581/ENSMUSG00000027954/ENSMUSG00000038615/ENSMUSG00000032715/ENSMUSG0 |
| 21 | 6 | ENSMUSG0 | 15 | ENSMUSG00000028128/ENSMUSG00000022346/ENSMUSG00000027954/ENSMUSG00000031668/ENSMUSG00000004040/ENSMUSG00000019997/ENSMUSG0 |
| 29 | 6 | ENSMUSG0 | 23 | ENSMUSG00000032193/ENSMUSG00000032501/ENSMUSG00000031770/ENSMUSG00000053581/ENSMUSG00000027954/ENSMUSG00000038615/ENSMUSG0 |
| 36 | 7 | ENSMUSG0 | 29 | ENSMUSG00000040152/ENSMUSG00000026360/ENSMUSG00000046062/ENSMUSG00000031960/ENSMUSG00000031490/ENSMUSG00000037331/ENSMUSG0 |
| 14 | 8 | ENSMUSG0 | 6 | ENSMUSG00000000555/ENSMUSG00000001131/ENSMUSG00000019997/ENSMUSG00000022817/ENSMUSG00000037815/ENSMUSG00000040945 |
| 34 | 12 | ENSMUSG0 | 22 | ENSMUSG00000024378/ENSMUSG00000022463/ENSMUSG00000025408/ENSMUSG00000022351/ENSMUSG00000042644/ENSMUSG00000045294/ENSMUSG0 |
| 7 | 1 | ENSMUSG0 | 6 | ENSMUSG00000005413/ENSMUSG00000025373/ENSMUSG00000033909/ENSMUSG00000049323/ENSMUSG00000024413/ENSMUSG00000026812 |
| 7 | 1 | ENSMUSG0 | 6 | ENSMUSG00000005413/ENSMUSG00000027995/ENSMUSG00000020458/ENSMUSG00000022508/ENSMUSG00000031451/ENSMUSG00000020075 |
| 7 | 4 | ENSMUSG0 | 3 | ENSMUSG00000040945/ENSMUSG00000009907/ENSMUSG00000031393 |
| 7 | 4 | ENSMUSG0 | 3 | ENSMUSG00000000567/ENSMUSG00000027954/ENSMUSG00000030341 |
| 6 | 1 | ENSMUSG0 | 5 | ENSMUSG00000027397/ENSMUSG00000031565/ENSMUSG00000042406/ENSMUSG00000022265/ENSMUSG00000056501 |
| 6 | 2 | ENSMUSG0 | 4 | ENSMUSG00000020458/ENSMUSG00000031785/ENSMUSG00000028613/ENSMUSG00000020745 |
| 6 | 2 | ENSMUSG0 | 4 | ENSMUSG00000020458/ENSMUSG00000031785/ENSMUSG00000028613/ENSMUSG00000020745 |
| 6 | 1 | ENSMUSG0 | 5 | ENSMUSG00000030536/ENSMUSG00000033955/ENSMUSG00000023951/ENSMUSG00000029816/ENSMUSG00000074934 |
| 6 | 2 | ENSMUSG0 | 4 | ENSMUSG00000041324/ENSMUSG00000039004/ENSMUSG00000070645/ENSMUSG00000018634 |
| 6 | 4 | ENSMUSG0 | 2 | ENSMUSG00000029467/ENSMUSG00000028161 |
| 6 | 4 | ENSMUSG0 | 2 | ENSMUSG00000038418/ENSMUSG00000027947 |
| 26 | 9 | ENSMUSG0 | 17 | ENSMUSG00000022346/ENSMUSG00000042406/ENSMUSG00000038615/ENSMUSG00000028893/ENSMUSG00000037012/ENSMUSG00000020889/ENSMUSG0 |
| 20 | 7 | ENSMUSG0 | 13 | ENSMUSG00000028128/ENSMUSG00000040152/ENSMUSG00000037411/ENSMUSG00000021950/ENSMUSG00000031778/ENSMUSG00000031451/ENSMUSG0 |
| 11 | 4 | ENSMUSG0 | 7 | ENSMUSG00000032193/ENSMUSG00000027995/ENSMUSG00000031778/ENSMUSG00000024789/ENSMUSG00000020889/ENSMUSG00000022037/ENSMUSG0 |
| 48 | 20 | ENSMUSG0 | 28 | ENSMUSG00000021670/ENSMUSG00000027995/ENSMUSG00000001467/ENSMUSG00000051439/ENSMUSG00000029482/ENSMUSG00000026596/ENSMUSG0 |
| 22 | 3 | ENSMUSG0 | 19 | ENSMUSG00000031770/ENSMUSG00000031668/ENSMUSG00000025408/ENSMUSG00000032115/ENSMUSG00000020484/ENSMUSG00000049792/ENSMUSG0 |
| 23 | 3 | ENSMUSG0 | 20 | ENSMUSG00000003032/ENSMUSG00000050335/ENSMUSG00000026596/ENSMUSG00000039899/ENSMUSG00000039004/ENSMUSG00000022508/ENSMUSG0 |
| 16 | 4 | ENSMUSG0 | 12 | ENSMUSG00000032193/ENSMUSG00000027995/ENSMUSG00000017929/ENSMUSG00000029050/ENSMUSG00000020889/ENSMUSG00000026728/ENSMUSG0 |
| 38 | 13 | ENSMUSG0 | 25 | ENSMUSG00000024912/ENSMUSG00000022346/ENSMUSG00000031565/ENSMUSG00000009376/ENSMUSG00000031490/ENSMUSG00000023067/ENSMUSG0 |
| 50 | 14 | ENSMUSG0 | 36 | ENSMUSG00000022346/ENSMUSG00000020917/ENSMUSG00000024378/ENSMUSG00000037071/ENSMUSG00000005069/ENSMUSG00000029482/ENSMUSG0 |
| 12 | 4 | ENSMUSG0 | 8 | ENSMUSG00000000567/ENSMUSG00000030849/ENSMUSG00000031849/ENSMUSG00000023951/ENSMUSG00000026836/ENSMUSG00000028179/ENSMUSG0 |
| 12 | 4 | ENSMUSG0 | 8 | ENSMUSG00000040152/ENSMUSG00000037411/ENSMUSG00000031849/ENSMUSG00000042594/ENSMUSG00000050965/ENSMUSG00000015839/ENSMUSG0 |
| 26 | 12 | ENSMUSG0 | 14 | ENSMUSG00000027995/ENSMUSG00000029675/ENSMUSG00000009376/ENSMUSG00000019970/ENSMUSG00000040021/ENSMUSG00000024533/ENSMUSG0 |
| 14 | 5 | ENSMUSG0 | 9 | ENSMUSG00000002489/ENSMUSG00000031714/ENSMUSG00000027996/ENSMUSG00000081683/ENSMUSG00000039384/ENSMUSG00000027177/ENSMUSG0 |
| 8 | 3 | ENSMUSG0 | 5 | ENSMUSG00000000567/ENSMUSG00000014444/ENSMUSG00000033161/ENSMUSG00000039952/ENSMUSG00000029361 |
| 8 | 3 | ENSMUSG0 | 5 | ENSMUSG00000004040/ENSMUSG00000045983/ENSMUSG00000030527/ENSMUSG00000027665/ENSMUSG00000050697 |
| 8 | 4 | ENSMUSG0 | 4 | ENSMUSG00000022367/ENSMUSG00000041959/ENSMUSG00000022425/ENSMUSG00000003814 |

0000033323/ENSMUSG00000022752/ENSMUSG00000024431/ENSMUSG00000074934/ENSMUSG00000005871
0000020745/ENSMUSG00000039952/ENSMUSG00000015839/ENSMUSG00000031548

0000037902
0000004040/ENSMUSG00000056216/ENSMUSG00000022508/ENSMUSG00000024789/ENSMUSG00000052040/ENSMUSG00000023951/ENSMUSG00000042594/ENSMUSG00000034201/ENSMUSG0000002
0000056758/ENSMUSG00000031778/ENSMUSG00000028641/ENSMUSG00000002489/ENSMUSG00000020484/ENSMUSG00000024789/ENSMUSG00000032340/ENSMUSG00000031451/ENSMUSG0000003
0000021950/ENSMUSG00000031778/ENSMUSG00000020484/ENSMUSG00000031451/ENSMUSG00000031849/ENSMUSG00000023951/ENSMUSG00000053797/ENSMUSG00000042594/ENSMUSG0000004
0000002489/ENSMUSG00000024789/ENSMUSG00000070645/ENSMUSG00000020889/ENSMUSG00000026796/ENSMUSG00000030059/ENSMUSG00000028161/ENSMUSG00000018634/ENSMUSG0000003

0000037851/ENSMUSG00000010755
0000024423/ENSMUSG00000028646/ENSMUSG00000026628
0000020484/ENSMUSG00000020601/ENSMUSG00000049792/ENSMUSG00000030059/ENSMUSG00000040021/ENSMUSG00000061436/ENSMUSG00000022037/ENSMUSG00000025234/ENSMUSG0000002
0000020349/ENSMUSG00000024789/ENSMUSG00000027996/ENSMUSG00000031523/ENSMUSG00000035199/ENSMUSG00000029249/ENSMUSG00000035900/ENSMUSG00000056071/ENSMUSG0000000
0000032715/ENSMUSG00000020484/ENSMUSG00000020601/ENSMUSG00000049792/ENSMUSG00000030530/ENSMUSG00000030059/ENSMUSG00000040021/ENSMUSG00000061436/ENSMUSG0000002
0000031668/ENSMUSG00000015846/ENSMUSG00000028893/ENSMUSG00000004040/ENSMUSG00000020423/ENSMUSG00000035093/ENSMUSG00000020594/ENSMUSG00000026728/ENSMUSG0000004

0000035941/ENSMUSG00000031778/ENSMUSG00000037012/ENSMUSG00000079111/ENSMUSG00000048490/ENSMUSG00000040021/ENSMUSG00000020745/ENSMUSG00000096054/ENSMUSG0000002

0000028673/ENSMUSG00000024085/ENSMUSG00000030747/ENSMUSG00000027665/ENSMUSG00000044167/ENSMUSG00000063229/ENSMUSG00000024431/ENSMUSG00000050697/ENSMUSG0000000
0000031849/ENSMUSG00000042594/ENSMUSG00000050965/ENSMUSG00000015839/ENSMUSG00000026797/ENSMUSG00000056071/ENSMUSG00000027712
0000032487
0000031778/ENSMUSG00000028641/ENSMUSG00000002489/ENSMUSG00000020484/ENSMUSG00000024789/ENSMUSG00000032340/ENSMUSG00000031451/ENSMUSG00000037012/ENSMUSG0000002
0000026389/ENSMUSG00000005087/ENSMUSG00000046223/ENSMUSG00000044337/ENSMUSG00000021070/ENSMUSG00000022037/ENSMUSG00000032487/ENSMUSG00000027230/ENSMUSG0000001
0000024789/ENSMUSG00000056501/ENSMUSG00000005087/ENSMUSG00000024098/ENSMUSG00000023951/ENSMUSG00000042594/ENSMUSG00000028042/ENSMUSG00000021149/ENSMUSG0000007
0000005125/ENSMUSG00000021379/ENSMUSG00000053398/ENSMUSG00000022037/ENSMUSG00000039952/ENSMUSG00000056071
0000030849/ENSMUSG00000040463/ENSMUSG00000019997/ENSMUSG00000029752/ENSMUSG00000026361/ENSMUSG00000045983/ENSMUSG00000005533/ENSMUSG00000006585/ENSMUSG0000002
0000038615/ENSMUSG00000003541/ENSMUSG00000032883/ENSMUSG00000031668/ENSMUSG00000025203/ENSMUSG00000028893/ENSMUSG00000045294/ENSMUSG00000004040/ENSMUSG0000002
0000031393/ENSMUSG00000042903
0000056071/ENSMUSG00000027712
0000032702/ENSMUSG00000041570/ENSMUSG00000038859/ENSMUSG00000044147/ENSMUSG00000031393/ENSMUSG00000025372/ENSMUSG00000002365/ENSMUSG00000005871
0000032717/ENSMUSG00000031548/ENSMUSG00000024182

24182/ENSMUSG00000031543
37012/ENSMUSG00000020889/ENSMUSG00000037447/ENSMUSG00000025340/ENSMUSG00000028161/ENSMUSG00000033717/ENSMUSG00000029249/ENSMUSG00000029361/ENSMUSG00000031548/E
47281/ENSMUSG00000028042/ENSMUSG00000050965/ENSMUSG00000063142/ENSMUSG00000068740/ENSMUSG00000015839/ENSMUSG00000026797/ENSMUSG00000056071/ENSMUSG00000027712
33717/ENSMUSG00000029249/ENSMUSG00000029361/ENSMUSG00000031548/ENSMUSG00000035828/ENSMUSG00000020684/ENSMUSG00000021794/ENSMUSG00000035621/ENSMUSG00000027712/E

28793/ENSMUSG00000032350/ENSMUSG00000038068/ENSMUSG00000025326/ENSMUSG00000028277/ENSMUSG00000024182
02083
22037/ENSMUSG00000025234/ENSMUSG00000028793/ENSMUSG00000032350/ENSMUSG00000038068/ENSMUSG00000009291/ENSMUSG00000025326/ENSMUSG00000028277/ENSMUSG00000024182
45983/ENSMUSG00000003135/ENSMUSG00000033499/ENSMUSG00000030738/ENSMUSG00000024423/ENSMUSG00000050965/ENSMUSG00000026254/ENSMUSG00000003814/ENSMUSG00000045896/E

21831/ENSMUSG00000030747/ENSMUSG00000039952/ENSMUSG00000030287/ENSMUSG00000002083/ENSMUSG00000043421/ENSMUSG00000027712/ENSMUSG00000034593

05886/ENSMUSG00000035621/ENSMUSG00000026628

20889/ENSMUSG00000037447/ENSMUSG00000025340/ENSMUSG00000028161/ENSMUSG00000033717/ENSMUSG00000029249/ENSMUSG00000029361/ENSMUSG00000031548/ENSMUSG00000044147/E
15839/ENSMUSG00000009291/ENSMUSG00000056071/ENSMUSG00000002083/ENSMUSG00000024807
79037/ENSMUSG00000056050/ENSMUSG00000029816/ENSMUSG00000024425/ENSMUSG00000085795/ENSMUSG00000045394

20745/ENSMUSG00000034154/ENSMUSG00000109511/ENSMUSG00000050965/ENSMUSG00000040945/ENSMUSG00000009907/ENSMUSG00000003814/ENSMUSG00000018707/ENSMUSG00000000184/E
25153/ENSMUSG00000032715/ENSMUSG00000029752/ENSMUSG00000037012/ENSMUSG00000020889/ENSMUSG00000030341/ENSMUSG00000022708/ENSMUSG00000039202/ENSMUSG00000004267/E

:NSMUSG00000044147/ENSMUSG00000035828/ENSMUSG00000036391/ENSMUSG00000020684/ENSMUSG00000056071/ENSMUSG00000021794/ENSMUSG00000035621/ENSMUSG00000027712

:NSMUSG00000022808/ENSMUSG00000034593

:NSMUSG00000040025/ENSMUSG00000020091/ENSMUSG00000022412/ENSMUSG00000021838/ENSMUSG00000056071/ENSMUSG00000026812

:NSMUSG00000035828/ENSMUSG00000036391/ENSMUSG00000021794/ENSMUSG00000035621/ENSMUSG00000027712

:NSMUSG00000031393/ENSMUSG00000042903
:NSMUSG00000030747/ENSMUSG00000032349/ENSMUSG00000030168/ENSMUSG00000053398/ENSMUSG00000026313/ENSMUSG00000038633/ENSMUSG00000032487/ENSMUSG00000024052/ENSMUS

;G00000039497/ENSMUSG00000063229/ENSMUSG00000026623/ENSMUSG00000050697/ENSMUSG00000034593

| Category | GOID | Description | GeneRatio | BgRatio | pvalue | padj | geneID | geneName |
|---|---|---|---|---|---|---|---|---|
| BP | GO:003192 | TOR signaling | 17\|1209 | 117\|1609 | 0.00641 | 0.04198 | ENSMUS | Slc7a1/Fnip2/Eif4ebp1/Larp1/Sesn2/Xbp1/Smcr8/Sik1/Gas6/Icr |
| BP | GO:004254 | response to hydrogen peroxide | 17\|1209 | 117\|1609 | 0.00641 | 0.04198 | ENSMUS | Hmox1/Met/Ppp1r15b/Hsph1/Rela/Axl/Kdm6b/Impact/Pycr1/ |
| BP | GO:004666 | female sex differentiation | 17\|1209 | 117\|1609 | 0.00641 | 0.04198 | ENSMUS | Serpine1/Inhba/Ncoa3/Tiparp/Cebpb/Vegfa/Nrip1/Axl/Zfx/Ptx3, |
| BP | GO:005081 | coagulation | 20\|1209 | 146\|1609 | 0.00642 | 0.04204 | ENSMUS | F3/Thbs1/Serpine1/Serping1/F13a1/Anxa8/Cd34/Cx3cl1/Gas6/ |
| BP | GO:000317 | heart valve morphogenesis | 9\|1209 | 46\|16097 | 0.00647 | 0.0421 | ENSMUS | Sox9/Efna1/Cyr61/Robo1/Nfatc1/Notch1/Robo2/Acvr1/Mdm4 |
| BP | GO:001059 | negative regulation of endothelial cell migration | 9\|1209 | 46\|16097 | 0.00647 | 0.0421 | ENSMUS | Thbs1/Klf4/Serpinf1/Spred1/Meox2/Pik3r2/Notch1/Vash1/Mec |
| BP | GO:004801 | vascular endothelial growth factor receptor signaling pathway | 9\|1209 | 46\|16097 | 0.00647 | 0.0421 | ENSMUS | Tnmd/Xbp1/Nrp1/Flt4/Vegfa/Nedd4/Tek/Hhex/Kdr |
| BP | GO:000762 | circadian rhythm | 24\|1209 | 186\|1609 | 0.00648 | 0.0421 | ENSMUS | Egr1/Atf4/Noct/Mybbp1a/Fbxw11/Klf10/Sik1/Maged1/Bhlhe40 |
| BP | GO:003515 | regulation of tube size | 21\|1209 | 156\|1609 | 0.00649 | 0.0421 | ENSMUS | Hmgcr/Rgs2/Cx3cl1/Tgfbr3/Egfr/Agtr2/Dusp5/Comp/Plod3/Npr |
| BP | GO:003529 | regulation of tube diameter | 21\|1209 | 156\|1609 | 0.00649 | 0.0421 | ENSMUS | Hmgcr/Rgs2/Cx3cl1/Tgfbr3/Egfr/Agtr2/Dusp5/Comp/Plod3/Npr |
| BP | GO:005088 | regulation of blood vessel size | 21\|1209 | 156\|1609 | 0.00649 | 0.0421 | ENSMUS | Hmgcr/Rgs2/Cx3cl1/Tgfbr3/Egfr/Agtr2/Dusp5/Comp/Plod3/Npr |
| BP | GO:009774 | regulation of blood vessel diameter | 21\|1209 | 156\|1609 | 0.00649 | 0.0421 | ENSMUS | Hmgcr/Rgs2/Cx3cl1/Tgfbr3/Egfr/Agtr2/Dusp5/Comp/Plod3/Npr |
| BP | GO:000600 | glucose metabolic process | 23\|1209 | 176\|1609 | 0.00651 | 0.04213 | ENSMUS | Myc/Igfbp3/Atf4/Nfe2l1/Igfbp4/Sesn2/C1qtnf3/Sik1/Hk1/Nr1d1 |
| BP | GO:007169 | anatomical structure maturation | 23\|1209 | 176\|1609 | 0.00651 | 0.04213 | ENSMUS | Xylt1/Fem1b/Cdkn1c/Cdh5/Ddit3/Abl2/Rflna/B4galt5/Xbp1/Igf |
| BP | GO:003527 | endocrine system development | 16\|1209 | 108\|1609 | 0.00666 | 0.04303 | ENSMUS | Sox9/Cdkn1c/Il6ra/Eif2ak3/Hmga2/Bmp6/Nog/Tgfbr1/Igf1r/Pbx |
| BP | GO:003362 | cell adhesion mediated by integrin | 11\|1209 | 63\|16097 | 0.00676 | 0.04322 | ENSMUS | Serpine1/Itga5/Piezo1/Efna1/Itgb5/Epha2/Itgbl1/Nov/Itga11/Ep |
| BP | GO:005081 | regulation of steroid biosynthetic process | 11\|1209 | 63\|16097 | 0.00676 | 0.04322 | ENSMUS | Egr1/Stard4/Atp1a1/Insig1/Bmp6/Nr1d1/Igf1r/Igf1/Dab2/Rest/ |
| BP | GO:006104 | negative regulation of wound healing | 11\|1209 | 63\|16097 | 0.00676 | 0.04322 | ENSMUS | Thbs1/Hmgcr/Serpine1/Cd109/Serping1/Cdkn1a/Cd34/Dusp10 |
| BP | GO:003292 | negative regulation of activin receptor signaling pathway | 4\|1209 | 11\|16097 | 0.00681 | 0.04322 | ENSMUS | Ski/Lemd3/Fst/Acvr1 |
| BP | GO:004356 | positive regulation of insulin-like growth factor receptor signal | 4\|1209 | 11\|16097 | 0.00681 | 0.04322 | ENSMUS | Igfbp5/Igfbp3/Igfbp4/Igf1 |
| BP | GO:006076 | epithelial cell proliferation involved in prostate gland developr | 4\|1209 | 11\|16097 | 0.00681 | 0.04322 | ENSMUS | Sox9/Serpinf1/Notch1/Apc |
| BP | GO:006129 | retina vasculature morphogenesis in camera-type eye | 4\|1209 | 11\|16097 | 0.00681 | 0.04322 | ENSMUS | Col4a1/Clic4/Nrp1/Rhoj |
| BP | GO:007027 | extracellular matrix constituent secretion | 4\|1209 | 11\|16097 | 0.00681 | 0.04322 | ENSMUS | Ctgf/Tnfrsf1a/Notch1/Creb3l1 |
| BP | GO:008610 | G protein-coupled receptor signaling pathway involved in hear | 4\|1209 | 11\|16097 | 0.00681 | 0.04322 | ENSMUS | Rgs2/Pde4b/Gnai2/Nos1 |
| BP | GO:009708 | vascular smooth muscle cell development | 4\|1209 | 11\|16097 | 0.00681 | 0.04322 | ENSMUS | Comp/Vegfa/Notch1/Hes1 |
| BP | GO:009742 | liver regeneration | 4\|1209 | 11\|16097 | 0.00681 | 0.04322 | ENSMUS | Sulf2/Hmox1/Cebpb/Ccnd1 |
| BP | GO:015007 | negative regulation of neuroinflammatory response | 4\|1209 | 11\|16097 | 0.00681 | 0.04322 | ENSMUS | Ldlr/Cx3cl1/Nr1d1/Igf1 |
| BP | GO:000018 | inactivation of MAPK activity | 5\|1209 | 17\|16097 | 0.00684 | 0.04322 | ENSMUS | Dusp1/Spred1/Dusp5/Dusp10/Prkca |
| BP | GO:002002 | hemoglobin metabolic process | 5\|1209 | 17\|16097 | 0.00684 | 0.04322 | ENSMUS | Klf4/Inhba/Cat/Alas2/Axin1 |
| BP | GO:003593 | corticosteroid hormone secretion | 5\|1209 | 17\|16097 | 0.00684 | 0.04322 | ENSMUS | Bmp6/Agtr2/Ren1/Crhr1/Dab2 |
| BP | GO:004688 | positive regulation of hormone biosynthetic process | 5\|1209 | 17\|16097 | 0.00684 | 0.04322 | ENSMUS | Egr1/Bmp6/Igf1r/Igf1/Dab2 |
| BP | GO:005148 | negative regulation of cytosolic calcium ion concentration | 5\|1209 | 17\|16097 | 0.00684 | 0.04322 | ENSMUS | Il6st/Gtf2i/Atp1a2/Nos1/Atp2b1 |
| BP | GO:007057 | negative regulation of neuron projection regeneration | 5\|1209 | 17\|16097 | 0.00684 | 0.04322 | ENSMUS | Xylt1/Rtn4/Thy1/Neo1/Epha4 |
| BP | GO:000194 | hair follicle development | 15\|1209 | 99\|16097 | 0.00687 | 0.0434 | ENSMUS | Igfbp5/Sox9/Inhba/Cd109/Fgfr2/Egfr/Rela/Notch1/Lgr5/Ptgs2/ |
| BP | GO:200005 | regulation of ubiquitin-dependent protein catabolic process | 20\|1209 | 147\|1609 | 0.00693 | 0.04368 | ENSMUS | Hspa1b/Hspa1a/Trib1/Herpud1/Zfand2a/Trib3/Trib2/Bag5/Lats |
| BP | GO:000761 | learning | 21\|1209 | 157\|1609 | 0.00698 | 0.0439 | ENSMUS | Hmgcr/Tlr2/Jun/Abl2/Sgk1/Synpo/Btg2/Nog/Foxp2/Cic/Nptx2/ |
| BP | GO:009872 | maintenance of cell number | 21\|1209 | 157\|1609 | 0.00698 | 0.0439 | ENSMUS | Sox9/Klf4/Fgfr1/Ncoa3/Hmga2/Stat3/Klf10/Ski/Nog/Cdc73/Igf1 |
| BP | GO:000721 | Notch signaling pathway | 22\|1209 | 167\|1609 | 0.007 | 0.0439 | ENSMUS | Sox9/Postn/Dlk1/Chac1/Sorbs2/Mmp14/Maml1/Stat3/Bcl6/Il6 |
| BP | GO:003514 | tube formation | 22\|1209 | 167\|1609 | 0.007 | 0.0439 | ENSMUS | Sox9/Abl2/Fgfr2/Ski/Sfrp2/Dlc1/Nog/Hectd1/Vegfa/Notch1/Plx |
| BP | GO:007207 | nephron tubule morphogenesis | 12\|1209 | 72\|16097 | 0.007 | 0.0439 | ENSMUS | Myc/Sox9/Lzts2/Agtr2/Maged1/Nog/Cd44/Vegfa/Pbx1/Hes1/Gr |
| BP | GO:004864 | muscle organ morphogenesis | 14\|1209 | 90\|16097 | 0.00702 | 0.04392 | ENSMUS | Col3a1/Tpm1/Fgfr2/Rxra/Mylk/Tgfbr3/Nog/Tgfbr1/Notch1/Col1 |
| BP | GO:000762 | locomotory behavior | 27\|1209 | 218\|1609 | 0.00702 | 0.04392 | ENSMUS | Egr1/Rcan1/Dnm1/Abl2/Selenop/Arrdc3/Npc1/Cxcl12/Pafah1b |
| BP | GO:000170 | endoderm formation | 7\|1209 | 31\|16097 | 0.00704 | 0.04397 | ENSMUS | Itga5/Inhba/Col5a2/Col12a1/Nog/Cdc73/Sox7 |
| BP | GO:009724 | amyloid-beta clearance | 7\|1209 | 31\|16097 | 0.00704 | 0.04397 | ENSMUS | Hmgcr/Ldlr/Cyp51/Cd36/Igf1r/Clu/Cltc |
| BP | GO:003026 | entry into host cell | 10\|1209 | 55\|16097 | 0.0072 | 0.04461 | ENSMUS | Lgals1/Npc1/Gas6/Axl/Ptx3/Ifitm3/Cav2/Exoc7/Uvrag/Vps18 |
| BP | GO:004440 | entry into host | 10\|1209 | 55\|16097 | 0.0072 | 0.04461 | ENSMUS | Lgals1/Npc1/Gas6/Axl/Ptx3/Ifitm3/Cav2/Exoc7/Uvrag/Vps18 |
| BP | GO:005077 | positive regulation of dendrite morphogenesis | 10\|1209 | 55\|16097 | 0.0072 | 0.04461 | ENSMUS | Met/Sgk1/Tiam1/Lrp8/Pafah1b1/Ptprd/Epha4/Rab21/Dbn1/Bai |
| BP | GO:005180 | entry into cell of other organism involved in symbiotic interact | 10\|1209 | 55\|16097 | 0.0072 | 0.04461 | ENSMUS | Lgals1/Npc1/Gas6/Axl/Ptx3/Ifitm3/Cav2/Exoc7/Uvrag/Vps18 |
| BP | GO:005182 | entry into other organism involved in symbiotic interaction | 10\|1209 | 55\|16097 | 0.0072 | 0.04461 | ENSMUS | Lgals1/Npc1/Gas6/Axl/Ptx3/Ifitm3/Cav2/Exoc7/Uvrag/Vps18 |
| BP | GO:009018 | regulation of kidney development | 10\|1209 | 55\|16097 | 0.0072 | 0.04461 | ENSMUS | Myc/Sox9/Egr1/Il6ra/Agtr2/Maged1/Nog/Vegfa/Grem1/Sox8 |
| BP | GO:005134 | positive regulation of hydrolase activity | 51\|1209 | 479\|1609 | 0.00721 | 0.04465 | ENSMUS | F3/Myc/Rgs16/Fgfr1/S100a10/Ppp1r15b/Efna1/Eif2ak3/Cyr61/ |

| Count | Up | Up_Gene_id | Down | Down_Gene_id |
|---|---|---|---|---|
| 17 | 2 | ENSMUSG0 | 15 | ENSMUSG00000041313/ENSMUSG00000061175/ENSMUSG00000031490/ENSMUSG00000037331/ENSMUSG00000028893/ENSMUSG00000020484/ENSMUSG0 |
| 17 | 5 | ENSMUSG0 | 12 | ENSMUSG00000005413/ENSMUSG00000009376/ENSMUSG00000046062/ENSMUSG00000024927/ENSMUSG00000018476/ENSMUSG00000024423/ENSMUSG0 |
| 17 | 5 | ENSMUSG0 | 12 | ENSMUSG00000037411/ENSMUSG00000041324/ENSMUSG00000027678/ENSMUSG00000034640/ENSMUSG00000056501/ENSMUSG00000023951/ENSMUSG0 |
| 20 | 7 | ENSMUSG0 | 13 | ENSMUSG00000028128/ENSMUSG00000040152/ENSMUSG00000037411/ENSMUSG00000021950/ENSMUSG00000031778/ENSMUSG00000031451/ENSMUSG0 |
| 9 | 6 | ENSMUSG0 | 3 | ENSMUSG00000000567/ENSMUSG00000027954/ENSMUSG00000026836 |
| 9 | 4 | ENSMUSG0 | 5 | ENSMUSG00000040152/ENSMUSG00000003032/ENSMUSG00000000753/ENSMUSG00000027351/ENSMUSG00000031393 |
| 9 | 7 | ENSMUSG0 | 2 | ENSMUSG00000020484/ENSMUSG00000023951 |
| 24 | 6 | ENSMUSG0 | 18 | ENSMUSG00000038418/ENSMUSG00000042406/ENSMUSG00000023087/ENSMUSG00000040463/ENSMUSG00000020271/ENSMUSG00000037465/ENSMUSG0 |
| 21 | 7 | ENSMUSG0 | 14 | ENSMUSG00000021670/ENSMUSG00000026360/ENSMUSG00000031778/ENSMUSG00000029287/ENSMUSG00000034765/ENSMUSG00000031849/ENSMUSG0 |
| 21 | 7 | ENSMUSG0 | 14 | ENSMUSG00000021670/ENSMUSG00000026360/ENSMUSG00000031778/ENSMUSG00000029287/ENSMUSG00000034765/ENSMUSG00000031849/ENSMUSG0 |
| 21 | 7 | ENSMUSG0 | 14 | ENSMUSG00000021670/ENSMUSG00000026360/ENSMUSG00000031778/ENSMUSG00000029287/ENSMUSG00000034765/ENSMUSG00000031849/ENSMUSG0 |
| 21 | 7 | ENSMUSG0 | 14 | ENSMUSG00000021670/ENSMUSG00000026360/ENSMUSG00000031778/ENSMUSG00000029287/ENSMUSG00000034765/ENSMUSG00000031849/ENSMUSG0 |
| 23 | 8 | ENSMUSG0 | 15 | ENSMUSG00000022346/ENSMUSG00000042406/ENSMUSG00000038615/ENSMUSG00000028893/ENSMUSG00000037012/ENSMUSG00000020889/ENSMUSG0 |
| 23 | 7 | ENSMUSG0 | 16 | ENSMUSG00000030657/ENSMUSG00000032244/ENSMUSG00000025408/ENSMUSG00000026596/ENSMUSG00000017929/ENSMUSG00000020484/ENSMUSG0 |
| 16 | 5 | ENSMUSG0 | 11 | ENSMUSG00000000567/ENSMUSG00000027947/ENSMUSG00000031668/ENSMUSG00000056758/ENSMUSG00000039004/ENSMUSG00000048616/ENSMUSG0 |
| 11 | 4 | ENSMUSG0 | 7 | ENSMUSG00000037411/ENSMUSG00000000555/ENSMUSG00000014444/ENSMUSG00000027954/ENSMUSG00000022817/ENSMUSG00000006445/ENSMUSG0 |
| 11 | 2 | ENSMUSG0 | 9 | ENSMUSG00000038418/ENSMUSG00000024378/ENSMUSG00000033161/ENSMUSG00000045294/ENSMUSG00000039004/ENSMUSG00000020889/ENSMUSG0 |
| 11 | 3 | ENSMUSG0 | 8 | ENSMUSG00000040152/ENSMUSG00000021670/ENSMUSG00000037411/ENSMUSG00000046186/ENSMUSG00000023067/ENSMUSG00000039384/ENSMUSG0 |
| 4 | 1 | ENSMUSG0 | 3 | ENSMUSG00000029050/ENSMUSG00000048661/ENSMUSG00000026836 |
| 4 | 4 | ENSMUSG0 | 0 | |
| 4 | 1 | ENSMUSG0 | 3 | ENSMUSG00000000567/ENSMUSG00000000753/ENSMUSG00000005871 |
| 4 | 3 | ENSMUSG0 | 1 | ENSMUSG00000037242 |
| 4 | 1 | ENSMUSG0 | 3 | ENSMUSG00000019997/ENSMUSG00000030341/ENSMUSG00000027230 |
| 4 | 1 | ENSMUSG0 | 3 | ENSMUSG00000026360/ENSMUSG00000028525/ENSMUSG00000029361 |
| 4 | 2 | ENSMUSG0 | 2 | ENSMUSG00000031849/ENSMUSG00000023951 |
| 4 | 1 | ENSMUSG0 | 3 | ENSMUSG00000006800/ENSMUSG00000005413/ENSMUSG00000056501 |
| 4 | 1 | ENSMUSG0 | 3 | ENSMUSG00000032193/ENSMUSG00000031778/ENSMUSG00000020889 |
| 5 | 1 | ENSMUSG0 | 4 | ENSMUSG00000027351/ENSMUSG00000034765/ENSMUSG00000039384/ENSMUSG00000050965 |
| 5 | 2 | ENSMUSG0 | 3 | ENSMUSG00000003032/ENSMUSG00000041324/ENSMUSG00000024182 |
| 5 | 2 | ENSMUSG0 | 3 | ENSMUSG00000039004/ENSMUSG00000070645/ENSMUSG00000018634 |
| 5 | 2 | ENSMUSG0 | 3 | ENSMUSG00000038418/ENSMUSG00000039004/ENSMUSG00000005533 |
| 5 | 2 | ENSMUSG0 | 3 | ENSMUSG00000021756/ENSMUSG00000029361/ENSMUSG00000019943 |
| 5 | 2 | ENSMUSG0 | 3 | ENSMUSG00000030657/ENSMUSG00000020458/ENSMUSG00000032340 |
| 15 | 7 | ENSMUSG0 | 8 | ENSMUSG00000000567/ENSMUSG00000041324/ENSMUSG00000046186/ENSMUSG00000030849/ENSMUSG00000024927/ENSMUSG00000032487/ENSMUSG0 |
| 20 | 5 | ENSMUSG0 | 15 | ENSMUSG00000032501/ENSMUSG00000031770/ENSMUSG00000053581/ENSMUSG00000032715/ENSMUSG00000020601/ENSMUSG00000049792/ENSMUSG0 |
| 21 | 7 | ENSMUSG0 | 14 | ENSMUSG00000021670/ENSMUSG00000027995/ENSMUSG00000026596/ENSMUSG00000019970/ENSMUSG00000020423/ENSMUSG00000048616/ENSMUSG0 |
| 21 | 4 | ENSMUSG0 | 17 | ENSMUSG00000000567/ENSMUSG00000003032/ENSMUSG00000031565/ENSMUSG00000027678/ENSMUSG00000056758/ENSMUSG00000004040/ENSMUSG0 |
| 22 | 13 | ENSMUSG0 | 9 | ENSMUSG00000000567/ENSMUSG00000040856/ENSMUSG00000027313/ENSMUSG00000050567/ENSMUSG00000004040/ENSMUSG00000022508/ENSMUSG0 |
| 22 | 7 | ENSMUSG0 | 15 | ENSMUSG00000000567/ENSMUSG00000026596/ENSMUSG00000030849/ENSMUSG00000029050/ENSMUSG00000027996/ENSMUSG00000031523/ENSMUSG0 |
| 12 | 5 | ENSMUSG0 | 7 | ENSMUSG00000022346/ENSMUSG00000000567/ENSMUSG00000035342/ENSMUSG00000048616/ENSMUSG00000005087/ENSMUSG00000023951/ENSMUSG0 |
| 14 | 9 | ENSMUSG0 | 5 | ENSMUSG00000030849/ENSMUSG00000015846/ENSMUSG00000029287/ENSMUSG00000048616/ENSMUSG00000007613 |
| 27 | 8 | ENSMUSG0 | 19 | ENSMUSG00000038418/ENSMUSG00000022951/ENSMUSG00000026596/ENSMUSG00000074794/ENSMUSG00000024413/ENSMUSG00000020745/ENSMUSG0 |
| 7 | 3 | ENSMUSG0 | 4 | ENSMUSG00000000555/ENSMUSG00000041324/ENSMUSG00000048616/ENSMUSG00000026361 |
| 7 | 1 | ENSMUSG0 | 6 | ENSMUSG00000021670/ENSMUSG00000032193/ENSMUSG00000001467/ENSMUSG00000005533/ENSMUSG00000022037/ENSMUSG00000047126 |
| 10 | 5 | ENSMUSG0 | 5 | ENSMUSG00000024413/ENSMUSG00000031451/ENSMUSG00000027832/ENSMUSG00000000058/ENSMUSG00000034216 |
| 10 | 5 | ENSMUSG0 | 5 | ENSMUSG00000024413/ENSMUSG00000031451/ENSMUSG00000027832/ENSMUSG00000000058/ENSMUSG00000034216 |
| 10 | 3 | ENSMUSG0 | 7 | ENSMUSG00000009376/ENSMUSG00000019970/ENSMUSG00000002489/ENSMUSG00000028613/ENSMUSG00000020745/ENSMUSG00000020132/ENSMUSG0 |
| 10 | 5 | ENSMUSG0 | 5 | ENSMUSG00000024413/ENSMUSG00000031451/ENSMUSG00000027832/ENSMUSG00000000058/ENSMUSG00000034216 |
| 10 | 5 | ENSMUSG0 | 5 | ENSMUSG00000024413/ENSMUSG00000031451/ENSMUSG00000027832/ENSMUSG00000000058/ENSMUSG00000034216 |
| 10 | 3 | ENSMUSG0 | 7 | ENSMUSG00000022346/ENSMUSG00000000567/ENSMUSG00000038418/ENSMUSG00000027947/ENSMUSG00000048616/ENSMUSG00000023951/ENSMUSG0 |
| 51 | 14 | ENSMUSG0 | 37 | ENSMUSG00000028128/ENSMUSG00000022346/ENSMUSG00000031565/ENSMUSG00000041959/ENSMUSG00000046062/ENSMUSG00000027954/ENSMUSG0 |

0000049323/ENSMUSG00000031451/ENSMUSG00000039662/ENSMUSG00000034135/ENSMUSG00000035992/ENSMUSG00000020091/ENSMUSG00000028646/ENSMUSG00000050697/ENSMUSG0000002
0000040268/ENSMUSG00000015839/ENSMUSG00000009630/ENSMUSG00000050697/ENSMUSG00000037902/ENSMUSG00000004207
0000048490/ENSMUSG00000079509/ENSMUSG00000027832/ENSMUSG00000040268/ENSMUSG00000031548/ENSMUSG00000025326
0000031849/ENSMUSG00000042594/ENSMUSG00000050965/ENSMUSG00000015839/ENSMUSG00000026797/ENSMUSG00000056071/ENSMUSG00000027712

0000030103/ENSMUSG00000020889/ENSMUSG00000048490/ENSMUSG00000022124/ENSMUSG00000063142/ENSMUSG00000021379/ENSMUSG00000025261/ENSMUSG00000038539/ENSMUSG0000003
0000004846/ENSMUSG00000021070/ENSMUSG00000063142/ENSMUSG00000032487/ENSMUSG00000033717/ENSMUSG00000032350/ENSMUSG00000029361/ENSMUSG00000019943
0000004846/ENSMUSG00000021070/ENSMUSG00000063142/ENSMUSG00000032487/ENSMUSG00000033717/ENSMUSG00000032350/ENSMUSG00000029361/ENSMUSG00000019943
0000004846/ENSMUSG00000021070/ENSMUSG00000063142/ENSMUSG00000032487/ENSMUSG00000033717/ENSMUSG00000032350/ENSMUSG00000029361/ENSMUSG00000019943
0000004846/ENSMUSG00000021070/ENSMUSG00000063142/ENSMUSG00000032487/ENSMUSG00000033717/ENSMUSG00000032350/ENSMUSG00000029361/ENSMUSG00000019943
0000030747/ENSMUSG00000027665/ENSMUSG00000044167/ENSMUSG00000063229/ENSMUSG00000024431/ENSMUSG00000050697/ENSMUSG00000005886/ENSMUSG00000035621/ENSMUSG0000002
0000032487/ENSMUSG00000039952/ENSMUSG00000028047/ENSMUSG00000040025/ENSMUSG00000031393/ENSMUSG00000026826/ENSMUSG00000074934/ENSMUSG00000028661/ENSMUSG0000003
0000007613/ENSMUSG00000005533/ENSMUSG00000018634/ENSMUSG00000022037/ENSMUSG00000024431
0000028661
0000005533/ENSMUSG00000029249/ENSMUSG00000024431
0000042594/ENSMUSG00000027712

0000005871/ENSMUSG00000034593
0000040021/ENSMUSG00000061436/ENSMUSG00000022037/ENSMUSG00000025234/ENSMUSG00000028793/ENSMUSG00000032350/ENSMUSG00000038068/ENSMUSG00000025326/ENSMUSG0000002
0000005442/ENSMUSG00000059991/ENSMUSG00000018634/ENSMUSG00000036564/ENSMUSG00000069895/ENSMUSG00000032487/ENSMUSG00000031393/ENSMUSG00000074923
0000037465/ENSMUSG00000029050/ENSMUSG00000048616/ENSMUSG00000026361/ENSMUSG00000029249/ENSMUSG00000044167/ENSMUSG00000034205/ENSMUSG00000031548/ENSMUSG0000003
0000021756/ENSMUSG00000007817/ENSMUSG00000036450
0000048616/ENSMUSG00000035247/ENSMUSG00000023951/ENSMUSG00000036606/ENSMUSG00000037820/ENSMUSG00000029009/ENSMUSG00000031548/ENSMUSG00000074934/ENSMUSG0000002
0000074934

0000061436/ENSMUSG00000063142/ENSMUSG00000018634/ENSMUSG00000000184/ENSMUSG00000031393/ENSMUSG00000026826/ENSMUSG00000005886/ENSMUSG00000026812/ENSMUSG0000000

0000025372

0000074934
0000031668/ENSMUSG00000026596/ENSMUSG00000030849/ENSMUSG00000031778/ENSMUSG00000004040/ENSMUSG00000002489/ENSMUSG00000019997/ENSMUSG00000020349/ENSMUSG0000001

26812

36432/ENSMUSG00000025326/ENSMUSG00000050697/ENSMUSG00000005886

26628
32096/ENSMUSG00000034593

24182

31540/ENSMUSG00000063382/ENSMUSG00000005871

26812

04207/ENSMUSG00000024182/ENSMUSG00000032096/ENSMUSG00000074923/ENSMUSG00000034593

18736/ENSMUSG00000024789/ENSMUSG00000027996/ENSMUSG00000031523/ENSMUSG00000035199/ENSMUSG00000081683/ENSMUSG00000023951/ENSMUSG00000027465/ENSMUSG00000006445/E

ENSMUSG00000040945/ENSMUSG00000062232/ENSMUSG00000000058/ENSMUSG00000020130/ENSMUSG00000029249/ENSMUSG00000035900/ENSMUSG00000042303/ENSMUSG00000039844/ENSMUS

;G00000056071/ENSMUSG00000002083/ENSMUSG00000026812/ENSMUSG00000004207/ENSMUSG00000035007/ENSMUSG00000002365

| Category | GOID | Description | GeneRatio | BgRatio | pvalue | padj | geneID | geneName |
|---|---|---|---|---|---|---|---|---|
| BP | GO:004654 | development of primary male sexual characteristics | 16\|1209 | 109\|16097 | 0.00728 | 0.04497 | ENSMUS | Sox9/Inhba/Adgrg1/Hmga2/Sfrp2/Ren1/Tgfbr1/Tmf1/Ccnd1/Tb |
| BP | GO:004682 | regulation of nucleocytoplasmic transport | 16\|1209 | 109\|16097 | 0.00728 | 0.04497 | ENSMUS | Ier3/Ppp2ca/Xbp1/Agtr2/Gas6/Tardbp/Cd36/Camk4/Pik3r2/Su |
| BP | GO:000327 | endocardial cushion formation | 6\|1209 | 24\|16097 | 0.00736 | 0.04534 | ENSMUS | Nog/Tgfbr1/Robo1/Notch1/Robo2/Acvr1 |
| BP | GO:007257 | hepatocyte proliferation | 6\|1209 | 24\|16097 | 0.00736 | 0.04534 | ENSMUS | Sulf2/Rtn4/Xbp1/Cebpb/Ptn/Mki67 |
| BP | GO:007257 | epithelial cell proliferation involved in liver morphogenesis | 6\|1209 | 24\|16097 | 0.00736 | 0.04534 | ENSMUS | Sulf2/Rtn4/Xbp1/Cebpb/Ptn/Mki67 |
| BP | GO:000991 | hormone transport | 37\|1209 | 325\|16097 | 0.00738 | 0.04539 | ENSMUS | Hmgcr/Inhba/Fgfr1/Igfbp3/Aacs/Hmga2/Bmp6/Egfr/Tiam1/Agtr |
| BP | GO:009723 | cellular response to toxic substance | 20\|1209 | 148\|16097 | 0.00746 | 0.04579 | ENSMUS | Hmox1/Egr1/Met/Hsph1/Cd36/Prdx6/Abtb2/Fbln5/Rela/Axl/Kc |
| BP | GO:003233 | regulation of chondrocyte differentiation | 9\|1209 | 47\|16097 | 0.00748 | 0.04579 | ENSMUS | Sox9/Bmp6/Rflna/Ctgf/Tgfbr1/Chadl/Rela/Loxl2/Grem1 |
| BP | GO:005501 | ventricular cardiac muscle tissue morphogenesis | 9\|1209 | 47\|16097 | 0.00748 | 0.04579 | ENSMUS | Tpm1/Fgfr2/Rxra/Tgfbr3/Nog/Tgfbr1/Notch1/Col11a1/Ly6e |
| BP | GO:003808 | peptidyl-tyrosine autophosphorylation | 8\|1209 | 39\|16097 | 0.00749 | 0.04579 | ENSMUS | Iqgap1/Abl2/Egfr/Igf1r/Vegfa/Ace/Kdr/Grem1 |
| BP | GO:200082 | regulation of heart morphogenesis | 8\|1209 | 39\|16097 | 0.00749 | 0.04579 | ENSMUS | Sox9/Nog/Tgfbr1/Robo1/Notch1/Robo2/Acvr1/Hes1 |
| BP | GO:000679 | sulfur compound metabolic process | 29\|1209 | 240\|16097 | 0.00751 | 0.04587 | ENSMUS | Sulf2/Chac1/Acly/Xylt1/Hspa9/Nfe2l1/Eif2ak3/Hmgcs1/Chpf/Ch |
| BP | GO:003223 | regulation of actin filament bundle assembly | 15\|1209 | 100\|16097 | 0.00754 | 0.04595 | ENSMUS | S100a10/Met/Tpm1/Synpo/Cx3cl1/Ctgf/Nrp1/Dlc1/Tgfbr1/Pik3 |
| BP | GO:009877 | skin epidermis development | 15\|1209 | 100\|16097 | 0.00754 | 0.04595 | ENSMUS | Igfbp5/Sox9/Inhba/Cd109/Fgfr2/Egfr/Rela/Notch1/Lgr5/Ptgs2/L |
| BP | GO:001063 | negative regulation of epithelial cell migration | 11\|1209 | 64\|16097 | 0.00762 | 0.04642 | ENSMUS | Thbs1/Klf4/Serpinf1/Spred1/Meox2/Tgfbr3/Pik3r2/Dusp10/Not |
| BP | GO:000910 | glycoprotein biosynthetic process | 30\|1209 | 251\|16097 | 0.00771 | 0.0469 | ENSMUS | Xylt1/Csgalnact2/Tet3/Rpn1/Chpf/Chst11/B4galt5/Npc1/Cytl1/ |
| BP | GO:006099 | regulation of dendritic spine development | 14\|1209 | 91\|16097 | 0.00774 | 0.047 | ENSMUS | Mapk6/Efna1/Ngef/Tiam1/Lrp8/Pafah1b1/Epha4/Dbn1/Arf6/Sha |
| BP | GO:007121 | cellular response to biotic stimulus | 25\|1209 | 199\|16097 | 0.00775 | 0.04703 | ENSMUS | Tlr2/Serpine1/Trib1/Cd14/Ppp1r15b/Eif2ak3/Ddit3/Bmp6/Pde |
| BP | GO:000224 | hematopoietic progenitor cell differentiation | 18\|1209 | 129\|16097 | 0.0078 | 0.04724 | ENSMUS | Inhba/Hspa9/Top2a/Vegfa/Notch1/Kcp/Eml1/Fst/Zbtb1/Fnip1/I |
| BP | GO:005072 | positive regulation of inflammatory response | 18\|1209 | 129\|16097 | 0.0078 | 0.04724 | ENSMUS | Ldlr/Tlr2/Serpine1/Lpl/Egfr/Jak2/Fabp4/Tnfrsf1a/Cebpb/Tgm2/F |
| BP | GO:004620 | nitric oxide metabolic process | 12\|1209 | 73\|16097 | 0.00782 | 0.04724 | ENSMUS | Tlr2/Klf4/Cd34/Jak2/Cd36/Igf1/Ddah2/Ptx3/Clu/Ptgs2/Nos1/Si |
| BP | GO:007209 | regulation of stem cell proliferation | 12\|1209 | 73\|16097 | 0.00782 | 0.04724 | ENSMUS | Fgfr1/Ncoa3/Hmga2/Cx3cl1/Vegfa/Nfatc1/Notch1/Ctnna1/Atxn |
| BP | GO:005080 | regulation of synapse structure or activity | 31\|1209 | 262\|16097 | 0.00789 | 0.04762 | ENSMUS | Tlr2/Thbs2/Sorbs2/Efna1/Tuba1a/Dnm1/Ngef/Tiam1/Lrp8/Eif4g |
| BP | GO:003425 | positive regulation of cellular amide metabolic process | 19\|1209 | 139\|16097 | 0.00795 | 0.0479 | ENSMUS | Thbs1/Efna1/Eif2ak3/Cyr61/Rxra/Vim/Larp4b/Eif3c/Impact/Rbm |
| BP | GO:000322 | ventricular cardiac muscle tissue development | 10\|1209 | 56\|16097 | 0.00819 | 0.04917 | ENSMUS | Col14a1/Tpm1/Fgfr2/Rxra/Tgfbr3/Nog/Tgfbr1/Notch1/Col11a1, |
| BP | GO:001046 | mesenchymal cell proliferation | 10\|1209 | 56\|16097 | 0.00819 | 0.04917 | ENSMUS | Myc/Sox9/Fgfr1/Lmna/Fgfr2/Foxp2/Vegfa/Syne1/Ptn/Kdr |
| BP | GO:200073 | regulation of stem cell differentiation | 10\|1209 | 56\|16097 | 0.00819 | 0.04917 | ENSMUS | Sox9/Hspa9/Ncoa3/Stat3/Notch1/Rest/Ptn/Nfe2l2/Ythdf2/Hes1 |
| BP | GO:000688 | ER to Golgi vesicle-mediated transport | 17\|1209 | 120\|16097 | 0.00824 | 0.04937 | ENSMUS | Hyou1/Insig1/Sec24d/Sec23a/Kdelr2/Gosr1/Copb2/Tbc1d20/Se |
| BP | GO:003476 | negative regulation of transmembrane transport | 17\|1209 | 120\|16097 | 0.00824 | 0.04937 | ENSMUS | Gem/Thbs1/Myc/Rgs2/Ppp2ca/Slc30a1/Arl6ip5/Actn2/Nedd4/F |
| BP | GO:002240 | molting cycle process | 15\|1209 | 101\|16097 | 0.00826 | 0.04939 | ENSMUS | Igfbp5/Sox9/Inhba/Cd109/Fgfr2/Egfr/Rela/Notch1/Lgr5/Ptgs2/L |
| BP | GO:002240 | hair cycle process | 15\|1209 | 101\|16097 | 0.00826 | 0.04939 | ENSMUS | Igfbp5/Sox9/Inhba/Cd109/Fgfr2/Egfr/Rela/Notch1/Lgr5/Ptgs2/L |

| Count | Up | Up_Gene_i | Down | Down_Gene_id |
|---|---|---|---|---|
| 16 | 4 | ENSMUSG0 | 12 | ENSMUSG00000000567/ENSMUSG00000041324/ENSMUSG00000031785/ENSMUSG00000056758/ENSMUSG00000027996/ENSMUSG00000070645/ENSMUSG0 |
| 16 | 6 | ENSMUSG0 | 10 | ENSMUSG00000003541/ENSMUSG00000020349/ENSMUSG00000020484/ENSMUSG00000031451/ENSMUSG00000002052/ENSMUSG00000047281/ENSMUSG0 |
| 6 | 3 | ENSMUSG0 | 3 | ENSMUSG00000048616/ENSMUSG00000007613/ENSMUSG00000026836 |
| 6 | 2 | ENSMUSG0 | 4 | ENSMUSG00000006800/ENSMUSG00000020458/ENSMUSG00000020484/ENSMUSG00000056501 |
| 6 | 2 | ENSMUSG0 | 4 | ENSMUSG00000006800/ENSMUSG00000020458/ENSMUSG00000020484/ENSMUSG00000056501 |
| 37 | 12 | ENSMUSG0 | 25 | ENSMUSG00000021670/ENSMUSG00000041324/ENSMUSG00000031565/ENSMUSG00000029482/ENSMUSG00000056758/ENSMUSG00000039004/ENSMUSG0 |
| 20 | 7 | ENSMUSG0 | 13 | ENSMUSG00000005413/ENSMUSG00000038418/ENSMUSG00000009376/ENSMUSG00000032724/ENSMUSG00000024927/ENSMUSG00000018476/ENSMUSG0 |
| 9 | 2 | ENSMUSG0 | 7 | ENSMUSG00000000567/ENSMUSG00000039004/ENSMUSG00000019997/ENSMUSG00000007613/ENSMUSG00000024927/ENSMUSG00000034205/ENSMUSG0 |
| 9 | 4 | ENSMUSG0 | 5 | ENSMUSG00000030849/ENSMUSG00000015846/ENSMUSG00000029287/ENSMUSG00000048616/ENSMUSG00000007613 |
| 8 | 3 | ENSMUSG0 | 5 | ENSMUSG00000030536/ENSMUSG00000026596/ENSMUSG00000005533/ENSMUSG00000023951/ENSMUSG00000074934 |
| 8 | 4 | ENSMUSG0 | 4 | ENSMUSG00000000567/ENSMUSG00000048616/ENSMUSG00000007613/ENSMUSG00000026836 |
| 29 | 7 | ENSMUSG0 | 22 | ENSMUSG00000006800/ENSMUSG00000027313/ENSMUSG00000020917/ENSMUSG00000030657/ENSMUSG00000024359/ENSMUSG00000038615/ENSMUSG0 |
| 15 | 8 | ENSMUSG0 | 7 | ENSMUSG00000041959/ENSMUSG00000009376/ENSMUSG00000031778/ENSMUSG00000019997/ENSMUSG00000031523/ENSMUSG00000007613/ENSMUSG0 |
| 15 | 7 | ENSMUSG0 | 8 | ENSMUSG00000000567/ENSMUSG00000041324/ENSMUSG00000046186/ENSMUSG00000030849/ENSMUSG00000024927/ENSMUSG00000032487/ENSMUSG0 |
| 11 | 4 | ENSMUSG0 | 7 | ENSMUSG00000040152/ENSMUSG00000003032/ENSMUSG00000000753/ENSMUSG00000027351/ENSMUSG00000029287/ENSMUSG00000039384/ENSMUSG0 |
| 30 | 5 | ENSMUSG0 | 25 | ENSMUSG00000030657/ENSMUSG00000042042/ENSMUSG00000034832/ENSMUSG00000030062/ENSMUSG00000032997/ENSMUSG00000017929/ENSMUSG0 |
| 14 | 3 | ENSMUSG0 | 11 | ENSMUSG00000042688/ENSMUSG00000027954/ENSMUSG00000026259/ENSMUSG00000002489/ENSMUSG00000028613/ENSMUSG00000020745/ENSMUSG0 |
| 25 | 3 | ENSMUSG0 | 22 | ENSMUSG00000027995/ENSMUSG00000037411/ENSMUSG00000032501/ENSMUSG00000051439/ENSMUSG00000046062/ENSMUSG00000031668/ENSMUSG0 |
| 18 | 6 | ENSMUSG0 | 12 | ENSMUSG00000041324/ENSMUSG00000024359/ENSMUSG00000023951/ENSMUSG00000058070/ENSMUSG00000033454/ENSMUSG00000035992/ENSMUSG0 |
| 18 | 5 | ENSMUSG0 | 13 | ENSMUSG00000032193/ENSMUSG00000027995/ENSMUSG00000037411/ENSMUSG00000024789/ENSMUSG00000030341/ENSMUSG00000056501/ENSMUSG0 |
| 12 | 4 | ENSMUSG0 | 8 | ENSMUSG00000027995/ENSMUSG00000003032/ENSMUSG00000024789/ENSMUSG00000027832/ENSMUSG00000022037/ENSMUSG00000032487/ENSMUSG0 |
| 12 | 4 | ENSMUSG0 | 8 | ENSMUSG00000031565/ENSMUSG00000027678/ENSMUSG00000056758/ENSMUSG00000031778/ENSMUSG00000023951/ENSMUSG00000037815/ENSMUSG0 |
| 31 | 14 | ENSMUSG0 | 17 | ENSMUSG00000027995/ENSMUSG00000027954/ENSMUSG00000026259/ENSMUSG00000002489/ENSMUSG00000028613/ENSMUSG00000045983/ENSMUSG0 |
| 19 | 6 | ENSMUSG0 | 13 | ENSMUSG00000040152/ENSMUSG00000027954/ENSMUSG00000031668/ENSMUSG00000015846/ENSMUSG00000026728/ENSMUSG00000033499/ENSMUSG0 |
| 10 | 5 | ENSMUSG0 | 5 | ENSMUSG00000030849/ENSMUSG00000015846/ENSMUSG00000029287/ENSMUSG00000048616/ENSMUSG00000007613 |
| 10 | 3 | ENSMUSG0 | 7 | ENSMUSG00000022346/ENSMUSG00000000567/ENSMUSG00000031565/ENSMUSG00000028063/ENSMUSG00000030849/ENSMUSG00000023951/ENSMUSG0 |
| 10 | 3 | ENSMUSG0 | 7 | ENSMUSG00000000567/ENSMUSG00000024359/ENSMUSG00000027678/ENSMUSG00000004040/ENSMUSG00000029249/ENSMUSG00000015839/ENSMUSG0 |
| 17 | 3 | ENSMUSG0 | 14 | ENSMUSG00000032115/ENSMUSG00000045294/ENSMUSG00000039234/ENSMUSG00000020986/ENSMUSG00000079111/ENSMUSG00000010392/ENSMUSG0 |
| 17 | 6 | ENSMUSG0 | 11 | ENSMUSG00000028214/ENSMUSG00000040152/ENSMUSG00000022346/ENSMUSG00000026360/ENSMUSG00000020349/ENSMUSG00000037434/ENSMUSG0 |
| 15 | 7 | ENSMUSG0 | 8 | ENSMUSG00000000567/ENSMUSG00000041324/ENSMUSG00000046186/ENSMUSG00000030849/ENSMUSG00000024927/ENSMUSG00000032487/ENSMUSG0 |
| 15 | 7 | ENSMUSG0 | 8 | ENSMUSG00000000567/ENSMUSG00000041324/ENSMUSG00000046186/ENSMUSG00000030849/ENSMUSG00000024927/ENSMUSG00000032487/ENSMUSG0 |

0000007613/ENSMUSG00000030059/ENSMUSG00000027465/ENSMUSG00000074656/ENSMUSG00000040268/ENSMUSG00000031548
0000109511/ENSMUSG00000032487/ENSMUSG00000026604/ENSMUSG00000024491

0000002489/ENSMUSG00000024789/ENSMUSG00000070645/ENSMUSG00000020889/ENSMUSG00000026796/ENSMUSG00000030059/ENSMUSG00000028161/ENSMUSG00000018634/ENSMUSG0000003
0000024423/ENSMUSG00000040268/ENSMUSG00000015839/ENSMUSG00000028211/ENSMUSG00000030287/ENSMUSG00000037902/ENSMUSG00000004207
0000074934

0000031668/ENSMUSG00000093930/ENSMUSG00000032997/ENSMUSG00000025153/ENSMUSG00000036356/ENSMUSG00000028521/ENSMUSG00000039662/ENSMUSG00000018339/ENSMUSG0000002
0000026812
0000005871/ENSMUSG00000034593
0000031393
0000024413/ENSMUSG00000036356/ENSMUSG00000043587/ENSMUSG00000028521/ENSMUSG00000004846/ENSMUSG00000045216/ENSMUSG00000020205/ENSMUSG00000023068/ENSMUSG0000002
0000044147/ENSMUSG00000024431/ENSMUSG00000031393/ENSMUSG00000025326/ENSMUSG00000025372
0000025408/ENSMUSG00000039004/ENSMUSG00000028525/ENSMUSG00000031778/ENSMUSG00000020484/ENSMUSG00000024789/ENSMUSG00000020889/ENSMUSG00000026361/ENSMUSG0000002
0000029249/ENSMUSG00000015839/ENSMUSG00000040025/ENSMUSG00000031548/ENSMUSG00000078671/ENSMUSG00000037902
0000037820/ENSMUSG00000050965/ENSMUSG00000032487/ENSMUSG00000016024/ENSMUSG00000035356/ENSMUSG00000056071/ENSMUSG00000022808
0000029361/ENSMUSG00000037902
0000069895/ENSMUSG00000045394
0000020745/ENSMUSG00000039953/ENSMUSG00000050965/ENSMUSG00000079037/ENSMUSG00000025582/ENSMUSG00000032593/ENSMUSG00000039952/ENSMUSG00000044147/ENSMUSG0000003
0000030738/ENSMUSG00000024423/ENSMUSG00000022037/ENSMUSG00000015839/ENSMUSG00000040025/ENSMUSG00000022412/ENSMUSG00000021838

0000096054
0000040025
0000032458/ENSMUSG00000027465/ENSMUSG00000026924/ENSMUSG00000056050/ENSMUSG00000036391/ENSMUSG00000032096/ENSMUSG00000002741/ENSMUSG00000031543
0000035199/ENSMUSG00000050965/ENSMUSG00000018634/ENSMUSG00000029361/ENSMUSG00000035640
0000005871/ENSMUSG00000034593
0000005871/ENSMUSG00000034593

33717/ENSMUSG00000029249/ENSMUSG00000029361/ENSMUSG00000031548/ENSMUSG00000035828/ENSMUSG00000020684/ENSMUSG00000021794/ENSMUSG00000035621/ENSMUSG00000027712/E

29201/ENSMUSG00000030747/ENSMUSG00000053398/ENSMUSG00000029009/ENSMUSG00000028179/ENSMUSG00000039497/ENSMUSG00000032350/ENSMUSG00000015839

21978/ENSMUSG00000038296/ENSMUSG00000029201/ENSMUSG00000020346/ENSMUSG00000024486/ENSMUSG00000039497/ENSMUSG00000048920/ENSMUSG00000028277/ENSMUSG00000039357/E

26728/ENSMUSG00000056501/ENSMUSG00000037447/ENSMUSG00000024927/ENSMUSG00000035673/ENSMUSG00000050965/ENSMUSG00000016024/ENSMUSG00000037902

31393/ENSMUSG00000025326/ENSMUSG00000025372

ENSMUSG00000022808/ENSMUSG00000034593

ENSMUSG00000024807/ENSMUSG00000034951
